# Social media use and adolescent health-risk behaviours: A systematic review and meta-analysis

**DOI:** 10.1101/2023.02.24.23286415

**Authors:** Ms Amrit Kaur Purba, Dr Rachel M Thomson, Dr Paul M Henery, Dr Anna Pearce, Professor Marion Henderson, Professor S Vittal Katikireddi

## Abstract

**Objectives:** To examine the association between social media (SM) use and health-risk behaviours: alcohol/drug/tobacco/electronic nicotine delivery system (ENDS) use, unhealthy dietary behaviour, inadequate physical activity, gambling, anti-social, sexual risk, and multiple risk behaviours in adolescents aged 10-19 years.

**Design:** Systematic review and meta-analysis.

**Data sources:** Embase, MEDLINE, APA PsycINFO, SocINDEX, CINAHL, SSRN, SocArXic, PsyArXiv, MedRxiv, and Google Scholar (01/1997-06/2022).

**Methods:** Included studies reported a SM exposure (time spent, frequency of use, exposure to health-risk behaviour content or other SM activities) and ≥1 relevant outcome. Screening and risk of bias (RoB) assessments were completed independently by two reviewers. Synthesis without meta-analysis (SWiM) based on effect direction and random-effects meta-analyses were used. Effect modification was explored using meta-regression and stratification. Certainty of evidence was assessed using GRADE (Grading of Recommendations, Assessment, Development and Evaluations).

**Results:** Of 17,077 studies screened, 126 were included (76 meta-analysed). The final sample included 1,431,534 adolescents (mean age:15.0 years).

SWiM indicated harmful associations between SM and all health-risk behaviours in most included studies, except inadequate physical activity where beneficial associations were reported in 63.6% of studies. Frequent (vs infrequent) SM use was associated with increased alcohol consumption (OR 1.48, 1.32 to 1.62; n=383,670), drug use (1.28, 1.05 to 1.56; n=117,646), tobacco use (1.78, 1.45 to 2.19; n=424,326), sexual risk (1.78, 1.49 to 2.13; n=47,325), anti-social behaviour (1.73, 1.44 to 2.06; n=54,993), multiple risk behaviours (1.75, 1.30 to 2.35; n=43,571), and gambling (2.84, 2.04 to 3.97; n=26,537). Exposure to health-risk behaviour content on SM (vs no exposure) was associated with increased odds of ENDS use (1.73, 1.34 to 2.23; n=721,322), unhealthy dietary behaviour (2.12, 1.87 to 2.39; n=9424), and alcohol consumption (2.43, 1.25 to 4.71; n=14,731). For alcohol consumption, stronger associations were identified for exposure to user-generated content (3.21, 2.37 to 4.33) vs marketer-generated content (2.18, 0.96 to 4.97). For time spent on SM, use for ≥2hrs/day (vs <2hrs) increased odds of alcohol consumption (2.13, 1.56 to 2.92; n=12,390). GRADE certainty was moderate for unhealthy dietary behaviour, low for alcohol use and very low for other investigated outcomes.

**Conclusions:** Social media use is associated with adverse adolescent health-risk behaviours, but further high quality research is needed to establish causality, understand effects on health inequalities, and determine which aspects of social media are most harmful. Given the pervasiveness of social media, efforts to understand and reduce the potential risks adolescents face may be warranted.

**Funding, competing interests, data sharing:** Funded by the Medical Research Council, Chief Scientist Office, NHS Research Scotland and the Wellcome Trust. All authors declare no competing interests. Template data forms, data extracted, and data analysed are available from the corresponding author on request.

**Systematic review registration:** PROSPERO: CRD42020179766.

## Introduction

Social media (SM) has revolutionised the communication landscape, with approximately 139 million adolescents using Instagram and 120.2 million using Facebook globally in 2022.^1, 2^ It can be defined as websites and applications which host numerous user activities including the creation and sharing of content, social networking, and microblogging. SM’s diverse and inherently social nature has supported adolescents’ need for autonomy, social connectedness, and relatedness.^3–6^ Recognised by the World Health Organisation as a powerful medium to promote health, the use of SM to elicit positive behaviour change is well documented, including increased physical activity, and healthy diets through increased interaction, increased accessibility to health information, and peer/social/emotional support.^7–9^

Despite its ubiquitous use and potential benefits, harmful effects on adolescent health-risk behaviours (e.g., substance use, sexual risk behaviour) at least partly due to aggravated peer-pressure and social norms are possible.^3, 10, 11^ Numerous pathways may exist between SM and health-risk behaviours (Figure-1). SM use might displace more traditional in-person interactions, thereby increasing physical inactivity. It can host marketer-generated (e.g., advertisements, influencers),^12–16^ and user-generated (e.g., user and peer posts) content displaying consumption of unhealthy commodities.^17, 18^ Exposure to such content on traditional media has been shown to influence adolescent health-risk behaviours (including substance use, unhealthy diet),^19, 20^ with experimental and longitudinal research suggesting online content also influences behaviours offline.^21–26^

**Figure-1.**
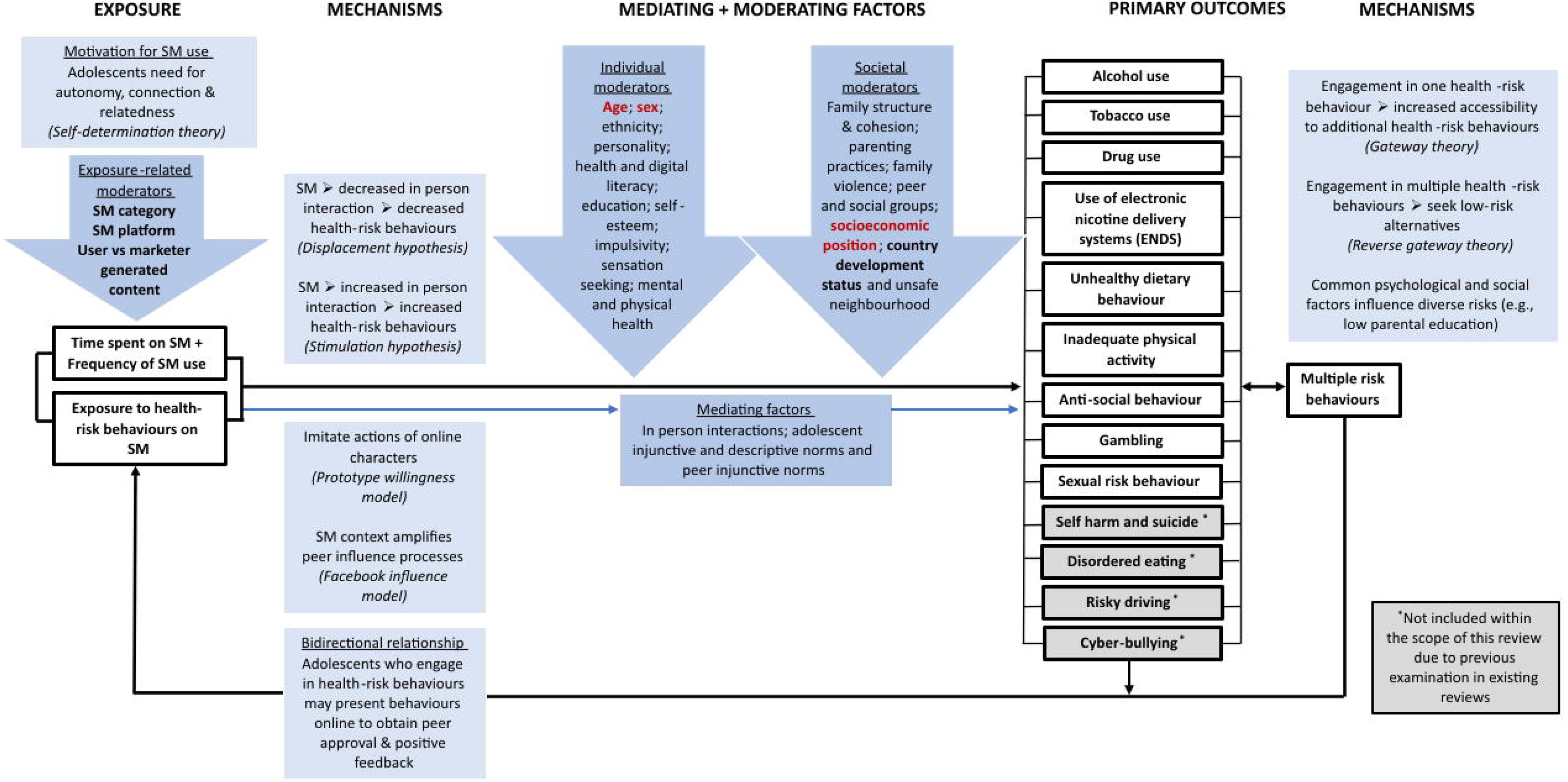
Logic model illustrating the pathways between social media and adolescent health risk behaviours. Variables considered important potential confounders in this study are indicated in red and were selected a priori by the researchers’ following expert advisory group consultation and identification of variables considered key confounders in the literature. Variables deemed potential effect modifiers for exploration in this study are indicated in bold. Abbreviations: SM = Social media.

Adolescence is a sensitive period for the adoption of lifelong behaviours – health consequences are therefore potentially immediate and lifelong.^27–29^ Immediate consequences include (but are not limited to) alcohol/drug-related injury, low educational attainment and depression (for alcohol and drug use), and sexually transmitted diseases and teenage pregnancy (for sexual risk behaviour).^30^ Yet, these represent relatively extreme outcomes and for most adolescents these behaviours, if experimental and short-lived, will have limited harms and can be considered a normal part of adolescent development. However some health behaviours, such as poor diet, inadequate physical activity and alcohol consumption, can be set in adolescence and carry lifelong consequences. ^27–29^ For anti-social behaviour, shown to be associated with adverse consequences such as criminality and psychosocial malfunctioning, the long-term effects extend to causing significant distress for others, emphasising the public health relevance of this risk behaviour.^31^

Existing reviews focus on university/college populations (and are therefore not representative of all adolescents); assess SM under the broad scope of digital media/internet use; do not assess risk of bias; and examine few health-risk behaviours (namely substance use and sexual risk behaviour).^20, 32–36^ Differential effects by socioeconomic position (SEP), specifically whether more disadvantaged groups are more susceptible to harm from SM, consequently resulting in a widening health inequalities, and between high and low-middle income countries have also not been explored.^23, 37–39^ Prior research investigating SM’s influence on adolescent mental health suggests age and sex differences, where greater negative effects exist for females and younger adolescents (compared to males and older adolescents), yet these potential differences are yet to be examined in relation to health-risk behaviours.^40, 41^ The most recent review by Vannucci et al. explored SMs association with adolescent substance use and risky sexual behaviour. The review’s synthesis of electronic media use (defined as electronic media with a direct component involving social interactions with others (2022 personal communication with A. Vannucci)) with SM and reliance on pooled correlations inhibits any explicit conclusions about the magnitude of associations resulting from SM use specifically.^42^ Due to the high risk of confounding and reverse causation in studies in this area (which largely rely on observational data) it is also important to assess the quality of evidence, which has been limited in other reviews.^32, 42^

Given the above, we aim to systematically review the evidence on SM use and adolescent health-risk behaviours, addressing the following objectives:

1. Explore how SM use is measured in studies examining its relationship with adolescent health-risk behaviours (alcohol, drug, tobacco, electronic nicotine delivery system (ENDS) use, unhealthy dietary behaviour, inadequate physical activity, gambling, anti-social behaviour, sexual risk behaviour, and multiple risk behaviours)
2. Investigate the association between time spent on SM and frequency of use on adolescent health-risk behaviours
3. Explore the association between exposure to health-risk behaviour content on SM and adolescent health-risk behaviours, and if any relationship differs by content viewed (user/marketer-generated)
4. Investigate if any relationships differ by SM platform/category used, age, sex, SEP, and development status of study setting
5. Evaluate the certainty of evidence using Grading of Recommendations Assessment, Development, and Evaluation (GRADE)

## Methods

We follow the Preferred Reporting Items for Systematic Reviews and Meta-Analyses (PRISMA) and Synthesis Without Meta-analysis (SWiM) reporting guidance.^43, 44^ We published a pre-specified protocol, including a logic model (Figure-1; further background in protocol^45, 46^) which was used to identify important confounders and effect modifiers. This study is registered with PROSPERO, CRD42020179766).^45^ Protocol deviations are reported in Appendix-1.

### Search methods for identification of studies

Embase, MEDLINE, APA PsycINFO, SocINDEX, CINAHL, SSRN, SocArXic, PsyArXiv, and MedRxiv were searched from 1^st^ January 1997 (first recognisable SM site ‘Six Degrees’ launched) to 6^th^ June 2022, using a comprehensive strategy developed with an information scientist (Appendix-2). We scrutinised the first thirty hits in Google Scholar, reference lists of included studies and relevant systematic reviews were screened, and subject experts contacted to identify additional, planned, ongoing, or unpublished studies. Filters for study types and geographical location/language limits were not applied.^47^ It was not possible to translate non-English language studies; these are reported in Appendix-3.^47^

### Study inclusion and exclusion criteria

The precise age range adolescence encompasses is debated. Following the World Health Organisation’s definition ^48, 49^ our population of interest was aged 10-19 years inclusive. Studies focussing on college or university participants (of all ages) were excluded due to the differing nature of SM use and health-risk behaviours in these groups. Studies including some non-college/university participants alongside college/university participants were included if relevant data on non-college/university participants could be extracted.^13, 22, 50^ The exposure of interest was use of any SM category in the SAGE Social Media Categorisation^51^ (social networking, microblogging, media-sharing, geo-location-based, bookmarking, social news, collaborative authoring sites, web conferencing and scheduling and meeting) (Appendix-4). Online (social) gambling (e.g., simulated gambling via Facebook) and online (social) gaming were eligible due to their inclusion of core SM functionalities, namely user interaction.^52–54^ SM dating platforms were excluded as most are restricted to users ≥18 years.^55–57^

SM exposures were classified into time spent (e.g., hours/day), frequency of use (e.g., daily, weekly, or general use), exposure to health-risk behaviour content (e.g., alcohol advertising on Facebook), and other SM activities (e.g., strategies to manage online presence). The process used to classify the SM category, platform, and type of health-risk behaviour content (user-generated/ marketer-generated) of reported exposures is provided in Appendix-4.

The comparator group was those with no or differing levels of time spent/frequency of use/exposure.

Outcome selection was guided by preliminary evidence,^58^ the logic model (Figure-1), and an advisory group (Appendix-5).^59^ Eligible outcomes were alcohol, drug, tobacco, electronic nicotine delivery systems (ENDS) use, sexual risk behaviour, gambling (not via SM, e.g., lottery, scratch cards), unhealthy dietary behaviour, inadequate physical activity, anti-social behaviour, and multiple risk behaviours (≥2 of the aforementioned behaviours) (Appendix-6).

Studies reporting quantitative data from primary research were eligible.

### Selection of studies

Records were de-duplicated in Mendeley^60^ and imported to Covidence^61^ for screening. Eligibility criteria were piloted on 100 studies and all titles/abstracts and full-texts were independently screened by AKP and a second reviewer (PMH, RT, AP, and MH), with conflicts resolved via consensus and/or discussion with a third reviewer (SVK). Where eligible studies contained overlapping or duplicate data, a set of decision rules (Appendix-7) considered alignment with our Population/Exposure/Comparator/Outcome criteria to select unique data for synthesis.

### Data extraction and risk of bias (RoB) assessment

Data were extracted in Excel (version 2025) by AKP and checked by a second reviewer (PMH, RT, AP, and MH) (Appendix-8). RoB assessment was conducted independently at datapoint/outcome level by AKP and a second reviewer using an adapted version of the Newcastle-Ottawa Scale for cross-sectional and cohort studies (NOS),^62^ and the Cochrane RoB-2 tool for randomised studies.^63^ The NOS was adapted to incorporate insights from the Cochrane ROBINS-I RoB tool, with assistance from GRADE Public Health Group members.^64^ This included assessing adjustment for pre-identified critical confounding domains (e.g., sex, age, and any measure of socioeconomic position (SEP) – e.g. parental academic qualifications), other justifiable confounders, attrition and missing data (Appendix-9). Conflicts were resolved via consensus and/or discussion with a third reviewer (SVK).

The RoB assessments informed data synthesis and certainty, assessed using GRADE.^59^

### Data synthesis

#### Synthesis without meta-analysis (SWiM)

Within SWiM effect direction was coded as beneficial or harmful for each outcome at the study level, with findings categorised as inconsistent if <70% of extracted datapoints reported a consistent effect direction.^44, 65^ As per Cochrane guidance, statistical significance was not taken into account.^66^ Sign tests assessed evidence of effect where there were ≥3 studies within a synthesis. Modified effect direction plots (created using *RStudio.V1.2.5.*^67^), displaying RoB results, were produced.^65^

#### Primary meta-analyses

Meta-analyses were performed by outcome for time spent on SM, frequency of SM use, and exposure to health-risk behaviour content, but not for other SM activities due to heterogeneity. Given anticipated heterogeneity in study designs, settings and measures, we used random-effect models, using the DerSimonian and Laird estimator.^68^ Heterogeneity was assessed with the I^2^ statistic.^69^ Continuous exposures (exposures assessed on a continuous scale) were analysed separately from binary exposures (Appendix-10).^69^ For binary exposures, odds ratios (ORs) were estimated.^69^ For continuous exposures, data were pooled to produce standardised beta coefficients (Std. Beta) or standardised mean differences (SMDs) for continuous outcomes and ORs for binary outcomes.

Where ≥10 studies were included in a meta-analysis, meta-regression explored heterogeneity by the following characteristics identified *a priori*: health-risk behaviour content viewed on SM (user vs marketer-generated), SM category (e.g., social networking), SM platform (e.g., Facebook), sex, average SEP of participants, development status of study setting (high vs low-middle income country (HIC vs LMIC)),^70^ and average age of participants (<16 vs ≥16 years, as existing evidence demonstrates risk behaviours tend to peak at age 16 years and the majority become acceptable (albeit not necessarily legal) from a societal perspective).^71^ Statistical analysis was performed using *Stata.V16.*^72^

#### Subgroup/sensitivity analyses

We stratified meta-analyses by the above characteristics if at least one subgroup had ≥2 studies and investigated potential bias by examining results by study design (cross-sectional vs cohort/randomised control trial (RCT)); adjustment for pre-identified critical confounding domains (age, sex, and SEP); RoB; and excluding datapoints with samples containing individuals outside our eligible age-range (10-19 years).

#### Publication bias

Publication bias/small study effects were assessed using funnel plots and the Egger’s test when ≥10 studies were meta-analysed.^73, 74^

### Certainty of the evidence

Certainty was assessed using GRADE,^59^ which combines information on RoB, imprecision, inconsistency, indirectness, and publication bias.^59^ As per GRADE, advisory group members ranked the importance of outcomes via an online survey (Appendix-5), and certainty for the top seven ranked outcomes (alcohol, drug, tobacco, ENDS use, sexual risk behaviour, gambling, and multiple risk behaviours) assessed using a four-category system (very low-high).^59^ Observational evidence automatically started at low with the ability to upgrade/downgrade.^59, 75^

### Patient and public involvement

Advisory group members included policy, non-governmental, and academic stakeholders who provided guidance during protocol development and the review stages (Appendix-5). Public and policymaker-facing summaries will be co-produced with additional public representatives, and advisory group members.

## Results

### Description of studies

Of 17,077 studies screened, 688 full-text studies were assessed, with 126 included (76 meta-analysed) (Figure-2). The final sample included 1,431,534 adolescents (mean age:15.0 years). Most included studies were cross-sectional (n=99; 78.6%), and investigated high-income countries (n=113; 89.7%),^70^ with 44 studies investigating US adolescents. Appendix-11 shows the geographical distribution of included study populations. Included and excluded study characteristics are presented in Appendix-11 and 12.

**Figure-2.**
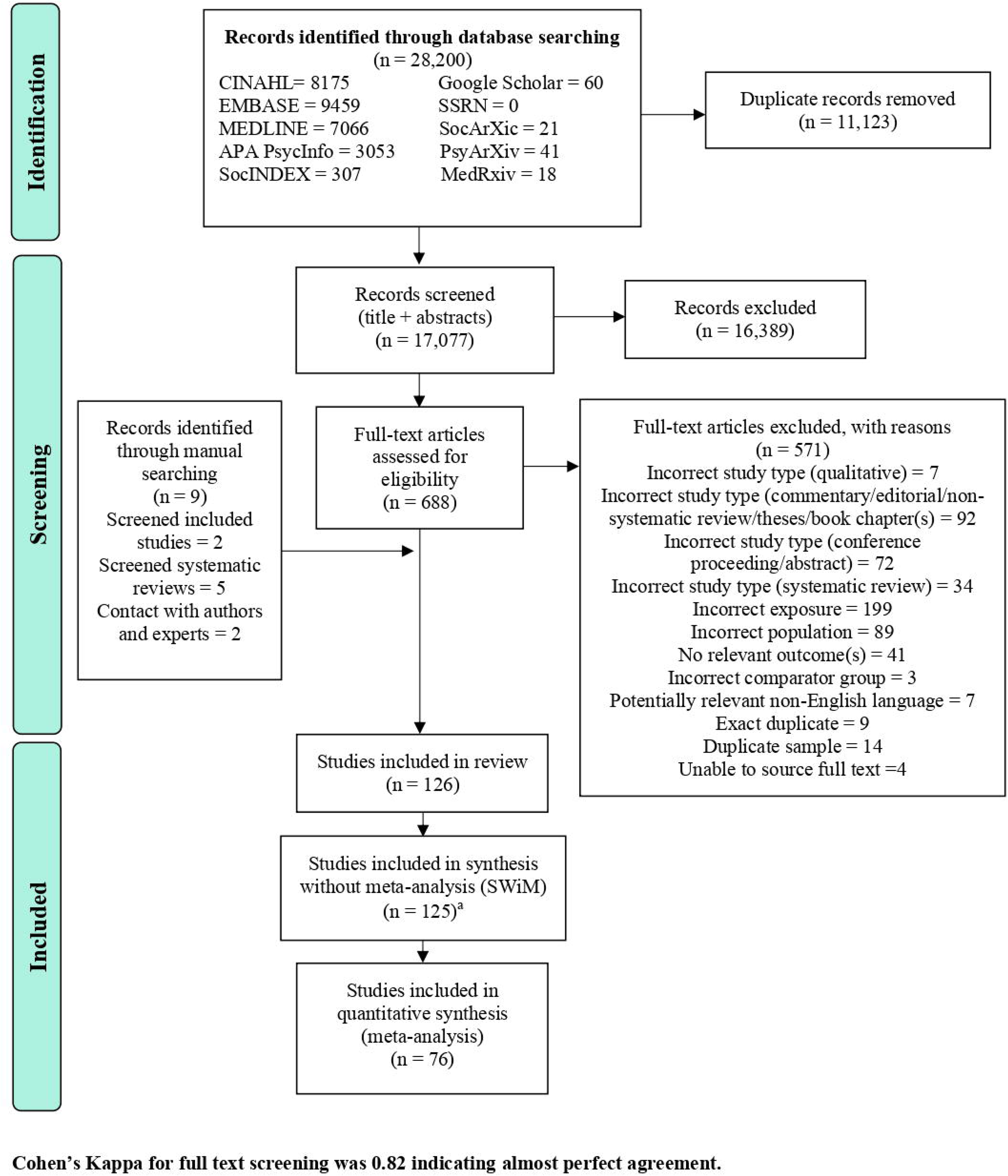
PRISMA flow diagram. ^a^ One study^89^ was not included in the synthesis without meta-analysis (SWiM) as this resulted in double counting of study participants; we were able to include estimates from this study in meta-analyses stratified by outcome where this issue did not occur. Abbreviations: APA = American Psychological Association; CINAHL = Cumulative Index to Nursing and Allied Health Literature; EMBASE = Excerpta Medica dataBASE; MEDLINE = Medical Literature Analysis and Retrieval System Online; and SSRN = Social Science Research Network.

For included cross-sectional and cohort studies (n=122), 46.7% (n=57) of studies were graded high RoB, 25.4% (n=31) moderate, and 27.9% (n=34) low. Of the four RCTs included, two were graded some concerns and two low RoB (Appendix-13). Reviewer RoB agreement was strong (κ=0.91).^76^

### Social media (SM) measures reported in included studies

Within included studies, many SM exposure measures were reported, with most investigating multiple measures (Appendix-14). All were incorporated in our exploration of how SM use is measured, therefore the number of datapoints reported differs across syntheses.

In total, 253 SM measures were reported: 53.4% (n=135) assessed frequency, 24.1% (n=61) exposure to health-risk behaviour content, 17.8% (n= 45) time spent, and 4.74% (n=12) other SM activities. Despite our broad definition of SM, most included studies assessed a narrow range of SM categories (or themselves adopted a broad definition). Social networking sites was the most common category investigated (55.7%; n=141). Of those SM measures investigating a specific platform (n=86), Facebook was most commonly investigated (n=40), followed by Twitter (n=10).

Of those measures assessing exposure to health-risk behaviour content, 59.0% (n=36) assessed marketer-generated, 26.2% (n=16) assessed user-generated content, and 14.8% (n=9) assessed both types of content. In total, 134 of the 253 SM measures provided sufficient information to differentiate between active (e.g., positing and commenting on posts) (n=90) and passive (e.g., observing others, content or watching videos) (n=44) use. Exposure ascertainment primarily used unvalidated adolescent self-report surveys (n=221) with a minority using data-driven codes, validated adolescent self-report questionnaires and/or clinical records (n=32).

### Social media (SM) use and health-risk behaviours

#### Alcohol use

Alcohol use was the most extensively studied outcome (Appendix-15). For time spent, 15/16 studies (93.8%) reported harmful associations (95% CI 71.7 to 98.9%; n=100,354; sign test p<0.001), 16/17 studies (94.1%) for frequency (73.0 to 99.0%; n=391,445; sign test p<0.001), and 11/12 studies (91.7%) for exposure to health-risk behaviour content (64.6 to 98.5%; n=24,451; sign test p=0.006). Other SM activities (had a Facebook account) was investigated by one study which reported a harmful association (20.7 to 100%; n=4485) (Figure-3 for effect direction plot).

**Figure-3.**
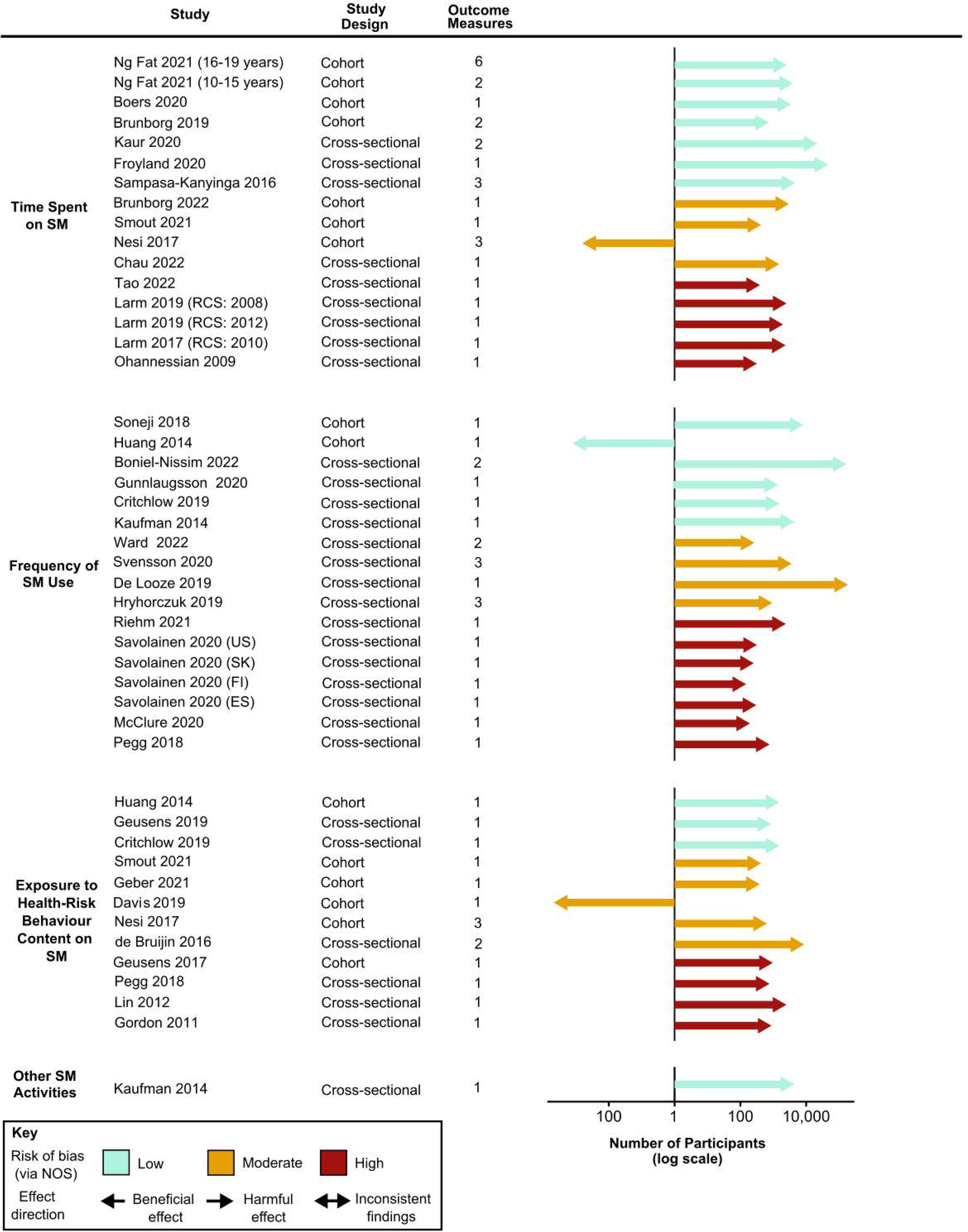
Effect direction plot for studies of the association between social media use and adolescent alcohol use, by social media exposure. Arrow size indicates sample size; arrow colour indicates study risk of bias. Sample size: represented by the size of the arrow, measured on a log scale. Outcome measure: number of outcome measures synthesised within each study. Studies organised by risk of bias grade, study design, and year of publication. Repeat cross-sectional studies, multiple study populations from different countries, and age subsets originating from the same study reported as separate studies. Abbreviations: ESP = Spain; FIN = Finland; KOR = South Korea; NOS = Assessed via adapted Newcastle Ottawa Scale; RCS = Repeat cross-sectional study; SM = Social media and USA = United States.

In meta-analyses, frequent/daily (vs infrequent/non-daily) SM use was associated with increased alcohol consumption (OR=1.48,1.36 to 1.62; I^2^=40.5%; n=383,670) (Figure-4A). In stratified analyses (Appendix-16, p162-166), effect sizes were larger for adolescents ≥16 years (1.80, 1.46 to 2.22 vs 1.35, 1.26 to 1.44 for those <16 years; p<0.01 for test of differences). Social networking sites were associated with increased alcohol consumption, whilst microblogging or media-sharing sites had an unclear relationship (p=0.03).

**Figure-4.**
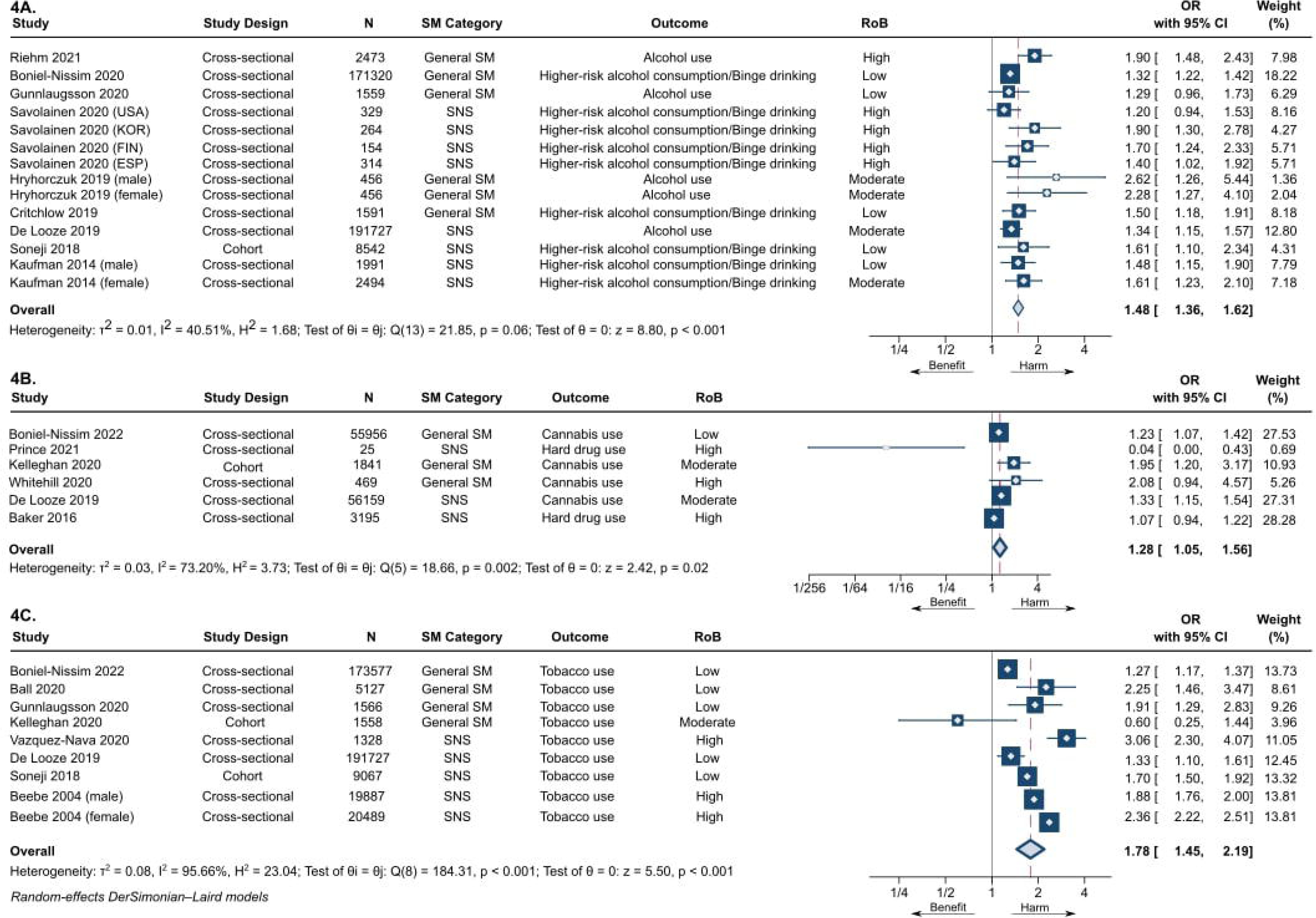
Forest plots for association between frequency of social media use and A) alcohol use, B) drug use, and C) tobacco use. Figure 4A presents forest plot for binary exposure (frequent/daily vs infrequent/non-daily) & binary/continuous alcohol use outcome meta-analysis, with odds ratio (OR) used as common metric. Total number of study participants = 383,670. Figure 4B presents forest plot for binary exposure (frequent/daily vs infrequent/non-daily) & binary/continuous drug use outcome meta-analysis, with odds ratio (OR) used as common metric. Total number of study participants = 117,645. Figure 4C presents forest plot for binary exposure (frequent vs infrequent) & binary/continuous tobacco use outcome meta-analysis, with odds ratio (OR) used as common metric. Total number of study participants = 424,326. Abbreviations: CI = Confidence interval; ESP = Spain; FIN = Finland; KOR = South Korea; N = Number of study participants; OR = Odds ratio; RoB = Risk of bias; SM = Social media and SNS = Social networking sites.

SM use for ≥2 hrs (vs <2 hrs/day) was associated with increased alcohol consumption (OR=2.13, 1.56 to 2.92; I^2^=81.6%; n=12,390), as was exposure (vs unexposed) to health-risk behaviour content (2.43, 1.25 to 4.71; I^2^=98.0%; n=14,731) (Appendix-16, p 167). Stratified analyses for time spent and exposure to health-risk behaviour content generally did not reveal important differences by age and SM category (Appendix-16, p 168-170). Associations were slightly stronger for exposure to health-risk behaviour content in user-generated (3.21, 2.37 to 4.33) vs marketer-generated content (2.18, 0.96 to 4.97; p=0.39) (Appendix-16, p 171). Meta-analyses for frequency, time spent and exposure to health-risk behaviour content (assessed on a continuous scale) demonstrated similar findings (Appendix-16, p 172-173). On stratification (Appendix-16, p 174-179), for exposure to health-risk behaviour content, associations were larger for adolescents ≥16 years (Std.Beta 0.35, 0.29 to 0.42 vs 0.09, 0.05 to 0.13 for those <16 years; p<0.001), indicating for every one standard deviation (SD) increase in exposure to health-risk behaviour content, alcohol consumption increased by 0.35 SD for older compared to 0.13 SD for younger adolescents.

#### Drug use

For drug use, across all exposures investigated, 86.6% of studies (n=13/15; 53.3% low/moderate RoB) reported harmful associations (Appendix-16, p 180). The pooled OR for frequent/daily use (vs infrequent/non-daily) was 1.28 (1.05 to 1.56, I^2^=73.2%; n=117,646) (Figure-4B). Stratification showed no clear differences (Appendix-16, p 182-184). Few studies (n=3) assessed time spent, with estimates suggestive of harm (OR=1.58, 0.91 to 2.75; I^2^=88.1%; n=7357 for ≤1hrs vs >1hr/day) (Appendix-16, p 185).

#### Tobacco use

For tobacco use, 88.9% (n=16/18; 50.0% low RoB) studies reported harmful associations of SM use (Appendix-16, p 186). Frequent (vs infrequent) use was associated with increased tobacco use (OR=1.78, 1.45 to 2.19; I^2^=95.7%; n=424,326) (Figure-4C), as was exposure to health-risk behaviour content (specifically marketer-generated content) (vs not exposed) (1.79, 1.63 to 1.96; I^2^=0.00%; n=22,882) (Appendix-16, p 188). In stratified analyses (Appendix-16, p 189-193) for frequency of use stronger associations were observed for LMICs (2.47, 1.56 to 3.91 vs 1.64, 1.31 to 2.06; p=0.12 for HICs), and for use of social networking sites (1.90, 1.57 to 2.30 vs 1.42, 1.06 to 1.90 for general SM; p=0.10).

#### Electronic nicotine delivery system (ENDS) use

Across all exposures investigated, 88.9% of studies (n=8/9; 77.8% low/moderate RoB) reported harmful associations on ENDS use (Appendix-16, p 194). Exposure to health-risk behaviour content (specifically marketer-generated content) (vs not exposed) was associated with increased ENDS use (OR=1.73, 1.34 to 2.23; I^2^=63.4%; n=721,322) (Appendix-16, p 195). No clear differences were identified on stratification (Appendix-16, p 196-197).

#### Sexual risk behaviour

After excluding one study with inconsistent findings, across all exposures investigated 90.3% (n=28/31; 67.7% high RoB) reported harmful associations on sexual risk behaviours (Appendix-16, p 198). Frequent/at all use (vs infrequent/not at all) was associated with increased sexual risk behaviours (e.g., sending a ‘sext’, transactional sex, and inconsistent condom use) (1.78, 1.49 to 2.13; I^2^=77.8%; n=47,325) (Figure-5A). Meta-regression (coeff. −0.36 [−0.68, −0.04]; p=0.03) (Appendix-16, p 276) and stratified analyses (Appendix-16, p 200-207) suggested stronger associations for adolescents <16 years vs those ≥16 years, but no moderation effects were seen for SM category (p=0.12) or study setting (p=0.52). Few studies assessed associations for time spent (Appendix-16, p 208).

**Figure-5.**
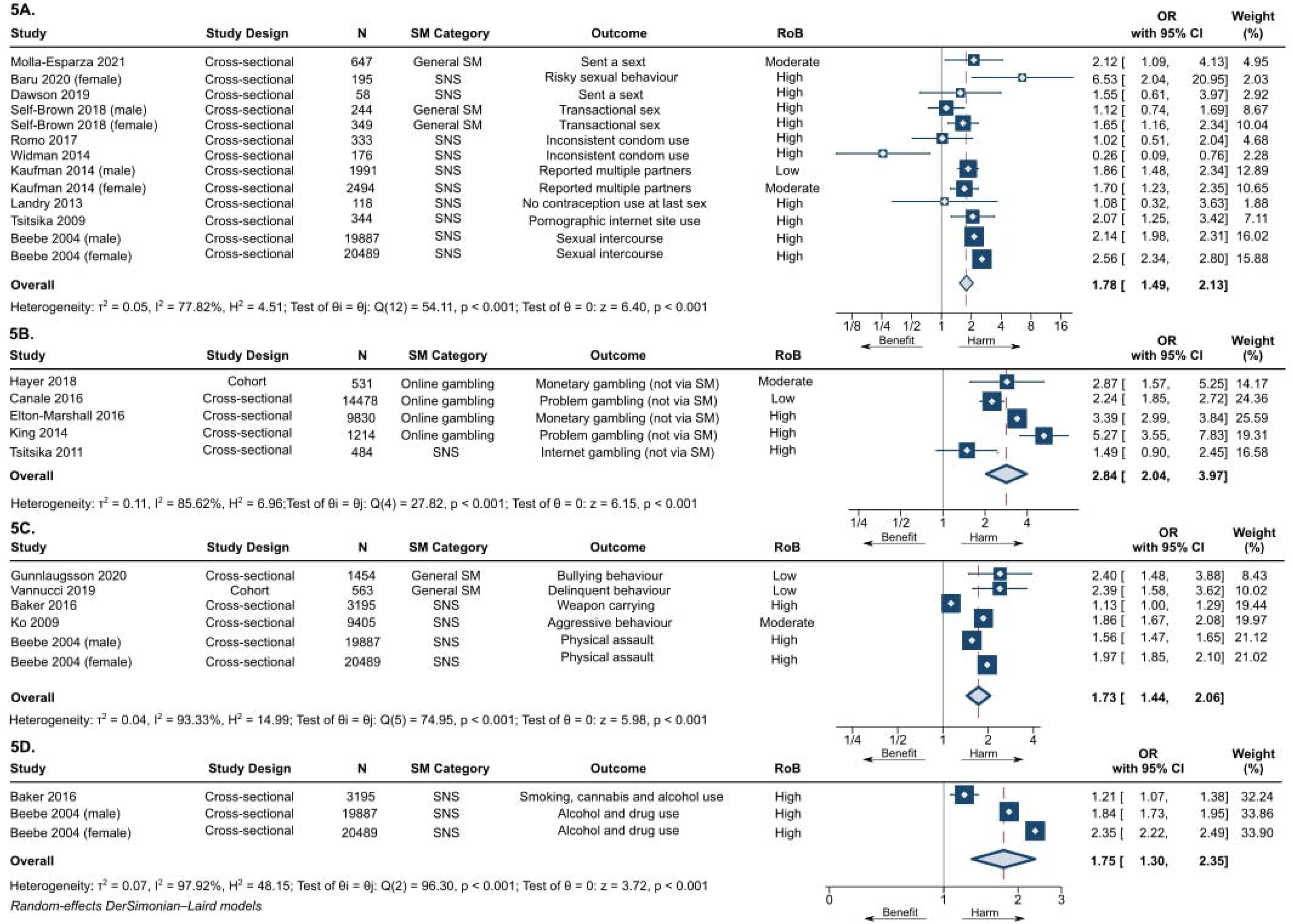
Forest plots for association between frequency of social media use and A) sexual risk behaviour, B) gambling, C) anti-social behaviour, and D) multiple risk behaviours. Figure 5A presents forest plot for binary exposure (frequent/at all vs infrequent/not at all) & binary/continuous sexual risk behaviour outcome meta-analysis, with odds ratio (OR) used as common metric. Total number of study participants = 47,325. Figure 5B presents forest plot for binary exposure (frequent/at all vs infrequent/not at all) & binary/continuous gambling outcome meta-analysis, with odds ratio (OR) used as common metric. Total number of study participants = 26,537. Figure 5C presents forest plot for binary exposure (frequent/at all vs infrequent/not at all) & binary/continuous anti-social behaviour outcome meta-analysis, with odds (OR) used as common metric. Total number of study participants = 54,993. Figure 5D presents forest plot for binary exposure (frequent/at all vs infrequent/not at all) & binary/continuous multiple risk behaviours outcome meta-analysis, with odds ratio (OR) used as common metric. Total number of study participants = 43,571. Abbreviations: CI = Confidence interval; N = Number of study participants; OR = Odds ratio; RoB = Risk of bias; SM = Social media and SNS = Social networking sites.

#### Gambling

After excluding one study demonstrating inconsistent findings, across all exposures investigated, all six studies investigating gambling reported harmful associations (Appendix-16, p 209). Frequent/at all use (vs infrequent/not at all) was associated with increased gambling (not via SM) (OR=2.84, 2.04 to 3.97; I^2^=85.6%; n=26,537) (Figure-5B). On differentiation by SM category, a relatively large association was found for online gambling via SM (3.22, 2.32 to 4.49), however associations were not present for social networking sites and general SM (Appendix-16, p 212).

#### Anti-social behaviour

Across all exposures investigated, all sixteen studies (43.8% low/moderate RoB) investigating anti-social behaviour demonstrated harmful associations (Appendix-16, p 213). Frequent/at all use (vs infrequent/not at all) was associated with increased anti-social behaviour (e.g., bullying, physical assault, and aggressive/delinquent behaviour) (OR=1.73, 1.44, 2.06; I^2^=75.0%; n=54,993) (Figure-5C), with time spent similarly associated with increased risk (Appendix-16, p 215). No subgroup differences were seen (Appendix-16, p 216-219).

#### Inadequate physical activity

For inadequate physical activity, after excluding three studies with inconsistent findings, 36.4% of studies (n=4/11; 72.7% low/moderate RoB) reported harmful associations across all exposures investigated (Appendix-16, p 220). No association between time spent (assessed on a continuous scale) and adolescent engagement in physical activity was seen (Std.Beta −0.00, −0.02 to 0.01; I^2^=59.8%; n=37,417) (Appendix-16, p 221), with no important differences across subgroups (Appendix-16, p 222-224).

#### Unhealthy dietary behaviour

Across all exposures investigated, all thirteen studies (incl. four RCTs: two rated low RoB and two some concerns) investigating unhealthy dietary behaviour demonstrated harmful associations, with most at low RoB (61.5%) (Appendix-16, p 225). Exposure to health-risk behaviour content (specifically marketer-generated content) was associated with increased unhealthy food consumption (OR=2.12, 1.87 to 2.39; I^2^=0.00%; n=9424) when compared to those unexposed (Appendix-16, p 226-277).

#### Multiple risk behaviours

For multiple risk behaviours, all nine studies demonstrated harmful associations across all exposures investigated (Appendix-16, p 228). The pooled OR for frequent/at all SM use (vs infrequent/not at all) was 1.75 (1.30 to 2.35; I^2^=96.3%; n=43,571) (Figure-5D), but the few studies precluded stratification.

#### Sensitivity analyses

For ENDS use, associations were stronger for cohort study datapoints (OR 2.13, 1.72 to 2.64 vs 1.43, 1.20 to 1.69 for cross-sectional datapoints; p=0.004) (Appendix-16, p 230) but no clear differences were seen for other outcomes (Appendix-16, p 231-242). Although based on few studies, for unhealthy dietary behaviour a stronger association was found for the RCT datapoint (3.21, 1.63 to 6.30 vs 2.08, 1.84 to 2.37 for cross-sectional datapoints; p=0.22) (Appendix-16, p 243).

When stratifying by adjustment for critical confounding domains no clear differences were identified (Appendix-16, p 244-255), with some exceptions. Associations were stronger for unadjusted vs adjusted datapoints for exposure to health-risk behaviour content and alcohol use (Std.Beta 0.28, 0.14 to 0.43 vs 0.07, 0.03 to 0.12; p=0.008) and for frequent use (vs infrequent) and tobacco use (OR 2.11, 1.73 to 2.58 vs 1.51, 1.23 to 1.85; p=0.02), though this was based on few studies (Appendix-16, p 255-256).

For alcohol use, effect sizes were generally stronger for moderate/high RoB datapoints (vs low) (Appendix-16, p 257-258), excluding time spent (≥2 vs <2 hrs/day) and exposure to health-risk behaviour content (vs not exposed) where low (compared to moderate/high) RoB datapoints displayed stronger associations (Appendix-16, p 259-260). For drug use, sexual risk and anti-social behaviour, no differences were detectable or low/moderate RoB datapoints showed stronger associations (compared to high) (Appendix-16, p 261-265). For tobacco use and gambling, stronger associations were found for high RoB datapoints or no clear differences were identified (Appendix-16, p 266-268). No clear differences by RoB were observed for the remaining outcomes (Appendix-16, p 269-270).

On exclusion of datapoints which overlapped the age-range of 10-19 years, there was a marginal reduction in effect size (Appendix-16, p 271) or no important differences were seen (Appendix-16, p 272-275).

#### Publication bias

Funnel plots and Egger’s test results suggested some publication bias in the meta-analysis investigating frequent/at all SM use (vs infrequent/not at all) and sexual risk behaviours (p=0.02; bias toward the null) (Appendix-17). Insufficient data precluded investigation of other outcomes.

#### Certainty of the evidence

As frequency was the most commonly investigated exposure and continuous and binary exposures reported similar effects, we focussed the GRADE assessment on the binary exposure, frequency of use. Where we report harmful effects on alcohol use with low certainty, and with drug, tobacco, ENDS use, sexual risk behaviours, gambling, and multiple risk behaviours with very low certainty We conducted a post hoc GRADE assessment for exposure to health-risk behaviour content (vs no exposure) and unhealthy dietary behaviour due to the substantial difference in quality of evidence observed (four RCTs), where we report moderate GRADE certainty (Table-1 and Appendix-18).^59^

**Table-1.** Condensed summary of findings and certainty of evidence (as per GRADE) ^a^ The risk in the intervention group (and its 95% confidence interval) is based on the assumed risk in the comparison group and the relative effect of the intervention (and its 95% CI). Abbreviations: CI = Confidence interval; GRADE = Grading of Recommendations Assessment, Development and Evaluation; and OR = Odds ratio. For full GRADE results, see Appendix-18. ^a^Downgraded by 1 level for risk of bias of included studies. ^b^Downgraded by 2 levels for inconsistency and risk of bias of included studies. ^c^Downgraded by 2 levels for imprecision and risk of bias of included studies. ^d^Downgraded by 2 levels for publication bias, and risk of bias of included studies. ^e^Downgraded by 2 levels for inconsistency and risk of bias of included studies. ^f^Downgraded by 1 level for indirectness.

| Table 1: Condensed summary of findings and certainty of evidence (as per GRADE) |  |  |  |  |  |
| --- | --- | --- | --- | --- | --- |
| Certainty of evidence for frequency of social media use (frequent vs infrequent) and seven priority outcome domains |  |  |  |  |  |
| Outcome | Number of participants (studies) | Anticipated absolute effects<sup>a</sup> (95% CI) |  | Effect estimate (95% CI) | Certainty of evidence |
|  |  | Risk with infrequent social media use | Risk with frequent social media use |  |  |
| Alcohol use | 383,670 (9 observational studies) | 48.9% of participants in the control group used alcohol | 58.6% of exposed group participants used alcohol (56.6 to 60.8%) | OR 1.48 (1.36 to 1.62) | Low |
| Drug use | 117,645 (6 observational studies) | 17.0% of participants in the control group used drugs | 20.8% of exposed group participants used drugs (17.7 to 24.2%) | OR 1.28 (1.05 to 1.56) | Very low<sup>a</sup> |
| Tobacco use | 424,326 (8 observational studies) | 12.1% of participants in the control group used tobacco | 19.6% of exposed group participants used tobacco (16.6 to 23.1%) | OR 1.78 (1.45 to 2.19) | Very low<sup>b</sup> |
| Electronic nicotine delivery system use (effect direction) | 18,047 (3 observational studies) | 66.7% of studies reported harmful association of social media use on adolescent use of electronic nicotine delivery systems (20.8 to 93.9%) |  |  | Very low<sup>c</sup> |
| Sexual risk behaviour | 47,325 (10 observational studies) | 37.0% of participants in the control group engaged in sexual risk behaviours | 51.1% of exposed group participants engaged in sexual risk behaviours (46.6 to 55.5%) | OR 1.78 (1.49 to 2.13) | Very low<sup>d</sup> |
| Gambling | 26,537 (5 observational studies) | 21.4% of participants in the control group engaged in gambling | 43.6% of exposed group participants engaged in gambling (35.7 to 52.0%) | OR 2.84 (2.04 to 3.97) | Very low<sup>a</sup> |
| Multiple risk behaviours | 43,571 (2 observational studies) | 41.3% of participants in the control group engaged in multiple risk behaviours | 55.2% of exposed group participants engaged in multiple risk behaviours (47.8 to 62.3%) | OR 1.75 (1.30 to 2.35) | Very low<sup>e</sup> |
| Certainty of evidence for exposure to health-risk behaviour content on social media (exposed vs unexposed) and unhealthy dietary behaviour |  |  |  |  |  |
| Outcome | Number of participants (studies) | Anticipated absolute effects<sup>a</sup> (95% CI) |  | Effect estimate (95% CI) | Certainty of evidence |
|  |  | Risk with infrequent social media use | Risk with frequent social media use |  |  |
| Unhealthy dietary behaviour (effect direction) | 521 (4 randomised control trials) | All studies report harmful effect of social media use on adolescent engagement in unhealthy dietary behaviours (51.0 to 100.0%) |  |  | Moderate<sup>f</sup> |

## Discussion

Our systematic review suggests social media (SM) use is adversely associated with several adolescent health-risk behaviours, including increased alcohol, drug, tobacco, ENDS use, gambling, sexual risk, anti-social, unhealthy dietary and multiple risk behaviours. Exposure to health-risk behaviour content on SM has the strongest evidence of harm, particularly in relation to alcohol use and unhealthy dietary behaviour (moderate GRADE certainty).

Review strengths include its comprehensive scope, inclusion of RCTs and adjusted estimates, assessment of risk of bias and certainty of the evidence using GRADE, and its explicit focus on SM specifically (rather than digital technologies).^59^ To our knowledge, no other review has synthesised the evidence of adolescent health-risk behaviours associated with SM categories, platforms, and content, and considered whether SM impacts vary across social groups. Generally, for alcohol use larger associations were seen for adolescents ≥16 years (vs <16 years), and for exposure to user-generated content (vs marketer-generated content). Whilst for tobacco use, larger associations were observed for low-middle income countries (vs high income countries). We followed a pre-registered protocol,^45^ with decisions about critical confounding domains and stratified analyses informed by a comprehensive literature review, logic model (Figure-1), and advisory group consultation. Searches covered the period 1997 to 2022. The nature of SM use has changed dramatically across this period, but the majority of data collection (66.9% of studies) occurred in the last eight years and so should be generalisable to the current SM environment.

However, objective SM measures were rare with self-report most common. When assessing frequency of use, most studies compared frequent vs infrequent, some daily vs non-daily, and some any use vs none. These exposure categories were combined in meta-analyses due to limited data availability, but frequency (assessed via continuous scale) reported similar findings. Some meta-analyses were based on few studies, yielding more uncertain estimates. However, meta-analysis is feasible even with two studies, and there is an argument that meta-analysis should be conducted where possible.^66, 77^ Meta-analysis was performed when three or more studies were available for a given synthesis and this was complemented with a narrative synthesis using the SWiM reporting guideline and effect direction plots.^44, 65, 77^ As recommended by Cochrane,^78^ adjustments for multiple tests were not conducted. Instead, effect sizes were the focus of interpretation where possible, outcomes and analyses of interest were prespecified in the published protocol,^45^ subgroup analyses were interpreted with caution, and results were not selected for emphasis on the basis of a statistically significant P value, with all conducted analyses presented. Moreover, although the review focus is harmful risk behaviours, SM may have positive/negligible harmful influences on some outcomes, such as physical activity and drug use, thus a holistic view should be taken when interpreting the review findings.

Our sensitivity analysis by confounder adjustment, focussed on critical confounding domains (age, sex, and SEP). We acknowledge other shared risk factors may exist between SM and health-risk behaviours (e.g., parental health-risk behaviours). Cross-sectional studies are subject to reverse causation, as reflected in the logic model (Figure-1). A bidirectional relationship may therefore exist, with adolescents who engage in health-risk behaviours more inclined to use social media to obtain peer approval and positive feedback. However, we identified harmful associations across study designs, including longitudinal studies which adjusted for baseline measures of outcomes and RCTs. Included RCTs involved random assignment of study participants to existing or manipulated SM posts (where all authors stipulated the means by which they tried to mimic the actual social media environment). For example, De Jans et al. identified a harmful association between exposure to manipulated Instagram posts showing a fictitious influencer promoting a snack low in nutritional value (vs high) and unhealthy snack consumption.^79^ A limitation of this study was its use of a fictitious influencer, which may limit its validity. Yet, Folkvord et al. overcame this limitation through use of existing Instagram posts demonstrating a popular social influencer consuming energy dense snacks (vs vegetables), subsequently finding participants exposed to the energy dense snack condition consumed less vegetables when compared to those participants exposed to the vegetable condition.^80^ The use of existing Instagram posts from a popular social influencer among the target group of participants helped improve external validity. Thus, the moderate GRADE certainty for included RCTs suggests a causal effect of health-risk behaviour content on unhealthy dietary behaviour although these studies still had limitations (e.g., a lack of real-time exposure to SM).

Previous reviews have focused on SM use to deliver behaviour change interventions, finding this avenue has potential. ^9, 81, 82^ Less attention has been paid to the implications of SM itself for health. Vannucci et al, identified cross-sectional correlations between SM use and substance use and risky sexual behaviour in adolescents, however were unable to separate out general electronic media use (electronic media with a direct component involving social interactions with others (2022 personal communication with A. Vannucci)) from SM use, although they did include some exploratory sensitivity analyses of potential differences by type of SM assessment.^42^ Whilst Curtis et al. reported correlations between alcohol-related SM content and alcohol consumption and alcohol-related problems in young adults, however did not explore if associations differed between exposure to user or marketer-generated content.^32^ Importantly, both reviews did not incorporate adjusted estimates or identify RCT evidence and did not formally assess risk of bias of the underlying evidence.^32, 42^

As SM reaches diverse populations, reporting of population characteristics and disaggregating results by socio-demographic groups should be prioritised. With most research conducted in the global North, research in low- and middle-income countries is needed.^34^ SWiM findings suggested SM use may present beneficial effects on adolescent engagement in physical activity, though meta-analysis (based on four cross-sectional studies) did not substantiate this conclusion. Further research on this outcome would allow health policy makers to potentially harness the benefits SM use could present on adolescent health. Moreover, many of the risk behaviours investigated can be experimental during adolescence, and the extent to which these behaviours affect health may vary. Longitudinal research tracking adolescents into adulthood would help study this. Well-conducted randomised trials studying risk-behaviours over and above unhealthy dietary behaviour would yield more robust evidence than currently available and have been shown to be feasible. Addressing the limitations of existing RCTs and use of real-time monitoring data of SM use would allow for more definitive causal conclusions on the effects of SM activity on adolescent health risk-behaviours.

The methodological limitations in the evidence may reflect limited access to data required to investigate SM’s health implications, adding weight to calls to compel SM corporations to share data with researchers.^83, 84^ In the absence of real-time objective data, the development of generalisable, validated measures of SM use (considering SM activities performed e.g., active/passive use) would facilitate comparability across studies. Awareness of the aspects of SM most harmful to adolescents (e.g., user/marketer-generated content), could support development and expedite introduction of the delayed UK Online Safety Bill, aimed at securing adolescents’ online safety.^83, 85^ The importance of exposure to marketer-generated content identified in this review in potentially promoting health risk behaviours highlights gaps in the Bill which largely focusses on user-generated content, and the unmet need for legislation targeting influencer marketing.^83, 86, 87^ Further research in this area could prove fruitful for informing regulation.

In adopting a multi-sector approach to securing adolescent online safety, digital-literacy school education and resource provision to parents, educators and health professionals to improve understanding of the different aspects of SM use (e.g., time spent, exposure to health-risk behaviour content) and the potential risks/benefits they present to adolescent health may be warranted.^88^

## Conclusion

Our review finds predominantly harmful associations between social media use and adolescent health-risk behaviours. However, this is based on a largely cross-sectional body of evidence, using self-report measures of social media use, and at risk of residual confounding due to many confounders remaining unadjusted for. Experimental and risk-taking behaviours are an inherent part of adolescence; however, as safeguards for a digital world are still evolving, application of the precautionary principle suggests action across academic, governmental, health and educational sectors to understand and reduce the risks adolescents may face from use of social media may be warranted.

## Contributors

AKP, SVK, AP and MH drafted the study protocol. AKP conducted literature searches, and all authors contributed to the screening process and selection of included studies. AKP conducted data extraction and risk of bias assessments; data were checked, and independent risk of bias scoring undertaken by SVK, AP, MH, RT, and PMH. AKP completed all data analysis, had full access to all the data in the study and takes responsibility for the integrity of the data and the accuracy of the data analysis. All authors critically reviewed and approved the manuscript. The corresponding author attests that all listed authors meet authorship criteria and that no other meeting the criteria have been omitted.

## Funding

This work was supported by the Medical Research Council (MC_UU_00022/2), Chief Scientist Office (SPHSU17), an NHS Research Scotland Senior Clinical Fellowship (SCAF/15/02) and the Wellcome Trust (218105/Z/19/Z, 205412/Z/16/Z). The funders played no active role in the design and conduct of the study; collection, management, analysis, and interpretation of the data; preparation, review, or approval of the manuscript; or decision to submit the manuscript for publication.

## Competing interests

All authors declare no support from any organisation for the submitted work; no financial relationships with any organisations that might have an interest in the submitted work in the previous three years; no other relationships or activities that could appear to have influenced the submitted work.

## Ethical approval

No ethics approval was requested, as the research solely extracted non-disclosive data from previously published studies in which informed consent was obtained by the primary investigators.

## Supporting information

PRISMA Checklist

## Data Availability

Data analysed were based on published data. Template data forms, the data extracted from included studies and data used for analyses are available from the corresponding author on reasonable request. The study protocol is published on PROSPERO: https://www.crd.york.ac.uk/prospero/display_record.php?RecordID=179766 (ID: CRD42020179766)

## Acknowledgements

The authors gratefully acknowledge the assistance of Ms Valerie Wells in designing and implementing the review search strategy, and Dr Hilary Thomson, and Dr Michele Hilton Boon, for providing guidance on the modifications made to the Newcastle Ottawa Scale and on usage of GRADE. We extend sincere thanks to the advisory group members for providing feedback on the protocol and provisional findings. We also thank Dr Andrew Baxter for his assistance with data visualisations.

## Transparency

The lead author (AKP) affirms that the manuscript is an honest, precise, and transparent account of the review reported, with no important aspects of the review omitted. Any discrepancies from the review as planned (and, registered) have been explained.

## Open access/Copyright statement

This is an open access article distributed in accordance with the terms of the Creative Commons Attribution (CC BY 4.0) license, which permits others to distribute, remix, adapt and build upon this work, for commercial use, provided the original work is properly cited. See: http://creativecommons.org/ licenses/by/4.0.

## Notes

### Competing Interest Statement

The authors have declared no competing interest.

### Clinical Protocols

https://www.crd.york.ac.uk/prospero/display_record.php?RecordID=179766

