## Supplementary Material for "Social media use and adolescent health-risk behaviours: A systematic review and meta-analysis"

**to**

Ms Amrit Kaur Purba (MPH)

Dr Rachel M Thomson (MPH)

Dr Paul M Henery (PhD)

Dr Anna Pearce (PhD)

Professor Marion Henderson (PhD)

Professor S Vittal Katikireddi (PhD)

### Appendix-1. Deviations from protocol

To improve the interpretability and comparability of the review findings, we made the following explanatory clarifications and minor protocol deviations.

Clarifications to and deviations from protocol as published:

- Updated the search to include all eligible studies from 01/1997-06/2022.
- Clarified online dating platforms were not included under the exposure social media use.
- Clarified randomized control trials were eligible for inclusion.
- Clarified the risk of bias tool to be used for randomised control trials is the Cochrane RoB-2 tool.
- We did not include ethnicity as a critical confounding factor when assessing study adjustment as the potential role of ethnicity as a confounder was not deemed substantial given the likely homogenous populations investigated within many included studies.
- We included online (social) gaming and online (social) gambling within included exposures following discussions with advisory group members, due to their emerging placement in social media platforms and the overlap in functionalities they share with social media.
- For planned subgroup analyses/meta-regression we originally stated if two or fewer studies were found in a given sub-category of a binary/multi-categorical moderator, formal moderation analysis would not be conducted for that specific variable. Due to the limited number of included studies, we allowed for more leniency using the data available and the decision was made to perform subgroup analyses/meta-regression if at least one subgroup had two or more studies, noting the requirement to interpret any conclusions with caution. The same rule was applied when conducting sensitivity analyses.
- Where duplicate data were identified across multiple studies, we prioritised inclusion of studies which had the longest period of follow up, followed by studies which had the largest most representative sample size, and then by most recent. We did not anticipate there would be many cohort studies identified during protocol development, thus we did not initially prioritise the inclusion of studies with longer follow up periods.
- Following discussions with statistical experts post publication of the protocol, the decision was made to combine binary exposure and binary/continuous outcomes in line with guidance provide by Cochrane^1^ expressed as odds ratios. For continuous exposure measures, we stated estimates would be converted to standardised regression coefficients/correlations. Where possible we converted regression coefficients to standardised regression coefficients. We used the recent method recommended by Mathur and Vanderwheele^2^ which facilitates the conversion of a Pearson correlation coefficient to standardised mean difference.
- We conducted a post-hoc GRADE assessment for the exposure to health-risk behaviour content on social media and outcome unhealthy dietary behaviour due to the substantial differences in the studies used to assess this exposure/outcome combination (specifically investigation by randomised control trials).

### Appendix-2. Details of search strategies conducted

All searches outlined in Table A were initially conducted on 30.04.2020 and repeated on 06.06.2022.

##### Table A. Sources searched and corresponding hits

| Sources searched | Date of initial search | Number of hits | Date of updated search | Number of hits | Total number of hits |
| --- | --- | --- | --- | --- | --- |
| Electronic databases | | | | | |
| CINAHL | 30.04.2020 | 6740 | 06.06.2022 | 1435 | 8175 |
| EMBASE | 30.04.2020 | 6896 | 06.06.2022 | 2563 | 9459 |
| MEDLINE | 30.04.2020 | 5253 | 06.06.2022 | 1813 | 7066 |
| APA PsychINFO | 30.04.2020 | 2545 | 06.06.2022 | 508 | 3053 |
| SocINDEX | 30.04.2020 | 245 | 06.06.2022 | 62 | 307 |
| Pre-print repositories | | | | | |
| SSRN | 30.04.2020 | 0 | 06.06.2022 | 0 | 0 |
| SocArXic | 30.04.2020 | 11 | 06.06.2022 | 10 | 21 |
| PsyArXiv | 30.04.2020 | 6 | 06.06.2022 | 35 | 41 |
| MedRxiv | 30.04.2020 | 18 | 06.06.2022 | 0 | 18 |
| Internet search engine | | | | | |
| Google Scholar | 30.04.2020 | 30 | 06.06.2022 | 30 | 60 |
| Total number of hits | | | | | **28,200** |
| Total number of hits following removal of duplicates | | | | | **17,077** |

##### Table B. Cumulative Index to Nursing and Allied Health Literature (CINAHL) search strategy

| String number | String |
| --- | --- |
| 1 | (MH "Adolescence+") OR (MH "Child+") OR (MH "Students+") OR (MH "Students, High School") OR (MH "Schools, Middle") OR (MH "Schools, Secondary") |
| 2 | TI ("young people" OR youth OR "school child*" OR teen* OR "young person*" OR "middle school" OR middle-school OR "secondary school" OR "high school"OR iGen OR "generation Z" OR "gen Z") OR AB ("young people" OR youth OR "school child*" OR teen* OR "young person*" OR "middle school" OR middle-school OR "secondary school" OR "high school" OR iGen OR "generation Z" OR "gen Z") |
| 3 | S1 OR S2 |
| 4 | (MH "Social Networking+") OR (MH "Social Media+") OR (MH "Smartphone") OR (MH "Internet+") OR (MH "Screen Time") OR (MH "Instant Messaging") |
| 5 | TI ("screen time" OR "social media" OR "social network* site" OR "social networking" OR "social-networking" OR "web 2.0" OR "online game*" OR "online gaming" OR "online social gaming" OR hashtag OR "instant messag*" OR instagram OR "Whats App" OR whatsapp OR facebook OR twitter OR linkedin OR youtube OR "you tube" OR tumblr OR vine OR snapchat OR myspace OR bebo OR reddit OR neknominate OR myspace OR wickr OR telegram OR whisper OR "kik messenger" OR "Tencent QQ" OR wechat OR meetup OR tiktok OR hinge OR happn OR bumble OR grindr OR Tinder OR "inner circle" OR periscope OR twitch) N2 (usage OR use*)) OR AB ("screen time" OR "social media" OR "social network* site" OR "social networking" OR "social-networking" OR "web 2.0" OR "online game*" OR "online gaming" OR "online social gaming" OR hashtag OR "instant messag*" OR instagram OR "Whats App" OR whatsapp OR facebook OR twitter OR linkedin OR youtube OR "you tube" OR tumblr OR vine OR snapchat OR myspace OR bebo OR reddit OR neknominate OR myspace OR wickr OR telegram OR whisper OR "kik messenger" OR "Tencent QQ" OR wechat OR meetup OR tiktok OR hinge OR happn OR bumble OR grindr OR Tinder OR "inner circle" OR periscope OR twitch) N2 (usage or use*)) |
| 6 | TI ("screen time" OR "social media" OR "social networking" OR "social-networking" OR "social network* site*" OR "web 2.0" OR "online game*" OR "online gaming" OR "online social gaming" OR hashtag OR "instant messag*" OR instagram OR "Whats App" OR whatsapp OR facebook OR twitter OR linkedin OR youtube OR "you tube" OR tumblr OR vine OR snapchat OR myspace OR bebo OR reddit OR neknominate OR myspace OR wickr OR telegram OR whisper OR "kik messenger" OR "Tencent QQ" OR wechat OR meetup OR tiktok OR hinge OR happn OR bumble OR grindr OR Tinder OR "inner circle" OR periscope OR twitch) OR AB ("screen time" OR "social media" OR "social networking" OR "social network* site*" OR "social-networking" OR "web 2.0" OR "online game*" OR "online gaming" OR "online social gaming" OR hashtag OR "instant messag*" OR instagram OR "Whats App" OR whatsapp OR facebook OR twitter OR linkedin OR youtube OR "you tube" OR tumblr OR vine OR snapchat OR myspace OR bebo OR reddit OR neknominate OR myspace OR wickr OR telegram OR whisper OR "kik messenger" OR "Tencent QQ" OR wechat OR meetup OR tiktok OR hinge OR happn OR bumble OR grindr OR Tinder OR "inner circle" OR periscope OR twitch) |
| 7 | S4 OR S5 OR S6 |
| 8 | (MH "Risk Taking Behavior+") OR (MH "Substance Abuse+") OR (MH "Substance Abusers+") |
| 9 | TI ("substance use*" OR "substance misuse*" OR risk-behav* OR "risk behav*" OR "risky behav*" OR "risk-taking behav*" OR "multiple risk behav*") OR AB ( "substance use*" OR "substance misuse*" OR risk-behav* OR "risk behav*" OR "risky behav*" OR "risk-taking behav*" OR "multiple risk behav*") |
| 10 | S8 OR S9 |
| 11 | (MH "Tobacco+") OR (MH "Smoking+") OR (MH "Tobacco Products+") |
| 12 | TI ("adolescent smok*" OR "chewing tobacco" OR "tobacco dependence" OR "tobacco use" OR "tobacco consumption" OR "tobacco snuff" OR cigarette OR "smoking initiation" OR "smoking behav*") OR AB ("adolescent smok*" OR "chewing tobacco" OR "tobacco dependence" OR "tobacco use" OR "tobacco consumption" OR "tobacco snuff" OR cigarette OR "smoking initiation" OR "smoking behav*") |
| 13 | S11 OR S12 |
| 14 | (MH "Electronic Cigarettes") |
| 15 | TI ("electronic nicotine delivery system*" OR e-cigarette* OR Juul OR vaping OR vape) OR AB ("electronic nicotine delivery system*" OR e-cigarette* OR Juul OR vaping OR vape) |
| 16 | S14 OR S15 |
| 17 | (MH "Drinking Behavior+") OR (MH "Alcohol Abuse+") OR (MH "Alcoholic Intoxication+") |
| 18 | TI ("underage drinking" OR "under-age drinking" OR "under age drinking" OR temperance OR "alcohol use*" OR "alcohol intake" OR "problem drinking" OR "alcoholism" OR "alcohol abstinence" OR "drinking behaviour" OR "alcohol consumption" OR "binge drinking") OR AB ("underage drinking" OR "under-age drinking" OR "under age drinking" OR temperance OR "alcohol use*" OR "alcohol intake" OR "problem drinking" OR "alcoholism" OR "alcohol abstinence" OR "drinking behaviour" OR "alcohol consumption" OR "binge drinking") |
| 19 | S17 OR S18 |
| 20 | (MH "Street Drugs+") OR (MH "Drugs") OR (MH "Cannabis") OR (MH "Cocaine+") |
| 21 | TI ("cannabis use*" OR "cannabis addict*" OR "illicit drug*" OR "drug abuse*" OR "drug use*" OR "drug misuse*" OR weed OR skunk OR marijuana OR "special k" OR crack OR methamphetamine* OR ecstasy OR heroin OR LSD OR steroid* OR ketamine OR MDMA OR GHB OR GBL) OR AB ("cannabis use*" OR "cannabis addict*" OR "illicit drug*" OR "drug abuse*" OR "drug use*" OR "drug misuse*" OR weed OR skunk OR marijuana OR "special k" OR crack OR methamphetamine* OR ecstasy OR heroin OR LSD OR steroid* OR ketamine OR MDMA OR GHB OR GBL) |
| 22 | S20 OR S21 |
| 23 | (MH "Juvenile Delinquency") OR (MH "Theft+") OR (MH "Disruptive Behavior") OR (MH "Gangs") OR (MH "Violence+") |
| 24 | TI ("antisocial behav*" OR "anti-social behav*" OR "social problem*" OR assault OR fighting OR steal* OR shoplift* OR vandal* OR "public nuisance") OR AB ("antisocial behav*" OR "anti-social behav*" OR "social problem*" OR assault OR fighting OR steal* OR shoplift* OR vandal* OR "public nuisance") |
| 25 | S23 OR S24 |
| 26 | (MH "Pregnancy in Adolescence+") OR (MH "Sexually Transmitted Diseases+") OR (MH "Pregnancy, Unwanted") OR (MH "Sex+") OR (MH "Unsafe Sex") OR (MH "HIV Infections+") |
| 27 | TI (sexting OR sex-text OR "sex text" OR "sexual behav*" OR "sexual intercourse" OR "sexually transmitted infection*" OR STIs OR STDs OR "teen* pregnancy" OR "unprotected sex" OR "first intercourse" OR "casual sexual relations*" OR "intimate sexual contact" OR "under age sex" OR "underage sex" OR "under-age sex" OR "underage pregnancy" OR "under-age pregnancy" OR "under age pregnancy") OR AB (sexting OR sex-text OR "sex text" OR "sexual behav*" OR "sexual intercourse" OR "sexually transmitted infection*" OR STIs OR STDs OR "teen* pregnancy" OR "unprotected sex" OR "first intercourse" OR "casual sexual relations*" OR "intimate sexual contact" OR "under age sex" OR "underage sex" OR "under-age sex" OR "underage pregnancy" OR "under-age pregnancy" OR "under age pregnancy") |
| 28 | S26 OR S27 |
| 29 | (MH "Gambling") |
| 30 | TI (gambling OR betting) OR AB (gambling OR betting) |
| 31 | S29 OR S30 |
| 32 | (MH "Sweetened Beverages") OR (MH "Fast Foods") OR (MH "Adolescent Nutrition") OR (MH "Eating Behavior+") |
| 33 | TI ("unhealthy diet*" OR "poor diet*" OR "dietary behav*" OR "sugary drink*" OR sweet* ) OR AB ("unhealthy diet*" OR "poor diet*" OR "dietary behav*" OR "sugary drink*" OR sweet* ) |
| 34 | S32 OR S33 |
| 35 | (MH "Physical Activity") OR (MH "Exercise+") OR (MH "Physical Fitness+") OR (MM "Life Style, Sedentary") |
| 36 | TI ( "physical inactiv*" OR "physical activ*" OR exercis* OR sport*) OR AB ( "physical inactiv*" OR "physical activ*" OR exercis* OR sport*) |
| 37 | S35 OR S36 |
| 38 | S10 OR S13 OR S16 OR S19 OR S22 OR S25 OR S28 OR S31 OR S34 OR S37 |
| 39 | S3 AND S7 AND S38 |

**Legend:** Initial search: date of search- 30.04.2020. Interface- EBSCOhost. Database and coverage- Cumulative Index to Nursing and Allied Health Literature (CINAHL),1981 to present. Limits applied- 01.01.1997 to 30.04.2020. Updated search: date of search- 06.06.2022. Interface- EBSCOhost. Database and coverage- Cumulative Index to Nursing and Allied Health Literature (CINAHL),1981 to present. Limits applied- 01.04.2020 to 31.06.2022.

##### Table C. Embase search strategy

| String number | String |
| --- | --- |
| 1 | adolescent/ or child/ or juvenile/ |
| 2 | middle school student/ or student/ or high school student/ |
| 3 | ("young people" or youth or "school child*" or teen* or "young person*" or "middle school" or middle-school or "secondary school" or "high school" or iGen or "generation Z" or "gen Z").ab,ti. |
| 4 | 1 or 2 or 3 |
| 5 | online social network/ or social media/ or smart phone/ or internet/ or screen time/ |
| 6 | (("screen time" or "social media" or "social networking" or "social-networking" or "social network* site*" or "web 2.0" or "online game*" or "online gaming" or "online social gaming" or hashtag or "instant messag*" or instagram or "Whats App" or whatsapp or facebook or twitter or linkedin or youtube or "you tube" or tumblr or vine or snapchat or myspace or bebo or reddit or neknominate or myspace or wickr or telegram or whisper or "kik messenger" or "Tencent QQ" or wechat or meetup or tiktok or hinge or happn or bumble or grindr or Tinder or "inner circle" or periscope or twitch) adj2 (usage or use*)).ab,ti. |
| 7 | ("screen time" or "social media" or "social networking" or "social-networking" or "social network* site*" or "web 2.0" or "online game*" or "online gaming" or "online social gaming" or hashtag or "instant messag*" or instagram or "Whats App" or whatsapp or facebook or twitter or linkedin or youtube or "you tube" or tumblr or vine or snapchat or myspace or bebo or reddit or neknominate or myspace or wickr or telegram or whisper or "kik messenger" or "Tencent QQ" or wechat or meetup or tiktok or hinge or happn or bumble or grindr or Tinder or "inner circle" or periscope or twitch).ab,ti. |
| 8 | 5 or 6 or 7 |
| 9 | high risk behavior/ or "substance use"/ or substance abuse/ |
| 10 | ("substance misuse*" or "substance use behav*" or "risk taking behav*" or "risk-taking behav*" or "risk behav*" or "risk-behav*" or "risky behav*" or "multiple risk behav*").ab,ti. |
| 11 | 9 or 10 |
| 12 | chewing tobacco/ or tobacco/ or smokeless tobacco/ or tobacco dependence/ or "tobacco use"/ or tobacco consumption/ or tobacco snuff/ or cigarette/ or cigarette smoking/ or adolescent smoking/ or smoking/ |
| 13 | ("smoking initiation" or "smoking behav*").ab,ti. |
| 14 | 12 or 13 |
| 15 | exp electronic cigarette/ |
| 16 | ("electronic nicotine delivery system*" or e-cigarette* or Juul or vaping or vape).ab,ti. |
| 17 | 15 or 16 |
| 18 | underage drinking/ or binge drinking/ or alcohol consumption/ or drinking behavior/ or alcohol abstinence/ or alcoholism/ or alcohol abuse/ |
| 19 | ("alcohol intoxication" or "problem drinking" or "alcohol intake" or "alcohol use*" or temperance or "under-age drinking" or "under age drinking" or "underage drinking").ab,ti. |
| 20 | 18 or 19 |
| 21 | "cannabis use"/ or cannabis addiction/ or illicit drug/ or drug abuse/ |
| 22 | ("street drug*" or "drug use*" or "drug misuse*" or weed or skunk or cannabis or marijuana or cocaine or "special k" or crack or methamphetamine* or ecstasy or heroin or LSD or steroid* or ketamine or MDMA or GHB or GBL).ab,ti. |
| 23 | 21 or 22 |
| 24 | antisocial behavior/ or social problem/ or assault/ or physical violence/ or gang/ or fighting/ or theft/ or juvenile delinquency/ |
| 25 | (steal* or shoplift* or vandal* or "public nuisance" or "physical assault" or "anti-social behav*").ab,ti. |
| 26 | 24 or 25 |
| 27 | adolescent pregnancy/ or sexting/ or sexually transmitted disease/ or unwanted pregnancy/ or sexual behavior/ or sexual intercourse/ or acquired immune deficiency syndrome/ or Human immunodeficiency virus/ |
| 28 | ("unwanted pregnancy" or "sexually transmitted infection*" or STIs or STDs or "teen* pregnancy" or "unprotected sex*" or "first intercourse" or "casual sexual relations*" or "intimate sexual contact" or "under age sex" or "underage sex*" or "under-age sex*" or " underage pregnancy" or " under age pregnancy" or "under-age pregnancy" or sex-text or "sex text" or "sexual behav*" or "sexual risk").ab,ti. |
| 29 | 27 or 28 |
| 30 | gambling/ |
| 31 | (betting or gambling).ab,ti. |
| 32 | 30 or 31 |
| 33 | unhealthy diet/ or sugar-sweetened beverage/ or fast food/ or adolescent nutrition/ |
| 34 | ("poor diet*" or "dietary behav*" or "eating behav*" or "sugary drink*" or sweet*).ab,ti. |
| 35 | 33 or 34 |
| 36 | physical inactivity/ or exercise/ or physical activity/ or fitness/ or sedentary lifestyle/ |
| 37 | ("physical inactiv*" or "physical activ*" or exercis* or sport*).ab,ti. |
| 38 | 36 or 37 |
| 39 | 11 or 14 or 17 or 20 or 23 or 26 or 29 or 32 or 35 or 38 |
| 40 | 4 and 8 and 39 |
| 41 | limit 40 to yr="1997 -Current" |

**Legend:** Initial search: date of search- 30.04.2020. Interface- Ovid. Database and coverage- Embase 1947 to present, updated daily. Limits applied- 1997 to 30.04.2020. Updated search: date of search- 06.06.2022. Interface- Ovid. Database and coverage- Embase 1947 to present, updated daily. Limits applied- 2020 to 06.06.2022.

##### Table D. MEDLINE search strategy

| String number | String |
| --- | --- |
| 1 | adolescent/ or child/ or students/ |
| 2 | ("young people" or youth or "school child*" or teen* or "young person*" or "middle school" or "middle-school" or "secondary school" or "high school" or iGen or "generation Z" or "gen Z").ab,ti. |
| 3 | 1 or 2 |
| 4 | online social networking/ or social media/ or smartphone/ or internet/ or screen time/ |
| 5 | (("screen time" or "social media" or "social networking" or "social-networking" or "social network* site*" or "web 2.0" or "online game*" or "online gaming" or "online social gaming" or hashtag or "instant messag*" or instagram or "Whats App" or whatsapp or facebook or twitter or linkedin or youtube or "you tube" or tumblr or vine or snapchat or myspace or bebo or reddit or neknominate or myspace or wickr or telegram or whisper or "kik messenger" or "Tencent QQ" or wechat or meetup or tiktok or hinge or happn or bumble or grindr or Tinder or "inner circle" or periscope or twitch) adj2 (usage or use*)).ab,ti. |
| 6 | ("screen time" or "social media" or "social networking" or "social-networking" or "social network* site*" or "web 2.0" or "online game*" or "online gaming" or "online social gaming" or hashtag or "instant messag*" or instagram or "Whats App" or whatsapp or facebook or twitter or linkedin or youtube or "you tube" or tumblr or vine or snapchat or myspace or bebo or reddit or neknominate or myspace or wickr or telegram or whisper or "kik messenger" or "Tencent QQ" or wechat or meetup or tiktok or hinge or happn or bumble or grindr or Tinder or "inner circle" or periscope or twitch).ab,ti. |
| 7 | 4 or 5 or 6 |
| 8 | Risk-Taking/ or exp Substance-Related Disorders/ |
| 9 | ("substance use*" or "substance abuse*" or "substance misuse*" or "risk taking behav*" or "risk-taking behav*" or "risk behav*" or "risk-behav*" or "risky behav*" or "multiple risk behav*").ab,ti. |
| 10 | 8 or 9 |
| 11 | exp "Tobacco Use"/ or exp Smoking/ or Tobacco/ |
| 12 | ("chewing tobacco" or "smokeless tobacco" or "tobacco dependence" or "tobacco consumption" or "tobacco snuff" or "cigarette smoking" or "adolescent smok*" or "smoking initiation" or "smoking behav*" or cigarette*).ab,ti. |
| 13 | 11 or 12 |
| 14 | Electronic Nicotine Delivery Systems/ |
| 15 | ("electronic cigarette*" or "e-cigarette*" or Juul or vaping or vape).ab,ti. |
| 16 | 14 or 15 |
| 17 | alcohol drinking/ or binge drinking/ or underage drinking/ or drinking behavior/ or alcohol abstinence/ or temperance/ or alcoholism/ |
| 18 | ("alcohol consumption" or "alcohol abuse*" or "alcohol intoxication" or "problem drinking" or "alcohol intake" or "alcohol use*" or "under-age drinking" or "under age drinking" or "underage drinking").ab,ti. |
| 19 | 17 or 18 |
| 20 | "Marijuana Use"/ or Marijuana Abuse/ or exp Illicit Drugs/ or exp Drug Misuse/ |
| 21 | ("cannabis use*" or "cannabis addict*" or "drug abuse*" or "street drug*" or "drug use*" or "drug misuse*" or weed or skunk or cannabis or marijuana or cocaine or "special k" or crack or methamphetamine* or ecstasy or heroin or LSD or steroid* or ketamine or MDMA or GHB or GBL).ab,ti. |
| 22 | 20 or 21 |
| 23 | social problems/ or juvenile delinquency/ or violence/ or theft/ |
| 24 | ("anti-social behav*" or "antisocial behav*" or assault or gang or fight* or steal* or shoplift* or vandal* or "public nuisance" or "physical assault").ab,ti. |
| 25 | 23 or 24 |
| 26 | pregnancy in adolescence/ or pregnancy, unwanted/ or sexual behavior/ or unsafe sex/ or exp Sexually Transmitted Diseases/ or exp HIV infections/ |
| 27 | (sexting or sex-text or "sex text" or "sexual intercourse" or "unwanted pregnancy" or "sexually transmitted infection*" or STIs or STDs or "teen* pregnancy" or "unprotected sex*" or "first intercourse" or "casual sexual relations*" or "intimate sexual contact" or "under age sex" or "underage sex*" or "under-age sex*" or "underage pregnancy" or "under age pregnancy" or "under-age pregnancy" or "sexual behav*" or "sexual risk").ab,ti. |
| 28 | 26 or 27 |
| 29 | Gambling/ |
| 30 | (betting or gambling).ab,ti. |
| 31 | 29 or 30 |
| 32 | Diet/ or Sugar-Sweetened Beverages/ or Fast Foods/ |
| 33 | ("unhealthy diet*" or "adolescent nutrition" or "poor diet*" or "dietary behav*" or "eating behav*" or "sugary drink*" or sweet*).ab,ti. |
| 34 | 32 or 33 |
| 35 | Sedentary Behavior/ or exp Exercise/ or exp Physical Fitness/ |
| 36 | ("physical inactiv*" or "physical activ*" or exercis* or sport*).ab,ti. |
| 37 | 35 or 36 |
| 38 | 10 or 13 or 16 or 19 or 22 or 25 or 28 or 31 or 34 or 37 |
| 39 | 3 and 7 and 38 |
| 40 | limit 39 to yr="1997 -Current" |

**Legend:** Initial search: date of search- 30.04.2020. Interface- Ovid. Database and coverage- Medical Literature Analysis and Retrieval System Online (MEDLINE)(R),1946 to present. Limits applied- 1997 to 30.04.2020. Updated search: date of search- 06.06.2022. Interface- Ovid. Database and coverage- Medical Literature Analysis and Retrieval System Online (MEDLINE)(R),1946 to present. Limits applied- 2020 to 06.06.2022.

##### Table E. APA PsycINFO search strategy

| String number | String |
| --- | --- |
| 1 | (DE "Middle School Students" OR DE "High School Students" OR DE "Students") |
| 2 | TI (adolescent* OR child* OR "young people" OR youth OR "school child*" OR teen* OR "young person*" OR "middle school" OR middle-school OR "secondary school" OR "high school" OR iGen OR "generation Z" OR "gen Z") OR AB (adolescent* OR child* OR "young people" OR youth OR "school child*" OR teen* OR "young person*" OR "middle school" OR middle-school OR "secondary school" OR "high school" OR iGen OR "generation Z" OR "gen Z") |
| 3 | S1 OR S2 |
| 4 | (DE "Online Social Networks" OR DE "Internet" OR DE "Social Media" OR DE "Smartphones" OR DE "Screen Time") |
| 5 | TI ("screen time" OR "social media" OR "social network* site*" OR"social networking" OR "social-networking" OR "web 2.0" OR "online game*" OR "online gaming" OR "online social gaming" OR hashtag OR "instant messag*" OR instagram OR "Whats App" OR whatsapp OR facebook OR twitter OR linkedin OR youtube OR "you tube" OR tumblr OR vine OR snapchat OR myspace OR bebo OR reddit OR neknominate OR myspace OR wickr OR telegram OR whisper OR "kik messenger" OR "Tencent QQ" OR wechat OR meetup OR tiktok OR hinge OR happn OR bumble OR grindr OR Tinder OR "inner circle" OR periscope OR twitch) N2 (usage OR use*)) OR AB ("screen time" OR "social media" OR "social network* site*" OR "social networking" OR "social-networking" OR "web 2.0" OR "online game*" OR "online gaming" OR "online social gaming" OR hashtag OR "instant messag*" OR instagram OR "Whats App" OR whatsapp OR facebook OR twitter OR linkedin OR youtube OR "you tube" OR tumblr OR vine OR snapchat OR myspace OR bebo OR reddit OR neknominate OR myspace OR wickr OR telegram OR whisper OR "kik messenger" OR "Tencent QQ" OR wechat OR meetup OR tiktok OR hinge OR happn OR bumble OR grindr OR Tinder OR "inner circle" OR periscope OR twitch) N2 (usage OR use*)) |
| 6 | TI ("screen time" OR "social media" OR "social network* site*" OR "social networking" OR "social-networking" OR "web 2.0" OR "online game*" OR "online gaming" OR "online social gaming" OR hashtag OR "instant messag*" OR instagram OR "Whats App" OR whatsapp OR facebook OR twitter OR linkedin OR youtube OR "you tube" OR tumblr OR vine OR snapchat OR myspace OR bebo OR reddit OR neknominate OR myspace OR wickr OR telegram OR whisper OR "kik messenger" OR "Tencent QQ" OR wechat OR meetup OR tiktok OR hinge OR happn OR bumble OR grindr OR Tinder OR "inner circle" OR periscope OR twitch) OR AB ("screen time" OR "social media" OR "social networking" OR "social network* site*" OR "social-networking" OR "web 2.0" OR "online game*" OR "online gaming" OR "online social gaming" OR hashtag OR "instant messag*" OR instagram OR "Whats App" OR whatsapp OR facebook OR twitter OR linkedin OR youtube OR "you tube" OR tumblr OR vine OR snapchat OR myspace OR bebo OR reddit OR neknominate OR myspace OR wickr OR telegram OR whisper OR "kik messenger" OR "Tencent QQ" OR wechat OR meetup OR tiktok OR hinge OR happn OR bumble OR grindr OR Tinder OR "inner circle" OR periscope OR twitch) |
| 7 | S4 OR S5 OR S6 |
| 8 | (DE "Risk Taking" OR DE "Substance Use Disorder") |
| 9 | TI ( "substance use*" OR "substance abuse*" OR "substance misuse*" OR "risk-behav*" OR "risk taking behav*" OR "risk-taking behav*" OR "risk behav*" OR "risky behav*" OR "multiple risk behav*") OR AB ("substance use*" OR "substance abuse*" OR "substance misuse*" OR "risk-behav*" OR "risk taking behav*" OR "risk-taking behav*" OR "risk behav*" OR "risky behav*" OR "multiple risk behav*") |
| 10 | S8 OR S9 |
| 11 | (DE "Tobacco Smoking" OR DE "Smokeless Tobacco" OR DE "Tobacco Use Disorder") |
| 12 | TI ("adolescent smok*" OR "chewing tobacco" OR "tobacco dependence" OR "tobacco use" OR "tobacco consumption" OR "tobacco snuff" OR cigarette* OR "smoking initiation" OR "smoking behav*") OR AB ("adolescent smok*" OR "chewing tobacco" OR "tobacco dependence" OR "tobacco use" OR "tobacco consumption" OR "tobacco snuff" OR cigarette* OR "smoking initiation" OR "smoking behav*") |
| 13 | S11 OR S12 |
| 14 | DE "Electronic Cigarettes" |
| 15 | TI ("electronic nicotine delivery system*" OR e-cigarette* OR Juul OR vaping OR vape) OR AB ( "electronic nicotine delivery system*" OR e-cigarette* OR Juul OR vaping OR vape) |
| 16 | S14 OR S15 |
| 17 | (DE "Underage Drinking" OR DE "Binge Drinking" OR DE "Drinking Behavior" OR DE "Alcohol Abuse" OR DE "Alcohol Drinking Patterns" OR DE "Alcohol Intoxication" OR DE "Sobriety" OR DE "Alcoholism" OR DE "Alcohol Use Disorder") |
| 18 | TI ("alcohol consumption" OR "alcohol abstinence" OR "alcohol abuse*" OR "alcohol use*" OR "problem drinking" OR "alcohol intake" OR temperance OR "under-age drinking" OR "under age drinking" OR "underage drinking") OR AB ("alcohol consumption" OR "alcohol abstinence" OR "alcohol abuse*" OR "alcohol use*" OR "problem drinking" OR "alcohol intake" OR temperance OR "under-age drinking" OR "under age drinking" OR "underage drinking") |
| 19 | S17 OR S18 |
| 20 | DE "Cannabis" OR DE "Hashish" OR DE "Marijuana" OR DE "Cannabinoids" OR DE "Cannabis Use Disorder" OR DE "Drug Abuse" OR DE "Cocaine" OR DE "Drug Usage" |
| 21 | TI ("street drug*" OR"cannabis use*" OR "cannabis addict*" OR "illicit drug*" OR "drug abuse*" OR "drug use*" OR "drug misuse*" OR weed OR skunk OR marijuana OR "special k" OR crack OR methamphetamine* OR ecstasy OR heroin OR LSD OR steroid* OR ketamine OR MDMA OR GHB OR GBL) OR AB ("street drug*" OR "cannabis use*" OR "cannabis addict*" OR "illicit drug*" OR "drug abuse*" OR "drug use*" OR "drug misuse*" OR weed OR skunk OR marijuana OR "special k" OR crack OR methamphetamine* OR ecstasy OR heroin OR LSD OR steroid* OR ketamine OR MDMA OR GHB OR GBL) |
| 22 | S20 OR S21 |
| 23 | DE "Antisocial Behavior" OR DE "Juvenile Delinquency" OR DE "Violence" OR DE "Gangs" OR DE "Social Issues" OR DE "Theft" OR DE "Vandalism" |
| 24 | TI ("anti-social behav*" OR shoplift* OR "social problem*" OR assault OR fighting OR steal* OR "public nuisance") OR AB ("anti-social behav*" OR shoplift* OR "social problem*" OR assault OR fighting OR steal* OR "public nuisance") |
| 25 | S23 OR S24 |
| 26 | DE "Adolescent Pregnancy"OR DE "Sexting" OR DE "Sexually Transmitted Diseases" OR DE "Sexual Risk Taking" OR DE "Sexual Intercourse (Human)" OR DE "AIDS" OR DE "HIV" |
| 27 | TI ("unwanted pregnancy" OR "sexually transmitted infection*" OR STIs OR STDs OR "teen* pregnancy" OR "unprotected sex" OR "first intercourse" OR "casual sexual relations*" OR "intimate sexual contact" OR "underage sex" OR "under-age sex" OR "under age sex" OR "underage pregnancy" OR "under-age pregnancy" OR "under age pregnancy" OR sex-text OR "sex text") OR AB ("unwanted pregnancy" OR "sexually transmitted infection*" OR STIs OR STDs OR "teen* pregnancy" OR "unprotected sex" OR "first intercourse" OR "casual sexual relations*" OR "intimate sexual contact" OR "underage sex" OR "under-age sex" OR "under age sex" OR "underage pregnancy" OR "under-age pregnancy" OR "under age pregnancy" OR sex-text OR "sex text") |
| 28 | S26 OR S27 |
| 29 | DE "Gambling" |
| 30 | TI ("betting OR gambling) OR AB ("betting OR gambling) |
| 31 | S29 OR S30 |
| 32 | DE "Diets" OR DE "Eating Behavior" OR DE "Fast Food" |
| 33 | TI ("unhealthy diet*" OR "poor diet*" OR "dietary behav*" OR "sugary drink*" OR sweet* OR "sugar-sweetened beverage*" OR "sugar sweetened beverage*" OR "adolescent nutrition") OR AB ("unhealthy diet*" OR "poor diet*" OR "dietary behav*" OR "sugary drink*" OR sweet* OR "sugar-sweetened beverage*" OR "sugar sweetened beverage*" OR "adolescent nutrition") |
| 34 | S32 OR S33 |
| 35 | (DE "Physical Activity" OR DE "Exercise" OR DE "Physical Fitness" OR DE "Sedentary Behavior") |
| 36 | TI ("physical inactiv*" OR "physical activ*" OR exercis* OR sport*) OR AB ("physical inactiv*" OR "physical activ" OR exercis* OR sport*) |
| 37 | S35 OR S36 |
| 38 | S10 OR S13 OR S16 OR S19 OR S22 OR S25 OR S28 OR S31 OR S34 OR S37 |
| 39 | S3 AND S7 AND S38 |

**Legend:** Initial search: date of search- 30.04.2020. Interface- EBSCOhost. Database and coverage- APA PsycINFO,1800s to present. Limits applied- 01.01.1997 to 30.04.2020. Updated search: date of search- 06.06.2022. Interface- EBSCOhost. Database and coverage- APA PsycINFO,1800s to present. Limits applied- 01.04.2020 to 31.06.2022.

##### Table F. SocINDEX search strategy

| String number | String |
| --- | --- |
| 1 | DE "STUDENTS" OR DE "MIDDLE school students" OR DE "HIGH school students" OR DE "ADOLESCENCE" OR DE "CHILDREN" OR DE "TEENAGERS" OR DE "YOUTH" |
| 2 | TI ("young people" OR "school child*" OR teen* OR "young person*" OR "middle school" OR middle-school OR "secondary school" OR "high school" OR iGen OR "generation Z" OR "gen Z") OR AB ("young people" OR "school child*" OR teen* OR "young person*" OR "middle school" OR middle-school OR "secondary school" OR "high school" OR iGen OR "generation Z" OR "gen Z") |
| 3 | S1 OR S2 |
| 4 | DE "SOCIAL media" OR DE "INTERNET" OR DE "SOCIAL networking mobile apps" |
| 5 | TI ("smart phone" OR smartphone OR "screen time" OR "social media" OR "social network* site*" OR "social networking" OR "social-networking" OR "web 2.0" OR "online game*" OR "online gaming" OR "online social gaming" OR hashtag OR "instant messag*" OR instagram OR "Whats App" OR whatsapp OR facebook OR twitter OR linkedin OR youtube OR "you tube" OR tumblr OR vine OR snapchat OR myspace OR bebo OR reddit OR neknominate OR myspace OR wickr OR telegram OR whisper OR "kik messenger" OR "Tencent QQ" OR wechat OR meetup OR tiktok OR hinge OR happn OR bumble OR grindr OR Tinder OR "inner circle" OR periscope OR twitch) N2 (usage or use*)) OR AB ("smart phone" OR smartphone OR "screen time" OR "social media" OR "social network* site*" OR "social networking" OR "social-networking" OR "web 2.0" OR "online game*" OR "online gaming" OR "online social gaming" OR hashtag OR "instant messag*" OR instagram OR "Whats App" OR whatsapp OR facebook OR twitter OR linkedin OR youtube OR "you tube" OR tumblr OR vine OR snapchat OR myspace OR bebo OR reddit OR neknominate OR myspace OR wickr OR telegram OR whisper OR "kik messenger" OR "Tencent QQ" OR wechat OR meetup OR tiktok OR hinge OR happn OR bumble OR grindr OR Tinder OR "inner circle" OR periscope OR twitch) N2 (usage or use*)) |
| 6 | TI ("smart phone" OR "smartphone" OR "screen time" OR "social media" OR "social network* site*" OR "social networking" OR "social-networking" OR "web 2.0" OR "online game*" OR "online gaming" OR "online social gaming" OR hashtag OR "instant messag*" OR instagram OR "Whats App" OR whatsapp OR facebook OR twitter OR linkedin OR youtube OR "you tube" OR tumblr OR vine OR snapchat OR myspace OR bebo OR reddit OR neknominate OR myspace OR wickr OR telegram OR whisper OR "kik messenger" OR "Tencent QQ" OR wechat OR meetup OR tiktok OR hinge OR happn OR bumble OR grindr OR Tinder OR "inner circle" OR periscope OR twitch) OR AB ("smart phone" OR "smartphone" OR "screen time" OR "social media" OR "social network* site*" OR "social networking" OR "social-networking" OR "web 2.0" OR "online game*" OR "online gaming" OR "online social gaming" OR hashtag OR "instant messag*" OR instagram OR "Whats App" OR whatsapp OR facebook OR twitter OR linkedin OR youtube OR "you tube" OR tumblr OR vine OR snapchat OR myspace OR bebo OR reddit OR neknominate OR myspace OR wickr OR telegram OR whisper OR "kik messenger" OR "Tencent QQ" OR wechat OR meetup OR tiktok OR hinge OR happn OR bumble OR grindr OR Tinder OR "inner circle" OR periscope OR twitch) |
| 7 | S4 OR S5 OR S6 |
| 8 | DE "RISK-taking behavior" OR DE "SUBSTANCE abuse" |
| 9 | TI ("substance misuse*" OR "substance use*" OR risk-behav* OR "risk behav*" OR "risky behav*" OR "risk-taking behav*" OR "risk taking behav*" OR "multiple risk behav*") OR AB ("substance misuse*" OR "substance use*" OR risk-behav* OR "risk behav*" OR "risky behav*" OR "risk-taking behav*" OR "risk taking behav*" OR "multiple risk behav*") |
| 10 | S8 OR S9 |
| 11 | DE "SMOKING" OR DE "TOBACCO use" OR DE "CIGARETTE smokers" OR DE "CIGARETTES" |
| 12 | TI ("adolescent smok*" OR "chewing tobacco" OR "tobacco dependence" OR "tobacco consumption" OR "tobacco snuff" OR "smoking initiation" OR "smoking behav*") OR AB ( ("adolescent smok*" OR "chewing tobacco" OR "tobacco dependence" OR "tobacco consumption" OR "tobacco snuff" OR "smoking initiation" OR "smoking behav*") |
| 13 | S11 OR S12 |
| 14 | TI ("electronic nicotine delivery system*" OR "electronic cigarette*" OR e-cigarette* OR Juul OR vaping OR vape) OR AB ("electronic nicotine delivery system*" OR "electronic cigarette*" OR e-cigarette* OR Juul OR vaping OR vape) |
| 15 | DE "UNDERAGE drinking" OR DE "BINGE drinking" OR DE "ALCOHOL drinking" OR DE "ALCOHOLIC intoxication" OR DE "DRINKING behavior" OR DE "ALCOHOLISM" OR DE "TEMPERANCE" OR DE "YOUTH & alcohol" |
| 16 | TI ("alcohol consumption" OR "alcohol abstinence" OR "alcohol abuse*" OR "under age drinking" OR "underage drinking" OR "under-age drinking" OR "alcohol use*" OR "alcohol intake" OR "problem drinking") OR AB ("alcohol consumption" OR "alcohol abstinence" OR "alcohol abuse*" OR "under age drinking" OR "underage drinking" OR "under-age drinking" OR "alcohol use*" OR "alcohol intake" OR "problem drinking") |
| 17 | S15 OR S16 |
| 18 | DE "MARIJUANA abuse" OR DE "DRUG abuse" OR DE "MARIJUANA" OR DE "DRUGS of abuse" OR DE "DRUGS" OR DE "COCAINE" OR DE "COCAINE abuse" |
| 19 | TI ("drug use*" OR "drug misuse*" OR "drug abuse*" OR "illicit drug*" OR "cannabis use*" OR "cannabis addict*" OR "illicit drug*" OR weed OR skunk OR marijuana OR "special k" OR crack OR methamphetamine* OR ecstasy OR heroin OR LSD OR steroid* OR ketamine OR MDMA OR GHB OR GBL) OR AB ("drug use*" OR "drug misuse*" OR "drug abuse*" OR "illicit drug*" OR "cannabis use*" OR "cannabis addict*" OR "illicit drug*"OR weed OR skunk OR marijuana OR "special k" OR crack OR methamphetamine* OR ecstasy OR heroin OR LSD OR steroid* OR ketamine OR MDMA OR GHB OR GBL) |
| 20 | S18 OR S19 |
| 21 | DE "JUVENILE delinquency" OR DE "SCHOOL violence" OR DE "GANGS" OR DE "SCHOOL vandalism" OR DE "YOUTH gangs" OR DE "SOCIAL problems" |
| 22 | TI ("antisocial behav*" OR "anti-social behav*" OR assault OR fighting OR steal* OR shoplift* OR vandal* OR "public nuisance") OR AB ("antisocial behav*" OR "anti-social behav*" OR assault OR fighting OR steal* OR shoplift* OR vandal* OR "public nuisance") |
| 23 | S21 OR S22 |
| 24 | DE "TEENAGE pregnancy" OR DE "SEXUALLY transmitted diseases" OR DE "UNWANTED pregnancy" OR "SEXUAL intercourse" OR DE "AIDS" |
| 25 | TI ("adolescent pregnancy" OR "teen* pregnancy" OR sexting OR sex-text OR "sex text" OR "sexual behav*" OR "human immunodeficiency virus" OR HIV OR "sexually transmitted infection*" OR STIs OR STDs OR "unprotected sex" OR "first intercourse" OR "casual sexual relations*" OR "intimate sexual contact" OR "under age sex" OR "under-age sex" OR "underage sex" OR "under age pregnancy" OR "underage pregnancy" OR "under-age pregnancy") OR AB ("adolescent pregnancy" OR "teen* pregnancy" OR sexting OR sex-text OR "sex text" OR "sexual behav*" OR "human immunodeficiency virus" OR HIV OR "sexually transmitted infection*" OR STIs OR STDs OR "unprotected sex" OR "first intercourse" OR "casual sexual relations*" OR "intimate sexual contact" OR "under age sex" OR "under-age sex" OR "underage sex" OR "under age pregnancy" OR "underage pregnancy" OR "under-age pregnancy") |
| 26 | S24 OR S25 |
| 27 | DE "GAMBLING behavior" |
| 28 | TI (betting OR gambling) OR AB (betting OR gambling) |
| 29 | S27 OR S28 |
| 30 | TI ("unhealthy diet*" OR "sugar-sweetened beverage*" OR "sugar sweetened beverage*" OR "fast food" OR "adolescent nutrition" OR "poor diet*" OR "dietary behav*" OR "eating behav*" OR "sugary drink*" OR sweet*) OR AB ("unhealthy diet*" OR "sugar-sweetened beverage*" OR "sugar sweetened beverage*" OR "fast food" OR "adolescent nutrition" OR "poor diet*" OR "dietary behav*" OR "eating behav*" OR "sugary drink*" OR sweet*) |
| 31 | DE "PHYSICAL fitness" OR DE "EXERCISE" |
| 32 | TI ("physical inactiv*" OR "physical activ*" OR exercis* OR sport* OR sedentary) OR AB ( ("physical inactiv*" OR "physical activ*" OR exercis* OR sport* OR sedentary) |
| 33 | S31 OR S32 |
| 34 | S10 OR S13 OR S14 OR S17 OR S20 OR S23 OR S26 OR S29 OR S30 OR S33 |
| 35 | S3 AND S7 AND S34 |

**Legend:** Initial search: date of search- 30.04.2020. Interface- EBSCOhost. Database and coverage- SocINDEX with Full Text, 1908 to present. Limits applied- 01.01.1997 to 30.04.2020. Updated search: date of search- 06.06.2022. Interface- EBSCOhost. Database and coverage- SocINDEX with Full Text, 1908 to present. Limits applied- 01.04.2020 to 31.06.2022.

##### Table G. Social Science Research Network (SSRN e-library vis SSRN) search strategy

| String | Initial search: records identified for screening | Updated search: records identified for screening |
| --- | --- | --- |
| (child* OR adolescent*) AND ("online social network*" OR "social media" OR "social network* site") AND risk behav* | 0 | 0 |
| (child* OR adolescent*) AND ("online social network*" OR "social media" OR "social network* site") AND (alcohol, OR drug*OR e-cigarette*OR smok* OR tobacco OR sex* OR diet* OR "physical inactiv*" OR antisocial OR anti-social OR gambl*) | 0 | 0 |
| (child* OR adolescent*) AND (facebook OR twitter OR instagram) AND risk behav* | 0 | 0 |
| (child* OR adolescent*) AND (facebook OR twitter OR instagram) AND (alcohol, OR drug*OR e-cigarette*OR smok* OR tobacco OR sex* OR diet* OR "physical inactiv*" OR antisocial OR anti-social OR gambl*) | 0 | 0 |

**Legend:** Initial search: date of search- 30.04.2020. Search/limits options- title, abstract and key words (no other limits available). Updated search: date of search- 06.06.2022. Search/limits options- title, abstract and key words (no other limits available).

##### Table H. SocArXic Preprints search strategy

| String | Initial search: records identified for screening | Updated search: records identified for screening |
| --- | --- | --- |
| (child* OR adolescent* OR student* OR juvenile OR youth OR "young people" OR teen*) AND ("online social network*" OR "social media" OR internet OR "smart phone" OR "screen time" OR "social network* site" OR "social networking" OR "social-networking" OR facebook OR twitter OR instagram) AND (risk behav* OR "substance use" OR alcohol OR drink* OR tobacco OR smok* OR drug* OR e-cigarette* OR cannabis OR "antisocial behav*" OR "sexually transmitted disease*" OR "sexual behav*" OR sexting OR gambl* OR "unhealthy diet*" OR "physical inactiv*" OR sedentary) | 11 | 10 |

**Legend:** Initial search: date of search- 30.04.2020. Search/limits- no limit/filters/advance search option available. Updated search: date of search- 06.06.2022. Search/limits- no limit/filters/advance search option available.

##### Table I. PsyArXiv Preprints search strategy

| String | Initial search: records identified for screening | Updated search: records identified for screening |
| --- | --- | --- |
| (child* OR adolescent* OR student* OR juvenile OR youth OR "young people" OR teen*) AND ("online social network*" OR "social media" OR internet OR "smart phone" OR "screen time" OR "social network* site" OR "social networking" OR "social-networking" OR facebook OR twitter OR instagram) AND (risk behav* OR "substance use" OR alcohol OR drink* OR tobacco OR smok* OR drug* OR e-cigarette* OR cannabis OR "antisocial behav*" OR "sexually transmitted disease*" OR "sexual behav*" OR sexting OR gambl* OR "unhealthy diet*" OR "physical inactiv*" OR sedentary) | 6 | 35 |

**Legend:** Initial search: date of search- 30.04.2020. Search/limits- no limit/filters/advance search option available. Updated search: date of search- 06.06.2022. Search/limits- no limit/filters/advance search option available.

##### Table J. MedRxiv Preprints search strategy

| String | Initial search: records identified for screening | Updated search: records identified for screening |
| --- | --- | --- |
| child* AND ("social network* site") AND risk behav* | 18 | 0 |
| child* AND ("social network* site") AND (alcohol, OR drug*OR e-cigarette*OR smok* OR tobacco OR sex* OR diet* OR "physical inactiv*" OR antisocial OR anti-social OR gambl*) | 0 | 0 |
| adolescent*AND ("social network* site") AND risk behav* | 0 | 0 |
| adolescent*AND ("social network* site") AND (alcohol, OR drug*OR e-cigarette*OR smok* OR tobacco OR sex* OR diet* OR "physical inactiv*" OR antisocial OR anti-social OR gambl*) | 0 | 0 |
| child* AND ("social media") AND risk behav* | 0 | 0 |
| child* AND ("social media") AND (alcohol, OR drug*OR e-cigarette*OR smok* OR tobacco OR sex* OR diet* OR "physical inactiv*" OR antisocial OR anti-social OR gambl*) | 0 | 0 |
| adolescent*AND ("social media") AND risk behav* | 0 | 0 |
| adolescent*AND ("social media") AND (alcohol, OR drug*OR e-cigarette*OR smok* OR tobacco OR sex* OR diet* OR "physical inactiv*" OR antisocial OR anti-social OR gambl*) | 0 | 0 |

**Legend:** Initial search: date of search- 30.04.2020. Search/limits- limited to 01.01.1997-30.04.2020; title, abstract and all terms. Updated search: date of search- 06.06.2022. Search/limits- limited to 30.04.2020-06.06.2022; title, abstract and all terms.

##### Table K. Google scholar via Google search strategy

| String | Initial search: records identified for screening | Updated search: records identified for screening |
| --- | --- | --- |
| (adolescent OR child) AND ("social media") AND ("risk behaviour" OR “risk behavior”) | First 30 records | First 30 records |

**Legend:** Initial search: date of search- 30.04.2020. Search/limits- limited to 1997-2020; advanced search function “find all words” selected.

Updated search: date of search- 06.06.2022. Search/limits- limited to 2020-2022; advanced search function “find all words” selected.

### Appendix-3. Potentially relevant non-English reports

##### Table A. Record of potentially relevant non-English records excluded at full-text screening

| Citation | Language |
| --- | --- |
| Blasco V and Bernal S. Patrón de uso de internet y control parental de redes sociales como predictor de sexting en adolescentes: una perspectiva de género. *Revista de Psicología y Educación*. 2019;14(1):16-26 | Spanish |
| Blazquez Barba M, Gomez Romero D, Frontaura Fernández I, Camacho Ojeda A, Rodriguez Salas FM, Toriz Cano H. Use of new technologies by adolescents in the search for health information. *Atencion Primaria*. 2018;50(9):547-552 | Spanish |
| González M, Fernández ME, Urturi A, et al. Use and risks of information and communication technologies in the adolescents from 13 to 18 years*. Acta Pediatrica Espanola*. 2015; 73:146-151 | Spanish |
| Marotta R, Rapetto U, Vismara MFM, et al. Impact and risks of new information technologies in adolescents: results of a survey conducted on 1534 subjects. G di *Neuropsichiatr dell’Età Evol*. 2018;38(1):9–13 | Spanish |
| Pedersen W. Mobile phones, web chat, and sex among Norwegian adolescents. *Tidsskr Nor Laegeforen*. 2004;1;124(13-14):1756-1759 | Norwegian |
| Richter M, Heilmann K, Moor I. The good, the bad and the ugly: the relationship between social media use, subjective health and risk behavior among children and adolescents. *Gesundheitswesen*. 2020;83(3):198-207 | German |
| Stulhofer A, Vukasović T, Perišić K, [Sušac](https://www.researchgate.net/profile/Nika-Susac-2) N, [Marjanović](https://www.researchgate.net/scientific-contributions/Bojan-MARJANOVIC-2093595794) B, Bauer M, et al. Internet and sexual compulsivity. *Socijalna Psihijatrija*. 2005; 33:190-200 | Croatian |

### Appendix-4. Process of social media categorisation

All social media categories outlined in the SAGE Social Media Categorisation^3^ (Table A) were eligible for inclusion. Online (social) gambling and online (social) gaming were deemed eligible exposures due to their inclusion of core social media functionalities, namely interaction between users.^4,5,6^

As the functionalities of social media platforms overlap, and social media platforms may fall into several social media categories, we made efforts to categorise using the initial premise/purpose of the platform if stated. For example, the social media platform Instagram possesses functionalities central to social networking sites however its initial premise/purpose was to facilitate media-sharing. Therefore, where a study reported Instagram use, this was classified under the social media category media-sharing. Where an included study reported use of social media overall, this was classified as general social media use. Thus, we attempted to apply a consistent process to classification drawing upon the information reported within studies. This was conducted during data extraction by the lead author (AKP) and then reviewed by a second reviewer.

A similar process was applied when classifying the type of health-risk behaviour content (user-generated or marketer-generated content) for those datapoints investigating exposure to health-risk behaviour content on social media. Where the exposure pertained to exposure to advertisements, marketing, or influencer content, marketer-generated content was selected. Where the exposure pertained to user/peer posts displaying risk behaviour content, user-generated content was selected. Where there was insufficient information reported to facilitate accurate classification, both marketer and user-generated content were selected and the datapoint was not used in any subsequent stratified analyses or meta-regression.

We also made efforts to classify reported exposures into those assessing active social media use (online behaviours which facilitate direct exchange among users, e.g., commenting, liking, sending messages and otherwise engaging with other users) and passive use (monitoring of others/content without direct engagement, e.g., browsing/scrolling).^7^

##### Table A. Social media categories eligible for inclusion

| Social media category | Definitions | Example platforms |
| --- | --- | --- |
| Social media categories outlined in SAGE social media categorisation | | |
| Social networking sites | Web-based services which facilitate individual construction of a public or semi-public profile within a bounded system, compose a list of other users with which they share a connection, and view and traverse their list of connections as well as those created by others within the system.^8^ | Facebook  Snapchat  Instagram  WhatsApp  Twitter  LinkedIn  WeChat  Chat rooms  Instant messaging |
| Microblogging sites | Services which are centred on short updates which are forwarded to anyone subscribed to receive the updates.^3^ | Twitter  Tumblr |
| Blogs and forums | Online forums which allow forum members to have conversations by posting messages. Blog comments are attached to blogs and usually the discussion is focussed on the topic of the blog post.^3^ | LiveJournal  WordPress |
| Media-sharing sites | Services which facilitate uploading and sharing of media including pictures and videos. The majority of services have other social features such as profiles, commenting etc.^3^ | YouTube  Pinterest  Instagram  Snapchat  Facebook |
| Geo-location-based sites | Services which allow users to connect and exchange messages based on their location.^9^ | Foursquare  Tinder |
| Bookmarking sites | A website which ranks references (bookmarks) to other websites contributed by users who use the site. Users can add comments to the bookmarks and make then private or public. The act of bookmarking indicates to others that an individual is interested in a given resource.^10^ | Delicious  StumbleUpon  Twitter |
| Social news sites | Services that allow individuals to post news items or links to outside articles and then facilitates user voting of the items. The voting is the primary social aspect, as items which get the most votes are displayed the most prominently. The community of users decide which news items are seen my more people.^3^ | Reddit  Digg |
| Collaborative authoring sites | Web-based services which allow users to create content and allow individuals with access to the service to modify, edit or review that content.^11^ | Wikipedia  Google Docs |
| Web conferencing | An umbrella term for types of online collaborative services including web-seminars (webinars), webcasts, and peer-level web meetings.^12^ | Skype  Zoom |
| Scheduling and meeting | Web-based services which facilitate group-based decisions regarding event.^3^ | Microsoft Outlook  Doodle  Google Calendar |
| Additional social media categories eligible for inclusion | | |
| Online gaming (social gaming) | A video game which offers online interactions with other players.^13^ | Multiplayer role-playing games (e.g., World of Warcraft)  Social instant games (e.g., Candy Crush) |
| Online gambling (social gambling) | Any kind of gambling conducted on social media/online platforms, which have varying degrees of social game features^4^ (not including gambling via internet websites with no social game/social media features).^4^ | Social networking sites real money gambling (e.g., Bingo Appy via Facebook)  Simulated gambling via social networking site applications (e.g., City Ville) |

### Appendix-5. Advisory group

We established an advisory group of experts and policy makers in the field of social media and adolescent health-risk behaviours to provide guidance during protocol development and the review stages. Recruited via expert stakeholders, members included patient/public representatives and stakeholders from policy, non-governmental, and academic sectors (Table A). In line with Grading of Recommendations Assessment, Development, and Evaluation (GRADE) approach,^14^ the advisory group members ranked pre-selected outcomes according to their relative importance on a 9-point Likert scale (categories: 1-3 – of limited importance; 4 to 6 – importance; 7 to 9 – critical),^14^ completed via an online survey (Table B). The review advisory group members were provided with detailed background information on the review. At the protocol stage, group members were requested to provide feedback on several factors including the relevance of the review’s question, population focus, search strategy, ongoing or published studies, and grey literature selection.^15^ Feedback was received during in person meetings, Zoom or via email. During the review stage advisory group members were contacted to identify relevant ongoing, planned, and unpublished studies.

##### Table A. Advisory group members

| Name | Organisation | Period of involvement |
| --- | --- | --- |
| Kirsty Blenkins | UK Health Security Agency, Office for Health Improvement and Disparities, London, UK | 2020 to present |
| Lee Carlton | Public Health Scotland, Glasgow, UK | 2020 to present |
| Neil Coles | We Are With You, Kent, UK | 2020 to February 2021 |
| Nicholas Hickmott | We Are With You, Kent, UK | 2020 to present |
| Professor John Holmes | Alcohol Policy, University of Sheffield, UK | 2020 to present |
| Rachel Macpherson | Scottish Government, Edinburgh, UK | 2020 to present |
| Dr Ross Whitehead | Public Health Scotland, Edinburgh, UK | 2020 to July 2021 |
| Dr Richard Purves | University of Stirling, Stirling, UK | 2020 to present |

##### Table B. Feedback from advisory group members (online survey)

| Rank outcomes according to their relative importance for the scope of the reviews and general public health decision-making in the context of social media use ^a,b^ | | | |
| --- | --- | --- | --- |
| Outcome | **Mean score** | **Rank** | **Rating** |
| Multiple risk behaviours | 7.50 | 1 | Critical |
| Alcohol use | 7.16 | 2 | Critical |
| Drug use | 7.00 | 3 | Critical |
| Tobacco use | 6.50 | 4 | Important |
| Use of ENDS | 5.83 | 5 | Important |
| Sexual risk behaviours | 5.60 | 6 | Important |
| Gambling | 5.16 | 7 | Important |
| Antisocial behaviour | 5.00 | 8 | Important |
| Inadequate physical activity | 5.00 | 8 | Important |
| Unhealthy dietary behaviours | 5.00 | 8 | Important |
| How well do the presented outcomes cover the review scope? | | | |
| Answers | **Rating** | **Number of responses** | |
| Important outcomes presented | 71% | 5 | |
| Important outcomes missing | 29% | 2 | |
| Comments on missing outcomes (2): | (1) Selling and advertising of illicit substances (2) Mental health-related outcomes and bullying | | |

**Legend:** ^a^ 9-point Likert scale (categories: 1 to 3- of limited importance; 4 to 6- important; 7 to 9 – critical). ^b^ Seven members of the advisory group responded to the survey. Abbreviations: ENDS = Electronic nicotine delivery systems.

### Appendix-6. Included outcomes

##### Table A. Definitions and illustrative examples of included outcomes

| Outcome | Definition | Illustrative examples |
| --- | --- | --- |
| Multiple risk behaviours | Two or more of the below outcomes. | Substance use (alcohol, tobacco, and drug use) |
| Alcohol use | The drinking of beverages containing ethyl alcohol.^16^ | Weekly alcohol use  Frequency of alcohol use  Problem, binge, or hazardous drinking |
| Drug use | Use of drugs for psychotropic rather than medical purposes, potentially including both legal and illegal substances.^17^ | Ever used cannabis  Illicit drug use  Frequency of drug use |
| Tobacco use | The practice of smoking tobacco and inhaling tobacco smoke.^18^ | Ever smoked a cigarette  Frequency of tobacco use |
| Use of electronic nicotine delivery systems (ENDS) | Umbrella term for vapes, vaporisers, vape pens, e-cigarettes, and e-pipes. ENDS are non-combustible tobacco products which use an e-liquid, containing nicotine.^19^ | Ever tried an e-cigarette  Frequency of e-cigarette use |
| Sexual risk behaviour | Initiation of sexual activity at an early age, engaging in unnatural or unprotected sexual intercourse, having sexual intercourse with multiple partners, engaging in paid or irregular or incentive-driven sex or sexual intercourse with an injecting drug user or under the influence (especially intoxication) of psychoactive substances, which may result in sexually transmitted infections, unintended/early pregnancies (or abortions), or legal or interpersonal conflicts.^20^ | Early age of sexual debut  Transactional sex  Unprotected sex  Sexual intercourse with multiple partners  Posting, sharing, or exchanging sexual content using electronic devices |
| Gambling (not via SM) | Placing something of value (usually but not always money) in hope of acquiring something of greater value.^21^ | Problem gambling  Pathological gambling  Internet gambling (not via social media) |
| Unhealthy dietary behaviour | Umbrella term referring to all phenomena related to food choice, eating behaviour, and dietary intake/nutrition. Disordered eating not considered^22^ | Low level of fruit and vegetable consumption  High fat/sugar/salt diet  Low-fibre diet |
| Inadequate physical activity | Doing no or very little physical activity at work, at home, for transport or in discretionary time.^23^ | Physically active for <60 minutes per day on <5 days a week  Low levels of physical activity |
| Anti-social behaviour | Any action which violates social norms in ways which reflect disregard for others, or which reflect the violation of another’s rights.^16^ | Violence  Criminal damage  Graffiti/vandalism  Aggregated assault  Assault with or without injury  Stealing/theft  Carrying a weapon |

### Appendix-7. Meta-analyses and synthesis without meta-analysis (SWiM) decision rules

The below guidance outlines the decision rules used when selecting datapoints/studies for inclusion in meta-analyses and synthesis without meta-analysis (SWiM).

**Duplicate studies (i.e., those looking at the same population, exposure combination, outcomes and during overlapping time period)**

- Select study with longest follow up period
- If studies have the same follow up period, select largest (or most representative)
- If studies are the same size, choose the most recent

Where a study includes multiple repeat cross-sectional samples, or investigates multiple study populations from different settings, these should be entered as separate datapoints, however, will be classified under the primary study from which they originate.

There may be instances of duplicate datapoints, but if some datapoints are unique (investigate different outcomes) between studies, the overall study should be retained, and individual duplicate datapoints will be removed at the next stage.

**Duplicate/overlapping datapoints**

Eight options:

1. Include in meta-analysis
2. Include in stratified analysis (sensitivity/subgroup analysis)
3. Include in meta-analysis and stratified analysis
4. Include in SWiM
5. Include in meta-analysis and SWiM
6. Include in stratified analysis and SWiM
7. Include in all analyses
8. Exclude from all analyses (where sex-stratified datapoints are selected for inclusion and whole sample datapoint is not used)

**Selection of datapoints for meta-analysis**

- In ensuring independence of data, only one effect size per outcome from each study should be used in each meta-analysis. However, datapoints not included in meta-analyses due to potential double counting of participants may be used within stratified analyses
- Meta-analysis should be performed for each exposure (time spent on social media, frequency of social media use and exposure to health-risk behaviour content on social media), by common metric (standardised beta, standardised mean difference, and odds ratio), and by outcome
- Meta-analysis should be conducted when ≥3 datapoints are available for a specific synthesis
- Meta-analyses should be conducted at the datapoint/outcome level, and all forest plots presented should report the risk of bias grade at the datapoint/outcome level

**Exposure**

*Multiple comparison groups:*

Select the largest comparator group, unless this would affect the comparability of results within studies, in which case select the most common/unifying comparison group. Where a common/unifying comparison group cannot be identified for a datapoint, report in SWiM.

*Same outcome assessed by multiple exposures:*

The most used exposure should be selected (i.e., the exposure for which most studies contribute a result). The below criteria should additionally be considered, in order of importance:

1. Ways of measuring the same construct: select validated rather than subjective/self-report exposure measure
2. Analysis: select exposure pertaining to adjusted estimate (i.e., adjusted for pre-specified critical confounding domains) or estimate which can be meta-analysed
3. Timepoint: select exposure pertaining to datapoint assessing the longest/last occasion measured unless not relevant

*Multiple exposure groups from a single datapoint:*

Approach taken to overcome a unit of analysis error for a datapoint that could contribute multiple, correlated comparisons, as per Cochrane guidance.^24^

1. Combine exposure groups to create a single pair-wise comparison comparable to other included datapoints in the meta-analysis
2. Select one pair of exposure groups and exclude the others, ensuring the groups selected are comparable with other included datapoints in the meta-analysis

Where a second datapoint originating from the same study, with the same exposure, is identified for inclusion in SWiM, the same groups used in the meta-analysis (via selection of one pair of exposure groups/combing exposure groups to create a single pair-wise comparison) should be used to enhance comparability.

*Different exposure periods (e.g., past week, current):*

Where possible, ensure exposure periods of datapoints to be included in meta-analysis align. Due to heterogeneity of exposures reported across studies, it is anticipated this may not be possible and each meta-analysis will include datapoints with varying exposure periods.

**Outcome**

If an adjusted datapoint cannot be converted to a common effect, and summary data or an unadjusted datapoint can be converted to a common effect, use this within meta-analysis. If neither the adjusted/unadjusted datapoint or summary data reported by the study can be converted to a common effect (e.g., change scores, outcome trajectory) report using SWiM.

*Multiple outcome measures for the same outcome:*

Where studies report multiple measures of the same behaviour (e.g., weekly alcohol consumption, frequency of binge drinking) the most common outcome measure should be selected (i.e., the outcome for which most studies contribute a result). The below criteria should additionally be considered, in order of importance:

1. Ways of measuring the same construct: select validated rather than subjective/self-report outcome measure
2. Analysis: select outcome measure pertaining to adjusted estimate (i.e., adjusted for pre-specified critical confounding domains) or estimate which can be meta-analysed
3. Timepoint: select outcome pertaining to datapoint assessing the longest/last occasion measured unless not relevant

*Different outcome periods (e.g., past week, past month use):*

Where possible, ensure that outcome periods of datapoints to be included in meta-analysis align. Due to heterogeneity of outcomes reported across studies, it is anticipated this may not be possible and each meta-analysis will include datapoints with varying outcome periods.

*Varying time points of follow up for an outcome:*

- Use the longest time point/last occasion measured unless not relevant (e.g., sexual intercourse measured at Time 1 (study sample aged 15) should be selected instead of sexual intercourse measured at Time 2 (study sample aged 16) where age of consent is 16 years in study setting)
- Where multiple time points are reported, these should be extracted separately, and sensitivity analysis conducted to explore any differences by study design (cross-sectional vs longitudinal)

**Sex**

- If sex-stratified datapoints reported, use both
- If whole sample estimate (male and female combined) alongside datapoint for a single sex reported, use the whole sample estimate
- If only one datapoint is reported pertaining to a single sex, use this

**Selection of datapoints for SWiM (vote counting based on effect direction)**

- Where effect estimates are incompletely reported or where study characteristics such as study design, exposures or outcomes are too diverse to provide a meaningful summary effect estimate, report datapoint using SWiM
- Where exposed and unexposed groups reported in a datapoint do not align with the meta-analysis exposed and unexposed group report using SWiM
- Datapoints reporting trajectory of outcome/change in outcome/change scores should not be used in meta-analyses and should be synthesised using SWiM
- Effect direction synthesis should be performed by exposure (time spent on social media, frequency of social media use, exposure to health-risk behaviour content and other social media activities), and by outcome domain
- Where multiple outcome measures are reported for the same exposure and same outcome, the direction of effects reported across outcome measures should be synthesised using the algorithm proposed by Cochrane,^25,26^ based on the proportion of effects which are in a consistent direction. Note it is possible for one study assessing one exposure and one outcome measure to demonstrate an unclear/conflicting/inconsistent effect^26^
- Where age subsets, study populations from different countries, and repeat cross sectional samples are reported in the same study, these should be entered as separate studies for purposes of SWiM, to maximise use of the available data
- SWiM should be conducted at the study level, and all effect direction plots presented, should report the study risk of bias grade

**Exposure**

*Same outcome assessed by multiple exposures:*

The most used exposure selected (i.e., the exposure for which most studies contribute a result). The below criteria will additionally be considered, in order of importance:

1. Ways of measuring the same construct: select validated rather than subjective/self-report exposure measure
2. Analysis: select exposure pertaining to adjusted estimate (i.e., adjusted for pre-specified critical confounding domains). If all datapoints fail to adjust for pre-specified critical confounders, consider adjustment for other justifiable confounders
3. Time point: select exposure pertaining to datapoint assessing the longest/last occasion measured unless not relevant

*Multiple exposure groups from a single datapoint:*

- Where a datapoint is reported in the meta-analysis and a pairwise comparison is selected/exposure groups are combined, when reporting this datapoint in SWiM ensure the same comparison is used to enhance comparability
- If a datapoint is to be reported in SWiM, and similar datapoints (with the same multiple exposure groups) originating from the same study have not been reported in meta-analyses or SWiM, then select a specific pairwise comparison/combine exposure groups, ensuring efforts are made to maximise use of all data, and the comparison aligns with other datapoints reported in SWiM synthesis

**Outcome**

*Multiple outcome measures reported for the same outcome:*

- Where there are multiple outcome measures investigating the same outcome, aggregate these using the effect direction algorithm: report direction of effect where ≥70% of outcomes report similar direction. If <70% of outcomes report consistent direction of effect, then report inconsistent findings^26^
- The largest sample size across all aggregated datapoints will be reported in the effect direction plot

*Multiple time points presented for an outcome from the same study:*

- Where there is a cross-sectional and cohort datapoint originating from same study investigating the exact same exposure and outcome, the direction of effect should be aggregated as above. The study design should be reported as a cohort study, and the associated cohort study risk of bias grade reported within the effect direction plot. The sample size reported should reflect that of the sample used in the cohort datapoint
- If datapoints, are reported for different follow up periods (e.g., time 1- time 2 and time 1- time 3) aggregate the direction of effect for all datapoints as above and report the associated risk of bias grade and sample size for the longest follow up period

**Sex**

If sex-stratified datapoints are reported, aggregate these when reporting effect direction and discard the whole sample estimate.

### Appendix-8. Data extraction form exemplar- study and datapoint level variables

##### Table A. Data extraction form: study level variables

| Field | Brief description | Guidance | Permissible entries |
| --- | --- | --- | --- |
| date | Date of data extraction by lead author | Not for completion by second-checker |  |
| source | Where did we find the study? | If this is a relevant publication screened in Covidence | Cov |
|  |  | If this is a relevant publication found via refence list of systematic review | SR |
|  |  | If this is a relevant publication identified via manual searching of reference lists of included studies | M |
|  |  | If this is a relevant publication identified via expert correspondence | E |
| study_ID | Internal reference number | Source = Covidence | #3343 use the number allocated within Covidence |
|  |  | Source = Systematic Review | #SR |
|  |  | Source = Reference list of included studies | #M |
|  |  | Source = Expert correspondence | #E |
| second_checker | Name of person doing second checking | To be completed by second-checker |  |
| second_checker_date | Date of second checking | To be completed by second-checker |  |
| first_author | Surname of first author |  |  |
| year_pub | Year of publication |  |  |
| published | Where was the study published | If study published in journal | Name of the journal |
| title | Title of the study |  |  |
| study_aim | Aim of study | Brief free-text description of the studies aim |  |
| author_contact | Study corresponding author contact details | Insert email of corresponding author |  |
| publication_cat | What type of publication is this? |  | Journal |
|  |  |  | Preprint |
| length | Was the study cross sectional or longitudinal? | No repeated measures | Cross-sectional |
|  |  | At least two waves of data collection on the same individuals | Longitudinal |
| country | Country of study | Where was the study carried out? (including location and social context) | e.g., Toronto Canada (5 Southern Toronto High Schools) |
| setting | Was the setting classified as a high or middle- or low-income country at the time of the study? | See World Bank Classification: https://datahelpdesk.worldbank.org/knowledgebase/articles/906519. If a study is looking at a range of countries which are a mixture of high- and low-income countries, select all options that apply | High income |
|  |  |  | Middle income |
|  |  |  | Low income |
| study_years | When did the study run? | Years in which study ran, including any follow-up. Can be expressed either as a range or a list (if certain years were excluded) |  |

| Field | Brief description | Guidance | Permissible entries |
| --- | --- | --- | --- |
| study_design | What kind of study? |  | Cross-sectional |
|  |  |  | Cohort |
|  |  |  | Randomised trial of intervention |
|  |  |  | Non-randomised trial of intervention |
|  |  |  | Cross-sectional analysis of cohort |
|  |  |  | Cross-sectional analysis of intervention |
|  |  |  | Natural experiment |
|  |  |  | Panel study |
|  |  |  | Repeat cross-sectional |
|  |  |  | Systematic review (primary data not available) |
| data_source_cat | Was primary or secondary data used in this study? |  | Primary |
|  |  |  | Secondary |
| data_source | If secondary data used, what is the name of the data source? | Insert name of data source |  |
| recruitment_  strategy | Recruitment setting/strategy | Free-text description of how participants were reached and recruited into study (e.g., sampling frame, sampling technique, location) |  |
| data_collection | Study data collection method | Free-text description of how and where data was collected from participants (e.g., online survey in home setting, paper survey conducted in classroom, telephone survey) |  |
| response_rate_% | Response rate as percentage | Those who completed baseline/those invited to participate |  |
| inc_criteria | Record study inclusion criteria | Description covering initial inclusion criteria for participation and analytical sample (if we are interested in a subset of the whole sample) |  |
| total_  participants | Total number of study participants in whole sample and analytical sample (if available) | If not reported for analytical sample extract for whole sample |  |
| average_age | Average age of whole sample and analytical sample (if available) | If not reported for analytical sample extract for whole sample | Report mean if available |
| age_range1 | Age range (indicator of spread) for whole sample and analytical sample (if available) | If not reported for analytical sample extract for whole sample | Report standard deviation if available |
| age_range2 | Sample age range covered in whole sample and analytical sample (if available) | If not reported for analytical sample extract for whole sample | Report range if available |
| percent_male | What percentage of the whole sample and analytical sample (if available) were male? | If not reported for analytical sample extract for whole sample |  |
| ethnicity | What is the ethnicity of study participants in the whole sample and analytical sample (if available) | If not reported for analytical sample extract for whole sample |  |
| sep | What is the socioeconomic position of the whole sample and analytical sample (if available) | Record the scale/measure used and the distribution amongst study participants in the analytical sample. If not reported for analytical sample extract for whole sample |  |
| conflicts_of_  interest | Any possible conflicts of interest? |  | Yes |
|  |  |  | No |
|  |  |  | Not reported (NR) |

| Field | Brief description | Guidance | Permissible entries |
| --- | --- | --- | --- |
| funding_source | Was the study funded by an organisation? | If yes | Record the name of funding bodies |
|  |  | If no | No |
|  |  | If not reported | Not reported (NR) |
| ethical_approval | Was ethnical approval obtained for the study? | If yes | Yes |
|  |  | If no | No |
|  |  | If not reported | Not reported (NR) |
|  |  | If not required (e.g., secondary data) | Not required |
| study_notes | Anything else worth recording in relation to study information |  |  |

##### Table B. Data extraction form: datapoint level variables

| Field | Brief description | Guidance | Permissible entries |
| --- | --- | --- | --- |
| dpID | Internal reference number for datapoint |  |  |
| dpnum | Datapoint number |  |  |
| fu_length | What was the length of follow up for this specific datapoint? | If cross-sectional study | Not applicable (NA) |
|  |  | For longitudinal studies, what was the length of follow-up for the specific datapoint? | Number of months/years |
| sg_gender/sex | For subgroup analysis, what is the gender/sex of the analytical sample this datapoint relates to? | If sex reported, select either Sex Male, Sex Female, Sex both. If gender reported, select either Gender Male, Gender Female, Gender Both | Gender Male |
|  |  |  | Gender Female |
|  |  |  | Gender Both |
|  |  |  | Sex Male |
|  |  |  | Sex Female |
|  |  |  | Sex Both |
| sg_age | For subgroup analysis, what is the average age of the analytical sample this datapoint relates to? | If not available for analytical sample, report for whole sample |  |
| sg_sep | For subgroup analysis, what is the socioeconomic position (SEP) of the analytical sample this datapoint refers to? | If not available for analytical sample, report for whole sample |  |
|  |  | If only includes those with low baseline SEP | Low SEP |
|  |  | If only includes those on high baseline SEP | High SEP |
|  |  | If includes a mix of low and high baseline SEP | Mixed- general population |
| sg_setting | For subgroup analysis what is the World Bank grouping of the country this datapoint refers to at the time of the study? | If not available for analytical sample, report for whole sample | High income |
|  |  |  | Middle income |
|  |  |  | Low income |
| exp_def | How was the exposure defined within the study? | Authors' description of social media use as per methods | e.g., frequency of social media networking site use, daily time spent using social media (hours/day) |
| exp_duration | When did data collection for the exposure occur? | Record when data collection for exposure occurred |  |
| time_period_exposure | What time period was the exposure measuring? | Record the time period for which the exposure measures | e.g., ever, current |

| Field | Brief description | Guidance | Permissible entries |
| --- | --- | --- | --- |
| sg_exp_cat | What social media category is under study for the datapoint? | Record which social media category is examined for specific data point. A number of social media platforms will fall under several social media types, for example, Twitter is a social networking site and a microblogging site.  If study authors state the specific type of social media record this, if they do not, make an assessment and record the social media type you think best represents the social media platform(s) under study, considering the initial premise of the platform.  Where it is impossible to determine which category is under study, select "Social Media" from the drop-down list | Social networking sites (e.g., Facebook, Whats App, Snapchat, Myspace, Whisper, Instant messaging) |
|  |  |  | Microblogging sites (e.g., Twitter, Tumblr) |
|  |  |  | Blogs and forums (e.g., Wordpress, Live journal, discussion boards, pin boards) |
|  |  |  | Media-sharing sites (e.g., YouTube, Pinterest, Instagram) |
|  |  |  | Geo-location- based sites (e.g., Foursquare, Tinder, Grindr) |
|  |  |  | Book marking sites (e.g., Delicious, Twitter, StumbleUpon) |
|  |  |  | Social news sites (e.g., Reddit, Digg) |
|  |  |  | Collaborative authoring sites (e.g., Wikipedia, Google Docs) |
|  |  |  | Web conferencing (e.g., Skype, zoom) |
|  |  |  | Scheduling and meeting (e.g., Microsoft outlook, Doodle, Google Calendar) |
|  |  |  | Online gambling |
|  |  |  | Online gaming |
|  |  |  | Online gaming |
|  |  |  | Social media |
| exp_platform | What social media platform is under study for the datapoint? | Record the specific social media platforms under study (e.g., Facebook, Reddit) for the specific datapoint or record the examples provided in relation to the datapoint under investigation | e.g., Facebook specified e.g., Examples: Facebook, Twitter |
| sg_exp_content | What type of social media content is understudy for the datapoint? | If user-generated content (e.g., content produced by the user, friends, others in the social media network) | User-generated |
|  |  | If marketer-generated content (e.g., advertisements & influencer content) | Marketer-generated |
|  |  | If content is both user-generated and marketer-generated content | User and Marketer-generated |
|  |  | If social media content is not specifically under investigation (e.g., time spent on social media/frequency of social media use) and we cannot distinguish what type of content the participant is exposed to | Not applicable (NA) |
| exp_ascertain | How was social media use measured for the datapoint? | Free-text description of measurement tool/instrument (e.g., specific scale, survey question, objective measures of social media usage tracked by mobile phones/electronic devices) For a scale, provide the name of the scale, upper and lower limits, and whether a high or low score is favourable and state definitions of any thresholds if appropriate  For survey questions, state the name of the survey, question, if it is self-report (or if a proxy has been used state this), question response options, whether a high or low score is favourable, and definitions of any thresholds/categories created if appropriate Record if objective/validated/self-report measure |  |

| Field | Brief description | Guidance | Permissible entries |
| --- | --- | --- | --- |
| exp_measure_  type | What type of measurement is the exposure? |  | Binary |
|  |  |  | Continuous |
|  |  |  | Categorical |
|  |  |  | Ordinal |
| exp_mean | Mean of exposure measure | Mean and/or proportion (n/%) of analytical sample or whole sample if not available  If exposure is ordinal/categorical, record number of those exposed in each exposure group |  |
| exp_SD | Standard deviation of exposure measure |  |  |
| outcome_  domain | What outcome does the datapoint report on? | Note 'multiple risk behaviours' should only be used where the analysis has specifically looked at 'multiple risk behaviours' as a single outcome (2 or more of the individual risk behaviours under investigation) | Alcohol use |
|  |  |  | Tobacco use |
|  |  |  | Drug use |
|  |  |  | Use of ENDS |
|  |  |  | Unhealthy dietary behaviour |
|  |  |  | Inadequate physical activity |
|  |  |  | Antisocial behaviour |
|  |  |  | Gambling |
|  |  |  | Sexual risk behaviour |
|  |  |  | Multiple risk behaviours |
| outcome_def | How was the outcome defined within the study? | Authors' description of outcome as per methods | e.g., frequency of drinking alcohol |
| outcome_  duration | When did data collection occur for the outcome? | Record when data collection occurred for outcome | e.g., 2004 (wave 2) |
| time_period_  outcome | What time period was the outcome measuring? | Record the time period for which the outcome measures | e.g., ever, current |
| outcome_  acertain | How was the outcome measured for the datapoint? | Free-text description of measurement tool/instrument For a scale, provide the name of the scale, upper and lower limits, and whether a high or low score is favourable and definitions of any thresholds if appropriate For survey questions, state the name of the survey, question, if it is self-report (or if a proxy has been used state this), question response options, whether a high or low score is favourable, and definitions of any thresholds/categories created if appropriate Record if validated tool/medical records/self-report/independent blind assessment | e.g., AUDIT-C. Response categories... |
| outcome_  measure_type | What type of measurement is the outcome? |  | Binary |
|  |  |  | Categorical |
|  |  |  | Continuous |
|  |  |  | Ordinal |

| Field | Brief description | Guidance | Permissible entries |
| --- | --- | --- | --- |
| outcome_mean | Mean of outcome measure | Mean and/or proportion (n/%) of analytical sample or whole sample if not available If exposure is ordinal/categorical, present number of those with outcome in each exposure group |  |
| outcome_SD | Standard deviation of outcome measure |  |  |
| analytical_  sample | Number of participants used in the analytical sample for the datapoint  Present for whole sample if possible |  |  |
| dp_measure | What effect measure is reported for the datapoint? | Where possible record adjusted measures for data extraction purposes if unavailable record unadjusted estimates Where both adjusted and unadjusted measures are presented, record adjusted estimates in data extraction form and state unadjusted estimates are available  If outcome measure is not listed, input as free text | Correlation coefficient (Pearson’s) |
|  |  |  | Correlation coefficient (Spearman’s) |
|  |  |  | Correlation coefficient (Point-biserial) |
|  |  |  | Correlation coefficient (Phi) |
|  |  |  | Standardised path coefficient (adjusted) |
|  |  |  | Standardised path coefficient (unadjusted) |
|  |  |  | Unstandardised path coefficient (adjusted) |
|  |  |  | Unstandardised path coefficient (unadjusted) |
|  |  |  | Standardised regression coefficient (adjusted) |
|  |  |  | Standardised regression coefficient (unadjusted) |
|  |  |  | Unstandardised regression coefficient (adjusted) |
|  |  |  | Unstandardised regression coefficient (unadjusted) |
|  |  |  | Standardised linear regression coefficient (adjusted) |
|  |  |  | Standardised linear regression coefficient (unadjusted) |
|  |  |  | Unstandardised linear regression coefficient (adjusted) |
|  |  |  | Unstandardised linear regression coefficient (unadjusted) |
|  |  |  | Odds ratio (adjusted |
|  |  |  | Odds ratio (unadjusted) |
|  |  |  | Risk Ratio (adjusted |
|  |  |  | Risk Ratio (unadjusted) |
|  |  |  | Mean |
|  |  |  | Median |
|  |  |  | Chi square |
|  |  |  | F-statistic |
|  |  |  | T-statistic |
|  |  |  | Raw summary data extracted |
| dp_analysis_  type | What type of analysis was conducted? | Insert brief statement on analysis method |  |
| analysis_desc | Describe the analysis used for investigation of the datapoint as per methods | Free-text description of analysis method used |  |

| Field | Brief description | Guidance | Permissible entries |
| --- | --- | --- | --- |
| incomplete_  outcome_ data | Was there any missing data (e.g., unit & item missingness)? How was this managed?  Were sampling/non-response weights used? | Describe the completeness of outcome data for each data point, including attrition (e.g., loss to follow up, withdrawn, non-response) and exclusions from the analysis.  Record if missing data handled appropriately or if weights (e.g., non-response and selection) were employed |  |
| dp_adjustment | If applicable, what confounders were adjusted for? | If adjusted estimates are presented, record all confounders controlled for |  |
| mediators_effect modifiers | Were mediators/effect modifiers investigated? | State if mediator/moderator investigated Record the name of the mediator/moderator | e.g., mediator: self-esteem |
| exp_group | What is the exposed group? |  |  |
| dp_point_est | The datapoint estimate of interest |  |  |
| n_numerator | People in numerator (with outcome) for group of interest | If available |  |
| n_denominator | People in denominator for group of interest | If available |  |
| other_denom | Other denominator | Use this to record the analytical sample or total number of individuals exposed for continuous exposures |  |
| lower_ci | Lower 95% confidence interval (CI) of main point estimate (if applicable) |  |  |
| upper_ci | Upper 95% CI of main point estimate (if applicable) |  |  |
| sd_se | Standard deviation/standard error (SE) of point estimate (if applicable) | If SE provided, note and mark (e.g., SE=) |  |
| t_z_stat | T or Z statistic (if applicable) |  |  |
| p_value | p value of main point estimate |  |  |
| sig_5% | Is the point estimate of interest significant at 5% level? | Record if significance level has been set at a level other than 5% | Yes |
|  |  |  | No, significance level set at (XXX) |
| comp_group | Description of comparator group | If exposure measure is continuous or point estimate is a correlation, state ‘Baseline' here to demonstrate the comparator group is those people with a different level of exposure than the exposed group | e.g., low social media use (<2 hours per day). |
| datapoint_notes | Location of data point extracted & study author reporting | Report the location of datapoint, numerators and denominators extracted within each study Record any issues regarding study author reporting |  |
| comp_point_est | Point estimate for comparator group |  |  |
| comp_  numerator | People in numerator (with outcome) for comparator group estimate | If available |  |
| comp_  denominator | People in denominator for comparator group estimate | If available |  |
| other_denom | Other denominator |  |  |
| comp_lower_ci | Lower 95% CI of comparator estimate (if applicable) |  |  |
| comp_upper_ci | Upper 95% CI of comparator estimate (if applicable) |  |  |

| Field | Brief description | Guidance | Permissible entries |
| --- | --- | --- | --- |
| comp_sd_se | Standard deviation/standard error of comparator estimate (if applicable) | If SE provided, note and mark (e.g., SE=) |  |
| comp_t_z_stat | T or Z statistic if applicable of comparator estimate (if applicable) |  |  |
| comp_pvalue | p value of comparator estimate |  |  |
| comp_sig5% | Is p value significant at 5% level? | Record if significance level has been set at a level other than 5% | Yes |
|  |  |  | No, significance level set at (XXX) |
| comp2… | Fields for second comparator group - replicate those for the first |  |  |
| other_pot_  relevant_ datapoints | Were other potentially relevant estimates reported for the exposure-outcome combination (datapoint) investigated? | If other relevant estimates recorded e.g., correlations/standardised estimates, record the estimates and state their location in paper  If unadjusted estimates provided state this |  |
| correspondence_  required | Do you need to contact the study authors for any reason (e.g., accessing original data, requesting additional info, verifying study details)? | Add information on required author correspondence |  |
| akp_notes |  | Lead author to insert any notes to aid interpretation/support data extracted |  |
| secondchecker_notes |  | Second checkers to insert any notes to aid interpretation/support data extracted |  |

### Appendix-9. Newcastle-Ottawa Scale (NOS) risk of bias assessment

For cross-sectional and cohort studies, an adapted version of the Newcastle-Ottawa Scale (NOS) for assessing risk of bias (RoB) in non-randomised studies was used.^27^ This appendix presents the adapted NOS used when assessing RoB of non-randomised studies and the algorithms used when assessing domain level and overall RoB of included datapoints. To ensure a standardised process to NOS RoB assessment, a detailed guidance document prepared by AKP was circulated to the review team. Those studies reporting baseline data from an interventional study were appraised as per cross-sectional studies. RoB assessment was conducted at the datapoint/outcome level and an overall RoB grade was assigned to each study through consideration of the most commonly reported RoB grade across included datapoints from a study, prioritising the overall grade assigned to datapoints which were investigated via the primary analysis conducted in a study.

#### Adapted Newcastle Ottawa Scale (NOS): cross-sectional studies

Used when assessing cross-sectional studies, cross-sectional analysis of cohort studies, cross-sectional analysis of intervention studies and repeat cross-sectional studies

***Domain A- Selection***

Selection- Representativeness of original sample

a) Good representativeness of the target population (e.g., all subjects, random sampling)
b) Selection process does not ensure representativeness, but it is clearly described (e.g., non- probability sampling)
c) Selected group of users, with the potential for selection bias
d) No description

Selection- Non-respondents

a) Comparability between respondents and non-respondents’ characteristics established and/or response rate ≥75% of original sample and (if applicable) those with and without missing data established and adjusted for
b) Comparability between respondents and non-respondents is unsatisfactory or response rate <75% or (if applicable) missing data addressed inappropriately
c) No description or unclear

##### Figure A. Algorithm to assess the Selection domain of the adapted Newcastle Ottawa Scale for cross-sectional studies

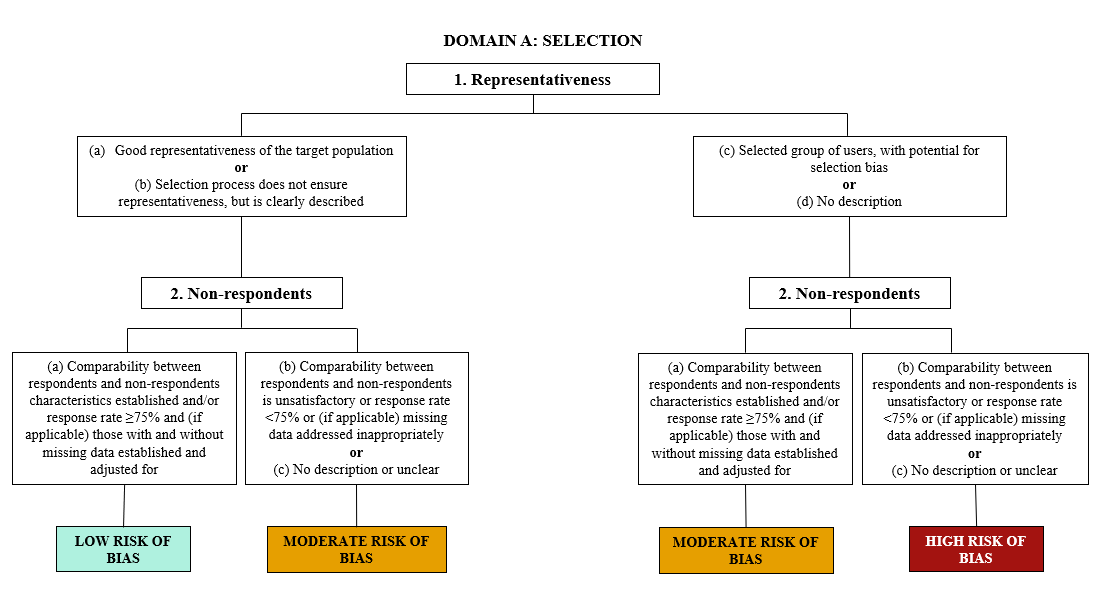

***Domain B- Exposure***

Exposure- Ascertainment of exposure

a) Objectively recorded social media usage data, independent of user reports
b) Validated measurement tool
c) Non-validated measurement tool, but the tool is available or described
d) No description or unclear

##### Figure B. Algorithm to assess the Exposure domain of the adapted Newcastle Ottawa Scale for cross-sectional studies

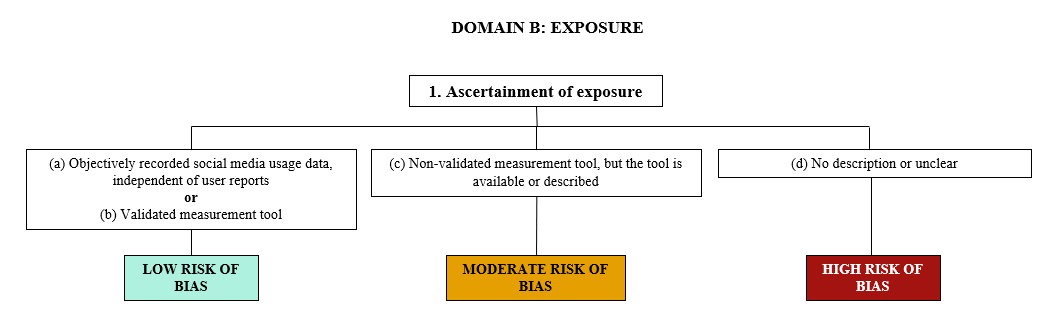

***Domain C- Comparability***

Comparability- Based on analysis of interest. Confounding factors are controlled.

(a) The study controls for key confounders age, sex/gender, and socioeconomic position (e.g., parental educational attainment, employment, income, area-level deprivation)
(b) The study controls for key confounders age, sex/gender, and socioeconomic position (e.g., parental educational attainment, employment, income, area-level deprivation) **and** the study controls for an alternative set of justifiable confounders (e.g., sensation seeking, peer influence or proxy measures for age, sex/gender, or socioeconomic position)
(c) The study controls for an alternative set of justifiable confounders (e.g., sensation seeking, peer influence or proxy measures for age, sex/gender, or socioeconomic position)
(d) No adjustment for potential confounders, no description or unclear

##### Figure C. Algorithm to assess the Comparability domain of the adapted Newcastle Ottawa Scale for cross-sectional studies

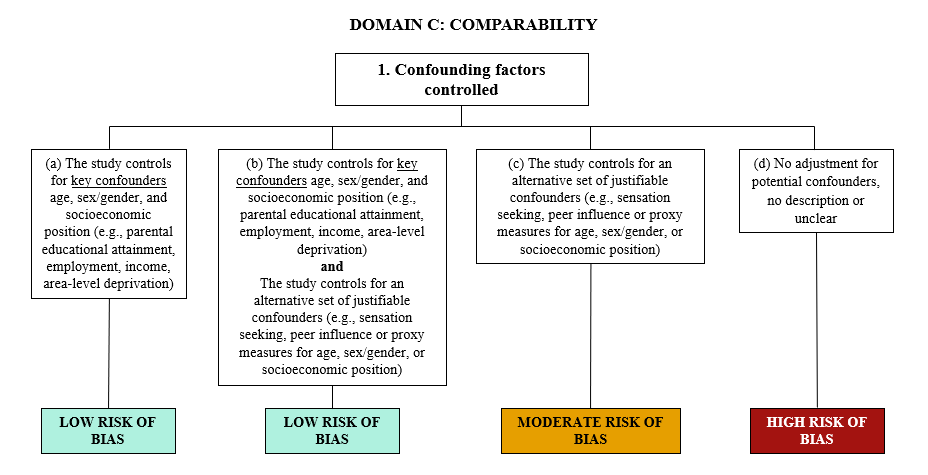

***Domain D- Outcome***

Outcome- Assessment of outcome

(a) Independent clinical assessment or validated measurement tool
(b) Medical/administrative records
(c) Self-report
(d) No description, or other inadequate

##### Figure D. Algorithm to assess the Outcome domain of the adapted Newcastle Ottawa Scale for cross-sectional studies

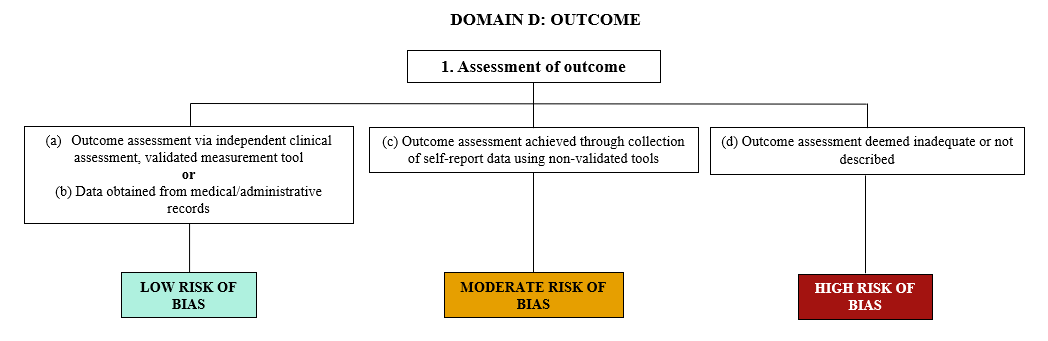

#### Adapted Newcastle Ottawa Scale (NOS): cohort studies

Used when assessing cohort and panel studies

***Domain A- Selection***

Selection- Representativeness of the original sample

a) Good representativeness of the target population (e.g., all subjects, random sampling)
b) Selection process does not ensure representativeness, but it is clearly described (e.g., non- probability sampling)
c) Selected group of users, with the potential for selection bias
d) No description

Selection- Selection of the comparator group

a) Drawn from the same community as the exposed cohort
b) Drawn from a different source
c) No description of the derivation of the non-exposed cohort

##### Figure E. Algorithm to assess the Selection domain of the adapted Newcastle Ottawa Scale for cohort studies

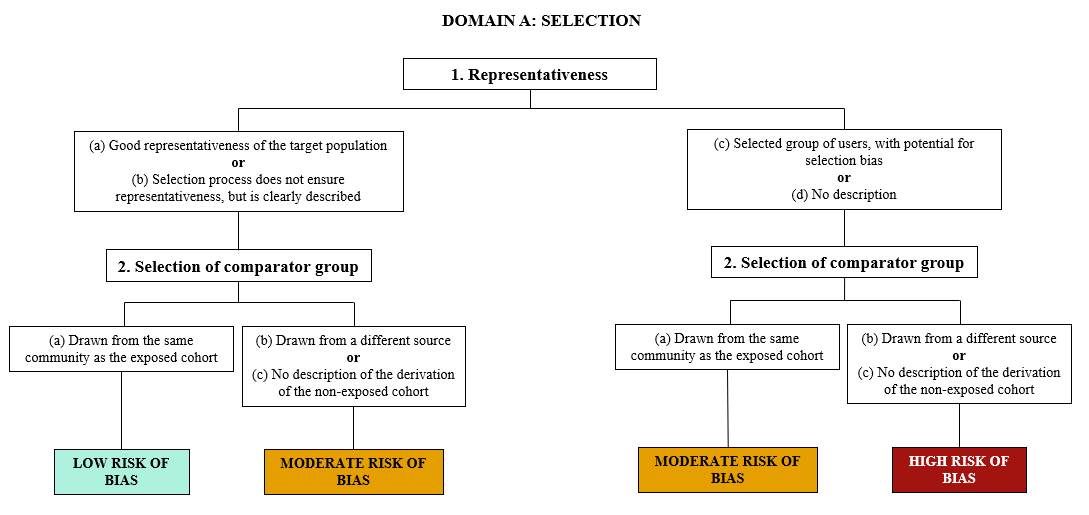

***Domain B- Exposure***

Exposure- Ascertainment of exposure

a) Objectively recorded social media usage data, independent of user reports
b) Validated measurement tool
c) Non-validated measurement tool, but the tool is available or described
d) No description or unclear

##### Figure F. Algorithm to assess the Exposure domain of the adapted Newcastle Ottawa Scale for cohort studies

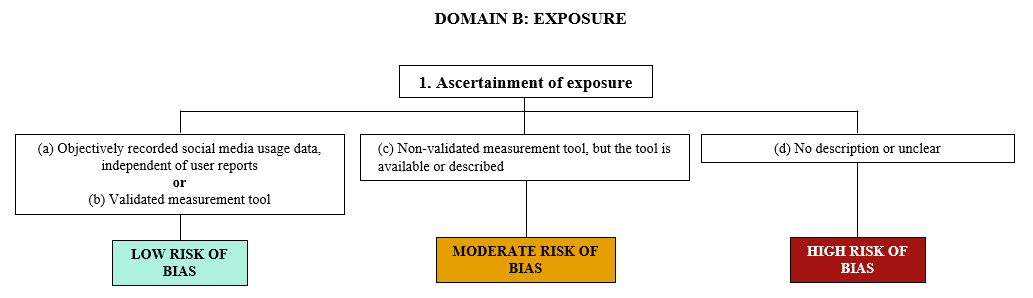

***Domain C- Comparability***

Comparability- Based on analysis of interest. Confounding factors are controlled.

a) The study controls for key confounders age, sex/gender, and socioeconomic position (e.g., parental educational attainment, employment, income, area-level deprivation)
b) The study controls for key confounders age, sex/gender, and socioeconomic position (e.g., parental educational attainment, employment, income, area-level deprivation) **and** the study controls for an alternative set of justifiable confounders (e.g., sensation seeking, peer influence or proxy measures for age, sex/gender, or socioeconomic position)
c) The study controls for an alternative set of justifiable confounders (e.g., sensation seeking, peer influence or proxy measures for age, sex/gender, or socioeconomic position)
d) No adjustment for potential confounders, no description or unclear

Comparability- Accounts for baseline measure of outcome

a) Yes
b) No

##### Figure G. Algorithm to assess the Comparability domain of the adapted Newcastle Ottawa Scale for cohort studies

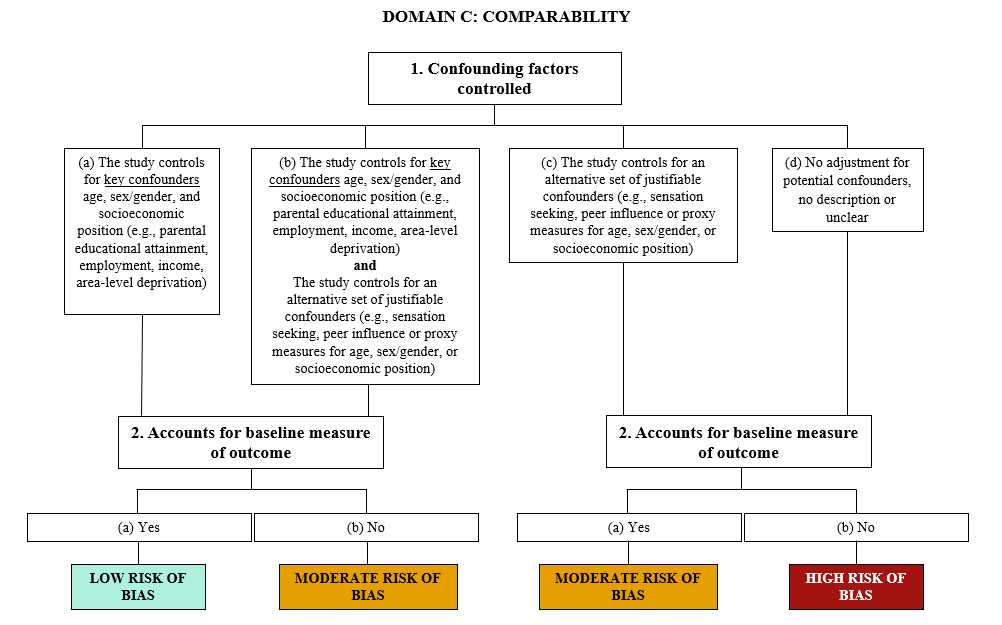

***Domain D- Outcome***

Outcome- Assessment of outcome

(a) Independent clinical assessment or validated measurement tool
(b) Medical/administrative records
(c) Self-report
(d) No description, or other inadequate

Outcome- Adequacy of follow up

a) Complete follow up (on all relevant variables)- all subjects accounted for
b) Subjects lost to follow up or due to missing data <25%, unlikely to introduce bias, or accounted for using weights, imputation etc.
c) Loss to follow up substantial (≥25%) and/or likely to introduce bias
d) Not described or unclear

##### Figure H. Algorithm to assess the Outcome domain of the adapted Newcastle Ottawa Scale for cohort studies

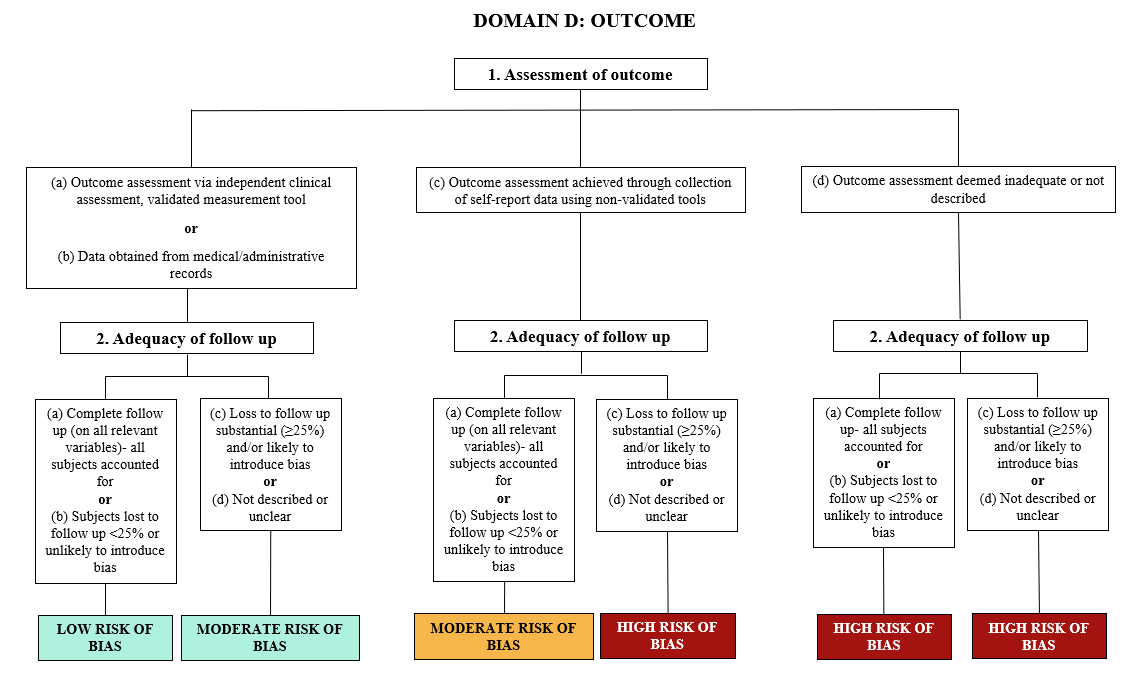

#### Assessing domain level and overall risk of bias (RoB) for included datapoints

For both cross-sectional studies and cohort studies, the algorithms presented above were used to grade each domain using the response options selected for each signalling question. Each domain was allocated either a low, moderate, or high RoB grade as illustrated in Table A.

##### Table A. Available domain level risk of bias (RoB) grades

| Domain | Risk of bias judgement | | |
| --- | --- | --- | --- |
| Selection | Low risk of bias | Moderate risk of bias | High risk of bias |
| Exposure | Low risk of bias | Moderate risk of bias | High risk of bias |
| Comparability | Low risk of bias | Moderate risk of bias | High risk of bias |
| Outcome | Low risk of bias | Moderate risk of bias | High risk of bias |

Once all domains were graded, Table B was used to reach an overall RoB judgement for each datapoint using the grades applied for each domain.

##### Table B. Algorithm to classify overall risk of bias (RoB) grade

| Overall risk of bias judgement | Criteria |
| --- | --- |
| Low risk of bias | Study is not judged to be at high risk of bias for any domain **and** is judged to be at low risk of bias for either the Exposure or Comparability domain |
| Moderate risk of bias | Study does not meet criteria for either High risk of bias or Low risk of bias |
| High risk of bias | Study is judged to be at high risk of bias in at least one domain |

### Appendix-10. Process for data transformations for meta-analysis

The majority of data transformations were conducted according to guidance within the Cochrane Handbook,^28,29^ and using the Campbell Collaboration online effect size calculator.^30^

For binary/dichotomous exposure measurements, continuous and dichotomous outcome data were combined. We opted to report these as odds ratios as most outcomes were originally reported on binary scales, therefore fewer assumptions were made when completing the conversions. For continuous exposure measurements- which are infrequently reported in systematic reviews, meaning best practice recommendations are not available within the Cochrane Handbook^28^ – we opted *not* to combine continuous and binary outcome data. This decision was taken (after discussion with systematic review and statistical experts) because the common statistical approaches to combine these data are grounded on the ability to make direct comparisons between a distinct exposed and unexposed group, and we did not feel it was interpretable or appropriate to utilise these for continuous exposures. Instead, continuous exposure measurements and continuous outcome data were pooled separately as standardised beta coefficients or standardised mean differences (SMDs), whilst continuous exposure and binary/dichotomous outcome data were pooled separately as odds ratios.

Beta coefficients were converted to standardised beta coefficients. Pearson correlation coefficients were converted to standardised mean differences adopting the method outlined by Mathur and VanderWeele.^2^ Here, where studies failed to report the standard deviation (SD) of the exposure, efforts were made to contact study authors to obtain the information required for transformation. Where this was not possible, as recommended a substitute estimate was extracted from a second comparable included study (n = 1) or subsample of the study used to estimate *r* and the N term within the formula replaced with the size of the second sample used to estimate the exposure SD.^2^

Where multiple exposure groups were presented for a datapoint, in overcoming any potential unit of analysis errors for a datapoint which could contribute multiple, correlated comparisons, efforts were made to combine groups to create a single pairwise comparison, as recommended by Cochrane,^24^ and following discussion with statistical experts. Where this was not possible one pair of exposure groups was selected, and the others excluded. Regardless of the option selected, groups were combined/selected ensuring comparability with other included datapoints within the meta-analysis. For subgroup analysis, in some instances the analysis includes datapoints not used within primary meta-analysis due to potential double counting of study participants. For example, where multiple datapoints were reported within the same study assessing different SM categories/platforms/content, they were included within separate subgroups.

As per the Cochrane Handbook, where studies reported a p value of <0.05 in the absence of the exact value and this was required to determine the standard error of an estimate, to facilitate inclusion in the meta-analysis, the p-value was assumed to be 0.05.^28^ Where data were insufficiently reported for standardisation or transformation, study investigators were contacted by email (n= 6 responses received).

### Appendix-11. Characteristics of included studies

##### Table A. Characteristics of included studies (n=126 studies; 338 datapoints)

| Author and year | Study design | Study period | Country | Equity | Participants | Mean age [range] | Risk of bias | N°. of dp | MA? | Exposure | Exposure measure | Outcome measure | N |
| --- | --- | --- | --- | --- | --- | --- | --- | --- | --- | --- | --- | --- | --- |
| Anastario 2020^31^ | Cross-sectional | NR | USA | High income country | Youth attending 5 schools located on or near a tribal reservation in Montana | 15.7  [14-18] | Mod | 2 | Yes | Freq. of SM use | Freq. of using Twitter to talk or learn about sex or any topic related to sex | No use of a condom at last sexual encounter | 146 |
|  |  |  |  |  |  |  |  |  |  | Freq. of SM use | Freq. of using Facebook to talk or learn about sex or any topic related to sex | No use of a condom at last sexual encounter | 146 |
| Baker 2016^32^ | Cross-sectional | 2009 | USA | High income country | Grade 6-12 urban school district students’ part of a federally funded project on school related initiatives | NR | High | 3 | Yes | Freq. of SM use | Freq. of SNS use | Soft drug use (smoking, marijuana,  alcohol) in the past month | 3195 |
|  |  |  |  |  |  |  |  |  |  | Freq. of SM use | Freq. of SNS use | Hard drug use (lifetime and past year) | 3195 |
|  |  |  |  |  |  |  |  |  |  | Freq. of SM use | Freq. of SNS use | Weapon carrying in the past month | 3195 |
| Baldwin 2018^33^ | Cross-sectional | 2014 | Australia | High income country with mixed SEP | Adolescents residing in New South Wales | NR [10-16] | Low | 7 | Yes | Exposure to health-risk behaviour content | Watched food/beverage brand YouTube videos | Freq. of unhealthy food consumption | 417 |
|  |  |  |  |  |  |  |  |  |  | Exposure to health-risk behaviour content | Seen favourite food advertised on SM | Freq. of unhealthy food consumption | 417 |
|  |  |  |  |  |  |  |  |  |  | Exposure to health-risk behaviour content | Liked a food/beverage brand on Facebook | Freq. of unhealthy food consumption | 204 |
|  |  |  |  |  |  |  |  |  |  | Exposure to health-risk behaviour content | Seen favourite food advertised on SM | Freq. of unhealthy drink consumption | 417 |
|  |  |  |  |  |  |  |  |  |  | Exposure to health-risk behaviour content | Seen favourite food advertised on SM | Freq. of unhealthy food & drink consumption | 407 |
|  |  |  |  |  |  |  |  |  |  | Freq. of SM use | Freq. of logging in, or checking Facebook account | Freq. of unhealthy food consumption | 204 |
|  |  |  |  |  |  |  |  |  |  | Freq. of SM use | Freq. of logging in, or checking Facebook account | Freq. of unhealthy drink consumption | 204 |

| Author and year | Study design | Study period | Country | Equity | Participants | Mean age [range] | Risk of bias | N°. of dp | MA? | Exposure | Exposure measure | Outcome measure | N |
| --- | --- | --- | --- | --- | --- | --- | --- | --- | --- | --- | --- | --- | --- |
| Ball 2020^34^ | Repeat cross-sectional | 2016 & 2018 | New Zealand | High income country with mixed SEP | Year 10 students’ part of the Youth Insights Survey | NR  [14-15] | Low | 2 | Yes | Freq. of SM use | Freq. of using SM (status updates, uploading photos or videos) in the past week | Current smoking (defined as smoking at least monthly) | 5127 |
|  |  |  |  |  |  |  |  |  |  | Freq. of SM use | Freq. of online gambling in the past week | Current smoking (defined as smoking at least monthly) | 5127 |
| Baru 2020^35^ | Cross-sectional | 2019 | Ethiopia | Low-middle income country  with mixed SEP | Sexually active unmarried young female internal migrants residing in Barayu Town | 18.9  [15-24] | High | 1 | Yes | Freq. of SM use | Freq. of SM use | Risk sexual behaviour (incl. multiple sexual partners; sex without condoms or inconsistent condom use; initiation of sex before the age of 18 years; sexual intercourse under the influence of substances) | 195 |
| Baumgartner 2012^36^ | Cohort | 2018 | Netherlands | High income country | Adolescents | 14.5 [12-18] | Mod | 1 | Yes | Freq. of SM use | Freq. of online communication | Online sexual risk behaviours (incl. searching for someone on the internet to have sex with; sending a photo or video in which they were partly naked to someone they knew only online) | 1762 |
| Bayraktar 2007^37^ | Cross-sectional | NR | Cyprus | HIC | Elementary and high school students residing in North Cyprus | 14.4 [NR] | High | 1 | No | Exposure to health-risk behaviour content | Online gaming: fighting games | Anti-social aggression | 686 |
| Beebe 2004^38^ | Cross-sectional | 2001 | USA | High income country | Grade 9 school students’ part of the Minnesota Student Survey | 14.7 [13-17] | High | 12 | Yes | Freq. of SM use | Presence of internet chat room use | Tobacco use in the past year | 40,376 |
|  |  |  |  |  |  |  |  |  |  | Freq. of SM use | Presence of internet chat room use | Alcohol/  drug use in the past year | 40,376 |
|  |  |  |  |  |  |  |  |  |  | Freq. of SM use | Presence of internet chat room use | Sexual intercourse ever | 40,376 |
|  |  |  |  |  |  |  |  |  |  | Freq. of SM use | Presence of internet chat room use | Physical assault in the past year | 40,376 |
|  |  |  |  |  |  |  |  |  |  | Freq. of SM use | Presence of internet chat room use | Vandalism in the past year | 40,376 |
|  |  |  |  |  |  |  |  |  |  | Freq. of SM use | Presence of internet chat room use | Truant in the past month | 40,376 |
| Boers 2020^39^ | Cohort | NR | Canada | High income country with low SEP | Grade 7 school students, part of the Co-Venture Preventure study | 12.7 [NR] | Low | 1 | Yes | Time spent on SM | Time spent on SM per day | Freq. of alcohol consumption | 3612 |

| Author and year | Study design | Study period | Country | Equity | Participants | Mean age [range] | Risk of bias | N°. of dp | MA? | Exposure | Exposure measure | Outcome measure | N |
| --- | --- | --- | --- | --- | --- | --- | --- | --- | --- | --- | --- | --- | --- |
| Boniel-Nissim 2022^40^ | Cross-sectional | 2017-2018 | 42 countries and regions across Europe, North America, and the Middle East | High income country with mixed SEP | School students, part of the Health Behaviour in School-aged Children Survey | 13.6  [11-15] | Low | 4 | Yes | Freq. of SM use | Freq. of online contact with others via SM *(via validated tool)* | Smoking (≥1 × in the last month) *(via validated tool)* | 173,577 |
|  |  |  |  |  |  |  |  |  |  | Freq. of SM use | Freq. of online contact with others via SM *(via validated tool)* | Alcohol consumption (≥3 × in the last month) *(via validated tool)* | 172,723 |
|  |  |  |  |  |  |  |  |  |  | Freq. of SM use | Freq. of online contact with others via SM *(via validated tool)* | Drunkenness (≥1 × in the last month) *(via validated tool)* | 171,320 |
|  |  |  |  |  |  |  |  |  |  | Freq. of SM use | Freq. of online contact with others via SM *(via validated tool)* | Cannabis use (≥1 × in the last month) *(via validated tool)* | 55,956 |
| Booker 2015^41^ | Cross-sectional | 2009 | UK | High income country with mixed SEP | Sample members of the youth panel of The UK Household Longitudinal Study | NR [10-15] | High | 1 | No | Time spent on SM | Time spent chatting on social websites on a normal school day | Sports participation | 4899 |
| Brunborg 2019^42^ | Cohort | 2014-2015 | Norway | High income country | Grade 8-10 and 1-2^nd^ year high school students’ part of the pilot Monitoring Young Lifestyles Project | 15.2 [13-17] | Low | 4 | Yes | Time spent on SM | Change in hrs of SM use per day (Δ = T2 – T1) | Change in episodic heavy drinking freq. | 763 |
|  |  |  |  |  |  |  |  |  |  | Time spent on SM | Change in hrs of SM use per day (Δ = T2 – T1) | Change in conduct problems *(via SDQ)* | 763 |
|  |  |  |  |  |  |  |  |  |  | Time spent on SM | Average number of hrs spent on SM per day in the past year | Episodic heavy drinking freq. in the past year | 763 |
|  |  |  |  |  |  |  |  |  |  | Time spent on SM | Average number of hrs spent on SM per day in the past year | Conduct problems in the past year *(via SDQ)* | 763 |
| Brunborg 2022^43^ | Cohort | 2017-2020 | Norway | High income country with mixed SEP | Middle school adolescents’ part of the MyLife Study | 14.3  [12.8-16.8] | Mod | 1 | No | Time spent on SM | Average number of hrs spent on SM per day | Change in alcohol use *(via AUDIT-C)* | 3096 |

| Author and year | Study design | Study period | Country | Equity | Participants | Mean age [range] | Risk of bias | N°. of dp | MA? | Exposure | Exposure measure | Outcome measure | N |
| --- | --- | --- | --- | --- | --- | --- | --- | --- | --- | --- | --- | --- | --- |
| Camenga 2018^44^ | Cohort | 2013-2014 | USA | High income country with mixed SEP | High and middle school students’ part of a longitudinal school-based cohort study | 14.1 [NR] | High | 4 | Yes | Exposure to health-risk behaviour content | Exposure to e-cigarette  advertisements on Facebook | Ever e-cigarette use | 1742 |
|  |  |  |  |  |  |  |  |  |  | Exposure to health-risk behaviour content | Exposure to e-cigarette advertisements on Twitter | Ever e-cigarette use | 1742 |
|  |  |  |  |  |  |  |  |  |  | Exposure to health-risk behaviour content | Exposure to e-cigarette advertisements on YouTube | Ever e-cigarette use | 1742 |
|  |  |  |  |  |  |  |  |  |  | Exposure to health-risk behaviour content | Exposure to e-cigarette advertisements on Pinterest/Google + | Ever e-cigarette use | 1742 |
| Canale 2016^45^ | Cross-sectional | 2013 | Italy | High income country with mixed SEP | High school students’ part of the European School Survey Project on Alcohol and Other Drugs Italia | 17.2 [15-19] | Low | 3 | Yes | Freq. of SM use | Freq. of using internet for leisure activities (e.g., online chatting) | Problem gambling *(via SOGS-RA)* | 14,478 |
|  |  |  |  |  |  |  |  |  |  | Freq. of SM use | Freq. of online gambling in past year | Problem gambling *(via SOGS-RA)* | 14,478 |
|  |  |  |  |  |  |  |  |  |  | Freq. of SM use | Freq. of using internet for leisure activities (e.g., online chatting) | At-risk gambling *(via SOGS-RA)* | 14,478 |
| Casaló 2022^46^ | Cross-sectional | 2016-2017 | Spain | High income country with mixed SEP | Secondary education students, part of the National Survey on Drug Use Among High School Students in Spain | NR  [14-18] | Low | 4 | Yes | Time spent on SM | Time spent on SNS per day | Sports frequency 1-3 days per year | 35,369 |
|  |  |  |  |  |  |  |  |  |  | Time spent on SM | Time spent on SNS per day | Sports frequency 1-3 days per month | 35,369 |
|  |  |  |  |  |  |  |  |  |  | Time spent on SM | Time spent on SNS per day | Sports frequency 1-4 days per week | 35,369 |
|  |  |  |  |  |  |  |  |  |  | Time spent on SM | Time spent on SNS per day | Sports frequency 5-7 days per week | 35,369 |
| Cavazos-Rehg 2014^47^ | Cross-sectional | 2011 | USA | High income country with mixed SEP | Grade 6-12 school students’ part of the National Youth Tobacco Survey | NR [11-17] | High | 1 | Yes | Exposure to health-risk behaviour content | Exposure to tobacco ads/promotions via Facebook/  Myspace in the past month | Used any form of tobacco in the past month | 15,673 |

| Author and year | Study design | Study period | Country | Equity | Participants | Mean age [range] | Risk of bias | N°. of dp | MA? | Exposure | Exposure measure | Outcome measure | N |
| --- | --- | --- | --- | --- | --- | --- | --- | --- | --- | --- | --- | --- | --- |
| Chang 2016^48^ | Cohort | 2010-2011 | Taiwan | High income country with mixed SEP | Grade 10 students from 26 high schools in Taipei City and New Taipei City | NR | High | 2 | Yes | Freq. of SM use | Freq. of online game use during past week | Incidence of unwanted online sexual solicitation perpetration in the past year | 1981 |
|  |  |  |  |  |  |  |  |  |  | Freq. of SM use | Freq. of chat room use during past week | Incidence of unwanted online sexual solicitation perpetration in the past year | 1981 |
| Chapin 2018^49^ | Cross-sectional | 2016-2017 | USA | High income country with mixed SEP | Middle and high school students’ part of the Empowering Latino Youth Project evaluation | 14.1 [12-18] | High | 2 | No | Other SM activities | Number of SM platforms used | Experience with electronic violence (perpetration) in the past month | 1167 |
|  |  |  |  |  |  |  |  |  |  | Other SM activities | Number of SM platforms used | Experience with face-to-face violence (perpetration) in the past month | 1167 |
| Chau 2022^50^ | Cross-sectional | 2010 | France | High income country with mixed SEP | Students attending 3 middle schools (2 public and 1 private) in the Lorraine region of North-eastern France | 13.5  [10-18] | Mod | 5 | Yes | Time spent on SM | Time spent on discussion forums and chatting online during a weekday | Alcohol use in the past month | 1559 |
|  |  |  |  |  |  |  |  |  |  | Time spent on SM | Time spent on discussion forums and chatting online during a weekday | Tobacco use in the past month | 1559 |
|  |  |  |  |  |  |  |  |  |  | Time spent on SM | Time spent on discussion forums and chatting online during a weekday | Cannabis use in the past month | 1559 |
|  |  |  |  |  |  |  |  |  |  | Time spent on SM | Time spent on discussion forums and chatting online during a weekday | Perpetrated violence *(via validated tool)* | 1559 |
|  |  |  |  |  |  |  |  |  |  | Time spent on SM | Time spent on discussion forums and chatting online during a weekday | Illicit drug use in the past month | 1559 |
| Chen 2019^51^ | Cross-sectional | 2018 | Belgium | High income country  with mixed SEP | School students’ part of the New Media Study | 16.4 [15-18] | High | 2 | Yes | Time spent on SM | Time spent on SM on a regular weekday and weekend day | Experience with risky (anti-social) selfie behaviour | 686 |
|  |  |  |  |  |  |  |  |  |  | Exposure to health-risk behaviour content | Exposure to risky selfie descriptive norms | Experience with risky selfie (anti-social) behaviour | 686 |
| Coates 2019^52^ | RCT | 2017 | UK | High income country | School students without food allergies | 10.1 [9-11] | Some concerns (RoB-2) | 1 | Yes | Exposure to health-risk behaviour content | Exposure to unhealthy mock Instagram influencer marketing *(objectively recorded)* | Caloric intake (kcal): consumption of unhealthy snacks *(objectively recorded)* | 117 |

| Author and year | Study design | Study period | Country | Equity | Participants | Mean age [range] | Risk of bias | N°. of dp | MA? | Exposure | Exposure measure | Outcome measure | N |
| --- | --- | --- | --- | --- | --- | --- | --- | --- | --- | --- | --- | --- | --- |
| Coyne 2013^53^ | Cross-sectional | 2010 | USA | High income country  with mixed SEP | Families with an adolescent aged 11-14 who used SNS who were part of a larger study on family life | 14.4 [11-14] | High | 1 | Yes | Time spent on SM | Time spent on SNS on a typical day | Delinquency *(via validated tool)* | 491 |
| Coyne 2018^54^ | Cohort | 2009-2014 | USA | High income country  with mixed SEP | Families with an adolescent aged 11-14 who used SNS who were part of the Flourishing Families Project | 13.5 [10-14] | High | 2 | No | Time spent on SM | Time spent on SNS on a typical day | Physical aggression | 457 |
|  |  |  |  |  |  |  |  |  |  | Time spent on SM | Time spent on SNS on a typical day | Relational aggression | 457 |
| Critchlow 2019^55^ | Cross-sectional | 2017 | UK | High income country with mixed SEP | Sample members of the UK Youth Alcohol Policy Survey | 15.2 [11-19] | Low | 3 | Yes | Freq. of SM use | SM apps used at least weekly in the past week | Higher-risk alcohol consumption in current drinkers *(via AUDIT-C)* | 1591 |
|  |  |  |  |  |  |  |  |  |  | Exposure to health-risk behaviour content | Participation with alcohol marketing on SM in the past month | Higher-risk alcohol consumption in current drinkers *(via AUDIT-C)* | 1591 |
|  |  |  |  |  |  |  |  |  |  | Exposure to health-risk behaviour content | Participation with user-created alcohol promotion on SM in the past month | Higher-risk alcohol consumption in current drinkers *(via AUDIT-C)* | 1591 |
| da Costa 2021^56^ | Cross-sectional | 2019 | Brazil | Low-middle income country with mixed SEP | High school students enrolled in high school courses integrated to professional courses, part of the Longitudinal Study of the Lifestyle of Adolescents | 16.3  [14-18] | Low | 3 | Yes | Time spent on SM | Time spent on SM on a typical weekday and weekend day | Sedentary behaviour in the last 4 days *(via Actigraph accelerometer)* | 718 |
|  |  |  |  |  |  |  |  |  |  | Time spent on SM | Time spent on SM on a typical weekday and weekend day | Light intensity physical activity in the last 4 days *(via Actigraph accelerometer)* | 718 |
|  |  |  |  |  |  |  |  |  |  | Time spent on SM | Time spent on SM on a typical weekday and weekend day | Moderate to vigorous physical activity in the last 4 days *(via Actigraph accelerometer)* | 718 |

| Author and year | Study design | Study period | Country | Equity | Participants | Mean age [range] | Risk of bias | N°. of dp | MA? | Exposure | Exposure measure | Outcome measure | N |
| --- | --- | --- | --- | --- | --- | --- | --- | --- | --- | --- | --- | --- | --- |
| Dai 2022^57^ | Cross-sectional | 2019 | China | High income country with mixed SEP | Junior, senior high and vocational high school students in Shanghai | 13.7  [13-18] | Low | 2 | Yes | Exposure to health-risk behaviour content | Exposure to e-cigarette advertisements on SM *(via validated tool)* | Ever e-cigarette use *(via validated tool)* | 708,765 |
|  |  |  |  |  |  |  |  |  |  | Exposure to health-risk behaviour content | Exposure to e-cigarette advertisements on SM *(via validated tool)* | E-cigarette use in the past month *(via validated tool)* | 708,765 |
| Davis 2019^58^ | Cohort | 2010-2016 | USA | HIC Mixed SEP | Grade 6-7 middle school students’ part of the CHOICE USA alcohol and drug use prevention program | 13.2 [12-15] | Mod | 1 | No | Exposure to health-risk behaviour content | Substance related media exposure via SM in the past 3 months | Freq. of alcohol use in the past month | 4840 |
| Dawson 2019^59^ | Cross-sectional | 2016-2017 | USA | High income country with mixed SEP | Students with previous diagnosis of ADHD part of the BEST Project | 14.5 [NR] | High | 3 | Yes | Freq. of SM use | Presence of SNS use | Ever sent a sext | 58 |
|  |  |  |  |  |  |  |  |  |  | Freq. of SM use | Number of participant posts on Facebook (posted by participant) over 2-month period *(objectively recorded)* | Ever sent a sext | 34 |
|  |  |  |  |  |  |  |  |  |  | Exposure to health-risk behaviour content | % of participant posts sharing inappropriate content on Facebook over 2-month period *(objectively recorded)* | Ever sent a sext | 34 |
| de Bruijn 2016^60^ | Cross-sectional | 2012 | Germany, Italy, Netherlands, and Poland | High income country | Urban and rural school students | 14.1 [NR] | Mod | 2 | Yes | Exposure to health-risk behaviour content | Ever use of an alcohol branded SM page | Onset of drinking | 9032 |
|  |  |  |  |  |  |  |  |  |  | Exposure to health-risk behaviour content | Ever used alcohol branded SM page | Binge drinking in the past month | 9032 |
| De Jans 2021^61^ | RCT | 2020 | Belgium | High income country with mixed SEP | School students from 3 primary schools | 10.0  [8-12] | Low | 1 | No | Exposure to health-risk behaviour content | Exposure to snack with low nutritional value (mini donut) on Instagram *(objectively recorded)* | Consumption of snack high in nutritional value (strawberries)  *(objectively recorded)* | 190 |
| De Looze 2019^62^ | Cross-sectional | 2002-2014 | European and North American Countries | High income country with mixed SEP | Sample members of the Health Behaviour in School-aged Children Survey | 13.5 [13.1-13.8] | Mod | 3 | Yes | Freq. of SM use | Freq. of electronic media communication with friends | Weekly alcohol use | 191,727 |
|  |  |  |  |  |  |  |  |  |  | Freq. of SM use | Freq. of electronic media communication with friends | Weekly smoking | 191,727 |
|  |  |  |  |  |  |  |  |  |  | Freq. of SM use | Freq. of electronic media communication with friends | Lifetime cannabis use | 56,159 |

| Author and year | Study design | Study period | Country | Equity | Participants | Mean age [range] | Risk of bias | N°. of dp | MA? | Exposure | Exposure measure | Outcome measure | N |
| --- | --- | --- | --- | --- | --- | --- | --- | --- | --- | --- | --- | --- | --- |
| Doornwaard 2014^63^ | Cross-sectional | 2012 | Netherlands | High income country | Elementary and high school students’ part of the Studies on Trajectories of Adolescent Relationships and Sexuality | 15.0 [11-18] | High | 1 | No | Exposure to health-risk behaviour content | Exposure to displays of sexual references on Facebook over 3-month period *(objectively recorded)* | Experience with sexual behaviours *(via validated tool)* | 104 |
| Doornwaard 2015^64^ | Cross-sectional | 2011 | Netherlands | High income country | Grade 7-10 students’ part of the Studies on Trajectories of Adolescent Relationships and Sexuality | 14.0 [11-17] | Mod | 4 | Yes | Time spent on SM | Time spent on SNS (most frequently used platform) per day | Ever experience with sexual behaviours | 1132 |
|  |  |  |  |  |  |  |  |  |  | Time spent on SM | Time spent on SNS (most frequently used platform) per day | Freq. of sex related online behaviours | 1132 |
| Elton-Marshall 2016^65^ | Cross-sectional | 2012-2013 | Canada | High income country with mixed SEP | Grade 9-12 school students’ part of the Canadian Youth Smoking Survey who responded to the Youth Gambling Survey supplement | 16.5 [13-19] | High | 2 | Yes | Freq. of SM use | Freq. of playing free simulated gambling games on Facebook in the past 3 months | Freq. of gambling for money (not via SM) | 9830 |
|  |  |  |  |  |  |  |  |  |  | Freq. of SM use | Freq. of online gambling participation in the past 3 months | Problem gambling severity *(via CAGI/GPSS)* | 3682 |
| Erreygers 2017^66^ | Cross-sectional | 2015 | Belgium | High income country with mixed SEP | Grade 7 school students’ part of a larger study | 13.6 [NR] | High | 1 | No | Freq. of SM use | Freq. of online gaming in the past 6 months | Performing online anti-social behaviours in the past month | 1720 |
| Floros 2013^67^ | Cross-sectional | 2010 | Greece | High income country with mixed SEP | High school students’ part of the Hippocrates Study | 15.1 [12-19] | High | 2 | No | Freq. of SM use | Freq. of using SNS in the past year | Internet gambling (not via SM) freq. in the past year | 2017 |
|  |  |  |  |  |  |  |  |  |  | Freq. of SM use | Freq. of using SNS in the past year | Pathological gambling past year *(via DSM-IV-MR-J)* | 2017 |
| Folkvord 2020^68^ | RCT | 2018 | Netherlands | High income country | Grade 1-2 secondary school students | 14.1  [13-16] | Low (RoB-2) | 1 | No | Exposure to health-risk behaviour content | Exposure to manipulated popular influencer Instagram post showing energy dense foods *(objectively recorded)* | Vegetable intake *(objectively recorded)* | 88 |

| Author and year | Study design | Study period | Country | Equity | Participants | Mean age [range] | Risk of bias | N°. of dp | MA? | Exposure | Exposure measure | Outcome measure | N |
| --- | --- | --- | --- | --- | --- | --- | --- | --- | --- | --- | --- | --- | --- |
| Froyland 2020^69^ | Cross-sectional | 2015 & 2018 | Norway | High income country | All junior and senior high school students in Oslo part of the Young in Oslo Surveys | NR  [13-18] | Low | 8 | Yes | Time spent on SM | Time spent on SM per day | Physical fighting (with and without weapons) in the past 12 months | 47,655 |
|  |  |  |  |  |  |  |  |  |  | Time spent on SM | Time spent on SM per day | School truancy in the past 12 months | 47,655 |
|  |  |  |  |  |  |  |  |  |  | Time spent on SM | Time spent on SM per day | Alcohol intoxication in the past 12 months | 47,655 |
|  |  |  |  |  |  |  |  |  |  | Time spent on SM | Time spent on SM per day | Cannabis use in the past 12 months | 47,655 |
| Gascoyne 2021^70^ | Cross-sectional | 2018 | Australia | High income country with mixed SEP | Secondary school students’ part of the National Secondary Students’ Diet and Activity Survey | NR  [12-17] | Low | 2 | Yes | Exposure to health-risk behaviour content | Liked/shared posts related to a food or drink product or brand (e.g., soft drink, fast food) | High intake of unhealthy food | 8708 |
|  |  |  |  |  |  |  |  |  |  | Exposure to health-risk behaviour content | Liked/shared posts related to a food or drink product or brand (e.g., soft drink, fast food) | High intake of unhealthy drinks | 8708 |
| Gazendam 2021^71^ | Cross-sectional | 2018 | Canada | High income country with mixed SEP | Grade 9-10 students, part of the Canadian Health Behaviour in School-aged Children Survey | 15.4  [NR] | Low | 2 | No | Time spent on SM | Time spent on SM per day | Early sexual intercourse (15 years or younger) | 6123 |
| Geber 2021^72^ | Cohort | 2019-2020 | Switzerland | High income country | 1st year students at 4 secondary schools | 15.1  [13-17] | Mod | 1 | Yes | Exposure to health-risk behaviour content | Exposure to alcohol related content on Instagram and Snapchat | Drinking behaviour | 402 |
| Geusens 2017^73^ | Cross-sectional | 2015 | Belgium | High income country with mixed SEP | 5^th^, 6^th^ and 7^th^ year secondary school students’ part of the Flemish Alcohol and Media Survey Research Project | 17.2 [16-20] | Mod | 2 | Yes | Exposure to health-risk behaviour content | Perceived number of friends sharing alcohol references online | Self-reported drinking behaviour *(via AUDIT)* | 2935 |
| Geusens 2017^74^ | Cohort | 2015-2016 | Belgium | High income country | 5^th^, 6^th^ and 7^th^ year secondary school students’ part of the Flemish Alcohol and Media Survey Research Project | 17.0  [16-20] | High | 2 | No | Exposure to health-risk behaviour content | Freq. of sharing alcohol references on SNS | Binge drinking in the last 12 months | 998 |
| Geusens 2019^75^ | Cross-sectional | 2014 | Belgium | High income country with mixed SEP | 3^rd^ and 4^th^ year secondary school students’ part of a larger study | 14.9 [14-16] | Low | 2 | Yes | Exposure to health-risk behaviour content | Freq. of exposure to peer alcohol references on SNS | Alcohol consumption | 886 |
|  |  |  |  |  |  |  |  |  |  | Exposure to health-risk behaviour content | Freq. of sharing of alcohol references on SNS | Alcohol consumption | 886 |

| Author and year | Study design | Study period | Country | Equity | Participants | Mean age [range] | Risk of bias | N°. of dp | MA? | Exposure | Exposure measure | Outcome measure | N |
| --- | --- | --- | --- | --- | --- | --- | --- | --- | --- | --- | --- | --- | --- |
| Gomez 2019^76^ | Cross-sectional | 2018 | Spain | High income country | Secondary and baccalaureate students | 14.4 [12-17] | High | 1 | No | Other SM activities | Signed up to more than 5 SNS | Online gambling & betting (not via SM) | 3772 |
| Gordon 2011^77^ | Cross-sectional | 2006-2007 | UK | High income country with mixed SEP | 2^nd^ year high school students’ part of the Assessing the Cumulative Impact of Alcohol Marketing on Youth Drinking Study | 13.0 [12-14] | High | 2 | Yes | Exposure to health-risk behaviour content | Awareness of alcohol marketing on SNS | Drinking status | 912 |
|  |  |  |  |  |  |  |  |  |  | Exposure to health-risk behaviour content | Used SNS containing alcohol brands or logos | Drinking status | 912 |
| Gregg 2018^78^ | Cross-sectional | 2015 | USA | High income country | High school students from 1 suburban high school | 16.2 [NR] | Mod | 1 | No | Freq. of SM use | Freq. of electronic communication | Freq. of sending sexts *(via SBS)* | 314 |
| Gunnlaugsson 2020^79^ | Cross-sectional | 2017 | Guinea-Bissau | Low-middle income country with mixed SEP | Students from 16 secondary schools in Bissau | NR  [14-19+] | Low | 3 | Yes | Freq. of SM use | Freq. of SM use *(via validated tool)* | Participated in bullying behaviour in the past 12 months *(via validated tool)* | 1454 |
|  |  |  |  |  |  |  |  |  |  | Freq. of SM use | Freq. of SM use *(via validated tool)* | Lifetime experience of smoking cigarettes *(via validated tool)* | 1566 |
|  |  |  |  |  |  |  |  |  |  | Freq. of SM use | Freq. of SM use *(via validated tool)* | Lifetime experience of drinking alcohol *(via validated tool)* | 1559 |
| Hamilton 2020^80^ | Cross-sectional | 2020 | USA | High income country with high SEP | Adolescent girls residing in Pennsylvania part of larger longitudinal study | 15.06  [12-17] | High | 1 | No | Time spent on SM | Time spent on SNS per day | Physical activity | 93 |
| Hayer 2018^81^ | Cohort | 2015-2016 | Germany | High income country | Grade 6-10 school students in Northern Germany | 13.4 [11-19] | Mod | 2 | Yes | Freq. of SM use | Freq. of participation in any simulated gambling on social networks in the past year | Freq. of monetary gambling (not via SM) in the past year | 531 |
|  |  |  |  |  |  |  |  |  |  | Freq. of SM use | Freq. of participation in simulated gambling from home on social networks in the past year | Freq. of participation in monetary gambling (not via SM) in the past year | 531 |
| Holtz 2011^82^ | Cross-sectional | 2007 | Austria | High income country | Rural and urban school students | 12.7 [10-14] | Mod | 1 | No | Freq. of SM use | Freq. of communicational internet use (e.g., chatrooms, social platforms like Myspace) | Delinquent and aggressive behaviours in the past 6 months *(via YSR)* | 205 |

| Author and year | Study design | Study period | Country | Equity | Participants | Mean age [range] | Risk of bias | N°. of dp | MA? | Exposure | Exposure measure | Outcome measure | N |
| --- | --- | --- | --- | --- | --- | --- | --- | --- | --- | --- | --- | --- | --- |
| Hryhorczuk 2019^83^ | Cross-sectional | 2011 | Ukraine | Low-middle income country | Sample members of the Family and Children of Ukraine Birth Cohort Study | 16.2 [15.1-18.2] | Mod | 6 | Yes | Freq. of SM use | Freq. of SM use | Used alcohol in the past month | 912 |
|  |  |  |  |  |  |  |  |  |  | Freq. of SM use | Freq. of SM use | Used alcohol in the past year | 917 |
|  |  |  |  |  |  |  |  |  |  | Freq. of SM use | Freq. of SM use | Ever used alcohol | 967 |
| Hrywna 2020^84^ | Cross-sectional | 2018 | USA | High income country | Grade 9-12 school students’ part of the New Jersey Youth Tobacco Survey | NR | Mod | 2 | Yes | Exposure to health-risk behaviour content | Liked/followed a tobacco brand on SM in the past year | Current use of  e-cigarette or Juul (use on ≥1 day of the past month) | 4183 |
|  |  |  |  |  |  |  |  |  |  | Exposure to health-risk behaviour content | Liked/followed a tobacco brand on SM in the past year | Frequent use of  e-cigarette or Juul (use on ≥20 days of the past month) | 4183 |
| Huang 2012^85^ | Cross-sectional | 2007 | China | High income country with mixed SEP | Grade 10 academic and vocational school students’ part of the Trans-disciplinary Tobacco and Alcohol Use Research Centre Study | 15.8 [13-19] | Low | 1 | No | Freq. of SM use | Freq. of social internet activity (online gaming, chatting with real friends, chatting with online friends) in the past week | Cigarette smoking in the past month | 2931 |
| Huang 2014^86^ | Cohort | 2010-2011 | USA | High income country with low SEP | Grade 10 school students’ part of the Social Network Study | 15.1 [NR] | Low | 4 | Yes | Freq. of SM use | Freq. of Myspace use | Alcohol use in the past month | 1315 |
|  |  |  |  |  |  |  |  |  |  | Freq. of SM use | Freq. of Facebook use | Ever smoking | 1315 |
|  |  |  |  |  |  |  |  |  |  | Exposure to health-risk behaviour content | Number of friends who posted risky pictures partying/  drinking | Alcohol use in the past month | 1315 |
|  |  |  |  |  |  |  |  |  |  | Exposure to health-risk behaviour content | Number of friends who posted risky pictures partying/  drinking | Ever smoking | 1315 |

| Author and year | Study design | Study period | Country | Equity | Participants | Mean age [range] | Risk of bias | N°. of dp | MA? | Exposure | Exposure measure | Outcome measure | N |
| --- | --- | --- | --- | --- | --- | --- | --- | --- | --- | --- | --- | --- | --- |
| Jeong 2022^87^ | Cross-sectional | 2020 | South Korea | High income country | Adolescents part of the Consumer Behaviour Survey for Food conducted by the Korea Rural Economic Institute | 16.1  [NR] | High | 2 | No | Freq. of SM use | Freq. of SM use | Eats food considering calories and nutrients *(via validated tool)* | 622 |
|  |  |  |  |  |  |  |  |  |  | Freq. of SM use | Freq. of SM use | Eats carefully selected food for one’s own health *(via validated tool)* | 622 |
| Jiang 2018^88^ | Cross-sectional | NR | China | High income country | Young inpatients enrolled at one of the largest addiction clinics in China | 16.8 [13-19] | High | 1 | No | Freq. of SM use | Freq. of online gaming *(via clinical records)* | Involvement in risk behaviours (e.g., skipping school, smoking) *(via clinical records)* | 467 |
| Kaufman 2014^89^ | Cross-sectional | 2012 | South Africa | Low-middle income country with mixed SEP | Grade 9 school students, part of a 2-year cluster- randomised trial | NR [12-20] | Low | 8 | Yes | Freq. of SM use | Freq. of SM use | Hazardous alcohol use in the past year *(via AUDIT)* | 4485 |
|  |  |  |  |  |  |  |  |  |  | Freq. of SM use | Freq. of SM use | Reported multiple partners in the past year | 4485 |
|  |  |  |  |  |  |  |  |  |  | Other SM activities | Has a Facebook account | Hazardous alcohol use in past year *(via AUDIT)* | 4485 |
|  |  |  |  |  |  |  |  |  |  | Other SM activities | Has a Facebook account | Reported multiple partners in the past year | 4485 |
| Kaur 2020^90^ | Cross-sectional | 2018 | USA | High income country with mixed SEP | Grade 8,10 and 12 school students’ part of the Monitoring the Future Survey | 15.1 [NR] | Low | 6 | No | Time spent on SM | Time spent on SNS per day | Binge drinking in the past 2 weeks | 22,980 |
|  |  |  |  |  |  |  |  |  |  | Time spent on SM | Time spent on SNS per day | Drinking in the past month | 23,150 |
|  |  |  |  |  |  |  |  |  |  | Time spent on SM | Time spent on SNS per day | Cannabis use in the past month | 23,167 |
|  |  |  |  |  |  |  |  |  |  | Time spent on SM | Time spent on SNS per day | Flavour vaping in the past month in 8^th^ and 10^th^ grade students | 6967 |
|  |  |  |  |  |  |  |  |  |  | Time spent on SM | Time spent on SNS per day | Cannabis vaping in the past month in 8^th^ and 10^th^ grade students | 7003 |
|  |  |  |  |  |  |  |  |  |  | Time spent on SM | Time spent on SNS per day | Nicotine vaping in the past month in 8^th^ and 10^th^ grade students | 6980 |

| Author and year | Study design | Study period | Country | Equity | Participants | Mean age [range] | Risk of bias | N°. of dp | MA? | Exposure | Exposure measure | Outcome measure | N |
| --- | --- | --- | --- | --- | --- | --- | --- | --- | --- | --- | --- | --- | --- |
| Kelleghan 2020^91^ | Cohort | 2015-2017 | USA | High income country with mixed SEP | High School students’ part of the Happiness & Health Study | 16.5  [NR] | Mod | 6 | Yes | Freq. of SM use | Freq. of SM posting (posting photos, video or statuses and sharing others content) | Any cannabis use initiation (incl. reported use of combustible cannabis, blunts, and edible, vaporized, or synthetic cannabis) | 1841 |
|  |  |  |  |  |  |  |  |  |  | Freq. of SM use | Freq. of SM posting (posting photos, video or statuses and sharing others content) | Combustible cannabis use initiation | 1841 |
|  |  |  |  |  |  |  |  |  |  | Freq. of SM use | Freq. of SM posting (posting photos, video or statuses and sharing others content) | Other cannabis use initiation (incl. reported use of edible, vaporized, or synthetic cannabis) | 1841 |
|  |  |  |  |  |  |  |  |  |  | Freq. of SM use | Freq. of SM posting (posting photos, video or statuses and sharing others content) | Any tobacco use initiation (incl. reported use of a few puffs of a cigarette, a whole cigarette, e-cigarettes with tobacco, smokeless tobacco, big cigars, little cigars/ cigarillos, and hookah water pipe) | 1558 |
|  |  |  |  |  |  |  |  |  |  | Freq. of SM use | Freq. of SM posting (posting photos, video or statuses and sharing others content) | Combustible cigarette use initiation (incl. reported use of a few puffs of a cigarette or a whole cigarette) | 1558 |
|  |  |  |  |  |  |  |  |  |  | Freq. of SM use | Freq. of SM posting (posting photos, video or statuses and sharing others content) | E-cigarette use initiation | 1558 |
| King 2014^92^ | Cross-sectional | 2012 | Australia | High income country | Secondary school students in Metropolitan region of Adelaide | 14.9 [12-17] | High | 1 | Yes | Freq. of SM use | Ever use of simulated gambling via SNS applications (Facebook apps) | Freq. of problem gambling *(via DSM-IV-MR-J)* | 1214 |
| Ko 2009^93^ | Cross-sectional | 2004 | Taiwan | High income country with mixed SEP | Junior high and senior high/vocational school students’ part of the Project for Health of Adolescents | 14.6-14.9 [NR] | Mod | 3 | Yes | Freq. of SM use | Ever online gaming | Aggressive behaviours in the past year | 9405 |
|  |  |  |  |  |  |  |  |  |  | Freq. of SM use | Ever online chatting | Aggressive behaviours in the past year | 9405 |
|  |  |  |  |  |  |  |  |  |  | Freq. of SM use | Ever online gambling | Aggressive behaviours in the past year | 9405 |

| Author and year | Study design | Study period | Country | Equity | Participants | Mean age [range] | Risk of bias | N°. of dp | MA? | Exposure | Exposure measure | Outcome measure | N |
| --- | --- | --- | --- | --- | --- | --- | --- | --- | --- | --- | --- | --- | --- |
| Kontostoli 2022^94^ | Cross-sectional | 2015-2016 | UK | High income country with mixed SEP | Adolescents part of the Millennium Cohort Study | 14.2  [NR] | Low | 6 | No | Time Spent on SM | Time spent browsing and updating SNS on a weekday | Moderate-to-vigorous physical activity *(via accelerometer)* | 4546 |
|  |  |  |  |  |  |  |  |  |  | Time Spent on SM | Time spent browsing and updating SNS on a weekday | Overall physical activity *(via accelerometer)* | 4546 |
|  |  |  |  |  |  |  |  |  |  | Time Spent on SM | Time spent browsing and updating SNS on a weekday | Sedentary behaviour | 3551 |
| Koutamanis 2015^95^ | Cross-sectional | 2012 | Netherlands | High income country | Families with ≥2 adolescents aged 10-15 | 12.6 [12-15] | High | 1 | Yes | Freq. of SM use | Freq. of online social exploration on SNS | Freq. of risky online self-presentation | 758 |
| Kwon 2022^96^ | Cross-sectional | 2017 | South Korea | High income country with mixed SEP | Middle and high school students’ part of the Korea Youth Health Risk Behaviour Web Based Online Survey | 15.0  [12-18] | Low | 4 | No | Freq. of SM use | Freq. of SNS use in the past 30 days | Moderate to vigorous aerobic physical activity at least 5 days per week or vigorous physical activity at least 3 days per week in the past 7 days | 53,133 |
|  |  |  |  |  |  |  |  |  |  | Freq. of SM use | Freq. of SNS use in the past 30 days | Muscle-strengthening activity for at least 3 days per week in past 7 days | 53,133 |
| Landry 2013^97^ | Cross-sectional | 2011-2012 | USA | High income country | Grade 9-10 Latino high school students in Maryland | 15.7 [13-19] | High | 2 | Yes | Freq. of SM use | Freq. of logging into SM sites | No contraception use at last sex | 118 |
|  |  |  |  |  |  |  |  |  |  | Other SM activities | Has a Facebook account | No contraception use at last sex | 118 |
| Larm 2017^98^ | Cross-sectional | 2010 | Sweden | High income country | Grade 9 primary school students’ part of the Survey of Adolescent Life in Vastmanland | NR [15-16] | High | 2 | Yes | Time spent on SM | Online social network  chatting | Alcohol use *(via AUDIT-C)* | 2439 |
| Larm 2019^99^ | Repeat cross-sectional | 2008  2010 2012 | Sweden | High income country | Grade 9 students’ part of the Survey of Adolescent Life in Vastmanland | NR [15-16] | High | 2 | Yes | Time spent on SM | Time spent on SM/  chatting per day | Drinking in the past year *(via AUDIT-C)* | 2605 (RCS: 2008)  2045 (RCS: 2012) |
| Lee 2015^100^ | Cross-sectional | 2012 | South Korea | High income country | Grade 1-3 middle school and Grade 5-6 elementary school students residing in Incheon | NR [11-16] | High | 1 | No | Time spent on SNS per day | Time spent on SNS per day | Negative SNS behaviours (e.g., real money trading in SNS games, exposed to porn on SNS) | 500 |

| Author and year | Study design | Study period | Country | Equity | Participants | Mean age [range] | Risk of bias | N°. of dp | MA? | Exposure | Exposure measure | Outcome measure | N |
| --- | --- | --- | --- | --- | --- | --- | --- | --- | --- | --- | --- | --- | --- |
| Lee 2019^101^ | Cohort | 2013-2016 | USA | High income country with mixed SEP | Non-institutionalised adolescents’ part of the Population Assessment of Tobacco and Health Study | NR [12-17] | High | 2 | No | Freq. of SM use | Freq. of visiting social networking account | Initiation of ENDS in the past year | 8704 |
| Lee 2021^102^ | Cross-sectional | 2017-2018 | China | High income country | Students from 1 secondary school | 18.4  [NR] | High | 2 | Yes | Time Spent on SM | Daytime use of social networks *(objectively recorded)* | Number of steps in the past 7 days *(via accelerometer)* | 32 |
|  |  |  |  |  |  |  |  |  |  | Time Spent on SM | Daytime use of social networks *(objectively recorded)* | Moderate to vigorous physical activity in the past week *(via accelerometer)* | 32 |
| Lee 2021^103^ | Cross-sectional | 2019 | USA | High income country | Middle and high school students’ part of the Florida Youth Tobacco Survey | NR  [NR] | Mod | 2 | No | Freq. of SM use | Freq. of Facebook use | Experimental vaping (vaped but not in the past month) | 10,475 |
|  |  |  |  |  |  |  |  |  |  | Freq. of SM use | Freq. of Facebook use | Current vaping in the past month | 10,475 |
| Lin 2012^104^ | Cross-sectional | NR | New Zealand | High income country with mixed SEP | Student’s part of a larger study | NR [13-14] | High | 2 | Yes | Exposure to health-risk behaviour content | Awareness of alcohol marketing on SNS | Drinking status in the past year | 2538 |
|  |  |  |  |  |  |  |  |  |  | Exposure to health-risk behaviour content | Used SNS containing alcohol brands or logos | Drinking status in the past year | 2538 |
| Lipsky 2017^105^ | Cohort | 2010-2014 | USA | HIC Mixed SEP | Grade 10 school students’ part of the NEXT Generation Health Study | 16.5 [NR] | Low | 3 | No | Time spent on SM | Time spent on social networking per day | Healthy eating (conformance to US Dietary Guidelines 2010) *(via HEI, ASA24)* | 566 |
|  |  |  |  |  |  |  |  |  |  | Time spent on SM | Time spent on social networking per day | Intake of empty calories *(via HEI, ASA24)* | 566 |
|  |  |  |  |  |  |  |  |  |  | Time spent on SM | Time spent on social networking per day | Intake of whole plant foods *(via ASA24)* | 566 |
| Longobardi 2021^106^ | Cross-sectional | NR | Italy | High income country | Grade 7-13 school students’ part of a larger study on SM use | 15  [NR] | High | 1 | Yes | Time spent on SM | Time spent on SM per day | Sexting and online exhibitionism *(via validated tool)* | 229 |
| McClure 2020^107^ | Cross-sectional | 2015- 2016 | USA | HIC Mixed SEP | Adolescents recruited from general paediatric clinics in New England | 14.5 [12-17] | High | 1 | No | Freq. of SM use | Freq. of SM use | Ever drinking | 202 |

| Author and year | Study design | Study period | Country | Equity | Participants | Mean age [range] | Risk of bias | N°. of dp | MA? | Exposure | Exposure measure | Outcome measure | N |
| --- | --- | --- | --- | --- | --- | --- | --- | --- | --- | --- | --- | --- | --- |
| Merrill 2019^108^ | Cross-sectional | 2015 | USA | High income country with mixed SEP | Grade 9-12 school students’ part of the Youth Risk Behaviour Surveillance System Survey | NR [12-18+ older] | High | 1 | Yes | Time spent on SM | Time spent on SM per day | Risky sexual behaviours in sexually active participants | 5603 |
| Michael 2016^109^ | Cross-sectional | 2015^c^ | Nigeria | Low-middle income country | Adolescents residing in Bayelsa State Capital | 15.1 [10-19] | High | 1 | No | Time spent on SM | Average time spent on SM per day | No use of contraception during sexual intercourse | 262 |
| Moitra 2022^110^ | Cross-sectional | 2021 | India | Low-middle income country with mixed SEP | Grade 6-10 students from 6 private schools and 4 government aided schools in Mumbai | 13.2  [10-15] | Low | 2 | Yes | Time spent on SM | Time spent using SNS on a typical weekend and weekday | Healthy eating habits | 1298 |
|  |  |  |  |  |  |  |  |  |  | Time spent on SM | Time spent using SNS on a typical weekend and weekday | Physical activity level *(via PAQ-C/-A)* | 1298 |
| Mojica 2014^111^ | Cross-sectional | 2010 | USA | High income country | Female adolescents recruited via Girls Scouts of Southwest Texas as part of an intervention planning grant | NR [11-14] | High | 3 | No | Time spent on SM | Average time spent on SNS per week | 5+ days of physically activity in the past week | 110 |
|  |  |  |  |  |  |  |  |  |  | Time spent on SM | Average time spent on SNS per week | Daily physical education class in an average week | 110 |
|  |  |  |  |  |  |  |  |  |  | Time spent on SM | Average time spent on SNS per week | Played on 1+ sports team in the past year | 110 |
| Molla-Esparza 2021^112^ | Cross-sectional | 2015 | Spain | High income country | Adolescents from 2 secondary charter schools and 2 state schools in the south of Valencia | 13.7  [12-18] | Mod | 3 | Yes | Freq. of SM use | Freq. of using SM platforms | Sent a sext | 647 |
|  |  |  |  |  |  |  |  |  |  | Other SM activities | Number of SM platforms used | Sent a sext | 647 |
|  |  |  |  |  |  |  |  |  |  | Other SM activities | Number of SM platforms used | Forwarded a sext | 647 |
| Nesi 2017^113^ | Cohort | 2009-2013 | USA | High income country with mixed SEP | Grade 6-8 middle school students’ part of a larger study | 15.8 [NR] | Mod | 6 | Yes | Exposure to health-risk behaviour content | Ever exposed to friends' SNS alcohol content | Initiation of drinking | 658 |
|  |  |  |  |  |  |  |  |  |  | Exposure to health-risk behaviour content | Ever exposed to friends' SNS alcohol content | Initiation of becoming drunk | 658 |
|  |  |  |  |  |  |  |  |  |  | Exposure to health-risk behaviour content | Ever exposed to friends' SNS alcohol content | Initiation of heavy episodic drinking | 658 |
|  |  |  |  |  |  |  |  |  |  | Time spent on SM | Average time on Facebook per day | Initiation of drinking | 658 |
|  |  |  |  |  |  |  |  |  |  | Time spent on SM | Average time on Facebook per day | Initiation of becoming drunk | 658 |
|  |  |  |  |  |  |  |  |  |  | Time spent on SM | Average time on Facebook per day | Initiation of heavy episodic drinking | 658 |

| Author and year | Study design | Study period | Country | Equity | Participants | Mean age [range] | Risk of bias | N°. of dp | MA? | Exposure | Exposure measure | Outcome measure | N |
| --- | --- | --- | --- | --- | --- | --- | --- | --- | --- | --- | --- | --- | --- |
| Nesi 2019^114^ | Cohort | 2015-2016 | USA | High income country with low SEP | Grade 7-8 rural, lower-middle class school students’ part of a larger study | 16.0 [NR] | High | 10 | Yes | Freq. of SM use | Freq. of daily SM use | No. of sexual partners in the past year | 716 |
|  |  |  |  |  |  |  |  |  |  | Freq. of SM use | Posted selfies on Instagram over 1 month period *(objectively recorded)* | No. of sexual partners in the past year | 233 |
|  |  |  |  |  |  |  |  |  |  | Freq. of SM use | Posted selfies on Instagram over 1 month period *(objectively recorded)* | Substance use (alcohol use and past year marijuana and cigarette use) | 233 |
|  |  |  |  |  |  |  |  |  |  | Other SM activities | Online status-seeking strategy use | No. of sexual partners in the past year | 716 |
|  |  |  |  |  |  |  |  |  |  | Other SM activities | Online status-seeking strategy use | Substance use (alcohol and past year marijuana and cigarette use) | 716 |
| Ng Fat 2021^115^ | Cohort | 2011-2016 | UK | High income country with mixed SEP | Sample members of the UK Household Longitudinal Survey | 12.5^g^ 17.5^h^  [10-19] | Low | 8 | Yes | Time spent on SM | Time spent on SM on a normal weekday | Increase in drink freq. in the past 3 years (in those 10-15 years) | 856 |
|  |  |  |  |  |  |  |  |  |  | Time spent on SM | Time spent on SM on a normal weekday | Increase in drink freq. in the past 3 years (in those 16-19 years) | 511 |
|  |  |  |  |  |  |  |  |  |  | Time spent on SM | Time spent on SM on a normal weekday | Increase in binge drink freq. in the past 3 years (in those 16-19 years) | 1057 |
|  |  |  |  |  |  |  |  |  |  | Time spent on SM | Time spent on SM on a normal weekday | Drank alcohol in the past month (in those 10-15 years) | 4093 |
|  |  |  |  |  |  |  |  |  |  | Time spent on SM | Time spent on SM on a normal weekday | Drank alcohol 1-3 times a month in the past month (in those 16-19 years) | 2689 |
|  |  |  |  |  |  |  |  |  |  | Time spent on SM | Time spent on SM on a normal weekday | Drank alcohol at least weekly in the past month (in those 16-19 years) | 2689 |
|  |  |  |  |  |  |  |  |  |  | Time spent on SM | Time spent on SM on a normal weekday | Binge drinking 1-2 times a month (in those 16-19 years) | 2687 |
|  |  |  |  |  |  |  |  |  |  | Time spent on SM | Time spent on SM on a normal weekday | Binge drinking ≥3 times a month (in those 16-19 years) | 2687 |

| Author and year | Study design | Study period | Country | Equity | Participants | Mean age [range] | Risk of bias | N°. of dp | MA? | Exposure | Exposure measure | Outcome measure | N |
| --- | --- | --- | --- | --- | --- | --- | --- | --- | --- | --- | --- | --- | --- |
| Ngqangashe 2021^116^ | RCT | NR | Belgium | High income country with mixed SEP | Students in Flanders part of a larger research project on food media use | 13.9  [12-14] | Some concerns | 1 | No | Exposure to health-risk behaviour content | Watched YouTube Tasty video portraying preparation of sweet snacks *(objectively recorded)* | Food choice behaviour (choosing fruit over a sweet snack) *(objectively recorded)* | 126 |
| Ohannessian 2009^117^ | Cross-sectional | 2006 | USA | High income country | Grade 9-10 high school students | 15.0 [14-16] | High | 2 | No | Time spent on SM | Time spent emailing and instant messaging on an average/typical day | Freq. of daily alcohol consumption in the past 6 months | 328 |
|  |  |  |  |  |  |  |  |  |  | Time spent on SM | Time spent emailing and instant messaging on an average/typical day | Smoking onset | 328 |
| Pegg 2018^118^ | Cross-sectional | 2014 | Australia | High income country | Year 12 students’ part of the Youth Activity Participation Survey | 17.3 [NR] | High | 2 | Yes | Exposure to health-risk behaviour content | SNS alcohol exposure in the past 6 months | Alcohol use in the past 6 months | 793 |
|  |  |  |  |  |  |  |  |  |  | Freq. of SM use | Freq. of SNS use (intensity) | Alcohol use in the past 6 months | 793 |
| Perez 2022^119^ | Cohort | 2014-2016 | USA | High income country with mixed SEP | Adolescents part of the Population Assessment of Tobacco and Health Study | NR  [12-17] | Low | 1 | No | Exposure to health-risk behaviour content | Exposure to tobacco related content on SM in the past 12 months (including e-cigarettes) | Dual use of e-cigarettes and at least one combustible product in the past month | 16,109,064 |
| Prince 2021^120^ | Cross-sectional | 2015-2017 | USA | High income country with mixed SEP | Grade 7-12 students living on or near reservations part of the Our Youth, Our Future Survey | 14.78  [NR] | High | 1 | Yes | Freq. of SM use | Freq. of Snapchat use | Past month opioid use | 25 |

| Author and year | Study design | Study period | Country | Equity | Participants | Mean age [range] | Risk of bias | N°. of dp | MA? | Exposure | Exposure measure | Outcome measure | N |
| --- | --- | --- | --- | --- | --- | --- | --- | --- | --- | --- | --- | --- | --- |
| Qutteina 2022^121^ | Cross-sectional | 2019 | Belgium | High income country with mixed SEP | Adolescents attending 18 secondary schools across Flanders | 15  [11-19] | Mod | 5 | No | Exposure to health-risk behaviour content | Exposure to non-core foods (energy dense, low nutrient: sweetened drinks, sweets, salty/savoury snacks) on SM | Freq. of sweet intake in the past month | 1002 |
|  |  |  |  |  |  |  |  |  |  | Exposure to health-risk behaviour content | Exposure to non-core foods (energy dense, low nutrient: sweetened drinks, sweets, salty/savoury snacks) on SM | Consumption of sweets in the past month (g/day) | 1002 |
|  |  |  |  |  |  |  |  |  |  | Exposure to health-risk behaviour content | Exposure to non-core foods (energy dense, low nutrient: sweetened drinks, sweets, salty/savoury snacks) on SM | Freq. of soft drink intake in the past month | 1002 |
|  |  |  |  |  |  |  |  |  |  | Exposure to health-risk behaviour content | Exposure to non-core foods (energy dense, low nutrient: sweetened drinks, sweets, salty/savoury snacks) on SM | Consumption of soft drinks in the past month (ml/day) | 1002 |
|  |  |  |  |  |  |  |  |  |  | Exposure to health-risk behaviour content | Exposure to non-core foods (energy dense, low nutrient: sweetened drinks, sweets, salty/savoury snacks) on SM | Freq. of fried food intake in the past month | 1002 |
| Riehm 2021^122^ | Cross-sectional | 2015 | USA | High income country with mixed SEP | Students from 10 public high schools in Los Angeles part of the Happiness & Health Study | 16.5  [NR] | High | 1 | Yes | Freq. of SM use | Freq. of posting own photos, images, videos, status updates, or blogs on SM | Ever use of alcohol | 2373 |
| Roditis 2016^123^ | Cross-sectional | 2014-2015 | USA | High income country | Grade 9 and 12 high school students residing in California | 16.1 [NR] | Mod | 2 | No | Other SM activities | Ever seen a message posted on SM about the risks/bad things of using marijuana | Ever use of marijuana | 786 |
|  |  |  |  |  |  |  |  |  |  | Exposure to health-risk behaviour content | Ever seen a message posted on SM about the benefits/  good things of using marijuana | Ever use of marijuana | 786 |

| Author and year | Study design | Study period | Country | Equity | Participants | Mean age [range] | Risk of bias | N°. of dp | MA? | Exposure | Exposure measure | Outcome measure | N |
| --- | --- | --- | --- | --- | --- | --- | --- | --- | --- | --- | --- | --- | --- |
| Romo 2017^124^ | Cross-sectional | 2014 | USA | High income country with mixed SEP | Adolescents recruited from 3 primary care paediatric clinics in Northern Manhattan | 16.0 [13-21] | High | 8 | Yes | Freq. of SM use | Freq. of SNS use *(via validated tool)* | Inconsistent condom use overall and at last sex | 333 |
|  |  |  |  |  |  |  |  |  |  | Freq. of SM use | Freq. of SM app use *(via validated tool)* | Inconsistent condom use overall and at last sex | 333 |
|  |  |  |  |  |  |  |  |  |  | Freq. of SM use | Freq. of SNS use *(via validated tool)* | 4 or more lifetime partners in sexually active participants | 333 |
|  |  |  |  |  |  |  |  |  |  | Freq. of SM use | Freq. of SNS use *(via validated tool)* | More than 1 partner in the past 3 months in sexually active participants | 333 |
|  |  |  |  |  |  |  |  |  |  | Freq. of SM use | Freq. of SNS use *(via validated tool)* | STI diagnosis ever in sexually active participants | 333 |
|  |  |  |  |  |  |  |  |  |  | Freq. of SM use | Freq. of SNS use *(via validated tool)* | History of no use of long-term acting reversible contraception in sexually active female participants | NR |
|  |  |  |  |  |  |  |  |  |  | Freq. of SM use | Freq. of SNS use *(via validated tool)* | History of no use of regular forms of hormonal contraception in sexually female active participants | NR |
|  |  |  |  |  |  |  |  |  |  | Freq. of SM use | Freq. of SNS use *(via validated tool)* | History of no use of emergency contraception in sexually active female participants | NR |
| Rutter 2021^125^ | Cross-sectional | 2019 | USA | High income country with mixed SEP | Adolescents in the USA | 14.6  [12-17] | Mod | 1 | No | Freq. of SM use | Freq. of SM use (checking and posting) | Physical activity | 4592 |
| Sampasa-  Kanyinga 2015^126^ | Cross-sectional | 2013 | Canada | High income country with mixed SEP | Grade 11-12 high school students’ part of the Ontario Student Drug Use and Health Survey | 15.2 [NR] | High | 2 | Yes | Time spent on SM | Time spent on SM websites either posting or browsing per day | Cannabis use in the past year | 5329 |
|  |  |  |  |  |  |  |  |  |  | Time spent on SM | Time spent on SM websites either posting or browsing per day | Tobacco use in the past year | 5329 |

| Author and year | Study design | Study period | Country | Equity | Participants | Mean age [range] | Risk of bias | N°. of dp | MA? | Exposure | Exposure measure | Outcome measure | N |
| --- | --- | --- | --- | --- | --- | --- | --- | --- | --- | --- | --- | --- | --- |
| Sampasa-  Kanyinga 2015^127^ | Cross-sectional | 2013 | Canada | High income country with mixed SEP | Grade 7-12 high school students’ part of the Ontario Student Drug Use and Health Survey | 15.2 [11-19] | Low | 3 | No | Time spent on SM | Time spent on SM websites either posting or browsing per day | Skipping breakfast in the past 5 days | 9858 |
|  |  |  |  |  |  |  |  |  |  | Time spent on SM | Time spent on SM websites either posting or browsing per day | Sugar-sweetened beverage consumption in the past 7 days | 9858 |
|  |  |  |  |  |  |  |  |  |  | Time spent on SM | Time spent on SM websites either posting or browsing per day | Energy drink consumption in the past 7 days | 9858 |
| Sampasa  Kanyinga 2016^128^ | Cross-sectional | 2013 | Canada | High income country with mixed SEP | Grade 7-12 high school students’ part of the Ontario Student Drug Use and Health Survey | 15.2 [11-20] | Low | 6 | Yes | Time spent on SM | Time spent on SM websites either posting or browsing per day | Occasional alcohol use in the past year | 4814 |
|  |  |  |  |  |  |  |  |  |  | Time spent on SM | Time spent on SM websites either posting or browsing per day | Regular alcohol use in the past year | 4814 |
|  |  |  |  |  |  |  |  |  |  | Time spent on SM | Time spent on SM websites either posting or browsing per day | Binge drinking in the past month | 4814 |
| Sampasa-Kanyinga 2016^129^ | Cross-sectional | 2013 | Canada | High income country with mixed SEP | Grade 7-12 public high school students’ part of the Ontario Student Drug Use and Health Survey | 15.3  [11-19] | Low | 2 | No | Time spent on SM | Time spent on SM websites either posting or browsing per day | Not meeting physical activity recommendation in past week (≥60 minutes per day of moderate to vigorous physical activity on all 7 days) | 9388 |
| Sampasa-  Kanyinga 2018^130^ | Cross-sectional | 2013 | Canada | High income country with mixed SEP | Grade 9-10 high school students’ part of the Ontario Student Drug Use and Health Survey | 16.1 [NR] | Mod | 1 | No | Time spent on SM | Time spent on SM websites either posting or browsing per day | Ever use of e-cigarettes | 2841 |
| Sandercock 2016^131^ | Cross-sectional | 2014 | UK | HIC Mixed SEP | Grade 6-11 junior and high school students residing in the East of England | 13.5 [NR] | Low | 5 | No | Time spent on SM | Time spent on SM on a normal day | Daily sedentary time | 678 |
|  |  |  |  |  |  |  |  |  |  | Time spent on SM | Time spent on SM on a normal day | High sedentary time | 678 |
|  |  |  |  |  |  |  |  |  |  | Time spent on SM | Time spent on SM on a normal day | Low cardio-respiratory fitness in female participants | 308 |

| Author and year | Study design | Study period | Country | Equity | Participants | Mean age [range] | Risk of bias | N°. of dp | MA? | Exposure | Exposure measure | Outcome measure | N |
| --- | --- | --- | --- | --- | --- | --- | --- | --- | --- | --- | --- | --- | --- |
| Savolainen 2020^132^ | Cross-sectional | 2017-2019 | USA, South Korea, Finland, and Spain | High income country | Adolescents in 4 countries across 4 continents | NR [15-17] | High | 20 | Yes | Freq. of SM use | Freq. of Facebook use *(via validated tool)* | Hazardous alcohol use *(via AUDIT-C)* | 329 (USA)  264 (KOR)  154 (FIN)  314 (ESP) |
|  |  |  |  |  |  |  |  |  |  | Freq. of SM use | Freq. of YouTube use *(via validated tool)* | Hazardous alcohol use *(via AUDIT-C)* | As above |
|  |  |  |  |  |  |  |  |  |  | Freq. of SM use | Freq. of Twitter use *(via validated tool)* | Hazardous alcohol use *(via AUDIT-C)* | As above |
|  |  |  |  |  |  |  |  |  |  | Freq. of SM use | Freq. of Instagram use *(via validated tool)* | Hazardous alcohol use *(via AUDIT-C)* | As above |
|  |  |  |  |  |  |  |  |  |  | Freq. of SM use | Freq. of instant messaging *(via validated tool)* | Hazardous alcohol use *(via AUDIT-C)* | As above |
| Self-Brown 2018^133^ | Cross-sectional | 2014 | Uganda | Low-middle income country with low SEP | Adolescents living in slums part of the Kampala Youth Survey | 17.0 [12-18] | High | 2 | Yes | Freq. of SM use | Presence of SM use *(via validated tool)* | Transactional Sex in sexually active youth *(via validated tool)* | 593 |
| Shan 2022^134^ | Cohort | 2013-2018 | USA | High income country with mixed SEP | Adolescents part of the Population Assessment of Health and Tobacco Study | NR  [12-14] | Low | 2 | Yes | Exposure to health-risk behaviour content | Followed tobacco brands (e.g., Marlboro, Newport, American Spirit, Vuse) on Facebook/Twitter or other SM sites | Initiation of cigarettes | 6557 |
|  |  |  |  |  |  |  |  |  |  | Exposure to health-risk behaviour content | Followed tobacco brands (e.g., Marlboro, Newport, American Spirit, Vuse) on Facebook/Twitter or other SM sites | Initiation of e-cigarettes | 6632 |

| Author and year | Study design | Study period | Country | Equity | Participants | Mean age [range] | Risk of bias | N°. of dp | MA? | Exposure | Exposure measure | Outcome measure | N |
| --- | --- | --- | --- | --- | --- | --- | --- | --- | --- | --- | --- | --- | --- |
| Sharma 2021^135^ | Cross-sectional | NR | India | Low-middle income country with mixed SEP | Grade 9-12 students in Barwala village, Delhi | NR | High | 1 | Yes | Exposure to health-risk behaviour content | Exposure to tobacco adverts on SM | Smokeless tobacco use | 652 |
| Shimoga 2019^136^ | Cross-sectional | 2014-2015 | USA | High income country with mixed SEP | Grade 8,10 and 12 school students’ part of the Monitoring the Future Survey | NR | Low | 1 | No | Freq. of SM use | Freq. of SM use | Freq. of vigorous physical activity | 19,543 |
| Smout 2021^137^ | Cohort | 2012-2015 | Australia | High income country | Middle school students’ part of the **CAP study** | 13.4  [13-16] | Mod | 2 | Yes | Time Spent on SM | Time spent on SM on a typical day | Days drinking per month | 441 |
|  |  |  |  |  |  |  |  |  |  | Exposure to health-risk behaviour content | Exposure to peer-generated content on SM depicting risky substance use | Days drinking per month | 441 |
| Soneji 2018^138^ | Cohort | 2013-2015 | USA | High income country with mixed SEP | Sample members of the Population Assessment of Tobacco and Health Study | NR [12-17] | Low | 4 | Yes | Freq. of SM use | Freq. of social networking account use | Initiation of binge drinking in the past month | 8542 |
|  |  |  |  |  |  |  |  |  |  | Freq. of SM use | Freq. of social networking account use | Initiation of tobacco use in the past month | 9067 |
|  |  |  |  |  |  |  |  |  |  | Freq. of SM use | Freq. of social networking account use | Increased frequency of tobacco product use | 11,996 |
|  |  |  |  |  |  |  |  |  |  | Freq. of SM use | Freq. of social networking account use | Progression from tobacco single-product to poly-product use (i.e., ≥ 2 products) | 10,928 |
| Stevens 2017^139^ | Cross-sectional | 2013-2014 | USA | High income country with low SEP | Sample members of a larger mixed methods study | 18.2 [13-24] | Low | 2 | No | Other SM activities | Exposure to contraception information on SNS in the past month | No condom use at last intercourse in sexually active participants | 172 |
|  |  |  |  |  |  |  |  |  |  | Other SM activities | Exposure to contraception information on SNS in the past month | No contraception use at last intercourse in sexually active participants | 175 |
| Suwanwong 2021^140^ | Cross-sectional | 2017 | Thailand | Low-middle income country | Adolescents part of the Cigarette Smoking and Alcohol Drinking Behaviour Survey | NR  [15-19] | High | 2 | No | Other SM activities | Exposure to Anti-smoking SM campaign | Smoking status (occasional smoker) | 5669 |
|  |  |  |  |  |  |  |  |  |  | Other SM activities | Exposure to Anti-smoking SM campaign | Smoking status (daily smoker) | 5851 |

| Author and year | Study design | Study period | Country | Equity | Participants | Mean age [range] | Risk of bias | N°. of dp | MA? | Exposure | Exposure measure | Outcome measure | N |
| --- | --- | --- | --- | --- | --- | --- | --- | --- | --- | --- | --- | --- | --- |
| Svensson 2020^141^ | Repeat cross-sectional | 2016-2019 | Sweden | High income country | Students from 17 secondary schools in 8 small municipalities in the county of Skåne part of the Öckerö project | NR  [14-15] | Mod | 3 | No | Freq. of SM use | Freq. of posting information on Facebook, Instagram, Snapchat, or other SM | Lifetime alcohol use | 3733 |
|  |  |  |  |  |  |  |  |  |  | Freq. of SM use | Freq. of posting information on Facebook, Instagram, Snapchat, or other SM | Drunkenness in the past year | 3733 |
|  |  |  |  |  |  |  |  |  |  | Freq. of SM use | Freq. of posting information on Facebook, Instagram, Snapchat, or other SM | Drunkenness in the past month | 3733 |
| Tao 2022^142^ | Cross-sectional | 2020-2021 | USA | High income country with mixed SEP | Adolescents | 16.5  [15-18] | High | 2 | Yes | Time Spent on SM | Average time on SM per week | Alcohol use disorder *(via AUDIT*) | 407 |
|  |  |  |  |  |  |  |  |  |  | Time Spent on SM | Average time on SM per week | Illicit drug use problems *(via validated tool)* | 407 |
| Trangenstein 2019^143^ | Cross-sectional | 2018 | USA | High income country with mixed SEP | Adolescents residing in USA states with legalised retail cannabis | NR [15-19] | High | 2 | No | Exposure to health-risk behaviour content | Liked/  followed cannabis business pages on Facebook,  Twitter and/or Instagram | Cannabis use in the past year | 482 |
|  |  |  |  |  |  |  |  |  |  | Exposure to health-risk behaviour content | Liked/  followed cannabis business pages on Facebook, Twitter and/or Instagram | Cannabis use in the past 28 days | 482 |
| Tsitsika 2009^144^ | Cross-sectional | 2007-2008 | Greece | High income country with mixed SEP | Grade 9-10 urban district school students in Athens | 14.9 [NR] | High | 1 | Yes | Freq. of SM use | Ever accessed the internet to visit chat rooms | Freq. of pornographic internet site use | 344 |
| Tsitsika 2011^145^ | Cross-sectional | 2007-2008 | Greens | High income country with mixed SEP | Grade 9-10 school students in the urban district of Athens | 14.9 [NR] | High | 1 | Yes | Freq. of SM use | Presence of internet chat room use | Internet gambling practices (not via SM) at least once per week | 484 |
| Vandenbosch 2016^146^ | Cross-sectional | 2010 | Belgium | High income country with mixed SEP | School students’ part of the MORES Panel Study | 15.4 [12-18] | High | 1 | Yes | Freq. of SM use | Freq. of chat room use | Use of erotic contact websites in the past 6 months | 1163 |
| Vannucci 2019^147^ | Cohort | 2016-2017 | USA | High income country with mixed SEP | Grade 7-8 middle school students’ part of the PANDA Research Project | 12.7 [11-14] | Low | 1 | Yes | Freq. of SM use | Freq. of SM use | Delinquent behaviours in the past 6 months *(via validated tool)* | 563 |

| Author and year | Study design | Study period | Country | Equity | Participants | Mean age [range] | Risk of bias | N°. of dp | MA? | Exposure | Exposure measure | Outcome measure | N |
| --- | --- | --- | --- | --- | --- | --- | --- | --- | --- | --- | --- | --- | --- |
| Vazquez-Nava 2020^148^ | Cross-sectional | NR | Mexico | Low-middle income country with mixed SEP | Urban school students in North-eastern Mexico | NR [13-19] | High | 1 | Yes | Freq. of SM use | Presence of use of social networks WhatsApp/Facebook *(via validated tool)* | Tobacco smoking *(via validated tool)* | 1328 |
| Vente 2020^149^ | Cross-sectional | 2016-2018 | USA | High income country | Adolescents seen at a paediatric clinic at an urban medical centre | 16.8  [12-21] | High | 2 | No | Freq. of SM use | Use of ≥4 SM applications per day | Sexting | 179 |
|  |  |  |  |  |  |  |  |  |  | Time Spent on SM | Time spent on SM per day | Sexting | 179 |
| Wana 2019^150^ | Cross-sectional | 2017 | Ethiopia | Low-middle income country | Pre-college students residing in Adama Town | NR | Mod | 1 | No | Freq. of SM use | Presence of SM use | Risky sexual behaviour | 346 |
| Ward 2022^151^ | Cross-sectional | 2017-2018 | USA | High income country | Adolescents in the Seattle metro area part of a larger longitudinal experimental study | 18.4  [15-20] | Mod | 3 | Yes | Freq. of SM use | Freq. of Facebook use per day | Past month typical drinks per week | 274 |
|  |  |  |  |  |  |  |  |  |  | Freq. of SM use | Freq. of Instagram use per day | Past month typical drinks per week | 274 |
|  |  |  |  |  |  |  |  |  |  | Freq. of SM use | Freq. of Facebook use per day | Past month peak drinks per occasion | 274 |
| Whitehill 2020^152^ | Cross-sectional | 2018 | USA | High income country with mixed SEP | Adolescents (aged 15-19) residing in US states with legalised retail cannabis | NR [15-19] | High | 4 | Yes | Freq. of SM use | Freq. of SM use | Lifetime cannabis use | 469 |
|  |  |  |  |  |  |  |  |  |  | Freq. of SM use | Presence of Facebook use | Lifetime cannabis use | 469 |
|  |  |  |  |  |  |  |  |  |  | Time spent on SM | Time spent on SM per day | Lifetime cannabis use | 469 |
|  |  |  |  |  |  |  |  |  |  | Freq. of SM use | Presence of Instagram use | Lifetime cannabis use | 469 |
| Widman 2014^153^ | Cross-sectional | 2014 | USA | High income country | Grade 9-10 high school students’ part of a larger study | 17.4 [16-19] | High | 2 | Yes | Freq. of SM use | Used technology based sexual communication to communicate with dating partners about using condoms | Inconsistent condom use in the past 6 months | 176 |
|  |  |  |  |  |  |  |  |  |  |  | Used technology based sexual communication to communicate with dating partners about risk of pregnancy | Inconsistent condom use in the past 6 months | 176 |
| Worku 2022 | Cross-sectional | 2021 | Ethiopia | Low-middle income country with low SEP | Female high school students at selected high schools of Yeka Sub-city, Addis Ababa | NR  [14-16] | Mod | 1 | No | Time Spent on SM | Stayed more than 2 hrs/day on SM | Low dietary diversity *(via FANTA)* | 284 |

| Author and year | Study design | Study period | Country | Equity | Participants | Mean age [range] | Risk of bias | N°. of dp | MA? | Exposure | Exposure measure | Outcome measure | N |
| --- | --- | --- | --- | --- | --- | --- | --- | --- | --- | --- | --- | --- | --- |
| Wulff 2021^154^ | Cross-sectional | 2015 | Germany | High income country | Adolescent obesity therapy participants | NR  [11-17] | Mod | 1 | No | Freq. of SM use | Freq. of WhatsApp use | Physical inactivity (exercised 0-2 days per week) | 228 |
| Yao 2022^155^ | Cross-sectional | NR | China | High income country | Grade 4-10 elementary and middle school students | 13.35  [6-18] | Mod | 2 | No | Exposure to health-risk behaviour content | Exposure to content (including text and pictures) about drinking or smoking (e.g., saw drinking-related information) | Tobacco and alcohol use | 1491 |

**Legend:** Where exposure ascertainment was via objectively recorded social media usage data, independent of user reports or via a validated measurement tool, this is stated in italics; where outcome ascertainment was via independent clinical assessment, a validated tool, or medical/administrative records, this is stated in italics. All other measures are self-report. An adapted Newcastle Ottawa Scale (NOS) was used to assess risk of bias for cross-sectional and cohort studies and the Cochrane risk of bias 2 tool for randomised trials (RoB-2) was used for randomised control trial studies. All included studies were assessed using synthesis without meta-analysis (SWiM), excluding one study^73^ which was included due to potential double counting of study participants; we were however able to include estimates from this studies in meta-analysis stratified by outcome where this issue did not occur. Abbreviations: ADHD = Attention deficit hyperactivity disorder; ASA24 = Dietary Assessment Tool; AUDIT/C = Alcohol Use Disorders Identification Test; CAGI/GPSS = Gambling Problem Severity Subscale of the Canadian Adolescent Gambling Index; dp = number of datapoints; DSM-IV-MR-J = Diagnostic and Statistical Manual of Mental Disorders-IV-Multiple Response- Adapted for Juveniles (assessment of adolescent gambling); ESP = Spain; FIN = Finland; Freq = Frequency; HEI = Healthy Eating Index; KOR = South Korea; MA = Study included in meta-analysis, subgroup or sensitivity analysis; Mod = Moderate RoB; N = Number of study participants; NR = Not reported; PAQ-C/A = Physical Activity Questionnaire for Children and Adolescents; RCS = Repeat cross-sectional study; RCT = Randomised control trial; RoB-2 = Cochrane risk of bias 2 tool for randomised trials; SBS = Sexting Behaviour Scale; SDQ = Strengths and Difficulties Questionnaire; SEP = Socioeconomic position; SOGS-RA = South Oaks Gambling Screen-Revised for Adolescents; T = Timepoint; UK = United Kingdom; USA = United States; WHO = World Health Organisation; YSR = Youth Self-Report and ∆ = Change.

##### Figure A. Map of geographical distribution of included study populations

**Key**

|  | >4 studies |
| --- | --- |
|  | 3 studies |
|  | 2 studies |
|  | 1 study |
|  | No data |

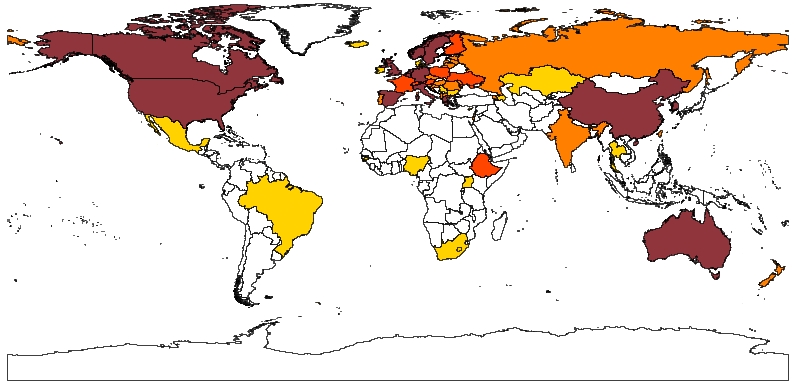

Legend: Studies undertaken in more than one country contribute multiple datapoints to the map. ^40,60,62,132,^

### Appendix-12. Characteristics of excluded studies

##### Table A. List of studies excluded at full text screening (n=571 studies) with reasons for exclusion

| **Author and year** | **Title** | **Publication source** | **Reason for exclusion** |
| --- | --- | --- | --- |
| Abara 2014 | Understanding internet sex-seeking behaviour and sexual risk among young men who have sex with men: evidence from a cross-sectional study. | Sexually Transmitted Infections | Incorrect population |
| Abdi 2015 | Personal, social, and environmental risk factors of problematic gambling among high school adolescents in Addis Ababa, Ethiopia | Journal of Gambling Studies | Incorrect exposure |
| Abed-Ali 2018 | Violence among high school female students in Baghdad city | Indian Journal of Public Health Research and Development | Incorrect exposure |
| Acar 2020 | Eating attitudes and physical appearance comparison with others in daily life versus on social media in adolescents | Journal of Adolescent Health | Incorrect study type: conference proceeding or abstract |
| Adam 2011 | When do online sexual fantasies become reality? The contribution of erotic chatting via the Internet to sexual risk-taking in gay and other men who have sex with men. | Health Education Research | Incorrect exposure |
| Adams 2010 | Correlates of physical activity in young American Indian children: lessons learned from the Wisconsin Nutrition and Growth Study | Journal of Public Health Management and Practice | Incorrect exposure |
| Adams 2019 | Predictors of overweight and obesity in American Indian families with young children | Journal of Nutrition Education and Behaviour | Incorrect exposure |
| Adebayo 2006 | Gender, internet use, and sexual behaviour orientation among young Nigerians | Cyberpsychology and Behaviour | Incorrect exposure |
| Afolabi 2015 | Media exposure and weight concern? | Child and Adolescent Health Issues: A tribute to the paediatrician Donald E Greydanus | Incorrect study type: commentary/editorial/non-systematic review/theses/book chapter(s) |
| Agaku 2014 | Trends in exposure to pro-tobacco advertisements over the Internet, in newspapers/magazines, and at retail stores among U.S. middle and high school students, 2000-2012 | Preventive Medicine | Incorrect exposure |
| Aggio 2012 | Temporal relationships between screen-time and physical activity with cardiorespiratory fitness in English schoolchildren: a 2-year longitudinal study. | Preventive Medicine | Incorrect exposure |
| Agurcia-Parker 2009 | An investigation into the relationship between screen time, consumption of advertised foods, and physical activity among Texas 4th grade elementary school children | Dissertation Abstracts International: Section B: The Sciences and Engineering | Incorrect study type: commentary/editorial/non-systematic review/theses/book chapter(s) |
| Ahern 2015 | Risky behaviours and social networking sites | Journal of Psychosocial Nursing & Mental Health Services | Incorrect study type: commentary/editorial/non-systematic review/theses/book chapter(s) |
| Aires 2010 | A 3-year longitudinal analysis of changes in fitness, physical activity, fatness, and screen time | Acta Paediatrica, International Journal of Paediatrics | Incorrect exposure |
| Aires 2010 | A 3-year longitudinal analysis of changes in Body Mass Index | International Journal of Sports Medicine | Incorrect exposure |

| **Author and year** | **Title** | **Publication source** | **Reason for exclusion** |
| --- | --- | --- | --- |
| Al-Ajlouni 2018 | Partner meeting venue typology and sexual risk behaviours among French men who have sex with men. | International Journal of STD & AIDS | Incorrect exposure |
| Albert 2018 | #consumingitall: Understanding the complex relationship between media consumption and eating behaviours | Dissertation Abstracts International: Section B: The Sciences and Engineering | Incorrect study type: commentary/editorial/non-systematic review/theses/book chapter(s) |
| Albury 2018 | Young people, digital media research and counter public sexual health | Sexualities | Incorrect study type: commentary/editorial/non-systematic review/theses/book chapter(s) |
| Alghadir 2020 | Differences among Saudi and expatriate students: Body composition indices, sitting time associated with media use and physical activity pattern | International Journal of Environmental Research and Public Health | Incorrect exposure |
| Alhabash 2021 | Trick or drink: offline and social media hierarchical normative influences on Halloween celebration drinking | Health Communication | Incorrect population |
| Al-Hamdani 2021 | Perceptions and experiences of vaping among youth and young adult e-cigarette users: considering age, gender, and tobacco use | Journal of Adolescent Health | Incorrect population |
| Al-Hamdani 2022 | Do perceptions and experiences of vaping among youth and young adults differ by device type? | Addiction Research & Theory | No relevant outcome(s) |
| Al-Hazzaa 2011 | Physical activity, sedentary behaviours, and dietary habits among Saudi adolescents relative to age, gender, and region | The International Journal of Behavioural Nutrition and Physical Activity | Incorrect exposure |
| Al-Hazzaa 2019 | Activity energy expenditure, screen time and dietary habits relative to gender among Saudi youth: interactions of gender with obesity status and selected lifestyle behaviours | Asia Pacific Journal of Clinical Nutrition | Incorrect exposure |
| Alhusaini 2020 | Cross-cultural variation in BMI, sedentary behaviour, and physical activity in international schoolgirls residing in Saudi Arabia | International Journal of Environmental Research and Public Health | Incorrect exposure |
| Allen 2017 | Mobile phone and internet use mostly for sex-seeking and associations with sexually transmitted infections and sample characteristics among black/African American and Hispanic/Latino men who have sex with men in 3 US cities | Sexually Transmitted Diseases | Incorrect population |
| Allen 2018 | The dirt on clean eating: a cross sectional analysis of dietary intake, restrained eating, and opinions about clean eating among women. | Nutrients | Incorrect population |
| Allender 2011 | Associations between activity-related behaviours and standardized BMI among Australian adolescents | Journal of Science and Medicine in Sport | Incorrect exposure |
| Alosaimi 2016 | Smartphone addiction among university students in Riyadh, Saudi Arabia. | Saudi Medical Journal | Incorrect population |

| **Author and year** | **Title** | **Publication source** | **Reason for exclusion** |
| --- | --- | --- | --- |
| Altenburg 2012 | Direction of the association between body fatness and self-reported screen time in Dutch adolescents | International Journal of Behavioural Nutrition and Physical Activity | Incorrect exposure |
| Altenburg 2017 | Actual and perceived weight status and its association with slimming and energy-balance related behaviours in 10- to 12-year-old European children: the ENERGY-project | Pediatric Obesity | Incorrect exposure |
| Alvarez-Jimenez 2019 | HORYZONS trial: Protocol for a randomised controlled trial of a moderated online social therapy to maintain treatment effects from first-episode psychosis services | BMJ Open | Incorrect study type: commentary/editorial/non-systematic review/theses/book chapter(s) |
| Ammouri 2004 | Correlates of exercise participation in adolescents | Correlates of Exercise Participation in Adolescents | Incorrect exposure |
| Amornsriwata-nakul 2017 | Are Thai children and youth sufficiently active? Prevalence and correlates of physical activity from a nationally representative cross-sectional study | International Journal of Behavioural Nutrition and Physical Activity | Incorrect exposure |
| Andrie 2019 | Gambling involvement and problem gambling correlates among European adolescents: results from the European Network for Addictive Behaviour study. | Social Psychiatry and Psychiatric Epidemiology | Incorrect exposure |
| Arie 2014 | Doctors and teachers receive new guidance on the internet's effect on young people's sex lives and relationships | BMJ (Clinical research ed.) | Incorrect study type: commentary/editorial/non-systematic review/theses/book chapter(s) |
| Armstrong 2018 | An exploration of how simulated gambling games may promote gambling with money | Journal of Gambling Studies | Incorrect study type: commentary/editorial/non-systematic review/theses/book chapter(s) |
| Arriscado 2014 | Factors associated with low adherence to a Mediterranean diet in healthy children in northern Spain | Appetite | Incorrect exposure |
| Arsad 2021 | A systematic review of immersive social media activities and risk factors for sexual boundary violations among adolescents | IUM Medical Journal Malaysia | Incorrect study type: systematic review |
| Arseniev-Koehler 2014 | Peer influence on undergraduates' intention to get drunk by communication formats | Alcoholism: Clinical and Experimental Research | Incorrect study type: conference proceeding or abstract |
| Asad 2015 | Screen-based behaviours of adolescents in Bangladesh | European Journal of Epidemiology | Incorrect exposure |
| Aschbrenner 2019 | Randomized trial of a lifestyle intervention for young adults with serious mental illness in community mental health centres | Schizophrenia Bulletin | Incorrect study type: conference proceeding or abstract |
| Ashford 2017 | Advertising exposure and use of e-cigarettes among female current and former tobacco users of childbearing age | Public Health Nursing | Incorrect population |

| **Author and year** | **Title** | **Publication source** | **Reason for exclusion** |
| --- | --- | --- | --- |
| Asut 2019 | Relationships between screen time, internet addiction and other lifestyle behaviours with obesity among secondary school students in the Turkish Republic of Northern Cyprus | Turkish Journal of Paediatrics | Incorrect exposure |
| Athauda 2020 | Factors influencing alcohol use among adolescents in South Asia: a systematic review | Journal of Studies on Alcohol and Drugs | Incorrect study type: systematic review |
| Atkin 2013 | Determinants of change in children's sedentary time | PLoS ONE | Incorrect exposure |
| Atkin 2021 | Adolescent time use and mental health: a cross-sectional, compositional analysis in the Millennium Cohort Study | BMJ Open | Incorrect exposure |
| Atkinson 2010 | Online behaviours of adolescents: victims, perpetrators, and Web 2.0. | Journal of Sexual Aggression | Incorrect study type: commentary/editorial/non-systematic review/theses/book chapter(s) |
| Attwood 2017 | Using a mobile health application to reduce alcohol consumption: a mixed-methods evaluation of the drinkaware track & calculate units’ application | BMC Public Health | Incorrect exposure |
| Atwood 2017 | Adolescent problematic digital behaviours associated with mobile devices | North American Journal of Psychology | Incorrect exposure |
| Bae 2018 | Selective exposure to misleading information in the new media environment by at-risk youth: A study of pro-smoking YouTube videos | Dissertation Abstracts International Section A: Humanities and Social Sciences | Incorrect study type: commentary/editorial/non-systematic review/theses/book chapter(s) |
| Bai 2016 | The associations of youth physical activity and screen time with fatness and fitness: The 2012 NHANES national youth fitness survey | PLoS ONE | Incorrect exposure |
| Baird 2016 | Social media and substance use | Journal Of Addictions Nursing | Incorrect study type: commentary/editorial/non-systematic review/theses/book chapter(s) |
| Baird 2019 | Teens and vaping: what you need to know | Journal of Addictions Nursing | Incorrect study type: commentary/editorial/non-systematic review/theses/book chapter(s) |
| Bakhali 2016 | Exploring the impact of information seeking behaviours of online health consumers in the Arab world | Studies in Health Technology & Informatics | Incorrect population |
| Balding 2015 | Young People into 2015: The health-related behaviour questionnaire results for over 78,000 young people | Education & Health | Incorrect study type: commentary/editorial/non-systematic review/theses/book chapter(s) |
| Bareghamyan 2021 | Sexual and reproductive health of adolescent girls | Akusherstvo i Ginekologiya (Russian Federation) | Unable to source full text |

| **Author and year** | **Title** | **Publication source** | **Reason for exclusion** |
| --- | --- | --- | --- |
| Barman-Adhikari 2016 | Social networking technology use and engagement in HIV-related risk and protective behaviours among homeless youth | Journal of Health Communication | Incorrect population |
| Barnes 2015 | Maternal correlates of objectively measured physical activity in girls | Maternal and Child Health Journal | Incorrect exposure |
| Barrere 2015 | Oncogenic human papillomavirus infections in 18- to 24-year-old female online daters | Sexually Transmitted diseases | Incorrect exposure |
| Bass III 2016 | Living life online: talking to parents about social media | Contemporary Paediatrics | Incorrect study type: commentary/editorial/non-systematic review/theses/book chapter(s) |
| Bauermeister 2014 | Sexting among young men who have sex with men: results from a national survey | Journal of Adolescent Health | Incorrect exposure |
| Baumgartner 2010 | Assessing causality in the relationship between adolescents' risky sexual online behaviour and their perceptions of this behaviour | Journal of Youth and Adolescence | Duplicate sample |
| Baumgartner 2012 | Unwanted online sexual solicitation and online sexual risk behaviour | Encyclopaedia of cyber behaviour, Vols. I - III. | Incorrect population |
| Bell 2015 | The debate over digital technology and young people | BMJ (Online) | Incorrect study type: commentary/editorial/non-systematic review/theses/book chapter(s) |
| Benotsch 2013 | Sexting, substance use, and sexual risk behaviour in young adults | Journal of Adolescent Health | Incorrect exposure |
| Berchtold 2018 | Daily internet time: towards an evidence-based recommendation? | European Journal of Public Health | Incorrect exposure |
| Bergman 2016 | The association between alcohol use disorder and social network site engagement among treatment seeking emerging adults | Alcoholism: Clinical and Experimental Research | Incorrect study type: conference proceeding or abstract |
| Bergman 2018 | Instagram participation and substance use among emerging adults: the potential perils of peer belonging | Cyberpsychology, Behaviour and Social Networking | Incorrect population |
| Bergman 2020 | Associations between substance use and Instagram participation to inform social network-based screening models: multimodal cross-sectional study | Journal of Medical Internet Research | Incorrect population |
| Berner 2013 | Lifestyle and depressive risk factors associated with problematic internet use in adolescents in an Arabian Gulf culture | Journal of Addiction Medicine | Incorrect population |

| **Author and year** | **Title** | **Publication source** | **Reason for exclusion** |
| --- | --- | --- | --- |
| Bernstein 2018 | Child and adolescent psychiatry case studies: a broad range of ethical dilemmas | Journal of the American Academy of Child and Adolescent Psychiatry | Incorrect study type: conference proceeding or abstract |
| Beullens 2016 | A conditional process analysis on the relationship between the use of social networking sites, attitudes, peer norms, and adolescents' intentions to consume alcohol | Media Psychology | No relevant outcome(s) |
| Beutel 2011 | Regular and problematic leisure-time Internet use in the community: results from a German population-based survey | Cyberpsychology, Behaviour and Social Networking | Incorrect population |
| Bevelander 2018 | Youth's social network structures and peer influences: study protocol MyMovez project - Phase I | BMC Public Health | Incorrect study type: commentary/editorial/non-systematic review/theses/book chapter(s) |
| Bharucha 2018 | Social network use and youth well-being: a study in India | Safer Communities | Incorrect study type: qualitative |
| Bhuyan 2019 | How vaping became fire: Snap streaks, social influencers, and bubble gum | American Sociological Association | Incorrect study type: conference proceeding or abstract |
| Bickham 2020 | Dating app use and sexual risk behaviours: examining aspects of use and motivation | Journal of Adolescent Health | Incorrect study type: conference proceeding or abstract |
| Biddle 2014 | Interventions designed to reduce sedentary behaviours in young people: a review of reviews | British Journal of Sports Medicine | Incorrect study type: systematic review |
| Bilgrami 2017 | Health implications of new-age technologies: a systematic review | Minerva Pediatrica | Incorrect study type: systematic review |
| Black 2013 | Actual versus perceived peer sexual risk behaviour in online youth social networks | Translational Behavioural Medicine | Incorrect exposure |
| Blanchard 2013 | Adolescent perceptions of digital play: A study in third-person effects | Dissertation Abstracts International Section A: Humanities and Social Sciences | Incorrect study type: commentary/editorial/non-systematic review/theses/book chapter(s) |
| Blasco 2019 | Pattern of internet use and parental monitoring of social networks as a predictor of sexting in adolescents: A gender per | Revista de Psicología y Educación | Potentially relevant non-English language |
| Blaszczynski 2016 | Mental health and online, land-based, and mixed gamblers | Journal of Gambling Studies | Incorrect population |
| Blaya 2015 | The young people and risk-taking on the Internet | Neuropsychiatrie de l'Enfance et de l'Adolescence | Incorrect study type: commentary/editorial/non-systematic review/theses/book chapter(s) |

| **Author and year** | **Title** | **Publication source** | **Reason for exclusion** |
| --- | --- | --- | --- |
| Blazquez Barba 2018 | Use of new technologies by adolescents in the search for health information | Atencion Primaria | Potentially relevant non-English language |
| Bleakley 2011 | A model of adolescents' seeking of sexual content in their media choices | Journal of Sex Research | Incorrect exposure |
| Bobkowski 2012 | 'Hit me up and we can get down': US youths' risk behaviours and sexual self-disclosure in MySpace profiles | Journal of Children and Media | Incorrect population |
| Boggs 2017 | The impact of exposure to alcohol advertisements on adolescents: A literature review | International Public Health Journal | Incorrect study type: commentary/editorial/non-systematic review/theses/book chapter(s) |
| Bonnaire 2012 | Internet gambling: What are the risks? | L'Encéphale | Incorrect study type: commentary/editorial/non-systematic review/theses/book chapter(s) |
| Borajy 2019 | Relationship of electronic device usage with obesity and speech delay in children | Family Medicine & Primary Care Review | Incorrect exposure |
| Borden 2019 | Vaping marketers take aim at youth through social media | Chest Physician | Incorrect study type: commentary/editorial/non-systematic review/theses/book chapter(s) |
| Bousono Serrano 2017 | Substance use or abuse, internet use, psychopathology, and suicidal ideation in adolescents | Adicciones | Incorrect exposure |
| Boyland 2016 | Advertising as a cue to consume: a systematic review and meta-analysis of the effects of acute exposure to unhealthy food and non-alcoholic beverage advertising on intake in children and adults | The American Journal of Clinical Nutrition | Incorrect study type: systematic review |
| Boyle 2018 | The social mindfeed project: Using objective assessment methods to better understand the nature of social-media based peer alcohol influence | Alcoholism: Clinical and Experimental Research | Incorrect study type: conference proceeding or abstract |
| Bozzola 2019 | Adolescence, smartphone and tablets: A review of the literature | Italian Journal of Pediatrics | Incorrect study type: conference proceeding or abstract |
| Brailovskaia 2020 | Relationship between depression symptoms, physical activity, and addictive social media use | CyberPsychology, Behavior & Social Networking | Incorrect exposure |
| Branley 2018 | Risky behaviour via social media: The role of reasoned and social reactive pathways. | Computers in Human Behaviour | Incorrect population |
| Braun-Courville 2009 | Exposure to sexually explicit web sites and adolescent sexual attitudes and behaviours | Journal of Adolescent Health | Incorrect exposure |

| **Author and year** | **Title** | **Publication source** | **Reason for exclusion** |
| --- | --- | --- | --- |
| Broaddus 2015 | Social media use and high-risk sexual behaviour among black men who have sex with men: a three-city study | AIDS and Behaviour | Incorrect population |
| Brown 2011 | Older and newer media: Patterns of use and effects on adolescents' health and wellbeing | Journal of Research on Adolescence | Incorrect study type: commentary/editorial/non-systematic review/theses/book chapter(s) |
| Brown 2012 | Too much, much too young | Therapy Today | Incorrect study type: commentary/editorial/non-systematic review/theses/book chapter(s) |
| Brunborg 2017 | Social media use and episodic heavy drinking among adolescents | Psychological Reports | Duplicate sample |
| Brunelle 2012 | Internet gambling, substance use, and delinquent behaviour: an adolescent deviant behaviour involvement pattern | Psychology of Addictive Behaviours | Incorrect exposure |
| Buchanan 2018 | The effects of digital marketing of unhealthy commodities on young people: A systematic review | Nutrients | Incorrect study type: systematic review |
| Buhi 2011 | Evaluating the internet as an std risk environment for teens: Findings from the communication, health, and teens (ch@t) study | Sexually Transmitted Infections | Incorrect study type: conference proceeding or abstract |
| Buhi 2013 | Evaluating the internet as a sexually transmitted disease risk environment for teens: Findings from the communication, health, and teens study | Sexually Transmitted Diseases | Incorrect exposure |
| Buhi 2013 | Teens, the internet, and STD Risk: Findings and lessons learned from the communication, health, and teens (CH@T) study | Sexually Transmitted Infections | Incorrect study type: conference proceeding or abstract |
| Bunnell 2015 | Intentions to smoke cigarettes among never-smoking US middle and high school electronic cigarette users: National youth tobacco survey, 2011-2013 | Nicotine and Tobacco Research | Incorrect exposure |
| Burgos 2013 | The relationship between risky behaviours and perceived victimization in individuals who participate in social networking websites | Dissertation Abstracts International: Section B: The Sciences and Engineering | Incorrect study type: commentary/editorial/non-systematic review/theses/book chapter(s) |
| Burke 2019 | The paradoxical outcomes of observing others' exercise behaviour on social network sites: friends' exercise posts, exercise attitudes, and weight concern | Health Communication | Incorrect population |
| Burns 2021 | Social media preference and condom use behaviours: an analysis of digital spaces with young African American males | Health Education & Behaviour | Incorrect study type: qualitative |
| Butdabut 2021 | Factors predicting sexual risk behaviours of adolescents in North-Eastern Thailand | Studies in Health Technology and Informatics | Incorrect exposure |

| **Author and year** | **Title** | **Publication source** | **Reason for exclusion** |
| --- | --- | --- | --- |
| Cabrera-Nguyen 2016 | Young adults' exposure to alcohol- and marijuana-related content on twitter | Journal Of Studies On Alcohol And Drugs | Incorrect population |
| Caravaca Sanchez 2016 | Prevalence and patterns of traditional bullying victimization and cyber-teasing among college population in Spain | BMC Public Health | Incorrect exposure |
| Card 2017 | Exploring the role of sex-seeking apps and websites in the social and sexual lives of gay, bisexual, and other men who have sex with men: a cross-sectional study | Sexual Health | Incorrect exposure |
| Carrotte 2015 | Predictors of "Liking" three types of health and fitness-related content on social media: A cross-sectional study | Journal of Medical Internet Research | Incorrect population |
| Carrotte 2016 | Who 'likes' alcohol? Young Australians' engagement with alcohol marketing via social media and related alcohol consumption patterns | Australian And New Zealand Journal of Public Health | Incorrect population |
| Castren 2022 | Risk factors for excessive social media use differ from those of gambling and gaming in Finnish youth | International Journal of Environmental Research and Public Health | Incorrect exposure |
| Castro-Calvo 2018 | Building bridges between substance and behavioural addictions: Alcohol consumption and their predictive power over internet and cybersex use and abuse in adolescents | Alcoholism: Clinical and Experimental Research | Incorrect study type: conference proceeding or abstract |
| Cavazos-Rehg 2021 | Exploring How Social Media Exposure and Interactions Are Associated with ENDS and Tobacco Use in Adolescents from the PATH Study | Nicotine and Tobacco Research | Duplicate sample |
| Cemelli 2016 | Video games impact lifestyle behaviors in adults | Topics in Clinical Nutrition | Incorrect population |
| Cen Chen-Sankey 2019 | E-cigarette marketing exposure and subsequent experimentation among youth and young adults | Pediatrics | Incorrect exposure |
| Chan 2017 | The role of gay identity confusion and outness in sex-seeking on mobile dating apps among men who have sex with men: a conditional process analysis | Journal of Homosexuality | Incorrect exposure |
| Chiao 2014 | Adolescent Internet use and its relationship to cigarette smoking and alcohol use: A prospective cohort study | Addictive Behaviours | Incorrect exposure |
| Children & Young People Now 2008 | Youth work support needed in new world of social networking | Children & Young People Now | Incorrect study type: commentary/editorial/non-systematic review/theses/book chapter(s) |
| Cho 2019 | Mechanisms of social media effects on attitudes toward e-cigarette use: motivations, mediators, and moderators in a national survey of adolescents | Journal of Medical Internet Research | No relevant outcome(s) |

| **Author and year** | **Title** | **Publication source** | **Reason for exclusion** |
| --- | --- | --- | --- |
| Chortatos 2020 | Comparing three screen-based sedentary behaviours' effect upon adolescents' participation in physical activity: The ESSENS study | PLoS ONE | Incorrect exposure |
| Clayton 2013 | Loneliness, anxiousness, and substance use as predictors of Facebook use | Computers in Human Behaviour | Incorrect population |
| Coates 2018 | Does social media food marketing influence children's food intake and preferences? | Obesity Facts | Incorrect study type: conference proceeding or abstract |
| Cook 2013 | Online network influences on emerging adults' alcohol and drug use | Journal Of Youth And Adolescence | Incorrect population |
| Cookingham 2015 | The impact of social media on the sexual and social wellness of adolescents | Journal of Paediatric and Adolescent Gynaecology | Incorrect study type: commentary/editorial/non-systematic review/theses/book chapter(s) |
| Coreas 2021 | Smoking susceptibility and tobacco media engagement among youth never smokers | Paediatrics | Duplicate sample |
| Cox 2021 | Profiles of parenting in the digital age: associations with adolescent alcohol and marijuana use | Journal of Studies on Alcohol and Drugs | Incorrect exposure |
| Cruz 2016 | Use of social networking applications (apps) and meeting sites in patients with acute HIV infection in a specialized clinic in Mexico City | Journal of the International AIDS Society | Incorrect study type: conference proceeding or abstract |
| Cruz 2019 | Tobacco marketing and subsequent use of cigarettes, e-cigarettes, and hookah in adolescents | Nicotine and Tobacco Research | Incorrect exposure |
| Cubitt 2014 | Social networking and risk-taking behaviour: the Lynx effect | Journal Of the International Society for Burn Injuries | Incorrect study type: commentary/editorial/non-systematic review/theses/book chapter(s) |
| Cui 2018 | Patterns of online and offline connectedness among gay, bisexual, and other men who have sex with men | AIDS & Behaviour | Incorrect exposure |
| Cureau 2018 | Associations of multiple unhealthy lifestyle behaviours with overweight/obesity and abdominal obesity among Brazilian adolescents: A country-wide survey | Nutrition, Metabolism and Cardiovascular Diseases | Incorrect exposure |
| Curlee 2021 | The role of social media use in adolescent alcohol use accounting for peer alcohol use | Dissertation Abstracts International: Section B: The Sciences and Engineering | Incorrect study type: commentary/editorial/non-systematic review/theses/book chapter(s) |
| Curtis 2018 | Meta-analysis of the association of alcohol-related social media use with alcohol consumption and alcohol-related problems in adolescents and young adults | Alcoholism: Clinical and Experimental Research | Incorrect study type: systematic review |

| **Author and year** | **Title** | **Publication source** | **Reason for exclusion** |
| --- | --- | --- | --- |
| da Costa 2020 | Association between lifestyle behaviours and health-related quality of life in a sample of Brazilian adolescents | International Journal of Environmental Research and Public Health | No relevant outcome(s) |
| da Costa 2022 | Movement behaviours and their association with depressive symptoms in Brazilian adolescents: A cross-sectional study | Journal of Sport and Health Science | Duplicate sample |
| Dai 2017 | Geographic variations in electronic cigarette advertisements on Twitter in the United States | International Journal of Public Health | Incorrect population |
| Dalisay 2022 | Exposure to tobacco and betel nut content on social media, risk perceptions, and susceptibility to peer influence among early adolescents in Guam | Addictive Behaviours Reports | No relevant outcome(s) |
| D'Angelo 2019 | Facebook-induced friend shift and identity shift: a longitudinal study of Facebook posting and collegiate drinking | Cyberpsychology, Behaviour and Social networking | Incorrect population |
| Das 2016 | Interventions for adolescent substance abuse: an overview of systematic reviews | Journal of Adolescent Health | Incorrect study type: systematic review |
| Davis 2021 | Temporal, sex-specific, social media-based alcohol influences during the transition to college | Substance Use & Misuse | Incorrect population |
| Dawson 2019 | Exploring technology-mediated social interactions among adolescents with ADHD | Dissertation Abstracts International: Section B: The Sciences and Engineering | Incorrect study type: commentary/editorial/non-systematic review/theses/book chapter(s) |
| de Bruijn 2016 | European longitudinal study on the relationship between adolescents' alcohol marketing exposure and alcohol use | Addiction | Incorrect exposure |
| Deforche 2015 | Changes in weight, physical activity, sedentary behaviour, and dietary intake during the transition to higher education: A prospective study | International Journal of Behavioural Nutrition and Physical Activity | Incorrect exposure |
| Depue 2015 | Encoded exposure to tobacco use in social media predicts subsequent smoking behaviour | American Journal of Health Promotion | Incorrect population |
| De-Sola 2019 | Cell phone use habits among the Spanish population: contribution of applications to problematic use | Frontiers in Psychiatry | Incorrect exposure |
| Diaz 2022 | Online tobacco advertising and current chew, dip, snuff, and snus use among youth and young adults, 2018-2019 | International Journal of Environmental Research and Public Health | Incorrect population |
| Divecha 2012 | Tweeting about testing: Do low-income, parenting adolescents and young adults use new media technologies to communicate about sexual health? | Perspectives on Sexual and Reproductive Health | Incorrect exposure |

| **Author and year** | **Title** | **Publication source** | **Reason for exclusion** |
| --- | --- | --- | --- |
| Doggett 2019 | Examining the association between exposure to various screen time sedentary behaviours and cannabis use among youth in the COMPASS study | Society of Social Medicine - Population Health | Incorrect exposure |
| Dolcini 2014 | A new window into adolescents' worlds: The impact of online social interaction on risk behaviour | Journal of Adolescent Health | Incorrect study type: commentary/editorial/non-systematic review/theses/book chapter(s) |
| Domingues-Montanari 2017 | Clinical and psychological effects of excessive screen time on children | Journal of Paediatrics and Child Health | Incorrect study type: commentary/editorial/non-systematic review/theses/book chapter(s) |
| Doornwaard 2015 | Adolescents' use of sexually explicit Internet material and their sexual attitudes and behaviour: Parallel development and directional effects | Developmental Psychology | Incorrect exposure |
| Dowdell 2011 | Original research: online social networking patterns among adolescents, young adults, and sexual offenders | The American Journal of Nursing | No relevant outcome(s) |
| Dowdell 2011 | Risky internet behaviours of middle-school students: Communication with online strangers and offline contact | Computers Informatics Nursing | No relevant outcome(s) |
| Dowdell 2022 | Problematic behaviours and predicting online risk behaviours in high school students | The Journal of School Nursing | Incorrect exposure |
| Dowell 2009 | Clustering of internet risk behaviours in a middle school student population | Journal of School Health | Incorrect exposure |
| Drescher 2011 | Caffeine and screen time in adolescence: associations with short sleep and obesity | Journal of Clinical Sleep Medicine | Incorrect exposure |
| Dubuc 2020 | Lifestyle habits predict academic performance in high school students: The adolescent student academic performance longitudinal study (ASAP) | International Journal of Environmental Research and Public Health | No relevant outcome(s) |
| Dumas 2019 | Am I cool now? Examining the relations between need for popularity, alcohol-related social media posts and heavy drinking among emerging adults | Alcoholism: Clinical and Experimental Research | Incorrect study type: conference proceeding or abstract |
| Dumas 2021 | Everyone loves my beer pong pics! examining feedback on social network sites and its role in shaping young adult binge drinking behaviour | Alcoholism: Clinical and Experimental Research | Incorrect study type: conference proceeding or abstract |
| Dumas 2021 | Likelihood of posting alcohol-related content scale | Alcoholism: Clinical and Experimental Research | Incorrect study type: conference proceeding or abstract |
| Dunaev 2016 | Seeking safe sex information: Social media use, gossip, and sexual health behaviour among minority youth | Journal of Adolescent Health | Incorrect study type: conference proceeding or abstract |

| **Author and year** | **Title** | **Publication source** | **Reason for exclusion** |
| --- | --- | --- | --- |
| Dunlop 2016 | Exposure to internet-based tobacco advertising and branding: results from population surveys of Australian youth 2010-2013 | Journal of Medical Internet Research | Incorrect exposure |
| Duplaga 2020 | The use of fitness influencers' websites by young adult women: a cross-sectional study | International Journal of Environmental Research and Public Health | Incorrect population |
| Durbin 2018 | Social media and adolescents: What are the health risks? | Clinical Advisor | Incorrect study type: commentary/editorial/non-systematic review/theses/book chapter(s) |
| Edler 2022 | The role of personality traits and social support in relations of health-related behaviours and depressive symptoms | BMC Psychiatry | Incorrect population |
| Effertz 2018 | The effect of online gambling on gambling problems and resulting economic health costs in Germany | European Journal of Health Economics | Incorrect population |
| Elavsky 2017 | Who are mobile app users from healthy lifestyle websites? Analysis of patterns of app use and user characteristics | Translational Behavioural Medicine | Incorrect exposure |
| Eleuteri 2017 | Identity, relationships, sexuality, and risky behaviours of adolescents in the context of social media | Journal of Physical Education, Recreation & Dance | Incorrect study type: commentary/editorial/non-systematic review/theses/book chapter(s) |
| Emery 2014 | Wanna know about vaping? Patterns of message exposure, seeking and sharing information about e-cigarettes across media platforms | Tobacco Control | Incorrect population |
| Emory 2019 | Lesbian, gay, bisexual, and transgender (LGBT) view it differently than non-LGBT: exposure to tobacco-related couponing, e-cigarette advertisements, and anti-tobacco messages on social and traditional media | Journal of the Society for Research on Nicotine and Tobacco | Incorrect population |
| Englander 2017 | Social media sex: Exploitation or everlasting love? | Journal of the American Academy of Child and Adolescent Psychiatry | Incorrect study type: conference proceeding or abstract |
| Epstein 2011 | Adolescent computer use and alcohol use: What are the role of quantity and content of computer use? | Addictive Behaviours | Incorrect exposure |
| Epstein-Ngo 2013 | Alcohol use, dating aggression, and mindfulness in high risk youth: Preliminary analyses | Alcoholism: Clinical and Experimental Research | Incorrect study type: conference proceeding or abstract |
| Epstein-Ngo 2014 | Alcohol, drugs, and other factors associated with digital dating violence among high risk urban youth | Alcoholism: Clinical and Experimental Research | Incorrect study type: conference proceeding or abstract |
| Erevik 2017 | Sharing of alcohol-related content on social networking sites: frequency, content, and correlates | Journal of Studies on Alcohol and Drugs | Incorrect population |

| **Author and year** | **Title** | **Publication source** | **Reason for exclusion** |
| --- | --- | --- | --- |
| Escobar-Chaves 2005 | Impact of the media on adolescent sexual attitudes and behaviours | Paediatrics | Incorrect study type: systematic review |
| Eugene 2015 | It's more than just a "sext"- a brief discussion on sexting activity among teens | Journal of Adolescent Health | Incorrect study type: commentary/editorial/non-systematic review/theses/book chapter(s) |
| Evans 2019 | Outcomes of the Adelante community social marketing campaign for Latino youth | Health Education Research | Incorrect exposure |
| Faulkner 2017 | 'Unintended' audiences of alcohol advertising: exposure and drinking behaviours among Australian adolescents | Journal of Substance Use | Incorrect exposure |
| Fielding-Singh 2021 | Tobacco product promotions remain ubiquitous and are associated with use and susceptibility to use among adolescents | Nicotine and Tobacco Research | Incorrect exposure |
| Fife 2019 | STI testing and documentation via a phone application (APP): Experience with the safe app | Sexually Transmitted Infections | Incorrect study type: conference proceeding or abstract |
| Floros 2015 | Adolescent online gambling in Cyprus: associated school performance and psychopathology | Journal of Gambling Studies | Incorrect exposure |
| Folkvord 2016 | Food advertising and eating behaviour in children | Current Opinion in Behavioural Sciences | Incorrect study type: commentary/editorial/non-systematic review/theses/book chapter(s) |
| Ford-Jones 2003 | Impact of media use on children and youth | Paediatrics and Child Health | Incorrect study type: commentary/editorial/non-systematic review/theses/book chapter(s) |
| Forsyth 2013 | The effect of the internet on teen and young adult tobacco use: A literature review | Journal of Paediatric Health Care | Incorrect study type: systematic review |
| Frankis 2017 | Regular STI testing amongst men who have sex with men and use social media is suboptimal - a cross-sectional study | International journal of STD & AIDS | Incorrect exposure |
| Friedman 2011 | GYT (Get Yourself Tested) Campaign: Getting young people talking and tested and sparking a social movement | Sexually Transmitted Infections | Incorrect study type: conference proceeding or abstract |
| Friedman 2013 | Do you GYT? Evaluation of the first two years of the united states' national get yourself tested campaign | Sexually Transmitted Infections | Incorrect study type: conference proceeding or abstract |
| Fung 2018 | Public health implications of image-based social media: a systematic review of Instagram, Pinterest, Tumblr, and Flickr | The Permanente Journal | Incorrect study type: systematic review |

| **Author and year** | **Title** | **Publication source** | **Reason for exclusion** |
| --- | --- | --- | --- |
| Gabrielli 2019 | A new recall of alcohol marketing scale for youth: measurement properties and associations with youth drinking status | Journal Of Studies On Alcohol And Drugs | Duplicate sample |
| Gainsbury 2014 | Are psychology university student gamblers representative of non-university students and general gamblers? a comparative analysis | Journal of Gambling Studies | Incorrect exposure |
| Gansner 2017 | "The internet made me do it": social media and potential for violence in adolescents | Psychiatric Times | Incorrect study type: commentary/editorial/non-systematic review/theses/book chapter(s) |
| Gebremeskel 2014 | Social media use and adolescent risk-taking behaviour | Journal of Adolescent Health | Incorrect study type: conference proceeding or abstract |
| Geisner 2012 | Differences between athletes and non-athletes in risk and health behaviours in graduating high school seniors | Journal of Child & Adolescent Substance Abuse | Incorrect exposure |
| Gentzke 2022 | Tobacco product use and associated factors among middle and high school students - National Youth Tobacco Survey, United States, 2021 | Centers for Disease Control and Prevention Surveillance Summaries | Incorrect exposure |
| Geusens 2016 | The association between social networking sites and alcohol abuse among Belgian adolescents: The role of attitudes and social norms | Journal of Media Psychology: Theories, Methods, and Applications | Duplicate sample |
| Gilliam 2014 | Digital media and sexually transmitted infections | Current Opinion in Obstetrics & Gynaecology | Incorrect study type: commentary/editorial/non-systematic review/theses/book chapter(s) |
| Godinho 2014 | Characteristics associated with media use in early adolescence | Cadernos de Saude Publica | Incorrect exposure |
| Gold 2011 | A systematic examination of the use of online social networking sites for sexual health promotion | BMC Public Health | Incorrect study type: systematic review |
| Golpe 2017 | The relationship between consumption of alcohol and other drugs and problematic Internet use among adolescents | Adicciones | Incorrect exposure |
| Gomez 2020 | Minors and online gambling: prevalence and related variables | Journal of Gambling Studies | Exact duplicate (exact same study in same journal) |
| Gommans 2015 | Frequent electronic media communication with friends is associated with higher adolescent substance use | International Journal of Public Health | Incorrect exposure |
| Gonzalez 2015 | Use and risks of information and communication technologies in the adolescents from 13 to 18 years | Acta Pediatrica Espanola | Potentially relevant non-English language |
| Govindappa 2014 | Internet use and risk-taking behaviours among adolescents | Indian Journal of Paediatrics | Incorrect study type: commentary/editorial/non-systematic review/theses/book chapter(s) |

| **Author and year** | **Title** | **Publication source** | **Reason for exclusion** |
| --- | --- | --- | --- |
| Grant 2014 | Social norms and social networking sites: The role of Facebook in predicting alcohol use among first-year undergraduate students | Alcoholism: Clinical and Experimental Research | Incorrect study type: conference proceeding or abstract |
| Greene 2020 | Social media use among adolescents being evaluated for sexual abuse | Journal of Paediatric and Adolescent Gynaecology | Incorrect study type: conference proceeding or abstract |
| Griffiths 2010 | Adolescent gambling on the internet: A review | International Journal of Adolescent Medicine and Health | Incorrect study type: commentary/editorial/non-systematic review/theses/book chapter(s) |
| Griffiths 2013 | Adolescent gambling via social networking sites: A brief overview | Education & Health | Incorrect study type: commentary/editorial/non-systematic review/theses/book chapter(s) |
| Groom 2021 | The influence of friends on teen vaping: a mixed-methods approach | International Journal of Environmental Research and Public Health | No relevant outcome(s) |
| Guerrero 2019 | Screen time and problem behaviours in children: exploring the mediating role of sleep duration | International Journal of Behavioural Nutrition & Physical Activity | Incorrect population |
| Gulec 2020 | Social media usage and health promoting lifestyle in profile related socio-demographic factors in Turkey | Health Promotion Perspectives | Incorrect population |
| Gumus 2021 | The relationship between adolescents' social media addiction and eating behaviours | Clinical Nutrition | Incorrect study type: conference proceeding or abstract |
| Gupta 2016 | A systematic review of the impact of exposure to internet-based alcohol-related content on young people's alcohol use behaviours | Alcohol and Alcoholism | Incorrect study type: systematic review |
| Gupta 2018 | The association between exposure to social media alcohol marketing and youth alcohol use behaviours in India and Australia | BMC Public Health | Incorrect population |
| Gutierrez 2013 | Internet and cell phone usage associated with risky situations of child sexual exploitation | Salud Mental | No relevant outcome(s) |
| Gutierrez 2015 | The impact of e-cigarette advertisements on e-cigarette initiation among middle and high school students | Drug and Alcohol Dependence | Incorrect study type: conference proceeding or abstract |
| Guy 2012 | Internet pornography and adolescent health: Early findings on effects of online pornography on adolescents show associations with risky behaviour | Medical Journal of Australia | Incorrect study type: commentary/editorial/non-systematic review/theses/book chapter(s) |
| Hadjipanayis 2019 | Social media and children: what is the paediatrician's role? | European Journal of Paediatrics | Incorrect study type: commentary/editorial/non-systematic review/theses/book chapter(s) |

| **Author and year** | **Title** | **Publication source** | **Reason for exclusion** |
| --- | --- | --- | --- |
| Hakim 2018 | Correlates of attempting to quit smoking among adults in Bangladesh | Addictive Behaviours Reports | Incorrect population |
| Hamm 2014 | A systematic review of the use and effectiveness of social media in child health | BMC Paediatrics | Incorrect study type: systematic review |
| Han 2021 | Identifying emerging predictors for adolescent electronic nicotine delivery systems use: A machine learning analysis of the Population Assessment of Tobacco and Health Study | Preventive Medicine | Duplicate sample |
| Hands 2011 | The associations between physical activity, screen time and weight from 6 to 14 yrs: The Raine Study | Journal of Science and Medicine in Sport | Incorrect exposure |
| Hansen 2018 | Electronic cigarette marketing and smoking behaviour in adolescence: A cross-sectional study | ERJ Open Research | Incorrect exposure |
| Hardon 2014 | Ethnographies of youth drug use in Asia | International Journal of Drug Policy | Incorrect study type: commentary/editorial/non-systematic review/theses/book chapter(s) |
| Hassan 2010 | Using technology to improve adolescent healthcare | Current Opinion in Paediatrics | Incorrect study type: commentary/editorial/non-systematic review/theses/book chapter(s) |
| Hendriks 2021 | Causal effects of alcohol-related Facebook posts on drinking behaviour: longitudinal experimental study | Journal of Medical Internet Research | Incorrect population |
| Henry 2009 | Food and beverage brands that market to children and adolescents on the Internet: a content analysis of branded web sites | Journal of Nutrition Education & Behaviour | Incorrect exposure |
| Henzel 2021 | Hooked on virtual social life. Problematic social media use and associations with mental distress and addictive disorders | PloS One | Incorrect exposure |
| Herbert 2017 | Exposure and engagement with tobacco- and e-cigarette related social media | Journal of Adolescent Health | Incorrect exposure |
| Hieftje 2013 | Electronic media-based health interventions promoting behaviour change in youth: A systematic review | JAMA Paediatrics | Incorrect study type: systematic review |
| Hill 2019 | Prevalence and correlates of lifetime and recent HIV testing among men who have sex with men (MSM) who use mobile geo-social networking applications in Greater Tokyo. | PLOS ONE | Incorrect exposure |
| Hingle 2013 | up34 stealth health: youth innovation, mobile technology, online social networking, and informal learning to promote physical activity | Journal of Nutrition Education & Behaviour | Incorrect study type: conference proceeding or abstract |

| **Author and year** | **Title** | **Publication source** | **Reason for exclusion** |
| --- | --- | --- | --- |
| Ho 2001 | Computer usage and its relationship with adolescent lifestyle in Hong Kong | Journal of Adolescent Health | Incorrect exposure |
| Hoare 2020 | Association of child and adolescent mental health with adolescent health behaviours in the UK Millennium Cohort | JAMA Network Open | Incorrect exposure |
| Hoffmann 2019 | High sedentary time in children is not only due to screen media use: A cross-sectional study | BMC Paediatrics | Incorrect exposure |
| Hollingdale 2014 | The effect of online violent video games on levels of aggression | PloS one | Incorrect population |
| Holloway 2014 | Acceptability of smartphone application-based HIV prevention among young men who have sex with men | AIDS and behaviour | Incorrect population |
| Holt 2012 | HIV testing, gay community involvement and internet use: social and behavioural correlates of HIV testing among Australian men who have sex with men | AIDS and behaviour | Incorrect population |
| Hospers 2002 | Chatters on the Internet: a special target group for HIV prevention | AIDS Care - Psychological and Socio-Medical Aspects of AIDS/HIV | Incorrect population |
| Hospers 2005 | A new meeting place: Chatting on the internet, e-dating and sexual risk behaviour among Dutch men who have sex with men | AIDS | Incorrect exposure |
| Howe 2016 | Gotta catch'em all! Pokemon GO and physical activity among young adults: difference in differences study | BMJ (Clinical research ed.) | Incorrect exposure |
| Huang 2012 | The effects of online and offline friendship networks and media use on alcohol and smoking behaviours | Alcoholism: Clinical and Experimental Research | Incorrect study type: conference proceeding or abstract |
| Huang 2014 | The interplay of friendship networks and social networking sites: longitudinal analysis of selection and influence effects on adolescent smoking and alcohol use | American Journal of Public Health | Incorrect study type: commentary/editorial/non-systematic review/theses/book chapter(s) |
| Huang 2017 | Trends and correlates of hookah use among high school students in North Carolina | North Carolina Medical Journal | Incorrect exposure |
| Hur 2013 | Growing up in the web of social networking: Adolescent development and social media | Adolescent Psychiatry | Incorrect study type: commentary/editorial/non-systematic review/theses/book chapter(s) |
| Hutton 2019 | mhealth interventions to reduce alcohol use in young people: a systematic review of the literature | Comprehensive Child and Adolescent Nursing | Incorrect study type: systematic review |

| **Author and year** | **Title** | **Publication source** | **Reason for exclusion** |
| --- | --- | --- | --- |
| Hwang 2009 | Being young and feeling blue in Taiwan: Examining adolescent depressive mood and online and offline activities | New Media & Society | No relevant outcome(s) |
| Ilakkuvan 2019 | Patterns of social media use and their relationship to health risks among young adults | The Journal of Adolescent Health | Incorrect population |
| Ioannidis 2018 | Problematic internet use as an age-related multifaceted problem: Evidence from a two-site survey | Addictive Behaviours | Incorrect population |
| Ishaque 2012 | Frequency of and factors leading to obesity and overweight in school children | Journal of Ayub Medical College, Abbottabad: JAMC | Incorrect exposure |
| Janikian 2015 | Adolescent gambling in seven European countries: Prevalence and related emotional and behavioural problems | Journal of Behavioural Addictions | Incorrect study type: conference proceeding or abstract |
| Jardine 2020 | The Dark Web and cannabis use in the United States: Evidence from a big data research design | International Journal of Drug Policy | Incorrect exposure |
| Jaronko 2019 | Leisure computer usage and perceived body weight, diet, and physical activity | Dissertation Abstracts International: Section B: The Sciences and Engineering | Incorrect study type: commentary/editorial/non-systematic review/theses/book chapter(s) |
| Jenkins 2019 | Youth appeal in recreational marijuana promotions across three social media platforms | Journal of Adolescent Health | Incorrect study type: conference proceeding or abstract |
| Johnson 2014 | Social media use and physical activity: Searching for opportunities to connect adolescents and older adults for health promotion | Journal of Adolescent Health | Incorrect study type: conference proceeding or abstract |
| Jones 2014 | The impact of health education transmitted via social media or text messaging on adolescent and young adult risky sexual behavior: A systematic review of the literature | Sexually Transmitted Diseases | Incorrect study type: systematic review |
| Jones 2016 | Association between young Australian's drinking behaviours and their interactions with alcohol brands on Facebook: results of an online survey | Alcohol and Alcoholism | Incorrect population |
| Jones Jayanetti 2018 | Pizza, burgers, and booze: online marketing and promotion of food and drink to university students | Australian and New Zealand journal of public health | Incorrect population |
| Jonsson 2015 | Online sexual behaviours among Swedish youth: associations to background factors, behaviours, and abuse | European Child and Adolescent Psychiatry | Incorrect exposure |
| Kairouz 2012 | Are online gamblers more at risk than offline gamblers? | Cyberpsychology, Behaviour and Social Networking | Incorrect population |

| **Author and year** | **Title** | **Publication source** | **Reason for exclusion** |
| --- | --- | --- | --- |
| Kandola 2021 | Prospective relationships of adolescents' screen-based sedentary behaviour with depressive symptoms: the Millennium Cohort Study | Psychological Medicine | No relevant outcome(s) |
| Kandola 2022 | Impact on adolescent mental health of replacing screen-use with exercise: a prospective cohort study | Journal of Affective Disorders | No relevant outcome(s) |
| Kaplan 2012 | Social networking and teen drug use: tremendous potential to help and potential to harm? | Psychiatric Times | Incorrect study type: commentary/editorial/non-systematic review/theses/book chapter(s) |
| Kazemi 2017 | Systematic review of surveillance by social media platforms for illicit drug use | Journal of Public Health | Incorrect study type: systematic review |
| Keihner 2009 | Psychosocial, socioeconomic, behavioural, and environmental risk factors for BMI and overweight among 9- to 11-year-old children | Californian Journal of Health Promotion | Incorrect exposure |
| Kemp 2020 | 'Social screens' and 'the mainstream': longitudinal competitors of non-organized physical activity in the transition from childhood to adolescence | The International Journal of Behavioural Nutrition and Physical Activity | Incorrect exposure |
| Kennewell 2022 | The relationships between school children's wellbeing, socio-economic disadvantage, and after-school activities: a cross-sectional study | BMC Paediatrics | Incorrect exposure |
| Kerekes 2021 | Changes in adolescents' psychosocial functioning and well-being as a consequence of long-term covid-19 restrictions | International Journal of Environmental Research and Public Health | No relevant outcome(s) |
| Kerr 2018 | Associations between problem alcohol use and active and passive social media posts | Journal of Adolescent Health | Incorrect study type: conference proceeding or abstract |
| Ketchoo 2013 | Smoking behaviour and associated factors of illicit cigarette consumption in a border province of southern Thailand | Tobacco Control | Incorrect exposure |
| Khajeheian 2018 | Effect of social media on child obesity: Application of structural equation modelling with the Taguchi method | International Journal of Environmental Research and Public Health | No relevant outcome(s) |
| Khundadze 2017 | Impact of internet gambling on mental and psychological health of children of various ages | Georgian Medical News | Incorrect exposure |
| Kim 2015 | International note: Teen users' problematic online behaviour: Using panel data from South Korea | Journal of Adolescence | Incorrect exposure |
| Kim 2017 | A path model of school violence perpetration: introducing online game addiction as a new risk factor | Journal of Interpersonal Violence | Incorrect exposure |

| **Author and year** | **Title** | **Publication source** | **Reason for exclusion** |
| --- | --- | --- | --- |
| Kim 2021 | Parental mental health and children's behaviours and media usage during COVID-19-related school closures | Journal of Korean Medical Science | Incorrect exposure |
| King 2007 | Surf and turf wars online--growing implications of Internet gang violence | Journal of Adolescent Health | Incorrect study type: commentary/editorial/non-systematic review/theses/book chapter(s) |
| Klainman 2015 | Comparison of smoking habits between Jewish and Arabic youth in Israel | European Journal of Preventive Cardiology | Incorrect study type: conference proceeding or abstract |
| Kleppang 2021 | Lifestyle habits and depressive symptoms in Norwegian adolescents: a national cross-sectional study | BMC Public Health | No relevant outcome(s) |
| Ko 2008 | The association between Internet addiction and problematic alcohol use in adolescents: the problem behaviour model | Cyberpsychology & Behaviour: the impact of the Internet, multimedia and virtual reality on behaviour and society | Incorrect exposure |
| Kocturk 2018 | A modern danger for adolescents: from online flirtation to sexual abuse. | Journal of Psychiatry & Neurological Sciences | Incorrect exposure |
| Korogoda 2016 | Developmental neuroscience explaining why adolescents engage in risky behaviours | Journal of Psychosocial Nursing & Mental Health Services | Incorrect study type: commentary/editorial/non-systematic review/theses/book chapter(s) |
| Kranzler 2019 | Youth social media use and health outcomes: #diggingdeeper | Journal of Adolescent Health | Incorrect study type: commentary/editorial/non-systematic review/theses/book chapter(s) |
| Krauss 2017 | Marijuana advertising exposure among current marijuana users in the U.S | Drug And Alcohol Dependence | Incorrect exposure |
| Kristiansen 2022 | Adolescent gambling advertising awareness: A national survey | International Journal of Social Welfare | Incorrect exposure |
| Kurten 2021 | Mothers matter: using regression tree algorithms to predict adolescents' sharing of drunk references on social media | International Journal of Environmental Research and Public Health | No relevant outcome(s) |
| Kuss 2017 | Social networking sites and addiction: Ten lessons learned | International Journal of Environmental Research and Public Health | Incorrect study type: commentary/editorial/non-systematic review/theses/book chapter(s) |
| Kwon 2020 | Factors associated with adolescents' internet use duration by suicidal ideation | International Journal of Environmental Research and Public Health | Incorrect exposure |
| LaBrie 2021 | An examination of the prospective associations between objectively assessed exposure to alcohol-related Instagram content, alcohol-specific cognitions, and first-year college drinking | Addictive Behaviours | Incorrect population |

| **Author and year** | **Title** | **Publication source** | **Reason for exclusion** |
| --- | --- | --- | --- |
| LaBrie 2021 | Prospective relationships between objectively assessed social media use, drinking norms, and alcohol consumption among first-year students | Journal of Studies on Alcohol and Drugs | Incorrect population |
| Lampert 2007 | Use of electronic media in adolescence: results of the German health interview and examination survey for children and adolescents | Bundesgesundheitsblatt - Gesundheitsforschung - Gesundheitsschutz (Federal Health Gazette - Health Research - Health Protection) | Incorrect exposure |
| Leatherdale 2010 | Factors associated with communication-based sedentary behaviours among youth: Are talking on the phone, texting, and instant messaging new sedentary behaviours to be concerned about? | Journal of Adolescent Health | Incorrect exposure |
| Lee 2002 | Internet and displacement effect: Children's media use and activities in Singapore | Journal of Computer-Mediated Communication | Incorrect exposure |
| Lee 2013 | Substance abuse precedes internet addiction | Addictive Behaviours | Incorrect exposure |
| Lee 2015 | Impact of the Internet use in the adolescence on the smoking and drinking in the early adult period: With the panel data | Journal of Behavioural Addictions | Incorrect study type: conference proceeding or abstract |
| Lee 2015 | The association between online health information-seeking behaviours and health behaviours among Hispanics in New York city: a community-based cross-sectional study | Journal of Medical Internet Research | Incorrect population |
| Lee 2017 | Longitudinal study shows that addictive Internet use during adolescence was associated with heavy drinking and smoking cigarettes in early adulthood | Acta Paediatric | Incorrect exposure |
| Lee 2019 | Social networking addiction and depressive symptoms among adolescents in Korea | Journal of Behavioural Addictions | Incorrect study type: conference proceeding or abstract |
| Lee 2020 | Youth and young adult use of pod-based electronic cigarettes from 2015 to 2019: a systematic review | JAMA Paediatrics | Incorrect study type: systematic review |
| Lees 2020 | Screen media activity does not displace other recreational activities among 9–10-year-old youth: a cross-sectional ABCD study R | BMC Public Health | Incorrect population |
| Lehmkuhl 2013 | The new media and their influence on children and adolescents | Kinder- und Jugendpsychiatrie und Psychotherapie (Child and Adolescent Psychiatry and Psychotherapy) | Incorrect study type: commentary/editorial/non-systematic review/theses/book chapter(s) |
| Lerman 2015 | Using the internet to meet people and adolescent sexual risk | Journal of Adolescent Health | Incorrect study type: conference proceeding or abstract |
| Leung 2018 | Exposure to electronic cigarette advertising and intention to use electronic cigarettes in Hong Kong adolescents | Tobacco Induced Diseases | Incorrect study type: conference proceeding or abstract |

| **Author and year** | **Title** | **Publication source** | **Reason for exclusion** |
| --- | --- | --- | --- |
| Leventhal 2018 | New tobacco products with fewer advertising restrictions and consequences for the current generation of youths | JAMA Paediatrics | Incorrect study type: commentary/editorial/non-systematic review/theses/book chapter(s) |
| Lewycka 2018 | Downwards trends in adolescent risk-taking behaviours in New Zealand: Exploring driving forces for change | Journal of Paediatrics and Child Health | Incorrect study type: commentary/editorial/non-systematic review/theses/book chapter(s) |
| Li 2017 | Condom use peer norms and self-efficacy as mediators between community engagement and condom use among Chinese men who have sex with men | BMC Public Health | Incorrect exposure |
| Lipsky 2016 | Behavioural and sociodemographic correlates of overall diet quality over 4 years in a national cohort of U.S. emerging adults | FASEB Journal | Incorrect study type: conference proceeding or abstract |
| Litt 2011 | Adolescent alcohol use: The roles of social norms and social networking sites | Alcoholism: Clinical and Experimental Research | Incorrect study type: conference proceeding or abstract |
| Litt 2018 | #drunktwitter: Examining the relations between alcohol-related Twitter content and alcohol willingness and use among underage young adults | Drug And Alcohol Dependence | Incorrect population |
| Litt 2019 | A longitudinal randomized experimental study examining the impact of social networking site abstainer and drinker content on normative perceptions | Alcoholism: Clinical and Experimental Research | Incorrect study type: conference proceeding or abstract |
| Liu 2017 | A pilot study of Pokémon go and players' physical activity | Games for Health Journal | Incorrect exposure |
| Liu 2021 | The feasibility of using Instagram data to predict exercise identity and physical activity levels: cross-sectional observational study | Journal of Medical Internet Research | Incorrect population |
| Livingstone 2008 | Taking risky opportunities in youthful content creation: Teenagers' use of social networking sites for intimacy, privacy, and self-expression | New Media & Society | Incorrect study type: qualitative |
| Livingstone 2015 | What difference does 'the digital' make to children's experiences of risk? | International Journal of Public Health | Incorrect study type: commentary/editorial/non-systematic review/theses/book chapter(s) |
| Lizandra 2019 | Screen time and moderate-to-vigorous physical activity changes and displacement in adolescence: A prospective cohort study | European Journal of Sport Science | Incorrect exposure |
| Long 2018 | Online and health risk behaviours in high school students: an examination of bullying | Paediatric Nursing | Incorrect exposure |
| Lorenzo-Blanco 2021 | E-cigarette use susceptibility among youth in Mexico: the roles of remote acculturation, parenting behaviours, and internet use frequency | Addictive Behaviours | Incorrect exposure |

| **Author and year** | **Title** | **Publication source** | **Reason for exclusion** |
| --- | --- | --- | --- |
| Lorimer 2016 | Young men who have sex with men's use of social and sexual media and sex-risk associations: cross-sectional, online survey across four countries | Sexually Transmitted Infections | Incorrect population |
| Lou 2012 | Media's contribution to sexual knowledge, attitudes, and behaviours for adolescents and young adults in Three Asian Cities | Journal of Adolescent Health | Incorrect exposure |
| Loukas 2019 | Electronic nicotine delivery systems marketing and initiation among youth and young adults | Paediatrics | Incorrect exposure |
| Love 2019 | How social media influences high school students to commit criminal offenses in South-eastern United States | Dissertation Abstracts International Section A: Humanities and Social Sciences | Incorrect study type: commentary/editorial/non-systematic review/theses/book chapter(s) |
| Lu 2018 | Cross-sectional and temporal associations between cyber dating abuse victimization and mental health and substance use outcomes | Journal Of Adolescence | Incorrect exposure |
| Lukhele 2016 | Multiple sexual partnerships and their correlates among Facebook users in Swaziland: an online cross-sectional study | African journal of AIDS research: AJAR | Incorrect population |
| Luo 2018 | Risk of HIV infection and its factors among men who have sex with men: a geosocial networking application-based survey in Beijing of China, 2017 | Chinese Journal of Preventive Medicine | Incorrect population |
| Luo 2020 | Comparison of HIV infection risk between 15 to 24 year-old student men who have sex with men and non-student men who have sex with men: a cross-sectional study | Chinese Journal of Preventive Medicine | Unable to source full text |
| Lwin 2017 | Media exposure and parental mediation on fast-food consumption among children in metropolitan and suburban Indonesia | Asia Pacific journal of clinical nutrition | Incorrect population |
| Lyons 2017 | Masculinities, alcohol consumption and social networking | Youth drinking cultures in a digital world: Alcohol, social media, and cultures of intoxication | Incorrect study type: commentary/editorial/non-systematic review/theses/book chapter(s) |
| Lyvers 2020 | Alexithymia, impulsivity, disordered social media use, mood, and alcohol use in relation to Facebook self-disclosure | Computers in Human Behaviour | Incorrect population |
| Maas 2019 | Online sexual experiences predict subsequent sexual health and victimization outcomes among female adolescents: a latent class analysis | Journal of Youth and Adolescence | Incorrect exposure |
| Macapagal 2018 | Hookup app use, sexual behaviour, and sexual health among adolescent men who have sex with men in the United States | Journal Of Adolescent Health | Incorrect exposure |
| Macapagal 2019 | Geosocial networking application use, characteristics of app-met sexual partners, and sexual behaviour among sexual and gender minority adolescents assigned male at birth | Journal of Sex Research | Incorrect exposure |

| **Author and year** | **Title** | **Publication source** | **Reason for exclusion** |
| --- | --- | --- | --- |
| MacMillan 2021 | Exploring factors associated with alcohol and/or substance use during the covid-19 pandemic | International Journal of Mental Health and Addiction | Incorrect population |
| Mahase 2019 | Social media can harm when use displaces sleep or exercise or involves bullying, finds study | The BMJ | Incorrect study type: commentary/editorial/non-systematic review/theses/book chapter(s) |
| Malheiros 2021 | Association between physical activity, screen time activities, diet patterns and daytime sleepiness in a sample of Brazilian adolescents | Sleep Medicine | No relevant outcome(s) |
| Marker 2019 | Exploring the myth of the chubby gamer: A meta-analysis on sedentary video gaming and body mass | Social Science and Medicine | Incorrect study type: systematic review |
| Marks 2015 | Friendship network characteristics are associated with physical activity and sedentary behaviour in early adolescence | PloS One | Incorrect exposure |
| Marotta 2018 | Impact and risks of new information technologies in adolescents: Results of a survey conducted on 1534 subjects | Giornale di Neuropsichiatria dell’Età Evolutiva | Potentially relevant non-English language |
| Marques 2018 | Facebook: risks and opportunities in Brazilian and Portuguese youths with different levels of psychosocial adjustment | The Spanish Journal of Psychology | No relevant outcome(s) |
| Masitah 2019 | Social media and adolescent macro nutrition intake | Annals of Nutrition and Metabolism | Incorrect study type: conference proceeding or abstract |
| Mata 2011 | Obesity in children and adolescents: Risks, causes, and therapy from a psychological perspective | Bundesgesundheitsblatt - Gesundheitsforschung - Gesundheitsschutz (Federal Health Gazette - Health Research - Health Protection) | Incorrect study type: commentary/editorial/non-systematic review/theses/book chapter(s) |
| Mayhew 2017 | Youth and sexually explicit internet material: Separating truth from fiction | Journal of the American Academy of Child and Adolescent Psychiatry | Incorrect study type: conference proceeding or abstract |
| McBride 2011 | Risks and benefits of social media for children and adolescents | Journal of Paediatric Nursing | Incorrect study type: commentary/editorial/non-systematic review/theses/book chapter(s) |
| McCarthy 2022 | The influence of unhealthy food and beverage marketing through social media and advergaming on diet-related outcomes in children-a systematic review | Obesity Reviews | Incorrect study type: systematic review |
| McClure 2013 | TV and internet alcohol marketing and underage alcohol use | Alcoholism: Clinical and Experimental Research | Incorrect study type: conference proceeding or abstract |
| McClure 2013 | Alcohol marketing receptivity, marketing-specific cognitions, and underage binge drinking | Alcoholism: Clinical and Experimental Research | Incorrect exposure |

| **Author and year** | **Title** | **Publication source** | **Reason for exclusion** |
| --- | --- | --- | --- |
| McClure 2016 | Internet alcohol marketing and underage alcohol use | Paediatrics | Incorrect exposure |
| McCreanor 2013 | Youth drinking cultures, social networking, and alcohol marketing: Implications for public health | Critical Public Health | Incorrect study type: commentary/editorial/non-systematic review/theses/book chapter(s) |
| McFarlane 2002 | Young adults on the Internet: risk behaviours for sexually transmitted diseases and HIV | Journal of Adolescent Health | Incorrect exposure |
| Meeus 2018 | Managing positive and negative media effects among adolescents: parental mediation matters but not always | Journal of Family Communication | Incorrect exposure |
| Melkevik 2010 | Is spending time in screen-based sedentary behaviours associated with less physical activity: A cross national investigation | International Journal of Behavioural Nutrition and Physical Activity | Incorrect exposure |
| Merkel 2018 | Social media use and physical activity: To share or not to share? | Journal of Physical Education, Recreation & Dance | Incorrect study type: commentary/editorial/non-systematic review/theses/book chapter(s) |
| Miller 2021 | Online peers and offline highs: an examination of online peer groups, social media homophily, and substance use | Journal of Psychoactive Drugs | Incorrect population |
| Mishu 2021 | Predictors of cigarette smoking, smokeless tobacco consumption, and use of both forms in adolescents in South Asia: a secondary analysis of the Global Youth Tobacco Surveys | Nicotine and Tobacco Research | Incorrect exposure |
| Mitchell 2007 | Youth internet users at risk for the most serious online sexual solicitations | American Journal of Preventive Medicine | No relevant outcome(s) |
| Mitchell 2014 | Rural Environments and Community Health (REACH): a randomised controlled trial protocol for an online walking intervention in rural adults | BMC Public Health | Incorrect study type: commentary/editorial/non-systematic review/theses/book chapter(s) |
| Mitchell 2019 | Physical inactivity in childhood from preschool to adolescence | ACSM's Health & Fitness Journal | Incorrect study type: commentary/editorial/non-systematic review/theses/book chapter(s) |
| Moitra 2021 | Screen time is associated with eating habits, sleep patterns, and adiposity measures in adolescents | Obesity Facts | Incorrect study type: conference proceeding or abstract |
| Moreno 2012 | Social networking sites and adolescent health | Paediatric Clinics of North America | Incorrect study type: commentary/editorial/non-systematic review/theses/book chapter(s) |
| Moreno 2014 | Influence of social media on alcohol use in adolescents and young adults | Alcohol Research: Current Reviews | Incorrect study type: commentary/editorial/non-systematic review/theses/book chapter(s) |

| **Author and year** | **Title** | **Publication source** | **Reason for exclusion** |
| --- | --- | --- | --- |
| Moreno 2016 | #Wasted: The intersection of substance use behaviours and social media in adolescents and young adults | Current Opinion in Psychology | Incorrect study type: commentary/editorial/non-systematic review/theses/book chapter(s) |
| Moreno 2019 | Testing young adults' reactions to Facebook cues and their associations with alcohol use | Substance Use & Misuse | Incorrect exposure |
| Morioka 2016 | Association between smoking and problematic internet use among Japanese adolescents: large-scale nationwide epidemiological study | Cyberpsychology, Behaviour and Social Networking | Incorrect exposure |
| Morioka 2017 | The association between alcohol use and problematic internet use: A large-scale nationwide cross-sectional study of adolescents in Japan | Journal of Epidemiology | Incorrect exposure |
| Mu 2015 | Internet use and adolescent binge drinking: Findings from the monitoring the future study | Addictive Behaviours Reports | Incorrect exposure |
| Mucci 2016 | Prevalence of internet addiction: A pilot study in a group of Italian students | European Neuropsychopharmacology | Incorrect study type: conference proceeding or abstract |
| Mukadi 2018 | Sexual behaviour of the school-going youth in the city of Likasi, democratic Republic of Congo | Pan African Medical Journal | Incorrect exposure |
| Munoz-Miralles 2016 | The problematic use of Information and Communication Technologies (ICT) in adolescents by the cross sectional JOITIC study | BMC Paediatrics | Incorrect exposure |
| Murray 2015 | A survey of the practices and perceptions of students in one catholic high school on the use of the internet regarding safety, cyberbullying, and sexting | Dissertation Abstracts International Section A: Humanities and Social Sciences | Incorrect study type: commentary/editorial/non-systematic review/theses/book chapter(s) |
| Namkoong 2017 | Communication, reasoning, and planned behaviours: unveiling the effect of interactive communication in an anti-smoking social media campaign | Health communication | Incorrect exposure |
| Nawi 2021 | Risk and protective factors of drug abuse among adolescents: a systematic review | BMC Public Health | Incorrect study type: systematic review |
| Negriff 2018 | Structural characteristics of the online social networks of maltreated youth and offline sexual risk behaviour | Child Abuse and Neglect | Incorrect comparator group |
| Negriff 2019 | The influence of online-only friends on the substance use of young adults with a history of childhood maltreatment | Substance Use & Misuse | Incorrect exposure |
| Nelson 2019 | Sexually explicit media use among 14-17-year-old sexual minority males in the U.S | Archives of Sexual Behaviour | Incorrect exposure |

| **Author and year** | **Title** | **Publication source** | **Reason for exclusion** |
| --- | --- | --- | --- |
| Ng Fat 2021 | Associations between social media usage and alcohol use among youths and young adults: findings from Understanding Society | Addiction | Exact duplicate (exact same study in same journal) |
| No Authors listed 2003 | Adolescents advertising and tobacco smoking | Medicine Today | Incorrect study type: commentary/editorial/non-systematic review/theses/book chapter(s) |
| No Authors listed 2010 | The "excess" generation | Rivista Italiana di Medicina dell'Adolescenza | Unable to source full text |
| No authors listed 2015 | Social media to combat youth drinking | Australian Nursing & Midwifery Journal | Incorrect study type: commentary/editorial/non-systematic review/theses/book chapter(s) |
| No authors listed 2016 | Social media as a new venue for aggression and bullying | Adolescent Medicine: State of the Art Reviews | Incorrect study type: commentary/editorial/non-systematic review/theses/book chapter(s) |
| No authors listed 2018 | The role of internet addiction on fatigue, sleep disturbances and poor life-style habits among adolescents | European Psychiatry | Incorrect study type: conference proceeding or abstract |
| Noel 2020 | Exposure to digital alcohol marketing and alcohol use: a systematic review | Journal of Studies on Alcohol and Drugs | Incorrect study type: systematic review |
| Noll 2013 | Association of maltreatment with high-risk internet behaviours and offline encounters | Paediatrics | No relevant outcome(s) |
| Norris Turner 2011 | Social media and chlamydia testing by university students: A pilot study | Sexually Transmitted Infections | Incorrect study type: conference proceeding or abstract |
| Nunez-Smith 2010 | Media exposure and tobacco, illicit drugs, and alcohol use among children and adolescents: a systematic review | Substance Abuse | Incorrect study type: systematic review |
| O'Brien 2021 | Relationship between gender, physical activity, screen time, body mass index and wellbeing in Irish children from social disadvantage | Child Care in Practice | Incorrect exposure |
| O'Cathail 2011 | Association of cigarette smoking with drug use and risk taking behaviour in Irish teenagers | Addictive Behaviours | Incorrect exposure |
| Ogunleye 2012 | Prevalence of high screen time in English youth: association with deprivation and physical activity | Journal of Public Health | Incorrect exposure |
| Ojanen 2014 | Investigating online harassment and offline violence among young people in Thailand: methodological approaches, lessons learned | Culture, Health & Sexuality | Incorrect study type: qualitative |

| **Author and year** | **Title** | **Publication source** | **Reason for exclusion** |
| --- | --- | --- | --- |
| O'Keeffe 2011 | Clinical report - The impact of social media on children, adolescents, and families | Pediatrics | Incorrect study type: commentary/editorial/non-systematic review/theses/book chapter(s) |
| O'Keeffe 2011 | The impact of social media on children, adolescents, and families | Paediatrics | Exact duplicate |
| Oksanen 2021 | Social media and access to drugs online: A nationwide study in the United States and Spain among adolescents and young adults | The European Journal of Psychology Applied to Legal Context | Incorrect population |
| Olafsdottir 2014 | Young children's screen activities, sweet drink consumption and anthropometry: Results from a prospective European study | European Journal of Clinical Nutrition | Incorrect exposure |
| Olaleye 2017 | Social-media use and sexual behaviour among in-school adolescents in Ibadan, Nigeria | Sexually Transmitted Infections | Incorrect study type: conference proceeding or abstract |
| O'Sullivan 2012 | Texts from last night: screen time, porn use, sexting, and chat as predictors of sexual intercourse experience among Canadian adolescents | Journal of Adolescent Health | Incorrect study type: conference proceeding or abstract |
| Ouellette 2019 | YouTube and risky behaviours in adolescents: The "choking game" | The American Journal of Emergency Medicine | Incorrect study type: commentary/editorial/non-systematic review/theses/book chapter(s) |
| Pahn 2019 | Impact of short message service (SMS) and social media on sexual intercourse of high school students in Cambodia | Journal of Korean Academy of Community Health Nursing | No relevant outcome(s) |
| Palamar 2020 | Posting, texting, and related social risk behaviour while high | Substance Abuse | Incorrect exposure |
| Palasinski 2013 | Can computer-mediated communication increase adolescents' sexually risky behaviours? | The American Psychologist | Incorrect study type: commentary/editorial/non-systematic review/theses/book chapter(s) |
| Palkar 2019 | Digitizing interventions: An internet-based approach to reach out to the "hidden network of men who have sex with men" in Mumbai, India | Journal of the International AIDS Society | Incorrect study type: conference proceeding or abstract |
| Park 2011 | The relation between screen time and health behaviours in Korean children | Obesity | Incorrect study type: conference proceeding or abstract |
| Park 2013 | A systematic review of social networking sites: Innovative platforms for health research targeting adolescents and young adults | Journal of Nursing Scholarship | Incorrect study type: systematic review |
| Parker 2021 | The use of digital platforms for adults' and adolescents' physical activity during the COVID-19 pandemic (our life at home): Survey study | Journal of Medical Internet Research | Incorrect exposure |

| **Author and year** | **Title** | **Publication source** | **Reason for exclusion** |
| --- | --- | --- | --- |
| Parkes 2013 | Are sexual media exposure, parental restrictions on media use and co-viewing TV and DVDs with parents and friends associated with teenagers' early sexual behaviour? | Journal of Adolescence | Incorrect exposure |
| Patel 2013 | Social media use and HIV risk behaviours in young men who have sex with men of colour in New York city: Implications for outreach and prevention | Journal of General Internal Medicine | Incorrect study type: conference proceeding or abstract |
| Patel 2016 | Social media use and HIV related risk behaviours in young black and Latino gay and bi men and transgender individuals in New York city: implications for online interventions | Journal of Urban Health: bulletin of the New York Academy of Medicine | Incorrect population |
| Patrick 2015 | Demographic and behavioural correlates of six sexting behaviours among Australian secondary school students | Sexual Health | Incorrect exposure |
| Patton 2014 | Social media as a vector for youth violence: A review of the literature | Computers in Human Behaviour | Incorrect study type: commentary/editorial/non-systematic review/theses/book chapter(s) |
| Paulos 2010 | DID video kill the radio star? - Assessing gambling and multimedia use in Luxembourg's high school students | European Psychiatry | Incorrect study type: conference proceeding or abstract |
| Pauwels 2016 | Differential online exposure to extremist content and political violence: Testing the relative strength of social learning and competing perspectives | Terrorism and Political Violence | Incorrect population |
| Pedersen 2004 | Mobile phones, web chat, and sex among Norwegian adolescents | Tidsskr Nor Laegeforen | Potentially relevant non-English language |
| Pedersen 2004 | Mobile phones, web chat, and sex: A study of Norwegian adolescents based on a representative sample | Tidsskrift for den Norske Laegeforening | Exact duplicate |
| Peek 2014 | The selfie in the digital age: from social media to sexting | Psychiatric Times | Incorrect study type: commentary/editorial/non-systematic review/theses/book chapter(s) |
| Peiper 2020 | Differential patterns of e-cigarette and tobacco marketing exposures among youth: Associations with substance use and tobacco prevention strategies | International Journal of Drug Policy | Incorrect exposure |
| Peter 2011 | The influence of sexually explicit internet material on sexual risk behaviour: a comparison of adolescents and adults | Journal of Health Communication | Incorrect exposure |
| Piguet 2015 | What keeps female problematic Internet users busy online? | European Journal of Paediatrics | Incorrect exposure |
| Pocs 2019 | Tobacco reduction on Facebook among 14-35-year-olds | Orv Hetil | Incorrect population |

| **Author and year** | **Title** | **Publication source** | **Reason for exclusion** |
| --- | --- | --- | --- |
| Pokhrel 2021 | Exposure to e-cigarette content on social media and e-cigarette use: An ecological momentary assessment study | Addictive Behaviours Reports | Incorrect population |
| Post 2021 | SARS-CoV-2 wave two surveillance in East Asia and the Pacific: longitudinal trend analysis | Journal of Medical Internet Research | Incorrect exposure |
| Potenza 2011 | Correlates of at-risk/problem internet gambling in adolescents | Journal of the American Academy of Child & Adolescent Psychiatry | Incorrect exposure |
| Prot 2014 | Long-term relations among prosocial-media use, empathy, and prosocial behaviour | Psychological Science | Incorrect exposure |
| Przybylski 2018 | Internet filtering and adolescent exposure to online sexual material | Cyberpsychology, Behaviour and Social Networking | Incorrect exposure |
| Pujazon-Zazik 2010 | Adolescents' self-presentation on a teen dating website: A risk content analysis | Journal of Adolescent Health | Incorrect study type: conference proceeding or abstract |
| Pujazon-Zazik 2010 | To tweet, or not to tweet: gender differences and potential positive and negative health outcomes of adolescents' social internet use | American Journal of Men's Health | Incorrect study type: commentary/editorial/non-systematic review/theses/book chapter(s) |
| Queiroz 2019 | Sexually transmitted infections and factors associated with condom use in dating app users in Brazil | Acta Paulista de Enfermagem | Incorrect population |
| Ra 2018 | Association of digital media use with subsequent symptoms of attention-deficit/hyperactivity disorder among adolescents | Journal of the American Medical Association | No relevant outcome(s) |
| Radanielina Hita 2018 | Parental mediation in the digital era: increasing children's critical thinking may help decrease positive attitudes toward alcohol | Journal of Health Communication | Incorrect population |
| Ragelienė 2021 | The role of peers, siblings and social media for children’s healthy eating socialization: A mixed methods study | Food Quality and Preference | No relevant outcome(s) |
| Raggatt 2019 | Correlates of reduced alcohol consumption among a sample of young Australians | Alcohol and Alcoholism | Incorrect population |
| Rankine 2016 | The association between online risk behaviours and real life sexual behaviours among African American female adolescents | Dissertation Abstracts International: Section B: The Sciences and Engineering | Incorrect study type: commentary/editorial/non-systematic review/theses/book chapter(s) |
| Reid 2014 | Social media use among adolescents: benefits and risks | Adolescent Psychiatry | Exact duplicate |
| Reid 2014 | Social media use among adolescents: Benefits and risks | Adolescent Psychiatry | Incorrect study type: commentary/editorial/non-systematic review/theses/book chapter(s) |

| **Author and year** | **Title** | **Publication source** | **Reason for exclusion** |
| --- | --- | --- | --- |
| Rial 2018 | Minors and problematic internet use: Evidence for better prevention | Computers in Human Behaviour | Incorrect exposure |
| Ribisl 2003 | The potential of the internet as a medium to encourage and discourage youth tobacco use | Tobacco Control | Incorrect study type: commentary/editorial/non-systematic review/theses/book chapter(s) |
| Rice 2010 | Internet use, social networking, and HIV/AIDS risk for homeless adolescents | Journal of Adolescent Health | Incorrect population |
| Rice 2016 | Social media and digital technology use among Indigenous young people in Australia: A literature review | International Journal for Equity in Health | Incorrect study type: systematic review |
| Richards 2015 | Impact of social media on the health of children and young people | Journal of Paediatrics and Child Health | Incorrect study type: systematic review |
| Richter 2020 | The good, the bad and the ugly: the relationship between social media use, subjective health and risk behaviour among children and adolescents | Gesundheitswesen (Bundesverband der Arzte des Offentlichen Gesundheitsdienstes (Germany)) | Potentially relevant non-English language |
| Richter 2021 | The good, the bad and the ugly: the relationship between social media use, subjective health and risk behaviour among children and adolescents | Gesundheitswesen | Exact duplicate (exact same study in same journal) |
| Ricketts 2015 | The effect of Internet related problems on the sexting behaviours of juveniles | American Journal of Criminal Justice | Incorrect exposure |
| Rideout 2002 | Generation Rx.com What are young people really doing online? | Marketing Health Services | Incorrect study type: commentary/editorial/non-systematic review/theses/book chapter(s) |
| Rodenhizer 2019 | The impacts of sexual media exposure on adolescent and emerging adults' dating and sexual violence attitudes and behaviours: a critical review of the literature | Trauma, Violence & Abuse | Incorrect study type: systematic review |
| Rodgers 2020 | A biopsychosocial model of social media use and body image concerns, disordered eating, and muscle-building behaviours among adolescent girls and boys | Journal of Youth and Adolescence | No relevant outcome(s) |
| Rodopman Arman 2015 | Defining social reciprocity deficits in internet addiction: Evaluation of problematic internet user (PIU) adolescents in an university outpatient clinic | European Child and Adolescent Psychiatry | Incorrect study type: conference proceeding or abstract |
| Romer 2017 | Digital media and risks for adolescent substance abuse and problematic gambling | Paediatrics | Incorrect study type: commentary/editorial/non-systematic review/theses/book chapter(s) |
| Romo 2016 | Associations between frequent social media and sexting with sexual risk behaviours in Uganda adolescents | Sexually Transmitted Diseases | Incorrect study type: conference proceeding or abstract |

| **Author and year** | **Title** | **Publication source** | **Reason for exclusion** |
| --- | --- | --- | --- |
| Romo 2016 | Understanding adolescent social media use: Association with sexual risk and parental monitoring factors that can influence protection | Journal of Adolescent Health | Incorrect study type: conference proceeding or abstract |
| Rosen 2014 | Media and technology use predicts ill-being among children, preteens, and teenagers independent of the negative health impacts of exercise and eating habits | Computers in Human Behaviour | Incorrect exposure |
| Rosengren 2020 | Online sex partner seeking and HIV testing frequency among young black sexual minority men | Journal of HIV/AIDS & Social Services | Incorrect population |
| Rosser 2013 | The effects of gay sexually explicit media on the HIV risk behaviour of men who have sex with men | AIDS and Behaviour | Incorrect exposure |
| Rounsefell 2020 | Social media, body image and food choices in healthy young adults: A mixed methods systematic review | Nutrition & Dietetics | Incorrect study type: systematic review |
| Rucker 2015 | Problematic Internet use is associated with substance use in young adolescents | Acta Paediatrica | Incorrect exposure |
| Russell 2022 | Social networking site use and alcohol use behaviors among adolescents: A latent profile analysis | Addictive Behaviors | Incorrect population |
| Ryu 2022 | Smartphone Usage Patterns and Dietary Risk Factors in Adolescents | The Journal of nutrition | Duplicate sample |
| Sabramani 2021 | Bullying and Its Associated Individual, Peer, Family and School Factors: Evidence from Malaysian National Secondary School Students | International journal of environmental research and public health | No relevant outcome(s) |
| Sampasa-Kanyinga 2020 | Sex differences in the relationship between social media use, short sleep duration, and body mass index among adolescents | Sleep Health | No relevant outcome(s) |
| Sande 2021 | Alcohol-related risks for slovene secondary school students on graduation trips: ten years later | Archives of Psychiatry Research | Incorrect exposure |
| Sano 2020 | Relationship between prolonged media usage and lifestyle habits among junior and senior high school students | Japanese Journal of Public Health | Incorrect exposure |
| Santistevan 2017 | Awareness of e-cigarettes and correlation of use among high school students | Dissertation Abstracts International Section A: Humanities and Social Sciences | Incorrect study type: commentary/editorial/non-systematic review/theses/book chapter(s) |
| Sarchiapone 2013 | The use of internet in prevention | European Psychiatry | Incorrect study type: conference proceeding or abstract |

| **Author and year** | **Title** | **Publication source** | **Reason for exclusion** |
| --- | --- | --- | --- |
| Saunders 2016 | A snapshot of the sexual experiences of bisexual black adolescent males over 1 year | Journal of Adolescent Health | Incorrect study type: conference proceeding or abstract |
| Savoia 2021 | Adolescents' exposure to online risks: gender disparities and vulnerabilities related to online behaviours | International Journal of Environmental Research and Public Health | No relevant outcome(s) |
| Savolainen 2020 | Online relationships and social media interaction in youth problem gambling: a four-country study | International Journal of Environmental Research and Public | Incorrect exposure |
| Savolainen 2021 | The role of online group norms and social identity in youth problem gambling | Computers in Human Behaviour | Incorrect population |
| Schafer 2022 | Stigma, social support, and substance use in diverse men who have sex with men and transgender women living with HIV in the US Southeast | Southern Medical Journal | Incorrect population |
| Scott 2016 | The social influence of friends' alcohol-related content posted on social media | Alcoholism: Clinical and Experimental Research | Incorrect study type: conference proceeding or abstract |
| Seidenberg 2017 | A national study of social media, television, radio, and internet usage of adults by sexual orientation and smoking status: implications for campaign design | International Journal of Environmental Research and Public Health | Incorrect population |
| Sela-Shayovitz 2012 | Gangs and the web: Gang members online behavior | Journal of Contemporary Criminal Justice | Incorrect study type: qualitative |
| Self-Brown 2021 | Individual and parental risk factors for sexual exploitation among high-risk youth in Uganda | Journal of Interpersonal Violence | Exact duplicate (exact same study in same journal) |
| Sevcikova 2013 | Predictors of online and offline sexual activities and behaviours among adolescents | Cyberpsychology, Behaviour and Social Networking | Incorrect exposure |
| Ševčíková 2016 | Girls' and boys' experience with teen sexting in early and late adolescence | Journal of Adolescence | Incorrect exposure |
| Sevic 2020 | The relationship between the use of social networking sites and sexually explicit material, the internalization of appearance ideals and body self-surveillance: results from a longitudinal study of male adolescents | Journal of Youth and Adolescence | No relevant outcome(s) |
| Shamu 2020 | Knowledge, attitudes, and practices of young adults towards HIV prevention: an analysis of baseline data from a community-based HIV prevention intervention study in two high HIV burden districts, South Africa | BMC Public Health | Incorrect population |
| Shapiro 2017 | Correlates of tinder use and risky sexual behaviours in young adults | Cyberpsychology, Behaviour and Social Networking | Incorrect population |

| **Author and year** | **Title** | **Publication source** | **Reason for exclusion** |
| --- | --- | --- | --- |
| She 2022 | Profiles of stress and coping associated with mental, behavioural, and internet use problems among adolescents during the COVID-19 pandemic: a stratified random sampling and cluster analysis | Frontiers in Public Health | Incorrect exposure |
| Shi 2011 | Weekend television viewing and video gaming are associated with less adolescent smoking | Journal of Substance Use | Incorrect exposure |
| Shuai 2021 | Influences of digital media use on children and adolescents with ADHD during COVID-19 pandemic | Globalization and Health | No relevant outcome(s) |
| Shukla 2019 | Sugar-sweetened beverages and screen time: partners in crime for adolescent obesity | Journal of Paediatrics | Incorrect study type: commentary/editorial/non-systematic review/theses/book chapter(s) |
| Silva 2016 | Type and quantity of physical activity and screen based activities of students from the 7th to the 12th grades: Characterization and association | Revista Portuguesa de Saude Publica | Incorrect exposure |
| Silva 2018 | Prevalence of overweight and obesity and associated factors in school children and adolescents in a medium-sized Brazilian city | Clinics (Sao Paulo, Brazil) | Incorrect exposure |
| Simon 2018 | Socioeconomic status and adolescent e-cigarette use: The mediating role of e-cigarette advertisement exposure | Preventive Medicine | Incorrect exposure |
| Sina 2022 | Social media and children's and adolescents' diets - a systematic review of the underlying social and physiological mechanisms | Advances in Nutrition | Incorrect study type: systematic review |
| Smith 2016 | Is sexual content in new media linked to sexual risk behaviour in young people? A systematic review and meta-analysis | Sexual Health | Incorrect study type: systematic review |
| Sobowale 2017 | Understanding the role of reward processing and depression in compulsive internet use among V adolescents | Journal of the American Academy of Child and Adolescent Psychiatry | Incorrect study type: conference proceeding or abstract |
| Soneji 2018 | Engagement with online tobacco marketing and associations with tobacco product use among US youth: findings from Wave 1 of the Population Assessment of Tobacco and Health Study | J Adolesc Heal | Duplicate sample |
| Soneji 2019 | Online tobacco marketing among US adolescent sexual, gender, racial, and ethnic minorities | Addictive Behaviours | No relevant outcome(s) |
| Spilkova 2017 | Predictors of excessive use of social media and excessive online gaming in Czech teenagers | Journal of Behavioural Addictions | Incorrect exposure |
| Stevens 2022 | On sex, drugs, and alcohol: A mixed-method analysis of youth posts on social media in the united states | Journal of Children and Media | Incorrect exposure |

| **Author and year** | **Title** | **Publication source** | **Reason for exclusion** |
| --- | --- | --- | --- |
| Stiglic 2019 | Effects of screentime on the health and well-being of children and adolescents: a systematic review of reviews | BMJ open | Incorrect study type: systematic review |
| Stoddard 2012 | Permissive norms and young adults' alcohol and marijuana use: the role of online communities | Journal of Studies on Alcohol and Drugs | Incorrect population |
| Strizek 2020 | Perceived problems with adolescent online gaming: national differences and correlations with substance use | Journal of Behavioural Addictions | Incorrect exposure |
| Stulhofer 2005 | Internet and sexual compulsivity | Socijalna Psihijatrija | Potentially relevant non-English language |
| Sun 2005 | Internet accessibility and usage among urban adolescents in Southern California: Implications for web-based health research | Cyberpsychology and Behaviour | Incorrect exposure |
| Suris 2014 | Problematic internet use and substance use in adolescence | Journal of Adolescent Health | Incorrect study type: conference proceeding or abstract |
| Suwarni 2019 | Determinants of the pornography exposure effects on Junior and Senior High School Adolescence in Sanggau District, West Kalimantan | Indian Journal of Public Health Research and Development | Incorrect exposure |
| Tadena 2020 | The influence of social media affinity on eating attitudes and body dissatisfaction in Philippine adolescents | Child Health Nursing Research | No relevant outcome(s) |
| Tahir 2020 | Does watching violent electronic and social media content lead to increased levels of aggression? A survey among adolescents in an urban slum of metropolitan Karachi | International Journal of Adolescent Medicine and Health | Incorrect exposure |
| Teunissen 2016 | Friends' drinking norms and male adolescents' alcohol consumption: The moderating role of performance-based peer influence susceptibility | Journal Of Adolescence | Incorrect exposure |
| Thammasarn 2020 | Effects of food fit for fun program with social media used on health literacy and obesity prevention behaviours among senior-primary school students, in Nakhon Ratchasima Province Thailand | Indian Journal of Public Health Research and Development | Incorrect exposure |
| Thompson 2005 | Addicted media: Substances on screen | Child and Adolescent Psychiatric Clinics of North America | Incorrect study type: commentary/editorial/non-systematic review/theses/book chapter(s) |
| Thonglua 2015 | The association between internet use and sexual attitudes and behaviours of the secondary school students in Bangkok | Journal of Sexual Medicine | No relevant outcome(s) |
| Thrasher 2016 | Prevalence and correlates of e-cigarette perceptions and trial among early adolescents in Mexico | Journal of Adolescent Health | Incorrect exposure |

| **Author and year** | **Title** | **Publication source** | **Reason for exclusion** |
| --- | --- | --- | --- |
| Tomic 2018 | Associations between Croatian adolescents' use of sexually explicit material and sexual behavior: does parental monitoring play a role? | Archives of Sexual Behavior | Incorrect exposure |
| Törrönen 2020 | How do social media-related attachments and assemblages encourage or reduce drinking among young people? | Journal of Youth Studies | Incorrect study type: qualitative |
| Trangenstein 2021 | Cannabis Marketing and Problematic Cannabis Use Among Adolescents | Journal of studies on alcohol and drugs | Duplicate sample |
| Trangenstein 2022 | Typology of Adolescents Exposed to Non-medical Cannabis Marketing and Associations with Consumption Patterns | Prevention science: the official journal of the Society for Prevention Research | Duplicate sample |
| Tucker 2013 | Cross-lagged associations between substance use-related media exposure and alcohol use during middle school | Alcoholism: Clinical and Experimental Research | Incorrect study type: conference proceeding or abstract |
| Tucker 2013 | Cross-lagged associations between substance use-related media exposure and alcohol use during middle school | Journal of Adolescent Health | Incorrect exposure |
| Twenge 2022 | Specification curve analysis shows that social media use is linked to poor mental health, especially among girls | Acta Psychologica | No relevant outcome(s) |
| Uhls 2017 | Benefits and costs of social media in adolescence | Pediatrics | Incorrect study type: commentary/editorial/non-systematic review/theses/book chapter(s) |
| Unger 2018 | Talking about tobacco on Twitter is associated with tobacco product use | Preventive Medicine | Incorrect population |
| van der Sanden 2021 | Predictors of using social media to purchase drugs in New Zealand: Findings from a large-scale online survey | International Journal of Drug Policy | Incorrect population |
| Van Hulst 2020 | Determinants of new onset cardiometabolic risk among normal weight children | International Journal of Obesity | Incorrect exposure |
| van Oosten 2015 | Exploring associations between exposure to sexy online self-presentations and adolescents' sexual attitudes and behaviour | Journal of Youth and Adolescence | No relevant outcome(s) |
| van Oosten 2017 | Sexy online self-presentation on social network sites and the willingness to engage in sexting: A comparison of gender and age | Journal of Adolescence | No relevant outcome(s) |
| van Oosten 2018 | The importance of adolescents' sexually outgoing self-concept: differential roles of self- and other-generated sexy self-presentations in social media | Cyberpsychology, behaviour and social networking | No relevant outcome(s) |

| **Author and year** | **Title** | **Publication source** | **Reason for exclusion** |
| --- | --- | --- | --- |
| Van Ouytsel 2016 | Cyber dating abuse: Research on young people's motives and the associations of the behaviour in Flanders, Belgium | Journal of the American Academy of Child and Adolescent Psychiatry | Incorrect study type: conference proceeding or abstract |
| Van Ouytsel 2019 | An exploratory study of sexting behaviours among heterosexual and sexual minority early adolescents | Journal of Adolescent Health | Incorrect exposure |
| Vandenbosch 2018 | Explaining the relationship between sexually explicit internet material and casual sex: a two-step mediation model | Archives of Sexual Behavior | Incorrect exposure |
| Vander Wyst 2019 | A social media intervention to improve nutrition knowledge and behaviours of low income, pregnant adolescents, and adult women | PLoS One | Incorrect exposure |
| Vannucci 2020 | Social media use and risky behaviours in adolescents: A meta-analysis | Journal of Adolescence | Incorrect study type: systematic review |
| Vaterlaus 2015 | #Gettinghealthy: The perceived influence of social media on young adult health behaviours. | Computers in Human Behaviour | Incorrect study type: qualitative |
| Vente 2017 | Social media as a vehicle for expression of self-harm and risk-taking behaviour in adolescents | Journal of the American Academy of Child and Adolescent Psychiatry | Incorrect study type: conference proceeding or abstract |
| Vente 2018 | Evaluating high-risk behaviours in adolescents on social media | Journal of Adolescent Health | Incorrect study type: conference proceeding or abstract |
| Villani 2001 | Impact of media on children and adolescents: A 10-year review of the research | Journal of the American Academy of Child and Adolescent Psychiatry | Incorrect study type: commentary/editorial/non-systematic review/theses/book chapter(s) |
| Viner 2019 | Roles of cyberbullying, sleep, and physical activity in mediating the effects of social media use on mental health and wellbeing among young people in England: a secondary analysis of longitudinal data | The Lancet Child and Adolescent Health | No relevant outcome(s) |
| Viner 2020 | Correction to Lancet Child Adolescent Health 2019: Roles of cyberbullying, sleep, and physical activity in mediating the effects of social media use on mental health and wellbeing among young people in England: a secondary analysis of longitudinal data | The Lancet Child and Adolescent Health | Incorrect study type: commentary/editorial/non-systematic review/theses/book chapter(s) |
| Vogel 2020 | Effects of social media on adolescents' willingness and intention to use e-cigarettes: an experimental investigation | Journal of the Society for Research on Nicotine and Tobacco | No relevant outcome(s) |
| Wahyuni 2020 | Determinants of adolescent’s high-risk sexual behaviour in SMK 8 and MegaRezky Health Vocational School Makassar | Enfermería Clínica | Unable to source full text |
| Wahyurin 2019 | Physical activity, screen time, and nutritional status in adolescents in Banyumas | Annals of Tropical Medicine and Public Health | Incorrect exposure |

| **Author and year** | **Title** | **Publication source** | **Reason for exclusion** |
| --- | --- | --- | --- |
| Walther 2014 | Nutrition, lifestyle factors, and mental health in adolescents and young adults living in Austria. | International Journal of Adolescent Medicine And Health | Incorrect exposure |
| Wang 2012 | Adolescent bullying involvement and psychosocial aspects of family and school life: A cross-sectional study from Guangdong province in China | PLoS ONE | Incorrect exposure |
| Watchirs Smith 2013 | Do new media affect adolescent sexual attitudes and behaviours? A systematic review | Sexually Transmitted Infections | Incorrect study type: conference proceeding or abstract |
| Welsh 2013 | The sugar-sweetened beverage wars: public health and the role of the beverage industry | Current Opinion in Endocrinology, Diabetes, and Obesity | Incorrect study type: commentary/editorial/non-systematic review/theses/book chapter(s) |
| Werneck 2018 | Social, behavioural, and biological correlates of cardiorespiratory fitness according to sex, nutritional status, and maturity status among adolescents. A cross-sectional study | Sao Paulo Medical Journal | Incorrect exposure |
| Westgate 2014 | "I will take a shot for every 'like' I get on this status": posting alcohol-related Facebook content is linked to drinking outcomes | Journal of Studies on Alcohol and Drugs | Incorrect population |
| White 2015 | Adolescents' and young adults' online risk taking: the role of gist and verbatim representations | Risk Analysis: an official publication of the Society for Risk Analysis | Incorrect exposure |
| Whitehill 2015 | Emerging adults' use of alcohol and social networking sites during a large street festival: A real-time interview study | Substance Abuse Treatment, Prevention, and Policy | Incorrect population |
| Whitehill 2020 | Exposure to cannabis marketing in social and traditional media and past-year use among adolescents in states with legal retail cannabis | Journal of Adolescent Health | Exact duplicate (exact same study in same journal) |
| Whiteley 2011 | African American adolescents and new media: Associations with HIV/STI risk behaviour and psychosocial variables | Ethnicity and Disease | Incorrect exposure |
| Wickel 2013 | Variables associated with active and inactive behaviour during the after-school period | Paediatric Exercise Science | Incorrect exposure |
| Willoughby 2022 | Social media, marijuana, and sex: an exploratory study of adolescents' intentions to use and college students' use of marijuana | Journal of Sex Research | Incorrect population |
| Winetrobe 2014 | Associations of unprotected anal intercourse with Grindr-met partners among Grindr-using young men who have sex with men in Los Angeles | AIDS Care | Incorrect population |
| Winther 2014 | Leisure time computer use and adolescent bone health: Findings from the tromso study-fit futures | Osteoporosis International | Incorrect study type: conference proceeding or abstract |

| **Author and year** | **Title** | **Publication source** | **Reason for exclusion** |
| --- | --- | --- | --- |
| Winther 2015 | Leisure time computer use and adolescent bone health-findings from the Tromso Study, Fit Futures: A cross-sectional study | BMJ Open | Incorrect exposure |
| Xu 2018 | The effect of using geosocial networking apps on the HIV incidence rate among men who have sex with men: eighteen-month prospective cohort study in Shenyang, China | Journal of Medical Internet Research | Incorrect exposure |
| Yau 2014 | Relationships between problematic Internet use and problem-gambling severity: Findings from a high-school survey | Addictive Behaviours | Incorrect exposure |
| Ybarra 2006 | Internet use among Ugandan adolescents: implications for HIV intervention | PLoS Medicine | No relevant outcome(s) |
| Ybarra 2008 | Linkages between internet and other media violence with seriously violent behaviour by youth | Paediatrics | Incorrect exposure |
| Ybarra 2014 | Sexual media exposure, sexual behaviour, and sexual violence victimization in adolescence | Clinical Paediatrics | Incorrect exposure |
| Ybarra 2015 | Can clans protect adolescent players of massively multiplayer online games from violent behaviours? | International Journal of Public Health | Incorrect comparator group |
| Ybarra 2016 | A national study of lesbian, gay, bisexual (LGB), and non-LGB youth sexual behaviour online and in-person | Archives of Sexual Behaviour | No relevant outcome(s) |
| Yonker 2015 | "Friending" teens: Systematic review of social media in adolescent and young adult health care | Journal of Medical Internet Research | Incorrect study type: systematic review |
| Yoo 2014 | Associations between overuse of the internet and mental health in adolescents | Nursing & Health Sciences | Incorrect exposure |
| Young 2011 | Online social networking technologies, HIV knowledge, and sexual risk and testing behaviours among homeless youth | AIDS and Behaviour | Incorrect population |
| Young 2013 | Social networking and diffusion of risks and interventions among youth | Sexually Transmitted Infections | Incorrect study type: conference proceeding or abstract |
| Young 2018 | HIV prevention and sex behaviours as organizing mechanisms in a Facebook group affiliation network among young black men who have sex with men | AIDS & Behaviour | Incorrect comparator group |
| Yu 2017 | Predictors and the distal outcome of general Internet use: The identification of children's developmental trajectories | The British Journal of Developmental Psychology | Incorrect exposure |

| **Author and year** | **Title** | **Publication source** | **Reason for exclusion** |
| --- | --- | --- | --- |
| Yusriani 2020 | Education through WhatsApp media in changing of smoking behaviour among senior high school students | National Public Health Journal | Incorrect exposure |
| Zhan 2019 | Electronic cigarette usage patterns: a case study combining survey and social media data | Journal of the American Medical Informatics Association | Incorrect population |
| Zheng 2021 | How does online e-cigarette advertisement promote youth’s e-cigarettes use? The mediating roles of social norm and risk perceptions | Health Communication | Exact duplicate (exact same study in same journal) |
| Zheng 2021 | How Does Online e-cigarette Advertisement Promote Youth's e-cigarettes Use? The Mediating Roles of Social Norm and Risk Perceptions | Health communication | Duplicate sample |
| Zheng 2021 | Social media and E-cigarette use among US youth: Longitudinal evidence on the role of online advertisement exposure and risk perception | Addictive Behaviours | Duplicate sample |
| Zhou 2014 | Internet use and its impact on engagement in leisure activities in China | PLoS ONE | Incorrect exposure |
| Zhu 2017 | Pro-smoking information scanning using social media predicts young adults' smoking behaviour | Computers in Human Behaviour | Incorrect population |
| Zonfrillo 2014 | NekNominate: a deadly, social media-based drinking dare | Clinical Pediatrics | Incorrect study type: commentary/editorial/non-systematic review/theses/book chapter(s) |

### Appendix-13. Risk of bias domain and overall grade for included datapoints and studies

##### Table A. Risk of bias domain and overall grades for included cross-sectional and cohort datapoints (n=334), and overall study risk of bias grade (n=122), assessed using adapted Newcastle Ottawa Scale

| Author and year | RoB assessment tool | Selection | Exposure | Comparability | Outcome | Overall datapoint RoB | Overall study RoB |
| --- | --- | --- | --- | --- | --- | --- | --- |
| Anastario 2020 | NOS: cross-sectional studies | Moderate | Moderate | Moderate | Moderate | Moderate | Moderate |
| Anastario 2020 | NOS: cross-sectional studies | Moderate | Moderate | Moderate | Moderate | Moderate |  |
| Baker 2016 | NOS: cross-sectional studies | High | High | High | Moderate | High | High |
| Baker 2016 | NOS: cross-sectional studies | High | High | High | Moderate | High |  |
| Baker 2016 | NOS: cross-sectional studies | High | High | High | Moderate | High |  |
| Baldwin 2018 | NOS: cross-sectional studies | Moderate | Moderate | Low | Moderate | Low | Low |
| Baldwin 2018 | NOS: cross-sectional studies | Moderate | Moderate | Low | Moderate | Low |  |
| Baldwin 2018 | NOS: cross-sectional studies | Moderate | Moderate | Low | Moderate | Low |  |
| Baldwin 2018 | NOS: cross-sectional studies | Moderate | Moderate | Low | Moderate | Low |  |
| Baldwin 2018 | NOS: cross-sectional studies | Moderate | Moderate | Low | Moderate | Low |  |
| Baldwin 2018 | NOS: cross-sectional studies | Moderate | Moderate | Low | Moderate | Low |  |
| Baldwin 2018 | NOS: cross-sectional studies | Moderate | Moderate | Low | Moderate | Low |  |
| Ball 2020 | NOS: cross-sectional studies | Low | Moderate | Low | Moderate | Low | Low |
| Ball 2020 | NOS: cross-sectional studies | Low | Moderate | Low | Moderate | Low |  |
| Baru 2020 | NOS: cross-sectional studies | High | Moderate | Moderate | Moderate | High | High |
| Baumgartner 2012 | NOS: cohort studies | Low | Moderate | Moderate | Moderate | Moderate | Moderate |
| Bayraktar 2007 | NOS: cross-sectional studies | High | High | High | Moderate | High | High |
| Beebe 2004 | NOS: cross-sectional studies | Moderate | Moderate | High | Moderate | High | High |
| Beebe 2004 | NOS: cross-sectional studies | Moderate | Moderate | High | Moderate | High |  |
| Beebe 2004 | NOS: cross-sectional studies | Moderate | Moderate | High | Moderate | High |  |
| Beebe 2004 | NOS: cross-sectional studies | Moderate | Moderate | High | Moderate | High |  |
| Beebe 2004 | NOS: cross-sectional studies | Moderate | Moderate | High | Moderate | High |  |
| Beebe 2004 | NOS: cross-sectional studies | Moderate | Moderate | High | Moderate | High |  |
| Beebe 2004 | NOS: cross-sectional studies | Moderate | Moderate | High | Moderate | High |  |
| Beebe 2004 | NOS: cross-sectional studies | Moderate | Moderate | High | Moderate | High |  |
| Beebe 2004 | NOS: cross-sectional studies | Moderate | Moderate | High | Moderate | High |  |
| Beebe 2004 | NOS: cross-sectional studies | Moderate | Moderate | High | Moderate | High |  |
| Beebe 2004 | NOS: cross-sectional studies | Moderate | Moderate | High | Moderate | High |  |
| Beebe 2004 | NOS: cross-sectional studies | Moderate | Moderate | High | Moderate | High |  |
| Boers 2020 | NOS: cohort studies | Low | Moderate | Low | Moderate | Low | Low |
| Booker 2015 | NOS: cross-sectional studies | Low | Moderate | High | Moderate | High | High |
| Boniel-Nissim 2022 | NOS: cross-sectional studies | Moderate | Low | Low | Low | Low | Low |
| Boniel-Nissim 2022 | NOS: cross-sectional studies | Moderate | Low | Low | Low | Low |  |
| Boniel-Nissim 2022 | NOS: cross-sectional studies | Moderate | Low | Low | Low | Low |  |
| Boniel-Nissim 2022 | NOS: cross-sectional studies | Moderate | Low | Low | Low | Low |  |

| Author and year | RoB assessment tool | Selection | Exposure | Comparability | Outcome | Overall datapoint RoB | Overall study RoB |
| --- | --- | --- | --- | --- | --- | --- | --- |
| Brunborg 2019 | NOS: cohort studies | Low | Moderate | Low | Moderate | Low | Low |
| Brunborg 2019 | NOS: cohort studies | Low | Moderate | Low | Low | Low |  |
| Brunborg 2019 | NOS: cross-sectional studies | Low | Moderate | High | Moderate | High |  |
| Brunborg 2019 | NOS: cross-sectional studies | Low | Moderate | High | Low | High |  |
| Brunborg 2022 | NOS: cohort studies | Low | Moderate | Moderate | Moderate | Moderate | Moderate |
| Camenga 2018 | NOS: cohort studies | Moderate | Moderate | Moderate | High | High | High |
| Canale 2016 | NOS: cross-sectional studies | Moderate | High | Low | Low | High | Low |
| Canale 2016 | NOS: cross-sectional studies | Moderate | High | Low | Low | High |  |
| Canale 2016 | NOS: cross-sectional studies | Moderate | Moderate | Low | Low | Low |  |
| Casaló 2022 | NOS: cross-sectional studies | Moderate | Moderate | Low | Moderate | Low | Low |
| Casaló 2022 | NOS: cross-sectional studies | Moderate | Moderate | Low | Moderate | Low |  |
| Casaló 2022 | NOS: cross-sectional studies | Moderate | Moderate | Low | Moderate | Low |  |
| Casaló 2022 | NOS: cross-sectional studies | Moderate | Moderate | Low | Moderate | Low |  |
| Cavazos-Rehg 2014 | NOS: cross-sectional studies | Moderate | Moderate | High | Moderate | High | High |
| Chang 2016 | NOS: cohort studies | Low | Moderate | High | High | High | High |
| Chang 2016 | NOS: cohort studies | Low | Moderate | High | High | High |  |
| Chapin 2018 | NOS: cross-sectional studies | Moderate | Moderate | High | Moderate | High | High |
| Chapin 2018 | NOS: cross-sectional studies | Moderate | Moderate | High | Moderate | High |  |
| Chau 2022 | NOS: cross-sectional studies | Low | Moderate | Moderate | Moderate | Moderate | Moderate |
| Chau 2022 | NOS: cross-sectional studies | Low | Moderate | Moderate | Moderate | Moderate |  |
| Chau 2022 | NOS: cross-sectional studies | Low | Moderate | Moderate | Moderate | Moderate |  |
| Chau 2022 | NOS: cross-sectional studies | Low | Moderate | Moderate | Moderate | Moderate |  |
| Chau 2022 | NOS: cross-sectional studies | Low | Moderate | Moderate | Moderate | Moderate |  |
| Chen 2019 | NOS: cross-sectional studies | High | Moderate | High | Moderate | High | High |
| Chen 2019 | NOS: cross-sectional studies | High | Moderate | High | Moderate | High |  |
| Coyne 2013 | NOS: cross-sectional studies | High | Moderate | High | Low | High | High |
| Coyne 2018 | NOS: cohort studies | Low | Moderate | High | Moderate | High | High |
| Coyne 2018 | NOS: cohort studies | Low | Moderate | High | Moderate | High |  |
| Critchlow 2019 | NOS: cross-sectional studies | Low | Moderate | Low | Low | Low | Low |
| Critchlow 2019 | NOS: cross-sectional studies | Low | Moderate | Low | Low | Low |  |
| Critchlow 2019 | NOS: cross-sectional studies | Low | Moderate | Low | Low | Low |  |
| da Costa 2021 | NOS: cross-sectional studies | Moderate | Moderate | Low | Low | Low | Low |
| da Costa 2021 | NOS: cross-sectional studies | Moderate | Moderate | Low | Low | Low |  |
| da Costa 2021 | NOS: cross-sectional studies | Moderate | Moderate | Low | Low | Low |  |
| Dai 2022 | NOS: cross-sectional studies | Low | Low | Moderate | Low | Low | Low |
| Dai 2022 | NOS: cross-sectional studies | Low | Low | Moderate | Low | Low |  |
| Davis 2019 | NOS: cohort studies | Moderate | Moderate | Moderate | Moderate | Moderate | Moderate |
| Dawson 2019 | NOS: cross-sectional studies | High | Moderate | High | Moderate | High | High |
| Dawson 2019 | NOS: cross-sectional studies | High | Low | High | Moderate | High |  |
| Dawson 2019 | NOS: cross-sectional studies | High | Low | High | Moderate | High |  |
| de Bruijn 2016 | NOS: cross-sectional studies | Low | Moderate | Moderate | Moderate | Moderate | Moderate |
| de Bruijn 2016 | NOS: cross-sectional studies | Low | Moderate | Moderate | Moderate | Moderate |  |

| Author and year | RoB assessment tool | Selection | Exposure | Comparability | Outcome | Overall datapoint RoB | Overall study RoB |
| --- | --- | --- | --- | --- | --- | --- | --- |
| De Looze 2019 | NOS: cross-sectional studies | Moderate | Moderate | Moderate | Moderate | Moderate | Moderate |
| De Looze 2019 | NOS: cross-sectional studies | Moderate | Moderate | Low | Moderate | Low |  |
| De Looze 2019 | NOS: cross-sectional studies | Moderate | Moderate | Moderate | Moderate | Moderate |  |
| Doornwaard 2014 | NOS: cross-sectional studies | High | Low | High | Low | High | High |
| Doornwaard 2015 | NOS: cross-sectional studies | Moderate | Moderate | High | Moderate | High | Moderate |
| Doornwaard 2015 | NOS: cross-sectional studies | Moderate | Moderate | High | Moderate | High |  |
| Doornwaard 2015 | NOS: cross-sectional studies | Moderate | Moderate | Moderate | Moderate | Moderate |  |
| Doornwaard 2015 | NOS: cross-sectional studies | Moderate | Moderate | Moderate | Moderate | Moderate |  |
| Elton-Marshall 2016 | NOS: cross-sectional studies | Moderate | Moderate | High | Moderate | High | High |
| Elton-Marshall 2016 | NOS: cross-sectional studies | Moderate | Moderate | High | Low | High |  |
| Erreygers 2017 | NOS: cross-sectional studies | Moderate | Moderate | High | Moderate | High | High |
| Floros 2013 | NOS: cross-sectional studies | Moderate | High | Moderate | Moderate | High | High |
| Floros 2013 | NOS: cross-sectional studies | Moderate | High | High | Low | High |  |
| Froyland 2020 | NOS: cross-sectional studies | Low | Moderate | Low | Moderate | Low | Low |
| Froyland 2020 | NOS: cross-sectional studies | Low | Moderate | Low | Moderate | Low |  |
| Froyland 2020 | NOS: cross-sectional studies | Low | Moderate | High | Moderate | High |  |
| Froyland 2020 | NOS: cross-sectional studies | Low | Moderate | High | Moderate | High |  |
| Froyland 2020 | NOS: cross-sectional studies | Low | Moderate | High | Moderate | High |  |
| Froyland 2020 | NOS: cross-sectional studies | Low | Moderate | High | Moderate | High |  |
| Froyland 2020 | NOS: cross-sectional studies | Low | Moderate | High | Moderate | High |  |
| Froyland 2020 | NOS: cross-sectional studies | Low | Moderate | High | Moderate | High |  |
| Gascoyne 2021 | NOS: cross-sectional studies | Moderate | Moderate | Low | Moderate | Low | Low |
| Gascoyne 2021 | NOS: cross-sectional studies | Moderate | Moderate | Low | Moderate | Low |  |
| Gazendam 2020 | NOS: cross-sectional studies | Moderate | Moderate | Low | Moderate | Low | Low |
| Gazendam 2020 | NOS: cross-sectional studies | Moderate | Moderate | Low | Moderate | Low |  |
| Geber 2021 | NOS: cohort studies | Low | Moderate | Moderate | Moderate | Moderate | Moderate |
| Geusens 2017 | NOS: cohort studies | Moderate | Moderate | Moderate | High | High | High |
| Geusens 2017 | NOS: cross-sectional studies | High | Moderate | High | Moderate | High |  |
| Geusens 2017 | NOS: cross-sectional studies | Moderate | Moderate | Moderate | Low | Moderate | Moderate |
| Geusens 2017 | NOS: cross-sectional studies | Moderate | Moderate | Moderate | Low | Moderate |  |
| Geusens 2019 | NOS: cross-sectional studies | Moderate | Moderate | Low | Moderate | Low | Low |
| Geusens 2019 | NOS: cross-sectional studies | Moderate | Moderate | Low | Moderate | Low |  |
| Gomez 2019 | NOS: cross-sectional studies | Moderate | High | High | High | High | High |
| Gordon 2011 | NOS: cross-sectional studies | High | Moderate | High | Moderate | High | High |
| Gordon 2011 | NOS: cross-sectional studies | High | Moderate | High | Moderate | High |  |
| Gregg 2018 | NOS: cross-sectional studies | Moderate | Moderate | Moderate | Low | Moderate | Moderate |
| Gunnlaugsson 2020 | NOS: cross-sectional studies | Moderate | Low | Moderate | Low | Low | Low |
| Gunnlaugsson 2020 | NOS: cross-sectional studies | Moderate | Low | Moderate | Low | Low |  |
| Gunnlaugsson 2020 | NOS: cross-sectional studies | Moderate | Low | Moderate | Low | Low |  |
| Hamilton 2020 | NOS: cross-sectional studies | Low | Moderate | High | Moderate | High | High |
| Hayer 2018 | NOS: cohort studies | Low | Moderate | Moderate | Moderate | Moderate | Moderate |
| Hayer 2018 | NOS: cohort studies | Low | Moderate | Moderate | Moderate | Moderate |  |
| Holtz 2011 | NOS: cross-sectional studies | Moderate | Moderate | Moderate | Low | Moderate | Moderate |

| Author and year | RoB assessment tool | Selection | Exposure | Comparability | Outcome | Overall datapoint RoB | Overall study RoB |
| --- | --- | --- | --- | --- | --- | --- | --- |
| Hryhorczuk 2019 | NOS: cross-sectional studies | Moderate | Moderate | Moderate | Moderate | Moderate | Moderate |
| Hryhorczuk 2019 | NOS: cross-sectional studies | Moderate | Moderate | Moderate | Moderate | Moderate |  |
| Hryhorczuk 2019 | NOS: cross-sectional studies | Moderate | Moderate | Moderate | Moderate | Moderate |  |
| Hryhorczuk 2019 | NOS: cross-sectional studies | Moderate | Moderate | Moderate | Moderate | Moderate |  |
| Hryhorczuk 2019 | NOS: cross-sectional studies | Moderate | Moderate | Moderate | Moderate | Moderate |  |
| Hryhorczuk 2019 | NOS: cross-sectional studies | Moderate | Moderate | Moderate | Moderate | Moderate |  |
| Hrywna 2020 | NOS: cross-sectional studies | Low | Moderate | Moderate | Moderate | Moderate | Moderate |
| Hrywna 2020 | NOS: cross-sectional studies | Low | Moderate | Moderate | Moderate | Moderate |  |
| Huang 2012 | NOS: cross-sectional studies | Moderate | Moderate | Low | Moderate | Low | Low |
| Huang 2014 | NOS: cohort studies | Low | Moderate | Low | Moderate | Low | Low |
| Huang 2014 | NOS: cohort studies | Low | Moderate | Low | Moderate | Low |  |
| Huang 2014 | NOS: cohort studies | Low | Moderate | Low | Moderate | Low |  |
| Huang 2014 | NOS: cohort studies | Low | Moderate | Low | Moderate | Low |  |
| Jeong 2022 | NOS: cross-sectional studies | Moderate | Moderate | High | Low | High | High |
| Jiang 2018 | NOS: cross-sectional studies | High | Low | Low | Low | High | High |
| Kaufman 2014 | NOS: cross-sectional studies | Moderate | Moderate | Low | Low | Low | Low |
| Kaufman 2014 | NOS: cross-sectional studies | Moderate | Moderate | Moderate | Low | Moderate |  |
| Kaufman 2014 | NOS: cross-sectional studies | Moderate | Moderate | Low | Low | Low |  |
| Kaufman 2014 | NOS: cross-sectional studies | Moderate | Moderate | Moderate | Low | Moderate |  |
| Kaufman 2014 | NOS: cross-sectional studies | Moderate | Moderate | Low | Moderate | Low |  |
| Kaufman 2014 | NOS: cross-sectional studies | Moderate | Moderate | Moderate | Moderate | Moderate |  |
| Kaufman 2014 | NOS: cross-sectional studies | Moderate | Moderate | Low | Moderate | Low |  |
| Kaufman 2014 | NOS: cross-sectional studies | Moderate | Moderate | Moderate | Moderate | Moderate |  |
| Kaur 2020 | NOS: cross-sectional studies | Low | Moderate | Moderate | Moderate | Low | Low |
| Kaur 2020 | NOS: cross-sectional studies | Low | Moderate | Moderate | Moderate | Low |  |
| Kaur 2020 | NOS: cross-sectional studies | Low | Moderate | Moderate | Moderate | Low |  |
| Kaur 2020 | NOS: cross-sectional studies | Low | Moderate | Moderate | Moderate | Low |  |
| Kaur 2020 | NOS: cross-sectional studies | Low | Moderate | Moderate | Moderate | Low |  |
| Kaur 2020 | NOS: cross-sectional studies | Low | Moderate | Moderate | Moderate | Low |  |
| Kelleghan 2020 | NOS: cohort studies | Low | Moderate | Moderate | Moderate | Moderate | Moderate |
| Kelleghan 2020 | NOS: cohort studies | Low | Moderate | Moderate | Moderate | Moderate |  |
| Kelleghan 2020 | NOS: cohort studies | Low | Moderate | Moderate | Moderate | Moderate |  |
| Kelleghan 2020 | NOS: cohort studies | Low | Moderate | Moderate | Moderate | Moderate |  |
| Kelleghan 2020 | NOS: cohort studies | Low | Moderate | Moderate | Moderate | Moderate |  |
| Kelleghan 2020 | NOS: cohort studies | Low | Moderate | Moderate | Moderate | Moderate |  |
| King 2014 | NOS: cross-sectional studies | High | Moderate | High | Low | High | High |
| Ko 2009 | NOS: cross-sectional studies | Low | Moderate | Moderate | Moderate | Moderate | Moderate |
| Ko 2009 | NOS: cross-sectional studies | Low | Moderate | Moderate | Moderate | Moderate |  |
| Ko 2009 | NOS: cross-sectional studies | Low | Moderate | Moderate | Moderate | Moderate |  |

| Author and year | RoB assessment tool | Selection | Exposure | Comparability | Outcome | Overall datapoint RoB | Overall study RoB |
| --- | --- | --- | --- | --- | --- | --- | --- |
| Kontostoli 2022 | NOS: cross-sectional studies | Moderate | Moderate | Low | Low | Low | Low |
| Kontostoli 2022 | NOS: cross-sectional studies | Moderate | Moderate | Low | Low | Low |  |
| Kontostoli 2022 | NOS: cross-sectional studies | Moderate | Moderate | Low | Moderate | Low |  |
| Kontostoli 2022 | NOS: cross-sectional studies | Moderate | Moderate | Low | Low | Low |  |
| Kontostoli 2022 | NOS: cross-sectional studies | Moderate | Moderate | Low | Low | Low |  |
| Kontostoli 2022 | NOS: cross-sectional studies | Moderate | Moderate | Low | Moderate | Low |  |
| Koutamanis 2015 | NOS: cross-sectional studies | Moderate | Moderate | High | Moderate | High | High |
| Kwon 2022 | NOS: cross-sectional studies | Moderate | Moderate | Low | Moderate | Low | Low |
| Kwon 2022 | NOS: cross-sectional studies | Moderate | Moderate | Low | Moderate | Low |  |
| Kwon 2022 | NOS: cross-sectional studies | Moderate | Moderate | Low | Moderate | Low |  |
| Kwon 2022 | NOS: cross-sectional studies | Moderate | Moderate | Low | Moderate | Low |  |
| Landry 2013 | NOS: cross-sectional studies | High | Moderate | Moderate | Moderate | High | High |
| Landry 2013 | NOS: cross-sectional studies | High | Moderate | High | Moderate | High |  |
| Larm 2017 | NOS: cross-sectional studies | Moderate | Moderate | High | Low | High | High |
| Larm 2017 | NOS: cross-sectional studies | Moderate | Moderate | High | Low | High |  |
| Larm 2019 | NOS: cross-sectional studies | High | Moderate | Moderate | Low | High | High |
| Larm 2019 | NOS: cross-sectional studies | High | Moderate | Moderate | Low | High |  |
| Lee 2015 | NOS: cross-sectional studies | High | Moderate | Moderate | Moderate | High | High |
| Lee 2019 | NOS: cohort studies | Low | Moderate | Moderate | High | High | High |
| Lee 2019 | NOS: cohort studies | Low | Moderate | Moderate | Moderate | Moderate |  |
| Lee 2021 | NOS: cross-sectional studies | High | Low | Moderate | Low | High | High |
| Lee 2021 | NOS: cross-sectional studies | High | Low | Moderate | Low | High |  |
| Lee 2021 | NOS: cross-sectional studies | Moderate | Moderate | Moderate | Moderate | Moderate | Moderate |
| Lee 2021 | NOS: cross-sectional studies | Moderate | Moderate | Moderate | Moderate | Moderate |  |
| Lin 2012 | NOS: cross-sectional studies | Low | Moderate | High | Moderate | High | High |
| Lin 2012 | NOS: cross-sectional studies | Low | Moderate | High | Moderate | High |  |
| Lipsky 2017 | NOS: cohort studies | Low | Moderate | Low | Low | Low | Low |
| Lipsky 2017 | NOS: cohort studies | Low | Moderate | Low | Low | Low |  |
| Lipsky 2017 | NOS: cohort studies | Low | Moderate | Low | Low | Low |  |
| Longobardi 2021 | NOS: cross-sectional studies | Moderate | High | High | Low | High | High |
| McClure 2020 | NOS: cross-sectional studies | High | Moderate | Low | Moderate | High | High |
| Merrill 2019 | NOS: cross-sectional studies | High | Moderate | Moderate | Moderate | High | High |
| Michael 2016 | NOS: cross-sectional studies | Moderate | High | High | High | High | High |
| Moitra 2022 | NOS: cross-sectional studies | Moderate | Moderate | Low | Moderate | Low | Low |
| Moitra 2022 | NOS: cross-sectional studies | Moderate | Moderate | Low | Low | Low |  |
| Mojica 2014 | NOS: cross-sectional studies | High | Moderate | Moderate | Moderate | High | High |
| Mojica 2014 | NOS: cross-sectional studies | High | Moderate | Moderate | Moderate | High |  |
| Mojica 2014 | NOS: cross-sectional studies | High | Moderate | Moderate | Moderate | High |  |
| Molla-Esparza 2021 | NOS: cross-sectional studies | Low | Moderate | Moderate | Moderate | Moderate | Moderate |
| Molla-Esparza 2021 | NOS: cross-sectional studies | Low | Moderate | Moderate | Moderate | Moderate |  |
| Molla-Esparza 2021 | NOS: cross-sectional studies | Low | Moderate | Moderate | Moderate | Moderate |  |

| Author and year | RoB assessment tool | Selection | Exposure | Comparability | Outcome | Overall datapoint RoB | Overall study RoB |
| --- | --- | --- | --- | --- | --- | --- | --- |
| Nesi 2017 | NOS: cohort studies | Moderate | Moderate | Moderate | Moderate | Moderate | Moderate |
| Nesi 2017 | NOS: cohort studies | Moderate | Moderate | Moderate | Moderate | Moderate |  |
| Nesi 2017 | NOS: cohort studies | Moderate | Moderate | Moderate | Moderate | Moderate |  |
| Nesi 2017 | NOS: cohort studies | Moderate | Moderate | Moderate | Moderate | Moderate |  |
| Nesi 2017 | NOS: cohort studies | Moderate | Moderate | Moderate | Moderate | Moderate |  |
| Nesi 2017 | NOS: cohort studies | Moderate | Moderate | Moderate | Moderate | Moderate |  |
| Nesi 2019 | NOS: cohort studies | Low | Moderate | High | Moderate | High | High |
| Nesi 2019 | NOS: cohort studies | Low | Moderate | High | Moderate | High |  |
| Nesi 2019 | NOS: cohort studies | Low | Moderate | High | Moderate | High |  |
| Nesi 2019 | NOS: cohort studies | Low | Low | High | Moderate | High |  |
| Nesi 2019 | NOS: cohort studies | Low | Low | High | Moderate | High |  |
| Nesi 2019 | NOS: cross-sectional studies | Moderate | Moderate | High | Moderate | High |  |
| Nesi 2019 | NOS: cross-sectional studies | Moderate | Moderate | High | Moderate | High |  |
| Nesi 2019 | NOS: cross-sectional studies | Moderate | Moderate | High | Moderate | High |  |
| Nesi 2019 | NOS: cross-sectional studies | Moderate | Low | High | Moderate | High |  |
| Nesi 2019 | NOS: cross-sectional studies | Moderate | Low | High | Moderate | High |  |
| Ng Fat 2021 | NOS: cohort studies | Low | Moderate | Low | Moderate | Low | Low |
| Ng Fat 2021 | NOS: cohort studies | Low | Moderate | Low | Moderate | Low |  |
| Ng Fat 2021 | NOS: cohort studies | Low | Moderate | Low | Moderate | Low |  |
| Ng Fat 2021 | NOS: cross-sectional studies | Low | Moderate | Low | Moderate | Low |  |
| Ng Fat 2021 | NOS: cross-sectional studies | Low | Moderate | Low | Moderate | Low |  |
| Ng Fat 2021 | NOS: cross-sectional studies | Low | Moderate | Low | Moderate | Low |  |
| Ng Fat 2021 | NOS: cross-sectional studies | Low | Moderate | Low | Moderate | Low |  |
| Ng Fat 2021 | NOS: cross-sectional studies | Low | Moderate | Low | Moderate | Low |  |
| Ohannessian 2009 | NOS: cross-sectional studies | High | Moderate | High | Moderate | High | High |
| Ohannessian 2009 | NOS: cross-sectional studies | High | Moderate | High | Moderate | High |  |
| Pegg 2018 | NOS: cross-sectional studies | High | Moderate | Moderate | Moderate | High | High |
| Pegg 2018 | NOS: cross-sectional studies | High | Moderate | Moderate | Moderate | High |  |
| Pérez 2022 | NOS: cohort studies | Low | Moderate | Low | Moderate | Low | Low |
| Prince 2021 | NOS: cross-sectional studies | Low | Moderate | High | Moderate | High | High |
| Qutteina 2022 | NOS: cross-sectional studies | Low | Moderate | Moderate | Low | Moderate | Moderate |
| Qutteina 2022 | NOS: cross-sectional studies | Low | Moderate | Moderate | Low | Moderate |  |
| Qutteina 2022 | NOS: cross-sectional studies | Low | Moderate | Moderate | Low | Moderate |  |
| Qutteina 2022 | NOS: cross-sectional studies | Low | Moderate | Moderate | Low | Moderate |  |
| Qutteina 2022 | NOS: cross-sectional studies | Low | Moderate | Moderate | Low | Moderate |  |
| Riehm 2021 | NOS: cross-sectional studies | Moderate | Moderate | High | Moderate | High | High |
| Roditis 2016 | NOS: cross-sectional studies | Low | Moderate | Moderate | Moderate | Moderate | Moderate |
| Roditis 2016 | NOS: cross-sectional studies | Low | Moderate | Moderate | Moderate | Moderate |  |

| Author and year | RoB assessment tool | Selection | Exposure | Comparability | Outcome | Overall datapoint RoB | Overall study RoB |
| --- | --- | --- | --- | --- | --- | --- | --- |
| Romo 2017 | NOS: cross-sectional studies | High | Low | Moderate | Low | High | High |
| Romo 2017 | NOS: cross-sectional studies | High | Low | Moderate | Low | High |  |
| Romo 2017 | NOS: cross-sectional studies | High | Low | Moderate | Low | High |  |
| Romo 2017 | NOS: cross-sectional studies | High | Low | Moderate | Low | High |  |
| Romo 2017 | NOS: cross-sectional studies | High | Low | Moderate | Low | High |  |
| Romo 2017 | NOS: cross-sectional studies | High | Low | Moderate | Low | High |  |
| Romo 2017 | NOS: cross-sectional studies | High | Low | Moderate | Low | High |  |
| Romo 2017 | NOS: cross-sectional studies | High | Low | Moderate | Low | High |  |
| Rutter 2021 | NOS: cross-sectional studies | Moderate | Moderate | Moderate | Moderate | Moderate | Moderate |
| Sampasa-Kanyinga 2015 | NOS: cross-sectional studies | Moderate | Moderate | Low | Moderate | Low | Low |
| Sampasa-Kanyinga 2015 | NOS: cross-sectional studies | Moderate | Moderate | Low | Moderate | Low |  |
| Sampasa-Kanyinga 2015 | NOS: cross-sectional studies | Moderate | Moderate | Low | Moderate | Low |  |
| Sampasa-Kanyinga 2015 | NOS: cross-sectional studies | Moderate | Moderate | High | Moderate | High | High |
| Sampasa-Kanyinga 2015 | NOS: cross-sectional studies | Moderate | Moderate | High | Moderate | High |  |
| Sampasa-Kanyinga 2016 | NOS: cross-sectional studies | Low | Moderate | Low | Moderate | Low | Low |
| Sampasa-Kanyinga 2016 | NOS: cross-sectional studies | Low | Moderate | Low | Moderate | Low |  |
| Sampasa-Kanyinga 2016 | NOS: cross-sectional studies | Low | Moderate | Low | Moderate | Low |  |
| Sampasa-Kanyinga 2016 | NOS: cross-sectional studies | Low | Moderate | Low | Moderate | Low |  |
| Sampasa-Kanyinga 2016 | NOS: cross-sectional studies | Low | Moderate | Low | Moderate | Low |  |
| Sampasa-Kanyinga 2016 | NOS: cross-sectional studies | Low | Moderate | Low | Moderate | Low |  |
| Sampasa-Kanyinga 2016 | NOS: cross-sectional studies | Moderate | Moderate | Low | Moderate | Low | Low |
| Sampasa-Kanyinga 2016 | NOS: cross-sectional studies | Moderate | Moderate | Low | Moderate | Low |  |
| Sampasa-Kanyinga 2018 | NOS: cross-sectional studies | Moderate | Moderate | Moderate | Moderate | Moderate | Moderate |
| Sandercock 2016 | NOS: cross-sectional studies | Moderate | Moderate | Low | Moderate | Low | Low |
| Sandercock 2016 | NOS: cross-sectional studies | Moderate | Moderate | Low | Moderate | Low |  |
| Sandercock 2016 | NOS: cross-sectional studies | Moderate | Moderate | Low | Moderate | Low |  |
| Sandercock 2016 | NOS: cross-sectional studies | Moderate | Moderate | Low | Moderate | Low |  |
| Sandercock 2016 | NOS: cross-sectional studies | Moderate | Moderate | Low | Low | Low |  |

| Author and year | RoB assessment tool | Selection | Exposure | Comparability | Outcome | Overall datapoint RoB | Overall study RoB |
| --- | --- | --- | --- | --- | --- | --- | --- |
| Savolainen 2020 | NOS: cross-sectional studies | High | Low | Moderate | Low | High | High |
| Savolainen 2020 | NOS: cross-sectional studies | High | Low | Moderate | Low | High |  |
| Savolainen 2020 | NOS: cross-sectional studies | High | Low | Moderate | Low | High |  |
| Savolainen 2020 | NOS: cross-sectional studies | High | Low | Moderate | Low | High |  |
| Savolainen 2020 | NOS: cross-sectional studies | High | Low | Moderate | Low | High |  |
| Savolainen 2020 | NOS: cross-sectional studies | High | Low | Moderate | Low | High |  |
| Savolainen 2020 | NOS: cross-sectional studies | High | Low | Moderate | Low | High |  |
| Savolainen 2020 | NOS: cross-sectional studies | High | Low | Moderate | Low | High |  |
| Savolainen 2020 | NOS: cross-sectional studies | High | Low | Moderate | Low | High |  |
| Savolainen 2020 | NOS: cross-sectional studies | High | Low | Moderate | Low | High |  |
| Savolainen 2020 | NOS: cross-sectional studies | High | Low | Moderate | Low | High |  |
| Savolainen 2020 | NOS: cross-sectional studies | High | Low | Moderate | Low | High |  |
| Savolainen 2020 | NOS: cross-sectional studies | High | Low | Moderate | Low | High |  |
| Savolainen 2020 | NOS: cross-sectional studies | High | Low | Moderate | Low | High |  |
| Savolainen 2020 | NOS: cross-sectional studies | High | Low | Moderate | Low | High |  |
| Savolainen 2020 | NOS: cross-sectional studies | High | Low | Moderate | Low | High |  |
| Savolainen 2020 | NOS: cross-sectional studies | High | Low | Moderate | Low | High |  |
| Savolainen 2020 | NOS: cross-sectional studies | High | Low | Moderate | Low | High |  |
| Savolainen 2020 | NOS: cross-sectional studies | High | Low | Moderate | Low | High |  |
| Savolainen 2020 | NOS: cross-sectional studies | High | Low | Moderate | Low | High |  |
| Self-Brown 2018 | NOS: cross-sectional studies | Low | Low | High | Low | High | High |
| Self-Brown 2018 | NOS: cross-sectional studies | Low | Low | High | Low | High |  |
| Shan 2022 | NOS: cohort studies | Low | Moderate | Low | Moderate | Low | Low |
| Shan 2022 | NOS: cohort studies | Low | Moderate | Low | Moderate | Low |  |
| Sharma 2021 | NOS: cross-sectional studies | Low | Moderate | High | High | High | High |
| Shimoga 2019 | NOS: cross-sectional studies | Moderate | Moderate | Low | Moderate | Low | Low |
| Smout 2021 | NOS: cohort studies | Low | Moderate | Moderate | Moderate | Moderate | Moderate |
| Smout 2021 | NOS: cohort studies | Low | Moderate | Moderate | Moderate | Moderate |  |
| Soneji 2018 | NOS: cohort studies | Low | Moderate | Low | Moderate | Low | Low |
| Soneji 2018 | NOS: cohort studies | Low | Moderate | Low | Moderate | Low |  |
| Soneji 2018 | NOS: cohort studies | Low | Moderate | Low | Moderate | Low |  |
| Soneji 2018 | NOS: cohort studies | Low | Moderate | Low | Moderate | Low |  |
| Stevens 2017 | NOS: cross-sectional studies | Moderate | Moderate | Low | Moderate | Low | Low |
| Stevens 2017 | NOS: cross-sectional studies | Moderate | Moderate | Low | Moderate | Low |  |
| Suwanwong 2021 | NOS: cross-sectional studies | High | Moderate | High | Moderate | High | High |
| Suwanwong 2021 | NOS: cross-sectional studies | High | Moderate | High | Moderate | High |  |
| Svensson 2020 | NOS: cross-sectional studies | Low | Moderate | Moderate | Moderate | Moderate | Moderate |
| Svensson 2020 | NOS: cross-sectional studies | Low | Moderate | Moderate | Moderate | Moderate |  |
| Svensson 2020 | NOS: cross-sectional studies | Low | Moderate | Moderate | Moderate | Moderate |  |
| Tao 2022 | NOS: cross-sectional studies | Low | Moderate | High | Low | High | High |
| Tao 2022 | NOS: cross-sectional studies | Low | Moderate | High | Low | High |  |
| Trangenstein 2019 | NOS: cross-sectional studies | High | Moderate | High | Moderate | High | High |
| Trangenstein 2019 | NOS: cross-sectional studies | High | Moderate | High | Moderate | High |  |

| Author and year | RoB assessment tool | Selection | Exposure | Comparability | Outcome | Overall datapoint RoB | Overall study RoB |
| --- | --- | --- | --- | --- | --- | --- | --- |
| Tsitsika 2009 | NOS: cross-sectional studies | Moderate | High | High | Moderate | High | High |
| Tsitsika 2011 | NOS: cross-sectional studies | Low | Moderate | High | Moderate | High | High |
| Vandenbosch 2016 | NOS: cross-sectional studies | Moderate | Moderate | High | Moderate | High | High |
| Vannucci 2019 | NOS: cohort studies | Low | Moderate | Low | Low | Low | Low |
| Vannucci 2019 | NOS: cross-sectional studies | Low | Moderate | High | Low | High |  |
| Vannucci 2019 | NOS: cross-sectional studies | Low | Moderate | High | Low | High |  |
| Vannucci 2019 | NOS: cross-sectional studies | Low | Moderate | High | Low | High |  |
| Vazquez-Nava 2020 | NOS: cross-sectional studies | Moderate | Low | High | Low | High | High |
| Vente 2020 | NOS: cross-sectional studies | Moderate | Moderate | High | Moderate | High | High |
| Vente 2020 | NOS: cross-sectional studies | Moderate | Moderate | High | Moderate | High |  |
| Wana 2019 | NOS: cross-sectional studies | Low | Moderate | Moderate | Moderate | Moderate | Moderate |
| Ward 2022 | NOS: cross-sectional studies | Moderate | Moderate | Moderate | Moderate | Moderate | Moderate |
| Ward 2022 | NOS: cross-sectional studies | Moderate | Moderate | Moderate | Moderate | Moderate |  |
| Ward 2022 | NOS: cross-sectional studies | Moderate | Moderate | Moderate | Moderate | Moderate |  |
| Whitehill 2020 | NOS: cross-sectional studies | High | Moderate | High | Moderate | High | High |
| Whitehill 2020 | NOS: cross-sectional studies | High | Moderate | High | Moderate | High |  |
| Whitehill 2020 | NOS: cross-sectional studies | High | Moderate | High | Moderate | High |  |
| Whitehill 2020 | NOS: cross-sectional studies | High | Moderate | High | Moderate | High |  |
| Widman 2014 | NOS: cross-sectional studies | High | Moderate | High | Moderate | High | High |
| Widman 2014 | NOS: cross-sectional studies | High | Moderate | High | Moderate | High |  |
| Worku 2022 | NOS: cross-sectional studies | Low | Moderate | Moderate | Low | Moderate | Moderate |
| Wulff 2021 | NOS: cross-sectional studies | Moderate | Moderate | Moderate | Low | Moderate | Moderate |
| Yao 2022 | NOS: cross-sectional studies | Moderate | Moderate | Moderate | Moderate | Moderate | Moderate |
| Yao 2022 | NOS: cross-sectional studies | Moderate | Moderate | Moderate | Moderate | Moderate |  |

**Legend:** Abbreviations: NOS = Adapted Newcastle Ottawa Scale and RoB = Risk of bias.

##### Table B. Risk of bias domain and overall grades for included randomised control trial datapoints (n=4), and overall study risk of bias grade (n=4), assessed using Cochrane Risk of Bias 2 tool

| Author and year | RoB assessment tool | Randomisation | Adherence | Missingness | Measurement | Reporting | Overall datapoint RoB | Overall study RoB |
| --- | --- | --- | --- | --- | --- | --- | --- | --- |
| Coates 2019 | RoB-2 | Low | Some concerns | Low | Low | Some concerns | Some concerns | Some concerns |
| De Jans 2021 | RoB-2 | Low | Low | Low | Low | Low | Low | Low |
| Folkvord 2020 | RoB-2 | Low | Low | Low | Low | Low | Low | Low |
| Ngqangashe 2021 | RoB-2 | Some concerns | Low | Low | Low | Low | Some concerns | Some concerns |

**Legend:** Abbreviations: RoB = Risk of bias and RoB-2 = Cochrane Risk of Bias 2 tool.

### Appendix-14. Social media measures reported in included studies

Table A below outlines the social media exposure measures (n=253) used to assess social media use across included studies. Within included studies, many social media exposure measures were reported and were incorporated in our exploration of how SM use is measured in relation to adolescent health-risk behaviours, therefore the number of datapoints reported differ from those included in the review synthesis. Please also note this table records the exposure measures in their original form as reported in included studies, in some instances the exposure measures may have been transformed/specific pairwise comparisons selected for inclusion in meta-analyses/SWiM as per the decisions rules outlined in Appendix-7.

##### Table A. Social media measures reported in included studies

| Exposure | Author and year | Exposure definition | Exposure ascertainment | Validated/  objectively recorded | Exposure measure  type | Exposure time period | SM platform | SM platform | SM category (active/  passive use) | SM content |
| --- | --- | --- | --- | --- | --- | --- | --- | --- | --- | --- |
| Time Spent on SM | Brunborg 2019 | Change in hrs of SM use per day (Δ = T2 – T1) | Self-report 2-item measure: 1-Frequency of SM use in the past 6/12 months. Participant asked to report on active SM use: reading, writing, watching pictures, making comments, or appointments on SM etc, and not merely the time logged on. Responses: every day to not at all. Responses recoded into the average number of days per month spent on SM.  2-How many hrs/day usually spent on SM. Responses: < 1 hr to >15 hrs/day in hourly increments.  Product of frequency (average days/month) and quantity (average hrs/day) divided by 30 to reflect the average number of hrs spent on SM per day. |  | Continuous | T1: past 12 months T2: past 6 months | Examples: Facebook, Snapchat, WhatsApp, Twitter, Instagram, and Kik | Mixed platforms | SNS  (active use) | NA |
| Time Spent on SM | Brunborg 2019 | Average number of hrs spent on SM per day | Self-report 2-item measure: 1-Frequency of SM use in the past 6/12 months. Respondents asked to report on active SM use: reading, writing, watching pictures, making comments, or appointments on SM etc, and not merely the time logged on. Responses: every day to not at all. Responses recoded into the average number of days per month spent on SM.  2-How many hrs per day usually spent on SM. Responses: < 1 hr to >15 hrs/day in hourly increments.  Product of frequency (average days/month) and quantity (average hrs/day) divided by 30 to reflect the average number of hrs spent on SM per day. |  | Continuous | Past 12 months | Examples: Facebook, Snapchat, WhatsApp, Twitter, Instagram, and Kik | Mixed platforms | SNS  (active use) | NA |

| Exposure | Author and year | Exposure definition | Exposure ascertainment | Validated/  objectively recorded | Exposure measure  type | Exposure time period | SM platform | SM platform | SM category  (active/  passive use) | SM content |
| --- | --- | --- | --- | --- | --- | --- | --- | --- | --- | --- |
| Time Spent on SM | Brunborg 2022 | Average number of hrs spent on SM per day | Self-report 2-item measure:  1-Frequency of active use of SM (e.g., Facebook, Snapchat, and Instagram) in the past 30 days  Responses: not at all to 5-days a week  2-How many hrs per day spent actively using SM  Responses: less than 1 hr to 10 hr or more  Product of frequency (days per month) and quantity (average hrs/day) divided by 30 to reflect average number of hrs spent on SM per day |  | Continuous | Past month | Examples: Facebook, Snapchat, and Instagram | Mixed platforms | General SM  (active use) | NA |
| Time Spent on SM | Boers 2020 | Time spent on SM per day | Self-report 1-item measure: 1-How much time spent on Facebook, Twitter or other SNS per day. Responses: 0–30 min, 30 min – 1 hr 30 min, 1 hr 30 min – 2 hrs 30 min, and ≥3 hrs 30 min/day. |  | Continuous | Current | Specified: Facebook, Twitter, and other SNS | Mixed platforms | SNS  (unclear) | NA |
| Time Spent on SM | Booker 2015 | Time spent chatting on social websites on a normal school day | Self-report 1-item measure: 1-How many hrs spent chatting or interacting with friends through a social web site like Bebo, Facebook, and Myspace on a normal school day. Responses: (1) none to (5) ≥7 hrs/day.  3 category variable constructed: <1 hr, 1 to 3 hrs, and ≥4 hrs/day. |  | Continuous | Current | Examples: Bebo, Facebook, and Myspace | Mixed platforms | SNS  (active use) | NA |
| Time Spent on SM | Casaló 2022 | Time spent on SNS per day | Self-report 1-item measure:  1-Time spent on SNS for fun (and not to do homework/work) per day  Responses: no time devoted, 30 mins or less, around an hr, 2-3 hrs, 4 or more hrs/day |  | Continuous | Current | NR | NR | SNS  (unclear) | NA |
| Time Spent on SM | Chau 2022 | Time spent on discussion forums and chatting online during a weekday | Self-report 1-item measure:  1-How many hours spent with discussion forums and chatting online during a weekday  Responses: (1) <2 hrs (2) 2-4 hrs (3) 5 or more hrs/day |  | Categorical | Current | NR | NR | Blogs & Forums  (active use) | NA |
| Time Spent on SM | Chau 2022 | Time spent on discussion forums and chatting online during a weekend day | Self-report 1-item measure:  1-How many hours spent with discussion forums and chatting online during a weekend day  Responses: (1) <2 hrs (2) 2-4 hrs (3) 5 or more hrs/day |  | Categorical | Current | NR | NR | Blogs & Forums  (active use) | NA |

| Exposure | Author and year | Exposure definition | Exposure ascertainment | Validated/  objectively recorded | Exposure measure  type | Exposure time period | SM platform | SM platform | SM category  (active/  passive use) | SM content |
| --- | --- | --- | --- | --- | --- | --- | --- | --- | --- | --- |
| Time Spent on SM | Chen 2019 | Time spent on SM on a regular weekday and weekend day | Self-report 8-item measure: 8 questions about time spent on Facebook, Instagram, and Snapchat on a regular weekday (Monday–Friday) and a weekend day (Saturday and Sunday).  Responses: (0) from 0 hrs to (10) the platform is opened throughout the day continuously.  Weighted score of average daily time for each SM platform computed: (time weekday × 5 + time weekend day × 2)/7.  SM use computed by the average score of the 3 platforms. |  | Continuous | Current | Specified: Facebook, Instagram, and Snapchat | Mixed platforms | SNS  (unclear) | NA |
| Time Spent on SM | Coyne 2013 | Time spent on SNS on a typical day | Self-report 1-item measure:  1-How much time spent on SNS in a typical day. Responses: 9-point scale from (1) none to (9) >8 hrs/day. |  | Continuous | Current | NR | NR | SNS  (unclear) | NA |
| Time Spent on SM | Coyne 2018 | Time spent on SNS on a typical day | Self-report 1-item measure, assessed at 6 timepoints over 6 years: 1-How much time spent on SNS, like Facebook, on a typical day. Responses: (1) none to (9) >8 hrs/day.  3 category variable constructed: peak users (low SM use that increases quickly after a few years and then returns to baseline), moderate users (steady SM use over time), and increasers (low SM use that increases gradually and ends high at the end of the study). |  | Categorical | Current | Examples: Facebook and Instagram | Mixed platforms | SNS  (unclear) | NA |
| Time Spent on SM | da Costa 2021 | Time spent on SM on a typical weekday and weekend day | Self-report 2-item measure:  1-Time spent engaged on SM on a typical weekday  2-Time spent engaged on SM on a typical weekend day  Daily time on SM estimated by weighting answers ([volume on weekdays x 5 + volume on weekend x 2]/7) hrs/day  4 category variable constructed: <2 hrs, ≥2 hrs, <4 hrs, and ≥4 hrs/day |  | Continuous | Current | NR | NR | General SM  (unclear) | NA |
| Time Spent on SM | Doornwaard 2015 | Time spent on SNS (most frequently used platform) per day | Self-report 1-item measure: 1-How much time actively spent each day on most used SNS.  Responses: not an SNS member, <15 mins, 15–30 mins, 30–60 mins, 1–2 hrs, 3–4 hrs, and >4 hrs/day. |  | Ordinal | Current | NR | NR | SNS  (active use) | NA |

| Exposure | Author and year | Exposure definition | Exposure ascertainment | Validated/  objectively recorded | Exposure measure  type | Exposure time period | SM platform | SM platform | SM category  (active/  passive use) | SM content |
| --- | --- | --- | --- | --- | --- | --- | --- | --- | --- | --- |
| Time Spent on SM | Froyland 2020 | Time spent on SM per day | Self-report 1-item measure:  1-How much time spent daily on SM (e.g., Facebook, Instagram, etc)  Responses: non, <30 min, 30 min-1 hr, 1-2 hrs, 2-3 hrs, >3 hrs. |  | Continuous | Current | Examples: Facebook and Instagram | Mixed platforms | General SM  (unclear) | NA |
| Time Spent on SM | Gazendam 2020 | Time spent on SM per day | Self-report 1-item measure:  1-How many hrs a day, in free time, spent using electronic devices such as computers, tablets (like iPad) or smartphone for other purposes (e.g., tweeting, Facebook, chatting)  Responses: none at all, about 30 mins, about 1 hr, about 2 hrs, about ≥3 hrs/day.  Variable dichotomised: <3hrs/ ≥3 hrs/day |  | Binary | Current | Examples: Facebook, chatting, and Twitter | Mixed platforms | General SM  (unclear) | NA |
| Time Spent on SM | Hamilton 2020 | Time spent on SNS per day | Daily diary self-report:  1-How much time spent on SNS (e.g., TikTok, Snapchat, Instagram) per day  Responses: none, <30 min, 0.5–1 hr, 1-2 hrs, 2-4 hrs, 4-6 hrs, and >6 hrs/day. |  | Continuous | Current | Examples: TikTok, Snapchat, and Instagram | Mixed platforms | SNS  (unclear) | NA |
| Time Spent on SM | Kaur 2020 | Time spent on SNS per day | Self-report 1-item measure: 1-How many hrs on an average day spent on social networking web sites like Facebook, Twitter, Instagram, etc. Responses: none, <1 hr, 1−2 hrs, 3−4 hrs, 5−6 hrs, 7−8 hrs, and ≥9 hrs/day.  4 category variable constructed: 0 hrs, <1 hr, 1-2 hrs and ≥3 hrs/day. |  | Binary | Current | Examples: Facebook, Twitter, and Instagram | Mixed platforms | SNS  (unclear) | NA |
| Time Spent on SM | Kontostoli 2020 | Time spent browsing and updating SNS on a weekday | Time use diary:  Participants recorded their behaviour in 10-min timeslots from 4 to 4 am the next day. For each 10-min timeslot, participants indicated their main activity, selecting from a pre-specified list of 44 activities, nested within 12 categories for both weekday and weekend day.  Participants were categorised according to whether they did (user) or did not (non-user) report time browsing and updating social networking sites. |  | Binary | Current | Examples: Facebook, Twitter, and Snapchat | Mixed platforms | SNS  (active and passive use) | NA |

| Exposure | Author and year | Exposure definition | Exposure ascertainment | Validated/  objectively recorded | Exposure measure  type | Exposure time period | SM platform | SM platform | SM category  (active/  passive use) | SM content |
| --- | --- | --- | --- | --- | --- | --- | --- | --- | --- | --- |
| Time Spent on SM | Kontostoli 2020 | Time spent browsing and updating SNS on a weekend day | Time use diary:  Participants recorded their behaviour in 10-min timeslots from 4 to 4 am the next day. For each 10-min timeslot, participants indicated their main activity, selecting from a pre-specified list of 44 activities, nested within 12 categories for both weekday and weekend day.  Participants were categorised according to whether they did (user) or did not (non-user) report time browsing and updating social networking sites. |  | Binary | Current | Examples: Facebook, Twitter, and Snapchat | Mixed platforms | SNS  (active and passive use) | NA |
| Time Spent on SM | Larm 2017 | Online social networking chatting | Self-report 2-item measure: 1-How often chats on online SNS such as Myspace, Facebook, and others. Responses: (1) never to (7) 6-7 days/week. 2-Average amount of time each day chatting on online SNS. Responses: (1) do not chat to (5) >5 hrs/day.  Summarised score from 0 to 10 derived from 2 items. |  | Continuous | Current | Examples: Myspace and Facebook | Mixed platforms | SNS  (active use) | NA |
| Time Spent on SM | Larm 2019 | Time spent on SM/chatting per day | Self-report 1-item measure: 1-How many hrs per day usually spent chatting on the internet/SM sites. Responses: not using computers, <1 hr/day, 1–2 hrs/day, 2–5 hrs/day, and >5 hrs/day.  Variable dichotomised: ≥2 hrs/day/<2 hrs/day. |  | Binary | Current | NR | NR | General SM  (active use) | NA |
| Time Spent on SM | Lee 2015 | Time spent on SNS per day | Self- report 7-item measure:  1- SNS usage time.  Responses: < 30 min, 30 min- 1 hr, 1-2 hrs, 2-3 hrs, and >3 hrs/day. |  | Binary | Current | NR | NR | SNS  (unclear) | NA |
| Time Spent on SM | Lee 2021 | Daytime use of social networks | Objective measure:  Smartphone assessed usage of social networks. Time spent on each session computed as the difference between the closing and opening time. Usage sessions of <1 second were discarded.  Daytime usage defined as the time from waking up to 1 hr before sleep, where sleeping time was identified using accelerometer data. | Yes | Continuous | Past week | Examples: FB, Twitter, Instagram, and Weibo | Mixed platforms | SNS  (unclear) | NA |
| Time Spent on SM | Lee 2021 | Bedtime use of social networks | Objective measure:  Smartphone assessed usage of social networks. Time spent on each session computed as the difference between the closing and opening time. Usage sessions of <1 second were discarded.  Bedtime usage defined as 1 hr before sleep, where sleeping time was identified using accelerometer data. | Yes | Continuous | Past week | Examples: FB, Twitter, Instagram, and Weibo | Mixed platforms | SNS  (unclear) | NA |

| Exposure | Author and year | Exposure definition | Exposure ascertainment | Validated/  objectively recorded | Exposure measure  type | Exposure time period | SM platform | SM platform | SM category (active/  passive use) | SM content |
| --- | --- | --- | --- | --- | --- | --- | --- | --- | --- | --- |
| Time Spent on SM | Lee 2021 | Use of social networks at wakeful moment during sleep | Objective measure:  Smartphone assessed usage of social networks. Time spent on each session computed as the difference between the closing and opening time. Usage sessions of <1 second were discarded. | Yes | Continuous | Past week | Examples: FB, Twitter, Instagram, and Weibo | Mixed platforms | SNS  (unclear) | NA |
| Time Spent on SM | Lipsky 2017 | Time spent on social networking per day | Self-report 2-item measure: 1-Time spent using a computer or cell phone for chatting online, internet, emailing, texting, tweeting, or similar social networking (other than for a job or school work) during weekdays. 2-Time spent using a computer or cell phone for chatting online, internet, emailing, texting, tweeting, or similar social networking (other than for a job or school work) during the weekend. Responses: 0.5 hrs, ~1, ~2, ~3, ~4, ~5, ~6, and ~≥7 hrs/day - converted to the number of hrs per day (ranging from 0 to 7). |  | Continuous | Current | NR | NR | SNS  (active use) | NA |
| Time Spent on SM | Longobardi 2021 | Time spent on SM per day | Self-report measure:  No information provided. |  | Continuous | Current | NR | NR | General SM  (unclear) | NA |
| Time Spent on SM | Merrill 2019 | Time spent on SM per day | Self-report 1-item measure: 1-How many hrs plays video or computer games or uses a computer for something that is not schoolwork (incl. Xbox, PlayStation, an iPod, an iPad or other tablet, a smartphone, YouTube, Facebook or other social networking tools, and the Internet) on an average school day.  Due to the interactive nature of the items included as “computer use,” this was relabelled “social media use” by study investigators.  Responses: 0 hrs per average school day, <1 hr, 1 hrs, 2 hrs, 3 hrs, 4 hrs, and ≥5 hrs/day. |  | Continuous | Current | Examples: Xbox, PlayStation, iPod, iPad or other tablet, smartphone, YouTube, Facebook or other social networking tools and the internet | Mixed platforms | General SM  (unclear) | NA |
| Time Spent on SM | Michael 2016 | Average time spent on SM per day | Self-report 1-item measure: 1-How much time spent on SM per day. Responses: 0-1 hrs, 2-3 hrs, 4-5 hrs, 6-7 hrs, ≥8 hrs/day  3 category variable constructed: low, moderate, and high. |  | Categorical | Current | Examples reported by participants Facebook, 2 go, Twitter, WhatsApp, and YouTube | Mixed platforms | SNS  (unclear) | NA |
| Time Spent on SM | Moitra 2022 | Time spent using SNS on a typical weekend and weekday | Self-report 1-item measure:  1-How much time spent on SNS on a typical weekend and weekday.  Responses: mins/day |  | Continuous | Past week | NR | NR | SNS  (unclear) | NA |
| Time Spent on SM | Mojica 2014 | Average time spent on SNS per week | Self-report 2-item measure: 1-Number of days SNS used in the past week. 2-Number of hrs per day of SNS use.  Items combined to create mean hrs per week. |  | Continuous | Current | NR | NR | SNS  (unclear) | NA |

| Exposure | Author and year | Exposure definition | Exposure ascertainment | Validated/  objectively recorded | Exposure measure  type | Exposure time period | SM platform | SM platform | SM category  (active/  passive use) | SM content |
| --- | --- | --- | --- | --- | --- | --- | --- | --- | --- | --- |
| Time Spent on SM | Nesi 2017 | Average time on Facebook per day | Self-report 1-item measure: 1-Average time spent on Facebook per day.  Responses: 7-point scale (1) <10 min to (7) ≥ 4 hrs/day. |  | Continuous | Current | Specified: Facebook | Facebook | SNS  (unclear) | NA |
| Time Spent on SM | Ng Fat 2021 | Time spent on SM on a normal weekday | Self-report 1-item measure: 1-How many hrs spent chatting or interacting with friends through social websites on a normal weekday. Responses: no-profile (those not on SM sites), non-daily user, <1 hr, 1–3 hrs, ≥4 hrs/day. |  | Ordinal | Current | Examples: Facebook, Myspace, and Bebo | Mixed platforms | SNS  (active use) | NA |
| Time Spent on SM | Ohannessian 2009 | Time spent emailing and instant messaging (IM) on an average/typical day | Self-report 1-item measure: 1-How much time spent emailing/instant messaging (IM) on an average/typical day. Responses: (1) none to (6) ≥4 hrs/day.  Variable dichotomised: high levels of email or IM use (≥1 hr/day)/low levels of email or IM use. |  | Binary | Current | NR | NR | SNS  (active use) | NA |
| Time Spent on SM | Sampasa-Kanyinga 2015 | Time spent on SM websites either posting or browsing per day | Self-report 1-item measure: 1-How many hrs a day spent on SM websites such as Facebook, Twitter, Instagram, Myspace, either posting or browsing.  Responses: do not use, visit these websites but not daily, <1 hr, about 1 hr, 2 hrs, 3–4 hrs, and ≥5 hrs/day. |  | Ordinal | Current | Examples: Facebook, Twitter, Instagram, and Myspace | Mixed platforms | SNS  (active and passive use) | NA |
| Time Spent on SM | Sampasa-Kanyinga 2015 | Time spent on SM websites either posting or browsing per day | Self-report 1-item measure: 1-How many hrs a day spent on SM websites such as Facebook, Twitter, Instagram, Myspace, either posting or browsing.  Responses: do not use, visit these websites but not daily, <1 hr, about 1 hr, 2 hrs, 3–4 hrs, and ≥5 hrs/day. |  | Ordinal | Current | Examples: Facebook, Twitter, Instagram, and Myspace | Mixed platforms | SNS  (active and passive use) | NA |
| Time Spent on SM | Sampasa-  Kanyinga 2016 | Time spent on SM websites either posting or browsing per day | Self-report 1-item measure: 1-How many hrs a day spent on SM websites such as Facebook, Twitter, Instagram, Myspace, either posting or browsing.  Responses: daily use (< 1 hr, about 1 hr, 2 hrs, 3-4 hrs, 5- 6 hrs, and ≥7 hrs/day).  3 category variable constructed: infrequent or no use of SNS (use SM, but not daily; use the Internet, but never visit SNS; and do not use the Internet), regular use (daily use ≤ 2 hrs), and heavy use (3-4 hrs, 5- 6 hrs, and ≥7 hrs/day). |  | Ordinal | Current | Examples: Facebook, Twitter, Instagram, and Myspace | Mixed platforms | SNS  (active and passive use) | NA |

| Exposure | Author and year | Exposure definition | Exposure ascertainment | Validated/  objectively recorded | Exposure measure  type | Exposure time period | SM platform | SM platform | SM category  (active/  passive use) | SM content |
| --- | --- | --- | --- | --- | --- | --- | --- | --- | --- | --- |
| Time Spent on SM | Tao 2022 | Average time on SM per week | Self-report 2-item measure:  1-In the past month, on average, approximately how many days in each week was time spent on SM (e.g., Instagram, Snapchat, Facebook, Tumble, Reddit)  Responses: 0 to 7 days/week.  2-In the past month, on average approximately how many hours in each day was time spent on SM (e.g., Instagram, Snapchat, Facebook, Twitter, Tumblr, Reddit).  Average hours of SM use per week calculated by multiplying the number of hrs of SM use a day (0-24) with days of SM use per week (0-7) |  | Continuous | Past month | Examples: Instagram, Snapchat, Facebook, Twitter, Tumblr, and Reddit | Mixed platforms | General SM  (unclear) | NA |
| Time Spent on SM | Vente 2020 | Time spent on SM per day | Self-report 1-item measure:  1-Total time spent on SM per day.  Variable dichotomised: >5 hrs of SM use per day/5 or less hrs of SM use. |  | Binary | Current | NR | NR | General SM  (unclear) | NA |
| Time Spent on SM | Worku 2022 | Stayed more than 2 hrs/day on SM | Self-report 1-item measure:  1-Long stay on SM.  Variable dichotomised: >2 / ≤ 2 hrs/day on SM |  | Binary | Current | NR | NR | General SM  (unclear) | NA |
| Time Spent on SM | Sampasa-  Kanyinga 2016 | Time spent on SM websites either posting or browsing per day | Self-report 1-item measure: 1-How many hrs a day spent on SM websites such as Facebook, Twitter, Instagram, Myspace, either posting or browsing.  Responses: do not use, visit these websites but not daily, <1 hr, about 1 hr, 2 hrs, 3–4 hrs, and ≥5 hrs/day. |  | Ordinal | Current | Examples: Facebook, Twitter, Instagram, and Myspace | Mixed platforms | SNS  (active and passive use) | NA |
| Time Spent on SM | Sampasa-Kanyinga 2018 | Time spent on SM websites either posting or browsing per day | Self-report 1-item measure: 1-How many hrs a day spent on SM websites such as Facebook, Twitter, Instagram, Myspace, either posting or browsing.  Responses: < 1 hr, about 1 hr, 2 hrs, 3-4 hrs, 5- 6 hrs, ≥7 hrs/day, visit these web sites but not daily, use the Internet but never visit these web sites, and do not use the Internet.  5 category variable constructed: Infrequent or no use of SNS (visit these web sites but not daily, use the internet but never visit these web sites, and do not use the internet), <1 hr, 1 hr, 2/hrs, and ≥3 hrs/day. |  | Ordinal | Current | Examples: Facebook, Twitter, Instagram, and Myspace | Mixed platforms | SNS  (active and passive use) | NA |

| Exposure | Author and year | Exposure definition | Exposure ascertainment | Validated/  objectively recorded | Exposure measure  type | Exposure time period | SM platform | SM platform | SM category  (active/  passive use) | SM content |
| --- | --- | --- | --- | --- | --- | --- | --- | --- | --- | --- |
| Time Spent on SM | Sandercock 2016 | Time spent on SM on a normal day | Self-report 2-item measure: 1-Use of SM. Responses: yes/no. 2- If yes, asked how long spent using SM on a normal day. Responses: <30 min, 30-60 min, 60-90 min, 90 min- 2 hrs, and >2 hrs/day. |  | Continuous | Current | NR | NR | General SM  (unclear) | NA |
| Time Spent on SM | Smout 2021 | Time spent on SM on a typical day | Self-report measure-  1-How many minutes spent on Facebook, Myspace, and other social networking sites on a typical day.  Responses greater than 12h per day truncated to 12h |  | Continuous | Current | Specified: Facebook, Myspace, and other social networking sites | Mixed platforms | SNS  (unclear) | NA |
| Time Spent on SM | Whitehill 2020 | Time spent on SM per day | Self-report measure:  Responses: <30 min, 30-60 min, 1-2 hrs, 2-4 hrs, and ≥ 4 hrs/day. No further information provided. |  | Ordinal | Current | NR | NR | General SM  (unclear) | NA |
| Freq. of SM use | Anastario 2020 | Freq. of use of Twitter to talk/learn about sex or any topic related to sex | Self-report 1-item measure:  1-How often do you use Twitter to talk or learn about sex or any topic related to sex? Responses: daily, a few days a week, every few weeks, less often, never use |  | Continuous | Current | Specified: Twitter | Twitter | Micro-blogging  (active and passive use) | NA |
| Freq. of SM use | Anastario 2020 | Freq. of use of Snapchat to talk/learn about sex or any topic related to sex | Self-report 1-item measure:  1-How often do you use Snapchat to talk or learn about sex or any topic related to sex? Responses: daily, a few days a week, every few weeks, less often, never use |  | Continuous | Current | Specified: Snapchat | Snapchat | SNS  (active and passive use) | NA |
| Freq. of SM use | Anastario 2020 | Freq. of use of Facebook to talk/learn about sex or any topic related to sex | Self-report 1-item measure:  1-How often do you use Facebook to talk or learn about sex or any topic related to sex? Responses: daily, a few days a week, every few weeks, less often, never use |  | Continuous | Current | Specified: Facebook | Facebook | SNS  (active and passive use) | NA |
| Freq. of SM use | Baker 2016 | Freq. of SNS use | Self-report 1- item measure: 1- How often used Myspace.com, Facebook.com, chat rooms or other online social networking websites.  Variable dichotomised: frequent SNS users (using such sites a few times per week, each week or everyday)/infrequent SNS users (using SNS never, a few times per year or a few times per month). |  | Binary | Current | Specified: Myspace, Facebook, chat rooms, and other SNS | Mixed platforms | SNS  (unclear) | NA |
| Freq. of SM use | Baldwin 2018 | Freq. of watching videos on YouTube | Self-report 1-item measure: 1-How often watch videos on YouTube. Reponses: never or rarely, less than once a week, a few times a week, once a day, a few times a day, and many times every day.  Variable dichotomised: at least daily users/less than daily users. |  | Binary | Current | Specified: YouTube | YouTube | Media-sharing  (passive use) | NA |

| Exposure | Author and year | Exposure definition | Exposure ascertainment | Validated/  objectively recorded | Exposure measure  type | Exposure time period | SM platform | SM platform | SM category  (active/  passive use) | SM content |
| --- | --- | --- | --- | --- | --- | --- | --- | --- | --- | --- |
| Freq. of SM use | Baldwin 2018 | Freq. of logging in, or checking Facebook account | Self-report 1-item measure: 1-How often login to, or check, Facebook account. Response options: never or rarely, less than once a week, a few times a week, once a day, a few times a day, and many times every day.  Variable dichotomised: at least daily users/less than daily users. |  | Binary | Current | Specified: Facebook | Facebook | SNS  (passive use) | NA |
| Freq. of SM use | Baru 2020 | Freq. of SM use | Self-report 1-item measure:  1-Frequency of SM use (Facebook, WhatsApp, IMO, Instagram etc)  Response: many times a day, several times a week, once a while |  | Ordinal | Current | Specified: Facebook, WhatsApp, instant messenger, and Instagram | Mixed platforms | SNS  (unclear) | NA |
| Freq. of SM use | Baumgartner 2012 | Freq. of online communication | Self-report 1-item measure: 1- How often use instant messaging, internet chats, and SNS. Responses: (0) never to (10) every day. |  | Continuous | Current | Specified: Instant messaging, internet chats, and SNS | Mixed platforms | SNS  (unclear) | **NA** |
| Freq. of SM use | Ball 2020 | Freq. of using SM (status updates, uploading photos or videos) | Self-report measure:  1-Internet activities used during the past 7 days.  Response options included SM activity (status updates and uploading photos or videos). |  | Binary | Past week | Examples: Facebook, Twitter, Instagram, Snapchat, and YouTube | NR | General SM  (active use) | NA |
| Freq. of SM use | Ball 2020 | Freq. of online gambling | Self-report measure:  1-Internet activities used during the past 7 days.  Response options included online gambling activity. |  | Binary | Past week | NR | NR | Online Gambling  (active use) | NA |
| Freq. of SM use | Boniel-Nissim 2022 | Freq. of online contact with others via SM | Self-report validated questionnaire:  Social Media Use Intensity Scale and Social Media Disorder Scale used to categorise participants into non-active users (online contact with others not at all or at most weekly and non-problematic user) and active users (online contact with others daily but not all the time and non-problematic user) | Yes | Binary | Current | NR | NR | General SM  (active and passive use) | NA |
| Freq. of SM use | Beebe 2004 | Presence of internet chat room use | Self-report 2-item measure: 1- Use the internet at home. 2- If yes, asked what internet is used for at home.  Those who checked the option “chat rooms” compared with those who did not. |  | Binary | Current | NR | NR | SNS  (active use) | NA |
| Freq. of SM use | Canale 2016 | Freq. of using internet for leisure activities | Self-report 1-item measure: 1-Internet used for leisure activities (e.g., online chatting, playing online games) Responses: yes/no. |  | Binary | Current | NR | NR | General SM  (active use) | NA |

| Exposure | Author and year | Exposure definition | Exposure ascertainment | Validated/  objectively recorded | Exposure measure  type | Exposure time period | SM platform | SM platform | SM category  (active/  passive use) | SM content |
| --- | --- | --- | --- | --- | --- | --- | --- | --- | --- | --- |
| Freq. of SM use | Canale 2016 | Freq. of online gambling in past year | Self-report 1-item measure: 1-How many occasions (if any) participated in online gambling activities. Responses: 7 options from 0 times to ≥40 times. Variable dichotomised: online gambler (anyone who had participated in online gambling at least once in the past 12 months)/non-online gamblers. |  | Binary | Past year | NR | NR | Online Gambling  (active use) | NA |
| Freq. of SM use | Chang 2016 | Freq. of chat room use during past week | Self-report 1-item measure: 1-How many days during the past week used chat rooms. Responses: 0 to 7 days. |  | Continuous | Past week | NR | NR | SNS  (active use) | NA |
| Freq. of SM use | Chang 2016 | Freq. of online game use during past week | Self-report 1-item measure: 1-How many days during the past week played online games. Responses: 0 to 7 days. |  | Continuous | Past week | NR | NR | Online Gaming  (active use) | NA |
| Freq. of SM use | Critchlow 2019 | SM apps used at least weekly | Self-report measure: 1-Which, if any, of the following apps used at least once a week: (1) Facebook; (2) Instagram; (3) Pinterest; (4) Snapchat; (5) Spotify; (6) Tumblr; (7) Twitter; (8) WhatsApp; (9) YouTube; and (10) Other, with free text box to write in.  Responses: yes/no/none of the above.  Cumulative score computed for SM apps used at least weekly (0–10), and 3 category variable constructed: high (6 or more apps), medium (4 or 5), and low use (3 or fewer). |  | Ordinal | Past week | Specified: Facebook, Instagram, Pinterest, Snapchat, Spotify, Tumblr, Twitter, WhatsApp, YouTube, and other | Mixed platforms | General SM  (unclear) | NA |
| Freq. of SM use | Dawson 2019 | Freq. of Facebook use | Self-report measure assessed via The Online Behaviour Demographic Questionnaire adapted from the Pew Research Centre's Internet, Science, and Tech self-report survey.^156^ Responses: (1) less than once a week to (6) almost constantly. |  | Continuous | Current | Specified: Facebook | Facebook | SNS  (unclear) | NA |
| Freq. of SM use | Dawson 2019 | Presence of SNS use | Self-report measure assessed via The Online Behaviour Demographic Questionnaire adapted from the Pew Research Centre’s Internet, Science, and Tech self-report survey.^156^ Responses: (0) not reported, and (1) reported. |  | Binary | Current | Examples: Instagram, Snapchat, Facebook, Twitter, Skype, Kik, Tumblr, Pinterest, Curious Cat, Discord, and Amino | Mixed platforms | SNS  (unclear) | NA |
| Freq. of SM use | Dawson 2019 | Presence of messaging app use | Self-report measure assessed via The Online Behaviour Demographic Questionnaire adapted from the Pew Research Centre’s Internet, Science, and Tech self-report survey.^156^ Responses: (0) not reported, and (1) reported. |  | Binary | Current | Example: WhatsApp | Whats-App | SNS  (unclear) | NA |

| Exposure | Author and year | Exposure definition | Exposure ascertainment | Validated/  objectively recorded | Exposure measure  type | Exposure time period | SM platform | SM platform | SM category  (active/  passive use) | SM content |
| --- | --- | --- | --- | --- | --- | --- | --- | --- | --- | --- |
| Freq. of SM use | Dawson 2019 | Presence of discussion board use | Self-report measure assessed via The Online Behaviour Demographic Questionnaire adapted from the Pew Research Centre’s Internet, Science, and Tech self-report survey.^156^ Responses: (0) not reported, and (1) reported. |  | Binary | Current | Example: Reddit | Reddit | Social News Sites  (unclear) | NA |
| Freq. of SM use | Dawson 2019 | Presence of anonymous sharing app use | Self-report measure assessed via The Online Behaviour Demographic Questionnaire adapted from the Pew Research Centre’s Internet, Science, and Tech self-report survey.^156^ Responses: (0) not reported, and (1) reported. |  | Binary | Current | Example: Whisper | Whisper | SNS  (unclear) | NA |
| Freq. of SM use | Dawson 2019 | Presence of Twitter use | Self-report measure assessed via The Online Behaviour Demographic Questionnaire adapted from the Pew Research Centre’s Internet, Science, and Tech self-report survey.^156^ Responses: (0) not reported, and (1) reported. |  | Binary | Current | Specified: Twitter | Twitter | Micro-blogging  (unclear) | **NA** |
| Freq. of SM use | Dawson 2019 | Presence of Instagram use | Self-report measure assessed via The Online Behaviour Demographic Questionnaire adapted from the Pew Research Centre’s Internet, Science, and Tech self-report survey.^156^ Responses: (0) not reported, and (1) reported. |  | Binary | Current | Specified: Instagram | Instagram | Media-sharing  (unclear) | NA |
| Freq. of SM use | Dawson 2019 | Presence of Snapchat use | Self-report measure assessed via The Online Behaviour Demographic Questionnaire adapted from the Pew Research Centre’s Internet, Science, and Tech self-report survey.^156^ Responses: (0) not reported, and (1) reported. |  | Binary | Current | Specified: Snapchat | Snapchat | SNS  (unclear) | NA |
| Freq. of SM use | Dawson 2019 | Presence of Facebook use | Self-report measure assessed via The Online Behaviour Demographic Questionnaire adapted from the Pew Research Centre’s Internet, Science, and Tech self-report survey.^156^ Responses: (0) not reported, and (1) reported. |  | Binary | Current | Specified: Facebook | Facebook | SNS  (unclear) | NA |
| Freq. of SM use | Dawson 2019 | Facebook activity: keeping up with friends (passive) | Self-report measure assessed via The Online Behaviour Demographic Questionnaire adapted from the Pew Research Centre’s Internet, Science, and Tech self-report survey.^156^  Facebook activity coded based on posting activity: (0) no to minimal activity, 0-25th percentile, and (3) heavy user, 75th+ percentile. Responses: (0) not reported, and (1) reported. |  | Binary | Current | Specified: Facebook | Facebook | SNS  (passive use) | NA |

| Exposure | Author and year | Exposure definition | Exposure ascertainment | Validated/  objectively recorded | Exposure measure  type | Exposure time period | SM platform | SM platform | SM category  (active/  passive use) | SM content |
| --- | --- | --- | --- | --- | --- | --- | --- | --- | --- | --- |
| Freq. of SM use | Dawson 2019 | Facebook activity: posting on own timeline (active) | Self-report measure assessed via The Online Behaviour Demographic Questionnaire adapted from the Pew Research Centre’s Internet, Science, and Tech self-report survey.^156^  Facebook activity coded based on posting activity: (0) no to minimal activity, 0-25th percentile, and (3) heavy user, 75th+ percentile. Responses: (0) not reported, and (1) reported. |  | Binary | Current | Specified: Facebook | Facebook | SNS  (active use) | NA |
| Freq. of SM use | Dawson 2019 | Facebook activity: commenting on friend’s posts (active) | Self-report measure assessed via The Online Behaviour Demographic Questionnaire adapted from the Pew Research Centre's Internet, Science, and Tech self-report survey.^156^  Facebook activity coded based on posting activity: (0) no to minimal activity, 0-25th percentile, and (3) heavy user, 75th+ percentile. Responses: (0) not reported, and (1) reported. |  | Binary | Current | Specified: Facebook | Facebook | SNS  (active use) | NA |
| Freq. of SM use | Dawson 2019 | Facebook activity: looking at videos or news stories (passive) | Self-report measure assessed via The Online Behaviour Demographic Questionnaire adapted from the Pew Research Centre’s Internet, Science, and Tech self-report survey.^156^  Facebook activity coded based on posting activity: (0) no to minimal activity, 0-25th percentile, and (3) heavy user, 75th+ percentile.  Responses: (0) not reported, and (1) reported. |  | Binary | Current | Specified: Facebook | Facebook | SNS  (passive use) | NA |
| Freq. of SM use | Dawson 2019 | Facebook interactions with friends they see daily | Self-report measure assessed via The Online Behaviour Demographic Questionnaire adapted from the Pew Research Centre’s Internet, Science, and Tech self-report survey.^156^ Responses: (0) not reported, and (1) reported. |  | Binary | Current | Specified: Facebook | Facebook | SNS  (active use) | NA |
| Freq. of SM use | Dawson 2019 | Facebook interactions with friends they see occasionally | Self-report measure assessed via The Online Behaviour Demographic Questionnaire adapted from the Pew Research Centre’s Internet, Science, and Tech self-report survey.^156^ Responses: (0) not reported, and (1) reported. |  | Binary | Current | Specified: Facebook | Facebook | SNS  (active use) | NA |
| Freq. of SM use | Dawson 2019 | Facebook interactions with online friends | Self-report measure assessed via The Online Behaviour Demographic Questionnaire adapted from the Pew Research Centre’s Internet, Science, and Tech self-report survey.^156^ Responses: (0) not reported, and(1) reported. |  | Binary | Current | Specified: Facebook | Facebook | SNS  (active use) | NA |
| Freq. of SM use | Dawson 2019 | Facebook interactions with family members | Self-report measure assessed via The Online Behaviour Demographic Questionnaire adapted from the Pew Research Centre’s Internet, Science, and Tech self-report survey.^156^ Responses: (0) not reported, and (1) reported. |  | Binary | Current | Specified: Facebook | Facebook | SNS  (active use) | NA |

| Exposure | Author and year | Exposure definition | Exposure ascertainment | Validated/  objectively recorded | Exposure measure  type | Exposure time period | SM platform | SM platform | SM category  (active/  passive use) | SM content |
| --- | --- | --- | --- | --- | --- | --- | --- | --- | --- | --- |
| Freq. of SM use | Dawson 2019 | Number of participant posts on Facebook (posted by participant) | Observationally coded measure, coded using Mikami and Szwedo's Facebook Coding Manual.^157^  Facebook profile coded over 2-month period to obtain total number of participant posted posts. | Yes | Continuous | Current | Specified: Facebook | Facebook | SNS  (active use) | NA |
| Freq. of SM use | Dawson 2019 | % of participant posts that were shared external material on Facebook | Observationally coded measure, coded using Mikami and Szwedo's Facebook Coding Manual.^157^  Facebook profile coded over 2-month period to obtain proportion of total participant posts that were “shared” external material (i.e., memes or web links copied and reposted from another location; types: emotional, animal related, intended humour, sports, motivational, news/politics, and music). | Yes | Continuous | Current | Specified: Facebook | Facebook | SNS  (active use) | NA |
| Freq. of SM use | Dawson 2019 | % of participant posts sharing accomplishments on Facebook | Observationally coded measure, coded using Mikami and Szwedo's Facebook Coding Manual.^157^  Facebook profile coded over 2-month period to obtain proportion of total participant posts sharing accomplishments (i.e., something that typically infers pride in some skillset or effort). | Yes | Continuous | Current | Specified: Facebook | Facebook | SNS  (active use) | NA |
| Freq. of SM use | Dawson 2019 | % of participant posts illustrating connection on Facebook | Observationally coded measure, coded using Mikami and Szwedo's Facebook Coding Manual.^157^  Facebook profile coded over 2-month period to obtain proportion of total participant posts illustrating connection with friends (e.g., meetings withing the year, specific plans for future meetings, or shared information (such as inside jokes). | Yes | Continuous | Current | Specified: Facebook | Facebook | SNS  (active use) | NA |
| Freq. of SM use | Dawson 2019 | % of participants posts sharing support on Facebook | Observationally coded measure, coded using Mikami and Szwedo's Facebook Coding Manual.^157^  Facebook profile coded over 2-month period to obtain proportion of total participant posts containing emotional support (i.e., posts offering encouragement, validation, compliments, or empathy). | Yes | Continuous | Current | Specified: Facebook | Facebook | SNS  (active use) | NA |
| Freq. of SM use | De Looze 2019 | Freq. of electronic media communication with friends | Self-report 3-item measure: Asked how often: 1- contacted friends using texting/SMS;  2- actively contacted friends using instant messaging (e.g., Facebook chat);  3- contacted friends using other SM, such as Facebook (posting on wall, not chat), Myspace, Twitter, apps (e.g., Instagram), games (e.g., Xbox), YouTube.  Responses: hardly ever or never, less than weekly, weekly, and daily.  Variable dichotomised: (0) less than daily/(1) daily. |  | Binary | Current | Examples: Blackberry Messaging, Facebook chat, Facebook, Myspace, Twitter, Instagram, Xbox, and YouTube | Mixed platforms | SNS  (active use) | NA |

| Exposure | Author and year | Exposure definition | Exposure ascertainment | Validated/  objectively recorded | Exposure measure  type | Exposure time period | SM platform | SM platform | SM category (active/  passive use) | SM content |
| --- | --- | --- | --- | --- | --- | --- | --- | --- | --- | --- |
| Freq. of SM use | Elton-Marshall 2016 | Freq. of playing free simulated gambling games on Facebook | Self-report 1-item measure:  1-Participation in any online gambling games on Facebook for fun (no money). Responses: not in the past 3 months, about once per month, 2–3 times per month, about once per week, 2–6 times per week, and daily.  Variable dichotomised: at least monthly but less than weekly (about once per month or 2–3 times per month)/at least weekly (about once per week, 2–6 times per week, or daily).  Overall prevalence based on any participation (indicated about once per month or more frequent). |  | Binary | Past 3 months | Specified: Facebook | Facebook | Online Gambling  (active use) | NA |
| Freq. of SM use | Elton-Marshall 2016 | Freq. of online gambling participation | Self-report measure: Online gamblers: respondents who indicated that they had gambled money or something of value in the past for any of 3 online gambling activities: internet poker, sports pools online, and slot machines online.  Land-based gamblers: respondents who had gambled money or something of value in the past 3 months but had not participated in any of the online gambling activities.  No further information reported. |  | Binary | Past 3 months | NR | NR | Online Gambling  (active use) | NA |
| Freq. of SM use | Erreygers 2017 | Freq. of online gaming | Self-report 2-item measure: 1-How often used online gaming (playing with others). 2-How often used online gaming (playing alone or against the computer). Responses: never, just a few times, 1-4 times per month, almost every day, multiple times per day, I don’t know and not applicable.  Confirmatory factor analyses used to generate online gaming factor. |  | Continuous | Past 6 months | NR | NR | Online Gaming  (active use) | NA |
| Freq. of SM use | Floros 2013 | Freq. of using SNS | Self-report measure: Internet activities measured on Likert scale for frequency. No further information reported. |  | Continuous | Past year | NR | NR | SNS  (unclear) | NA |
| Freq. of SM use | Floros 2013 | Freq. of online discussions in real time (IRC, MSN etc) | Self-report measure: Internet activities measured on Likert scale for frequency. No further information reported. |  | Continuous | Past year | NR | NR | SNS  (active use) | NA |
| Freq. of SM use | Floros 2013 | Freq. of online discussions with posts on boards, forums | Self-report measure: Internet activities measured on Likert scale for frequency. No further information reported. |  | Continuous | Past year | NR | NR | Blogs & Forums  (active use) | NA |

| Exposure | Author and year | Exposure definition | Exposure ascertainment | Validated/  objectively recorded | Exposure measure  type | Exposure time period | SM platform | SM platform | SM category (active/  passive use) | SM content |
| --- | --- | --- | --- | --- | --- | --- | --- | --- | --- | --- |
| Freq. of SM use | Gregg 2018 | Freq. of electronic communication | Self-report 3-item measure: 1-How long used SM on a normal school day. Responses: from (0) never, to (7) >4 hrs/day. 2-How long used SM on a normal non-school day. Responses: from (0) never, to (7) >4 hrs/day. 3- How many text messages sent on an average day.  Responses: from (0) I do not text to (6) >300.  Responses added to produce overall estimate of SM use. Higher scores indicated more frequent use of SM. |  | Continuous | Current | NR | NR | General SM  (unclear) | NA |
| Freq. of SM use | Gunnlaugsson 2020 | Freq. of SM use | Self-report 1-item validated measure:  1-Experience of communicating in the last 12 months on SM with friends, family, and people the respondent would like to know.  Responses: everyday, 2-3 times a week, every week, less than monthly, and never. | Yes | Binary | Past year | NR | NR | General SM  (active use) | NA |
| Freq. of SM use | Hayer 2018 | Freq. of participation in any simulated gambling on social networks | Self-report 1-item measure: 1-Frequency of participation in any simulated gambling on social networks. Responses (5 options): from not at all to more than 8 times a month.  Variable dichotomised: participation/no participation. |  | Binary | Past year | NR | NR | Online Gambling  (active use) | NA |
| Freq. of SM use | Hayer 2018 | Freq. of participation in any simulated gambling via apps | Self-report 1-item measure: 1- Frequency of participation in any simulated gambling via apps. Responses (5 options): from not at all to more than 8 times a month.  Variable dichotomised: participation/no participation. |  | Binary | Past year | NR | NR | Online Gambling  (active use) | NA |
| Freq. of SM use | Hayer 2018 | Freq. of participation in simulated gambling from home on social networks | Self-report 1-item measure: 1-Frequency of participation in simulated gambling from home on social networks. Responses (5 options): from not at all to more than 8 times a month.  Variable dichotomised: participation/no participation. |  | Binary | Past year | NR | NR | Online Gambling  (active use) | NA |
| Freq. of SM use | Hayer 2018 | Freq. of participation in simulated gambling from home via apps | Self-report 1-item measure: 1-Frequency of participation in simulated gambling from home via apps.  Responses (5 options): from not at all to more than 8 times a month.  Variable dichotomised: participation/no participation. |  | Binary | Past year | NR | NR | Online Gambling  (active use) | NA |
| Freq. of SM use | Hayer 2018 | Freq. of participation in simulated gambling when out and about on social networks | Self-report 1-item measure: 1-Frequency of participation in simulated gambling when out and about on social networks.  Responses (5 options): from not at all to more than 8 times a month.  Variable dichotomised: participation/no participation. |  | Binary | Past year | NR | NR | Online Gambling  (active use) | NA |

| Exposure | Author and year | Exposure definition | Exposure ascertainment | Validated/  objectively recorded | Exposure measure  type | Exposure time period | SM platform | SM platform | SM category  (active/  passive use) | SM content |
| --- | --- | --- | --- | --- | --- | --- | --- | --- | --- | --- |
| Freq. of SM use | Hayer 2018 | Freq. of participation in simulated gambling when out and about via apps | Self-report 1-item measure: 1-Frequency of participation in simulated gambling when out and about via apps.  Responses (5 options): from not at all to more than 8 times a month.  Variable dichotomised: participation/no participation. |  | Binary | Past year | NR | NR | Online Gambling  (active use) | NA |
| Freq. of SM use | Holtz 2011 | Freq. of online gaming | Self-report 3-item measure: Frequency of playing:  1-first person shooters online; 2-playing online role-playing games, and; 3-playing other games. Responses: 1 (never) to 5 (very often).  Latent factor generated. |  | Continuous | Current | NR | NR | Online Gaming  (active use) | NA |
| Freq. of SM use | Holtz 2011 | Freq. of communicational internet use (e.g., chatrooms, social platforms like Myspace) | Self-report 3-item measure: Frequency of use of: 1-email; 2-use of chatrooms; 3-use of social platforms like Myspace. Responses: (1) never to (5) very often.  Latent factor created. |  | Continuous | Current | Example: Myspace | Myspace | SNS  (active and passive use) | NA |
| Freq. of SM use | Hryhorczuk 2019 | Freq. of SM use | Self-report measure: 1-How free time spent. Adolescents who responded they use SM frequently or sometimes compared to those who said that they never use SM. |  | Binary | Current | NR | NR | General SM  (unclear) | NA |
| Freq. of SM use | Huang 2012 | Freq. of social internet activity (online gaming, chatting with real friends, chatting with online friends) | Self-report measure: 1-How often conducted the following computer and internet-based activities: online games, chatting with friends in real daily life and chatted with friends met online.  The average of the items played online games, chatting with friends in real daily life, and chatted with friends met online taken and loaded to create factor 'Social internet activity' and appropriate scale. |  | Continuous | Past week | NR | NR | SNS  (active use) | NA |
| Freq. of SM use | Huang 2014 | Freq. of Facebook use | Self-report 1-item measure: 1-How frequently SNS Facebook visited. Responses: never, rarely (about once a month or less), occasionally (about once a week or less), frequently (about once every 2-3 days), and very frequently (about once a day or more). |  | Ordinal | Past month | Specified: Facebook | Facebook | SNS  (unclear) | NA |

| Exposure | Author and year | Exposure definition | Exposure ascertainment | Validated/  objectively recorded | Exposure measure  type | Exposure time period | SM platform | SM platform | SM category  (active/  passive use) | SM content |
| --- | --- | --- | --- | --- | --- | --- | --- | --- | --- | --- |
| Freq. of SM use | Huang 2014 | Freq. of Myspace use | Self-report 1-item measure: 1-How frequently SNS Myspace visited. Responses: never, rarely (about once a month or less), occasionally (about once a week or less), frequently (about once every 2-3 days), and very frequently (about once a day or more). |  | Ordinal | Past month | Specified: Myspace | Myspace | SNS  (unclear) | NA |
| Freq. of SM use | Jeong 2022 | Freq. of SM use | Self-report 1-item measure:  1-SM usage frequency.  Response: not at all, sometimes, and often  Variable dichotomised: SM users (sometimes, often)/non-SM user (not at all). |  | Binary | Current | NR | NR | General SM  (unclear) | NA |
| Freq. of SM use | Jiang 2018 | Freq. of online gaming | Exposure ascertained via clinical records: 5 items assessed- online gaming history, frequency of online gaming, degree of involvement, number of gaming buddies, and amount of time of online gaming on average. Responses: 5-point Likert scale. | Yes | Continuous | Current | NR | NR | Online Gaming (active use) | NA |
| Freq. of SM use | Kaufman 2014 | Freq. of SM use | Self-report 1-item measure: 1-How often used Facebook, Mxit, or other social networks. Responses: every day, every 2–3 days, once a week, once every 2–3 weeks, never and almost never.  Variable dichotomised: SM used every day/ did not use SM every day. |  | Binary | Current | Examples: Facebook, Mxit, and other social networks | Mixed platforms | SNS  (unclear) | NA |
| Freq. of SM use | Kelleghan 2020 | Freq. of SM posting (posting photos, video or statuses and sharing others content) | Self-report 1-item measure:  1-How often posted own photographs, images, videos, status updates, or blogs over past week.  Responses: 0 times, 1-2 times per week, 1-2 times per day, and many times per day.  Variable dichotomised: high frequency use (multiple times per day)/less frequent use (0 times, 1-2 times per week, 1-2 times per day) |  | Binary | Current | NR | NR | General SM  (active use) | NA |
| Freq. of SM use | King 2014 | Ever use of simulated gambling via SNS applications | Self-report 1-item measure: 1-Ever tried simulated gambling via SNS applications (Facebook). Responses: yes/no. |  | Binary | Ever | Specified: Facebook | Facebook | Online Gambling  (active use) | NA |
| Freq. of SM use | Ko 2009 | Ever online chatting | Self-report 1-item measure: 1-Ever participated in online chatting.  No further information provided. |  | Binary | Ever | NR | NR | SNS  (active use) | NA |
| Freq. of SM use | Ko 2009 | Ever online gaming | Self-report 1-item measure: 1-Ever participated in online gaming.  No further information provided. |  | Binary | Ever | NR | NR | Online Gaming  (active use) | NA |
| Freq. of SM use | Ko 2009 | Ever online gambling | Self-report 1-item measure: 1-Ever participated in online gambling. No further information provided. |  | Binary | Ever | NR | NR | Online Gambling  (active use) | NA |

| Exposure | Author and year | Exposure definition | Exposure ascertainment | Validated/  objectively recorded | Exposure measure  type | Exposure time period | SM platform | SM platform | SM category  (active/  passive use) | SM content |
| --- | --- | --- | --- | --- | --- | --- | --- | --- | --- | --- |
| Freq. of SM use | Koutamanis 2015 | Freq. of online social exploration on SNS | Self-report 4-item measure: How often following things done on SNS: 1-invited someone to become friends; 2-commented on a message or picture of someone they don’t know that well; 3-sent a message to someone they don’t know that well; 4-asked someone whether they want to do something fun with them. Responses: never, almost never, sometimes, often, and very often.  Average of 4 items used to create composite scale. |  | Continuous | Current | NR | NR | SNS  (active use) | NA |
| Freq. of SM use | Kwon 2022 | Freq. of SNS use | Self-report measure:  1-Frequency of smartphone use at weekday and weekend day, and specific content accessed (e.g., SNS) |  | Continuous | Past month | Examples: Blogs, Instagram, Twitter, and Facebook | Mixed platforms | SNS  (unclear) | NA |
| Freq. of SM use | Landry 2013 | Freq. of logging into SM sites | Self-report measure: If internet used, how often, and if had accounts on any of the following SM sites: Facebook, Myspace, Twitter, Yahoo, YouTube, My Yearbook, Tumblr, Google buzz, Flickr, Ustream, and Other.  A count variable created for the number of SM accounts. If participant had an account, they were asked about their frequency of internet use and logging in to SM sites. Responses: several times a day, about once a day, 3 to 5 days week, 1 to 2 days a week, every few weeks, and less often.  Frequency of logging in to SM sites dichotomised: daily log-in/less frequent log-in. |  | Binary | Current | Specified: Facebook, Myspace, Twitter, Yahoo, YouTube, My Yearbook, Tumblr, Google buzz, Flickr, Ustream, and other | Mixed platforms | SNS  (unclear) | NA |
| Freq. of SM use | Lee 2019 | Freq. of visiting social networking account | Self-report 1-item measure: 1-How often visited Facebook, Google Plus, Myspace, Twitter, or other social networking account. Responses: no account/use, monthly or less, weekly, and daily. |  | Ordinal | Current | Examples: Facebook, Google plus, Myspace, and Twitter | Mixed platforms | SNS  (unclear) | NA |
| Freq. of SM use | Lee 2021 | Freq. of Facebook use | Self-report 1-item measure:  1-How often Facebook visited.  Responses: never, every few months, every few weeks, 1-2 days per week, 3-5 days per week, once per day, and several times per day.  3 category variable constructed: never/non-daily (every few months, every few weeks, 1-2 days per week, and 3-5 days per week)/daily (once per day, and several times per day) |  | Ordinal | Current | Specified: Facebook | Facebook | SNS  (unclear) | NA |

| Exposure | Author and year | Exposure definition | Exposure ascertainment | Validated/  objectively recorded | Exposure measure  type | Exposure time period | SM platform | SM platform | SM category  (active/  passive use) | SM content |
| --- | --- | --- | --- | --- | --- | --- | --- | --- | --- | --- |
| Freq. of SM use | Lee 2021 | Freq. of Instagram use | Self-report 1-item measure:  1-How often Instagram visited.  Responses: never, every few months, every few weeks, 1-2 days per week, 3-5 days per week, once per day, and several times per day.  3 category variable constructed: never/non-daily (every few months, every few weeks, 1-2 days per week, and 3-5 days per week)/daily (once per day, and several times per day) |  | Ordinal | Current | Specified: Instagram | Instagram | Media-sharing  (unclear) | NA |
| Freq. of SM use | Lee 2021 | Freq. of Twitter use | Self-report 1-item measure:  1-How often Twitter visited.  Responses: never, every few months, every few weeks, 1-2 days per week, 3-5 days per week, once per day, and several times per day.  3 category variable constructed: never/non-daily (every few months, every few weeks, 1-2 days per week, and 3-5 days per week)/daily (once per day, and several times per day) |  | Ordinal | Current | Specified: Twitter | Twitter | Micro-blogging  (unclear) | NA |
| Freq. of SM use | Lee 2021 | Freq. of Snapchat use | Self-report 1-item measure:  1-How often Snapchat visited.  Responses: never, every few months, every few weeks, 1-2 days per week, 3-5 days per week, once per day, and several times per day.  3 category variable constructed: never/non-daily (every few months, every few weeks, 1-2 days per week, and 3-5 days per week)/daily (once per day, and several times per day) |  | Ordinal | Current | Specified: Snapchat | Snapchat | SNS  (unclear) | NA |
| Freq. of SM use | McClure 2020 | Freq. of SM use | Self-report 1-item measure: 1-How often SM used. Responses: never, rarely, once in a while, about once a day, and many times a day.  Mean score calculated. |  | Continuous | Current | NR | NR | General SM  (unclear) | NA |
| Freq. of SM use | Molla-Esparza 2021 | Freq. of using SM platforms | Self-report 1-item measure:  1-How often SM used.  Responses: daily, several days a week, several days a month, almost never, never  Variable dichotomised: low usage frequency (never, almost never, and several days a month)/high usage frequency (several days a week and daily) |  | Binary | Current | NR | NR | General SM  (unclear) | NA |

| Exposure | Author and year | Exposure definition | Exposure ascertainment | Validated/  objectively recorded | Exposure measure  type | Exposure time period | SM platform | SM platform | SM category  (active/  passive use) | SM content |
| --- | --- | --- | --- | --- | --- | --- | --- | --- | --- | --- |
| Freq. of SM use | Nesi 2019 | Posted photos with peers on Instagram | Observationally coded measure: 1-Photos posted with peers, where both participant and same-age peers depicted.  Sum of the number of photos with peers posted during the 3-month coding period taken. | Yes | Continuous | During coding period | Specified: Instagram | Instagram | Media-sharing  (active use) | NA |
| Freq. of SM use | Nesi 2019 | Freq. of daily SM use | Self-report 1-item measure: 1-Average daily frequency of SM use defined as any website/app that involves social interaction, i.e., Facebook, Instagram, Tumblr, Snapchat. Responses: (0) I don’t use this to (6) ≥5 hrs/day. |  | Continuous | Current | Examples: texting, Facebook, Instagram, and Snapchat | Mixed platforms | SNS  (active use) | NA |
| Freq. of SM use | Nesi 2019 | Posted selfies on Instagram | Observationally coded measure: 1-Selfies, or photos of the participant alone. Sum of the number of selfies posted during the 3-month coding period taken. | Yes | Continuous | During coding period | Specified: Instagram | Instagram | Media-sharing  (active use) | NA |
| Freq. of SM use | Pegg 2018 | Freq. of SNS use (intensity) | Self-report 1-item measure: 1-How many hrs per week spent on SNS. Responses: 0 hrs/week to ≥30 hrs/week. |  | Continuous | Current | NR | NR | SNS  (unclear) | NA |
| Freq. of SM use | Prince 2021 | Freq. of Snapchat use | Self-report 1-item measure:  1-How often Snapchat used.  Responses: never, rarely, sometimes, and often  Variable dichotomised: sometimes/often. |  | Binary | Current | Specified: Snapchat | Snapchat | SNS  (unclear) | NA |
| Freq. of SM use | Riehm 2021 | Freq. of checking SM sites | Self-report 1-item measure:  1-How often checked SM sites.  Responses: none, 1 to 2 times per week, 1 to 2 times per day, and many times per day.  Variable dichotomised: high frequency engagement (many times per day)/lower frequency engagement |  | Binary | Current | Examples: Facebook, Twitter, and Instagram | NR | General SM  (passive use) | NA |
| Freq. of SM use | Riehm 2021 | Freq. of posting own photos, images, videos, status updates, or blogs on SM | Self-report 1-item measure:  1-How often posted own photos, images, videos, status updates, or blogs on SM.  Responses: none, 1 to 2 times per week, 1 to 2 times per day, and many times per day.  Variable dichotomised: high frequency engagement (many times per day)/lower frequency engagement |  | Binary | Current | NR | NR | General SM  (active use) | NA |
| Freq. of SM use | Riehm 2021 | Freq. of liking or commenting on other people’s statuses, wall posts, pictures, etc on SM | Self-report 1-item measure:  1-How often liked or commented on other people’s statuses, wall posts, pictures, etc on SM.  Responses: none, 1 to 2 times per week, 1 to 2 times per day, and many times per day.  Variable dichotomised: high frequency engagement (many times per day)/lower frequency engagement |  | Binary | Current | NR | NR | General SM  (active use) | NA |

| Exposure | Author and year | Exposure definition | Exposure ascertainment | Validated/  objectively recorded | Exposure measure  type | Exposure time period | SM platform | SM platform | SM category  (active/  passive use) | SM content |
| --- | --- | --- | --- | --- | --- | --- | --- | --- | --- | --- |
| Freq. of SM use | Riehm 2021 | Freq. of sharing other people's photos, images, videos, status updates, blogs, articles, news, or websites on SM | Self-report 1-item measure:  1-How often shared other people’s photos, images, videos, status updates, blogs, articles, news, or websites on SM.  Responses: none, 1 to 2 times per week, 1 to 2 times per day, and many times per day.  Variable dichotomised: high frequency engagement (many times per day)/lower frequency engagement |  | Binary | Current | NR | NR | General SM  (active use) | NA |
| Freq. of SM use | Romo 2017 | Freq. of SM app use | Self-report 1-item measure via validated questionnaire: 1-How often visited/used apps per day or week.  Variable dichotomised: frequent use of visiting online SM apps (>10 times per day)/infrequent use (≤10 times per day). | Yes | Binary | Current | NR | NR | General SM  (unclear) | NA |
| Freq. of SM use | Romo 2017 | Freq. of SNS use | Self-report 1-item measure via validated questionnaire: 1-How often SNS visited/used per day or week.  Variable dichotomised: frequent use of visiting online SNS (>10 times per day)/infrequent use (≤ 10 times per day). | Yes | Binary | Current | NR | NR | SNS  (unclear) | NA |
| Freq. of SM use | Rutter 2021 | Freq. of SM use (checking and posting) | Self-report measure:  Panel of surveys assessing SM use and SM rules.  No further information reported. |  | Continuous | Current | NR | NR | General SM  (unclear) | NA |
| Freq. of SM use | Savolainen 2020 | Freq. of Facebook use | Self-report 1-item measure via validated questionnaire: 1-How often Facebook used. Responses: I do not use, seldom, daily, and several times a day.  Variable dichotomised: daily user/non-daily user. | Yes | Binary | Current | Specified: Facebook | Facebook | SNS  (unclear) | NA |
| Freq. of SM use | Savolainen 2020 | Freq. of YouTube use | Self-report 1-item measure via validated questionnaire: 1-How often YouTube used. Responses: I do not use, seldom, daily, and several times a day.  Variable dichotomised: daily user vs non-daily user. | Yes | Binary | Current | Specified: YouTube | YouTube | Media-sharing  (unclear) | NA |
| Freq. of SM use | Savolainen 2020 | Freq. of Twitter use | Self-report 1-item measure via validated questionnaire: 1-How often Twitter used. Responses: I do not use, seldom, daily, and several times a day.  Variable dichotomised: daily user vs non-daily user. | Yes | Binary | Current | Specified: Twitter | Twitter | Micro-blogging  (unclear) | NA |
| Freq. of SM use | Savolainen 2020 | Freq. of Instagram use | Self-report 1-item measure via validated questionnaire: 1-How often Instagram used. Responses: I do not use, seldom, daily, and several times a day.  Variable dichotomised: daily user vs non-daily user. | Yes | Binary | Current | Specified: Instagram | Instagram | Media-sharing  (unclear) | NA |

| Exposure | Author and year | Exposure definition | Exposure ascertainment | Validated/  objectively recorded | Exposure measure  type | Exposure time period | SM platform | SM platform | SM category  (active/  passive use) | SM content |
| --- | --- | --- | --- | --- | --- | --- | --- | --- | --- | --- |
| Freq. of SM use | Savolainen 2020 | Freq. of instant messaging | Self-report 1-item measure via validated questionnaire:  1-How often instant messaging used (e.g., WhatsApp/Snapchat).  Responses: I do not use, seldom, daily, and several times a day.  Variable dichotomised: daily user vs non-daily user. | Yes | Binary | Current | Specified: Instant messaging. Examples: WhatsApp, and Snapchat | Instant Messaging | SNS  (active use) | NA |
| Freq. of SM use | Self-Brown 2018 | Presence of SM use | Self-report 1-item measure via validated questionnaire: 1-Used any type of SM (e.g., Twitter, Facebook). Responses: yes/no. | Yes | Binary | Current | Examples: Twitter and Facebook | Mixed platforms | General SM  (unclear) | NA |
| Freq. of SM use | Shimoga 2019 | Freq. of SM use | Self-report 1-item measure:  1-How often social networking websites like Facebook, Twitter, Instagram, etc visited. Responses: never, a few times a year, one to two times a month, once a week, and every day. |  | Ordinal | Current | Examples: Twitter, Facebook, and Instagram | Mixed platforms | SNS  (unclear) | NA |
| Freq. of SM use | Soneji 2018 | Freq. of social networking account use | Self-report 1-item measure: 1-How often Facebook, Google Plus, Myspace, Twitter, or other social networking account visited. Responses: several times a day, daily, weekly, monthly, and less. |  | Ordinal | Current | Examples: Facebook, Google plus, Myspace, Twitter, and other | Mixed platforms | SNS  (unclear) | NA |
| Freq. of SM use | Svensson 2020 | Freq. of posting information on Facebook, Instagram, Snapchat, or other SM | Self-report 1-item measure:  1-How often use a computer, mobile phone, or tablet to post information about yourself on Facebook, Instagram, Snapchat, or other SM.  Responses: never, once a month, about once a week, several times a day, everyday |  | Continuous | Current | Examples: Facebook, Instagram, Snapchat, or other SM | Mixed platforms | General SM  (active use) | NA |
| Freq. of SM use | Svensson 2020 | Freq. of staying in contact with and staying informed about friends via Facebook, Instagram or similar | Self-report 1-item measure:  1-How often use a computer, mobile phone, or tablet to stay in contact with and stay informed about friends via Facebook, Instagram or similar.  Responses: never, once a month, about once a week, several times a day, everyday |  | Continuous | Current | Examples: Facebook, Instagram, or similar | Mixed platforms | General SM  (active and passive use) | NA |
| Freq. of SM use | Tsitsika 2009 | Ever accessing the internet to visit chat rooms | Self-report measure: Primary objects of interest via internet. No further info provided |  | Binary | Current | NR | NR | SNS  (active use) | NA |
| Freq. of SM use | Tsitsika 2011 | Presence of internet chat room use | Self-report measure: Use of internet chat rooms.  No further information provided. |  | Binary | Current | NR | NR | SNS  (active use) | NA |
| Freq. of SM use | Vandenbosch 2016 | Freq. of chat room use | Self-report 1-item measure: 1-How often usually visited chat rooms. Responses: (1) never to (8) all day long.  3 category variable constructed: non-users, infrequent (less than once a month, and frequent (monthly to daily). |  | Ordinal | Current | NR | NR | SNS  (active use) | NA |

| Exposure | Author and year | Exposure definition | Exposure ascertainment | Validated/  objectively recorded | Exposure measure  type | Exposure time period | SM platform | SM platform | SM category  (active/  passive use) | SM content |
| --- | --- | --- | --- | --- | --- | --- | --- | --- | --- | --- |
| Freq. of SM use | Vannucci 2019 | Freq. of SM use | Self-report measure via Technology Use Questionnaire: 1-How often used a range of SM platforms on a typical day (discussion boards, FB, Google +, Instagram, Pin Boards, Snapchat, Tumblr, Twitter).  Responses: (0) never to (8) almost constantly.  Total number of platforms used calculated by coding each platform as either (0) never used and (1) used at least once or more, and then summing usage scores.  2-How much time, overall, they spent using SM platforms on a typical day (in hrs).  Latent profile analyses used to identify latent subgroups of SM: high SM use (frequent daily overall use across platforms), high Instagram/Snapchat use (hourly use of Instagram + Snapchat use only, with low use of all other SM platforms), and low SM use (less than once daily use of all SM platforms). |  | Categorical | Current | Examples: Discussion boards, FB, Google +, Instagram, Pin Boards, Snapchat, Tumblr, and Twitter | Mixed platforms | General SM  (unclear) | NA |
| Freq. of SM use | Vannucci 2019 | Freq. of Google+ use | Self-report 1-item measure: 1-How often used Google + on a typical day.  Responses: never, less than once a week, once a week, several times a week, once a day, several times a day, once an hr, several times an hr, and almost constantly. |  | Continuous | Current | Specified: Google + | Google+ | SNS  (unclear) | NA |
| Freq. of SM use | Vannucci 2019 | Freq. of use of pin boards | Self-report 1-item measure: 1-How often used pin boards on a typical day.  Responses: never, less than once a week, once a week, several times a week, once a day, several times a day, once an hr, several times an hr, and almost constantly. |  | Continuous | Current | Specified: Pin board | Discussion & Pin Boards | Blogs & Forums  (unclear) | NA |
| Freq. of SM use | Vannucci 2019 | Freq. of Facebook use | Self-report 1-item measure: 1-How often used Facebook on a typical day.  Responses: never, less than once a week, once a week, several times a week, once a day, several times a day, once an hr, several times an hr, and almost constantly. |  | Continuous | Current | Specified: Facebook | Facebook | SNS  (unclear) | NA |
| Freq. of SM use | Vannucci 2019 | Freq. of Instagram use | Self-report 1-item measure: 1- How often used Instagram on a typical day.  Responses: never, less than once a week, once a week, several times a week, once a day, several times a day, once an hr, several times an hr, and almost constantly. |  | Continuous | Current | Specified: Instagram | Instagram | SNS  (unclear) | NA |
| Freq. of SM use | Vannucci 2019 | Freq. of Snapchat use | Self-report 1-item measure: 1-How often used Snapchat on a typical day.  Responses: never, less than once a week, once a week, several times a week, once a day, several times a day, once an hr, several times an hr, and almost constantly. |  | Continuous | Current | Specified: Snapchat | Snapchat | SNS  (unclear) | NA |

| Exposure | Author and year | Exposure definition | Exposure ascertainment | Validated/  objectively recorded | Exposure measure  type | Exposure time period | SM platform | SM platform | SM category  (active/  passive use) | SM content |
| --- | --- | --- | --- | --- | --- | --- | --- | --- | --- | --- |
| Freq. of SM use | Vannucci 2019 | Freq. of Twitter use | Self-report 1-item measure: 1-How often used Twitter on a typical day.  Responses: never, less than once a week, once a week, several times a week, once a day, several times a day, once an hr, several times an hr, and almost constantly. |  | Continuous | Current | Specified: Twitter | Twitter | Micro-blogging  (unclear) | NA |
| Freq. of SM use | Vannucci 2019 | Freq. of Tumblr use | Self-report 1-item measure: 1-How often used Tumblr on a typical day.  Responses: never, less than once a week, once a week, several times a week, once a day, several times a day, once an hr, several times an hr, and almost constantly. |  | Continuous | Current | Specified: Tumblr | Tumblr | Micro-blogging  (unclear) | NA |
| Freq. of SM use | Vannucci 2019 | Freq. of use of discussion boards | Self-report 1-item measure: 1-How often used discussion boards on a typical day.  Responses: never, less than once a week, once a week, several times a week, once a day, several times a day, once an hr, several times an hr, and almost constantly. |  | Continuous | Current | Specified: Discussion boards | Discussion & Pin Boards | Blogs & forums  (unclear) | NA |
| Freq. of SM use | Vazquez-Nava 2020 | Presence of use of social networks (WhatsApp/  Facebook) | Self- report measure via validated questionnaire: 1-Use of online messaging platforms such as WhatsApp or Facebook to communicate with their friends and other people. | Yes | Binary | Current | Specified: WhatsApp and Facebook | Mixed platforms | SNS  (active use) | NA |
| Freq. of SM use | Vente 2020 | Any SM per day | Self-report measure:  Total time spent on SM per day, and number and type of SM application used. |  | Binary | Current | NR | NR | General SM  (unclear) | NA |
| Freq. of SM use | Vente 2020 | Use of ≥4 SM applications per day | Self-report measure:  Total time spent on SM per day, and number and type of SM application used. |  | Binary | Current | NR | NR | General SM  (unclear) | NA |
| Freq. of SM use | Wana 2019 | Presence of SM use | Self-report measure:  If SM user, what platforms used, frequency of SM use, and the purpose of using SM. |  | Binary | Current | Examples: Facebook, Viber, WhatsApp, YouTube, and Instagram | Mixed platforms | SNS  (unclear) | NA |
| Freq. of SM use | Ward 2022 | Freq. of Facebook use per day | Self-report 1-item measure:  1-How many times per day Facebook checked.  Responses: 0 to 7+ times. |  | Continuous | Current | Specified: Facebook | Facebook | SNS  (unclear) | NA |
| Freq. of SM use | Ward 2022 | Freq. of Snapchat use per day | Self-report 1-item measure:  1-How many times per day Snapchat checked.  Responses: 0 to 7+ times. |  | Continuous | Current | Specified: Snapchat | Snapchat | SNS  (unclear) | NA |
| Freq. of SM use | Ward 2022 | Freq. of Instagram use per day | Self-report 1-item measure:  1-How many times per day Instagram checked.  Responses: 0 to 7+ times. |  | Continuous | Current | Specified: Instagram | Instagram | Media-sharing  (unclear) | NA |
| Freq. of SM use | Whitehill 2020 | Freq. of SM use | Self-report 1-item measure: Responses: never, monthly, a few times a month, weekly, a few times a week, once a day, and more than once a day. No further information provided. |  | Ordinal | Current | NR | NR | General SM  (unclear) | NA |

| Exposure | Author and year | Exposure definition | Exposure ascertainment | Validated/  objectively recorded | Exposure measure  type | Exposure time period | SM platform | SM platform | SM category (active/  passive use) | SM content |
| --- | --- | --- | --- | --- | --- | --- | --- | --- | --- | --- |
| Freq. of SM use | Whitehill 2020 | Presence of Facebook use | No information reported. |  | Binary | Current | Specified: Facebook | Facebook | SNS  (unclear) | NA |
| Freq. of SM use | Whitehill 2020 | Presence of Twitter use | No information reported. |  | Binary | Current | Specified: Twitter | Twitter | Micro-blogging  (unclear) | NA |
| Freq. of SM use | Whitehill 2020 | Presence of Instagram use | No information reported. |  | Binary | Current | Specified: Instagram | Instagram | Media-sharing  (unclear) | NA |
| Freq. of SM use | Widman 2014 | Used technology based sexual communication to communicate with dating partners about using condoms | Self-report 1-item measure: 1-Ever used private technology (i.e., “electronically interacting with someone in a way that is not visible to the public, such as Snapchat, or private Facebook messaging”) to communicate with dating partners about using condoms. Dating partners defined as a boy/girlfriend or someone with whom participants had a romantic or sexual relationship. |  | Binary | Ever | Examples: texting, Snapchat, and Facebook | Mixed platforms | SNS  (active use) | NA |
| Freq. of SM use | Widman 2014 | Used technology based sexual communication to communicate with dating partners about using other forms of birth control | Self-report 1-item measure: 1-Ever used private technology (i.e., “electronically interacting with someone in a way that is not visible to the public, such as Snapchat, or private Facebook messaging”) to communicate with dating partners about using other forms of birth control. Dating partners defined as a boy/girlfriend or someone with whom participants had a romantic or sexual relationship. |  | Binary | Ever | Examples: texting, Snapchat, and Facebook | Mixed platforms | SNS  (active use) | NA |
| Freq. of SM use | Wulff 2021 | Freq. of WhatsApp use | Self-report 1-item measure:  1-Frequency of social network use (e.g., Twitter, Facebook) and used functions (e.g., texting, view pictures, posting content)  Responses: never, at least once a month, at least once a week, daily, more than 1 hr/day. |  | Binary | Current | Specified: WhatsApp | WhatsApp | SNS  (active and passive use) | NA |
| Freq. of SM use | Wulff 2021 | Freq. of YouTube use | Self-report 1-item measure:  1-Frequency of social network use (e.g., Twitter, Facebook) and used functions (e.g., texting, view pictures, posting content)  Responses: never, at least once a month, at least once a week, daily, more than 1 hr/day. |  | Binary | Current | Specified: YouTube | YouTube | Media-sharing  (active and passive use) | NA |
| Freq. of SM use | Wulff 2021 | Freq. of Instagram use | Self-report 1-item measure:  1-Frequency of social network use (e.g., Twitter, Facebook) and used functions (e.g., texting, view pictures, posting content)  Responses: never, at least once a month, at least once a week, daily, more than 1 hr/day. |  | Binary | Current | Specified: Instagram | Instagram | Media-sharing  (active and passive use) | NA |

| Exposure | Author and year | Exposure definition | Exposure ascertainment | Validated/  objectively recorded | Exposure measure  type | Exposure time period | SM platform | SM platform | SM category  (active/  passive use) | SM content |
| --- | --- | --- | --- | --- | --- | --- | --- | --- | --- | --- |
| Freq. of SM use | Wulff 2021 | Freq. of Facebook use | Self-report 1-item measure:  1-Frequency of social network use (e.g., Twitter, Facebook) and used functions (e.g., texting, view pictures, posting content)  Responses: never, at least once a month, at least once a week, daily, more than 1 hr/day. |  | Binary | Current | Specified: Facebook | Facebook | SNS  (active and passive use) | NA |
| Freq. of SM use | Wulff 2021 | Freq. of Google+ use | Self-report 1-item measure:  1-Frequency of social network use (e.g., Twitter, Facebook) and used functions (e.g., texting, view pictures, posting content)  Responses: never, at least once a month, at least once a week, daily, more than 1 hr/day. |  | Binary | Current | Specified: Google + | Google+ | SNS  (active and passive use) | NA |
| Freq. of SM use | Wulff 2021 | Freq. of Twitter use | Self-report 1-item measure:  1-Frequency of social network use (e.g., Twitter, Facebook) and used functions (e.g., texting, view pictures, posting content)  Responses: never, at least once a month, at least once a week, daily, more than 1 hr/day. |  | Binary | Current | Specified: Twitter | Twitter | Micro-blogging  (active and passive use) | NA |
| Freq. of SM use | Widman 2014 | Used technology based sexual communication to communicate with dating partners about HIV/AIDS | Self-report 1-item measure: 1-Ever used private technology (i.e., “electronically interacting with someone in a way that is not visible to the public, such as Snapchat, or private Facebook messaging”) to communicate with dating partners about HIV/AIDS. Dating partners defined as a boy/girlfriend or someone with whom participants had a romantic or sexual relationship. |  | Binary | Ever | Examples: texting, Snapchat, and Facebook | Mixed platforms | SNS  (active use) | NA |
| Freq. of SM use | Widman 2014 | Used technology based sexual communication to communicate with dating partners about STIs | Self-report 1-item measure: 1-Ever used private technology (i.e., “electronically interacting with someone in a way that is not visible to the public, such as Snapchat, or private Facebook messaging”) to communicate with dating partners about STIs. Dating partners defined as a boy/girlfriend or someone with whom participants had a romantic or sexual relationship. |  | Binary | Ever | Examples: texting, Snapchat, and Facebook | Mixed platforms | SNS  (active use) | NA |
| Freq. of SM use | Widman 2014 | Used technology based sexual communication to communicate with dating partners about risk of pregnancy | Self-report 1-item measure: 1-Ever used private technology (i.e., “electronically interacting with someone in a way that is not visible to the public, such as Snapchat, or private Facebook messaging”) to communicate with dating partners about risk of pregnancy. Dating partners defined as a boy/girlfriend or someone with whom participants had a romantic or sexual relationship. |  | Binary | Ever | Examples: texting, Snapchat, and Facebook | Mixed platforms | SNS  (active use) | NA |

| Exposure | Author and year | Exposure definition | Exposure ascertainment | Validated/  objectively recorded | Exposure measure  type | Exposure time period | SM platform | SM platform | SM category  (active/  passive use) | SM content |
| --- | --- | --- | --- | --- | --- | --- | --- | --- | --- | --- |
| Freq. of SM use | Widman 2014 | Used technology based sexual communication to communicate with dating partners about sexual limits | Self-report 1-item measure: 1-Ever used private technology (i.e., “electronically interacting with someone in a way that is not visible to the public, such as Snapchat, or private Facebook messaging”) to communicate with dating partners about sexual limits. Dating partners defined as a boy/girlfriend or someone with whom participants had a romantic or sexual relationship. |  | Binary | Ever | Examples: texting, Snapchat, and Facebook | Mixed platforms | SNS  (active use) | NA |
| Exposure to health-risk behaviour content | Baldwin 2018 | Watched food/  beverage brand YouTube videos | Self-report 1-item measure: 1-Ever watched any commercials/ads for food or drink products on YouTube. Responses: yes/no. |  | Binary | Ever | Specified: YouTube | YouTube | Media-sharing  (passive use) | Marketer-gen |
| Exposure to health-risk behaviour content | Baldwin 2018 | Seen favourite food advertised on SM | Self-report 1-item measure: 1-Had seen favourite food brands advertised on SM. Responses: yes/no. |  | Binary | NR | NR | NR | General SM  (passive use) | Marketer-gen |
| Exposure to health-risk behaviour content | Baldwin 2018 | Liked a food/  beverage brand on Facebook | Self-report 1-item measure: 1-Ever liked any food/beverage companies or brands on Facebook (e.g., they liked or shared any of these pages’ content). Responses: yes/no. |  | Binary | Ever | Specified: Facebook | Facebook | SNS  (active use) | Marketer-gen |
| Exposure to health-risk behaviour content | Baldwin 2018 | Entered a food/  beverage brand competition on Facebook | Self-report 1-item measure: 1-Ever entered a food/beverage brand competition/contest on Facebook. Responses: yes/no. |  | Binary | Ever | Specified: Facebook | Facebook | SNS  (active use) | Marketer-gen |
| Exposure to health-risk behaviour content | Bayraktar 2007 | Online gaming: murdering games | Self-report measure: Completed a set of questionnaires related to internet experience, internet sites used (chatting sites, music sites, popstar sites, played games online etc), and reasons for internet usage etc.  No further information provided. |  | Continuous | Current | NR | NR | Online Gaming  (active use) | Marketer-gen |
| Exposure to health-risk behaviour content | Bayraktar 2007 | Online gaming: fighting games | Self-report measure: Completed a set of questionnaires related to internet experience, internet sites used (chatting sites, music sites, popstar sites, played games online etc), and reasons for internet usage etc.  No further information provided. |  | Continuous | Current | NR | NR | Online Gaming  (active use) | Marketer-gen |
| Exposure to health-risk behaviour content | Bayraktar 2007 | Online gaming: bombing games | Self-report measure: Completed a set of questionnaires related to internet experience, internet sites used (chatting sites, music sites, popstar sites, played games online etc), and reasons for internet usage etc.  No further information provided. |  | Continuous | Current | NR | NR | Online Gaming  (active use) | Marketer-gen |

| Exposure | Author and year | Exposure definition | Exposure ascertainment | Validated/  objectively recorded | Exposure measure  type | Exposure time period | SM platform | SM platform | SM category  (active/  passive use) | SM content |
| --- | --- | --- | --- | --- | --- | --- | --- | --- | --- | --- |
| Exposure to health-risk behaviour content | Camenga 2018 | Exposure to  e-cigarette advertisements on Facebook | Self-report 1-item measure: 1-Recently seen advertisements on Facebook. Responses: yes/no. |  | Binary | Current | Specified: Facebook | Facebook | SNS  (passive use) | Marketer-gen |
| Exposure to health-risk behaviour content | Camenga 2018 | Exposure to  e-cigarette advertisements on Twitter | Self-report 1-item measure 1-Recently seen advertisements on Twitter. Responses: yes/no. |  | Binary | Current | Specified: Twitter | Twitter | Micro-blogging  (passive use) | Marketer-gen |
| Exposure to health-risk behaviour content | Camenga 2018 | Exposure to  e-cigarette advertisements on YouTube | Self-report 1-item measure 1-Recently seen advertisements on YouTube. Responses: yes/no. |  | Binary | Current | Specified: YouTube | YouTube | Media-sharing  (passive use) | Marketer-gen |
| Exposure to health-risk behaviour content | Camenga 2018 | Exposure to  e-cigarette advertisements on Pinterest/  Google + | Self-report 1-item measure 1-Recently seen advertisements on Pinterest/Google +. Responses: yes/no. |  | Binary | Current | Specified: Google + and Pinterest | Mixed platforms | Media-sharing  (passive use) | Marketer-gen |
| Exposure to health-risk behaviour content | Cavazos-Rehg 2014 | Exposure to tobacco ads/  promotions via Facebook or  Myspace | Self-report 2-item measure: If during the past 30 days had: 1-received coupons from a tobacco company through…; 2-received ads from a tobacco company through…. Participants could select one or more responses from the following choices: the mail, E-mail, the Internet, Facebook, Myspace, a text message.  Responses for Facebook and Myspace combined to represent variable. |  | Binary | Past month | Specified: Facebook and Myspace | Facebook & Myspace | SNS  (unclear) | Marketer-gen |
| Exposure to health-risk behaviour content | Chen 2019 | Exposure to risky selfie descriptive norms | Self-report 1-item measure: 1-How many of friends posted risky selfies. Responses: (1) nobody to (5) everybody. |  | Continuous | Current | NR | NR | General SM  (passive use) | User-gen |
| Exposure to health-risk behaviour content | Coates 2019 | Exposure to unhealthy mock Instagram influencer marketing | Exposed to mock Instagram profiles for 2 SM influencers (male and female). Profiles consisted of the Instagram banner and 6 images (3 test and 3 filler unbranded non-food items) of influencer holding a product (unhealthy e.g., choc cookies; healthy e.g., banana; branded non-food e.g., sneakers). Images obtained and edited from influencer YouTube channels.  Participant randomly exposed to 1 of 3 mock Instagram profiles of an influencer holding a product: unhealthy snacks, healthy snacks, or branded non-food items.  Counterbalancing of participants to condition, and influencer order (man first or woman first), was conducted by using randomizer.org. | Yes | Categorical | Current | Specified: Instagram | Instagram | SNS  (passive use) | Marketer-gen |

| Exposure | Author and year | Exposure definition | Exposure ascertainment | Validated/  objectively recorded | Exposure measure  type | Exposure time period | SM platform | SM platform | SM category (active/  passive use) | SM content |
| --- | --- | --- | --- | --- | --- | --- | --- | --- | --- | --- |
| Exposure to health-risk behaviour content | Critchlow 2019 | Participation with alcohol marketing on SM | Self-report 5-item measure: If had: 1-liked an alcohol brand on SM, such as Twitter, Facebook or Instagram;  2-shared something related to an alcohol drinks brand, such as a status, Tweet, or picture;  3-followed an alcohol brand on social media;  4-entered a competition run by an alcoholic drink brand online or on social media; and  5-searched for alcoholic drinks adverts on websites, such as YouTube.  Responses: yes/no/none of the above. A cumulative score was computed (0–5).  3 category variable constructed: no participation with any marketing, participation with 1 form of marketing, or participation with ≥2 forms of marketing. |  | Ordinal | Past month | Examples: Twitter, Facebook, Instagram, and SM | Mixed platforms | General SM  (active use) | Marketer-gen |
| Exposure to health-risk behaviour content | Critchlow 2019 | Participation with user-created alcohol promotion on SM | Self-report 1-item measure: 1-Updated status or uploaded pictures of themselves or friends drinking an alcoholic drink.  Responses: yes/no. |  | Binary | Past month | NR | NR | General SM  (active use) | User-gen |
| Exposure to health-risk behaviour content | Dai 2022 | Exposure to e-cigarette advertisements on SM | Self-report 1-item validated measure:  1-Seen e-cigarette advertisements on SM in the past 30 days. Responses: never, sometimes (once or twice), and often (more than 3 times).  Variable dichotomised: yes (sometimes/often)/no (never). | Yes | Binary | Past month | NR | NR | General SM  (passive use) | Marketer-gen |
| Exposure to health-risk behaviour content | Davis 2019 | Substance-related media exposure via SM | Self-report 2-item measure: 1,2-How often saw or heard pictures or comments on a SNS (e.g., Facebook) showing someone or talking about someone who is drunk. Responses: (0) not at all to (6) every day. Average of 2 items taken for analysis. |  | Continuous | Past 3 months | Example: Facebook | Facebook | SNS  (passive use) | User + Marketer-generated |
| Exposure to health-risk behaviour content | Dawson 2019 | % of participant posts sharing inappropriate content on Facebook | Observationally coded measure coded using Mikami and Szwedo's Facebook Coding Manual.^157^  Facebook profile coded over 2-month period to obtain proportion of total participant posts shared containing inappropriate content (i.e., profanity, substance use, sexual behaviour, violence, or other illegal behaviour). | Yes | Continuous | Current | Specified: Facebook | Facebook | SNS  (active use) | User-gen |
| Exposure to health-risk behaviour content | Dawson 2019 | % of participant posts containing relational aggression on Facebook | Observationally coded measure coded using Mikami and Szwedo's Facebook Coding Manual.^157^ Facebook profile coded over 2-month period to obtain proportion of total participant posts containing relational aggression (i.e., comments meant to criticize, ostracize, or embarrass a person or group of people). | Yes | Continuous | Current | Specified: Facebook | Facebook | SNS  (active use) | User-gen |

| Exposure | Author and year | Exposure definition | Exposure ascertainment | Validated/  objectively recorded | Exposure measure  type | Exposure time period | SM platform | SM platform | SM category (active/  passive use) | SM content |
| --- | --- | --- | --- | --- | --- | --- | --- | --- | --- | --- |
| Exposure to health-risk behaviour content | Dawson 2019 | % of participant friend posts containing relational aggression on Facebook | Observationally coded measure coded using Mikami and Szwedo's Facebook Coding Manual.^157^ Facebook profile coded over 2-month period to obtain proportion of friend posts on participant timeline containing content on relational aggression (i.e., comments meant to criticize, ostracise, or embarrass a person or group of people). | Yes | Continuous | Current | Specified: Facebook | Facebook | SNS  (passive use) | User-gen |
| Exposure to health-risk behaviour content | Dawson 2019 | % of participant friend posts containing inappropriate content | Observationally coded measure coded using Mikami and Szwedo's Facebook Coding Manual.^157^ Facebook profile coded over 2-month period to obtain proportion of friend posts on participant timeline containing inappropriate content (i.e., profanity, substance use, sexual behaviour, violence, or other illegal behaviour). | Yes | Continuous | Current | Specified: Facebook | Facebook | SNS  (passive use) | User-gen |
| Exposure to health-risk behaviour content | de Bruijn 2016 | Ever used alcohol branded SM page | Self-report 1-item measure:  1-Ever used a profile page on sites such as Hyves, Facebook, MSN, or Myspace containing an alcohol brand or logo.  Responses: never, rarely/sometimes, and often/very often. |  | Binary | Ever | Examples: Hyves, Facebook, MSN, and Myspace | Mixed platforms | SNS  (passive use) | Marketer-gen |
| Exposure to health-risk behaviour content | De Jans 2021 | Exposure to snack with low nutritional value (mini donut) on Instagram | Exposed to 1 of 2 individual Instagram posts:  1-Post portraying snack high in nutritional value (i.e., strawberries).  2-Post portraying snack low in nutritional value (i.e., donuts) | Yes | Binary | Current | Specified: Instagram | Instagram | Media-sharing  (passive use) | Marketer-gen |
| Exposure to health-risk behaviour content | Doornwaard 2014 | Exposure to displays of sexual references on Facebook | Observationally coded measure using codebook based on procedures used in previous content analyses of SNS.^158^  Reviewers analysed visible elements on participant Facebook timeline via content analysis: status updates, images, comments and downloaded icons to investigate sexual references on Facebook related to safe sex, risky sex, sexual behaviour, sexualised personal descriptions, revealing personal images, sexual paraphernalia, and romance.  Sexual references defined as any textual or visual depiction of sexual activity or sexually suggestive behaviour. References did not need to be created by the owner (participant), they included posts, comments, tags by friends on the participant’s profile.  Sexual reference displayers compared with non-sexual reference displayers. | Yes | Binary | During coding period | Specified: Facebook | Facebook | SNS  (active and passive use) | User-gen |

| Exposure | Author and year | Exposure definition | Exposure ascertainment | Validated/  objectively recorded | Exposure measure  type | Exposure time period | SM platform | SM platform | SM category (active/  passive use) | SM content |
| --- | --- | --- | --- | --- | --- | --- | --- | --- | --- | --- |
| Exposure to health-risk behaviour content | Folkvord 2020 | Exposure to manipulated popular influencer Instagram post showing energy dense foods | Exposed to 1 of 2 popular SM influencers on Instagram.  1-Post showing vegetables (control condition)  2-Post showing energy dense snacks (experimental condition)  3-Post showing non-food products | Yes | Binary | Current | Specified: Instagram | Instagram | Media-sharing  (passive use) | Marketer-gen |
| Exposure to health-risk behaviour content | Gascoyne 2021 | Seen an advertisement for a food or drink product on SM (e. g. Facebook, Instagram) | Self-report 1-item measure:  1-How often in last month saw a food or drink product on SM (e. g. Facebook, Instagram).  Responses: not in the last month, 1–3 times a month, 1–3 times a week and daily or almost daily. |  | Ordinal | Past month | Examples: Facebook and Instagram | Mixed platforms | General SM  (passive use) | Marketer-gen |
| Exposure to health-risk behaviour content | Gascoyne 2021 | Liked/ shared posts related to a food or drink product or brand (e.g., soft drink, fast food) | Self-report 1-item measure:  1-How often in last month ‘liked’ or ‘shared’ posts related to a food or drink product or brand (e.g., soft drink, fast food).  Responses: not in the last month, 1–3 times a month, 1–3 times a week and daily or almost daily. |  | Ordinal | Past month | Examples: Facebook and Instagram | Mixed platforms | General SM  (active use) | Marketer-gen |
| Exposure to health-risk behaviour content | Gerber 2021 | Exposure to alcohol related content on Instagram and Snapchat | Self-report 2-item measure:  1-How often see content in which alcohol is present on Instagram.  2-How often see content in which alcohol is present on Snapchat.  Responses: (1) never to (5) very often.  Mean of both items calculated, representing exposure to alcohol-related content on SM. |  | Continuous | Current | Specified: Instagram and Snapchat | Mixed platforms | Media-sharing  (passive use) | User + Marketer-generated |
| Exposure to health-risk behaviour content | Gerber 2021 | Shared alcohol related content on Instagram and Snapchat | Self-report 2-item measure:  1-How often post or send content on Instagram depicting alcohol.  2-How often post or send content on Snapchat depicting alcohol.  Responses: (1) never to (5) very often.  Mean of both items calculated, representing exposure to alcohol-related content on SM. |  | Continuous | Current | Specified: Instagram and Snapchat | Mixed platforms | Media-sharing  (active use) | User + Marketer-generated |

| Exposure | Author and year | Exposure definition | Exposure ascertainment | Validated/  objectively recorded | Exposure measure  type | Exposure time period | SM platform | SM platform | SM category  (active/  passive use) | SM content |
| --- | --- | --- | --- | --- | --- | --- | --- | --- | --- | --- |
| Exposure to health-risk behaviour content | Geusens 2017 | Frequency of sharing alcohol references on SNS | Self-report 8-item measure: How often privately shared the following things on any social medium and how often publicly shared the following things on any social medium: 1- photos or video clips referring to alcohol use; 2- textual updates referring to alcohol use; 3- photos or video clips in which they or their friends were drunk, and; 4- textual updates while they were drunk.  ‘Private sharing’ was defined as ‘references shared with a limited amount of people, e.g., through communication via direct messaging, private groups, or group chats’.  Responses: (0) never to (6) several times a day. |  | Continuous | Ever | NR | NR | SNS  (active use) | User-gen |
| Exposure to health-risk behaviour content | Geusens 2017 | Perceived number of friends sharing alcohol references online | Self-report 4-item measure:  How many of their friends shared:  1-photos or video clips referring to alcohol use;  2-textual updates referring to alcohol use;  3-photos or video clips in which they are drunk, or;  4-textual updates while they are drunk.  Responses: (0) none to (4) all of them. |  | Continuous | Current | NR | NR | SNS  (passive use) | User-gen |
| Exposure to health-risk behaviour content | Geusens 2019 | Frequency of exposure to peer alcohol references on SNS | Self-report 3-item measure: How often saw videos or images: 1-on YouTube or similar sites or;  2-on other SNS, such as Facebook or Twitter, about youth drinking alcohol and; 3-youth being drunk.  Responses: (0) never to (8) all day long.  Factor analysis used to load all 3 items loaded onto one scale. |  | Continuous | Current | Examples: YouTube, Facebook, Twitter, and other SNS | Mixed platforms | SNS  (passive use) | User-gen |
| Exposure to health-risk behaviour content | Geusens 2019 | Frequency of sharing of alcohol references on SNS | Self-report 1-item measure: How often shared videos/images: 1-on YouTube or similar sites or;  2-on other SNS, such as Facebook or Twitter, about youth drinking alcohol and;  3- youth being drunk.  Responses: (0) never to (8) all day long.  Upper scale points collapsed after calculating the composite score with 0 (never), 1 (a few times per year), 2 (once per month), and 3 (more than once per month). |  | Continuous | Current | Examples: YouTube, Facebook, Twitter, and other SNS | Mixed platforms | SNS  (active use) | User-gen |

| Exposure | Author and year | Exposure definition | Exposure ascertainment | Validated/  objectively recorded | Exposure measure  type | Exposure time period | SM platform | SM platform | SM category  (active/  passive use) | SM content |
| --- | --- | --- | --- | --- | --- | --- | --- | --- | --- | --- |
| Exposure to health-risk behaviour content | Gordon 2011 | Awareness of alcohol marketing on SNS | Self-report 1-item measure: 1-Had seen any alcohol marketing on SNS. Responses: yes/no/don't know. |  | Binary | Current | NR | NR | SNS  (passive use) | Marketer-gen |
| Exposure to health-risk behaviour content | Gordon 2011 | Used SNS containing alcohol brands or logos | Self-report 1-item measure: 1-Has used SNS containing alcohol brands or logos. Responses: yes/no/don't know. |  | Binary | Current | NR | NR | SNS  (passive use) | Marketer-gen |
| Exposure to health-risk behaviour content | Hrywna 2020 | Liked or followed a tobacco brand on SM | Self-report 1-item measure: 1- Had visited, followed, liked, or become a fan of a tobacco brand on sites like Instagram, Twitter, Facebook, or YouTube. |  | Binary | Past year | Examples: Instagram, Twitter, Facebook, and YouTube | Mixed platforms | SNS  (active and passive use) | Marketer-gen |
| Exposure to health-risk behaviour content | Huang 2014 | Number of friends who posted risky pictures partying or drinking | Self-report measure: Egocentric (personal) networks created for each participant, participant asked to name 7 best friends regardless of where they live or go to school and provide basic information about each of them (alters).  Friends’ online behaviours were assessed by asking whether alters ever “posted pictures of themselves partying or drinking alcohol online”  Indicator friends’ online risk behaviour created using total number of alters for these items. |  | Continuous | Ever | NR | NR | SNS  (passive use) | User-gen |
| Exposure to health-risk behaviour content | Lin 2012 | Used SNS containing alcohol brands or logos | Self-report 1-item measure: 1-Used SNS containing alcohol brands or logos. Responses: yes/no/don't know. |  | Binary | Current | NR | NR | SNS  (passive use) | Marketer-gen |
| Exposure to health-risk behaviour content | Lin 2012 | Awareness of alcohol marketing  on SNS | Self-report measure: If they could think of any makes or brands of alcohol that they had seen or heard advertised recently. Followed by a set of questions assessing awareness across 15 types of marketing channels (including SNS). Response: yes/no/don't know. |  | Binary | Current | NR | NR | SNS  (passive use) | Marketer-gen |
| Exposure to health-risk behaviour content | Nesi 2017 | Ever exposed to friends' SNS alcohol content | Self-report 2-item measure:  Whether a friend had ever:  1-posted a picture of themselves with alcohol, or;  2-posted a status, picture, or link about drinking alcohol.  Items combined, with endorsement of either item coded as (1) and endorsement of neither as (0). |  | Binary | Ever | NR | NR | SNS  (passive use) | User-gen |

| Exposure | Author and year | Exposure definition | Exposure ascertainment | Validated/  objectively recorded | Exposure measure  type | Exposure time period | SM platform | SM platform | SM category  (active/  passive use) | SM content |
| --- | --- | --- | --- | --- | --- | --- | --- | --- | --- | --- |
| Exposure to health-risk behaviour content | Nesi 2017 | Exposure to SNS alcohol content posted by self | Self-report 5-item measure:  If had: 1-posted status, picture, or link about alcohol; 2-posted picture of self with alcohol;  3-tagged friends in photos with alcohol;  4-posted picture of self, passed out or vomiting as result of alcohol;  5-posted picture of friend passed out or vomiting as a result of alcohol. |  | Binary | Ever | NR | NR | SNS  (active use) | User-gen |
| Exposure to health-risk behaviour content | Ngqangashe 2021 | Watched YouTube Tasty video portraying preparation of sweet snacks | Exposed to 1 of 2 short-form ‘Tasty’ culinary videos on YouTube.  1-Video portraying sweet snacks (The Best Fudgy Brownies Ever, Chocolate Peanut Brownies (Buckeye Brownies), and 6 Ways to Make Better Boxed Brownies.  2-Video portraying fruits and vegetables (4 Make-Ahead Vegetable-Packed Smoothies, 4 Healthier Desserts, and Fruit Salad Four Ways). | Yes | Binary | Current | Specified: YouTube | YouTube | Media-sharing  (passive use) | Marketer-gen |
| Exposure to health-risk behaviour content | Pegg 2018 | SNS alcohol exposure | Self-report 1-item measure: 1-How often in the previous 6 months friends posted pictures, updates, or wall posts that showed or talked about them drinking alcohol.  Responses: (1) none to (8) ≥31 times. |  | Continuous | Past 6 months | NR | NR | SNS  (active and passive use) | User-gen |
| Exposure to health-risk behaviour content | Pérez 2022 | Exposure to tobacco related content on SM in the past 12 months (including e-cigarettes) | Self-report 1-item measure:  1-In past 12 months, posted content about tobacco products (including e-cigarettes) on any SM sites.  Responses: yes/no. |  | Binary | Past year | NR | NR | General SM  (active use) | User + Marketer-generated |
| Exposure to health-risk behaviour content | Pérez 2022 | Posted tobacco related content on SM in the past 12 months (including e-cigarettes) | Self-report 1-item measure:  1-In past 12 months, seen content posted about tobacco products (including e-cigarettes) on SM sites.  Responses: yes/no. |  | Binary | Past year | NR | NR | General SM  (passive use) | User + Marketer-generated |
| Exposure to health-risk behaviour content | Qutteina 2022 | Exposure to non-core foods (energy dense, low nutrient: sweetened drinks, sweets, salty/savoury snacks) on SM | Self-report measure:  1-How often saw food messages posted by friends, influencers, and celebrities as well as messages posted by brands.  Responses: (1) not at all to (5) very often. |  | Ordinal | Current | NR | NR | General SM  (passive use) | User + Marketer-generated |

| Exposure | Author and year | Exposure definition | Exposure ascertainment | Validated/  objectively recorded | Exposure measure  type | Exposure time period | SM platform | SM platform | SM category (active/  passive use) | SM content |
| --- | --- | --- | --- | --- | --- | --- | --- | --- | --- | --- |
| Exposure to health-risk behaviour content | Qutteina 2022 | Exposure to branded non-core foods (energy dense, low nutrient: sweetened drinks, sweets, salty/savoury snacks) on SM | Self-report measure:  1-How often saw food messages posted by friends, influencers, and celebrities as well as messages posted by brands.  Responses: (1) not at all to (5) very often. |  | Ordinal | Current | NR | NR | General SM  (passive use) | User + Marketer-generated |
| Exposure to health-risk behaviour content | Roditis 2016 | Ever seen a message posted on SM about the benefits or good things of using marijuana | Self-report 1-item measure: 1-Ever seen a message posted on SM about benefits or good things related to using marijuana. Responses: yes/no. |  | Binary | Ever | NR | NR | General SM  (passive use) | User + Marketer-generated |
| Exposure to health-risk behaviour content | Shan 2022 | Followed tobacco brands on Facebook/Twitter or other SM sites | Self-report 1-item measure:  1-In past 12 months, liked or followed any of the following brands (e.g., Marlboro, Newport, American Spirit, Vuse) on Facebook, Twitter, or other SM sites.  Responses; yes/no |  | Binary | Past year | Examples: Facebook and Twitter | Mixed platforms | General SM  (active use) | Marketer-gen |
| Exposure to health-risk behaviour content | Sharma 2021 | Exposure to tobacco adverts on SM | Self-report measure:  Modified Global Youth Tobacco Survey (GYTS) questionnaire.  Responses: yes/no. No further information reported. |  | Binary | Current | NR | NR | General SM  (passive use) | Marketer-gen |
| Exposure to health-risk behaviour content | Smout 2021 | Exposure to peer-generated content on SM depicting risky substance use | Self-report 1-item measure:  1-See pictures of kids drunk, passed out or using drugs on these sites?  Responses: yes/no |  | Binary | Current | Specified: Facebook, Myspace, and other SNS | Mixed platforms | SNS  (passive use) | User-gen |
| Exposure to health-risk behaviour content | Trangenstein 2019 | Liked/follow cannabis business pages on Facebook, Twitter and/or Instagram | Self-report measure: 1-Engagement: if like/follow any cannabis business pages on Facebook, Instagram and/or Twitter and if so, which ones. Engagement defined as liking, following, and/or commenting on a cannabis business page.  Responses: yes/no. |  | Binary | Current | Specified: Facebook, Twitter, and Instagram | Mixed platforms | SNS  (active use) | Marketer-gen |
| Exposure to health-risk behaviour content | Trangenstein 2019 | Liked/follow cannabis business pages on Facebook | Self-report 1-item measure: 1-Engagement: if like/follow any cannabis business pages on Facebook. Engagement defined as liking, following, and/or commenting on a cannabis business page.  Responses: yes/no. |  | Binary | Current | Specified: Facebook | Facebook | SNS  (active use | Marketer-gen |

| Exposure | Author and year | Exposure definition | Exposure ascertainment | Validated/  objectively recorded | Exposure measure  type | Exposure time period | SM platform | SM platform | SM category  (active/  passive use) | SM content |
| --- | --- | --- | --- | --- | --- | --- | --- | --- | --- | --- |
| Exposure to health-risk behaviour content | Trangenstein 2019 | Liked/follow cannabis business pages on Twitter | Self-report 1-item measure: 1-Engagement: if like/follow any cannabis business pages on Twitter. Engagement defined as liking, following, and/or commenting on a cannabis business page.  Responses: yes/no. |  | Binary | Current | Specified: Twitter | Twitter | Micro-blogging  (active use) | Marketer-gen |
| Exposure to health-risk behaviour content | Trangenstein 2019 | Liked/follow cannabis business pages on Instagram | Self-report 1-item measure: 1-Engagement: if like/follow any cannabis business pages on Instagram. Engagement defined as liking, following, and/or commenting on a cannabis business page.  Responses: yes/no. |  | Binary | Current | Specified: Instagram | Instagram | Media-sharing  (active use) | Marketer-gen |
| Exposure to health-risk behaviour content | Yao 2022 | Exposure to content (including text and pictures) about drinking or smoking (e.g., saw drinking-related information) | Self-report 1-item measure:  1-How often exposed to content (including text and pictures) about drinking or smoking (e.g., saw drinking-related information) in three popular Chinese social media platforms—WeChat Moment, Qzone, and Weibo  Responses: (1) never to (5) always) with higher scores indicating higher frequency of SM exposure to tobacco and alcohol content |  | Continuous | Current | Specified: WeChat Moment, Qzone, and Weibo | Mixed Platforms | General SM  (passive use) | User + Marketer-generated |
| Exposure to health-risk behaviour content | Whitehill 2020 | Cumulative frequency of exposure to cannabis promotions (Facebook, Twitter, and Instagram) | Self-report 3-item measure: When using (1-Facebook, 2-Twitter, and 3-Instagram), how often saw ads or promotions for cannabis or related products. Responses: never, rarely, sometimes, most of the time, always, and do not use platform.  Variable dichotomised for each platform: rarely, sometimes, most of the time, and always/never and I do not use (1-Facebook, 2-Twitter, 3-Instagram). Binary variables summed to give counts of exposure across Facebook, Twitter and Instagram giving cumulative exposure. |  | Continuous | Current | Specified: Facebook, Twitter, and Instagram | Mixed platforms | SNS  (passive use) | Marketer-gen |
| Exposure to health-risk behaviour content | Whitehill 2020 | Frequency of exposure to cannabis promotions on Facebook | Self-report 1-item measure: 1-When using Facebook, how often saw ads or promotions for cannabis or related products. Responses: never, rarely, sometimes, most of the time, always, and do not use platform.  Variable dichotomised: rarely, sometimes, most of the time, and always/never and I do not use Facebook. |  | Binary | Current | Specified: Facebook | Facebook | SNS  (passive use) | Marketer-gen |

| Exposure | Author and year | Exposure definition | Exposure ascertainment | Validated/  objectively recorded | Exposure measure  type | Exposure time period | SM platform | SM platform | SM category  (active/  passive use) | SM content |
| --- | --- | --- | --- | --- | --- | --- | --- | --- | --- | --- |
| Exposure to health-risk behaviour content | Whitehill 2020 | Frequency of exposure to cannabis promotions on Twitter | Self-report 1-item measure: 1-When using Twitter, how often saw ads or promotions for cannabis or related products. Responses: never, rarely, sometimes, most of the time, always, and do not use platform.  Variable dichotomised: rarely, sometimes, most of the time, and always/ never and I do not use Twitter. |  | Binary | Current | Specified: Twitter | Twitter | Micro-blogging  (passive use) | Marketer-gen |
| Exposure to health-risk behaviour content | Whitehill 2020 | Frequency of exposure to cannabis promotions on Instagram | Self-report 1-item measure: 1-When using Instagram, how often saw ads or promotions for cannabis or related products. Responses: never, rarely, sometimes, most of the time, always, and do not use platform.  Variable dichotomised: rarely, sometimes, most of the time, and always/ never and I do not use Instagram. |  | Binary | Current | Specified: Instagram | Instagram | Media-sharing  (passive use) | Marketer-gen |
| Other SM activities | Chapin 2018 | Number of SM platforms used | Self-report measure: Asked to circle which SM platforms used and to identify which were favourites.  A space was provided for students write in other platforms they were using. |  | Continuous | Current | Examples: Texting, Facebook, Twitter, Instagram, Pinterest, and Snapchat | Mixed platforms | SNS  (unclear) | NA |
| Other SM activities | Gomez 2019 | Signed up to more than 5 SNS | Self-report measure:  No further information reported. |  | Binary | NR | NR | NR | SNS  (unclear) | NA |
| Other SM activities | Kaufman 2014 | Has a Facebook account | Self-report 1-item measure: 1-Had a Facebook account.  Responses: yes/no. |  | Binary | Current | Specified: Facebook | Facebook | SNS  (unclear) | NA |
| Other SM activities | Kaufman 2014 | Has a Mxit account | Self-report 1-item measure: 1-Had a Mxit account. Responses: yes/no. |  | Binary | Current | Specified: Mxit | Mxit | SNS  (unclear) | NA |
| Other SM activities | Landry 2013 | Has a Facebook account | Self-report measure: If internet used, how often, and if had accounts on any of the following SM sites: Facebook, Myspace, Twitter, Yahoo, YouTube, My yearbook, Tumblr, Google buzz, Flickr, Ustream, and other.  Responses: yes/no. |  | Binary | Current | Specified: Facebook, Myspace, Twitter, Yahoo, YouTube, My Yearbook, Tumblr, Google buzz, Flickr, Ustream, and other | Facebook | SNS  (unclear) | NA |
| Other SM activities | Landry 2013 | Has a SM account | Self-report measure:  If internet used, how often, and if had accounts on any of the following SM sites: Facebook, Myspace, Twitter, Yahoo, YouTube, My yearbook, Tumblr, Google buzz, Flickr, Ustream, and other.  Responses: yes/no. |  | Binary | Current | Specified: Facebook, Myspace, Twitter, Yahoo, YouTube, My Yearbook, Tumblr, Google buzz, Flickr, Ustream, and other | Mixed platforms | SNS  (unclear) | NA |
| Other SM activities | Molla-Esparza 2021 | Number of SM platforms used | Self-report 1-item measure:  1-Number of SM platforms used. |  | Continuous | Current | NR | NR | General SM  (unclear) | NA |

| Exposure | Author and year | Exposure definition | Exposure ascertainment | Validated/  objectively recorded | Exposure measure  type | Exposure time period | SM platform | SM platform | SM category (active/  passive use) | SM content |
| --- | --- | --- | --- | --- | --- | --- | --- | --- | --- | --- |
| Other SM activities | Nesi 2019 | Online status-seeking strategy use | Self-report 2-item measure: Rated use of strategies to manage their online presence and accumulate online status indicators: 1-“I purposefully post on social media during ‘high traffic’ times (i.e., times that I know most people will see it) so that my posts/photos get more likes and comments” and;  2-“If something I post does not get a lot of likes or comments, I might take it down.” Responses: from (1) not at all true to (5) extremely true. |  | Continuous | Current | NR | NR | SNS  (unclear) | NA |
| Other SM activities | Roditis 2016 | Ever seen a message posted on SM about the risks or bad things of using marijuana | Self-report 1-item measure: 1-Ever seen a message posted on SM about the risks or bad things related to using marijuana. Responses: yes/no. |  | Binary | Ever | NR | NR | General SM  (passive use) | NA |
| Other SM activities | Stevens 2017 | Exposure to contraception information on SNS | Self-report measure: In past 30 days, where had heard about pregnancy prevention among young people. Options included SNS. Responses: yes/no. |  | Binary | Past month | NR | NR | SNS  (unclear) | NA |
| Other SM activities | Stevens 2017 | Exposure to HIV/STD information on SNS | Self-report measure: In past 30 days, where had heard about HIV or STDs. Options included SNS. Responses: yes/no |  | Binary | Past month | NR | NR | SNS  (unclear) | NA |
| Other SM activities | Suwanwong 2021 | Exposure to Anti-smoking SM campaign | Self-report 1-item measure:  1-In past 30 days, seen any information about anti-smoking in SM.  Responses: yes/no. |  | Binary | Past month | NR | NR | General SM  (passive use) | NA |

**Legend:** Abbreviations: Freq = Frequency; IM = Instant messaging; Hrs = Hours; Marketer-gen = Marketer-generated content; Min = Minutes; NA = Not applicable; NR = Not reported; SM = Social media; SNS = Social networking sites; T = Timepoint and User-gen = User-generated content.

### Appendix-15. Exposure and outcome combinations amenable to meta-analysis

##### Table A. List of outcomes and exposures for which sufficient data were available to undertake meta-analysis, subgroup analysis/meta-regression^a^ or sensitivity analysis

| **Outcome** | **Exposure** | **Exposure measure type** | **Outcome measure type** | **Common metric** | **Meta-analysis** | **Prespecified sensitivity analyses** | | | | **Prespecified subgroup analyses/meta-regression**^a^ | | | | | | |
| --- | --- | --- | --- | --- | --- | --- | --- | --- | --- | --- | --- | --- | --- | --- | --- | --- |
|  |  |  |  |  |  | **Study design** | **Excl. age overlap** | **Adjust-ment** | **RoB** | **Sex** | **Age** | **Income** | **SEP** | **SM platform** | **SM category** | **SM content** |
| Alcohol use | Time spent on SM | Continuous | Continuous | Std. beta | ✓ | ✓ |  | ✓ |  |  |  |  |  |  |  |  |
|  |  | Continuous | Continuous | Std. mean difference | ✓ |  |  |  |  |  |  |  |  |  | ✓ |  |
|  |  | Continuous | Binary | Odds ratio |  |  |  |  |  |  |  |  |  |  |  |  |
|  |  | Binary | Binary/  continuous | Odds ratio | ✓ | ✓ | ✓ | ✓ | ✓ |  | ✓ |  |  |  | ✓ |  |
|  | Frequency of SM use | Continuous | Continuous | Std. beta | ✓ | ✓ |  | ✓ |  |  | ✓ |  |  |  | ✓ |  |
|  |  | Continuous | Continuous | Std. mean difference |  |  |  |  |  |  |  |  |  |  |  |  |
|  |  | Continuous | Binary | Odds ratio |  |  |  |  |  |  |  |  |  |  |  |  |
|  |  | Binary | Binary/  continuous | Odds ratio | ✓ | ✓ | ✓ | ✓ | ✓ | ✓ | ✓ | ✓ |  | ✓ | ✓ |  |
|  | Exposure to health-risk behaviour content | Continuous | Continuous | Std. beta | ✓ | ✓ | ✓ | ✓ | ✓ |  | ✓ |  | ✓ |  | ✓ |  |
|  |  | Continuous | Continuous | Std. mean difference |  |  |  |  |  |  |  |  |  |  |  |  |
|  |  | Continuous | Binary | Odds ratio |  |  |  |  |  |  |  |  |  |  |  |  |
|  |  | Binary | Binary/  continuous | Odds ratio | ✓ | ✓ |  | ✓ | ✓ |  |  |  |  |  | ✓ | ✓ |

| **Outcome** | **Exposure** | **Exposure measure type** | **Outcome measure type** | **Common metric** | **Meta-analysis** | **Prespecified sensitivity analyses** | | | | **Prespecified subgroup analyses/meta-regression**^a^ | | | | | | |
| --- | --- | --- | --- | --- | --- | --- | --- | --- | --- | --- | --- | --- | --- | --- | --- | --- |
|  |  |  |  |  |  | **Study design** | **Excl. age overlap** | **Adjust-ment** | **RoB** | **Sex** | **Age** | **Income** | **SEP** | **SM platform** | **SM category** | **SM content** |
| Sexual risk behaviour | Time spent on SM | Continuous | Continuous | Std. beta | ✓ |  |  |  |  |  |  |  |  |  |  |  |
|  |  | Continuous | Continuous | Std. mean difference | ✓ |  |  |  |  |  |  |  |  |  |  |  |
|  |  | Continuous | Binary | Odds ratio |  |  |  |  |  |  |  |  |  |  |  |  |
|  |  | Binary | Binary/  continuous | Odds ratio |  |  |  |  |  |  |  |  |  |  |  |  |
|  | Frequency of SM use | Continuous | Continuous | Std. beta |  |  |  |  |  |  |  |  |  |  |  |  |
|  |  | Continuous | Continuous | Std. mean difference | ✓ | ✓ |  |  |  |  | ✓ |  |  |  |  |  |
|  |  | Continuous | Binary | Odds ratio | ✓ | ✓ |  |  | ✓ |  |  |  |  |  | ✓ |  |
|  |  | Binary | Binary/  continuous | Odds ratio | ✓ |  | ✓ | ✓ | ✓ | ✓ | ✓ | ✓ | ✓ |  | ✓ |  |
|  | Exposure to health-risk behaviour content | Continuous | Continuous | Std. beta |  |  |  |  |  |  |  |  |  |  |  |  |
|  |  | Continuous | Continuous | Std. mean difference |  |  |  |  |  |  |  |  |  |  |  |  |
|  |  | Continuous | Binary | Odds ratio |  |  |  |  |  |  |  |  |  |  |  |  |
|  |  | Binary | Binary/  continuous | Odds ratio |  |  |  |  |  |  |  |  |  |  |  |  |
| Anti-social behaviour | Time spent on SM | Continuous | Continuous | Std. beta |  |  |  |  |  |  |  |  |  |  |  |  |
|  |  | Continuous | Continuous | Std. mean difference | ✓ |  |  |  |  |  | ✓ |  |  |  | ✓ |  |
|  |  | Continuous | Binary | Odds ratio |  |  |  |  |  |  |  |  |  |  |  |  |
|  |  | Binary | Binary/  continuous | Odds ratio |  |  |  |  |  |  |  |  |  |  |  |  |
|  | Frequency of SM use | Continuous | Continuous | Std. beta |  |  |  |  |  |  |  |  |  |  |  |  |
|  |  | Continuous | Continuous | Std. mean difference |  |  |  |  |  |  |  |  |  |  |  |  |
|  |  | Continuous | Binary | Odds ratio |  |  |  |  |  |  |  |  |  |  |  |  |
|  |  | Binary | Binary/  continuous | Odds ratio | ✓ | ✓ | ✓ | ✓ | ✓ |  |  | ✓ |  |  | ✓ |  |
|  | Exposure to health-risk behaviour content | Continuous | Continuous | Std. beta |  |  |  |  |  |  |  |  |  |  |  |  |
|  |  | Continuous | Continuous | Std. mean difference |  |  |  |  |  |  |  |  |  |  |  |  |
|  |  | Continuous | Binary | Odds ratio |  |  |  |  |  |  |  |  |  |  |  |  |
|  |  | Binary | Binary/  continuous | Odds ratio |  |  |  |  |  |  |  |  |  |  |  |  |

| **Outcome** | **Exposure** | **Exposure measure type** | **Outcome measure type** | **Common metric** | **Meta-analysis** | **Prespecified sensitivity analyses** | | | | **Prespecified subgroup analyses/meta-regression**^a^ | | | | | | |
| --- | --- | --- | --- | --- | --- | --- | --- | --- | --- | --- | --- | --- | --- | --- | --- | --- |
|  |  |  |  |  |  | **Study design** | **Excl. age overlap** | **Adjust-ment** | **RoB** | **Sex** | **Age** | **Income** | **SEP** | **SM platform** | **SM category** | **SM content** |
| Tobacco use | Time spent on SM | Continuous | Continuous | Std. beta |  |  |  |  |  |  |  |  |  |  |  |  |
|  |  | Continuous | Continuous | Std. mean difference |  |  |  |  |  |  |  |  |  |  |  |  |
|  |  | Continuous | Binary | Odds ratio |  |  |  |  |  |  |  |  |  |  |  |  |
|  |  | Binary | Binary/  continuous | Odds ratio |  |  |  |  |  |  |  |  |  |  |  |  |
|  | Frequency of SM use | Continuous | Continuous | Std. beta |  |  |  |  |  |  |  |  |  |  |  |  |
|  |  | Continuous | Continuous | Std. mean difference |  |  |  |  |  |  |  |  |  |  |  |  |
|  |  | Continuous | Binary | Odds ratio |  |  |  |  |  |  |  |  |  |  |  |  |
|  |  | Binary | Binary/  continuous | Odds ratio | ✓ | ✓ | ✓ | ✓ | ✓ |  | ✓ | ✓ |  |  | ✓ |  |
|  | Exposure to health-risk behaviour content | Continuous | Continuous | Std. beta |  |  |  |  |  |  |  |  |  |  |  |  |
|  |  | Continuous | Continuous | Std. mean difference |  |  |  |  |  |  |  |  |  |  |  |  |
|  |  | Continuous | Binary | Odds ratio |  |  |  |  |  |  |  |  |  |  |  |  |
|  |  | Binary | Binary/  continuous | Odds ratio | ✓ | ✓ |  | ✓ | ✓ |  |  | ✓ |  |  | ✓ |  |
| Drug use | Time spent on SM | Continuous | Continuous | Std. beta |  |  |  |  |  |  |  |  |  |  |  |  |
|  |  | Continuous | Continuous | Std. mean difference | ✓ |  |  |  |  |  |  |  |  |  |  |  |
|  |  | Continuous | Binary | Odds ratio |  |  |  |  |  |  |  |  |  |  |  |  |
|  |  | Binary | Binary/  continuous | Odds ratio | ✓ |  |  |  | ✓ |  |  |  |  |  |  |  |
|  | Frequency of SM use | Continuous | Continuous | Std. beta |  |  |  |  |  |  |  |  |  |  |  |  |
|  |  | Continuous | Continuous | Std. mean difference |  |  |  |  |  |  |  |  |  |  |  |  |
|  |  | Continuous | Binary | Odds ratio |  |  |  |  |  |  |  |  |  |  |  |  |
|  |  | Binary | Binary/  continuous | Odds ratio | ✓ | ✓ |  | ✓ | ✓ |  | ✓ |  |  | ✓ | ✓ |  |
|  | Exposure to health-risk behaviour content | Continuous | Continuous | Std. beta |  |  |  |  |  |  |  |  |  |  |  |  |
|  |  | Continuous | Continuous | Std. mean difference |  |  |  |  |  |  |  |  |  |  |  |  |
|  |  | Continuous | Binary | Odds ratio |  |  |  |  |  |  |  |  |  |  |  |  |
|  |  | Binary | Binary/  continuous | Odds ratio |  |  |  |  |  |  |  |  |  |  |  |  |

| **Outcome** | **Exposure** | **Exposure measure type** | **Outcome measure type** | **Common metric** | **Meta-analysis** | **Prespecified sensitivity analyses** | | | | **Prespecified subgroup analyses/meta-regression**^a^ | | | | | | |
| --- | --- | --- | --- | --- | --- | --- | --- | --- | --- | --- | --- | --- | --- | --- | --- | --- |
|  |  |  |  |  |  | **Study design** | **Excl. age overlap** | **Adjust-ment** | **RoB** | **Sex** | **Age** | **Income** | **SEP** | **SM platform** | **SM category** | **SM content** |
| Inadequate physical activity | Time spent on SM | Continuous | Continuous | Std. beta | ✓ |  |  | ✓ | ✓ |  | ✓ | ✓ |  |  | ✓ |  |
|  |  | Continuous | Continuous | Std. mean difference |  |  |  |  |  |  |  |  |  |  |  |  |
|  |  | Continuous | Binary | Odds ratio |  |  |  |  |  |  |  |  |  |  |  |  |
|  |  | Binary | Binary/  continuous | Odds ratio |  |  |  |  |  |  |  |  |  |  |  |  |
|  | Frequency of SM use | Continuous | Continuous | Std. beta |  |  |  |  |  |  |  |  |  |  |  |  |
|  |  | Continuous | Continuous | Std. mean difference |  |  |  |  |  |  |  |  |  |  |  |  |
|  |  | Continuous | Binary | Odds ratio |  |  |  |  |  |  |  |  |  |  |  |  |
|  |  | Binary | Binary/  continuous | Odds ratio |  |  |  |  |  |  |  |  |  |  |  |  |
|  | Exposure to health-risk behaviour content | Continuous | Continuous | Std. beta |  |  |  |  |  |  |  |  |  |  |  |  |
|  |  | Continuous | Continuous | Std. mean difference |  |  |  |  |  |  |  |  |  |  |  |  |
|  |  | Continuous | Binary | Odds ratio |  |  |  |  |  |  |  |  |  |  |  |  |
|  |  | Binary | Binary/  continuous | Odds ratio |  |  |  |  |  |  |  |  |  |  |  |  |
| Unhealthy dietary behaviour | Time spent on SM | Continuous | Continuous | Std. beta |  |  |  |  |  |  |  |  |  |  |  |  |
|  |  | Continuous | Continuous | Std. mean difference |  |  |  |  |  |  |  |  |  |  |  |  |
|  |  | Continuous | Binary | Odds ratio |  |  |  |  |  |  |  |  |  |  |  |  |
|  |  | Binary | Binary/  continuous | Odds ratio |  |  |  |  |  |  |  |  |  |  |  |  |
|  | Frequency of SM use | Continuous | Continuous | Std. beta |  |  |  |  |  |  |  |  |  |  |  |  |
|  |  | Continuous | Continuous | Std. mean difference |  |  |  |  |  |  |  |  |  |  |  |  |
|  |  | Continuous | Binary | Odds ratio |  |  |  |  |  |  |  |  |  |  |  |  |
|  |  | Binary | Binary/  continuous | Odds ratio |  |  |  |  |  |  |  |  |  |  |  |  |
|  | Exposure to health-risk behaviour content | Continuous | Continuous | Std. beta |  |  |  |  |  |  |  |  |  |  |  |  |
|  |  | Continuous | Continuous | Std. mean difference |  |  |  |  |  |  |  |  |  |  |  |  |
|  |  | Continuous | Binary | Odds ratio |  |  |  |  |  |  |  |  |  |  |  |  |
|  |  | Binary | Binary/  continuous | Odds ratio | ✓ | ✓ |  |  |  |  |  |  |  |  | ✓ |  |

| **Outcome** | **Exposure** | **Exposure measure type** | **Outcome measure type** | **Common metric** | **Meta-analysis** | **Prespecified sensitivity analyses** | | | | **Prespecified subgroup analyses/meta-regression**^a^ | | | | | | |
| --- | --- | --- | --- | --- | --- | --- | --- | --- | --- | --- | --- | --- | --- | --- | --- | --- |
|  |  |  |  |  |  | **Study design** | **Excl. age overlap** | **Adjust-ment** | **RoB** | **Sex** | **Age** | **Income** | **SEP** | **SM platform** | **SM category** | **SM content** |
| Gambling | Time spent on SM | Continuous | Continuous | Std. beta |  |  |  |  |  |  |  |  |  |  |  |  |
|  |  | Continuous | Continuous | Std. mean difference |  |  |  |  |  |  |  |  |  |  |  |  |
|  |  | Continuous | Binary | Odds ratio |  |  |  |  |  |  |  |  |  |  |  |  |
|  |  | Binary | Binary/  continuous | Odds ratio |  |  |  |  |  |  |  |  |  |  |  |  |
|  | Frequency of SM use | Continuous | Continuous | Std. beta |  |  |  |  |  |  |  |  |  |  |  |  |
|  |  | Continuous | Continuous | Std. mean difference |  |  |  |  |  |  |  |  |  |  |  |  |
|  |  | Continuous | Binary | Odds ratio |  |  |  |  |  |  |  |  |  |  |  |  |
|  |  | Binary | Binary/  continuous | Odds ratio | ✓ | ✓ |  | ✓ | ✓ |  | ✓ |  |  |  | ✓ |  |
|  | Exposure to health-risk behaviour content | Continuous | Continuous | Std. beta |  |  |  |  |  |  |  |  |  |  |  |  |
|  |  | Continuous | Continuous | Std. mean difference |  |  |  |  |  |  |  |  |  |  |  |  |
|  |  | Continuous | Binary | Odds ratio |  |  |  |  |  |  |  |  |  |  |  |  |
|  |  | Binary | Binary/  continuous | Odds ratio |  |  |  |  |  |  |  |  |  |  |  |  |
| Multiple risk behaviours | Time spent on SM | Continuous | Continuous | Std. beta |  |  |  |  |  |  |  |  |  |  |  |  |
|  |  | Continuous | Continuous | Std. mean difference |  |  |  |  |  |  |  |  |  |  |  |  |
|  |  | Continuous | Binary | Odds ratio |  |  |  |  |  |  |  |  |  |  |  |  |
|  |  | Binary | Binary/  continuous | Odds ratio |  |  |  |  |  |  |  |  |  |  |  |  |
|  | Frequency of SM use | Continuous | Continuous | Std. beta |  |  |  |  |  |  |  |  |  |  |  |  |
|  |  | Continuous | Continuous | Std. mean difference |  |  |  |  |  |  |  |  |  |  |  |  |
|  |  | Continuous | Binary | Odds ratio |  |  |  |  |  |  |  |  |  |  |  |  |
|  |  | Binary | Binary/  continuous | Odds ratio | ✓ |  |  |  |  |  |  |  |  |  |  |  |
|  | Exposure to health-risk behaviour content | Continuous | Continuous | Std. beta |  |  |  |  |  |  |  |  |  |  |  |  |
|  |  | Continuous | Continuous | Std. mean difference |  |  |  |  |  |  |  |  |  |  |  |  |
|  |  | Continuous | Binary | Odds ratio |  |  |  |  |  |  |  |  |  |  |  |  |
|  |  | Binary | Binary/  continuous | Odds ratio |  |  |  |  |  |  |  |  |  |  |  |  |

| **Outcome** | **Exposure** | **Exposure measure type** | **Outcome measure type** | **Common metric** | **Meta-analysis** | **Prespecified sensitivity analyses** | | | | **Prespecified subgroup analyses/meta-regression**^a^ | | | | | | |
| --- | --- | --- | --- | --- | --- | --- | --- | --- | --- | --- | --- | --- | --- | --- | --- | --- |
|  |  |  |  |  |  | **Study design** | **Excl. age overlap** | **Adjust-ment** | **RoB** | **Sex** | **Age** | **Income** | **SEP** | **SM platform** | **SM category** | **SM content** |
| Use of ENDS | Time spent on SM | Continuous | Continuous | Std. beta |  |  |  |  |  |  |  |  |  |  |  |  |
|  |  | Continuous | Continuous | Std. mean difference |  |  |  |  |  |  |  |  |  |  |  |  |
|  |  | Continuous | Binary | Odds ratio |  |  |  |  |  |  |  |  |  |  |  |  |
|  |  | Binary | Binary/  continuous | Odds ratio |  |  |  |  |  |  |  |  |  |  |  |  |
|  | Frequency of SM use | Continuous | Continuous | Std. beta |  |  |  |  |  |  |  |  |  |  |  |  |
|  |  | Continuous | Continuous | Std. mean difference |  |  |  |  |  |  |  |  |  |  |  |  |
|  |  | Continuous | Binary | Odds ratio |  |  |  |  |  |  |  |  |  |  |  |  |
|  |  | Binary | Binary/  continuous | Odds ratio |  |  |  |  |  |  |  |  |  |  |  |  |
|  | Exposure to health-risk behaviour content | Continuous | Continuous | Std. beta |  |  |  |  |  |  |  |  |  |  |  |  |
|  |  | Continuous | Continuous | Std. mean difference |  |  |  |  |  |  |  |  |  |  |  |  |
|  |  | Continuous | Binary | Odds ratio |  |  |  |  |  |  |  |  |  |  |  |  |
|  |  | Binary | Binary/  continuous | Odds ratio | ✓ | ✓ |  | ✓ | ✓ |  |  |  |  | ✓ | ✓ |  |

**Legend:** ^a^ Where ≥10 studies were included in a meta-analysis; meta-regression was conducted. Abbreviations: ENDS = Electronic nicotine delivery systems; RoB = Risk of bias; SM = Social media; and Std. = Standardised.

### Appendix-16. Synthesis without meta-analysis (SWiM), meta-analyses, meta-regression, subgroup, and sensitivity analyses

#### Alcohol use

##### Effect direction plot

Figure A demonstrates the effect direction in those studies investigating alcohol use, by exposure. Six studies investigated more than one exposure.^55,86,89,113,118,159^ For time spent on social media, 15/16 studies (93.8%) reported harmful associations (95% CI 71.7 to 98.9%; participant n=100,354; sign test p < 0.001), 16/17 studies (94.1%) reported harmful associations for frequency of social media use (73.0 to 99.0%; participant n=391,445; sign test p < 0.001), and 11/12 studies (91.7%) reported harmful associations for exposure to health-risk behaviour content on social media (64.6 to 98.5%; participant n=24,451; sign test p=0.006). Other social media activities was investigated by one study which demonstrated a harmful association (20.7 to 100%; participant n=4485; insufficient data to conduct sign test).

##### Figure A. Effect direction plot for studies of the association between social media use and adolescent alcohol use, by social media exposure. Arrow size indicates sample size; arrow colour indicates study risk of bias

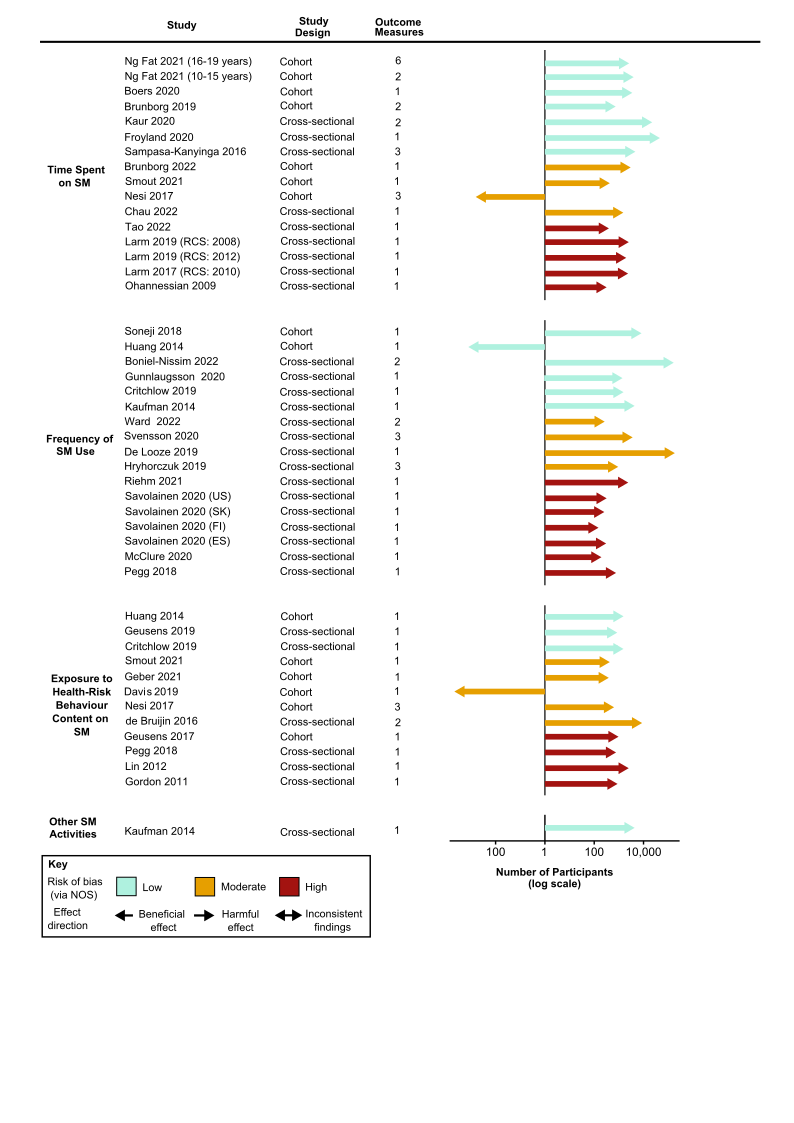

**Legend:** Sample size: represented by the size of the arrow, measured on a log scale. Outcome measure: number of outcome measures synthesised within each study. Studies organised by risk of bias grade, study design, and year of publication. Repeat cross-sectional studies, multiple study populations from different countries, and age subsets originating from the same study reported as separate studies. Abbreviations: ESP = Spain; FIN = Finland; KOR = South Korea; NOS = Assessed via adapted Newcastle Ottawa Scale; RCS = Repeat cross-sectional study; SM = Social media and USA = United States.

##### Forest plots for meta-analyses and subgroup analyses

##### Figure B. Forest plot for association between frequency of social media use and alcohol use

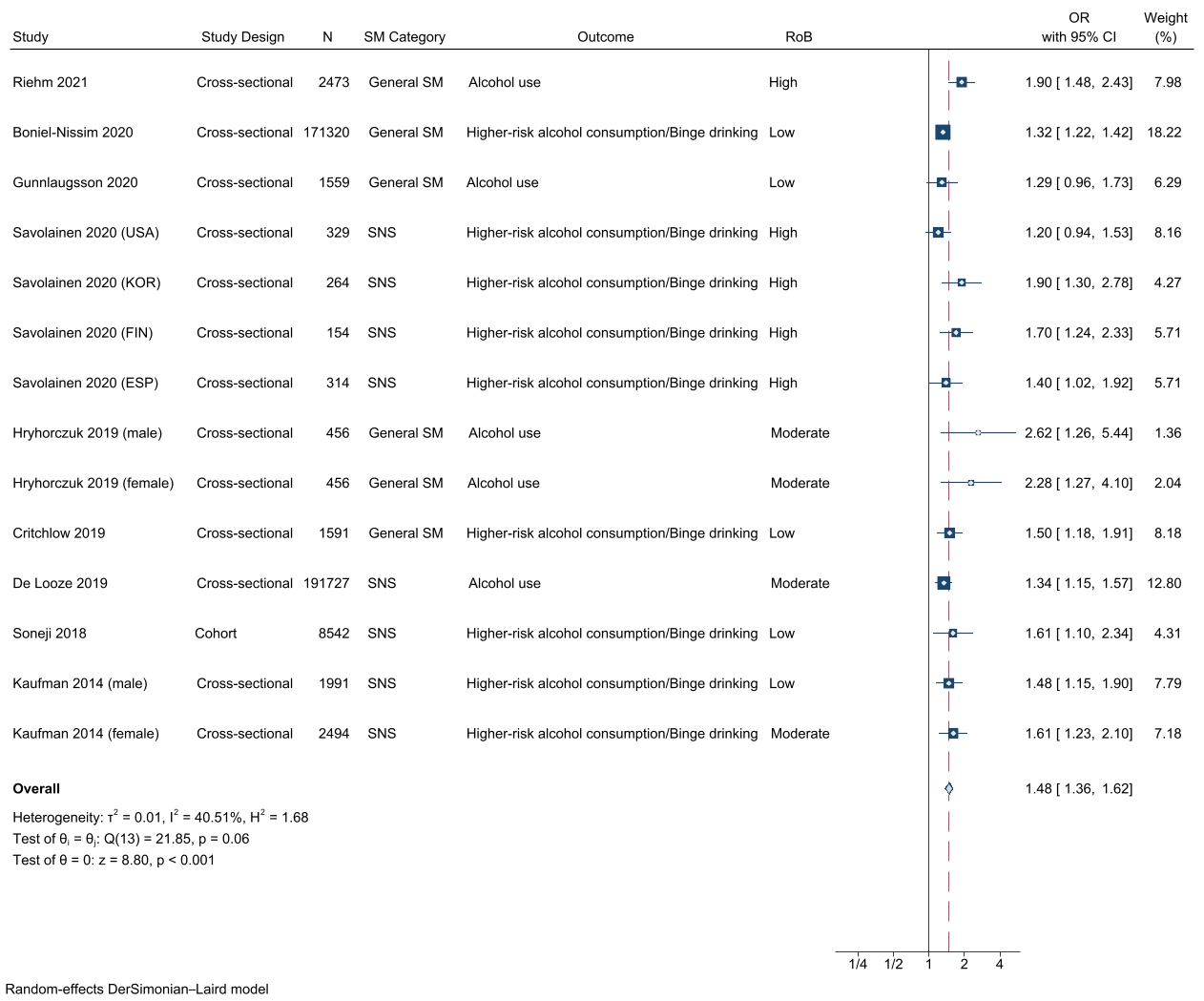
**Legend:** Figure presents forest plot for binary exposure (frequent/daily vs infrequent/non-daily) & binary/continuous outcome meta-analysis, with odds ratio (OR) used as common metric. Total number of study participants = 383,670. Abbreviations: CI = Confidence interval; ESP = Spain; FIN = Finland; KOR = South Korea; N = Number of study participants; OR = Odds ratio; RoB = Risk of bias; SM = Social media and SNS = Social networking sites.

##### Figure C. Forest plot for association between frequency of social media use and alcohol use, stratified by average age of study participants

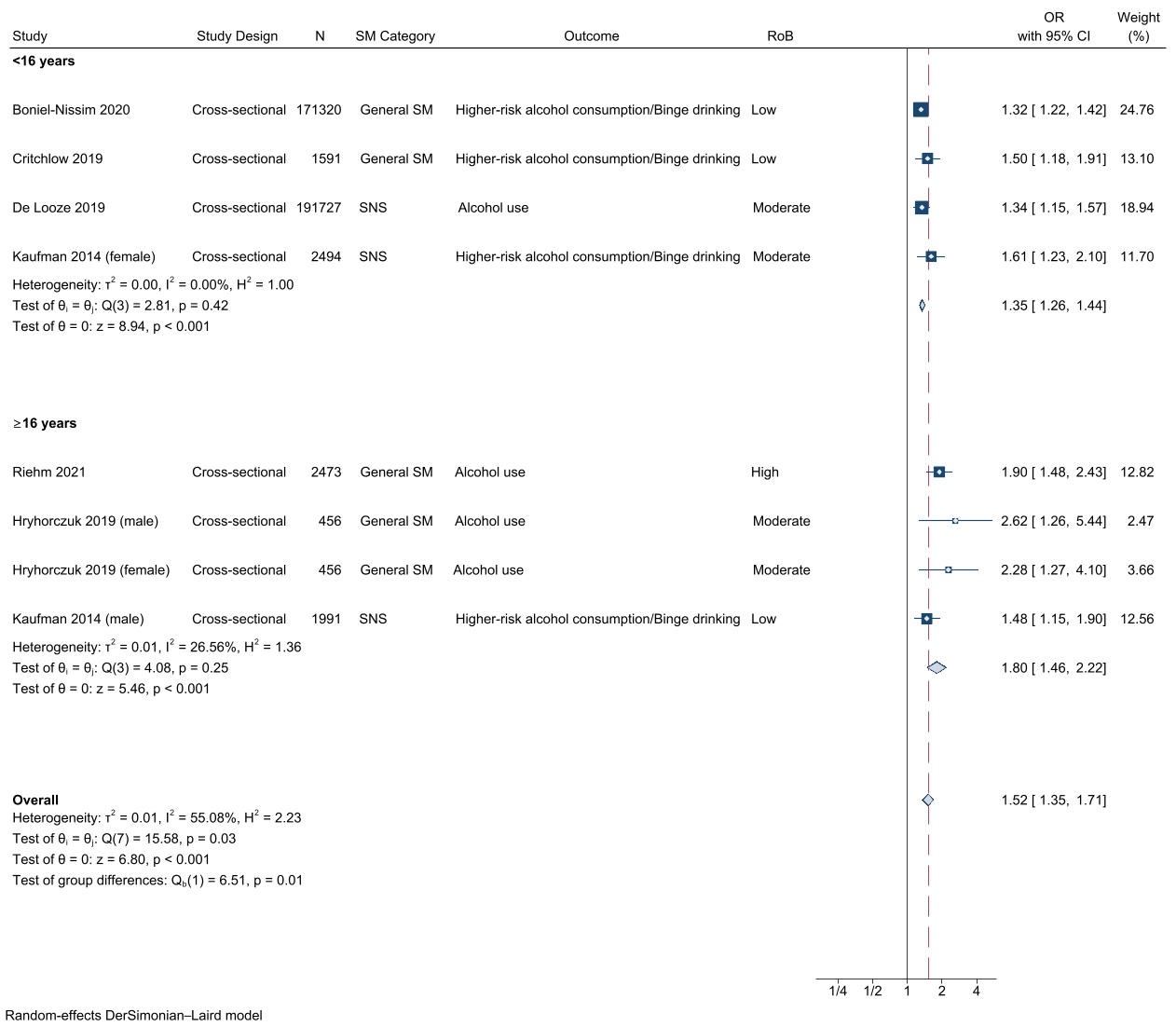

**Legend:** Figure presents forest plot for binary exposure (frequent/daily vs infrequent/non-daily) & binary/continuous outcome subgroup analysis, with odds ratio (OR) used as common metric. Total number of study participants = 372,508. Abbreviations: CI = Confidence interval; N = Number of study participants; OR = Odds ratio; RoB = Risk of bias; SM = Social media and SNS = Social networking sites.

##### Figure D. Forest plot for association between frequency of social media use alcohol use, stratified by development status of study setting^a^

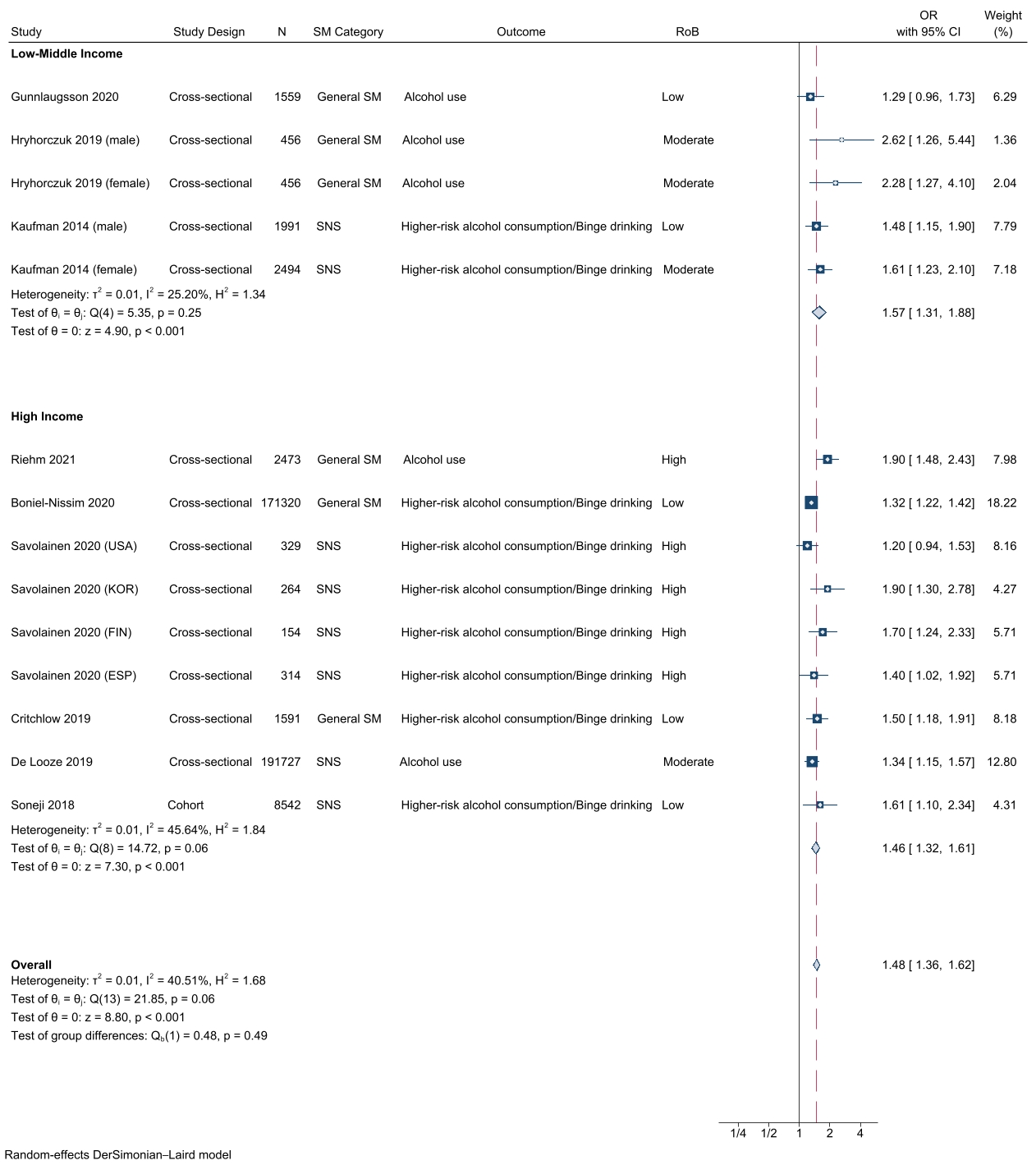

**Legend:** Figure presents forest plot for binary exposure (frequent/daily vs infrequent/non-daily) & binary/continuous outcome subgroup analysis, with odds ratio (OR) used as common metric. ^a^ Development status classified as per the World Bank Country Income Level Classification.^160^ Total number of study participants = 383,670. Abbreviations: CI = Confidence interval; ESP = Spain; FIN = Finland; KOR = South Korea; N = Number of study participants; OR = Odds ratio; RoB = Risk of bias; SM = Social media; SNS = Social networking sites and USA = United States.

##### Figure E. Forest plot for association between frequency of social media use and alcohol use, stratified by social media category

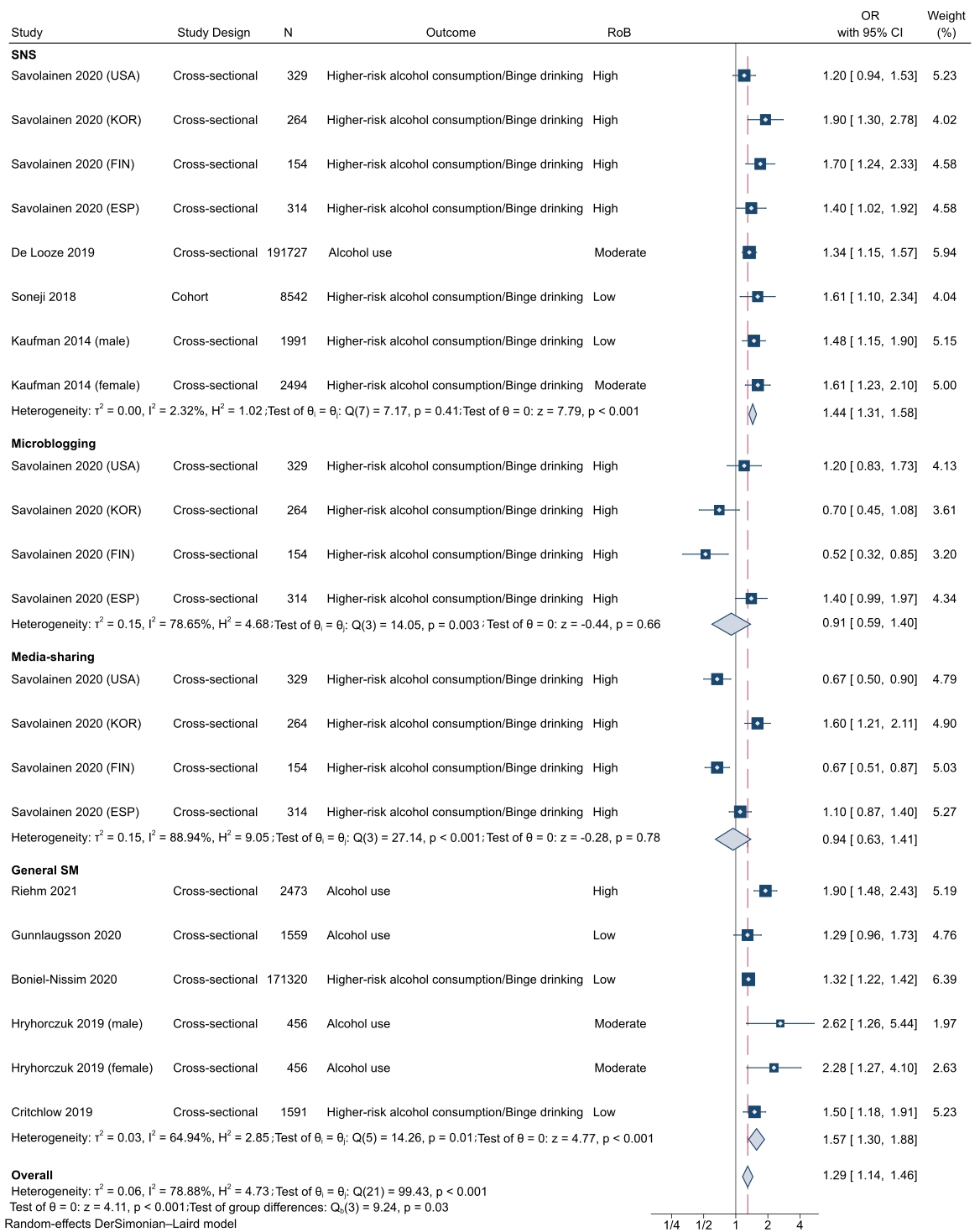

**Legend:** Figure presents forest plot for binary exposure (frequent/daily vs infrequent/non-daily) & binary/continuous outcome subgroup analysis, with odds ratio (OR) used as common metric. Total number of study participants = 385,792. Abbreviations: CI = Confidence interval; ESP = Spain; FIN = Finland; KOR = South Korea; N = Number of study participants; OR = Odds ratio; RoB = Risk of bias; SM = Social media and SNS = Social networking sites.

##### Figure F. Forest plot for association between frequency of social media use and alcohol use, stratified by social media platform

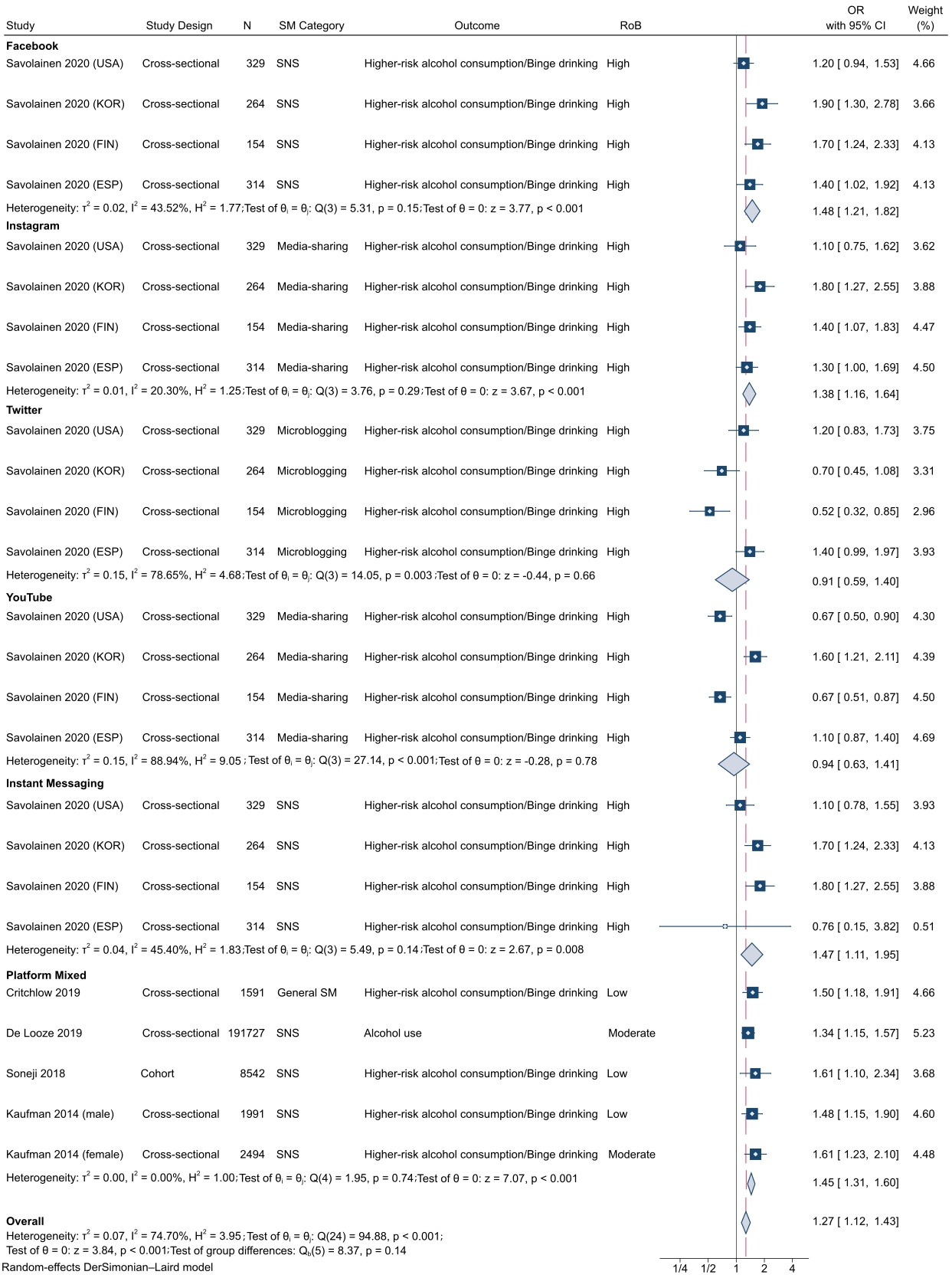

**Legend:** Figure presents forest plot for binary exposure (frequent/daily vs infrequent/non-daily) & binary/continuous outcome subgroup analysis, with odds ratio (OR) used as common metric. Total number of study participants = 211,650. Abbreviations: CI = Confidence interval; ESP = Spain; FIN = Finland; KOR = South Korea; N = Number of study participants; OR = Odds ratio; RoB = Risk of bias; SM = Social media and SNS = Social networking sites.

##### Figure G. Forest plot for association between frequency of social media use and alcohol use, stratified by sex

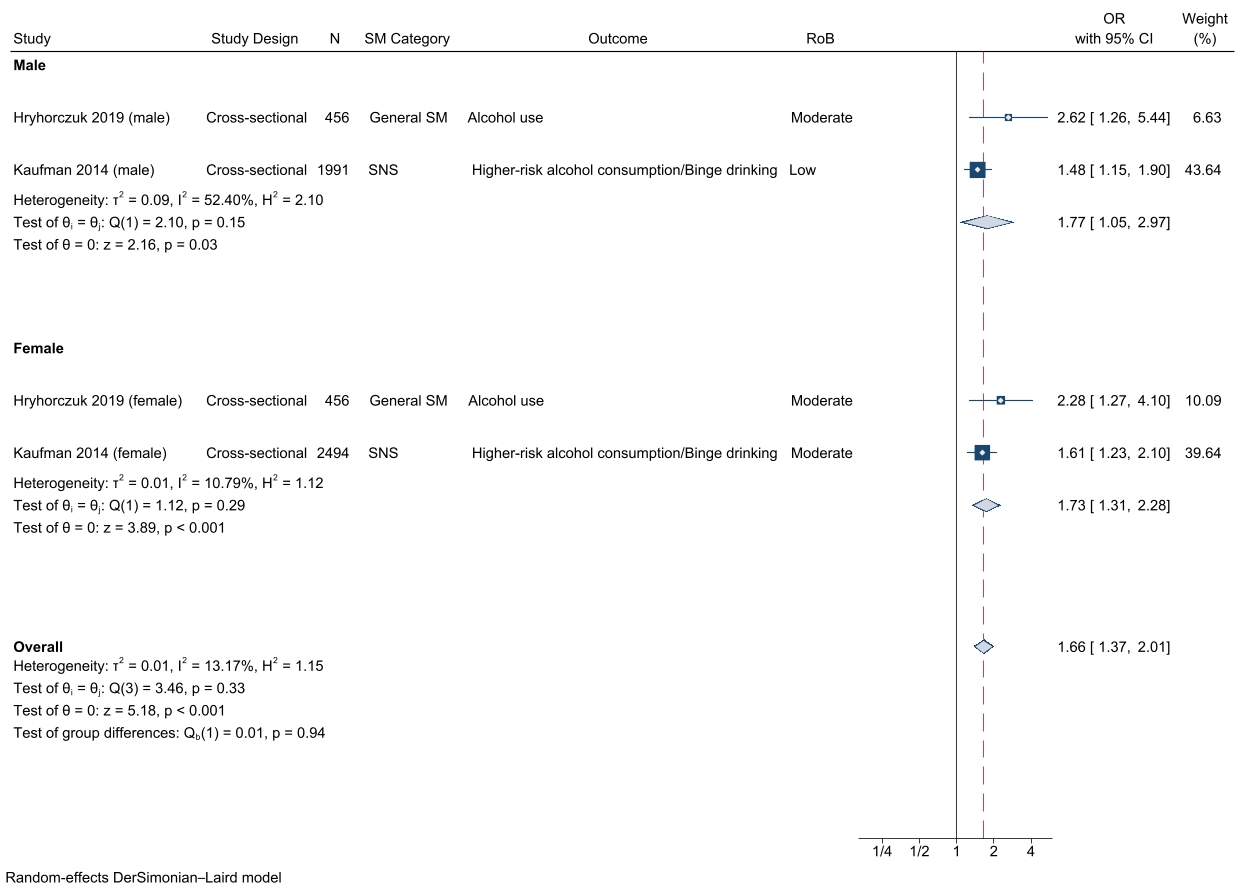

**Legend:** Figure presents forest plot for binary exposure (frequent/daily vs infrequent/non-daily) & binary/continuous outcome subgroup analysis, with odds ratio (OR) used as common metric. Total number of study participants = 5397. Abbreviations: CI = Confidence interval; N = Number of study participants; OR = Odds ratio; RoB = Risk of bias; SM = Social media and SNS = Social networking sites.

##### Figure H. Forest plot for association between time spent on social media and alcohol use

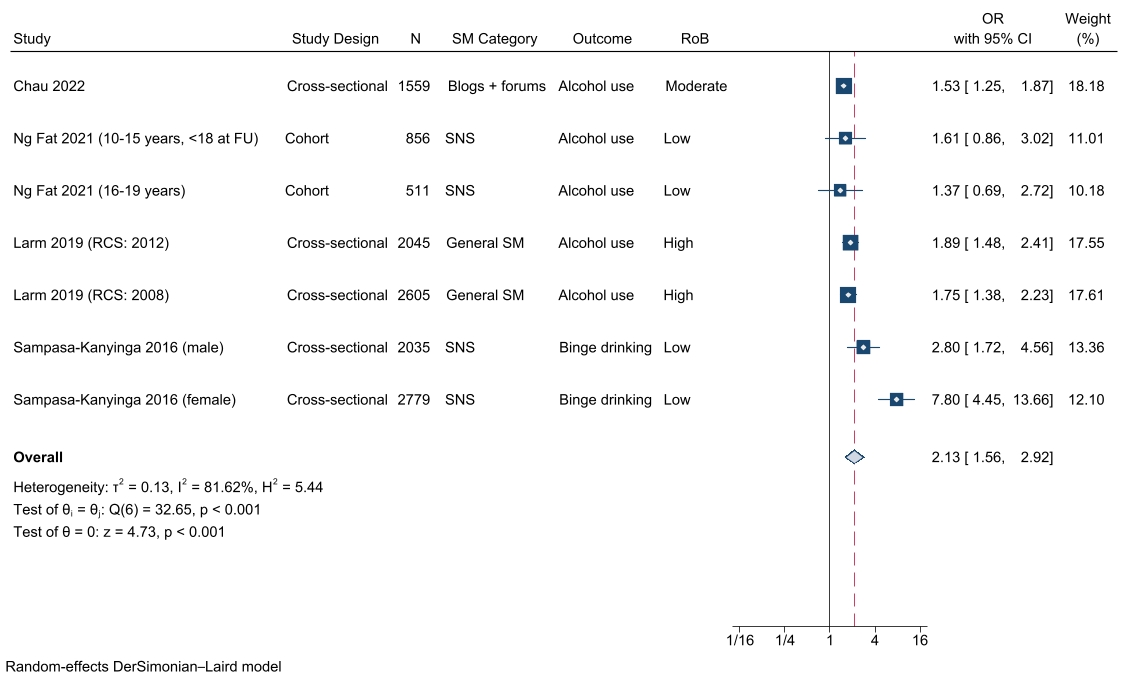
**Legend:** Figure presents forest plot for binary exposure (≥2 vs <2 hrs/day social media use) & binary/continuous outcome meta-analysis, with odds ratio (OR) used as common metric. Total number of study participants = 12,390. Abbreviations: CI = Confidence interval; FU = Follow up; hrs = Hours; N = Number of study participants; OR = Odds ratio; RCS = Repeat cross-sectional study; RoB = Risk of bias; SM = Social media and SNS = Social networking sites.

##### Figure I. Forest plot for association between exposure to health-risk behaviour content on social media and alcohol use

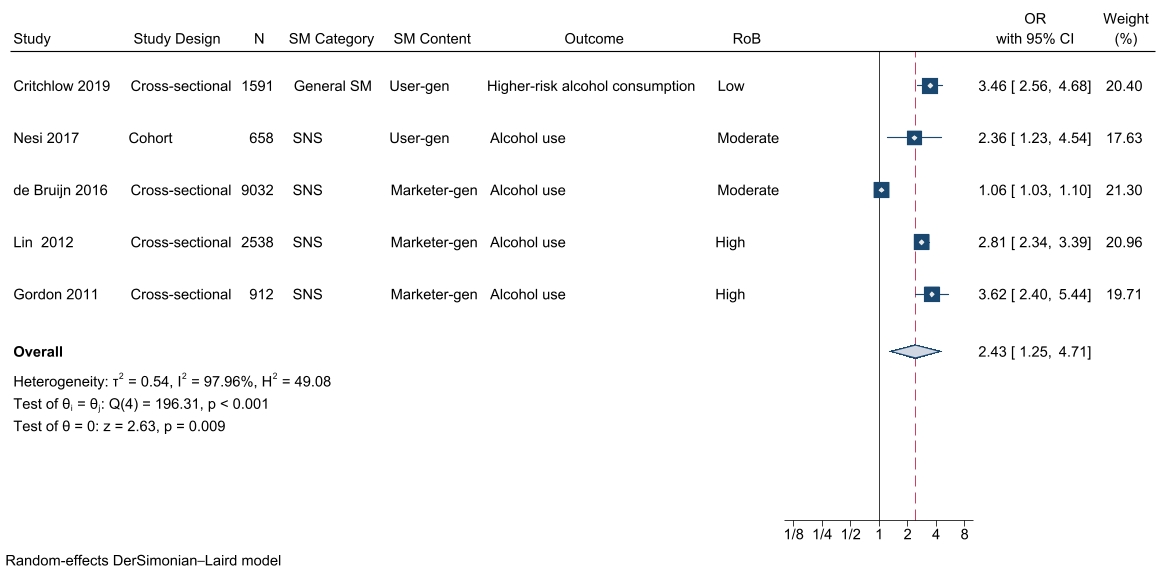

**Legend:** Figure presents forest plot for binary exposure (exposed vs unexposed) & binary/continuous outcome meta-analysis, with odds ratio (OR) used as common metric. Total number of study participants = 14,731. Abbreviations: CI = Confidence interval; Marketer-gen = Marketer-generated content; N = Number of study participants; OR = Odds ratio; RoB = Risk of bias; SM = Social media; SNS = Social networking sites and User-gen = User-generated content.

##### Figure J. Forest plot for association between time spent on social media and alcohol use, by average age of study participants

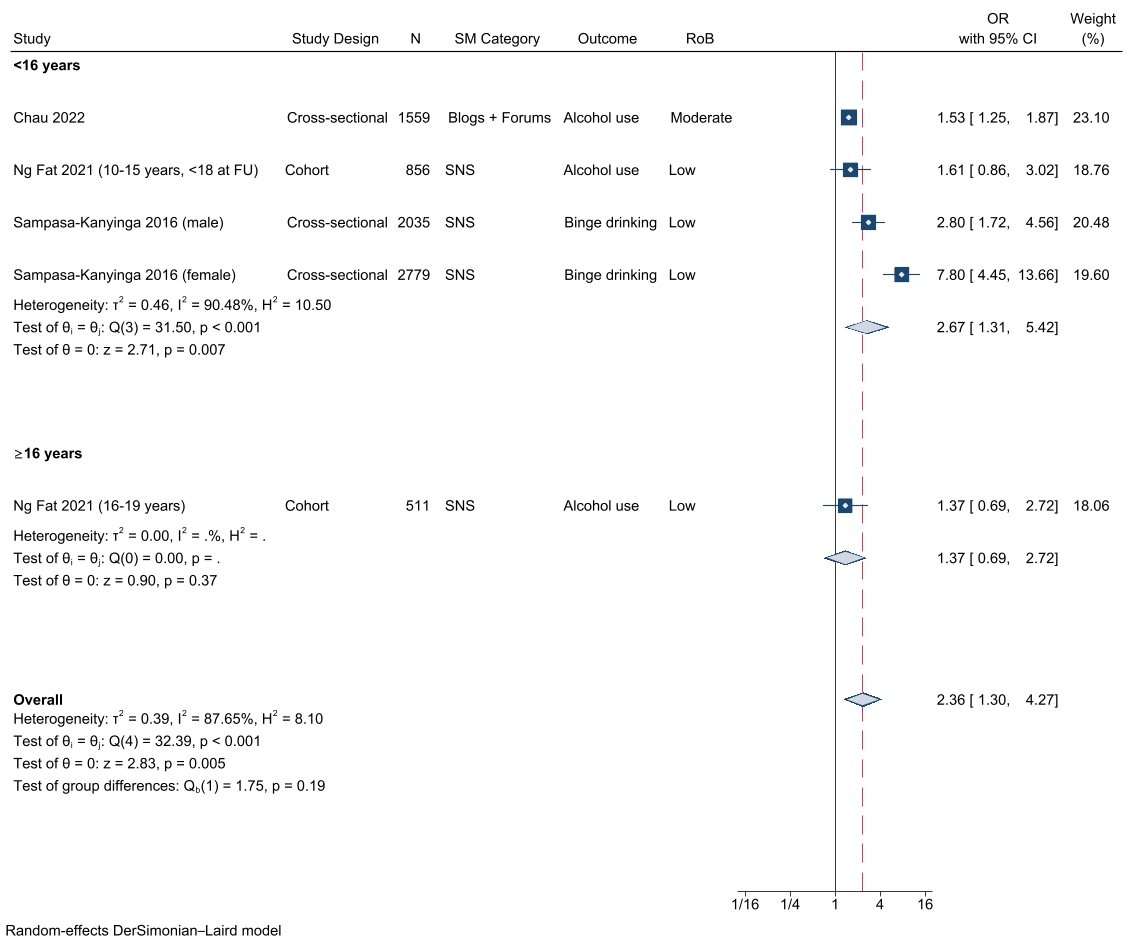

**Legend:** Figure presents forest plot for binary exposure (≥2 vs <2 hrs/day social media use) & binary/continuous outcome subgroup analysis, with odds ratio (OR) used as common metric. Total number of study participants = 7740. Abbreviations: CI = Confidence interval; FU = Follow up; hrs = Hours; N = Number of study participants; OR = Odds ratio; RoB = Risk of bias; SM = Social media and SNS = Social networking sites.

##### Figure K. Forest plot for association between time spent on social media and alcohol use, by social media category

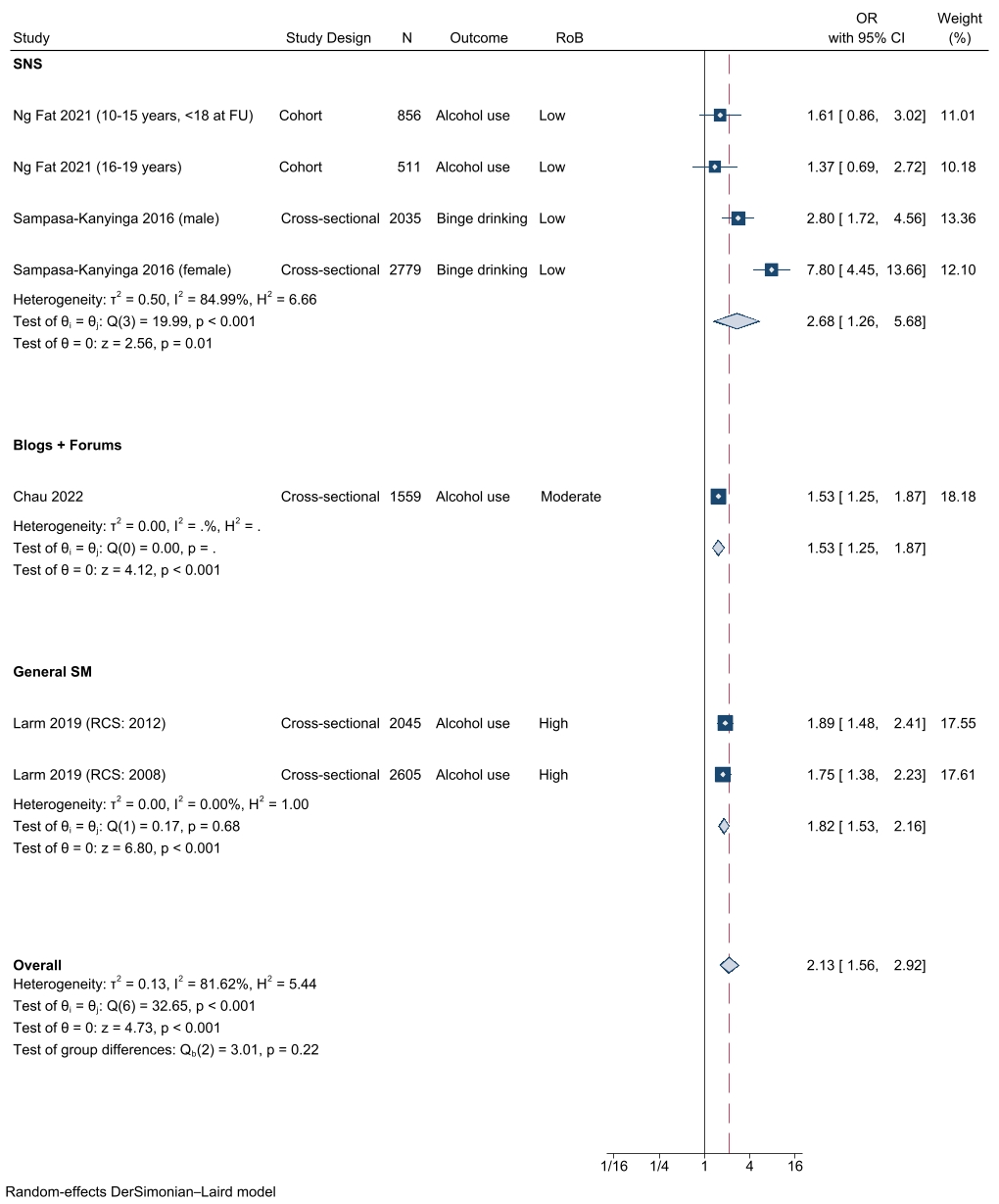

**Legend:** Figure presents forest plot for binary exposure (≥2 vs <2 hrs/day social media use) & binary/continuous outcome sensitivity analysis, with odds ratio (OR) used as common metric. Total number of study participants = 12,390. Abbreviations: CI = Confidence interval; FU = Follow up; hrs = Hours; N = Number of study participants; OR = Odds ratio; RCS = Repeat cross-sectional study; RoB = Risk of bias; SM = Social media and SNS = Social networking sites.

##### Figure L. Forest plot for association between exposure to health-risk behaviour content on social media and alcohol use, by social media category

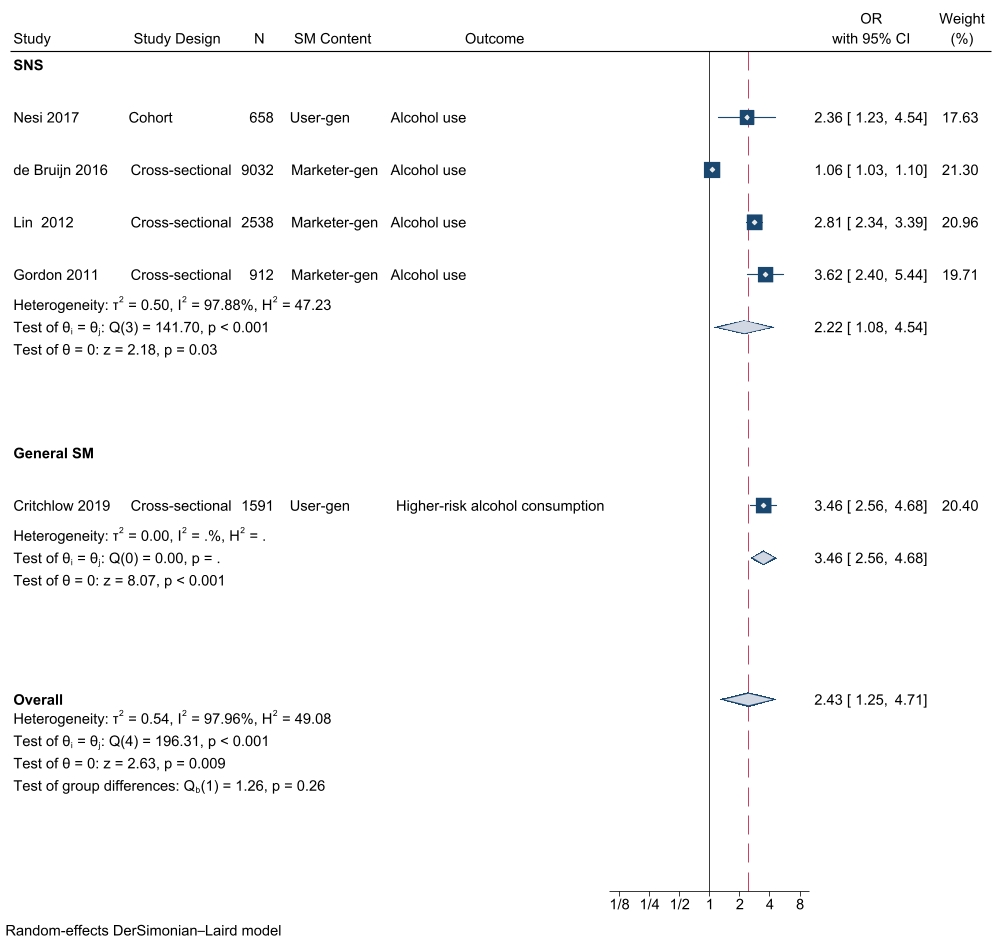

**Legend:** Figure presents forest plot for binary exposure (exposed vs unexposed) & binary/continuous outcome subgroup analysis, with odds ratio (OR) used as common metric. Total number of study participants = 14,731. Abbreviations: CI = Confidence interval; Marketer-gen = Marketer-generated content; N = Number of study participants; OR = Odds ratio; RoB = Risk of bias; SM = Social media; SNS = Social networking sites and User-gen = User-generated content.

##### Figure M. Forest plot for association between exposure to health-risk behaviour content on social media and alcohol use, by social media content

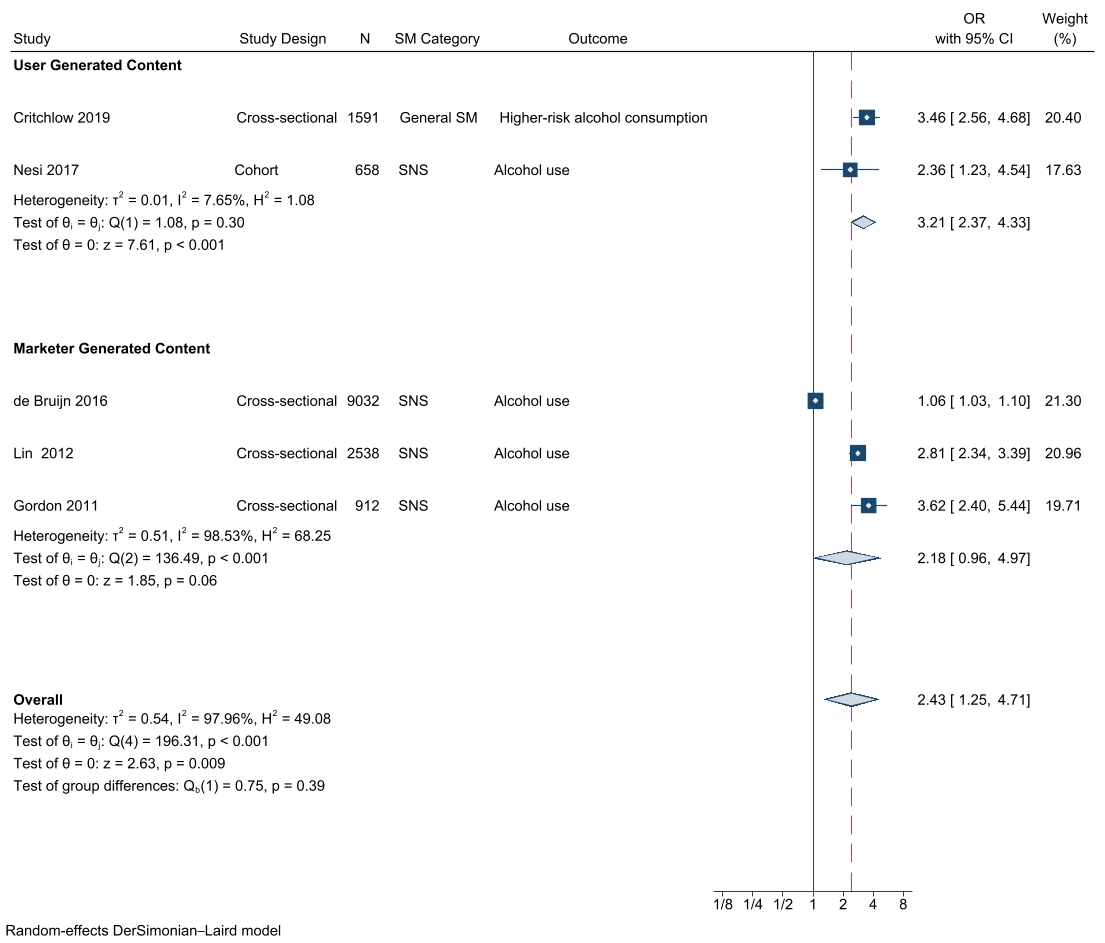

**Legend:** Figure presents forest plot for binary exposure (exposed vs unexposed) & binary/continuous outcome subgroup analysis, with odds ratio (OR) used as common metric. Total number of study participants = 14,731. Abbreviations: CI = Confidence interval; N = Number of study participants; OR = Odds ratio; RoB = Risk of bias; SM = Social media and SNS = Social networking sites.

##### Figure N. Forest plot for association between frequency of social media use and alcohol use

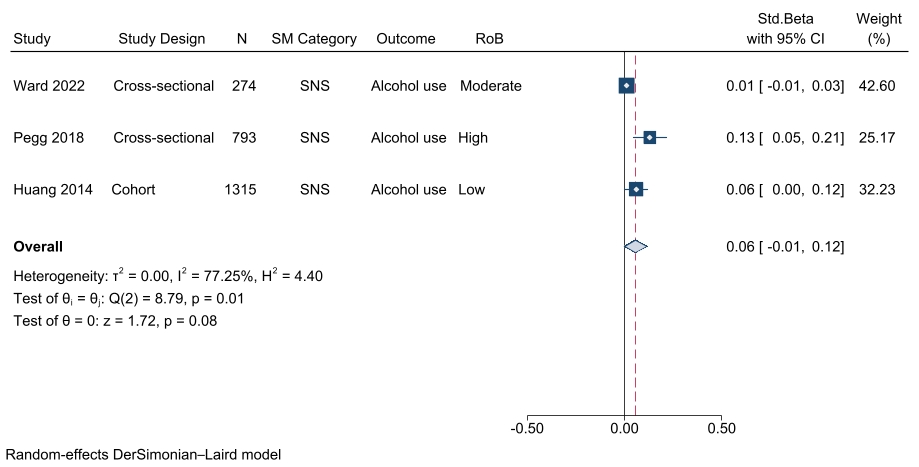

**Legend:** Figure presents forest plot for continuous exposure & continuous outcome meta-analysis, with standardised beta (Std.Beta) used as common metric. Total number of study participants = 2382. Abbreviation: CI = Confidence interval; N = Number of study participants;

RoB = Risk of bias; SM = Social media; SNS = Social networking sites and Std. Beta = Standardised beta.

##### Figure O. Forest plot for association between time spent on social media and alcohol use

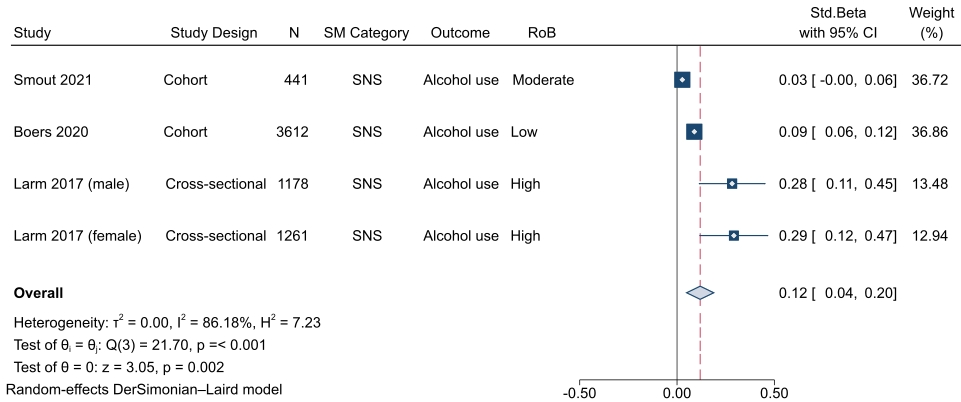
**Legend:** Figure presents forest plot for continuous exposure & continuous outcome meta-analysis, with standardised beta (Std.Beta) used as common metric. Total number of study participants = 6492. Abbreviation: CI = Confidence interval; N = Number of study participants; RoB = Risk of bias; SM = Social media; SNS = Social networking sites and Std. Beta = Standardised beta.

##### Figure P. Forest plot for association between time spent on social media and alcohol use

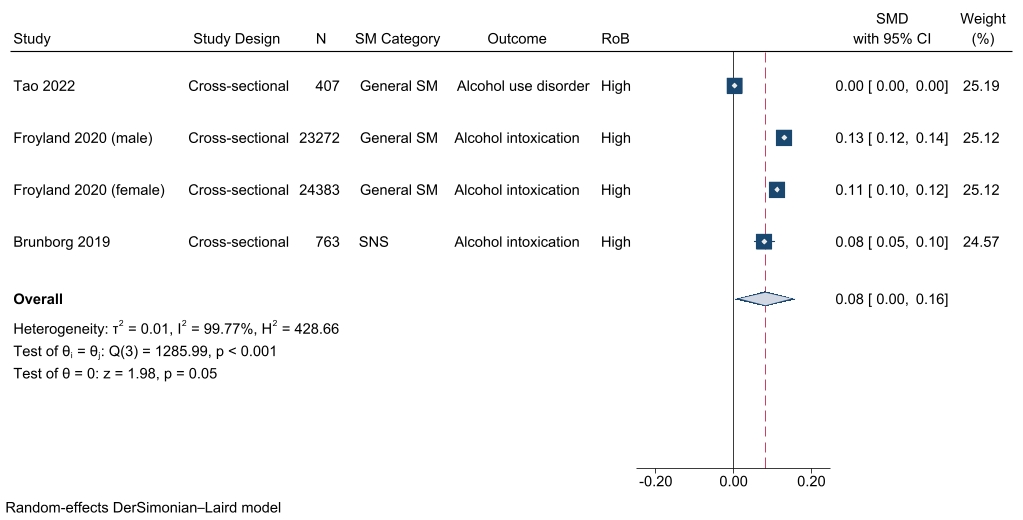

**Legend:** Figure presents forest plot for continuous exposure & continuous outcome meta-analysis, with standardised mean difference (SMD) used as common metric. Total number of study participants = 48,825. Abbreviation: CI = Confidence interval; N = Number of study participants; RoB = Risk of bias; SM = Social media; SMD = Standardised mean difference and SNS = Social networking sites.

##### Figure Q. Forest plot for associations between exposure to health-risk behaviour content on social media and alcohol use

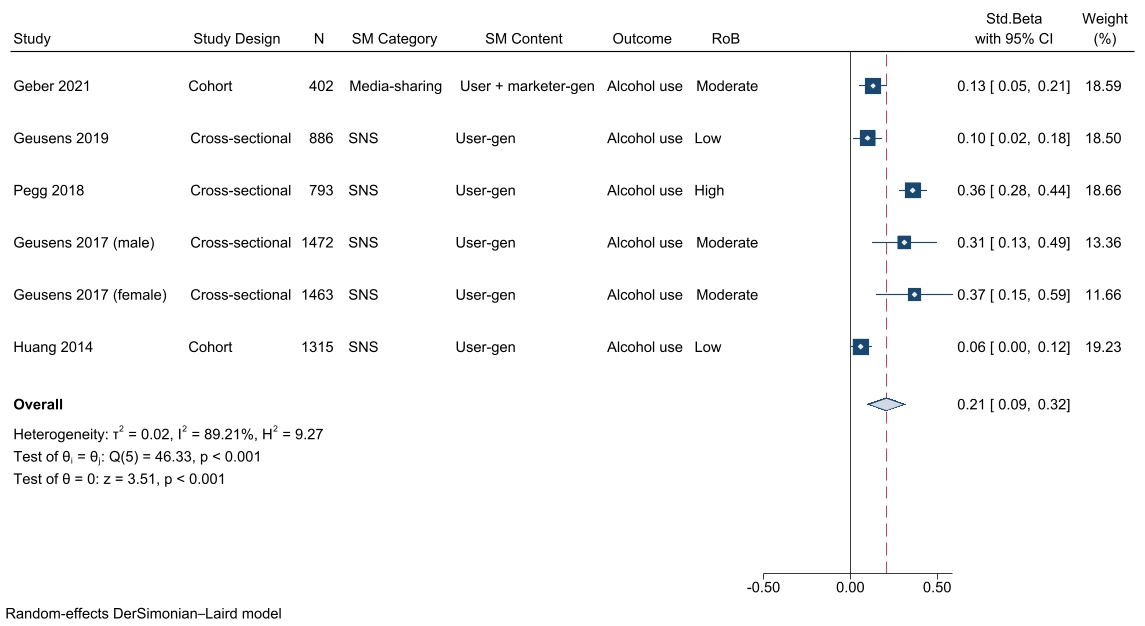

**Legend:** Figure presents forest plot for continuous exposure & continuous outcome meta-analysis, with standardised beta (Std.Beta) used as common metric. Total number of study participants = 6331. Abbreviation: CI = Confidence interval; N = Number of study participants; Marketer-gen = Marketer-generated content; RoB = Risk of bias; SM = Social media; SNS = Social networking sites; Std. Beta = Standardised beta and User-gen = User-generated content.

##### Figure R. Forest plot for association between exposure to health-risk behaviour content on social media and alcohol use, by average age of study participants

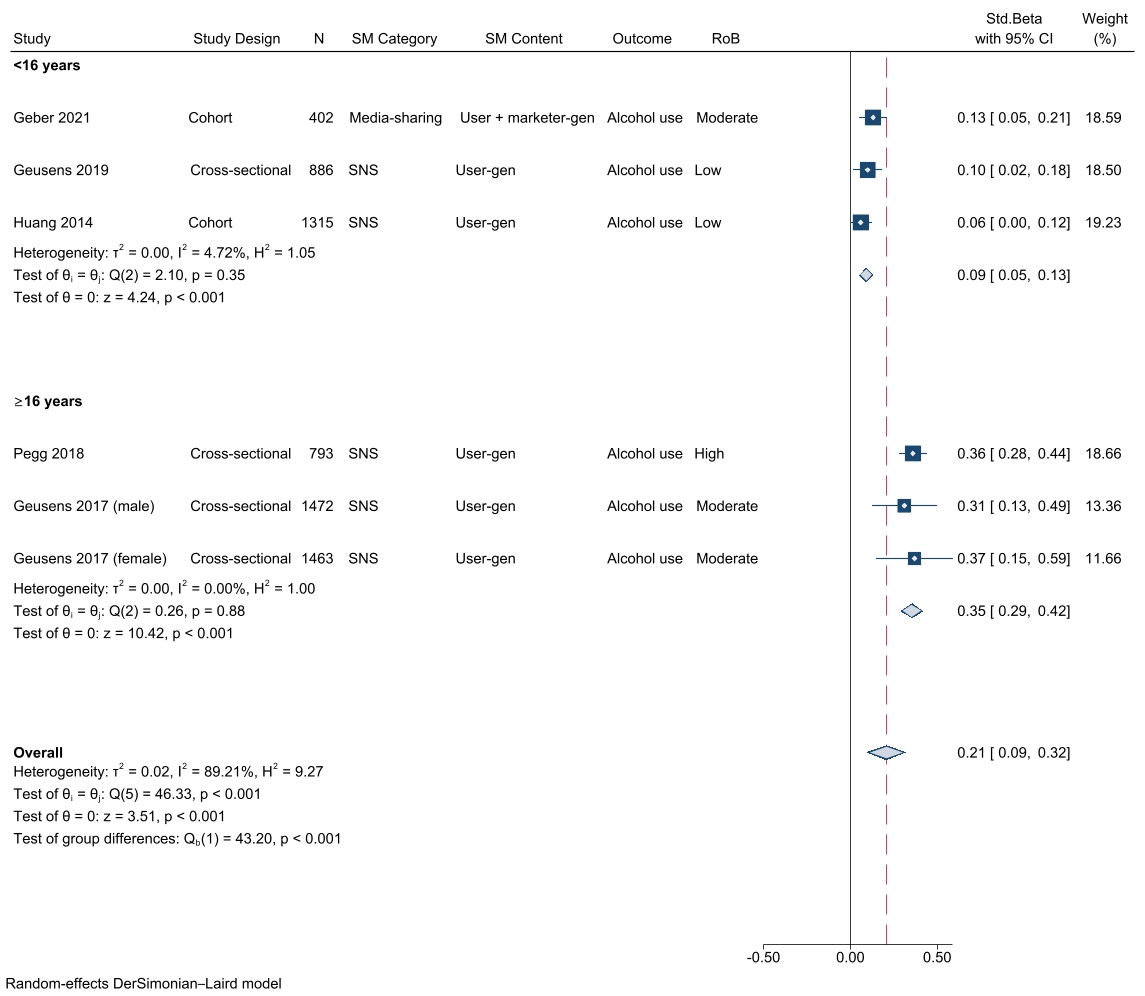

**Legend:** Figure presents forest plot for continuous exposure & continuous outcome subgroup analysis, with standardised beta (Std.Beta) used as common metric. Total number of study participants = 6331. Abbreviation: CI = Confidence interval; N = Number of study participants; Marketer-gen = Marketer-generated content; RoB = Risk of bias; SM = Social media; SNS = Social networking sites; Std. Beta = Standardised beta and User-gen = User-generated content.

##### Figure S. Forest plot for association between frequency of social media use and alcohol use, by average age of study participants

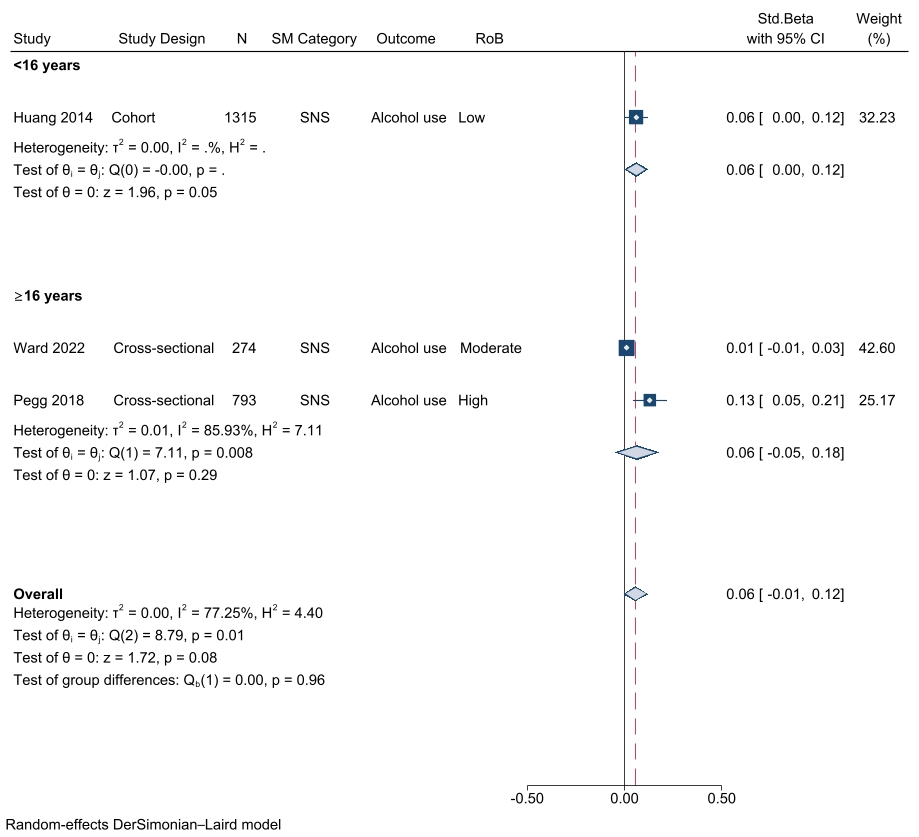

**Legend:** Figure presents forest plot for continuous exposure & continuous outcome subgroup analysis, with standardised beta (Std.Beta) used as common metric. Total number of study participants = 2382. Abbreviation: CI = Confidence interval; N = Number of study participants; RoB = Risk of bias; SM = Social media; SNS = Social networking sites and Std. Beta = Standardised beta.

##### Figure T. Forest plot for association between frequency of social media use and alcohol use, by social media category

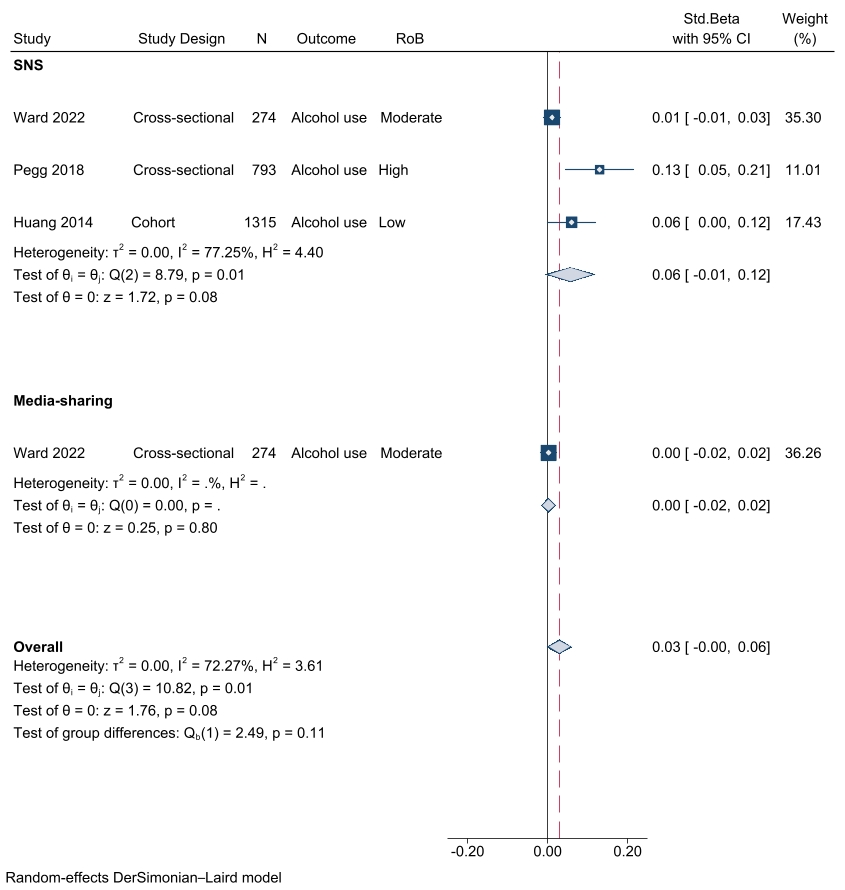

**Legend:** Figure presents forest plot for continuous exposure & continuous outcome subgroup analysis, with standardised beta (Std.Beta) used as common metric. Total number of study participants = 2656. Abbreviation: CI = Confidence interval; N = Number of study participants; RoB = Risk of bias; SM = Social media; SNS = Social networking sites and Std. Beta = Standardised beta.

##### Figure U. Forest plot for association between time spent on social media and alcohol use, by social media category

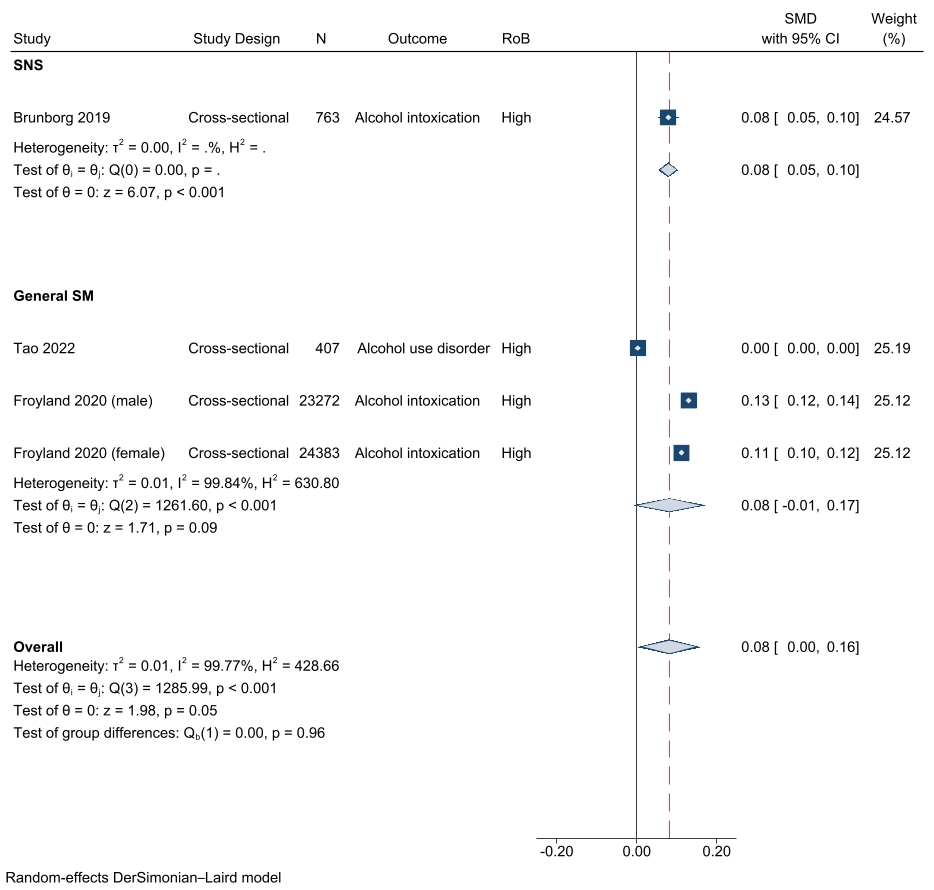

**Legend:** Figure presents forest plot for continuous exposure & continuous outcome subgroup analysis, with standardised mean difference (SMD) used as common metric. Total number of study participants = 48,825. Abbreviation: CI = Confidence interval; N = Number of study participants; RoB = Risk of bias; SM = Social media; SMD = Standardised mean difference and SNS = Social networking sites.

##### Figure V. Forest plot for association between exposure to health-risk behaviour content on social media and alcohol use, by average socioeconomic position of study participants

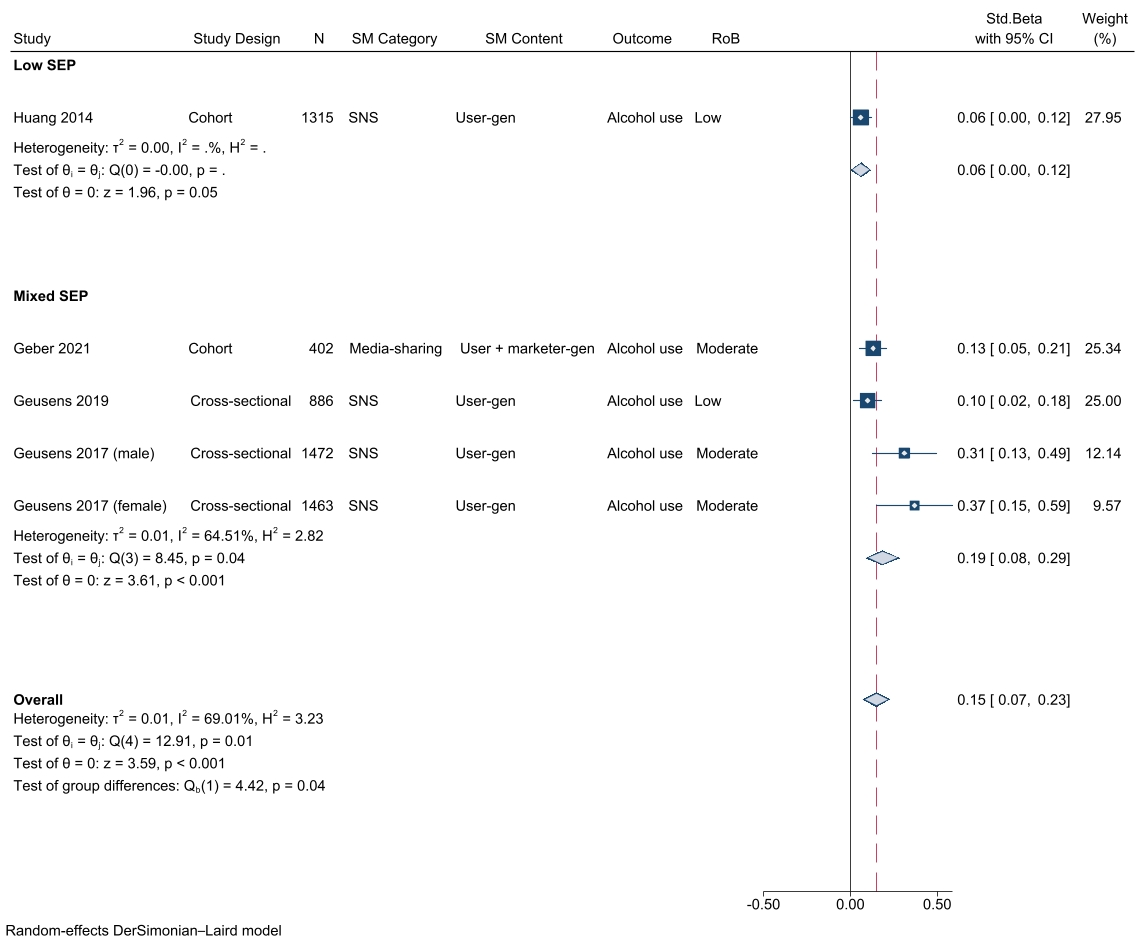

**Legend:** Figure presents forest plot for continuous exposure & continuous outcome subgroup analysis, with standardised beta (Std.Beta) used as common metric. Total number of study participants = 5538. Abbreviation: CI = Confidence interval; N = Number of study participants; RoB = Risk of bias; SEP = Socioeconomic position; SM = Social media; SNS = Social networking sites; Std. Beta = Standardised beta and User-gen = User-generated content.

##### Figure W. Forest plot for association between exposure to health-risk behaviour content on social media and alcohol use, by social media category

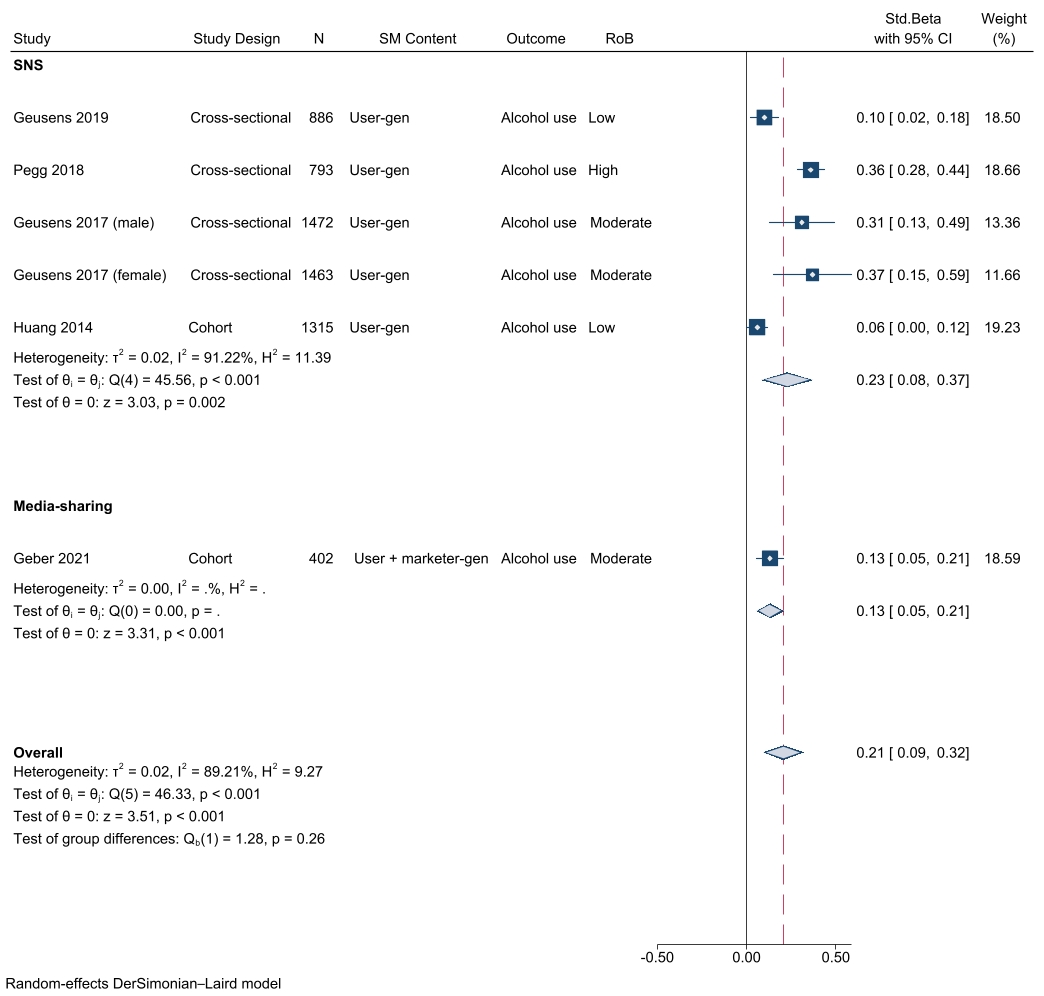

**Legend:** Figure presents forest plot for continuous exposure & continuous outcome subgroup analysis, with standardised beta (Std.Beta) used as common metric. Total number of study participants = 6331. Abbreviation: CI = Confidence interval; N = Number of study participants; Marketer-gen = Marketer-generated content; RoB = Risk of bias; SM = Social media; SNS = Social networking sites; Std. Beta = Standardised beta and User-gen = User-generated content.

#### Drug use

##### Effect direction plot

Figure X demonstrates the effect direction in those studies (n=13) investigating drug use, by exposure. Two studies investigated more than one exposure.^123,152^ For time spent on social media, 5/6 studies (83.3%) reported harmful associations (95% CI 43.6 to 97.0%; participant n=727,788; sign test p=0.22). For frequency of social media use 5/6 studies (83.3%) demonstrated harmful associations (43.6 to 97.0%; participant n=117,645; sign test p=0.22) and for exposure to health-risk behaviour content on social media all studies demonstrated harmful associations (34.2 to 100.0%; study n=2; participant n=1268; insufficient data to conduct sign test). Other social media activities was investigated by one study which demonstrated a harmful association (20.7 to 100.0%; participant n=786; insufficient data to conduct sign test).

##### Figure X. Effect direction plot for studies of the association between social media use and adolescent drug use, by social media exposure. Arrow size indicates sample size; arrow colour indicates study risk of bias

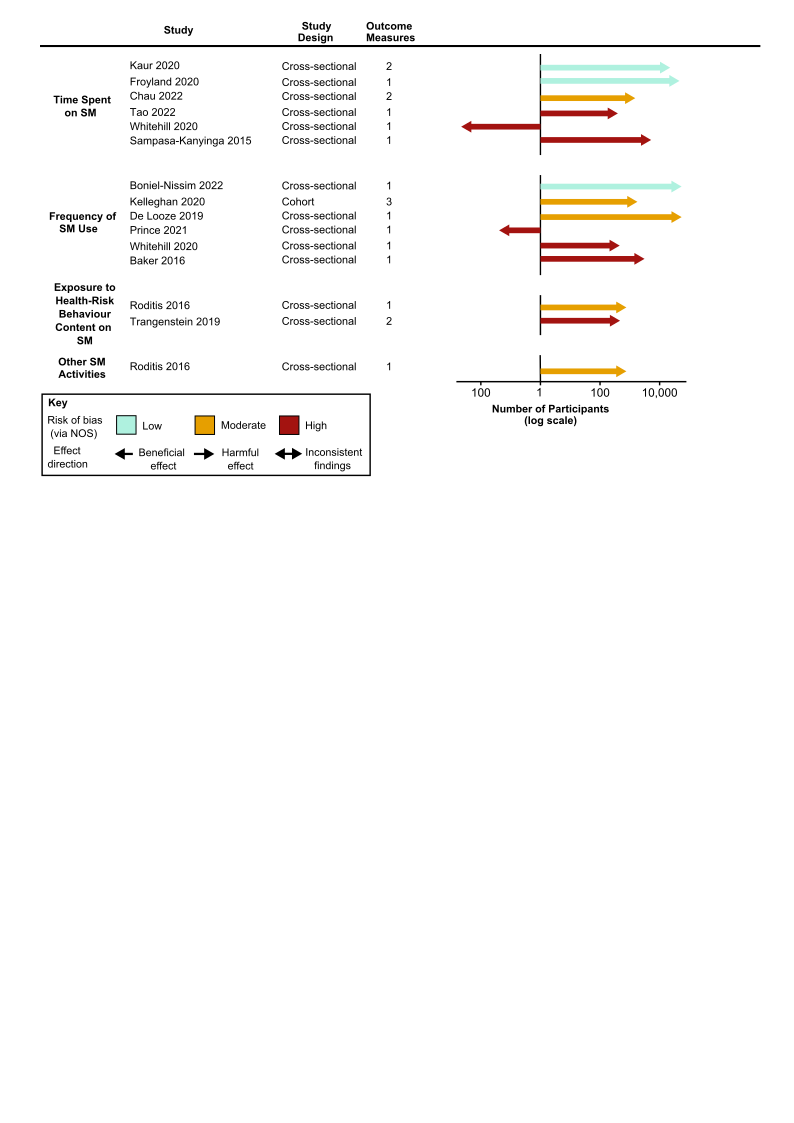

**Legend:** Sample size: represented by the size of the arrow, measured on a log scale. Outcome measure: number of outcome measures synthesised within each study. Studies organised by risk of bias grade, study design, and year of publication. Repeat cross-sectional studies, multiple study populations from different countries, and age subsets originating from the same study reported as separate studies. Abbreviations: NOS = Assessed via adapted Newcastle Ottawa Scale and SM = Social media.

##### Forest plots for meta-analyses and subgroup analyses

##### Figure Y. Forest plot for association between frequency of social media use and drug use

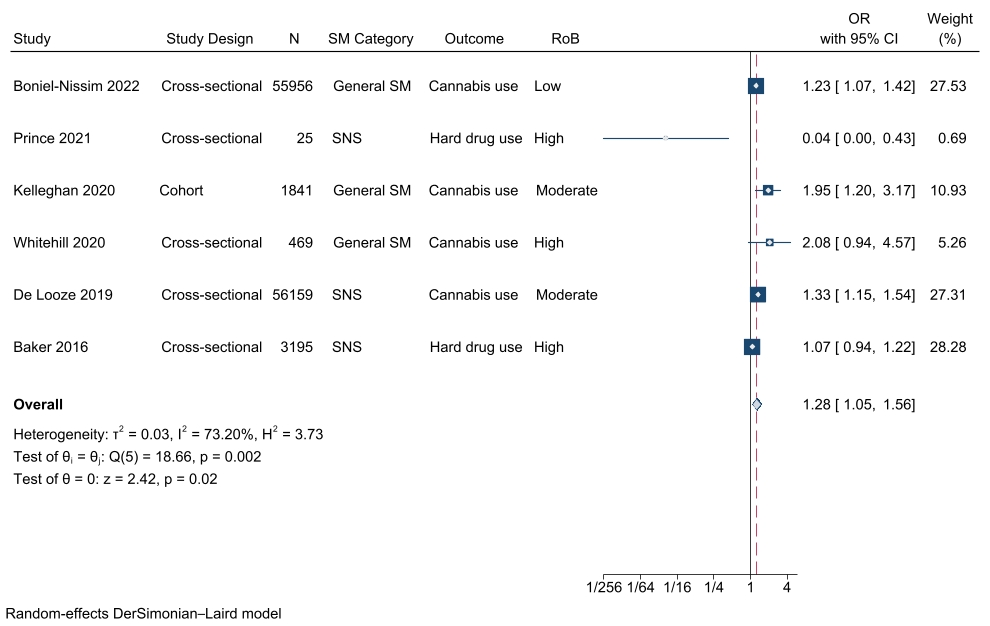

**Legend:** Figure presents forest plot for binary exposure (frequent/daily vs infrequent/non-daily) & binary/continuous outcome meta-analysis, with odds ratio (OR) used as common metric. Total number of study participants = 117,645. Abbreviations: CI = Confidence interval; N = Number of study participants; OR = Odds ratio; RoB = Risk of bias; SM = Social media and SNS = Social networking sites.

##### Figure Z. Forest plot for association between frequency of social media use and drug use, by average age of study participants

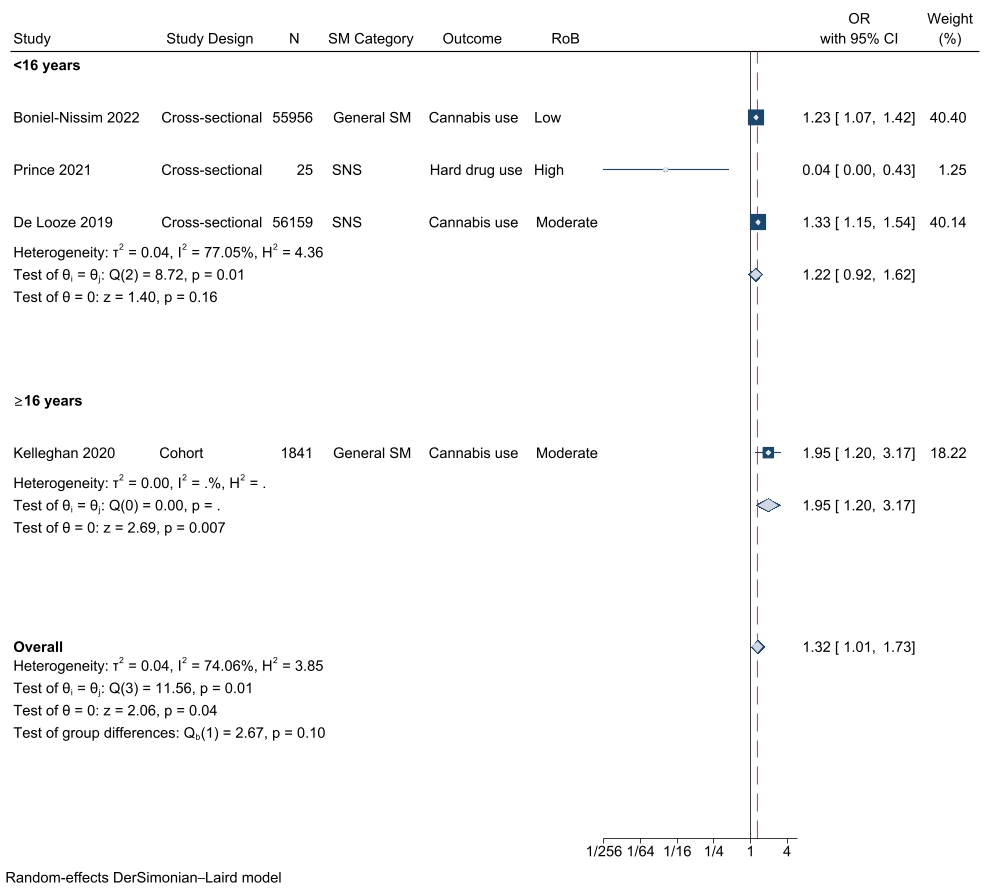

**Legend:** Figure presents forest plot for binary exposure (frequent/daily vs infrequent/non-daily) & binary/continuous outcome meta-analysis, with odds ratio (OR) used as common metric. Total number of study participants = 113,981. Abbreviations: CI = Confidence interval; N = Number of study participants; OR = Odds ratio; RoB = Risk of bias; SM = Social media and SNS = Social networking sites.

##### Figure AA. Forest plot for association between frequency of social media use and drug use, by social media platform

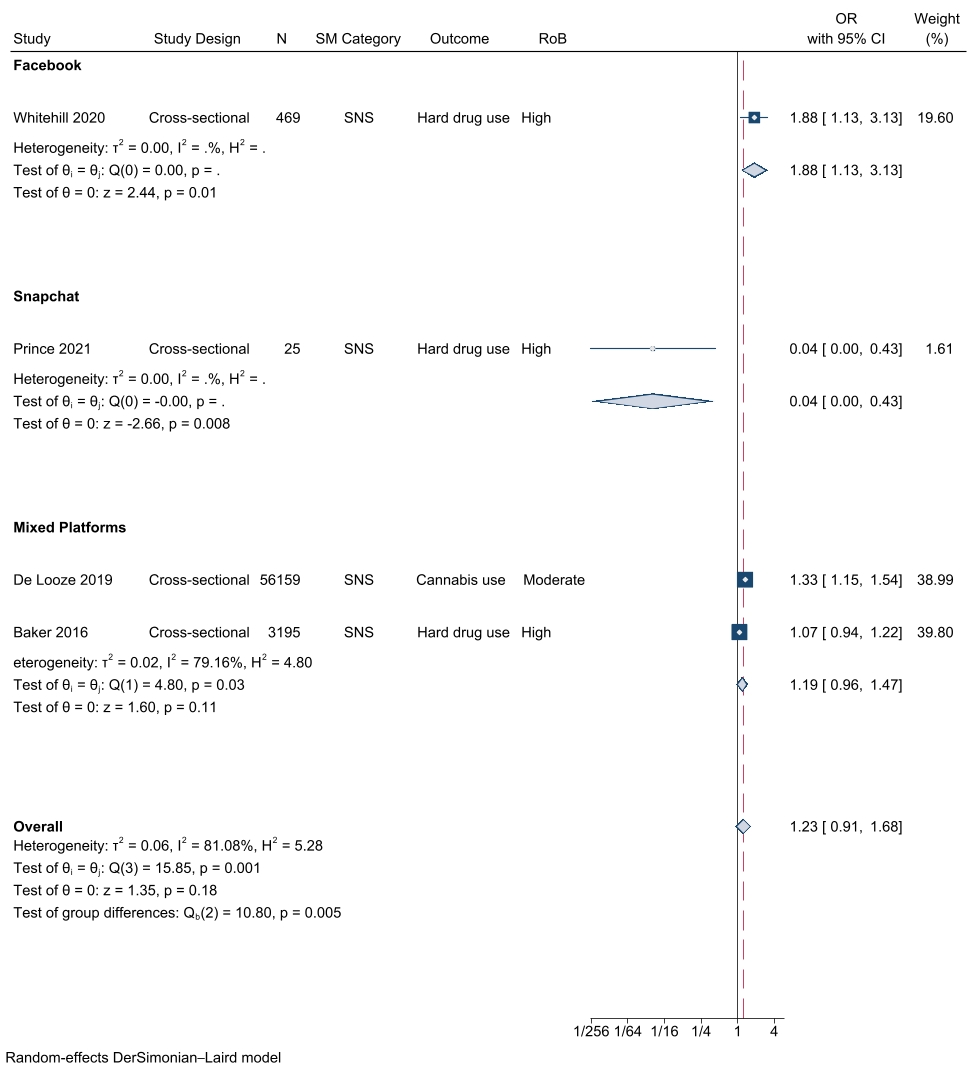

**Legend:** Figure presents forest plot for binary exposure (frequent/daily vs infrequent/non-daily) & binary/continuous outcome meta-analysis, with odds ratio (OR) used as common metric. Total number of study participants = 59,848. Abbreviations: CI = Confidence interval; N = Number of study participants; OR = Odds ratio; RoB = Risk of bias; SM = Social media and SNS = Social networking sites.

##### Figure AB. Forest plot for association between frequency of social media use and drug use, by social media category

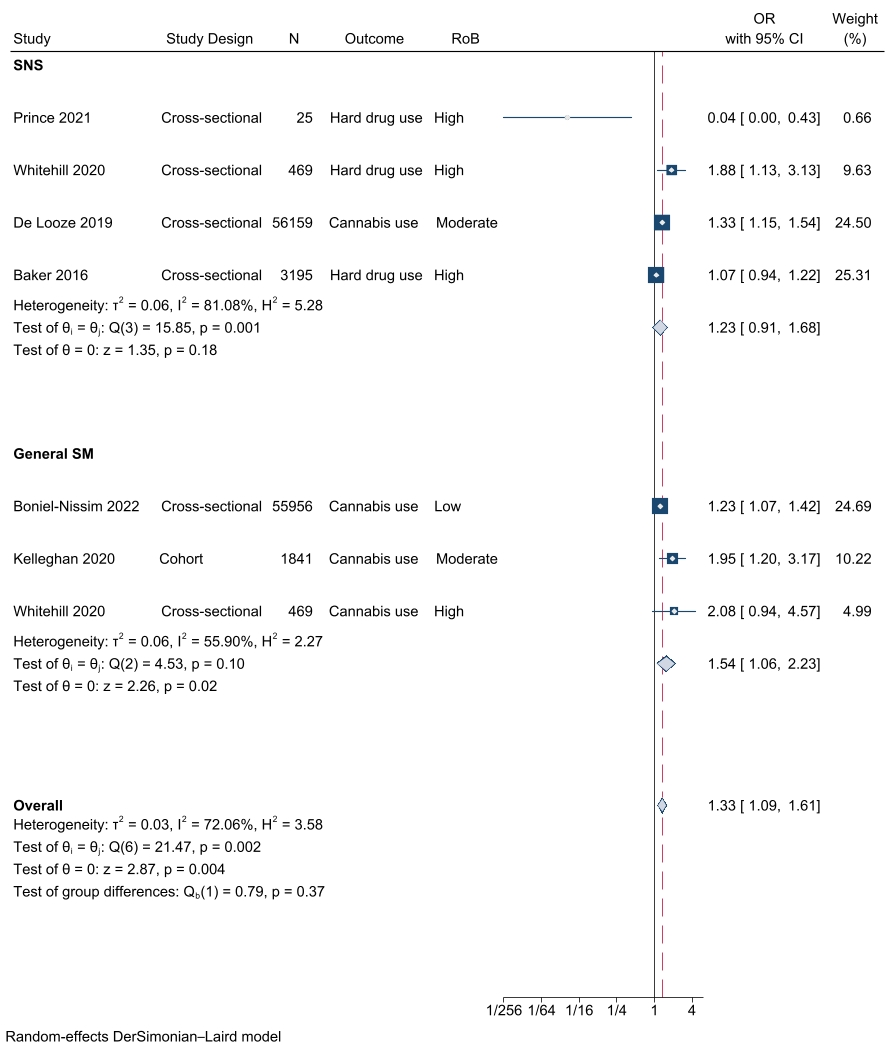

**Legend:** Figure presents forest plot for binary exposure (frequent/daily vs infrequent/non-daily) & binary/continuous outcome meta-analysis, with odds ratio (OR) used as common metric. Total number of study participants = 118,114. Abbreviations: CI = Confidence interval; N = Number of study participants; OR = Odds ratio; RoB = Risk of bias; SM = Social media and SNS = Social networking sites.

##### Figure AC. Forest plot for association between time spent on social media and drug use

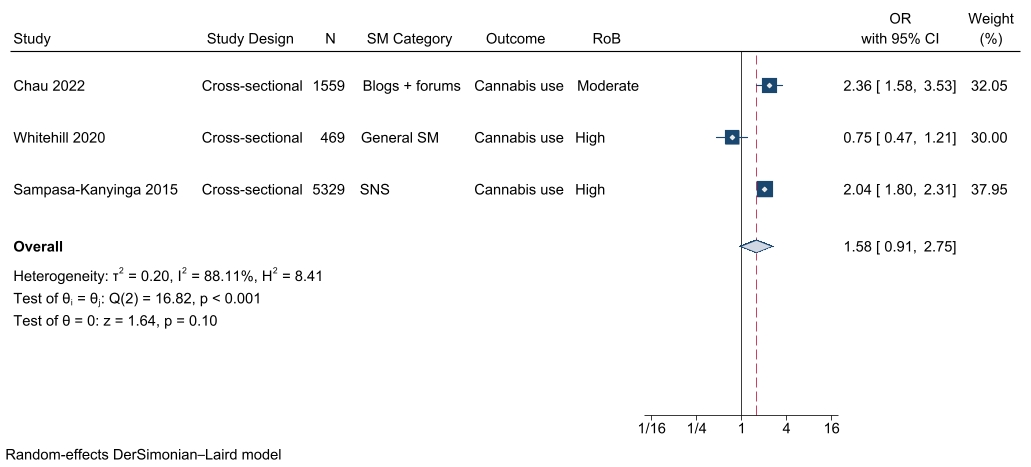

**Legend:** Figure presents forest plot for binary exposure (≤1hrs/day vs >1hr/day) & binary/continuous outcome meta-analysis, with odds ratio (OR) used as common metric. Total number of study participants = 7357. Abbreviations: CI = Confidence interval; N = Number of study participants; OR = Odds ratio; RoB = Risk of bias; SM = Social media and SNS = Social networking sites.

##### Figure AD. Forest plot for association between time spent on social media and drug use

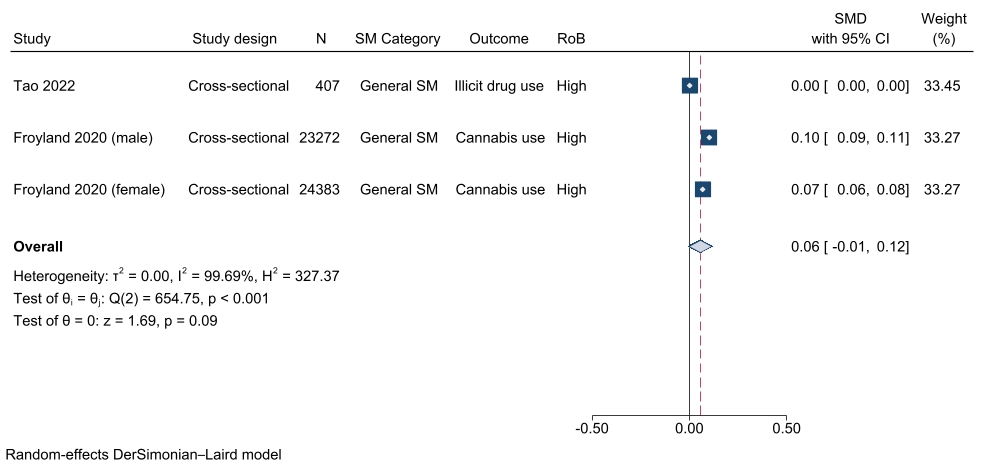

**Legend:** Figure presents forest plot for continuous exposure & continuous meta-analysis, with standardised mean difference (SMD) used as common metric. Total number of study participants = 48,062. Abbreviations: CI = Confidence interval; N = Number of study participants; OR = Odds ratio; RoB = Risk of bias; SM = Social media and SMD = Standardised mean difference.

#### Tobacco use

##### Effect direction plot

Figure AE illustrates the effect direction in those studies investigating tobacco use, by exposure. One study investigated more than one exposure.^86^ For time spent on social media all studies demonstrated harmful associations of social media use (95% CI 43.9 to 100%; study n=3; participant n=7216; sign test p=0.25), as did all studies investigating exposure to health-risk behaviour content (51.0 to 100%; study n = 4; participant n=24,197; sign test p=0.13). For frequency of social media use 9/10 studies (90.0%) demonstrated harmful associations (59.6 to 98.2%; participant n=431,501; sign test p=0.02). Other social media activities was investigated by one study which demonstrated a harmful association (0.00 to 79.3%; participant n=5851; insufficient data to conduct sign test).

##### Figure AE. Effect direction plot for studies of the association between social media use and adolescent tobacco use, by social media exposure. Arrow size indicates sample size; arrow colour indicates study risk of bias

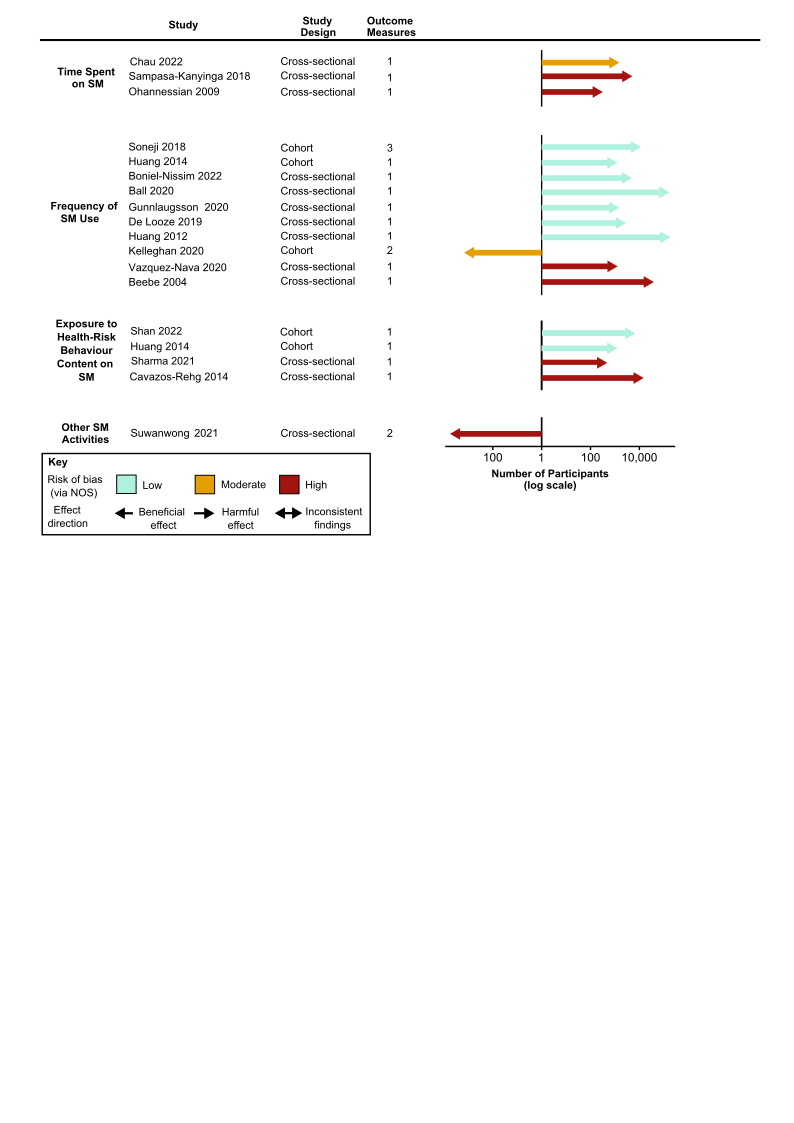

**Legend:** Sample size: represented by the size of the arrow, measured on a log scale. Outcome measure: number of outcome measures synthesised within each study. Studies organised by risk of bias grade, study design, and year of publication. Repeat cross-sectional studies, multiple study populations from different countries, and age subsets originating from the same study reported as separate studies. Abbreviations: NOS = Assessed via adapted Newcastle Ottawa Scale and SM = Social media.

##### Forest plots for meta-analyses and subgroup analyses

##### Figure AF. Forest plot for association between frequency of social media use and tobacco use

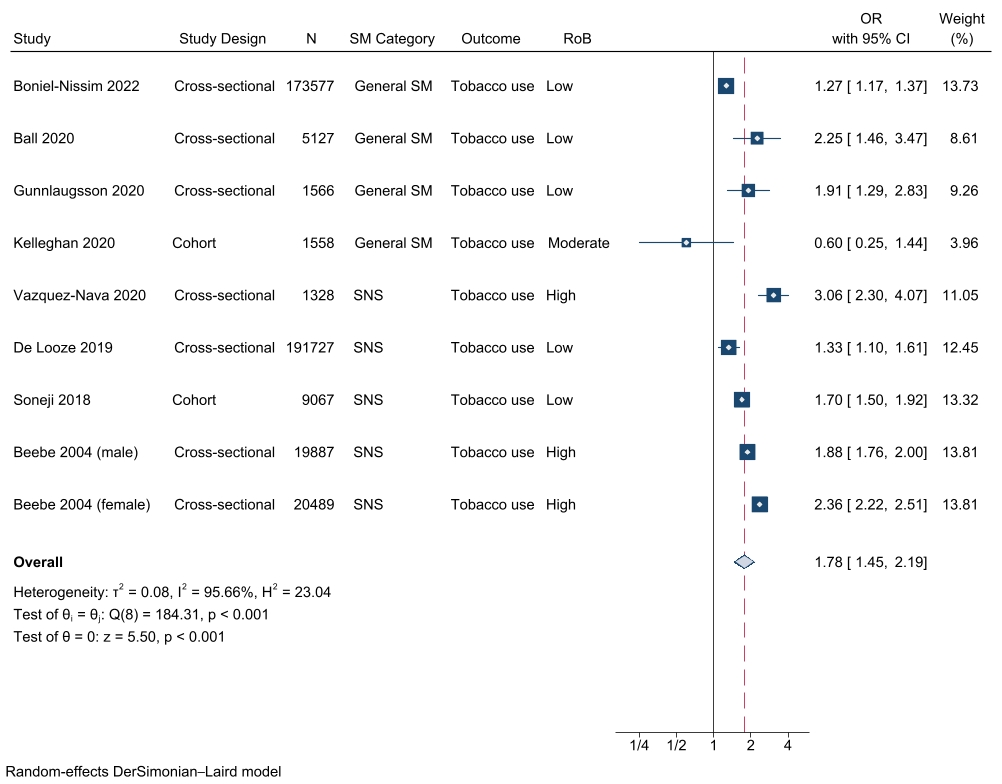

**Legend:** Figure presents forest plot for binary exposure (frequent vs infrequent) & binary/continuous outcome meta-analysis, with odds ratio (OR) used as common metric. Total number of study participants = 424,326. Abbreviations: CI = Confidence interval; N = Number of study participants; OR = Odds ratio; RoB = Risk of bias; SM = Social media and SNS = Social networking sites.

##### Figure AG. Forest plot for association between exposure to health-risk behaviour content on social media and tobacco use

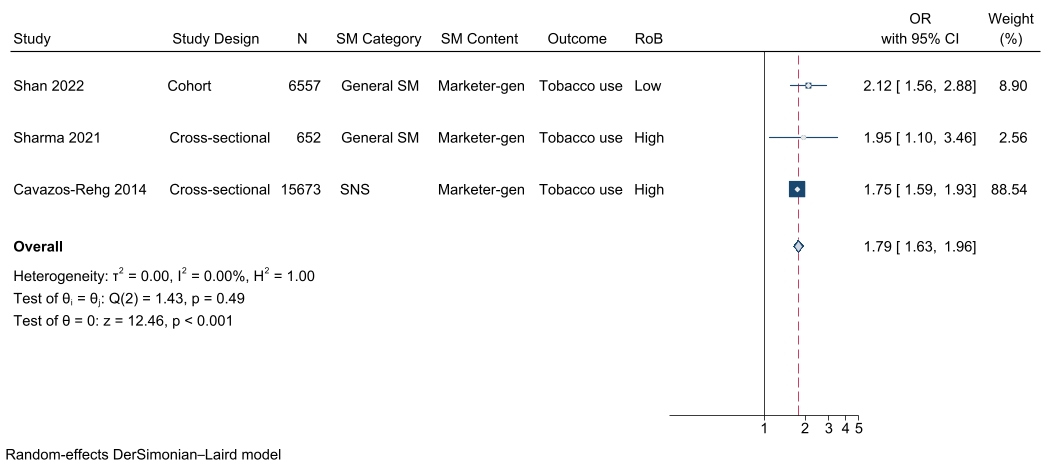

**Legend:** Figure presents forest plot for binary exposure (exposed vs unexposed) & binary/continuous outcome meta-analysis, with odds ratio (OR) used as common metric. Total number of study participants = 22,882. Abbreviations: CI = Confidence interval; Markter-gen = Marketer-generated content; N = Number of study participants; OR = Odds ratio; RoB = Risk of bias; SM = Social media and SNS = Social networking sites.

##### Figure AH. Forest plot for association between frequency of social media use and tobacco use, by average age of study participants

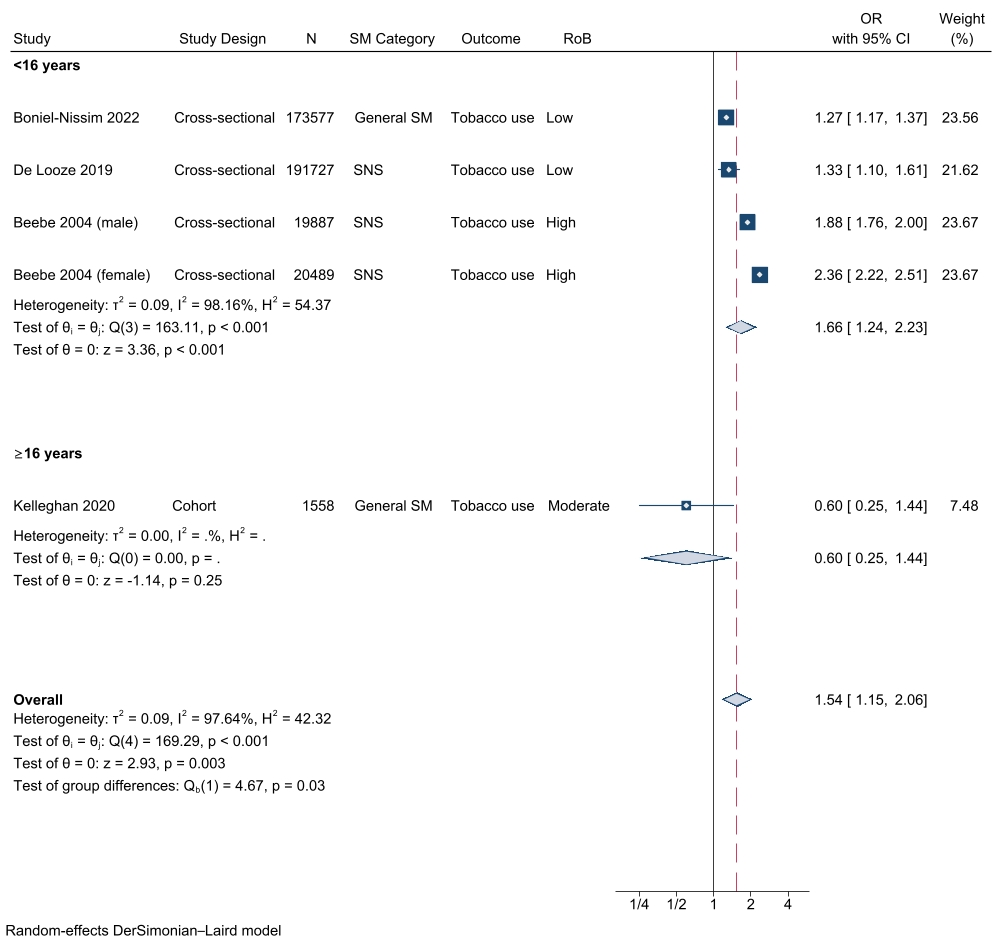

**Legend:** Figure presents forest plot for binary exposure (frequent vs infrequent) & binary/continuous outcome subgroup analysis, with odds ratio (OR) used as common metric. Total number of study participants = 407,238. Abbreviations: CI = Confidence interval; N = Number of study participants; OR = Odds ratio; RoB = Risk of bias; SM = Social media and SNS = Social networking sites

##### Figure AI. Forest plot for association between frequency of social media use and tobacco use, by development status of study setting^a^

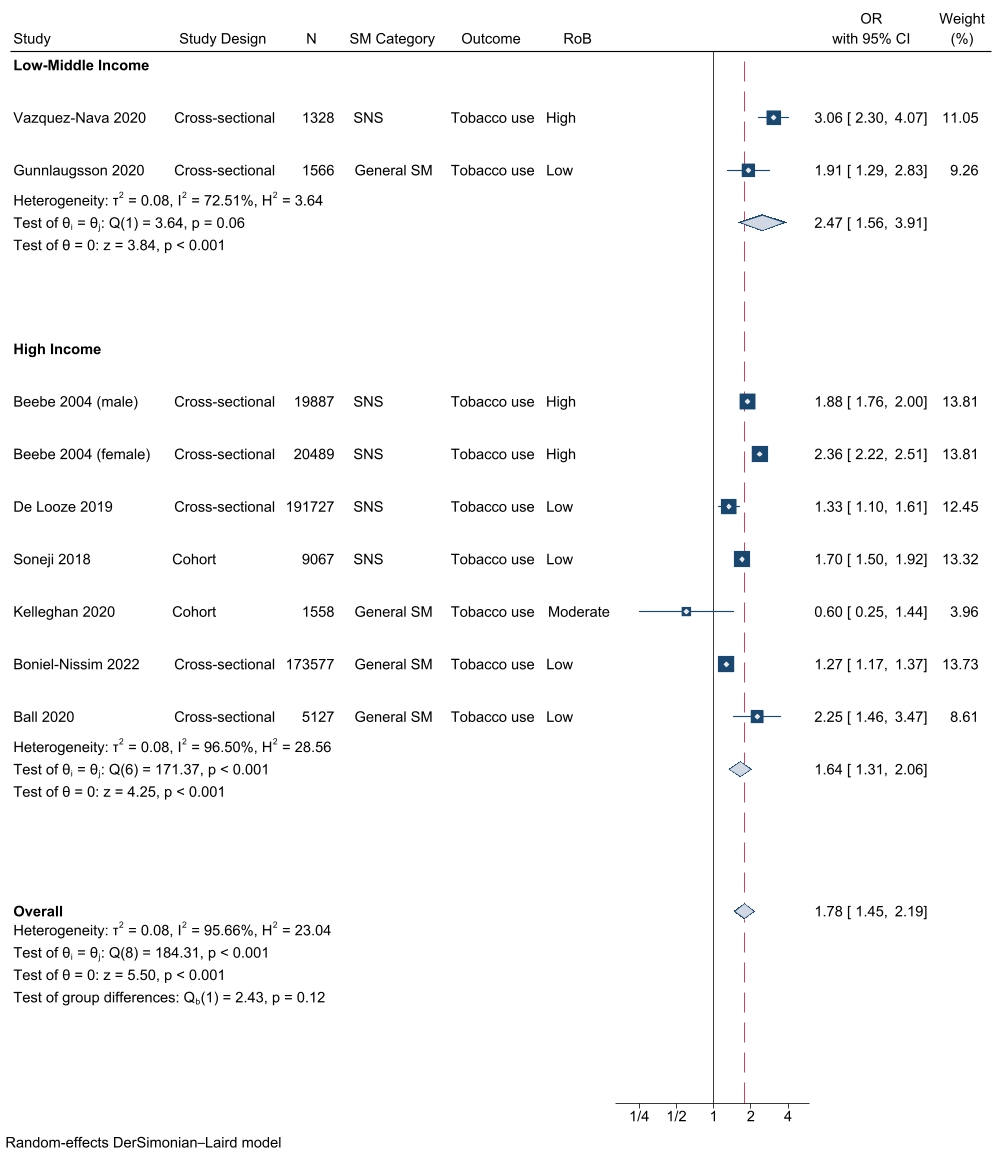

**Legend:** Figure presents forest plot for binary exposure (frequent vs infrequent) & binary/continuous outcome subgroup analysis, with odds ratio (OR) used as common metric. ^a^ Development status classified as per the World Bank Country Income Level Classification.^160^ Total number of study participants = 424,326. Abbreviations: CI = Confidence interval; N = Number of study participants; OR = Odds ratio; RoB = Risk of bias; SM = Social media and SNS = Social networking sites.

##### Figure AJ. Forest plot for association between frequency of social media use and tobacco use, by social media category

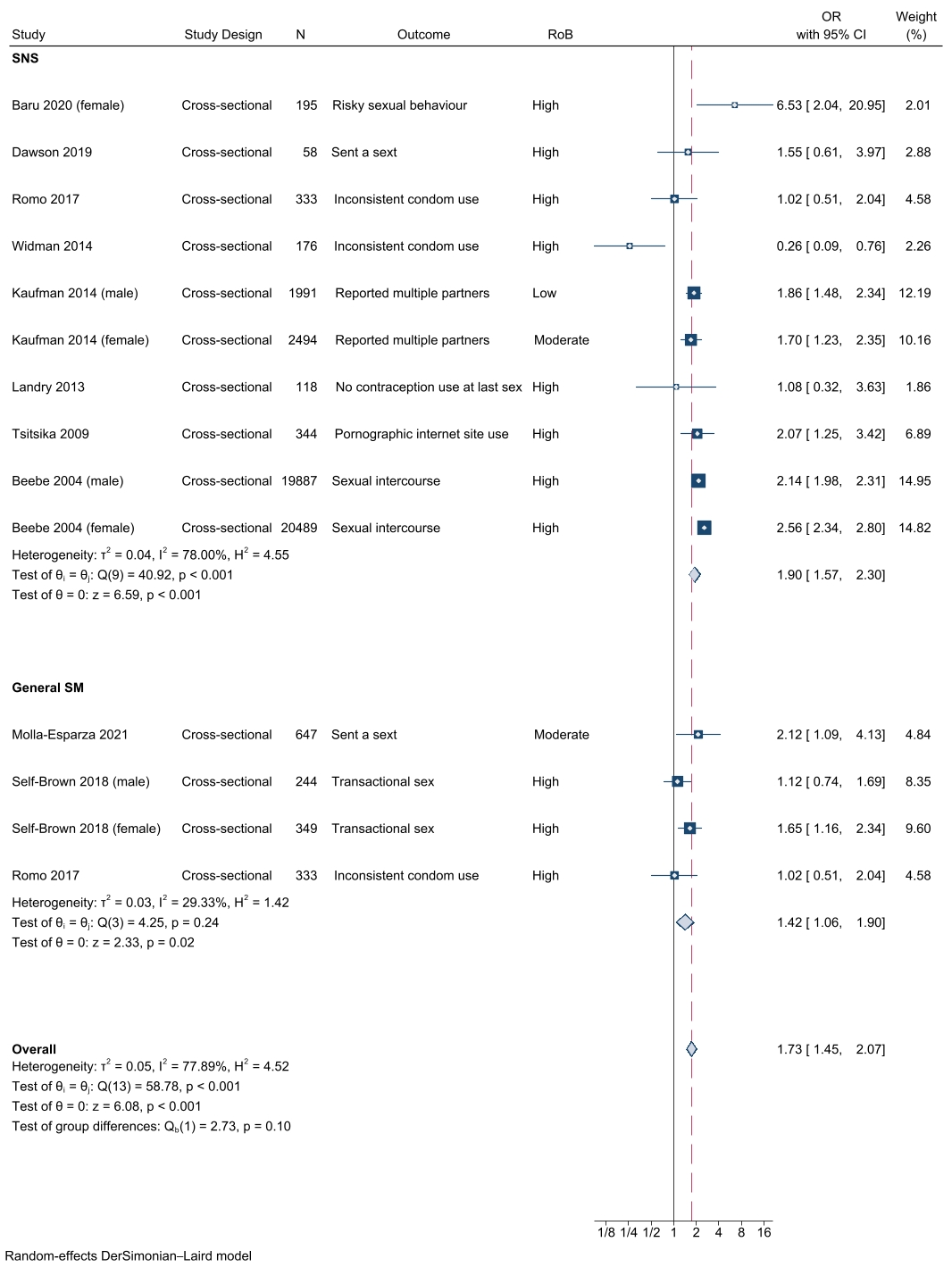

**Legend:** Figure presents forest plot for binary exposure (frequent vs infrequent) & binary/continuous outcome subgroup analysis, with odds ratio (OR) used as common metric. Total number of study participants = 47,658. Abbreviations: CI = Confidence interval; N = Number of study participants; OR = Odds ratio; RoB = Risk of bias; SM = Social media and SNS = Social networking sites

##### Figure AK. Forest plot for association between exposure to health-risk behaviour content on social media and tobacco use, by development status of study setting^a^

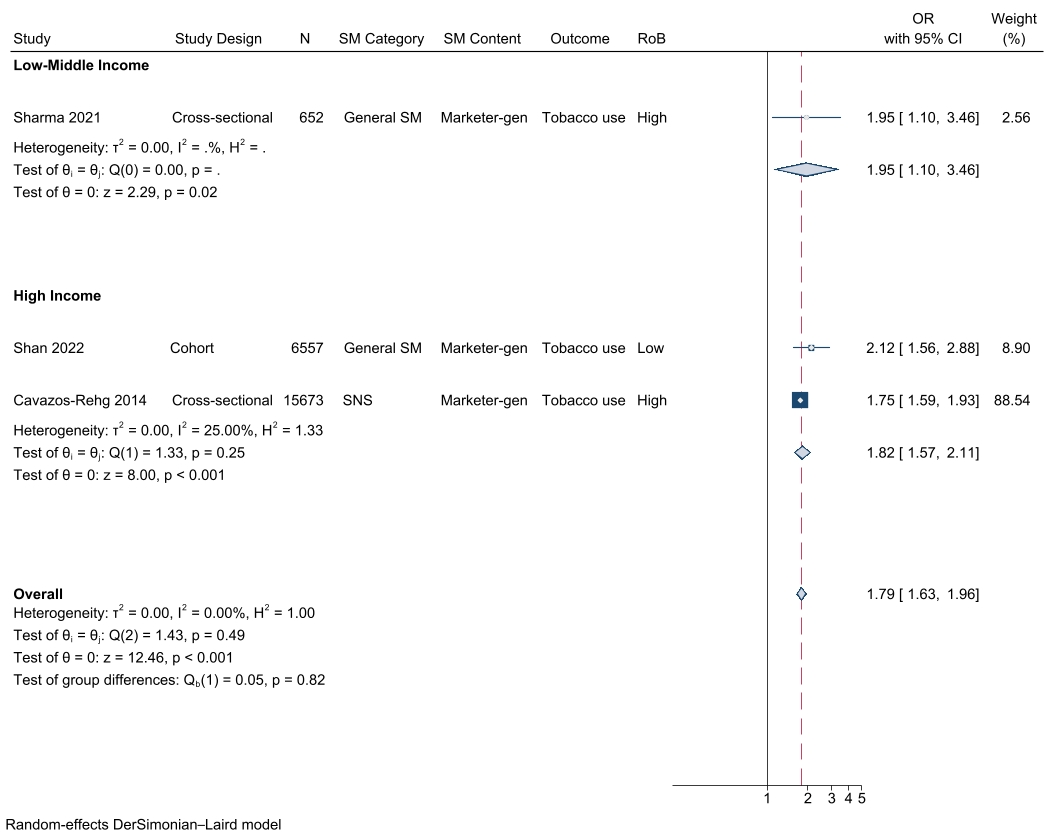

**Legend:** Figure presents forest plot for binary exposure (exposed vs unexposed) & binary/continuous outcome sensitivity analysis, with odds ratio (OR) used as common metric. ^a^ Development status classified as per the World Bank Country Income Level Classification.^160^ Total number of study participants = 22,882. Abbreviations: CI = Confidence interval; Marketer-gen = Marketer-generated content; N = Number of study participants; OR = Odds ratio; RoB = Risk of bias; SM = Social media and SNS = Social networking sites.

##### Figure AL. Forest plot for association between exposure to health-risk behaviour content on social media and tobacco use, by social media category

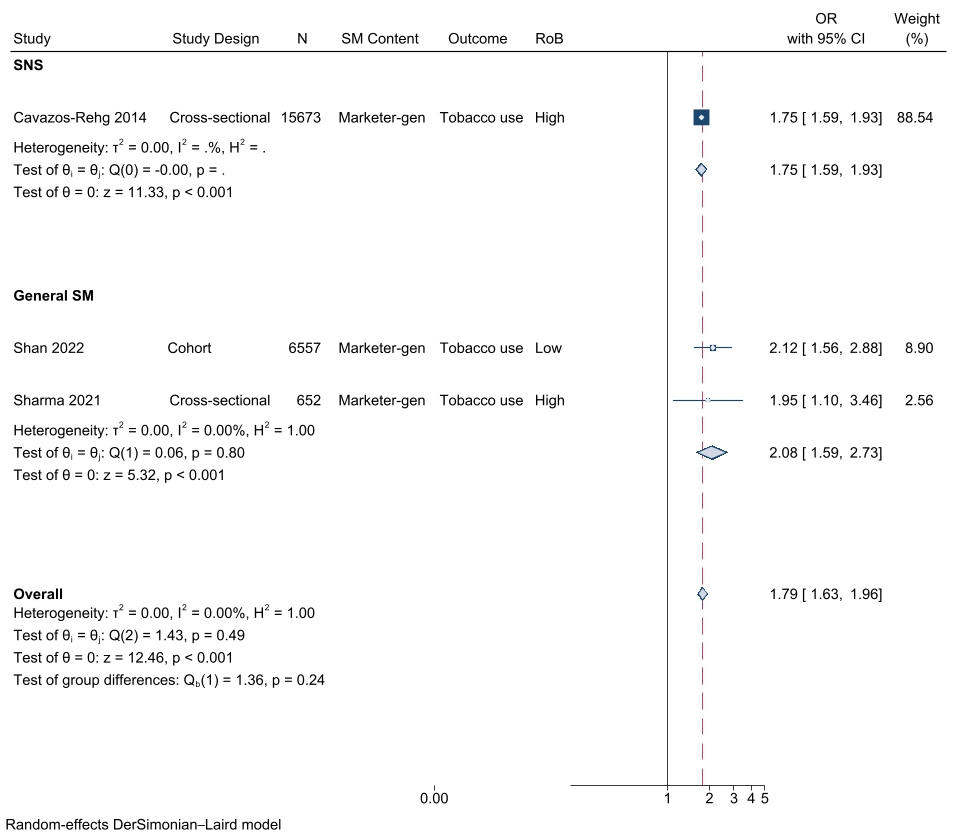

**Legend:** Figure presents forest plot for binary exposure (exposed vs unexposed) & binary/continuous outcome sensitivity analysis, with odds ratio (OR) used as common metric. Total number of study participants = 22,882. Abbreviations: CI = Confidence interval; Marketer-gen = Marketer-generated content; N = Number of study participants; OR = Odds ratio; RoB = Risk of bias; SM = Social media and SNS = Social networking sit

#### Use of electronic nicotine delivery systems (ENDS)

##### Effect direction plot

Figure AM demonstrates the effect direction in those studies (n=9) investigating use of electronic nicotine delivery systems (ENDS), by exposure. For time spent on social media, all studies reported harmful associations (95% CI 34.2 to 100.0%; study n=2; participant n=9821; insufficient data to conduct sign test), for frequency of social media use, 2/3 studies (66.7%) demonstrated harmful associations (20.8 to 93.9%; participant n=18,047; sign test p=1.00) and for exposure to health-risk behaviour content on social media all studies reported harmful effects (51.0 to 100.0%; study n=4; participant n=721,322; sign test p=1.00).

##### Figure AM. Effect direction plot for studies of the association between social media and adolescent use of electronic nicotine delivery systems, by social media exposure. Arrow size indicates sample size; arrow colour indicates study risk of bias

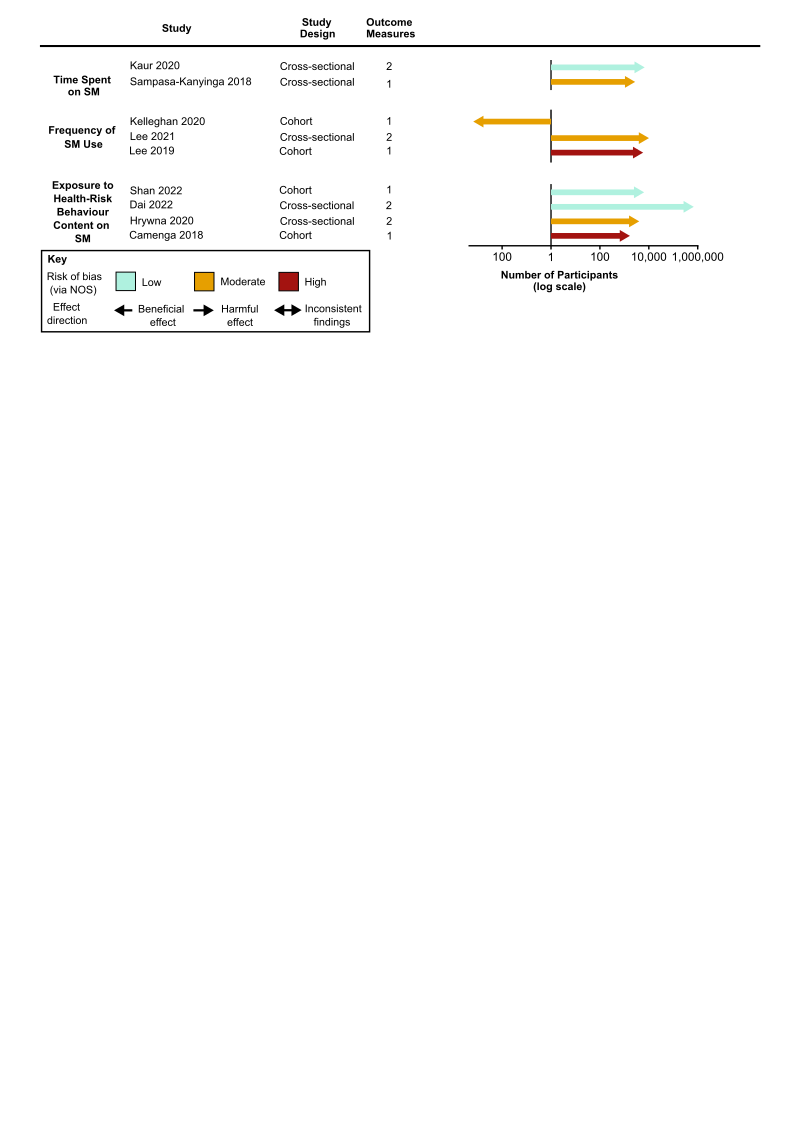

**Legend:** Sample size: represented by the size of the arrow, measured on a log scale. Outcome measure: number of outcome measures synthesised within each study. Studies organised by risk of bias grade, study design, and year of publication. Repeat cross-sectional studies, multiple study populations from different countries, and age subsets originating from the same study reported as separate studies. Abbreviations: NOS = Assessed via adapted Newcastle Ottawa Scale and SM = Social media.

##### Forest plots for meta-analyses and subgroup analyses

##### Figure AN. Forest plot for association between exposure to health-risk behaviour content on social media and use of electronic nicotine delivery systems

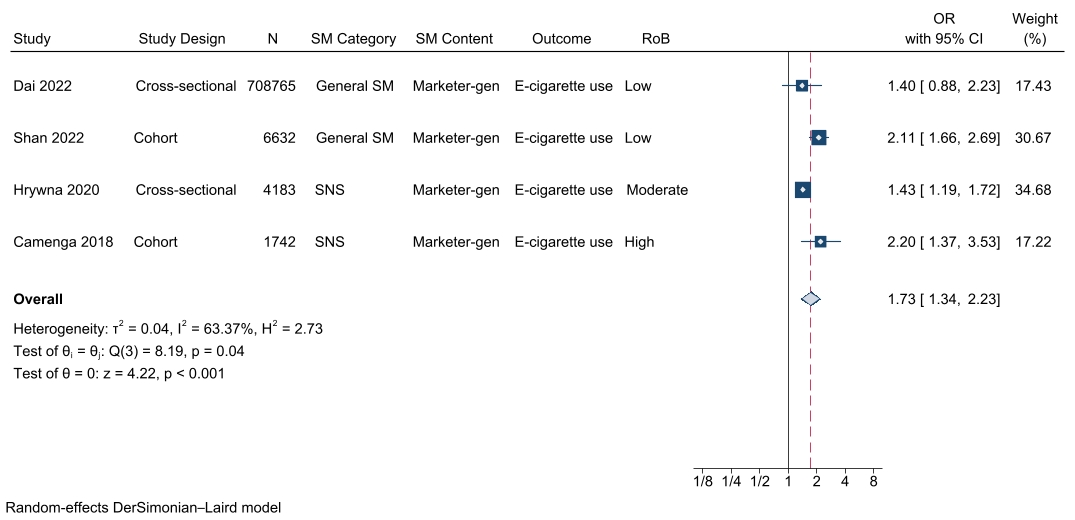

**Legend:** Figure presents forest plot for binary exposure (exposed vs unexposed) & binary/continuous outcome meta-analysis, with odds ratio (OR) used as common metric. Total number of study participants = 721,322. Abbreviations: CI = Confidence interval; N = Number of study participants; OR = Odds ratio; RoB = Risk of bias; SM = Social media and SNS = Social networking sites

##### Figure AO. Forest plot for association between exposure to health-risk behaviour content on social media and use of electronic nicotine delivery systems, by social media category

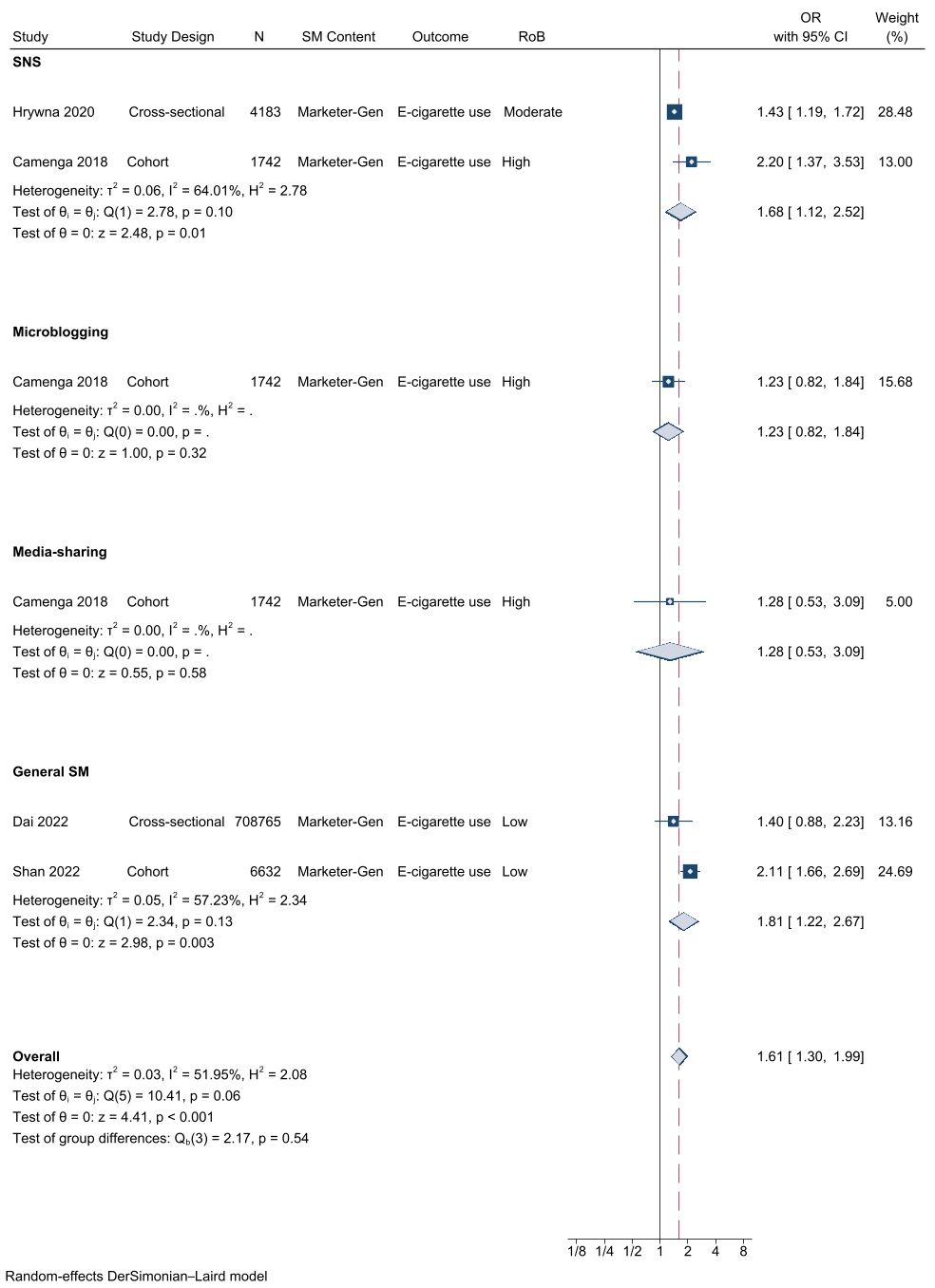

**Legend:** Figure presents forest plot for binary exposure (exposed vs unexposed) & binary/continuous outcome subgroup analysis, with odds ratio (OR) used as common metric. Total number of study participants = 724,716. Abbreviations: CI = Confidence interval; N = Number of study participants; OR = Odds ratio; RoB = Risk of bias; SM = Social media and SNS = Social networking sites

##### Figure AP. Forest plot for association between exposure to health-risk behaviour content on social media and use of electronic nicotine delivery systems, by social media platform

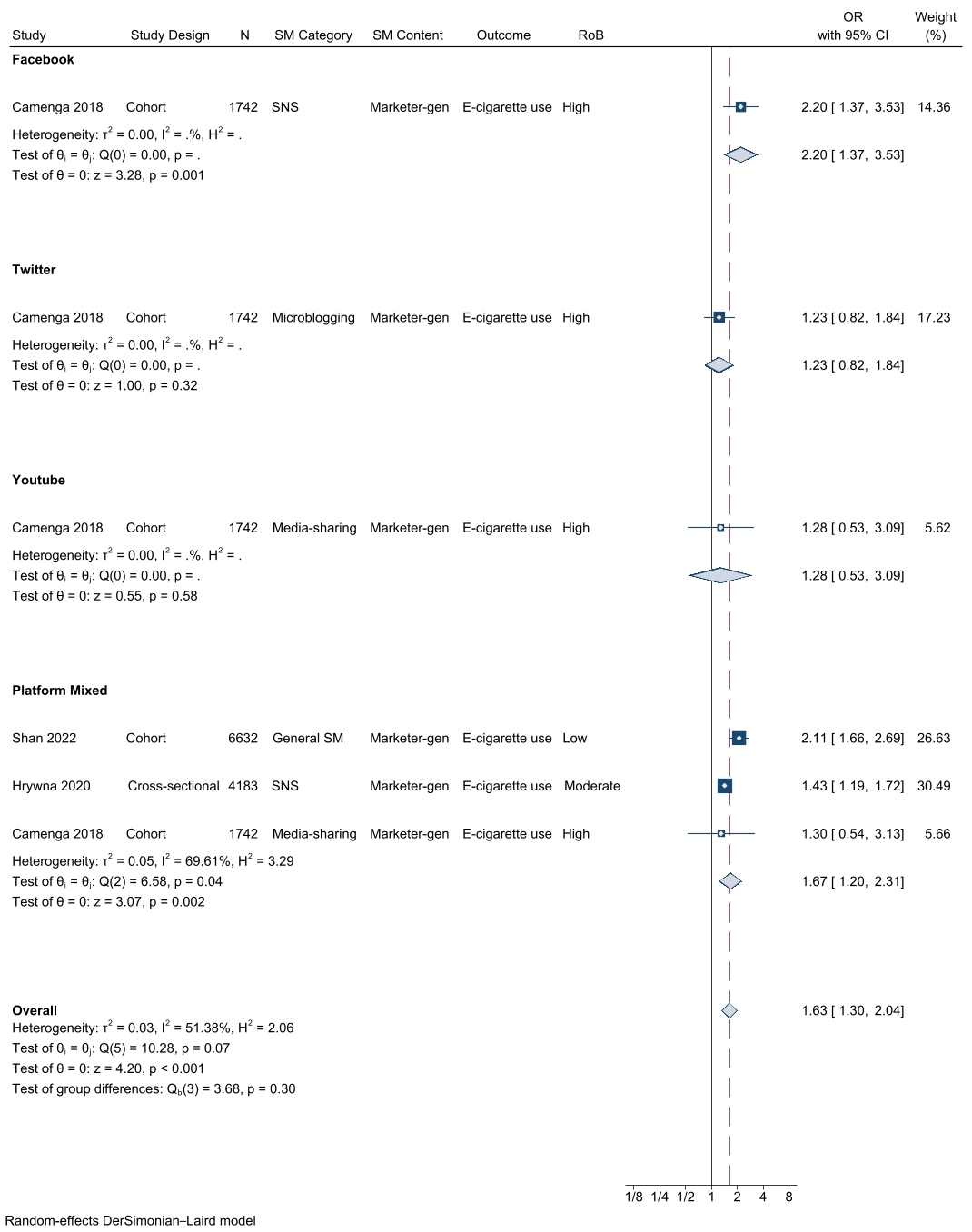

**Legend:** Figure presents forest plot for binary exposure (exposed vs unexposed) & binary/continuous outcome subgroup analysis, with odds ratio (OR) used as common metric. Total number of study participants = 17,783. Abbreviations: CI = Confidence interval; N = Number of study participants; OR = Odds ratio; RoB = Risk of bias; SM = Social media and SNS = Social networking sites

#### Sexual risk behaviour

##### Effect direction plot

Figure AQ demonstrates the effect direction in those studies investigating sexual risk behaviour, by exposure. Six studies investigated more than one exposure type. ^59,89,97,114,149,112^ After excluding 1 study with inconsistent findings (participant n=333),^124^ for time spent on social media, 5/6 studies (83.3%) reported harmful associations (95% CI 43.6 to 97.0%; participant n=13,528; sign test p=0.22), 17/18 studies (94.5%) reported harmful associations for frequency of social media use (74.2 to 99.0%; participant n=53,850; sign test p < 0.001), all studies reported harmful associations for exposure to health-risk behaviour content on social media (34.2 to 100.0%; study n=2; participant n=138; insufficient data to conduct sign test), and 4/5 studies (80.0%) reported harmful associations of engagement in other social media activities (37.6 to 96.4%; participant n=6141; sign test p=0.38).

##### Figure AQ. Effect direction plot for studies of the association between social media use and adolescent engagement in sexual risk behaviour, by social media exposure. Arrow size indicates sample size; arrow colour indicates study risk of bias

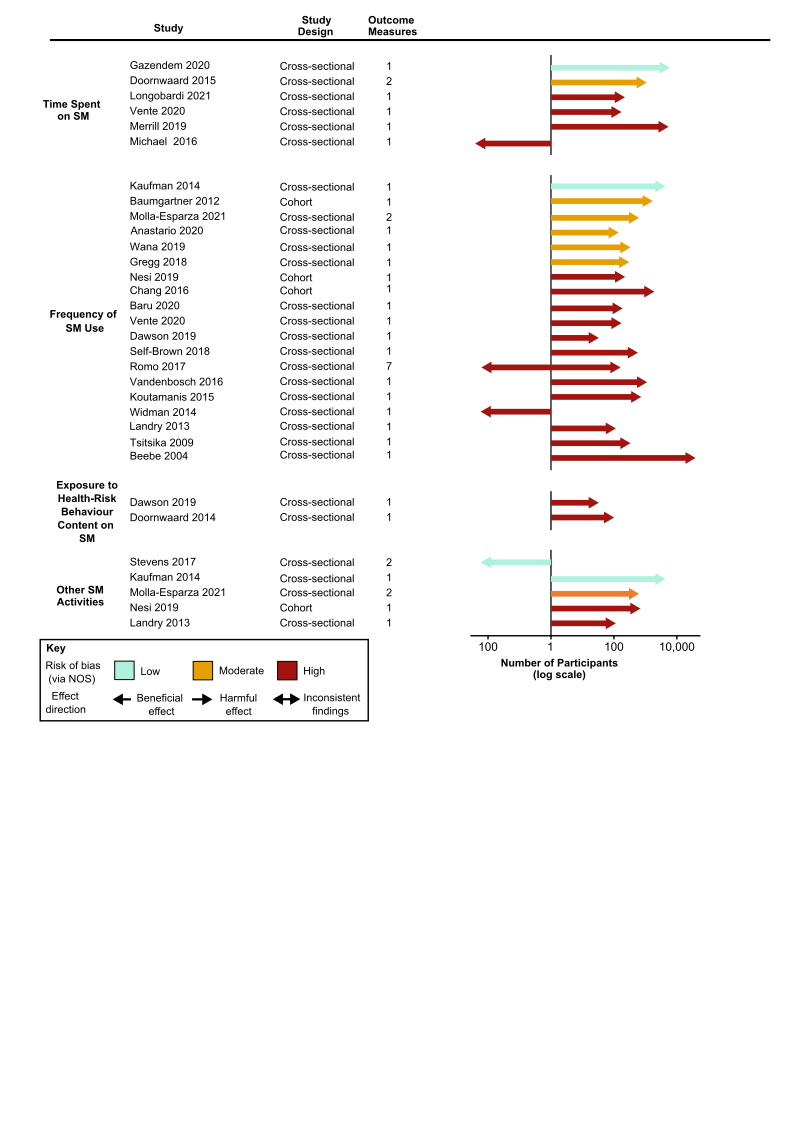

**Legend:** Sample size: represented by the size of the arrow, measured on a log scale. Outcome measure: number of outcome measures synthesised within each study. Studies organised by risk of bias grade, study design, and year of publication. Repeat cross-sectional studies, multiple study populations from different countries, and age subsets originating from the same study reported as separate studies. Abbreviations: NOS = Assessed via adapted Newcastle Ottawa Scale and SM = Social media. Note: Gregg et al, exposure measure assessed texting alongside social media use.

##### Forest plots for meta-analyses and subgroup analyses

##### Figure AR. Forest plot for association between frequency of social media use and sexual risk behaviour

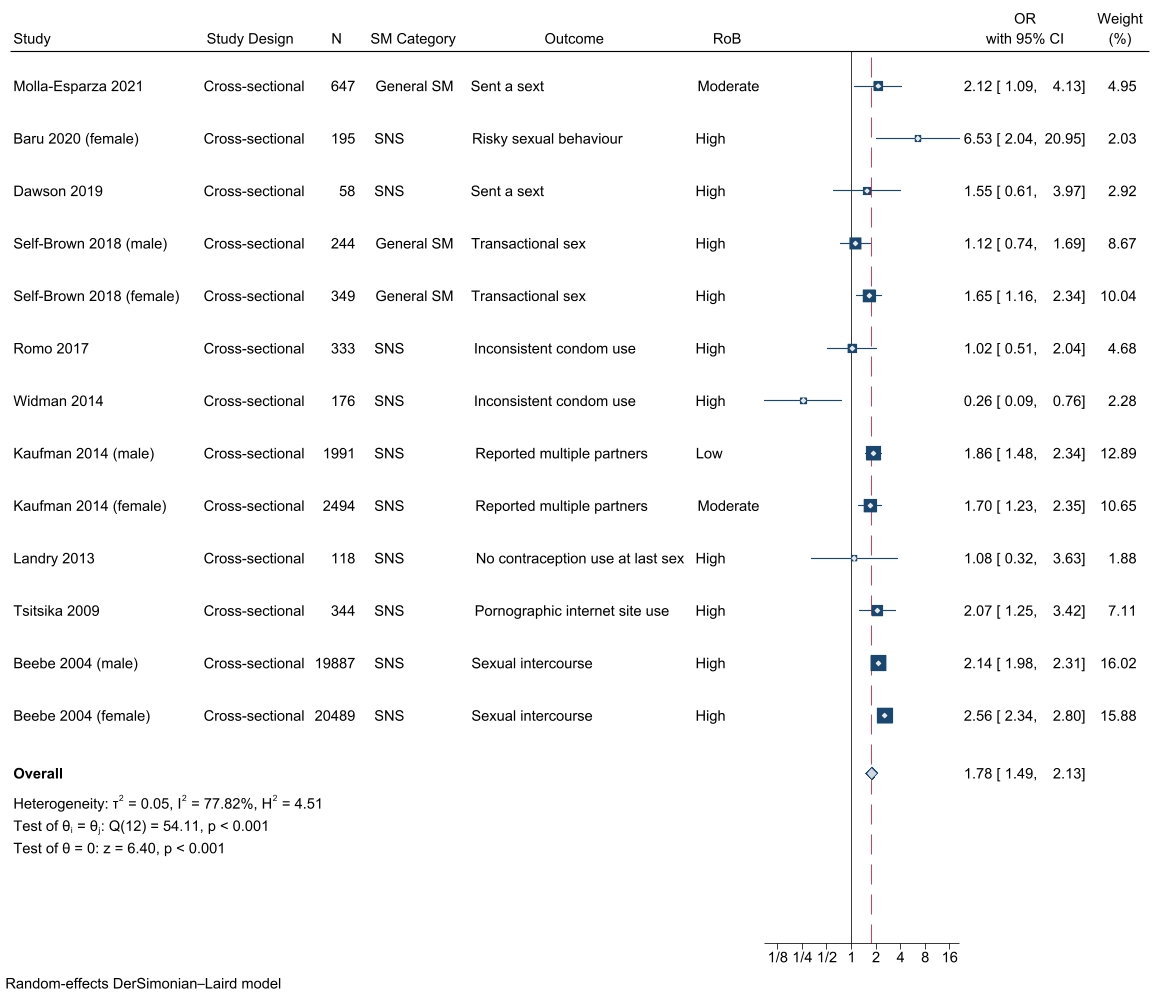
**Legend:** Figure presents forest plot for binary exposure (frequent/at all vs infrequent/not at all) & binary/continuous outcome meta-analysis, with odds ratio (OR) used as common metric. Total number of study participants = 47,325. Abbreviations: CI = Confidence interval; N = Number of study participants; OR = Odds ratio; RoB = Risk of bias; SM = Social media and SNS = Social networking sites.

##### Figure AS. Forest plot for association between frequency of social media use and sexual risk behaviour, average age of study participants

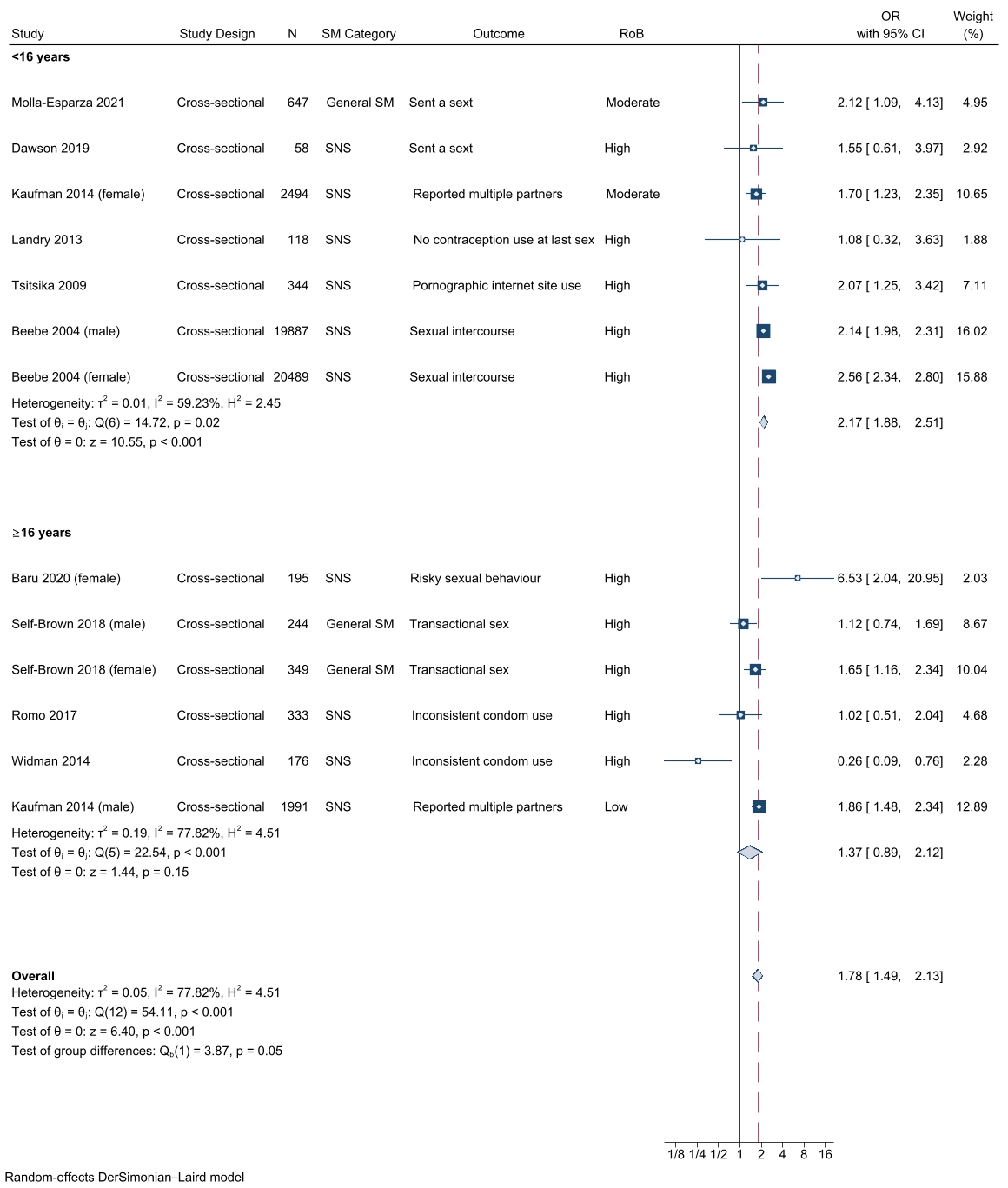

**Legend:** Figure presents forest plot for binary exposure (frequent/at all vs infrequent/not at all) & binary/continuous outcome subgroup analysis, with odds ratio (OR) used as common metric. Total number of study participants = 47,325. Abbreviations: CI = Confidence interval; N = Number of study participants; OR = Odds ratio; RoB = Risk of bias; SM = Social media and SNS = Social networking sites.

##### Figure AT. Forest plot for association between frequency of social media use and sexual risk behaviour, by social media category

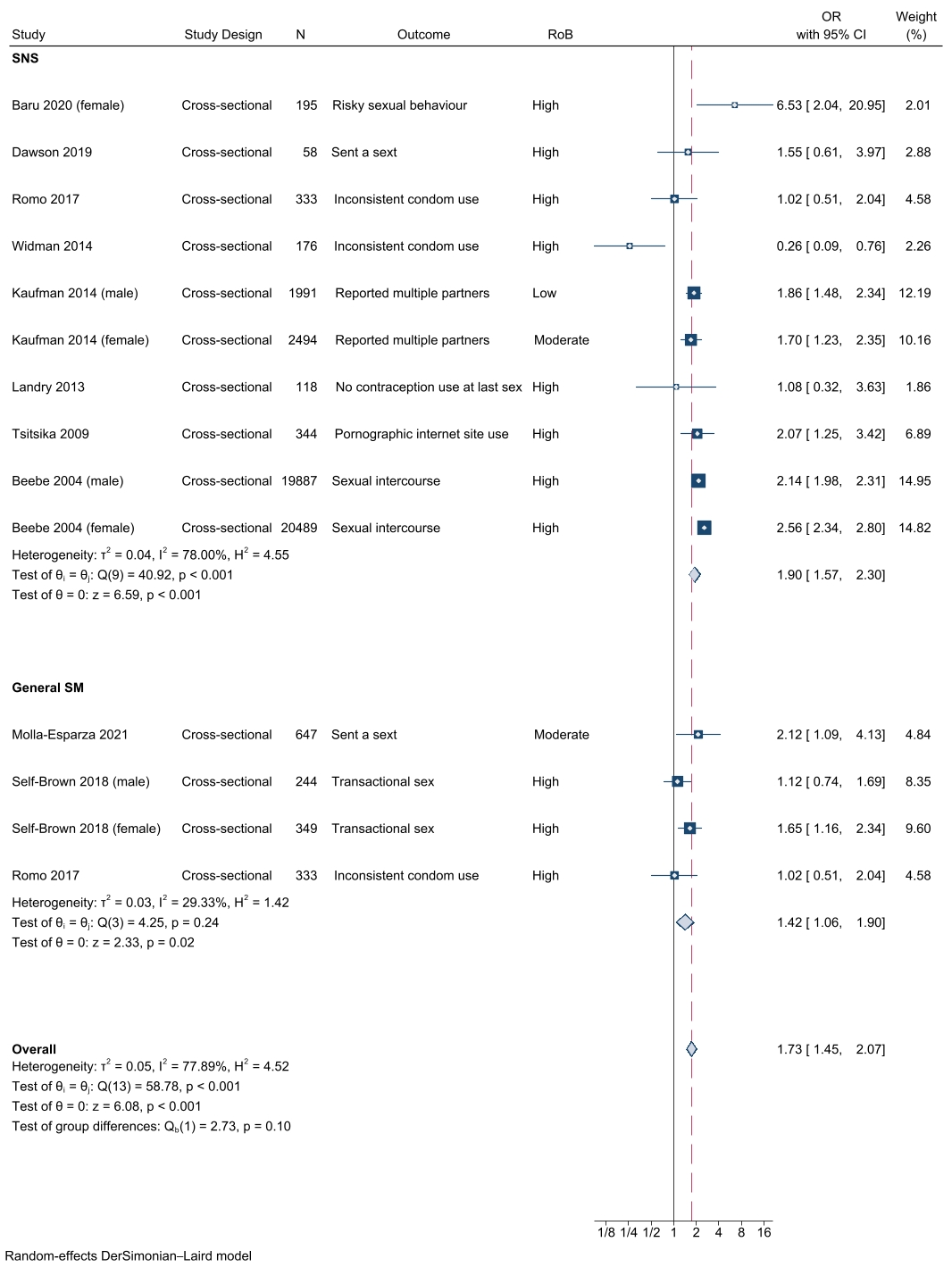

**Legend:** Figure presents forest plot for binary exposure (frequent/at all vs infrequent/not at all) & binary/continuous outcome subgroup analysis, with odds ratio (OR) used as common metric. Total number of study participants = 47,658. Abbreviations: CI = Confidence interval; N = Number of study participants; OR = Odds ratio; RoB = Risk of bias; SM = Social media and SNS = Social networking sites.

##### Figure AU. Forest plot for association between frequency of social media use and sexual risk behaviour, by development status of study setting^a^

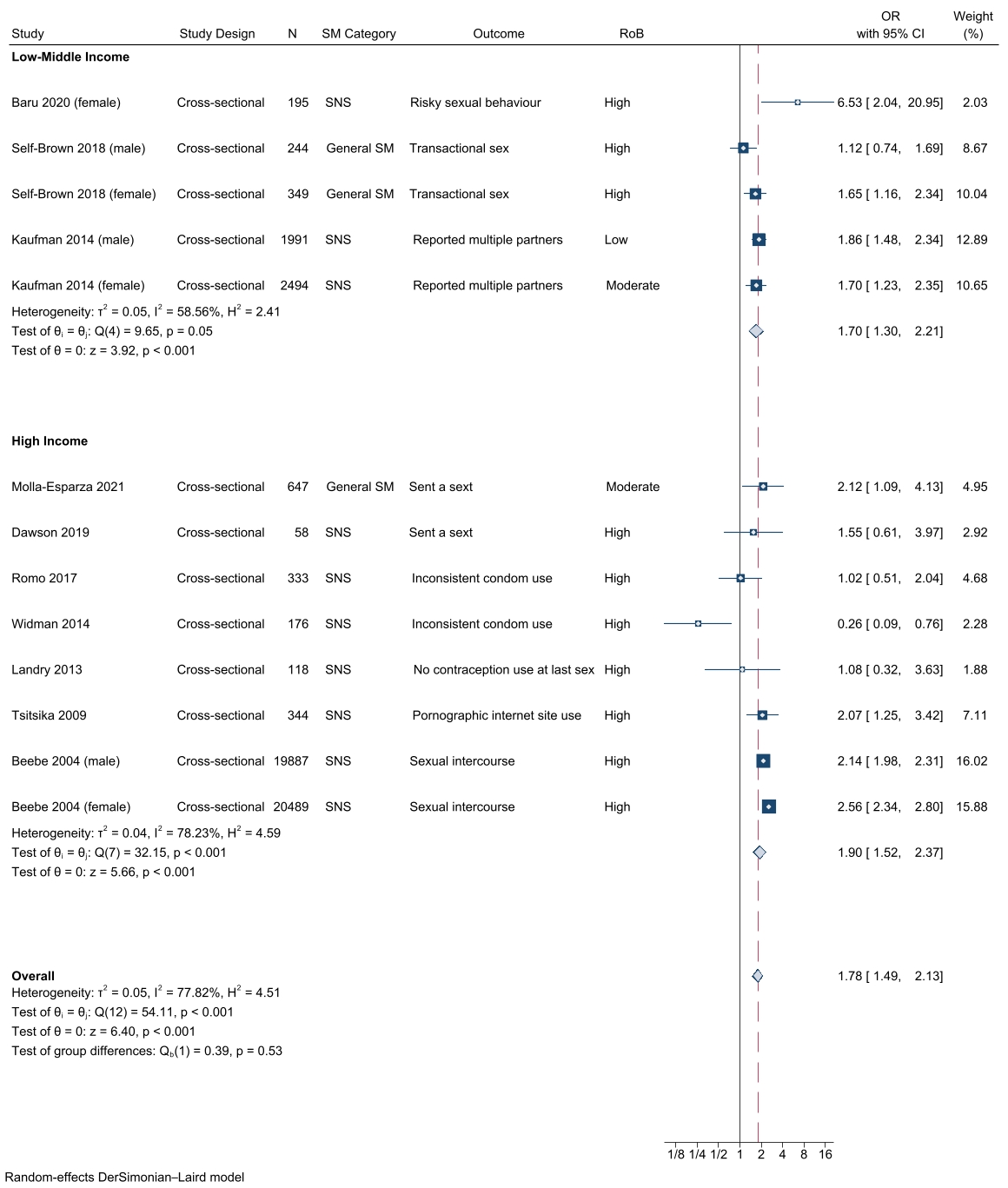

**Legend:** Figure presents forest plot for binary exposure (frequent/at all vs infrequent/not at all) & binary/continuous outcome subgroup analysis, with odds ratio (OR) used as common metric. ^a^ Development status classified as per the World Bank Country Income Level Classification.^160^ Total number of study participants = 47,325. Abbreviations: CI = Confidence interval; N = Number of study participants; OR = Odds ratio; RoB = Risk of bias; SM = Social media and SNS = Social networking sites.

##### Figure AV. Forest plot for association between frequency of social media use and sexual risk behaviour, by sex

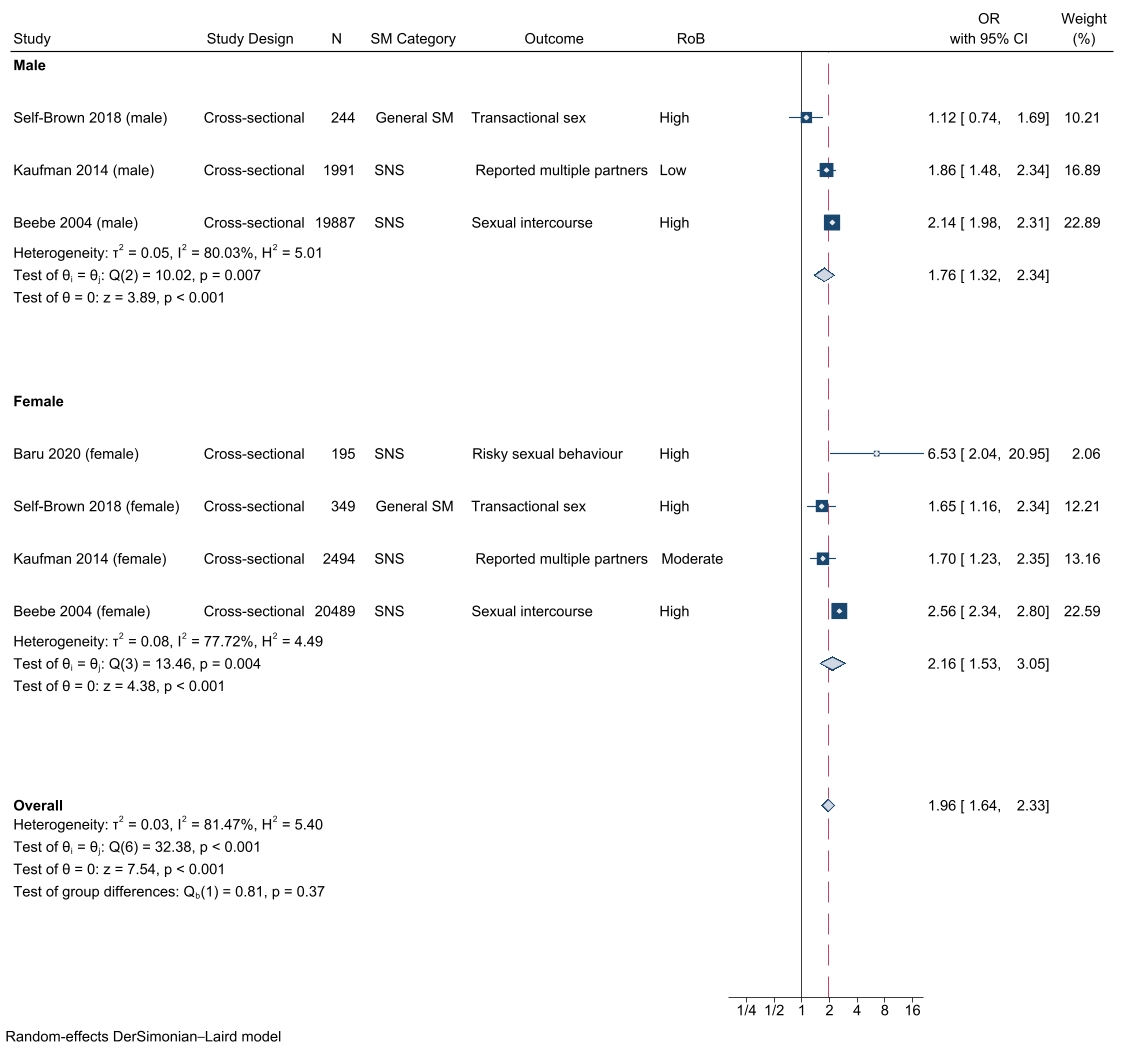

**Legend:** Figure presents forest plot for binary exposure (frequent/at all vs infrequent/not at all) & binary/continuous outcome subgroup analysis, with odds ratio (OR) used as common metric. Total number of study participants = 45,649. Abbreviations: CI = Confidence interval; N = Number of study participants; OR = Odds ratio; RoB = Risk of bias; SM = Social media and SNS = Social networking sites.

##### Figure AW. Forest plot for association between frequency of social media use sexual risk behaviour, by average socioeconomic position of study participants

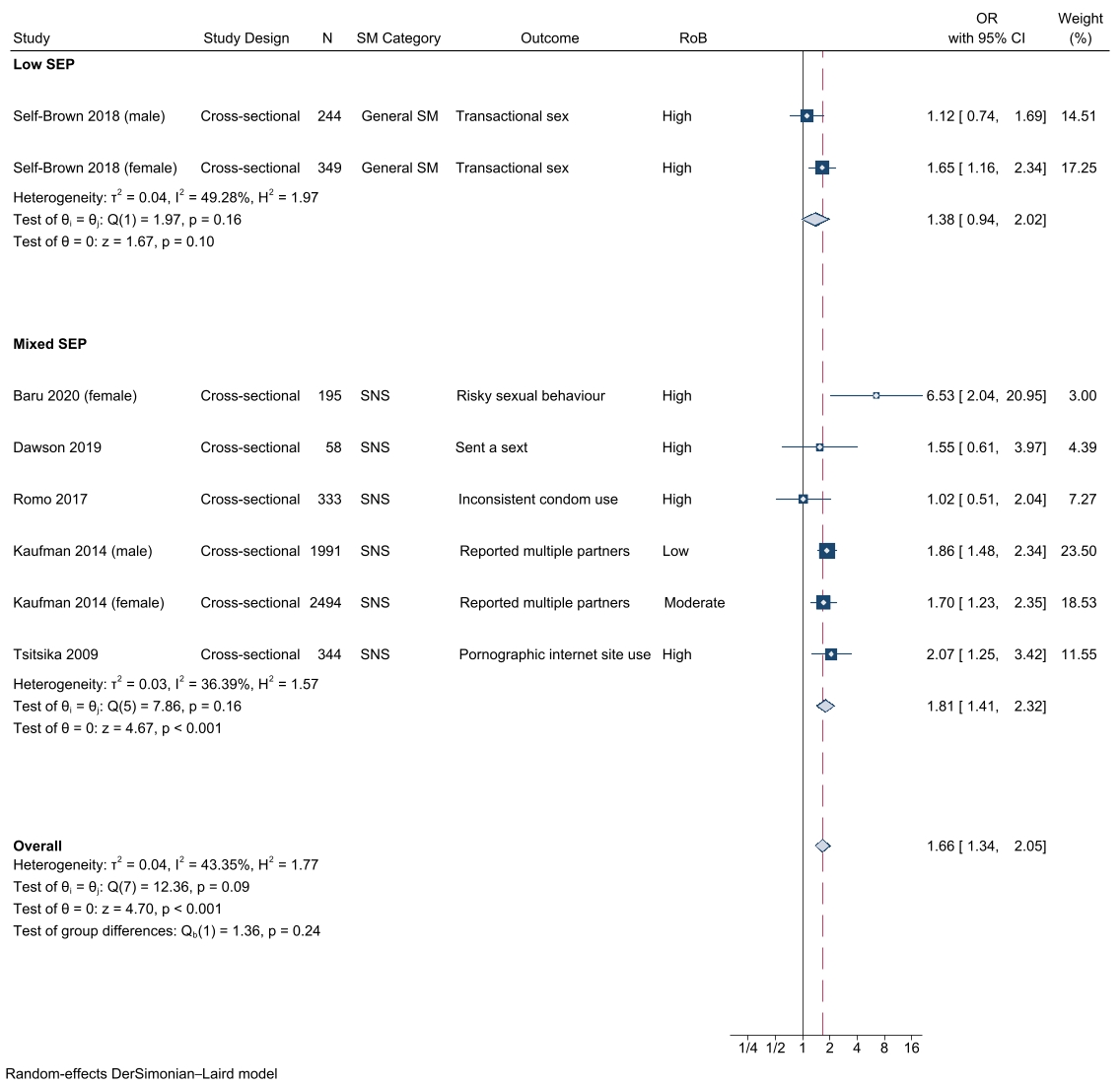

**Legend:** Figure presents forest plot for binary exposure (frequent/at all vs infrequent/not at all) & binary/continuous outcome subgroup analysis, with odds ratio (OR) used as common metric. Total number of study participants = 6008. Abbreviations: CI = Confidence interval; N = Number of study participants; OR = Odds ratio; RoB = Risk of bias; SM = Social media and SNS = Social networking sites.

##### Figure AX. Forest plot for association between frequency of social media use and sexual risk behaviour

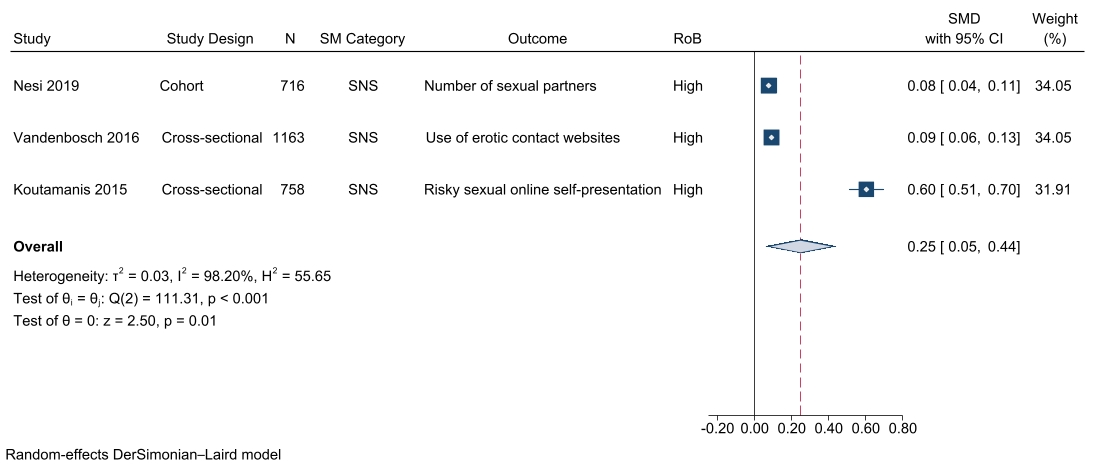

**Legend:** Figure presents forest plot for continuous exposure & continuous outcome meta-analysis, with standardised mean difference (SMD) used as common metric. Total number of study participants = 2637. Abbreviations: CI = Confidence interval; N = Number of study participants; RoB = Risk of bias; SM = Social media; SMD = Standardised mean difference and SNS = Social networking sites.

##### Figure AY. Forest plot for association between frequency of social media use and sexual risk behaviour, by average age of study participants

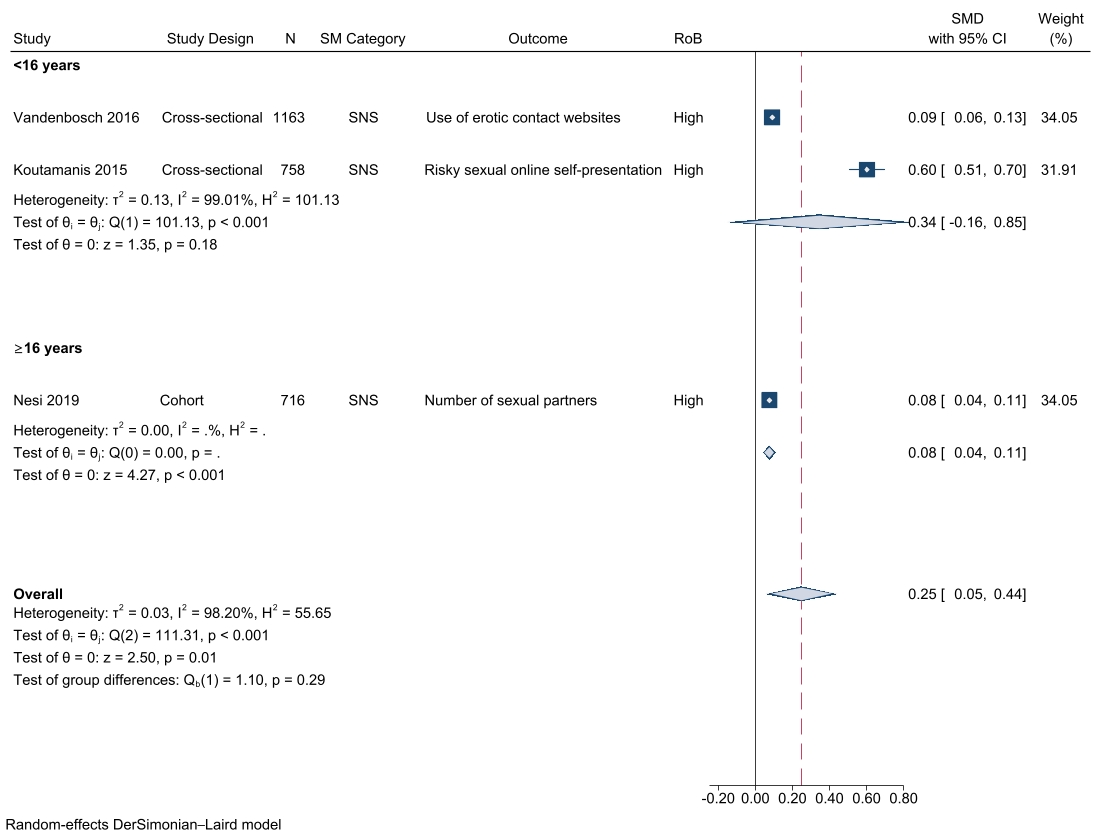

**Legend:** Figure presents forest plot for continuous exposure & continuous outcome subgroup analysis, with standardised mean difference (SMD) used as common metric. Total number of study participants = 2637. Abbreviations: CI = Confidence interval; N = Number of study participants; RoB = Risk of bias; SM = Social media; SMD = Standardised mean difference and SNS = Social networking sites.

##### Figure AZ. Forest plot for association between frequency of social media use and sexual risk behaviour

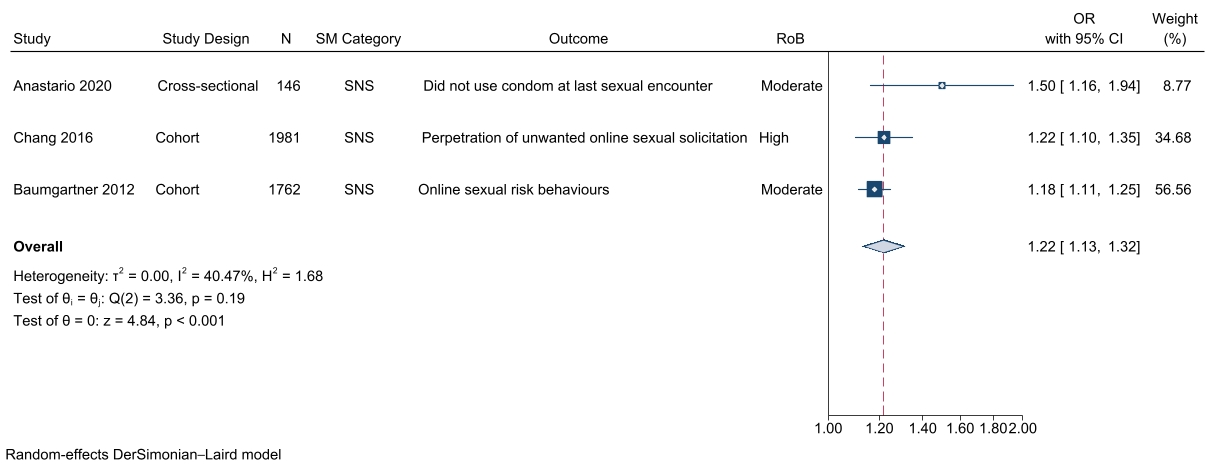

**Legend:** Figure presents forest plot for continuous exposure & binary outcome meta-analysis, with odds ratio (OR) used as common metric. Total number of study participants = 3889. Abbreviations: CI = Confidence interval; N = Number of study participants; OR = Odds ratio; RoB = Risk of bias; SM = Social media and SNS = Social networking sites.

##### Figure BA. Forest plot for association between frequency of social media use and sexual risk behaviour, by social media category

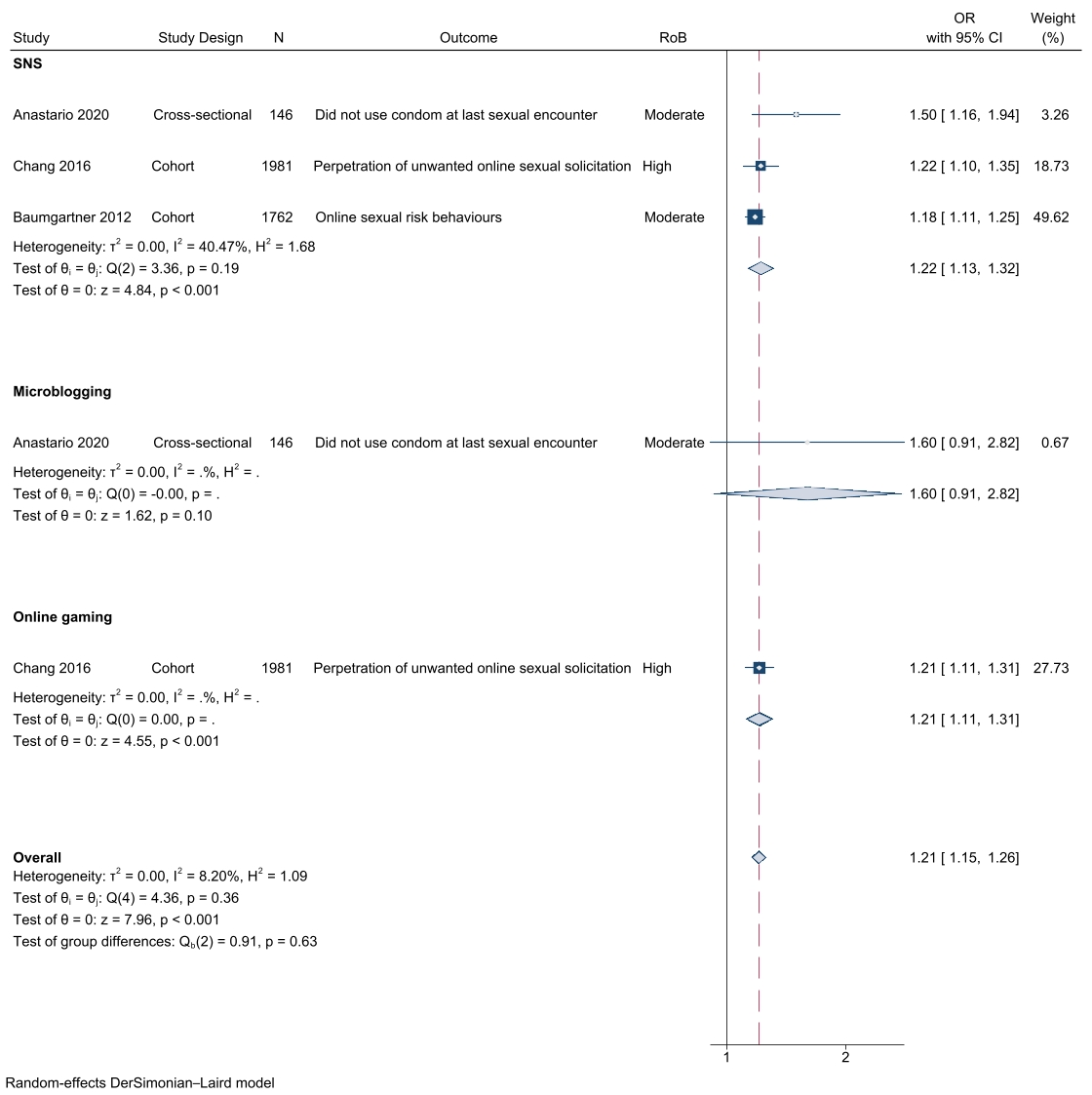

**Legend:** Figure presents forest plot for continuous exposure & binary outcome subgroup analysis, with odds ratio (OR) used as common metric. Total number of study participants = 6016. Abbreviations: CI = Confidence interval; N = Number of study participants; OR = Odds ratio; SM = Social media and SNS = Social networking sites.

##### Figure BB. Forest plot for association between time spent on social media and sexual risk behaviour

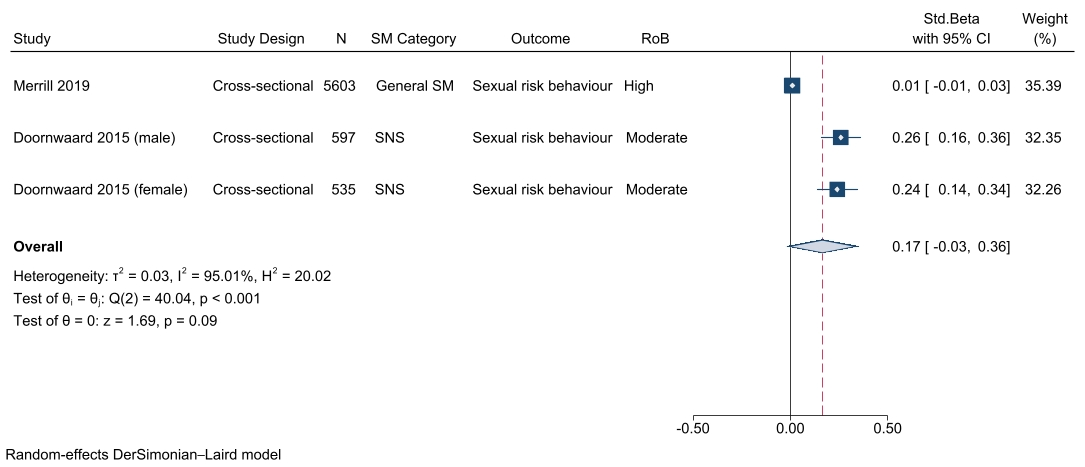

**Legend:** Figure presents forest plot for continuous exposure & continuous outcome meta-analysis, with standardised beta (Std.Beta) used as common metric. Total number of study participants = 6735. Abbreviations: CI = Confidence interval; N = Number of study participants; RoB = Risk of bias; SM = Social media; SNS = Social networking sites and Std.Beta = Standardised beta.

##### Figure BC. Forest plot for association between time spent on social media and sexual risk behaviour

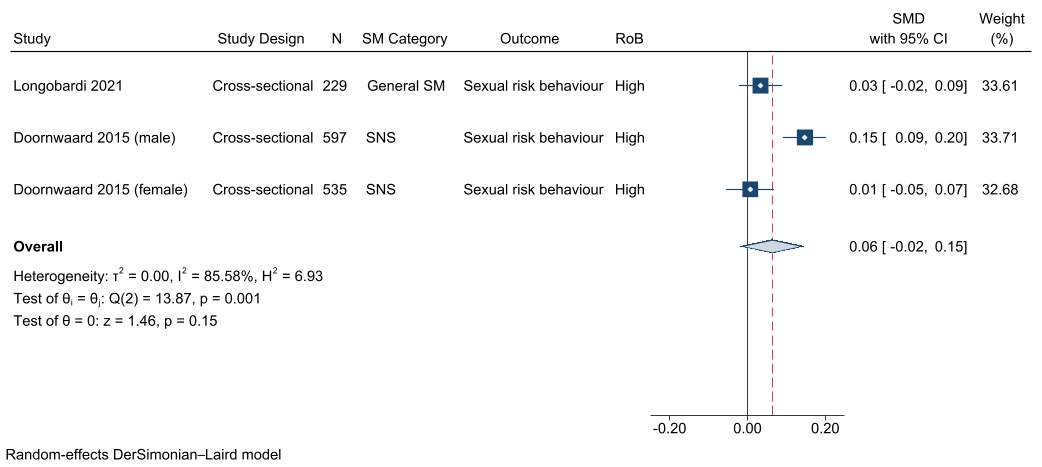
**Legend:** Figure presents forest plot for continuous exposure & continuous outcome meta-analysis, with standardised mean difference (SMD) used as common metric. Total number of study participants = 1361. Abbreviations: CI = Confidence interval; N = Number of study participants; RoB = Risk of bias; SM = Social media; SMD = Standardised mean difference and SNS = Social networking sites.

#### Gambling

##### Effect direction plot

Figure BD shows the effect direction in those studies investigating gambling behaviour, by exposure. After excluding 1 study demonstrating inconsistent effects (participant n=14,478),^45^ for frequency of social media use all studies reported harmful associations (95% CI 56.6 to 100.0%; study n=5; participant n=7928; sign test p=0.06). Other social media activities was investigated by one study which demonstrated a harmful association on gambling behaviours (20.7 to 100.0%; participant n=3772; insufficient data to conduct sign test).

##### Figure BD. Effect direction plot for studies of the association between social media use and adolescent gambling, by social media exposure. Arrow size indicates sample size; arrow colour indicates study risk of bias

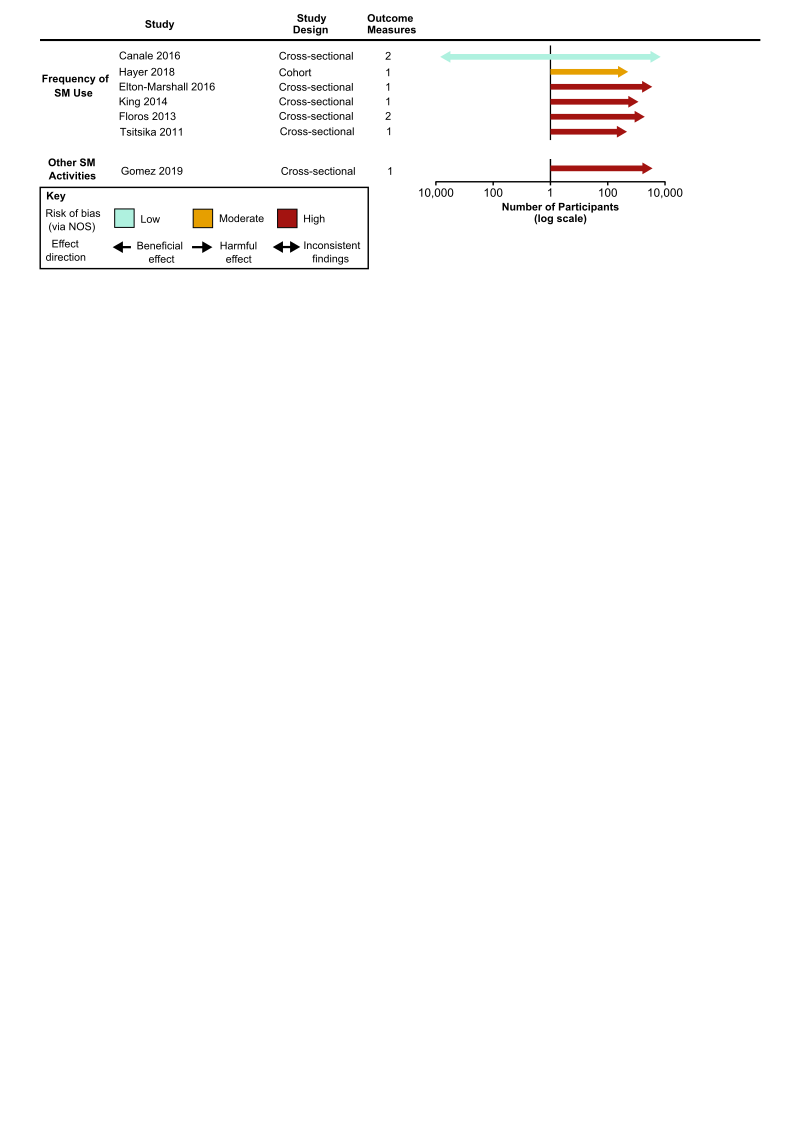

**Legend:** Sample size: represented by the size of the arrow, measured on a log scale. Outcome measure: number of outcome measures

synthesised within each study. Studies organised by risk of bias grade, study design, and year of publication. Repeat cross-sectional studies, multiple study populations from different countries, and age subsets originating from the same study reported as separate studies. Abbreviations: NOS = Assessed via adapted Newcastle Ottawa Scale and SM = Social media.

##### Forest plots for meta-analyses and subgroup analyses

##### Figure BE. Forest plot for association between frequency of social media use and gambling (not via social media)

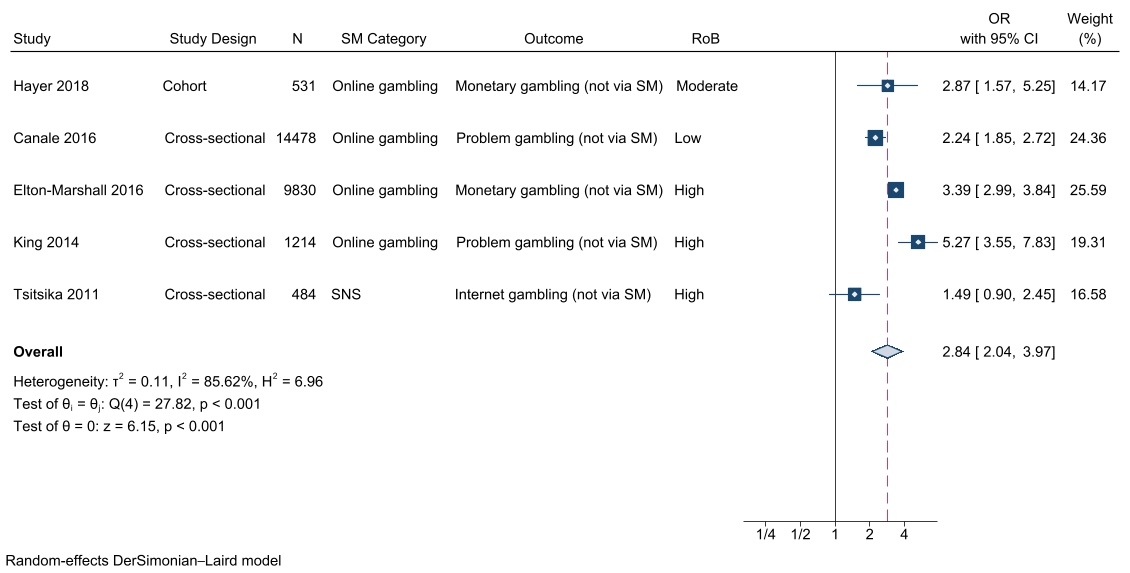

**Legend:** Figure presents forest plot for binary exposure (frequent/at all vs infrequent/not at all) & binary/continuous outcome meta-analysis, with odds ratio (OR) used as common metric. Total number of study participants = 26,537. Abbreviations: CI = Confidence interval; N = Number of study participants; OR = Odds ratio; RoB = Risk of bias; SM = Social media and SNS = Social networking sites

##### Figure BF. Forest plot for association between frequency of social media use and gambling (not via social media), by average age of study participants

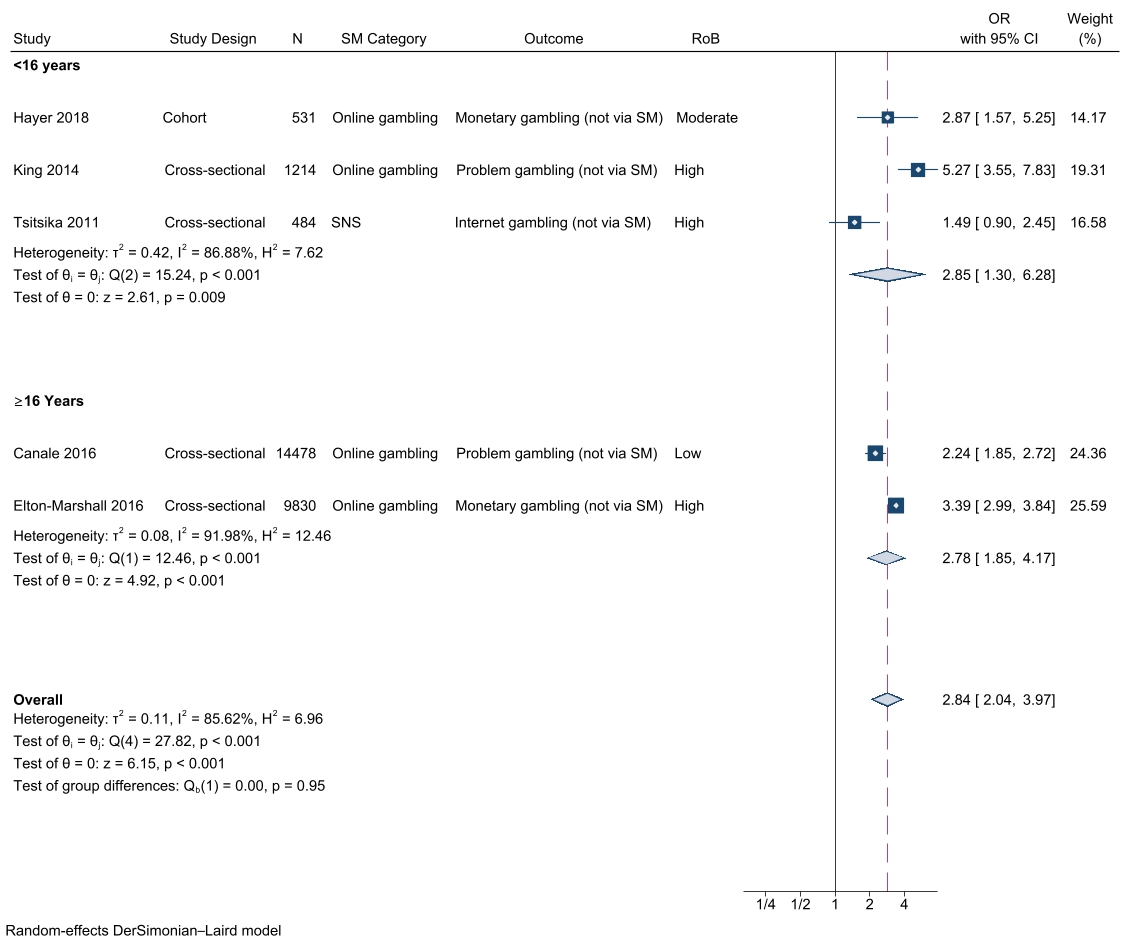

**Legend:** Figure presents forest plot for binary exposure (frequent/at all vs infrequent/not at all) & binary/continuous outcome subgroup analysis, with odds ratio (OR) used as common metric. Total number of study participants = 26,537. Abbreviations: CI = Confidence interval; N = Number of study participants; OR = Odds ratio; RoB = Risk of bias; SM = Social media and SNS = Social networking sites.

##### Figure BG. Forest plot for association between frequency of social media use and gambling (not via social media), by social media category

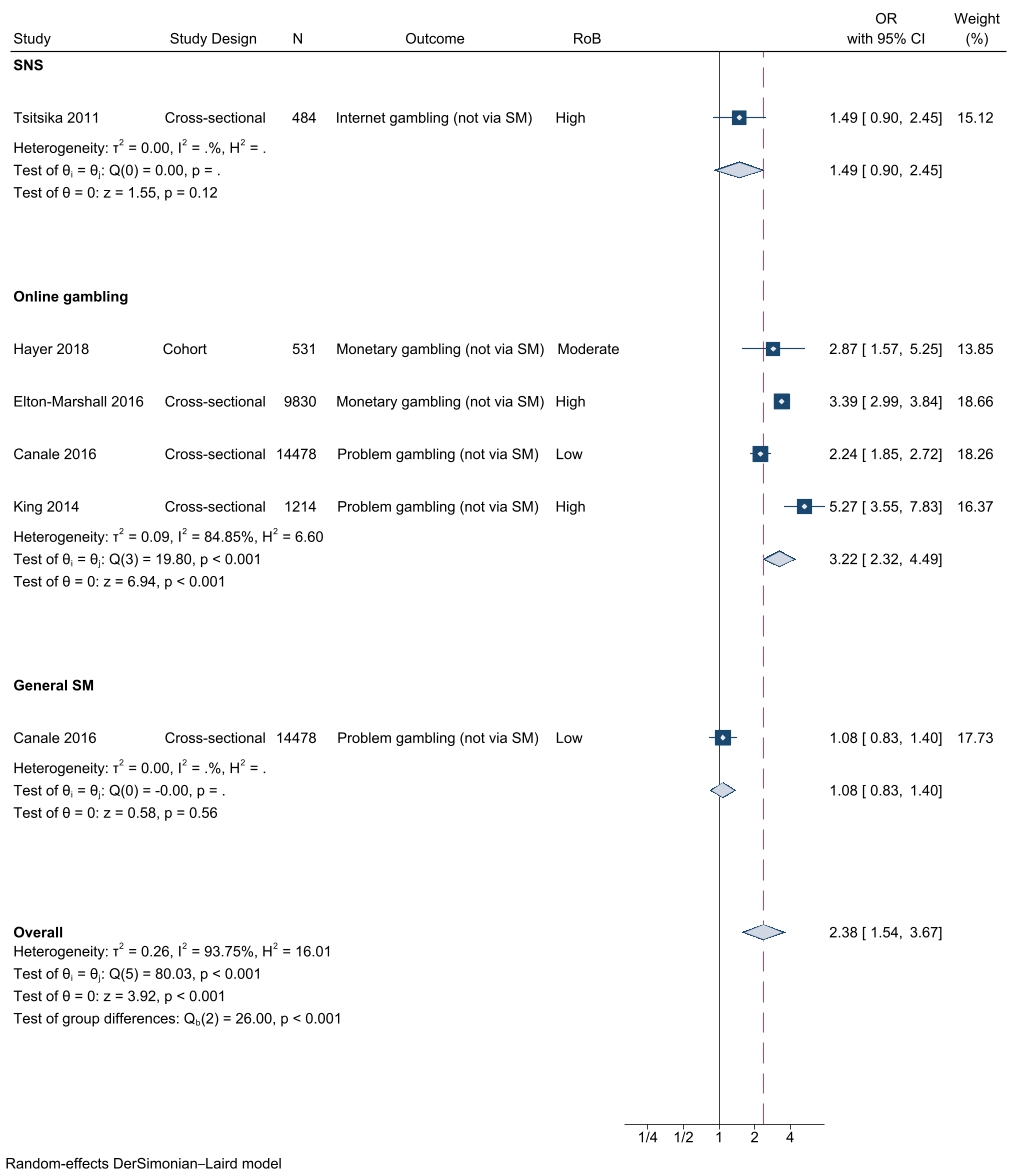

**Legend:** Figure presents forest plot for binary exposure (frequent/at all vs infrequent/not at all) & binary/continuous outcome subgroup analysis, with odds ratio (OR) used as common metric. Total number of study participants = 41,015. Abbreviations: CI = Confidence interval; N = Number of study participants; OR = Odds ratio; RoB = Risk of bias; SM = Social media and SNS = Social networking sites.

#### Anti-social behaviour

##### Effect direction plot

Figure BH demonstrates the effect direction in those studies investigating anti-social risk behaviour, by exposure. One study investigated more than one exposure type.^51^Across all investigated exposures, all studies demonstrated harmful associations of social media use (time spent on social media: 95% CI 61.0 to 100.0%, study n=6, participant n=51,611, sign test p=0.03; frequency of social media use: 64.6 to 100.0%, study n=7, participant n=56,918, sign test p=0.02; and exposure to health-risk behaviour content on social media: 34.2 to 100.0%, study n=2, participant n=1372, insufficient data to conduct sign test). Other social media activities was investigated by one study, which demonstrated a harmful effect (20.7 to 100.0%; participant n=1167; insufficient data to conduct sign test.

##### Figure BH. Effect direction plot for studies of the association between social media use and adolescent engagement in anti-social behaviour, by social media exposure. Arrow size indicates sample size; arrow colour indicates study risk of bias

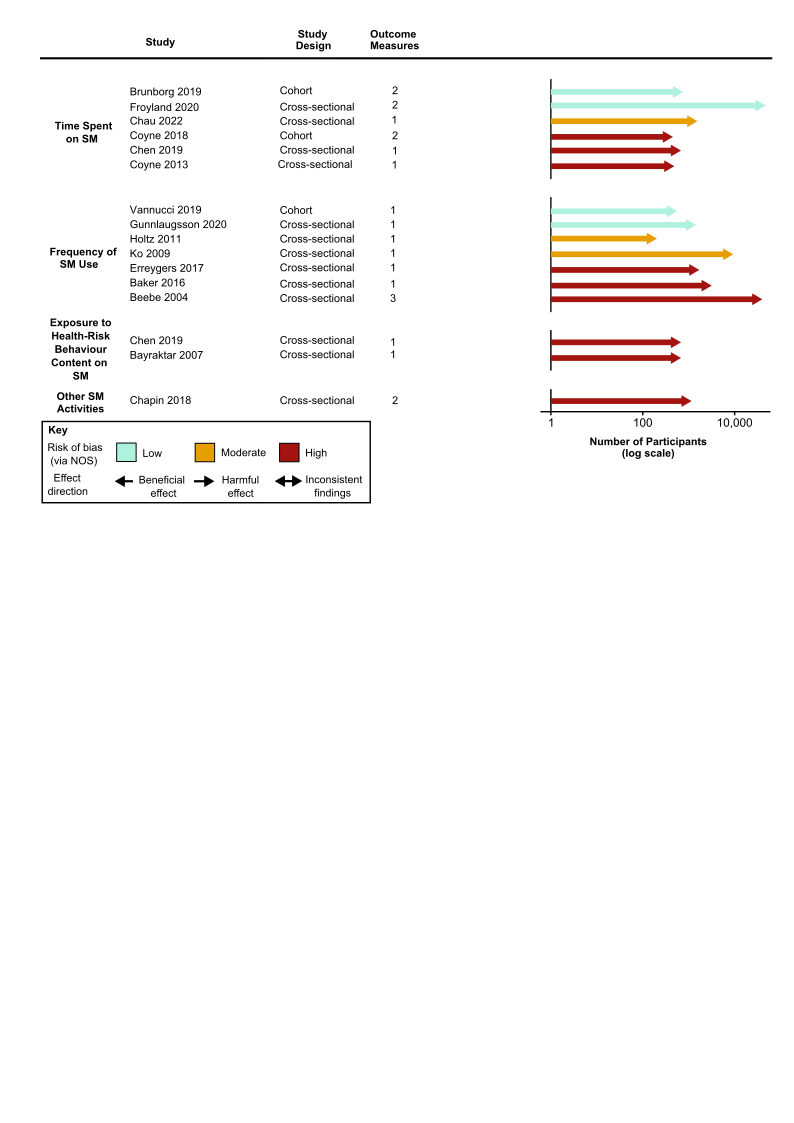

**Legend:** Sample size: represented by the size of the arrow, measured on a log scale. Outcome measure: number of outcome measures synthesised within each study. Studies organised by risk of bias grade, study design, and year of publication. Repeat cross-sectional studies, multiple study populations from different countries, and age subsets originating from the same study reported as separate studies. Abbreviations: NOS = Assessed via adapted Newcastle Ottawa Scale and SM = Social media.

##### Forest plots for meta-analyses and subgroup analyses

##### Figure BI. Forest plot for association between frequency of social media use and anti-social behaviour

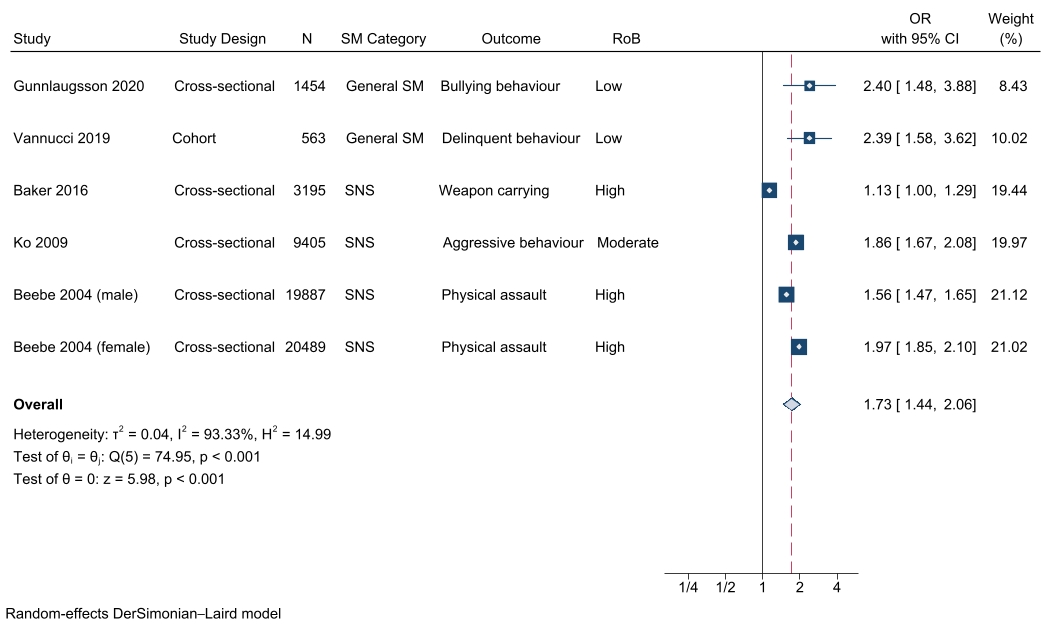

**Legend:** Figure presents forest plot for binary exposure (frequent/at all vs infrequent/not at all) & binary/continuous outcome meta-analysis, with odds (OR) used as common metric. Total number of study participants = 54,993. Abbreviations: CI = Confidence interval; N = Number of study participants; OR = Odds ratio; RoB = Risk of bias; SM = Social media and SNS = Social networking sites.

##### Figure BJ. Forest plot for association between time spent on social media and anti-social behaviour

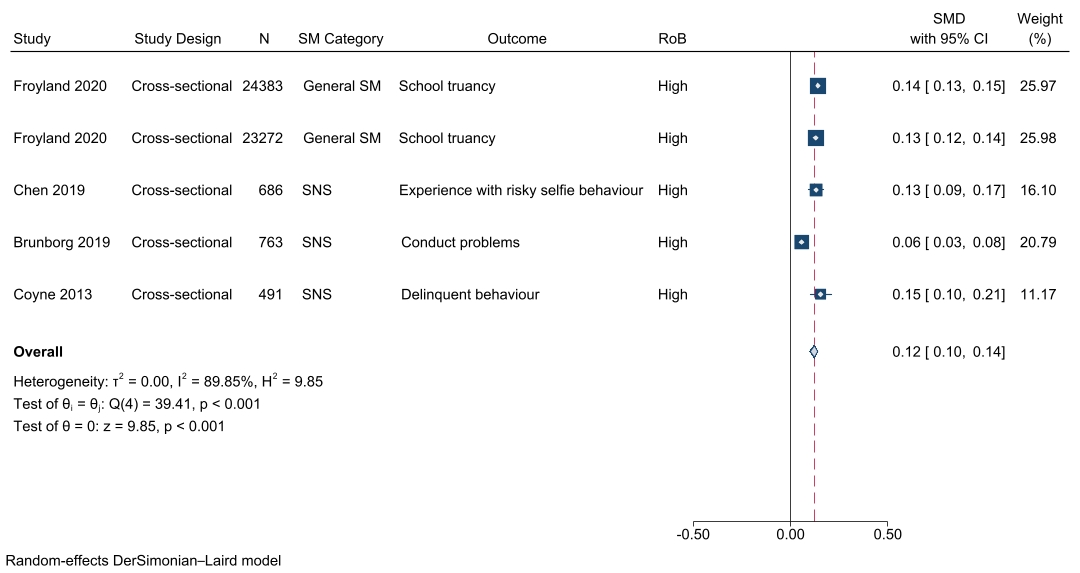

**Legend:** Figure presents forest plot for continuous exposure & continuous outcome meta-analysis, with standardised mean difference (SMD) used as common metric. Total number of study participants = 49,595. Abbreviations: CI = Confidence interval; N = Number of study participants; RoB = Risk of bias; SM = Social media; SMD = Standardised mean difference and SNS = Social networking sites.

##### Figure BK. Forest plot for association between frequency of social media use and anti-social behaviour, by development status of study setting^a^

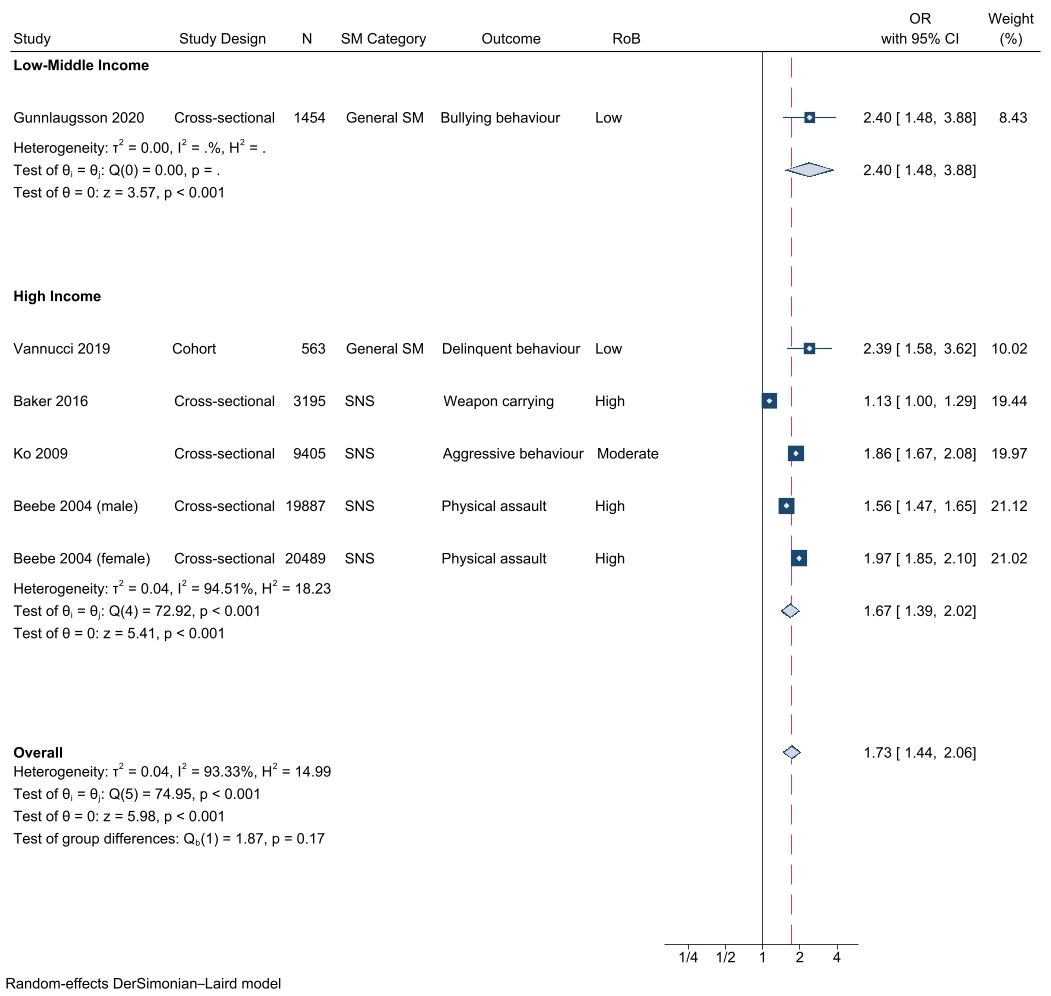

**Legend:** Figure presents forest plot for binary exposure (frequent/at all vs infrequent/not at all) & binary/continuous outcome subgroup analysis, with odds (OR) used as common metric. ^a^ Development status classified as per the World Bank Country Income Level Classification.^160^ Total number of study participants = 54,993. Abbreviations: CI = Confidence interval; N = Number of study participants; OR = Odds ratio; RoB = Risk of bias; SM = Social media and SNS = Social networking sites

##### Figure BL. Forest plot for association between frequency of social media use and anti-social behaviour, by social media category

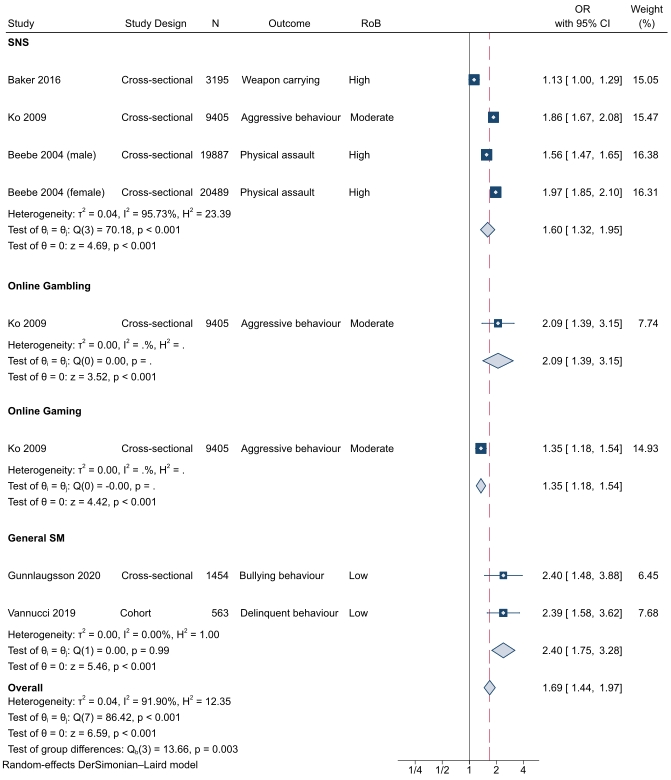

**Legend:** Figure presents forest plot for binary exposure (frequent/at all vs infrequent/not at all) & binary/continuous outcome subgroup analysis, with odds (OR) used as common metric. Total number of study participants = 73,803. Abbreviations: CI = Confidence interval; N = Number of study participants; OR = Odds ratio; RoB = Risk of bias; SM = Social media and SNS = Social networking site

##### Figure BM. Forest plot for association between time spent on social media and anti-social behaviour, by average age of study participants

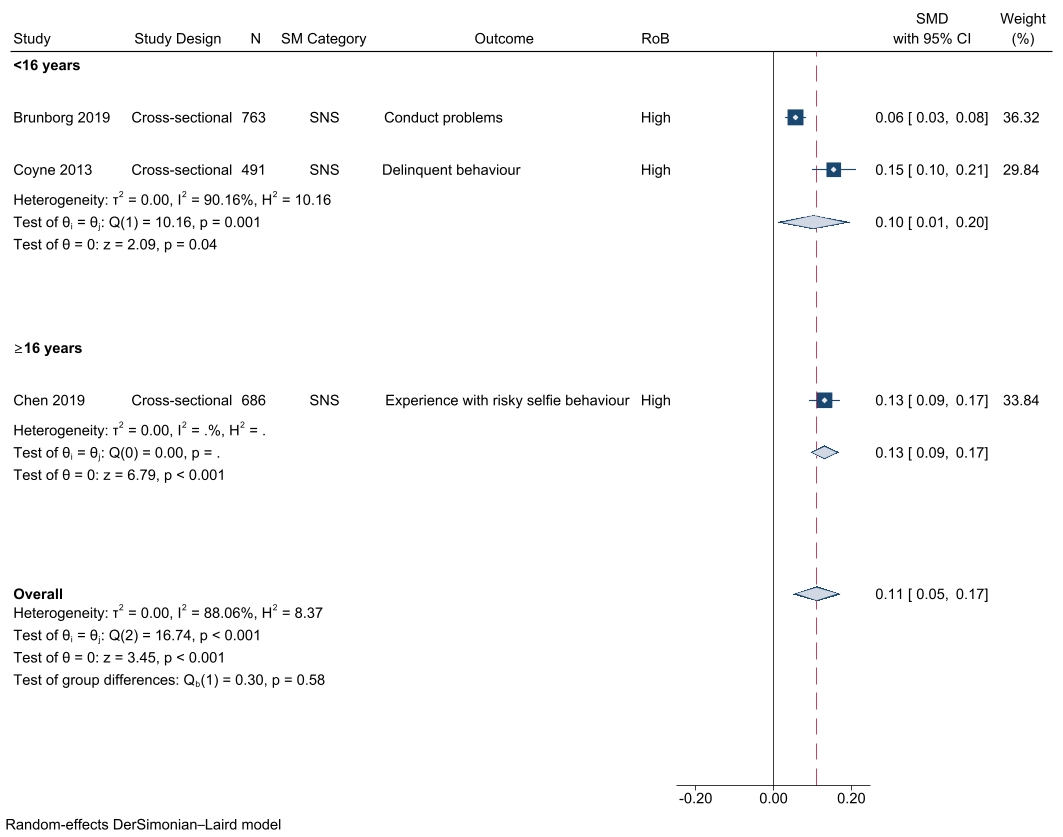

**Legend:** Figure presents forest plot for continuous exposure & continuous outcome subgroup analysis, with standardised mean difference (SMD) used as common metric. Total number of study participants = 1940. Abbreviations: CI = Confidence interval; N = Number of study participants; RoB = Risk of bias; SM = Social media; SMD = Standardised mean difference and SNS = Social networking sites.

##### Figure BN. Forest plot for association between time spent on social media and anti-social behaviour, by social media category

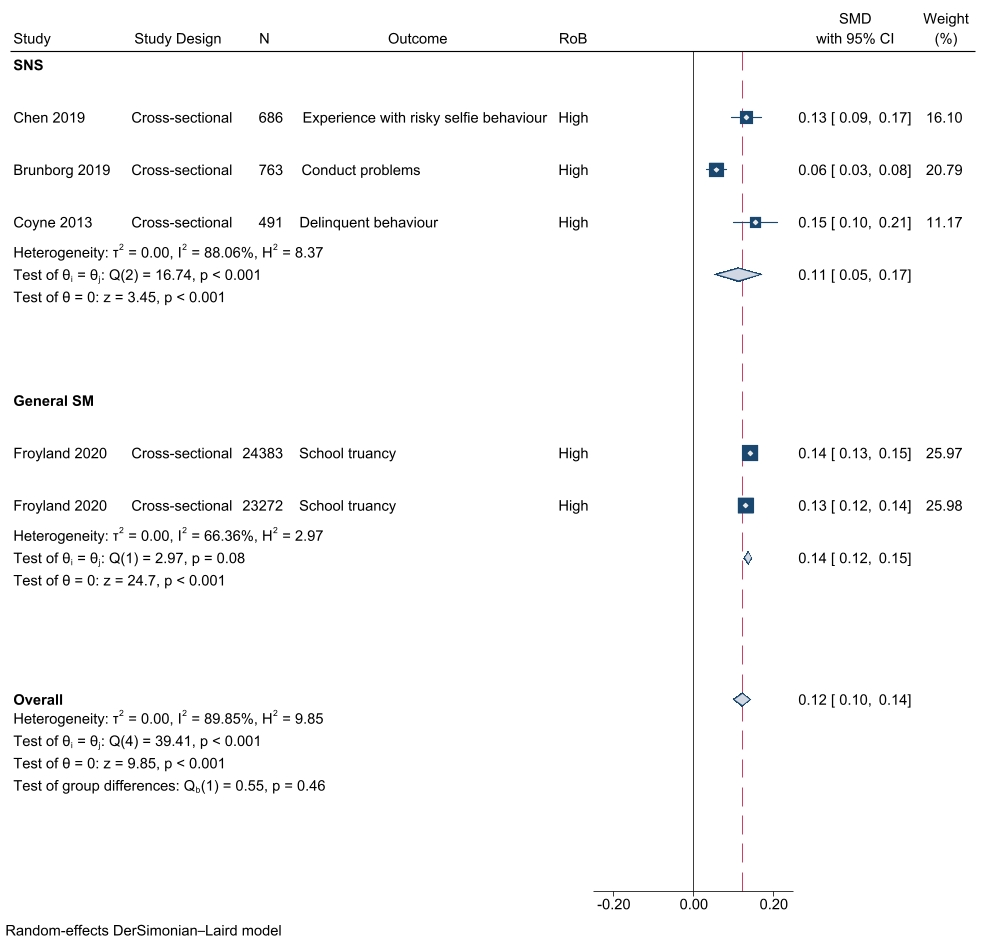

**Legend:** Figure presents forest plot for continuous exposure & continuous outcome meta-analysis, with standardised mean difference (SMD) used as common metric. Total number of study participants = 49,595. Abbreviations: CI = Confidence interval; N = Number of study participants; RoB = Risk of bias; SM = Social media; SMD = Standardised mean difference and SNS = Social networking sites.

#### Inadequate physical activity

##### Effect direction plot

Figure BO demonstrates the effect direction in those studies (n=14) investigating inadequate physical activity, by exposure. After excluding those with inconsistent findings (n=3),^111,136,94^ for time spent on social media, 4/8 of studies reported harmful associations (95% CI 21.5 to 78.5%; participant n = 52,475; sign test p=1.00), whilst for frequency of social media use no study reported a harmful association (0.00 to 56.1%; study n=3; participant n=57,953; sign test p=0.25).

##### Figure BO. Effect direction plot for studies of the association between social media use and adolescent inadequate physical activity, by social media exposure. Arrow size indicates sample size; arrow colour indicates study risk of bias

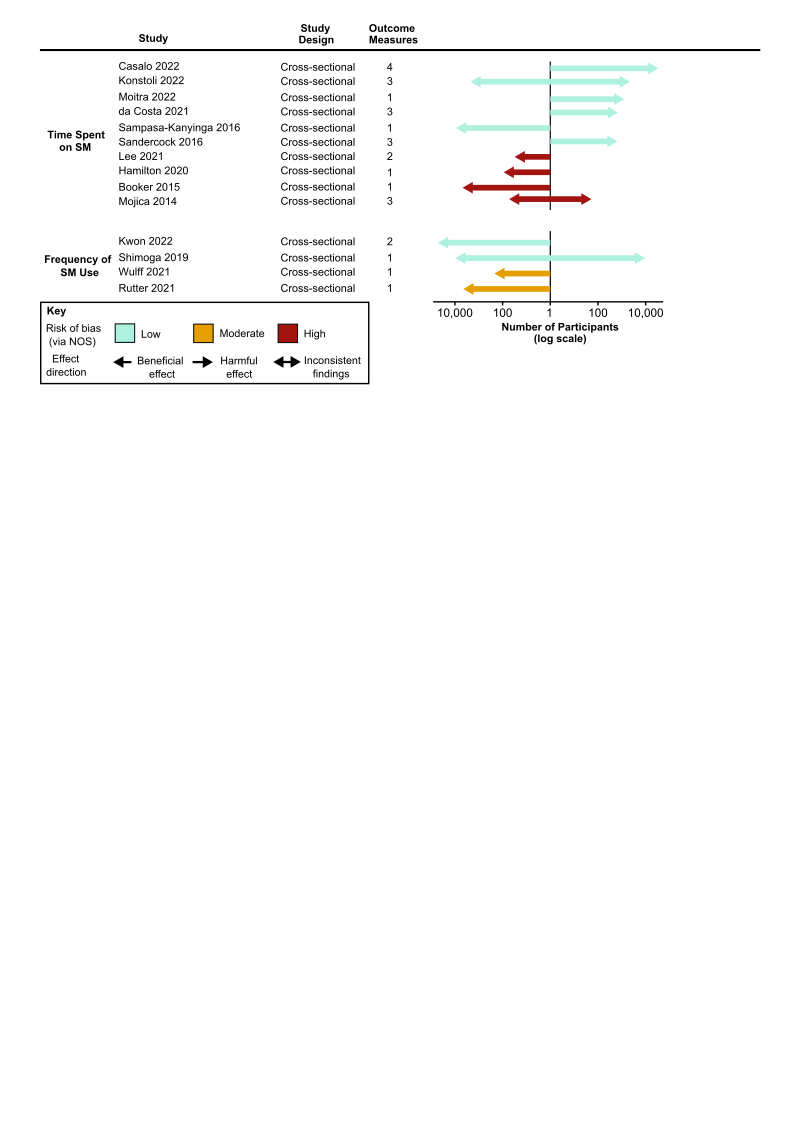

**Legend:** Sample size: represented by the size of the arrow, measured on a log scale. Outcome measure: number of outcome measures synthesised within each study. Studies organised by risk of bias grade, study design, and year of publication. Repeat cross-sectional studies, multiple study populations from different countries, and age subsets originating from the same study reported as separate studies. Shimoga 2019 assessed frequency of social media use and one outcome (physical activity) across three subgroups. Two of the three subgroups showed increased frequency of social media resulted in decreased physical activity, and one subgroup showed increased frequency of social media resulted in increased physical activity, thus this study was classified as demonstrating inconsistent findings. Abbreviations: NOS = Assessed via adapted Newcastle Ottawa Scale and SM = Social media.

##### Forest plots for meta-analyses and subgroup analyses

##### Figure BP. Forest plot for association between time spent on social media and inadequate physical activity

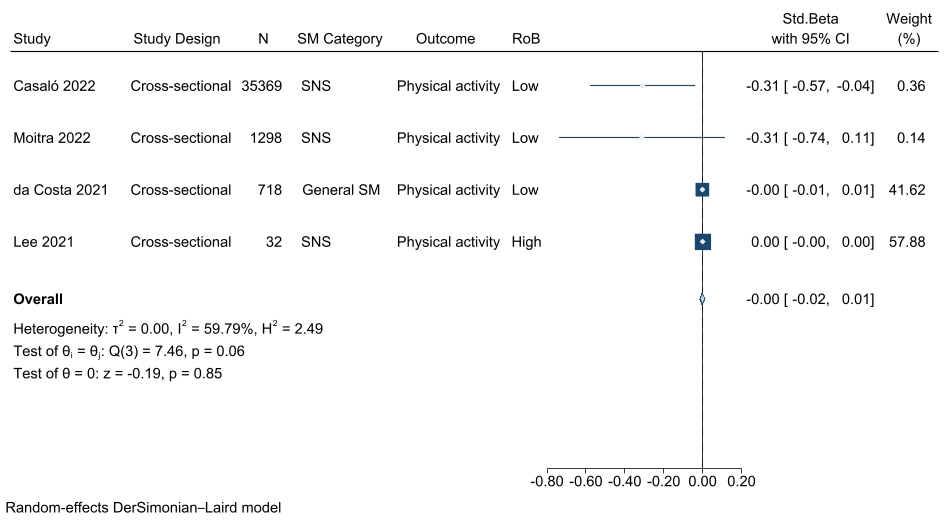

**Legend:** Figure presents forest plot for continuous exposure & continuous outcome meta-analysis, with standardised beta (Std.Beta) used as common metric. Total number of study participants =37,417. Abbreviations: CI = Confidence interval; N = Number of study participants; RoB = Risk of bias; SM = Social media; SNS = Social networking sites and Std.Beta = Standardised beta.

##### Figure BQ. Forest plot for association between time spent on social media and inadequate physical activity, by average age of study participants

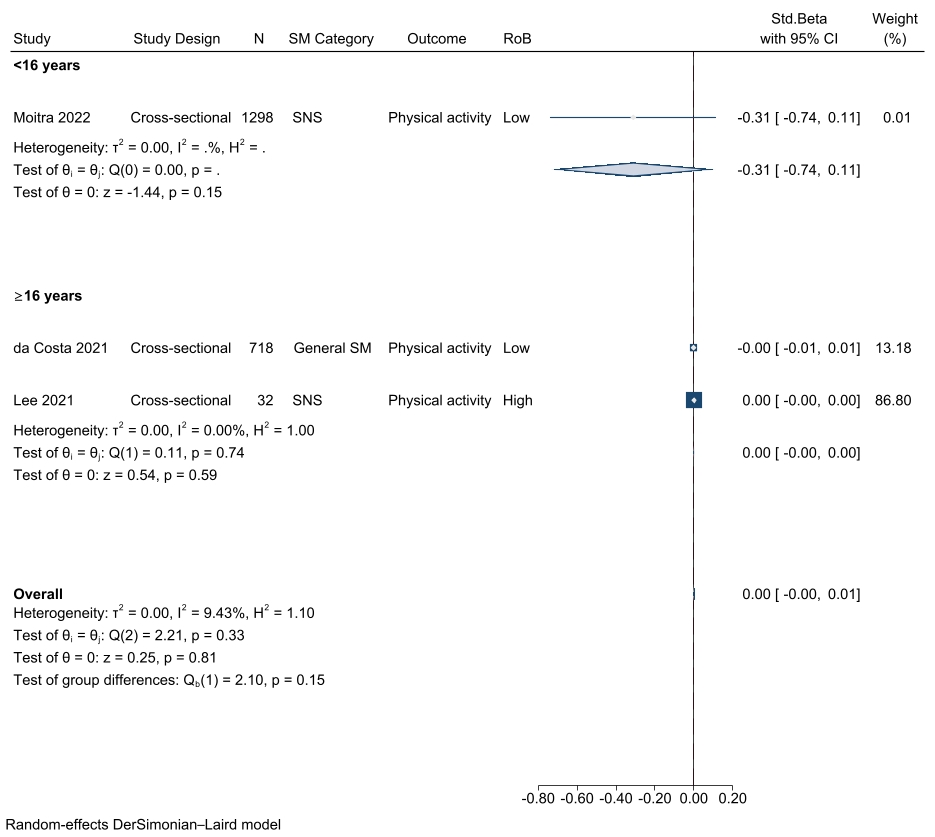

**Legend:** Figure presents forest plot for continuous exposure & continuous outcome subgroup analysis, with standardised beta (Std.Beta) used as common metric. Total number of study participants = 2048. Abbreviations: CI = Confidence interval; N = Number of study participants; RoB = Risk of bias; SM = Social media; SNS = Social networking sites and Std.Beta = Standardised beta.

##### Figure BR. Forest plot for association between time spent on social media and inadequate physical activity, by development status of study setting^a^

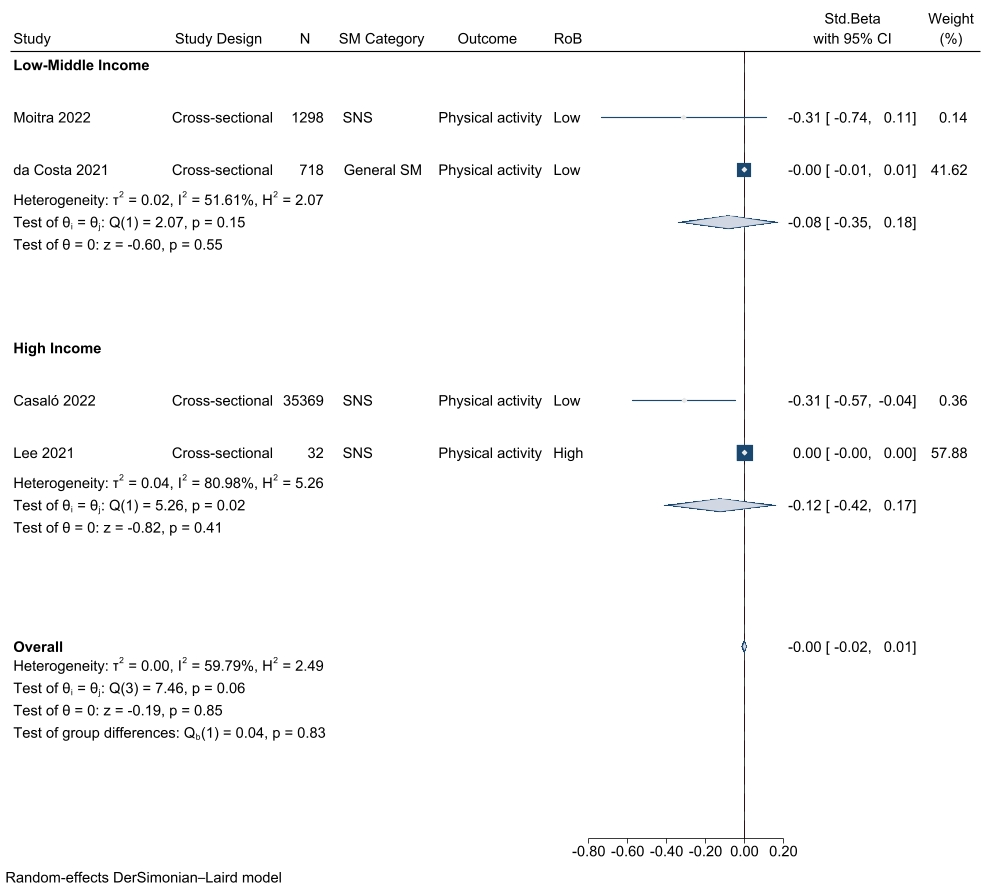

**Legend:** Figure presents forest plot for continuous exposure & continuous outcome subgroup analysis, with standardised beta (Std.Beta) used as common metric. ^a^ Development status classified as per the World Bank Country Income Level Classification.^160^ Total number of study participants = 37,417. Abbreviations: CI = Confidence interval; N = Number of study participants; RoB = Risk of bias; SM = Social media; SNS = Social networking sites and Std.Beta = Standardised beta.

##### Figure BS. Forest plot for association between time spent on social media and inadequate physical activity, by social media category

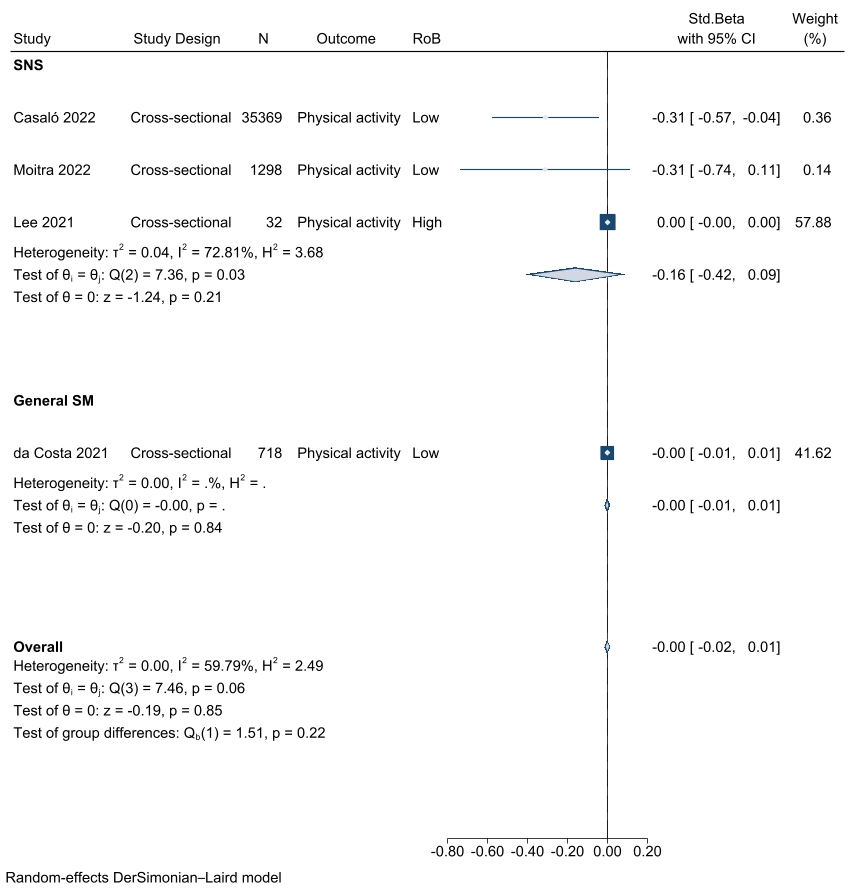

**Legend:** Figure presents forest plot for continuous exposure & continuous outcome subgroup analysis, with standardised beta (Std.Beta) used as common metric. Total number of study participants = 37,417. Abbreviations: CI = Confidence interval; N = Number of study participants; RoB = Risk of bias; SM = Social media; SNS = Social networking sites and Std.Beta = Standardised beta.

#### Unhealthy dietary behaviour

##### Effect direction plot

Figure BT demonstrate the effect direction in those studies investigating unhealthy dietary behaviours, by exposure. Two studies investigated more than one exposure.^33,161^ For time spent on social media and frequency of social media use, all studies reported harmful associations (time spent on social media: 95% CI 51.0 to 100.0%, study n=4, participant n=12,006, sign test p=0.13; frequency of social media use: 34.2 to 100.0%, study n=2, participant n = 826, insufficient data to conduct sign test). The relationship between exposure to health-risk behaviour content on social media and unhealthy dietary behaviours was investigated by four RCT’s (two rated low risk of bias (RoB) and two rated some concerns, via the Cochrane RoB-2 Tool), and three cross-sectional studies (two rated low RoB and one moderate). Considering all seven studies together, all studies reported harmful associations of social media (64.6 to 100.0%; study n=7; participant n=10,648; sign test p=0.02). When differentiating by study design, all RCT’s reported harmful effects (51.0 to 100.0%; study n=4; participant n=521; sign test p=0.13) and all cross-sectional studies reported harmful associations (43.9 to 100.0%; study n=3; participant n=10,127; sign test p=0.25).

##### Figure BT. Effect direction plot for studies of the association between social media use and adolescent unhealthy dietary behaviour, by social media exposure. Arrow size indicates sample size; arrow colour indicates study risk of bias

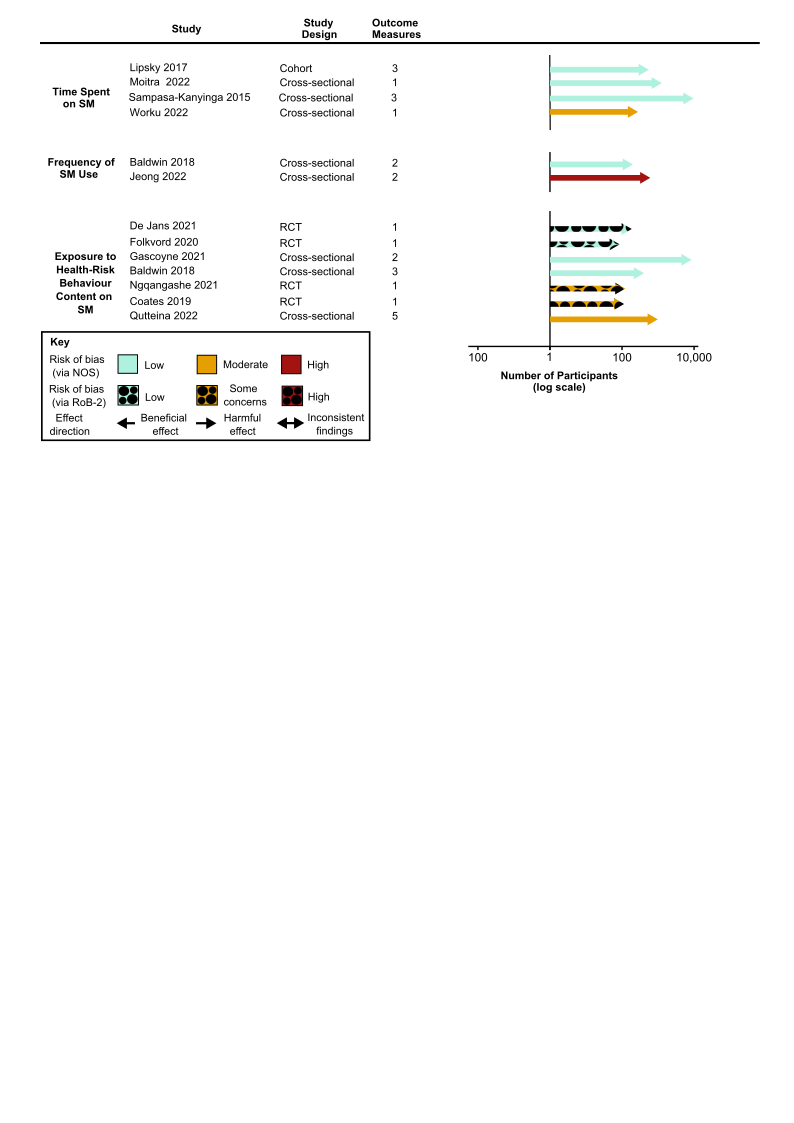

**Legend:** Sample size: represented by the size of the arrow, measured on a log scale. Outcome measure: number of outcome measures synthesised within each study. Studies organised by risk of bias grade, study design, and year of publication. Repeat cross-sectional studies, multiple study populations from different countries, and age subsets originating from the same study reported as separate studies. Abbreviations: NOS = Assessed via adapted Newcastle Ottawa Scale; RCT = Randomised Control Trial; RoB-2 = Assessed via Cochrane Risk of Bias 2 Tool; and SM = Social media.

##### Forest plots for meta-analyses and subgroup analyses

##### Figure BU. Forest plot for association between exposure to health-risk behaviour content on social media and unhealthy dietary behaviour

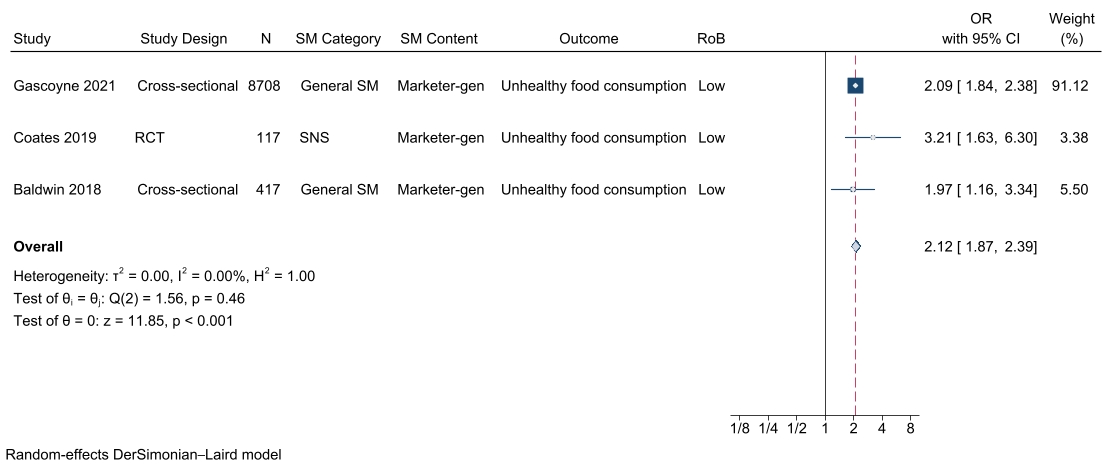

**Legend:** Figure presents forest plot for binary exposure (exposed vs unexposed) & binary/continuous outcome meta-analysis, with odds ratio (OR) used as common metric. Total number of study participants = 9242. Abbreviations: CI = Confidence interval; N = Number of study participants; OR = Odds ratio; RoB = Risk of bias; SM = Social media and SNS = Social networking sites.

##### Figure BV. Forest plot for association between exposure to health-risk behaviour content on social media and unhealthy dietary behaviour, by social media category

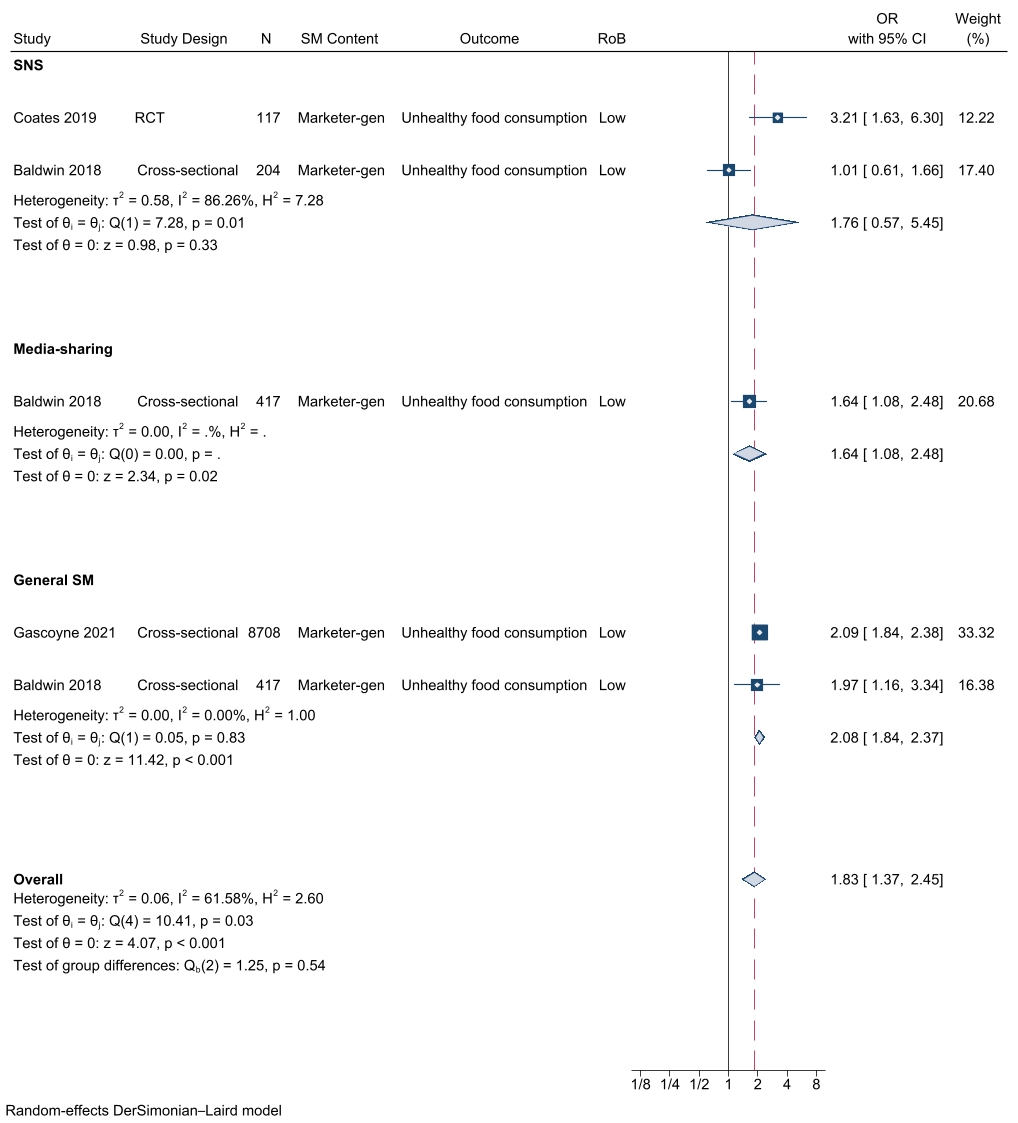

**Legend:** Figure presents forest plot for binary exposure (exposed vs unexposed) & binary/continuous outcome subgroup analysis, with odds ratio (OR) used as common metric. Total number of study participants = 9863. Abbreviations: CI = Confidence interval; N = Number of study participants; OR = Odds ratio; RoB = Risk of bias; SM = Social media and SNS = Social networking sites

#### Multiple risk behaviours

##### Effect direction plot

Figure BW demonstrates the effect direction in those studies (n=9) investigating multiple risk behaviours, by exposure. One study investigated more than one exposure type.^114^ For time spent on social media, the one study investigated reported a harmful association (95% CI 20.7 to 100.0%; participant n=500; insufficient data to conduct sign test), for frequency of social media use all studies demonstrated harmful associations (51.0 to 100.0%; study n=4; participant n=44,271; sign test p=0.13). Similarly, for exposure to health-risk behaviour content on social media, all studies demonstrated harmful associations (43.9 to 100.0%; study n=3; participant n=16,110,555; sign test p=0.25) and for other social media activities, the one study investigated reported a harmful association (20.7 to 100.0%; participant n=716; insufficient data to conduct sign test).

##### Figure BW. Effect direction plot for studies of the association between social media use and adolescent engagement in multiple risk behaviours, by social media exposure. Arrow size indicates sample size; arrow colour indicates study risk of bias

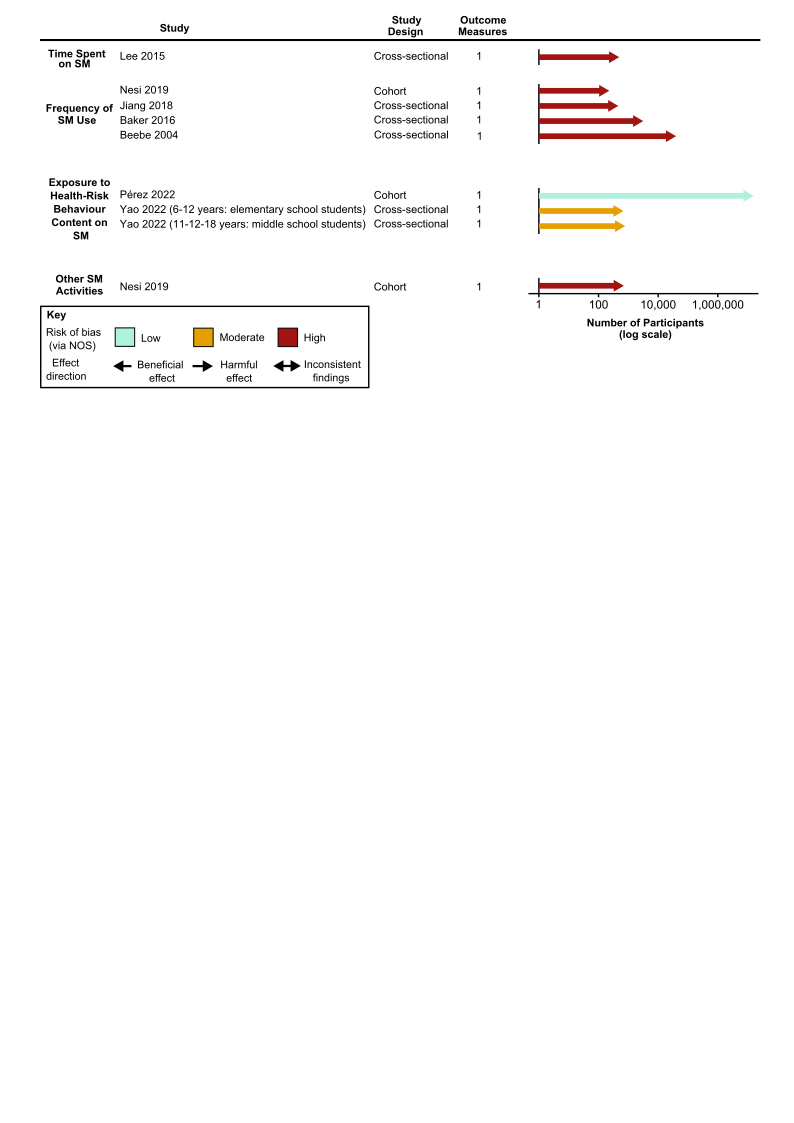

**Legend:** Sample size: represented by the size of the arrow, measured on a log scale. Outcome measure: number of outcome measures synthesised within each study. Studies organised by risk of bias grade, study design, and year of publication. Repeat cross-sectional studies, multiple study populations from different countries, and age subsets originating from the same study reported as separate studies. Abbreviations: NOS = Assessed via adapted Newcastle Ottawa Scale and SM = Social media.

##### Forest plots for meta-analyses and subgroup analyses

##### Figure BX. Forest plot for association between frequency of social media use and multiple risk behaviours

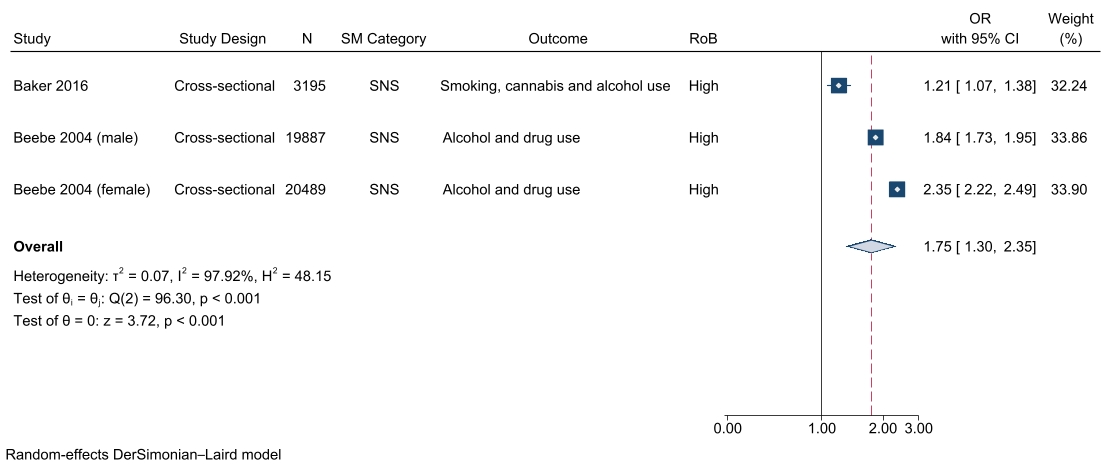

**Legend:** Figure presents forest plot for binary exposure (frequent/at all vs infrequent/not at all) & binary/continuous outcome meta-analysis, with odds ratio (OR) used as common metric. Total number of study participants = 43,571. Abbreviations: CI = Confidence interval; N = Number of study participants; OR = Odds ratio; RoB = Risk of bias; SM = Social media and SNS = Social networking sites.

#### Sensitivity analyses

##### Figure BY. Forest plot for association between exposure to health-risk behaviour content on social media and use of electronic nicotine delivery systems, by study design

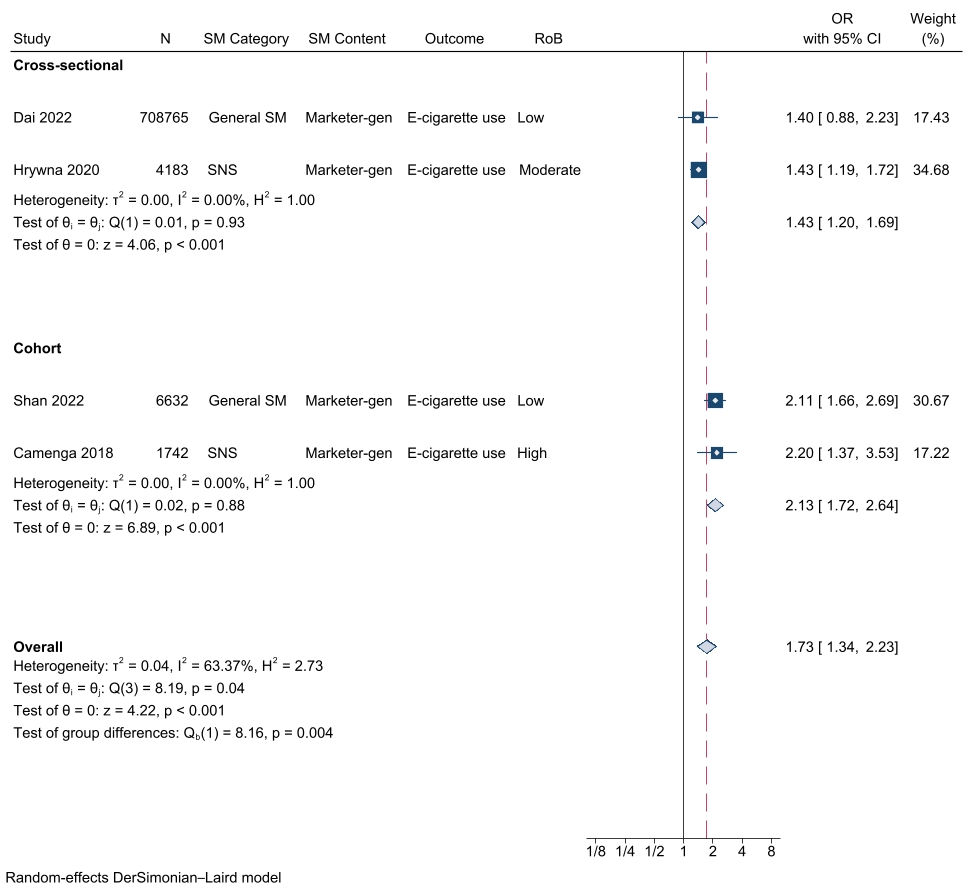

**Legend:** Figure presents forest plot for binary exposure (exposed vs unexposed) & binary/continuous outcome sensitivity analysis, with odds ratio (OR) used as common metric. Total number of study participants = 721,322. Abbreviations: CI = Confidence interval; N = Number of study participants; OR = Odds ratio; RoB = Risk of bias; SM = Social media and SNS = Social networking sites

##### Figure BZ. Forest plot for association between frequency of social media use and alcohol use, by study design

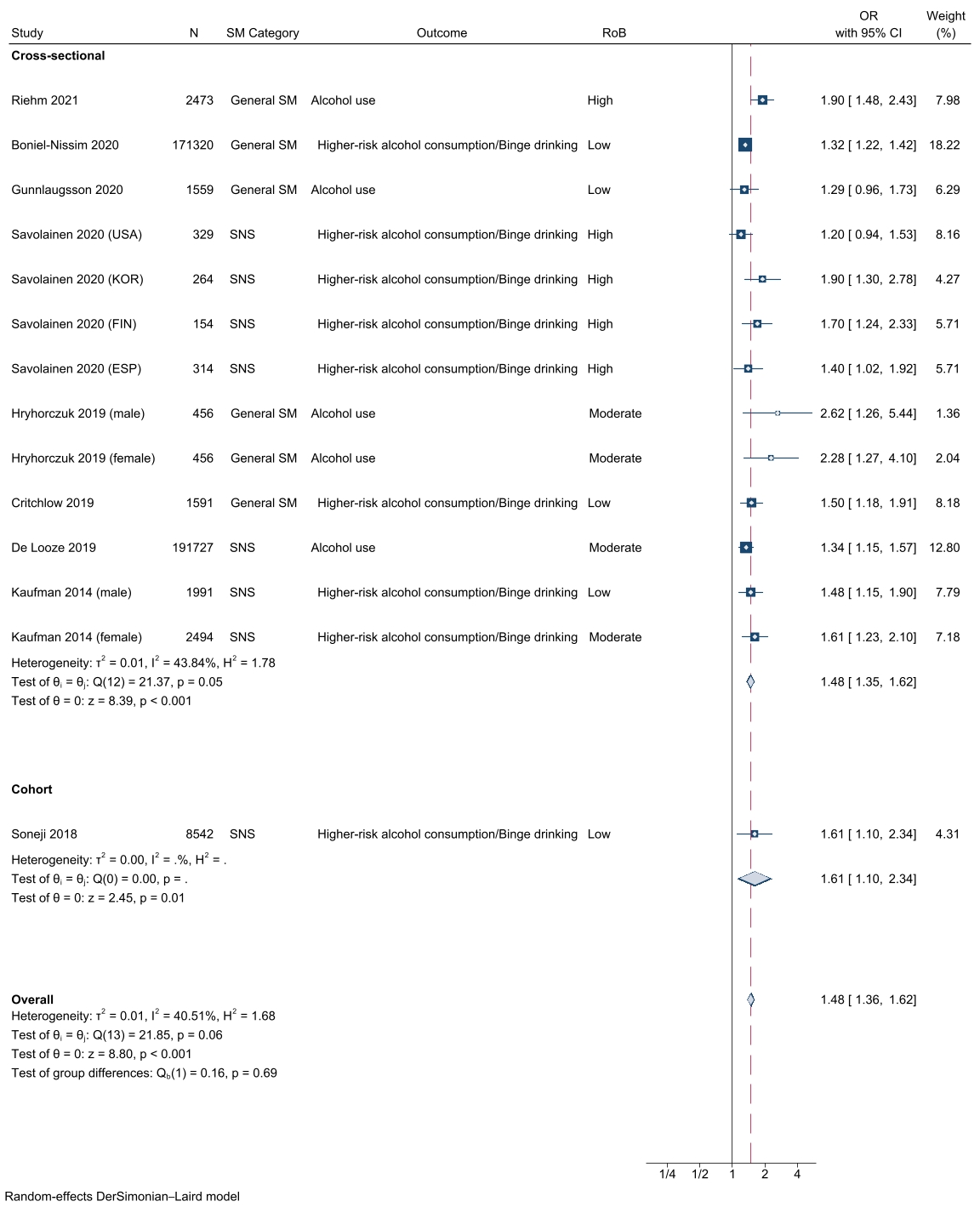

**Legend:** Figure presents forest plot for binary exposure (frequent/daily vs infrequent/non-daily) & binary/continuous outcome sensitivity analysis, with odds ratio (OR) used as common metric. Total number of study participants = 383,670. Abbreviations: CI = Confidence interval; ESP = Spain; FIN = Finland; KOR = South Korea; N = Number of study participants; OR = Odds ratio; RoB = Risk of bias; SM = Social media and SNS = Social networking sites.

##### Figure CA. Forest plot for association between frequency of social media use and alcohol use, by study design

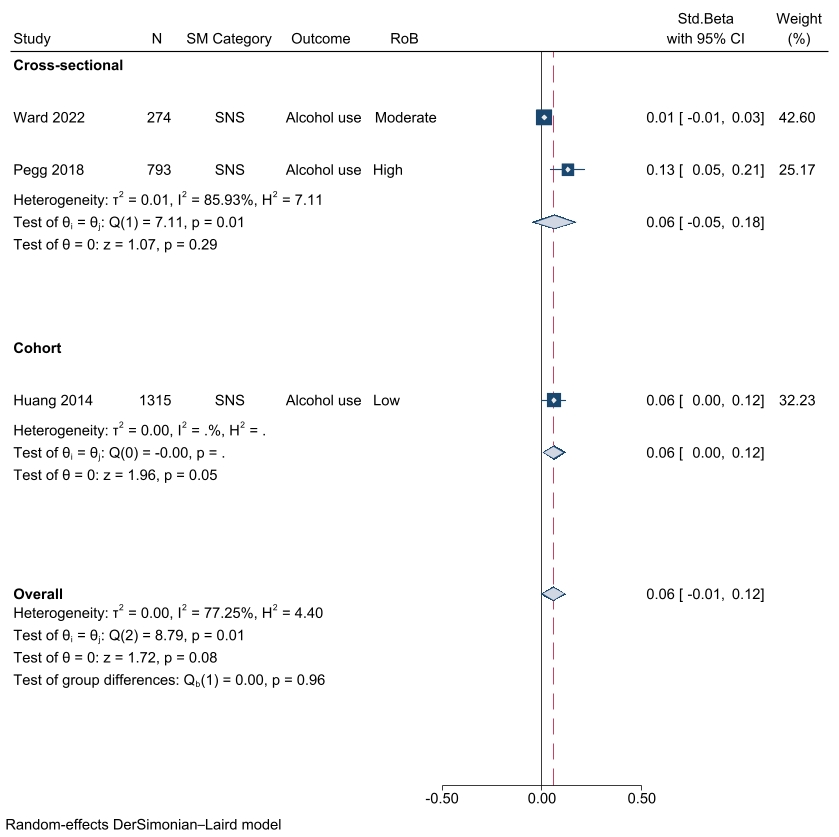

**Legend:** Figure presents forest plot for continuous exposure & continuous outcome sensitivity analysis, with standardised beta (Std.Beta) used as common metric. Total number of study participants = 2382. Abbreviation: CI = Confidence interval; N = Number of study participants; RoB = Risk of bias; SM = Social media; SNS = Social networking sites and Std. Beta = Standardised beta.

##### Figure CB. Forest plot for association between time spent on social media and alcohol use, by study design

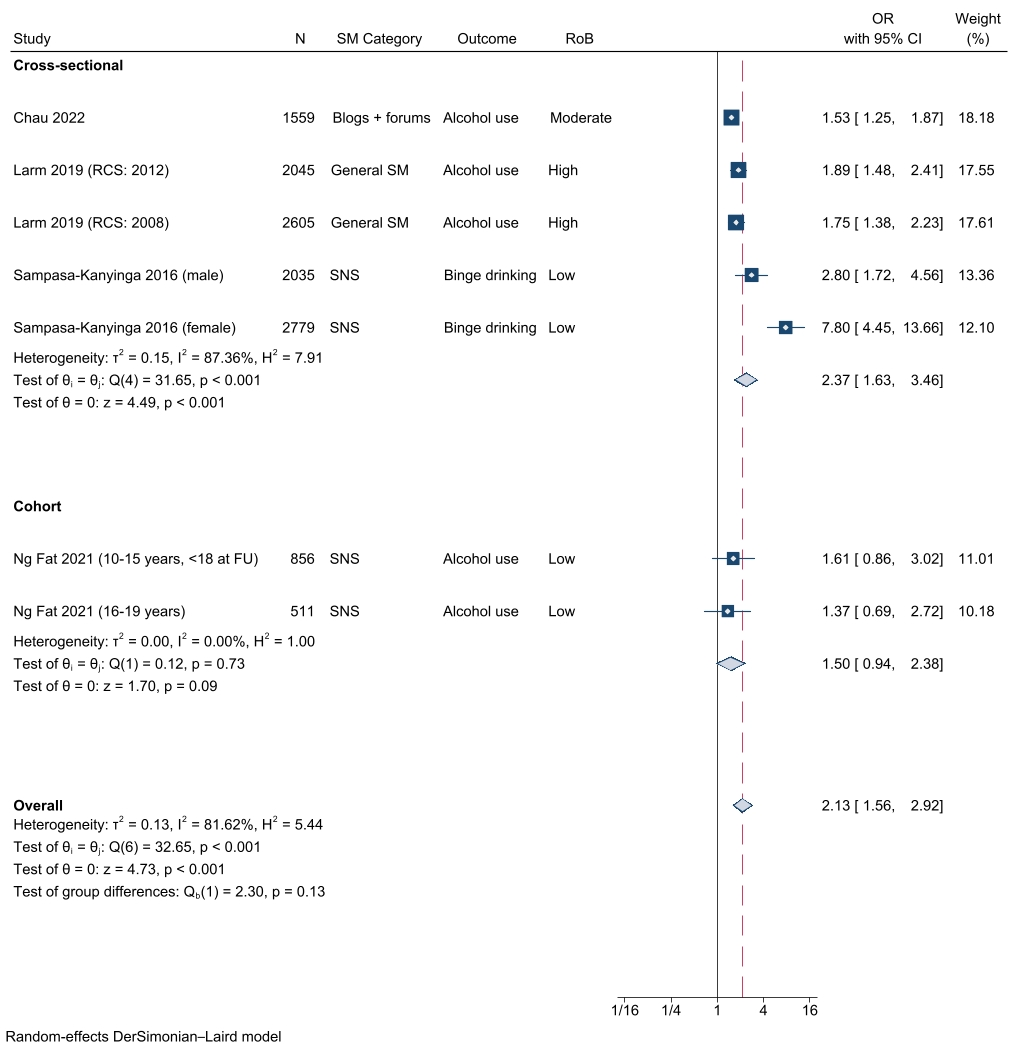

**Legend:** Figure presents forest plot for binary exposure (≥2 vs <2 hrs/day social media use) & binary/continuous outcome sensitivity analysis, with odds ratio (OR) used as common metric. Total number of study participants = 12,390. Abbreviations: CI = Confidence interval; FU = Follow up; hrs = Hours; N = Number of study participants; OR = Odds ratio; RCS = Repeat cross-sectional study; RoB = Risk of bias; SM = Social media and SNS = Social networking sites.

##### Figure CC. Forest plot for association between time spent on social media and alcohol use, by study design

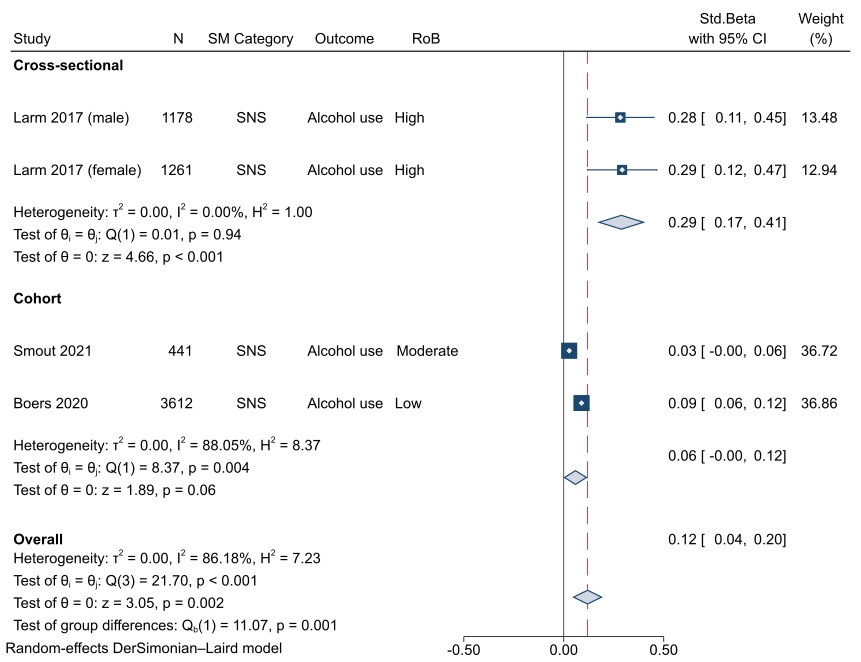

**Legend:** Figure presents forest plot for continuous exposure & continuous outcome sensitivity analysis, with standardised beta (Std.Beta) used as common metric. Total number of study participants = 6492. Abbreviation: CI = Confidence interval; N = Number of study participants; RoB = Risk of bias; SM = Social media; SNS = Social networking sites and Std. Beta = Standardised beta.

##### Figure CD. Forest plot for association between exposure to health-risk behaviour content on social media and alcohol use, by study design

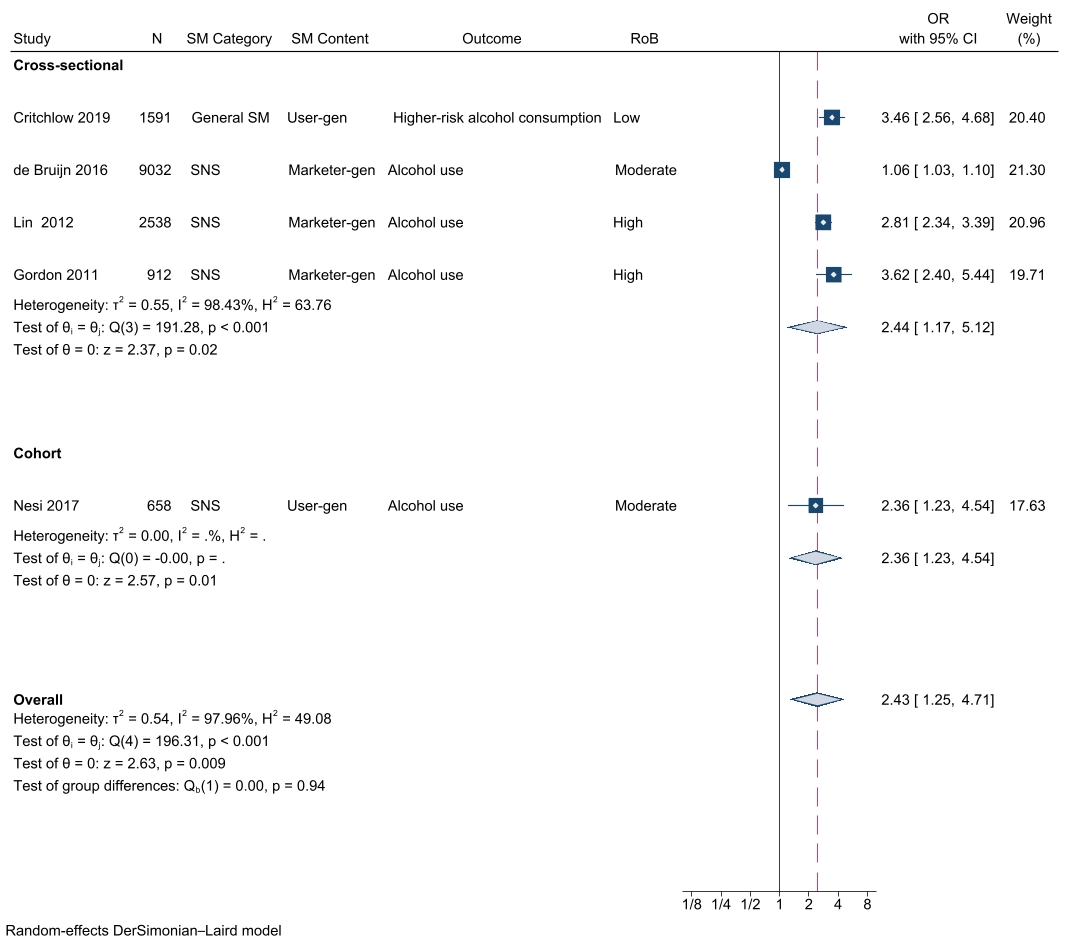

**Legend:** Figure presents forest plot for binary exposure (exposed vs unexposed) & binary/continuous outcome sensitivity analysis, with odds ratio (OR) used as common metric. Total number of study participants = 14,731. Abbreviations: CI = Confidence interval; Marketer-gen = Marketer-generated content; N = Number of study participants; OR = Odds ratio; RoB = Risk of bias; SM = Social media; SNS = Social networking sites and User-gen = User-generated content.

##### Figure CE. Forest plot for associations between exposure to health-risk behaviour content on social media and alcohol use, by study design

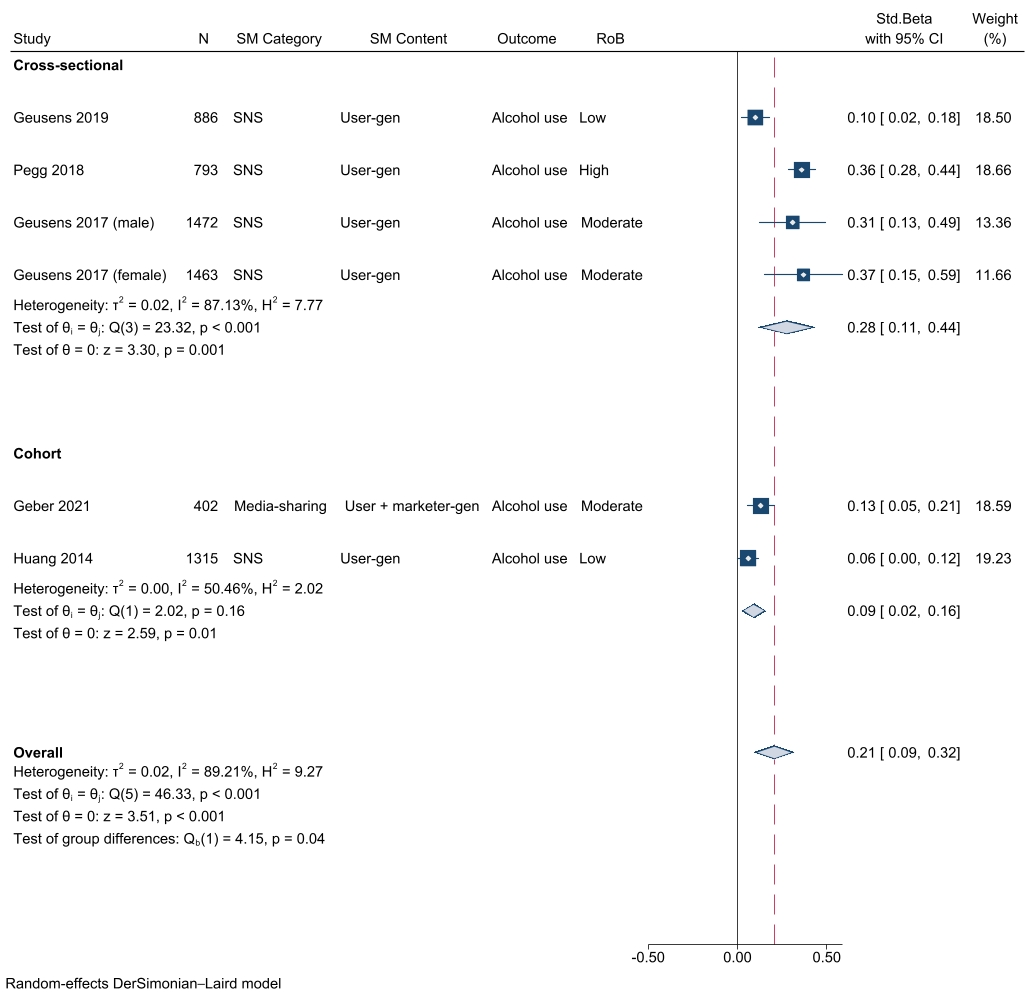

**Legend:** Figure presents forest plot for continuous exposure & continuous outcome sensitivity analysis, with standardised beta (Std.Beta) used as common metric. Total number of study participants = 6331. Abbreviation: CI = Confidence interval; N = Number of study participants; Marketer-gen = Marketer-generated content; RoB = Risk of bias; SM = Social media; SNS = Social networking sites; Std. Beta = Standardised beta and User-gen = User-generated content.

##### Figure CF. Forest plot for association between frequency of social media use and drug use, by study design

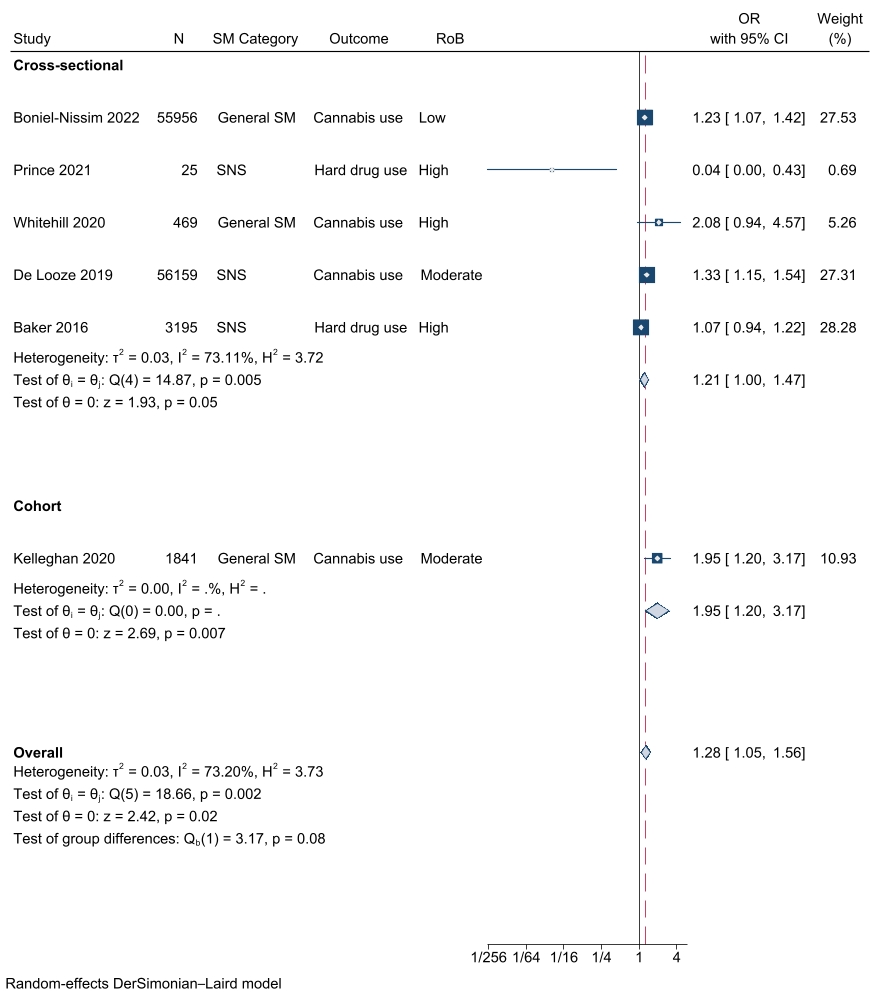

**Legend:** Figure presents forest plot for binary exposure (frequent/daily vs infrequent/non-daily) & binary/continuous outcome sensitivity analysis, with odds ratio (OR) used as common metric. Total number of study participants = 117,645. Abbreviations: CI = Confidence interval; N = Number of study participants; OR = Odds ratio; RoB = Risk of bias; SM = Social media and SNS = Social networking sites.

##### Figure CG. Forest plot for association between exposure to health-risk behaviour content on social media and tobacco use, by study design

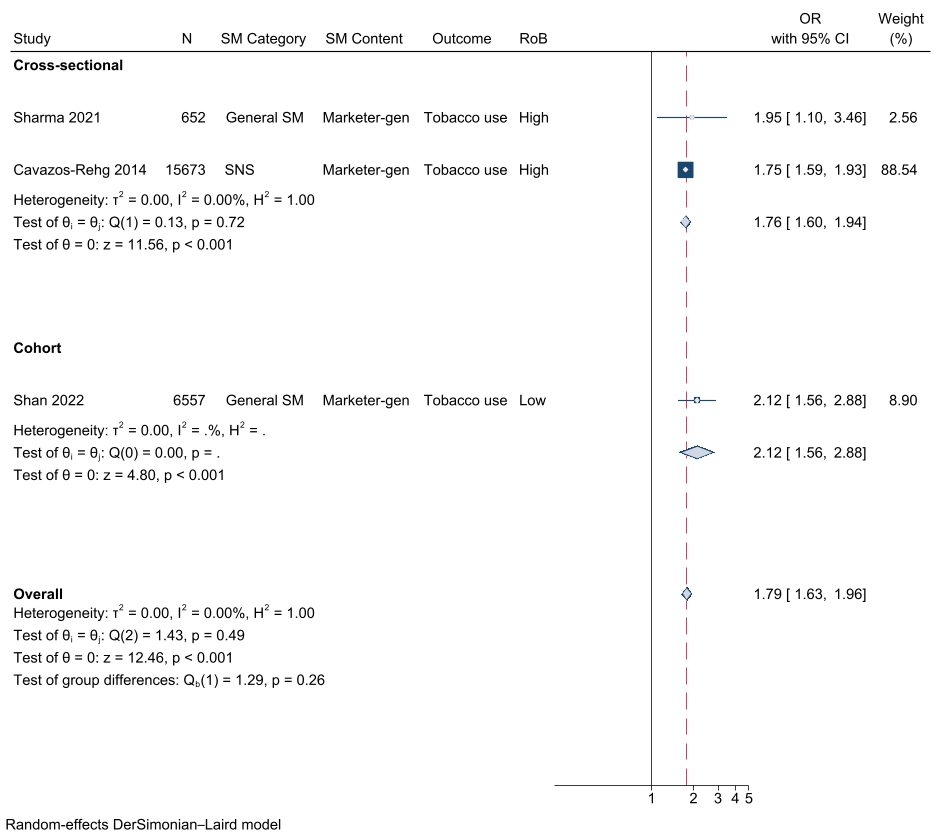

**Legend:** Figure presents forest plot for binary exposure (exposed vs unexposed) & binary/continuous outcome sensitivity analysis, with odds ratio (OR) used as common metric. Total number of study participants = 22,882. Abbreviations: CI = Confidence interval; Markter-gen = Marketer-generated content; N = Number of study participants; OR = Odds ratio; RoB = Risk of bias; SM = Social media and SNS = Social networking sites.

##### Figure CH. Forest plot for association between frequency of social media use and sexual risk behaviour, by study design

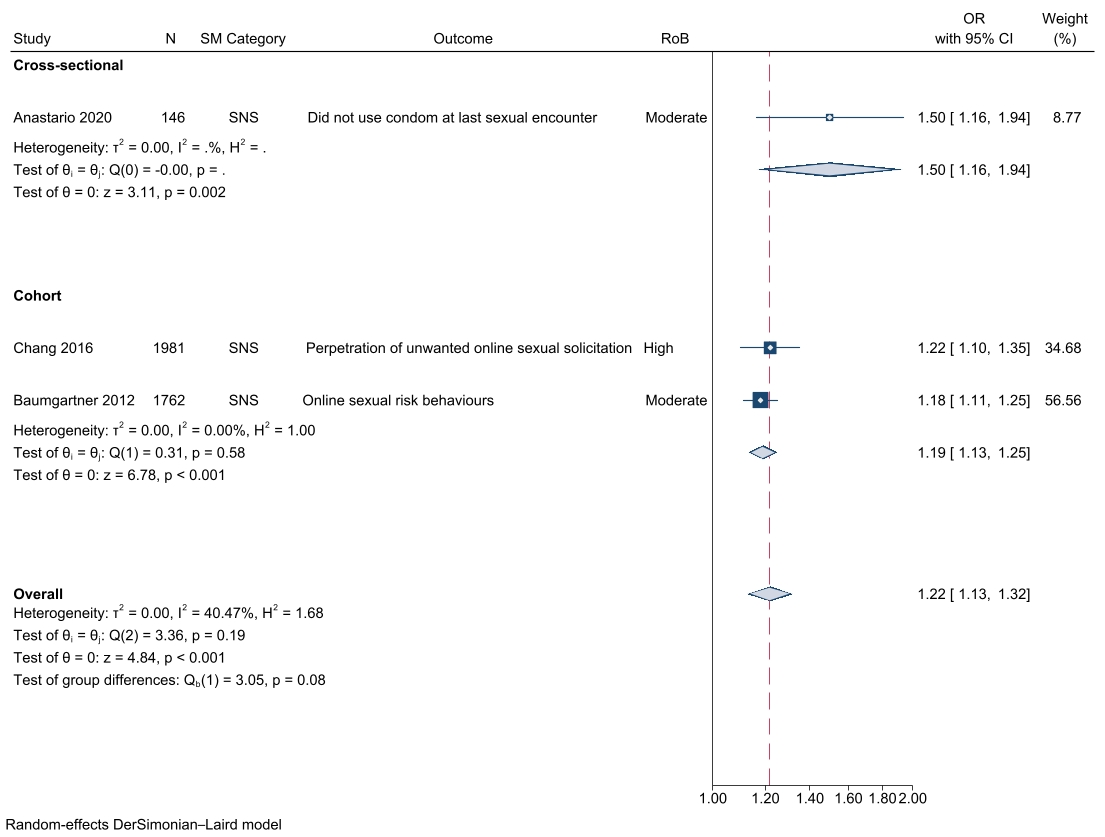

**Legend:** Figure presents forest plot for continuous exposure & binary outcome sensitivity analysis, with odds ratio (OR) used as common metric. Total number of study participants = 3889. Abbreviations: CI = Confidence interval; N = Number of study participants; OR = Odds ratio; RoB = Risk of bias; SM = Social media and SNS = Social networking sites.

##### Figure CI. Forest plot for association between frequency of social media use and sexual risk behaviour, by study design

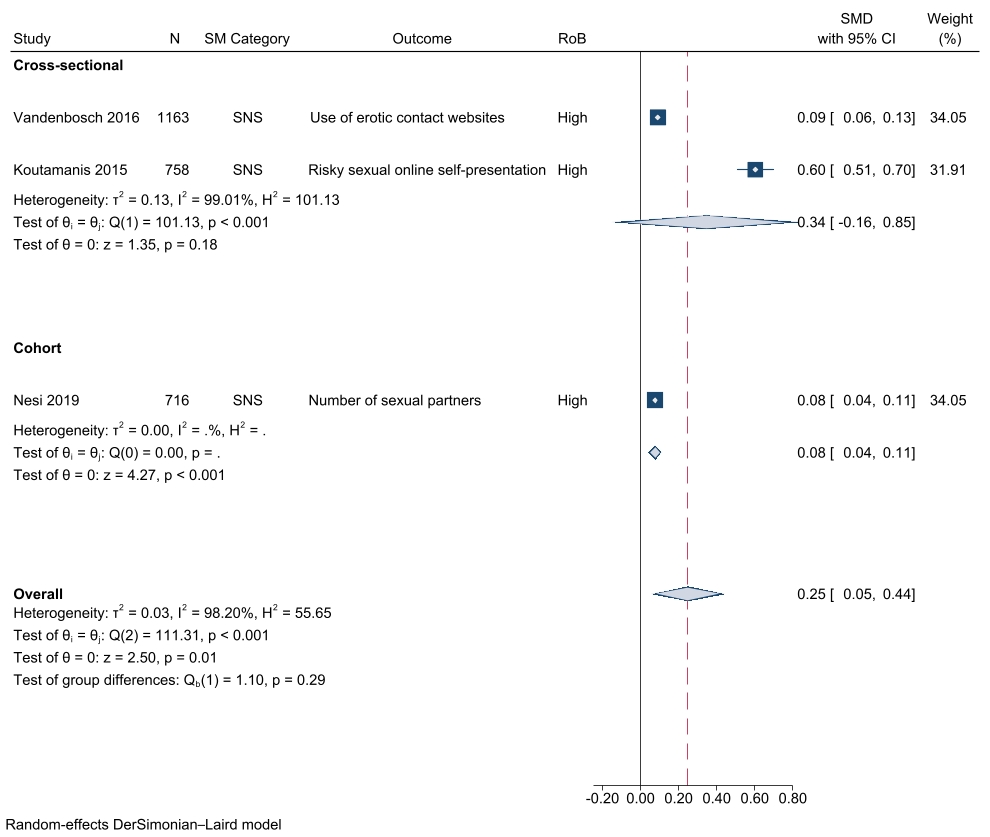

**Legend:** Figure presents forest plot for continuous exposure & continuous outcome sensitivity analysis, with standardised mean difference (SMD) used as common metric. Total number of study participants = 2637. Abbreviations: CI = Confidence interval; N = Number of study participants; RoB = Risk of bias; SM = Social media; SMD = Standardised mean difference and SNS = Social networking sites.

##### Figure CJ. Forest plot for association between frequency of social media use and gambling, by study design

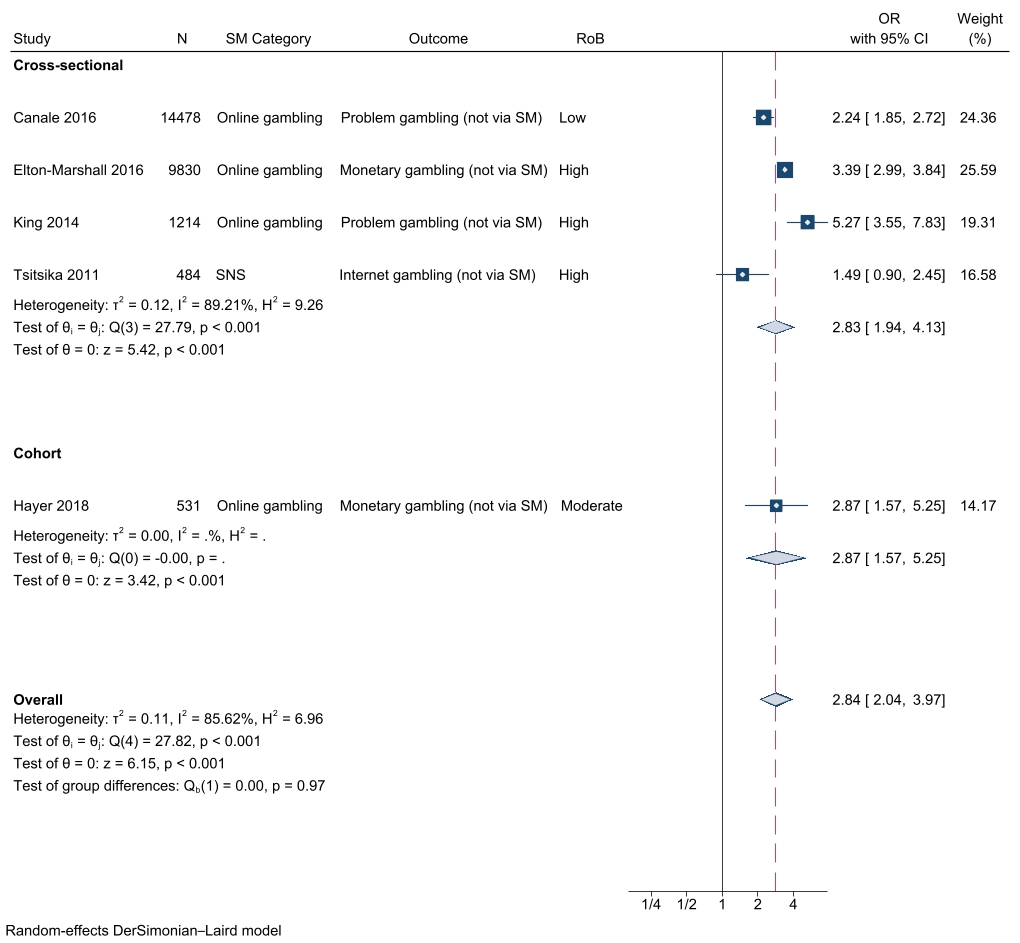

**Legend:** Figure presents forest plot for binary exposure (frequent/at all vs infrequent/not at all) & binary/continuous outcome sensitivity analysis, with odds ratio (OR) used as common metric. Total number of study participants = 26,537. Abbreviations: CI = Confidence interval; N = Number of study participants; OR = Odds ratio; RoB = Risk of bias; SM = Social media and SNS = Social networking sites.

##### Figure CK. Forest plot for association between frequency of social media use and anti-social behaviour, by study design

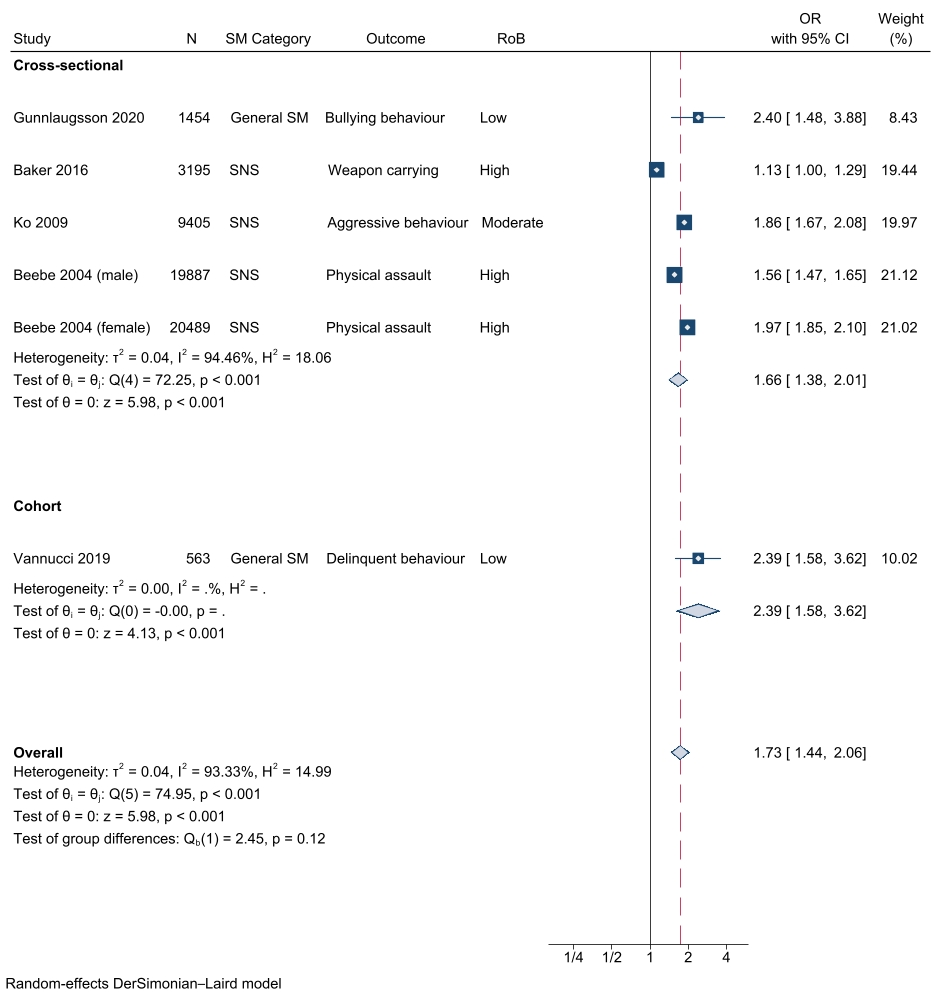

**Legend:** Figure presents forest plot for binary exposure (frequent/at all vs infrequent/not at all) & binary/continuous outcome sensitivity analysis, with odds (OR) used as common metric. Total number of study participants = 54,993. Abbreviations: CI = Confidence interval; N = Number of study participants; OR = Odds ratio; RoB = Risk of bias; SM = Social media and SNS = Social networking sites

##### Figure CL. Forest plot for association between exposure to health-risk behaviour content on social media and unhealthy dietary behaviour, by study design

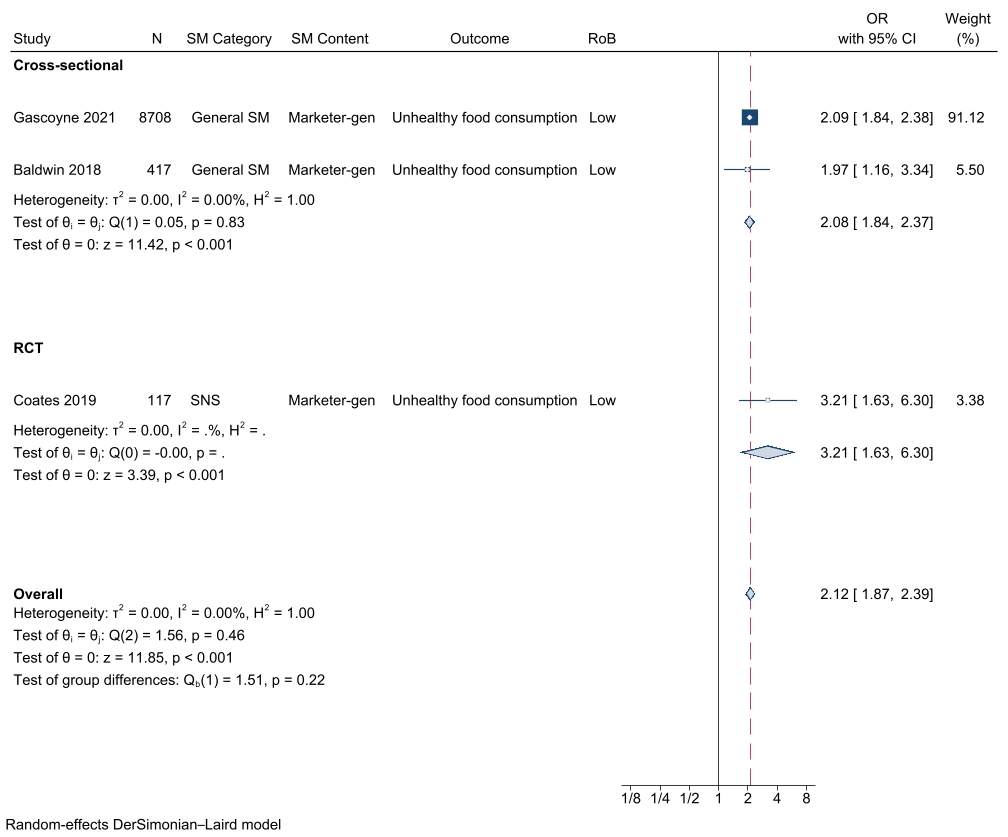

**Legend:** Figure presents forest plot for binary exposure (exposed vs unexposed) & binary/continuous outcome sensitivity analysis, with odds ratio (OR) used as common metric. Total number of study participants = 9242. Abbreviations: CI = Confidence interval; N = Number of study participants; OR = Odds ratio; RoB = Risk of bias; SM = Social media and SNS = Social networking sites.

##### Figure CM. Forest plot for association between frequency of social media use and alcohol use, by adjustment for critical confounding domains^a^

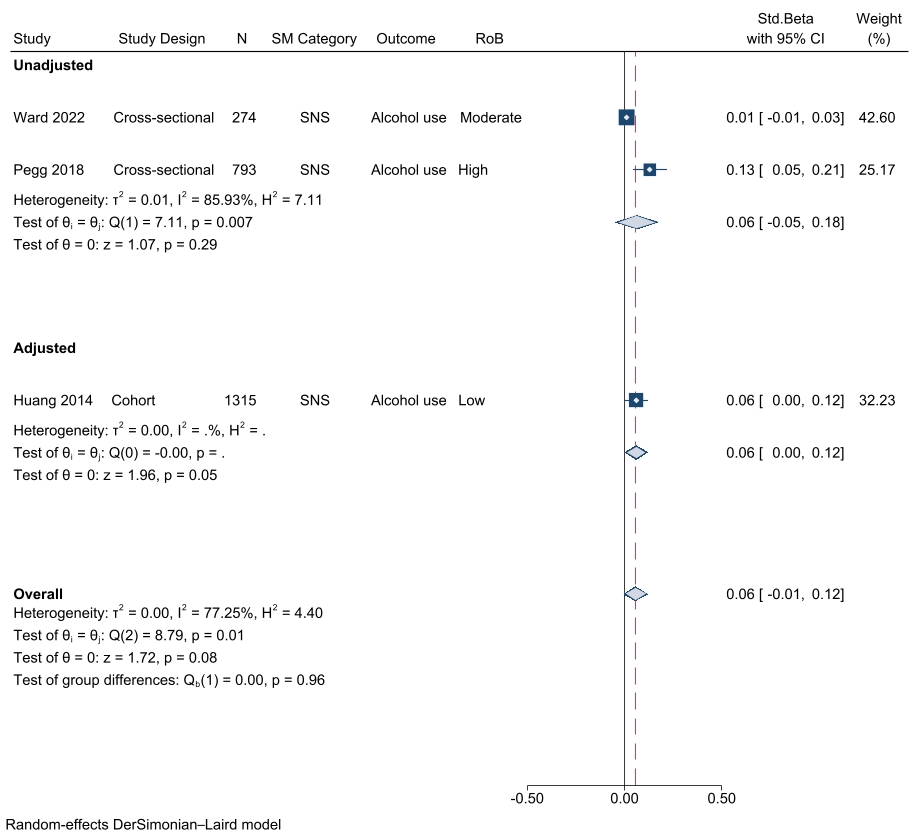

**Legend:** Figure presents forest plot for continuous exposure & continuous outcome sensitivity analysis, with standardised beta (Std.Beta) used as common metric. ^a^ Critical confounding domains: age, sex, and socioeconomic position (SEP). Total number of study participants = 2382. Abbreviation: CI = Confidence interval; N = Number of study participants; RoB = Risk of bias; SM = Social media; SNS = Social networking sites and Std. Beta = Standardised beta.

##### Figure CN. Forest plot for association between time spent on social media and alcohol use, by adjustment for critical confounding domains^a^

**Legend:** Figure presents forest plot for continuous exposure & continuous outcome sensitivity analysis, with standardised beta (Std.Beta) used as common metric. ^a^ Critical confounding domains: age, sex, and socioeconomic position (SEP). Total number of study participants = 6492. Abbreviation: CI = Confidence interval; N = Number of study participants; RoB = Risk of bias; SM = Social media; SNS = Social networking sites and Std. Beta = Standardised beta.

##### Figure CO. Forest plot for association between exposure to health-risk behaviour content on social media and alcohol use, by adjustment for critical confounding domains^a^

**Legend:** Figure presents forest plot for binary exposure (exposed vs unexposed) & binary/continuous outcome sensitivity analysis, with odds ratio (OR) used as common metric. ^a^ Critical confounding domains: age, sex, and socioeconomic position (SEP). Total number of study participants = 14,731. Abbreviations: CI = Confidence interval; Marketer-gen = Marketer-generated content; N = Number of study participants; OR = Odds ratio; RoB = Risk of bias; SM = Social media; SNS = Social networking sites and User-gen = User-generated content.

##### Figure CP. Forest plot for association between frequency of social media use and drug use, by adjustment for critical confounding domains^a^

**Legend:** Figure presents forest plot for binary exposure (frequent/daily vs infrequent/non-daily) & binary/continuous outcome sensitivity analysis, with odds ratio (OR) used as common metric. ^a^ Critical confounding domains: age, sex, and socioeconomic position (SEP). Total number of study participants = 117,645. Abbreviations: CI = Confidence interval; N = Number of study participants; OR = Odds ratio; RoB = Risk of bias; SM = Social media and SNS = Social networking sites.

##### Figure CQ. Forest plot for association between exposure to health-risk behaviour content on social media and tobacco use, by adjustment for critical confounding domains^a^

**Legend:** Figure presents forest plot for binary exposure (exposed vs unexposed) & binary/continuous outcome sensitivity analysis, with odds ratio (OR) used as common metric. ^a^ Critical confounding domains: age, sex, and socioeconomic position (SEP). Total number of study participants = 22,882. Abbreviations: CI = Confidence interval; Markter-gen = Marketer-generated content; N = Number of study participants; OR = Odds ratio; RoB = Risk of bias; SM = Social media and SNS = Social networking sites.

##### Figure CR. Forest plot for association between exposure to health-risk behaviour content on social media and use of electronic nicotine delivery systems, by adjustment for critical confounding domains^a^

**Legend:** Figure presents forest plot for binary exposure (exposed vs unexposed) & binary/continuous outcome sensitivity analysis, with odds ratio (OR) used as common metric. ^a^ Critical confounding domains: age, sex, and socioeconomic position (SEP)**.** Total number of study participants = 721,322. Abbreviations: CI = Confidence interval; N = Number of study participants; OR = Odds ratio; RoB = Risk of bias; SM = Social media and SNS = Social networking sites

##### Figure CS. Forest plot for association between frequency of social media use and sexual risk behaviour, by adjustment for critical confounding domains^a^

**Legend:** Figure presents forest plot for binary exposure (frequent/at all vs infrequent/not at all) & binary/continuous outcome sensitivity analysis, with odds ratio (OR) used as common metric. ^a^ Critical confounding domains: age, sex, and socioeconomic position (SEP). Total number of study participants = 47,325. Abbreviations: CI = Confidence interval; N = Number of study participants; OR = Odds ratio; RoB = Risk of bias; SM = Social media and SNS = Social networking sites.

##### Figure CT. Forest plot for association between frequency of social media use and gambling, by adjustment for critical confounding domains^a^

**Legend:** Figure presents forest plot for binary exposure (frequent/at all vs infrequent/not at all) & binary/continuous outcome sensitivity analysis, with odds ratio (OR) used as common metric. ^a^ Critical confounding domains: age, sex, and socioeconomic position (SEP). Total number of study participants = 26,537. Abbreviations: CI = Confidence interval; N = Number of study participants; OR = Odds ratio; RoB = Risk of bias; SM = Social media and SNS = Social networking sites

##### Figure CU. Forest plot for association between frequency of social media use and anti-social behaviour, by adjustment for critical confounding domains^a^

**Legend:** Figure presents forest plot for binary exposure (frequent/at all vs infrequent/not at all) & binary/continuous outcome sensitivity analysis, with odds (OR) used as common metric. ^a^ Critical confounding domains: age, sex, and socioeconomic position (SEP). Total number of study participants = 54,993. Abbreviations: CI = Confidence interval; N = Number of study participants; OR = Odds ratio; RoB = Risk of bias; SM = Social media and SNS = Social networking sites

##### Figure CV. Forest plot for association between time spent on social media and inadequate physical activity, by adjustment for critical confounding domains^a^

**Legend:** Figure presents forest plot for continuous exposure & continuous outcome sensitivity analysis, with standardised beta (Std.Beta) used as common metric. ^a^ Critical confounding domains: age, sex, and socioeconomic position (SEP). Total number of study participants =37,417. Abbreviations: CI = Confidence interval; N = Number of study participants; RoB = Risk of bias; SM = Social media; SNS = Social networking sites and Std.Beta = Standardised beta.

##### Figure CX. Forest plot for association between time spent on social media and alcohol use, by adjustment for critical confounding domains^a^

**Legend:** Figure presents forest plot for binary exposure (≥2 vs <2 hrs/day social media use) & binary/continuous outcome sensitivity analysis, with odds ratio (OR) used as common metric. ^a^ Critical confounding domains: age, sex, and socioeconomic position (SEP). Total number of study participants = 12,390. Abbreviations: CI = Confidence interval; FU = Follow up; hrs = Hours; N = Number of study participants; OR = Odds ratio; RCS = Repeat cross-sectional study; RoB = Risk of bias; SM = Social media and SNS = Social networking sites.

##### Figure CY. Forest plot for association between exposure to health-risk behaviour content on social media and alcohol use, by adjustment for critical confounding domains^a^

**Legend:** Figure presents forest plot for continuous exposure & continuous outcome sensitivity analysis, with standardised beta (Std.Beta) used as common metric. ^a^ Critical confounding domains: age, sex, and socioeconomic position (SEP). Total number of study participants = 6331. Abbreviation: CI = Confidence interval; N = Number of study participants; Marketer-gen = Marketer-generated content; RoB = Risk of bias; SM = Social media; SNS = Social networking sites; Std. Beta = Standardised beta and User-gen = User-generated content.

##### Figure CZ. Forest plot for association between frequency of social media use and tobacco use, by adjustment for critical confounding domains^a^

**Legend:** Figure presents forest plot for binary exposure (frequent vs infrequent) & binary/continuous outcome sensitivity analysis, with odds ratio (OR) used as common metric. ^a^ Critical confounding domains: age, sex, and socioeconomic position (SEP)**.** Total number of study participants = 424,326. Abbreviations: CI = Confidence interval; N = Number of study participants; OR = Odds ratio; RoB = Risk of bias; SM = Social media and SNS = Social networking sites

##### Figure DA. Forest plot for association between frequency of social media use and alcohol use, by risk of bias grade

**Legend:** Figure presents forest plot for binary exposure (frequent/daily vs infrequent/non-daily) & binary/continuous outcome sensitivity analysis, with odds ratio (OR) used as common metric. Total number of study participants = 383,670. Abbreviations: CI = Confidence interval; ESP = Spain; FIN = Finland; KOR = South Korea; N = Number of study participants; OR = Odds ratio; SM = Social media and SNS = Social networking sites.

##### Figure DB. Forest plot for association between exposure to health-risk behaviour content on social media and alcohol use, by risk of bias grade

**Legend:** Figure presents forest plot for continuous exposure & continuous outcome sensitivity analysis, with standardised beta (Std.Beta) used as common metric. Total number of study participants = 6331. Abbreviation: CI = Confidence interval; N = Number of study participants; Marketer-gen = Marketer-generated content; SM = Social media; SNS = Social networking sites; Std. Beta = Standardised beta and User-gen = User-generated content.

##### Figure DC. Forest plot for association between time spent on social media and alcohol use, by risk of bias grade

**Legend:** Figure presents forest plot for binary exposure (≥2 vs <2 hrs/day social media use) & binary/continuous outcome sensitivity analysis, with odds ratio (OR) used as common metric. Total number of study participants = 12,390. Abbreviations: CI = Confidence interval; FU = Follow up; hrs = Hours; N = Number of study participants; OR = Odds ratio; RCS = Repeat cross-sectional study; SM = Social media and SNS = Social networking sites.

##### Figure DD. Forest plot for association between exposure to health-risk behaviour content on social media and alcohol use, by risk of bias grade

**Legend:** Figure presents forest plot for binary exposure (exposed vs unexposed) & binary/continuous outcome sensitivity analysis, with odds ratio (OR) used as common metric. ^a^ Critical confounding domains: age, sex, and socioeconomic position (SEP). Total number of study participants = 14,731. Abbreviations: CI = Confidence interval; Marketer-gen = Marketer-generated content; N = Number of study participants; OR = Odds ratio; SM = Social media; SNS = Social networking sites and User-gen = User-generated content.

##### Figure DE. Forest plot for association between frequency of social media use and drug use, by risk of bias grade

**Legend:** Figure presents forest plot for binary exposure (frequent/daily vs infrequent/non-daily) & binary/continuous outcome sensitivity analysis, with odds ratio (OR) used as common metric. Total number of study participants = 117,645. Abbreviations: CI = Confidence interval; N = Number of study participants; OR = Odds ratio; SM = Social media and SNS = Social networking sites.

##### Figure DF. Forest plot for association between time spent on social media and drug use, by risk of bias grade

**Legend:** Figure presents forest plot for binary exposure (≤1hrs vs >1hr/day) & binary/continuous outcome meta-analysis, with odds ratio (OR) used as common metric. Total number of study participants = 7357. Abbreviations: CI = Confidence interval; N = Number of study participants; OR = Odds ratio; SM = Social media and SNS = Social networking sites.

##### Figure DG. Forest plot for association between frequency of social media use and sexual risk behaviour, by risk of bias grade

**Legend:** Figure presents forest plot for binary exposure (frequent/at all vs infrequent/not at all) & binary/continuous outcome sensitivity analysis, with odds ratio (OR) used as common metric. Total number of study participants = 47,325. Abbreviations: CI = Confidence interval; N = Number of study participants; OR = Odds ratio; SM = Social media and SNS = Social networking sites.

##### Figure DH. Forest plot for association between frequency of social media use and sexual risk behaviour, by risk of bias grade

**Legend:** Figure presents forest plot for continuous exposure & binary outcome sensitivity analysis, with odds ratio (OR) used as common metric. Total number of study participants = 3889. Abbreviations: CI = Confidence interval; N = Number of study participants; OR = Odds ratio; SM = Social media and SNS = Social networking sites.

##### Figure DI. Forest plot for association between frequency of social media use and anti-social behaviour, by risk of bias grade

**Legend:** Figure presents forest plot for binary exposure (frequent/at all vs infrequent/not at all) & binary/continuous outcome sensitivity analysis, with odds (OR) used as common metric. Total number of study participants = 54,993. Abbreviations: CI = Confidence interval; N = Number of study participants; OR = Odds ratio; SM = Social media and SNS = Social networking sites

##### Figure DJ. Forest plot for association between frequency of social media use and tobacco use, by risk of bias grade

**Legend:** Figure presents forest plot for binary exposure (frequent vs infrequent) & binary/continuous outcome sensitivity analysis, with odds ratio (OR) used as common metric. Total number of study participants = 424,326. Abbreviations: CI = Confidence interval; N = Number of study participants; OR = Odds ratio; SM = Social media and SNS = Social networking sites.

##### Figure DK. Forest plot for association between exposure to health-risk behaviour content on social media and tobacco use, by risk of bias

**Legend:** Figure presents forest plot for binary exposure (exposed vs unexposed) & binary/continuous outcome sensitivity analysis, with odds ratio (OR) used as common metric. Total number of study participants = 22,882. Abbreviations: CI = Confidence interval; Marketer-gen = Marketer-generated content; N = Number of study participants; OR = Odds ratio; SM = Social media and SNS = Social networking sites.

##### Figure DL. Forest plot for association between frequency of social media use and gambling, by risk of bias grade

**Legend:** Figure presents forest plot for binary exposure (frequent/at all vs infrequent/not at all) & binary/continuous sensitivity analysis, with odds ratio (OR) used as common metric. Total number of study participants = 26,537. Abbreviations: CI = Confidence interval; N = Number of study participants; OR = Odds ratio; SM = Social media and SNS = Social networking sites.

##### Figure DM. Forest plot for association between exposure to health-risk behaviour content on social media and use of electronic nicotine delivery systems, by risk of bias grade

**Legend:** Figure presents forest plot for binary exposure (exposed vs unexposed) & binary/continuous outcome sensitivity analysis with odds ratio (OR) used as common metric. Total number of study participants = 721,322. Abbreviations: CI = Confidence interval; N = Number of study participants; OR = Odds ratio; SM = Social media and SNS = Social networking sites.

##### Figure DN. Forest plot for association between time spent on social media and inadequate physical activity, by risk of bias grade

**Legend:** Figure presents forest plot for continuous exposure & continuous outcome sensitivity analysis, with standardised beta (Std.Beta) used as common metric. Total number of study participants =37,417. Abbreviations: CI = Confidence interval; N = Number of study participants; SM = Social media; SNS = Social networking sites and Std.Beta = Standardised beta.

##### Figure DO. Forest plot for association between time spent on social media and alcohol use, excluding datapoints which overlap 10-19 years

**Legend:** Figure presents forest plot for binary exposure (≥2 vs <2 hrs/day social media use) & binary/continuous outcome sensitivity analysis, with odds ratio (OR) used as common metric. Total number of study participants = 7576. Abbreviations: CI = Confidence interval; FU = Follow up; hrs = Hours; N = Number of study participants; OR = Odds ratio; RCS = Repeat cross-sectional study; RoB = Risk of bias; SM = Social media and SNS = Social networking sites.

##### Figure DP. Forest plot for association between frequency of social media use and alcohol use, excluding datapoints which overlap 10-19 years

**Legend:** Figure presents forest plot for binary exposure (frequent/daily vs infrequent/non-daily) & binary/continuous outcome sensitivity analysis, with odds ratio (OR) used as common metric. Total number of study participants = 377,626. Abbreviations: CI = Confidence interval; ESP = Spain; FIN = Finland; KOR = South Korea; N = Number of study participants; OR = Odds ratio; RoB = Risk of bias; SM = Social media and SNS = Social networking sites.

##### Figure DQ. Forest plot for association between exposure to health-risk behaviour content on social media and alcohol use, excluding datapoints which overlap 10-19 years 0.21 0.09-0.32

**Legend:** Figure presents forest plot for continuous exposure & continuous outcome sensitivity analysis, with standardised beta (Std.Beta) used as common metric. Total number of study participants = 3396. Abbreviation: CI = Confidence interval; N = Number of study participants; Marketer-gen = Marketer-generated content; RoB = Risk of bias; SM = Social media; SNS = Social networking sites; Std. Beta = Standardised beta and User-gen = User-generated content.

##### Figure DR. Forest plot for association between frequency of social media use and tobacco use, excluding datapoints which overlap 10-19 years

**Legend:** Figure presents forest plot for binary exposure (frequent vs infrequent) & binary/continuous outcome sensitivity analysis, with odds ratio (OR) used as common metric. Total number of study participants = 422,760. Abbreviations: CI = Confidence interval; N = Number of study participants; OR = Odds ratio; RoB = Risk of bias; SM = Social media and SNS = Social networking sites.

##### Figure DS. Forest plot for association between frequency of social media use and sexual risk behaviour, excluding datapoints which overlap 10-19 years

**Legend:** Figure presents forest plot for binary exposure (frequent/at all vs infrequent/not at all) & binary/continuous outcome sensitivity analysis, with odds ratio (OR) used as common metric. Total number of study participants = 42,312. Abbreviations: CI = Confidence interval; N = Number of study participants; OR = Odds ratio; RoB = Risk of bias; SM = Social media and SNS = Social networking sites.

##### Figure DT. Forest plot for association between frequency of social media use and anti-social behaviour, excluding datapoints which overlap 10-19 years

**Legend:** Figure presents forest plot for binary exposure (frequent/at all vs infrequent/not at all) & binary/continuous outcome sensitivity analysis, with odds (OR) used as common metric. Total number of study participants = 53,539. Abbreviations: CI = Confidence interval; N = Number of study participants; OR = Odds ratio; RoB = Risk of bias; SM = Social media and SNS = Social networking sites.

##### Table A. Summary of meta-regression findings

| **Exposure** | **Outcome** | **Study level variable** | **Coefficient (95% CI)** | **p-value** | **Related forest plot** |
| --- | --- | --- | --- | --- | --- |
| Frequency of SM use (binary exposure) | Tobacco use | SM category (reference category- SNS) | General SM: -0.27 (-0.61, 0.07) | 0.12 | Figure AJ |
| Frequency of SM use  (binary exposure) | Sexual risk behaviour | Average age of study participants (reference category- <16 years) | **≥16 years: -0.36 (-0.68, -0.04)** | **0.03** | Figure AS |
|  |  | SM category (reference category- General SM) | SNS: 0.30 (-0.07, 0.67) | 0.12 | Figure AT |
|  |  | Development status of study setting (reference category- Low-Middle income) | High income: 0.11 (-0.23, 0.45) | 0.52 | Figure AU |

**Legend:** Results with p < 0.05 in bold. Abbreviations: 95% CI = 95% confidence interval; SM = Social media and SNS = Social networking sites.

### Appendix-17. Assessment of publication bias/small study effects

##### Figure A. Contour enhanced funnel plot for meta-analysis of studies investigating the effect of frequency of social media use (frequent vs infrequent) on sexual risk behaviour, and Egger’s test result

### Appendix-18. Summary of findings and certainty of evidence

##### Table A. Summary of findings and certainty of evidence for seven priority outcomes (as per GRADE) with reasons for upgrading/downgrading of the evidence

| **Population/setting**: Adolescents aged 10-19 years in high and low-middle income settings  **Intervention**: Frequent social media use  **Comparison**: Infrequent social media use | | | | | | |
| --- | --- | --- | --- | --- | --- | --- |
| **Outcome** | **Anticipated absolute effects^a^ (95% CI)** | | **Relative effect (95% CI)** | **No. of participants**  **(studies)** | **Certainty of the evidence (GRADE)** | **Comments** |
|  | **Risk with infrequent social media use** | **Risk with frequent social media use** |  |  |  |  |
| Alcohol use | 48.9% of participants in the control group used alcohol | 58.6% of exposed group participants used alcohol (56.6 to 60.8%) | **OR 1.48** (1.36 to 1.62) | 383,670 (9 observational studies) | ⊕⊕⊝⊝ **Low**  ^a,b,c,d,e^ | Frequent social media use increases adolescent alcohol use.  **Absolute effect calculated from Riehm 2021.**^122^ |
| Drug use | 17.0% of participants in the control group used drugs | 20.8% of exposed group participants used drugs (17.7 to 24.2%) | **OR 1.28** (1.05 to 1.56) | 117,645 (6 observational studies) | ⊕⊝⊝⊝ **Very low**  ^f,g,h^ | Downgraded for RoB.  Frequent social media use may increase adolescent drug use.  **Absolute effect calculated from Whitehill 2020.**^152^ |
| Tobacco use | 12.1% of participants in the control group used tobacco | 19.6% of exposed group participants used tobacco (16.6 to 23.1%) | **OR 1.78** (1.45 to 2.19) | 424,326 (8 observational studies) | ⊕⊝⊝⊝ **Very low**  ^i,j,k^ | Downgraded for RoB and inconsistency.  Frequent social media use may increase adolescent tobacco use.  **Absolute effect calculated from Vazquez-Nava 2020.**^148^ |
| Electronic nicotine delivery system use | 66.7% of studies demonstrated a harmful effect of social media use on adolescent use of electronic nicotine delivery systems (95% CI 20.8 to 93.9%) | | | 18,047 (3 observational studies) | ⊕⊕⊝⊝ **Very low**  ^l,m^ | Downgraded for RoB and imprecision.  Frequent social media use increases adolescent use of ENDS. |
| Sexual risk behaviour | 37.0% of participants in the control group engaged in sexual risk behaviours | 51.1% of exposed group participants engaged in sexual risk behaviours (46.6 to 55.5%) | **OR 1.78** (1.49 to 2.13) | 47,325 (10 observational studies) | ⊕⊝⊝⊝ **Very low**  ^n,o,p,q,r^ | Downgraded for RoB and publication bias.  Frequent social media use may increase in adolescent sexual risk behaviours.  **Absolute effect calculated from Self-Brown 2018**.^133^ |
| Gambling | 21.4% of participants in the control group engaged in gambling | 43.6% of exposed group participants engaged in gambling (35.7 to 52.0%) | **OR 2.84** (2.04 to 3.97) | 26,537 (5 observational studies) | ⊕⊝⊝⊝ **Very low**  ^s,t,u^ | Downgraded for RoB.  Frequent social media use may increase adolescent gambling.  **Absolute effect calculated from King 2012.**^92^ |

| **Outcome** | **Anticipated absolute effects^a^ (95% CI)** | | **Relative effect (95% CI)** | **No. of participants**  **(studies)** | **Certainty of the evidence** | **Comments** |
| --- | --- | --- | --- | --- | --- | --- |
|  | **Risk with infrequent SM use** | **Risk with frequent SM use** |  |  |  |  |
| Multiple risk behaviours | 41.3% of participants in the control group engaged in multiple risk behaviours | 55.2% of exposed group participants engaged in multiple risk behaviours (47.8 to 62.3%) | **OR 1.75** (1.30 to 2.35) | 43,571 (2 observational studies) | ⊕⊝⊝⊝ **Very low**  ^v,x,y^ | Downgraded for RoB and inconsistency.  Frequent social media use may increase adolescent engagement in multiple risk behaviours.  **Absolute effect calculated from Beebe 2005.**^38^ |
| **GRADE Working Group grades of evidence** **High certainty** ⊕⊕⊕⊕**:** We are very confident that the true effect lies close to that of the estimate of the effect **Moderate certainty** ⊕⊕⊕⊝**:** We are moderately confident in the effect estimate: The true effect is likely to be close to the estimate of the effect, but there is a possibility that it is substantially different **Low certainty** ⊕⊕⊝⊝**:** Our confidence in the effect estimate is limited: The true effect may be substantially different from the estimate of the effect **Very low certainty** ⊕⊝⊝⊝**:** We have very little confidence in the effect estimate: The true effect is likely to be substantially different from the estimate of effect | | | | | | |
| **Explanations for upgrading/downgrading the evidence**  ^a^ Not downgraded for RoB, as most studies were low RoB, assessed exposure and outcome via validated tools, and no difference in effect size in high RoB studies was observed on stratification.  ^b^ Not downgraded for inconsistency as heterogeneity was moderate (I^2^ = 40.5%) and all 95% confidence intervals overlapped.  ^c^ Not downgraded for indirectness as only 1/9 studies assessed text messaging as well as social media use.  ^d^ Not downgraded for imprecision as 95% confidence interval did not cross the null effect, was narrow and did not include appreciable harm or benefit.  ^e^ Unable to assess publication bias via a funnel plot due to insufficient data, however as a systematic search was conducted the chance of publication bias is reduced.  ^f^ Downgraded for RoB as half of the studies were rated high RoB, only one study assessed exposure and outcome via validated tools, and all but one study failed to adjust for critical confounding domains.  ^g^ Not downgraded for indirectness as majority of included studies specifically assessed social media use.  ^h^ Not downgraded for imprecision as 95% confidence interval did not cross the null effect, was narrow and did not include appreciable harm or benefit.  ^i^ Downgraded for RoB, as although majority of included studies were low/moderate RoB, the contributing high RoB studies report notably larger effect sizes.  ^j^ Downgraded for serious inconsistency as heterogeneity of included studies was considerable (I^2^ = 95.7%).  ^k^ Not downgraded for imprecision as 95% confidence interval did not cross null effect, was narrow and did not include appreciable harm or benefit.  ^l^ Downgraded for RoB as all studies were moderate/high RoB, failed to adjust for critical confounding domains, and assessed exposure and outcome via non-validated tools.  ^m^ Downgraded for serious imprecision as wide 95% confidence interval suggests lack of confidence in estimate.  ^n^ Downgraded for RoB, as majority of studies were high RoB, failed to adjust for critical confounding domains, and assessed exposure and outcome via non-validated tools.  ^o^ Not downgraded for serious inconsistency as heterogeneity was reduced when stratification was performed by socioeconomic position, age, social media category and development status of study setting.  ^p^ Not downgraded for indirectness as only 1/10 studies assessed text messaging as well as social media use.  ^q^ Not downgraded for imprecision as 95% confidence interval did not cross null effect, was narrow and did not include appreciable harm or benefit.  ^r^ Downgraded for suspected publication bias given impression from asymmetric contour enhanced funnel plot.  ^s^ Downgraded for RoB as majority of studies were high RoB, failed to adjust for critical confounding domains, and assessed exposure via non-validated tools.  ^t^ Not downgraded for indirectness as no concerns were raised regarding population, intervention, comparator, direct comparisons, or outcome.  ^u^ Not downgraded for imprecision as 95% confidence interval does not cross null effect and does not include appreciable harm or benefit.  ^v^ Downgraded for RoB as all studies were high RoB, failed to adjust for critical confounding domains, and assessed exposure and outcome via non-validated tools.  ^w^ Downgraded for serious inconsistency as heterogeneity of included studies was considerable (I^2^ = 97.9%) and confidence intervals show no or minimal overlap.  ^x^ Not downgraded for imprecision as 95% confidence interval did not cross null effect, was narrow and did not include appreciable harm or benefit. | | | | | | |

**Legend:** ^a^ The risk in the intervention group (and its 95% confidence interval) is based on the assumed risk in the comparison group and the relative effect of the intervention (and its 95% CI). Abbreviations: CI = Confidence interval; OR = Odds ratio and RoB = Risk of bias.

##### Table B. Post-hoc analysis: summary of findings and certainty of evidence for unhealthy dietary behaviour (as per GRADE) with reasons for upgrading/downgrading of the evidence

| **Outcome** | **Effect direction** | **No. of participants**  **(studies)** | **Certainty of the evidence (GRADE)** | **Comments** |
| --- | --- | --- | --- | --- |
| Unhealthy dietary behaviour (effect direction) | All studies demonstrated a harmful effect of social media use on adolescent engagement in unhealthy dietary behaviours (51.0 to 100.0%) | 521 (4 randomised control trials) | ⊕⊕⊕⊝  **Moderate**  ^a,b^ | Downgraded for indirectness.  Exposure to health-risk behaviour content on social media increases adolescent engagement in unhealthy dietary behaviours. |
| **GRADE Working Group grades of evidence** **High certainty** ⊕⊕⊕⊕**:** We are very confident that the true effect lies close to that of the estimate of the effect **Moderate certainty** ⊕⊕⊕⊝**:** We are moderately confident in the effect estimate: The true effect is likely to be close to the estimate of the effect, but there is a possibility that it is substantially different **Low certainty** ⊕⊕⊝⊝**:** Our confidence in the effect estimate is limited: The true effect may be substantially different from the estimate of the effect **Very low certainty** ⊕⊝⊝⊝**:** We have very little confidence in the effect estimate: The true effect is likely to be substantially different from the estimate of effect | | | | |
| **Explanations for upgrading/downgrading the evidence**  ^a^ Not downgraded for RoB as all studies were RCT's, with two rated low RoB and two some concerns.  ^b^ Downgraded for serious indirectness of comparator, as two studies used a comparator group pertaining to healthy food exposure and the remaining two used a comparator group pertaining to exposure to non-food items. | | | | |

**Legend:** Abbreviations: RoB = Risk of bias.

### Appendix-19. PRISMA checklists

Table A. PRISMA 2020 checklist for systematic reviews and meta-analysis

| **Section and Topic** | **Item #** | **Checklist item** | **Location where item is reported** |
| --- | --- | --- | --- |
| **TITLE** | | |  |
| Title | 1 | Identify the report as a systematic review. | Manuscript: Title page  Manuscript: Abstract  Manuscript: Methods |
| **ABSTRACT** | | |  |
| Abstract | 2 | See the PRISMA 2020 for Abstracts checklist. | Manuscript: Abstract  Appendix-19: Table B. PRISMA checklists |
| **INTRODUCTION** | | |  |
| Rationale | 3 | Describe the rationale for the review in the context of existing knowledge. | Manuscript: Introduction |
| Objectives | 4 | Provide an explicit statement of the objective(s) or question(s) the review addresses. | Manuscript: Introduction |
| **METHODS** | | |  |
| Eligibility criteria | 5 | Specify the inclusion and exclusion criteria for the review and how studies were grouped for the syntheses. | Manuscript: Study inclusion and exclusion criteria  Appendix-5: Process of social media categorisation |
| Information sources | 6 | Specify all databases, registers, websites, organisations, reference lists and other sources searched or consulted to identify studies. Specify the date when each source was last searched or consulted. | Manuscript: Search methods for identification of studies  Manuscript: Figure-2 PRISMA study flow chart  Appendix-3: Details of search strategies conducted |
| Search strategy | 7 | Present the full search strategies for all databases, registers and websites, including any filters and limits used. | Manuscript: Search methods for identification of studies  Appendix-3: Details of search strategies conducted |
| Selection process | 8 | Specify the methods used to decide whether a study met the inclusion criteria of the review, including how many reviewers screened each record and each report retrieved, whether they worked independently, and if applicable, details of automation tools used in the process. | Manuscript: Study inclusion and exclusion criteria  Manuscript: Selection of studies  Manuscript: Data extraction and RoB assessment |
| Data collection process | 9 | Specify the methods used to collect data from reports, including how many reviewers collected data from each report, whether they worked independently, any processes for obtaining or confirming data from study investigators, and if applicable, details of automation tools used in the process. | Manuscript: Selection of studies  Manuscript: Data extraction and RoB assessment  Appendix-8: Data extraction form exemplar |
| Data items | 10a | List and define all outcomes for which data were sought. Specify whether all results that were compatible with each outcome domain in each study were sought (e.g. for all measures, time points, analyses), and if not, the methods used to decide which results to collect. | Manuscript: Study inclusion and exclusion criteria  Appendix-6: Included outcomes  Appendix-7: Meta-analyses and synthesis without meta-analysis (SWiM) decision rules |
|  | 10b | List and define all other variables for which data were sought (e.g. participant and intervention characteristics, funding sources). Describe any assumptions made about any missing or unclear information. | Manuscript: Study inclusion and exclusion criteria  Appendix-8: Data extraction form exemplar |
| Study risk of bias assessment | 11 | Specify the methods used to assess risk of bias in the included studies, including details of the tool(s) used, how many reviewers assessed each study and whether they worked independently, and if applicable, details of automation tools used in the process. | Manuscript: Data extraction and RoB assessment  Appendix-9: Newcastle-Ottawa Scale (NOS) risk of bias assessment |
| Effect measures | 12 | Specify for each outcome the effect measure(s) (e.g. risk ratio, mean difference) used in the synthesis or presentation of results. | Manuscript: Data synthesis  Appendix: Process for data transformations for meta-analysis |
| Synthesis methods | 13a | Describe the processes used to decide which studies were eligible for each synthesis (e.g. tabulating the study intervention characteristics and comparing against the planned groups for each synthesis (item #5)). | Manuscript: Data synthesis  Appendix-7: Meta-analyses and synthesis without meta-analysis (SWiM) decision rules |
|  | 13b | Describe any methods required to prepare the data for presentation or synthesis, such as handling of missing summary statistics, or data conversions. | Manuscript: Data synthesis  Appendix-7: Meta-analyses and synthesis without meta-analysis (SWiM) decision rules  Appendix-10: Process for data transformations for meta-analysis |
|  | 13c | Describe any methods used to tabulate or visually display results of individual studies and syntheses. | Manuscript: Data synthesis  Appendix-7: Meta-analyses and synthesis without meta-analysis (SWiM) decision rules |
|  | 13d | Describe any methods used to synthesize results and provide a rationale for the choice(s). If meta-analysis was performed, describe the model(s), method(s) to identify the presence and extent of statistical heterogeneity, and software package(s) used. | Manuscript: Data synthesis  Appendix-15: Meta-analyses, meta-regression, subgroup, and sensitivity analyses |
|  | 13e | Describe any methods used to explore possible causes of heterogeneity among study results (e.g. subgroup analysis, meta-regression). | Manuscript: Data synthesis |
|  | 13f | Describe any sensitivity analyses conducted to assess robustness of the synthesized results. | Manuscript: Data synthesis |
| Reporting bias assessment | 14 | Describe any methods used to assess risk of bias due to missing results in a synthesis (arising from reporting biases). | Manuscript: Data synthesis |
| Certainty assessment | 15 | Describe any methods used to assess certainty (or confidence) in the body of evidence for an outcome. | Manuscript: Certainty of the evidence |
| **RESULTS** | | |  |
| Study selection | 16a | Describe the results of the search and selection process, from the number of records identified in the search to the number of studies included in the review, ideally using a flow diagram. | Manuscript: Results  Manuscript: Figure-2 PRISMA study flow diagram |
|  | 16b | Cite studies that might appear to meet the inclusion criteria, but which were excluded, and explain why they were excluded. | Appendix-12: Characteristics of excluded studies |
| Study characteristics | 17 | Cite each included study and present its characteristics. | Manuscript: Results  Appendix-11: Characteristics of included studies |
| Risk of bias in studies | 18 | Present assessments of risk of bias for each included study. | Manuscript: Results  Appendix-13: Risk of bias domain and overall grade for included datapoints and studies  Appendix-18: Summary of findings and certainty of the evidence |
| Results of individual studies | 19 | For all outcomes, present, for each study: (a) summary statistics for each group (where appropriate) and (b) an effect estimate and its precision (e.g. confidence/credible interval), ideally using structured tables or plots. | Manuscript: Results  Manuscript: Figures 4A-C  Manuscript: Figures 5A-D  Appendix-16: Synthesis without meta-analysis, meta-analyses, meta-regression, subgroup, and sensitivity analyses |
| Results of syntheses | 20a | For each synthesis, briefly summarise the characteristics and risk of bias among contributing studies. | Manuscript: Results  Appendix-13: Risk of bias domain and overall grade for included datapoints and studies  Appendix-18: Summary of findings and certainty of the evidence S13 Appendix-14: Social media measures reported in included studies |
|  | 20b | Present results of all statistical syntheses conducted. If meta-analysis was done, present for each the summary estimate and its precision (e.g. confidence/credible interval) and measures of statistical heterogeneity. If comparing groups, describe the direction of the effect. | Manuscript: Results  Manuscript: Figures 4A-C  Manuscript: Figures 5A-D  Appendix-16: Synthesis without meta-analysis, meta-analyses, meta-regression, subgroup, and sensitivity analyses |
|  | 20c | Present results of all investigations of possible causes of heterogeneity among study results. | Manuscript: Results  Appendix-16: Synthesis without meta-analysis, meta-analyses, meta-regression, subgroup, and sensitivity analyses  Appendix-17: Assessment of publication bias/small study effects |
|  | 20d | Present results of all sensitivity analyses conducted to assess the robustness of the synthesized results. | Manuscript: Results  Appendix-16: Synthesis without meta-analysis, meta-analyses, meta-regression, subgroup, and sensitivity analyses |
| Reporting biases | 21 | Present assessments of risk of bias due to missing results (arising from reporting biases) for each synthesis assessed. | Manuscript: Results  Appendix-13: Risk of bias domain and overall grade for included datapoints and studies  Appendix-18: Summary of findings and certainty of the evidence S13 |
| Certainty of evidence | 22 | Present assessments of certainty (or confidence) in the body of evidence for each outcome assessed. | Manuscript: Certainty of evidence  Appendix-18: Summary of findings and certainty of the evidence |
| **DISCUSSION** | | |  |
| Discussion | 23a | Provide a general interpretation of the results in the context of other evidence. | Manuscript: Discussion |
|  | 23b | Discuss any limitations of the evidence included in the review. | Manuscript: Discussion |
|  | 23c | Discuss any limitations of the review processes used. | Manuscript: Discussion |
|  | 23d | Discuss implications of the results for practice, policy, and future research. | Manuscript: Discussion |
| **OTHER INFORMATION** | | |  |
| Registration and protocol | 24a | Provide registration information for the review, including register name and registration number, or state that the review was not registered. | Manuscript: Methods  Appendix-20: Registered/published protocols |
|  | 24b | Indicate where the review protocol can be accessed, or state that a protocol was not prepared. | Manuscript: Methods  Appendix-20: Registered/published protocols |
|  | 24c | Describe and explain any amendments to information provided at registration or in the protocol. | Manuscript: Methods  Appendix-2: Deviations from protocol |
| Support | 25 | Describe sources of financial or non-financial support for the review, and the role of the funders or sponsors in the review. | Manuscript: Funding |
| Competing interests | 26 | Declare any competing interests of review authors. | Manuscript: Competing interests |
| Availability of data, code and other materials | 27 | Report which of the following are publicly available and where they can be found: template data collection forms; data extracted from included studies; data used for all analyses; analytic code; any other materials used in the review. | Manuscript: Data sharing |

Table B. PRISMA 2020 structured abstract checklist

| **Section and Topic** | **Item #** | **Checklist item** | **Reported (Yes/No)** |
| --- | --- | --- | --- |
| **TITLE** | | |  |
| Title | 1 | Identify the report as a systematic review. | Yes |
| **BACKGROUND** | | |  |
| Objectives | 2 | Provide an explicit statement of the main objective(s) or question(s) the review addresses. | Yes |
| **METHODS** | | |  |
| Eligibility criteria | 3 | Specify the inclusion and exclusion criteria for the review. | Yes |
| Information sources | 4 | Specify the information sources (e.g. databases, registers) used to identify studies and the date when each was last searched. | Yes |
| Risk of bias | 5 | Specify the methods used to assess risk of bias in the included studies. | Yes |
| Synthesis of results | 6 | Specify the methods used to present and synthesise results. | Yes |
| **RESULTS** | | |  |
| Included studies | 7 | Give the total number of included studies and participants and summarise relevant characteristics of studies. | Yes |
| Synthesis of results | 8 | Present results for main outcomes, preferably indicating the number of included studies and participants for each. If meta-analysis was done, report the summary estimate and confidence/credible interval. If comparing groups, indicate the direction of the effect (i.e. which group is favoured). | Yes |
| **DISCUSSION** | | |  |
| Limitations of evidence | 9 | Provide a brief summary of the limitations of the evidence included in the review (e.g. study risk of bias, inconsistency and imprecision). | Yes |
| Interpretation | 10 | Provide a general interpretation of the results and important implications. | Yes |
| **OTHER** | | |  |
| Funding | 11 | Specify the primary source of funding for the review. | Yes |
| Registration | 12 | Provide the register name and registration number. | Yes |

### Appendix-20. Registered/published protocols

**University of Glasgow published protocol**

https://www.gla.ac.uk/media/Media_718614_smxx.pdf

**PROSPERO registered and updated protocol**

PROSPERO ID: [CRD42020179766](https://www.crd.york.ac.uk/prospero/display_record.php?ID=CRD42020179766)

<https://www.crd.york.ac.uk/prospero/display_record.php?ID=CRD42020179766>
